# Discordant Evidence on Corticosteroids in Sepsis: A Meta-Research Study

**DOI:** 10.64898/2026.08.20.26360343

**Authors:** Stephanie Weibel, Hanna Düngfelder, Tamara Pscheidl, Manuel Krone, Patrick Meybohm

## Abstract

**Background:** Despite numerous randomized controlled trials (RCTs) and systematic reviews (SRs), current sepsis guidelines continue to issue only weak recommendations for corticosteroids. We examined the clinical scope, underlying study pools, and mortality conclusions of SRs evaluating corticosteroids for sepsis.

**Methods:** We conducted a meta-research study of SRs on corticosteroids in sepsis (2015– 2025), extracting SR characteristics, mortality results, and included RCTs. Study-pool overlap was assessed using an SR×RCT inclusion matrix, Jaccard similarity (J), and hierarchical clustering. SRs and RCTs were classified according to standardized Population, Intervention, Comparison, Outcome (PICO) profiles. We explored discordance in short-term mortality conclusions among clinically comparable SRs and potential associations with study-pool composition, target populations, and methodological characteristics.

**Results:** Forty-two SRs including 121 unique RCTs were identified. More than half of pairwise SR comparisons shared no RCTs, and only three pairs showed high overlap (J>0.8). SRs addressing similar intervention and target population profiles frequently relied on different study pools. Among 38 SRs with short-term mortality meta-analyses, 15 (39%) reported benefit and 23 (61%) no evidence of effect. Discordance occurred exclusively among SRs evaluating broad, non-specific corticosteroid strategies; conclusions were consistent for hydrocortisone plus fludrocortisone (benefit) and hydrocortisone, ascorbic acid, and thiamine (no evidence of effect). SRs including sepsis ± shock populations more frequently reported benefit than those restricted to septic shock (62% vs 22%), although estimates were imprecise. No single methodological or clinical factor consistently explained discordance.

**Conclusions:** SRs addressing apparently similar clinical questions frequently synthesized different underlying evidence bases and reported discordant conclusions. Guideline developers should therefore consider not only methodological quality and reported PICO, but also whether the RCTs included in an SR adequately represent the intended clinical question. Clinically coherent evidence syntheses may improve the interpretability of pooled treatment effects and support more targeted corticosteroid therapy in sepsis.

**Key messages:**

- SRs addressing clinically similar questions frequently relied on different and only partially overlapping sets of RCTs.
- Discordant mortality conclusions occurred exclusively among SRs evaluating non-specific broad corticosteroid strategies, whereas SRs of hydrocortisone plus fludrocortisone and hydrocortisone, ascorbic acid, and thiamine reported consistent findings.
- Differences in target populations (sepsis ± shock vs septic shock) may contribute to discordant findings, although no single methodological or clinical factor consistently explained the observed discordance.
- Clinical guideline developers should evaluate the clinical scope and underlying evidence base of SRs, not only their methodological quality, when formulating recommendations.

## Background

Sepsis is a life-threatening syndrome caused by a dysregulated host response to infection and remains a leading cause of mortality worldwide [1–4]. Clinical management relies on timely evidence-based decisions, making high-quality systematic reviews (SRs) and clinical practice guidelines essential for informing treatment recommendations [1, 2, 5].

Corticosteroids are among the most widely studied adjunctive therapies in sepsis [6]. Over the past six decades, more than 100 randomized controlled trials (RCTs) and numerous SRs have evaluated their effects on mortality and other patient-important outcomes, yielding conflicting conclusions [7–14]. Nevertheless, although the SR underpinning current international guideline recommendations concluded that corticosteroids reduce short-term mortality in sepsis patients with moderate-certainty evidence [11], these guidelines continue to issue only weak or conditional recommendations for corticosteroid therapy in septic shock [15, 16].

Several factors may contribute to the cautious guideline recommendations. First, the balance between desirable and undesirable effects, including potential adverse events associated with corticosteroid therapy, may influence the strength of recommendations despite moderate-certainty evidence for reduced short-term mortality [11]. Second, substantial clinical heterogeneity exists across the available evidence, with differences in corticosteroid intervention strategies (drug type, dose, timing, duration, and combination regimens) as well as in the included sepsis populations, both of which may contribute to variation in treatment effects across RCTs and SRs [6]. Third, the rapidly increasing number of published SRs has raised concerns regarding their methodological quality, consistency, and clinical applicability across medical disciplines [17]. Even when addressing apparently similar clinical questions, SRs may differ in their exact Population, Intervention, Comparison, and Outcome (PICO) definitions, eligibility criteria, search strategies, and synthesis methods, potentially resulting in multiple overlapping SRs with conflicting results that are difficult to compare and prioritize for guideline development [18].

Understanding how SRs addressing similar clinical questions differ in their underlying evidence base and conclusions is essential for interpreting the available evidence and informing future evidence syntheses and guideline development. Accordingly, this meta-research study aimed to characterize the evidence landscape of SRs evaluating corticosteroids for sepsis, including their study pool composition, clinical scope, quality, and mortality conclusions, and to explore potential sources of discordance.

## Methods

### Study design, eligibility, and study selection

We conducted a meta-research study of SRs evaluating corticosteroids in sepsis according to a reporting guideline and a prospectively published protocol [19, 20]. The unit of analysis was the SR. We included SRs of patients with suspected or confirmed sepsis or septic shock investigating corticosteroid therapy as a primary intervention, either alone or in combination with other agents. All published updates of eligible SRs were retained.

We searched MEDLINE (via PubMed) and Epistemonikos from January 2015 to 20 June 2025. Two reviewers independently screened titles/abstracts and full texts, with disagreements resolved by discussion with a third reviewer. Detailed eligibility criteria and the full search strategy are provided in the Supplementary Methods and the protocol [20].

### Data extraction and quality assessment of systematic reviews

We extracted SR characteristics, mortality data, certainty assessments, PICO eligibility criteria, and all included RCTs, including whether individual RCTs contributed to mortality meta-analyses. For RCTs with available full-text publications, we additionally extracted eligibility criteria and baseline characteristics relevant to sepsis status. Data were extracted by one reviewer and independently cross-checked by a second reviewer.

Reporting quality and methodological rigor were assessed using PRISMA 2020 [21] and AMSTAR 2 [22], respectively, as described in the protocol [20]. Assessments were performed by one reviewer and verified by a second reviewer.

### Study-pool similarity and clustering

We constructed an SR×RCT inclusion matrix and quantified pairwise study-pool overlap using the Jaccard similarity coefficient (J). Hierarchical clustering using average linkage (UPGMA) was applied to Jaccard distance (1−J), with clusters defined using a distance cut-off of 0.5.

### PICO classification of systematic reviews

We derived post hoc standardized PICO profiles from the reported SR eligibility criteria using multidimensional binary coding. Domains included age, target population, intervention strategy, corticosteroid type, dose, timing/schedule, and comparator. Primary analyses focused on intervention strategy (e.g., non-specific broad corticosteroids, hydrocortisone plus fludrocortisone [HC+FC], and hydrocortisone, ascorbic acid, and thiamine [HAT]) and target population (particularly sepsis ± shock vs septic shock only). Detailed coding definitions are provided in the Supplementary Methods.

### Classification of RCT populations

We first classified RCTs with available full-text publications according to their reported target population based on the original eligibility criteria (e.g. sepsis ± shock, sepsis without shock, septic shock only, or other populations). We then performed an exploratory post hoc classification of RCTs with available full-text publications according to compatibility with contemporary consensus definitions (Sepsis-3 for adults and Phoenix Sepsis Score criteria for pediatric populations [23, 24]). Using reported eligibility criteria and baseline characteristics, RCTs were classified as compatible, likely compatible, unclear, likely incompatible, or incompatible. Classification was based exclusively on aggregate information reported in the original publications; no patient-level reclassification was performed. Detailed classification criteria are provided in the Supplementary Methods. Because this analysis was not prespecified, all analyses based on this classification were considered exploratory.

### PICO–study-pool alignment and mortality discordance

We linked PICO profiles to study-pool clusters to assess whether SRs addressing similar clinical questions relied on similar underlying RCT evidence bases. Discordance was defined as differences in short-term mortality conclusions among clinically comparable SRs, categorized as benefit, no evidence of effect, or harm according to statistical significance.

Potential sources of discordance were explored according to study-pool composition and overlap, population definitions, inclusion of likely incompatible or incompatible RCTs, methodological characteristics, certainty of evidence (GRADE), methodological confidence (AMSTAR 2), and search date. Categorical variables were compared using Fisher’s exact test and continuous variables using the Wilcoxon rank-sum test. All analyses were exploratory.

### Sensitivity analysis

We repeated the short-term mortality meta-analyses of Annane et al. [7], the SR with the largest study pool, and Pitre et al. [11], which informed current international guideline recommendations [15, 16], after progressively restricting their RCT evidence bases according to the post hoc sepsis compatibility classification. Analyses included (1) compatible RCTs only, (2) compatible and likely compatible RCTs, and (3) compatible, likely compatible, and unclear RCTs. RCTs classified as likely incompatible or incompatible were additionally analyzed separately. Meta-analyses used the original statistical methods of the respective SRs. Further details are provided in the Supplementary Methods.

### Statistical analysis

Analyses were performed in R. Given their exploratory nature and the limited number of SRs, results were interpreted primarily according to patterns and consistency rather than formal hypothesis testing.

## Results

### Identification and characteristics of SRs

The systematic search identified 6,856 records, of which 42 SRs investigating corticosteroids in sepsis published between 2015 and 2025 were included (Supplementary Figure S1; Supplementary Tables S1-S3).

Three SRs were Cochrane reviews, which were updates of the same review (Annane 2015 [25], 2019 [26], and 2025 [7]). The number of included RCTs per SR varied substantially (range 0–87; median 10, interquartile range (IQR) 7–21) (Supplementary Table S4). The most comprehensive SR was Annane 2025 (87 RCTs) [7]; one SR [27] did not include any RCTs but only non-randomized studies.

Reporting was limited, with 34 of 42 SRs (81%) reporting fewer than 50% of the applicable PRISMA 2020 items. Methodological confidence according to AMSTAR 2 was rated as critically low in 38 of 42 SRs (90%) (Supplementary Table S5-S6; Supplementary Figures S2-S3).

### Structure of the evidence base and study pool similarity

Across the 42 SRs, 121 unique included RCTs were identified which were published between 1963 and 2023 (Supplementary Table S7). Three landmark RCTs (Annane 2002 [8], Annane 2018 [28], Sprung 2008 [13]) were included in about half of all SRs, whereas 28 RCTs were included in only a single SR, indicating substantial variability in study selection (Supplementary Table S8, Supplementary Figure S4).

Pairwise comparison of study pools across 861 SR pairs indicated substantial fragmentation of the evidence base. More than half of SR pairs shared no RCTs, whereas only three pairs showed high overlap in included studies (J > 0.8; [29, 30], [11, 12], [31, 32]) (**Figure 1**, Supplementary Table S9).

**Figure 1.**
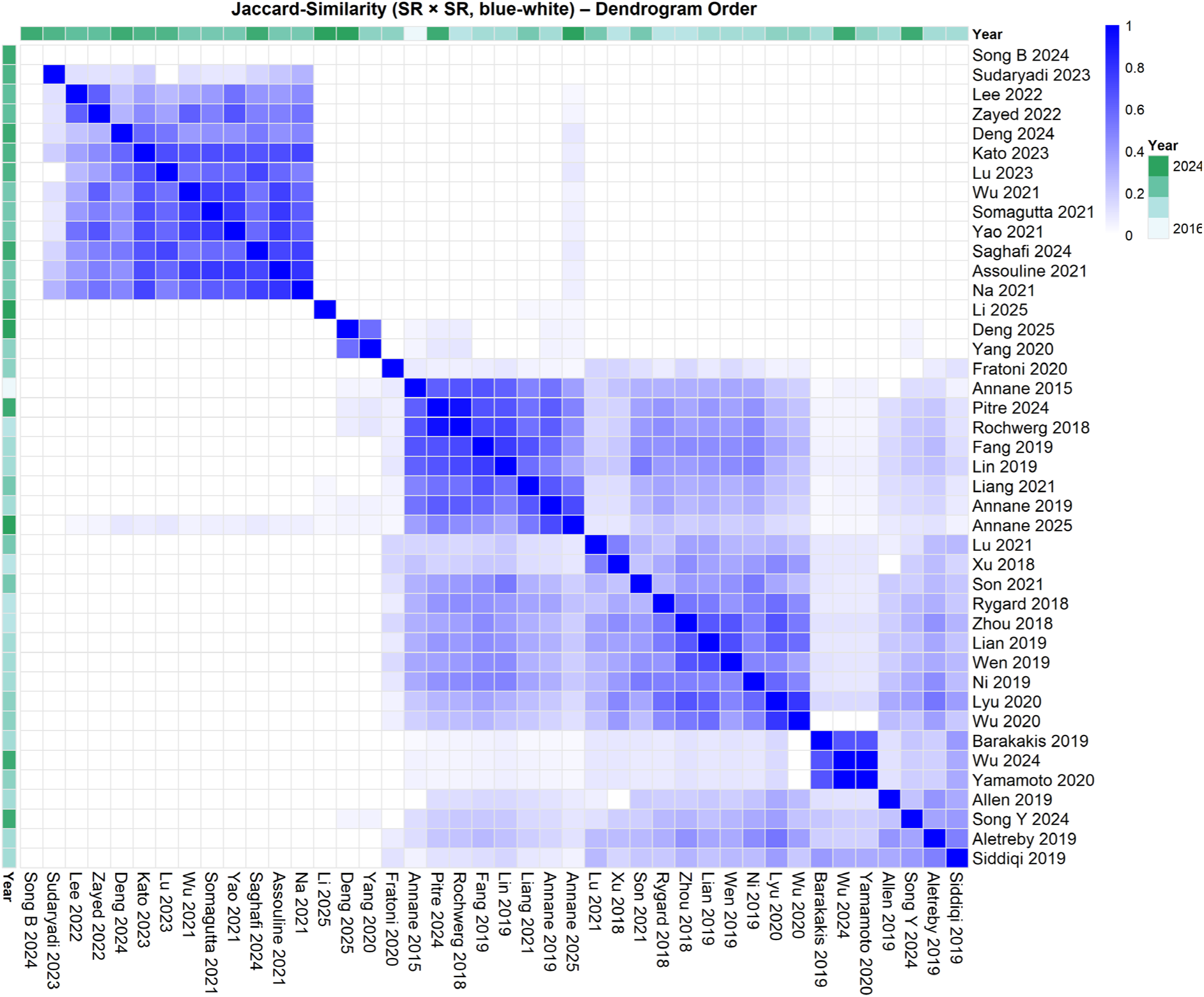
Pairwise similarity of systematic reviews based on included RCTs. Heatmap shows the pairwise Jaccard similarity between systematic reviews (SRs) based on study pool similarity, calculated from the binary SR×RCT inclusion matrix. Color intensity (white to blue) indicates increasing overlap (0 = no shared RCTs; 1 = identical study pool). SRs are displayed in dendrogram order derived from hierarchical clustering using average linkage on the distance metric 1−J. Publication year of each SR is shown as an annotation bar.

Cluster analysis identified 17 clusters of SRs based on study pool similarity (**Figure 2**, Supplementary Table S10), including four clusters (C) with more than two SRs with moderate to high study pool overlap (J ≥ 0.5): (1) C3: rank 1, n=8, [7, 10–12, 25, 26, 33, 34]; (2) C4: rank 2, n=8, [31, 32, 35–40]; (3) C11: rank 3, n=6, [41–46]; (4) C5: rank 4, n=3, [29, 30, 47].

**Figure 2.**
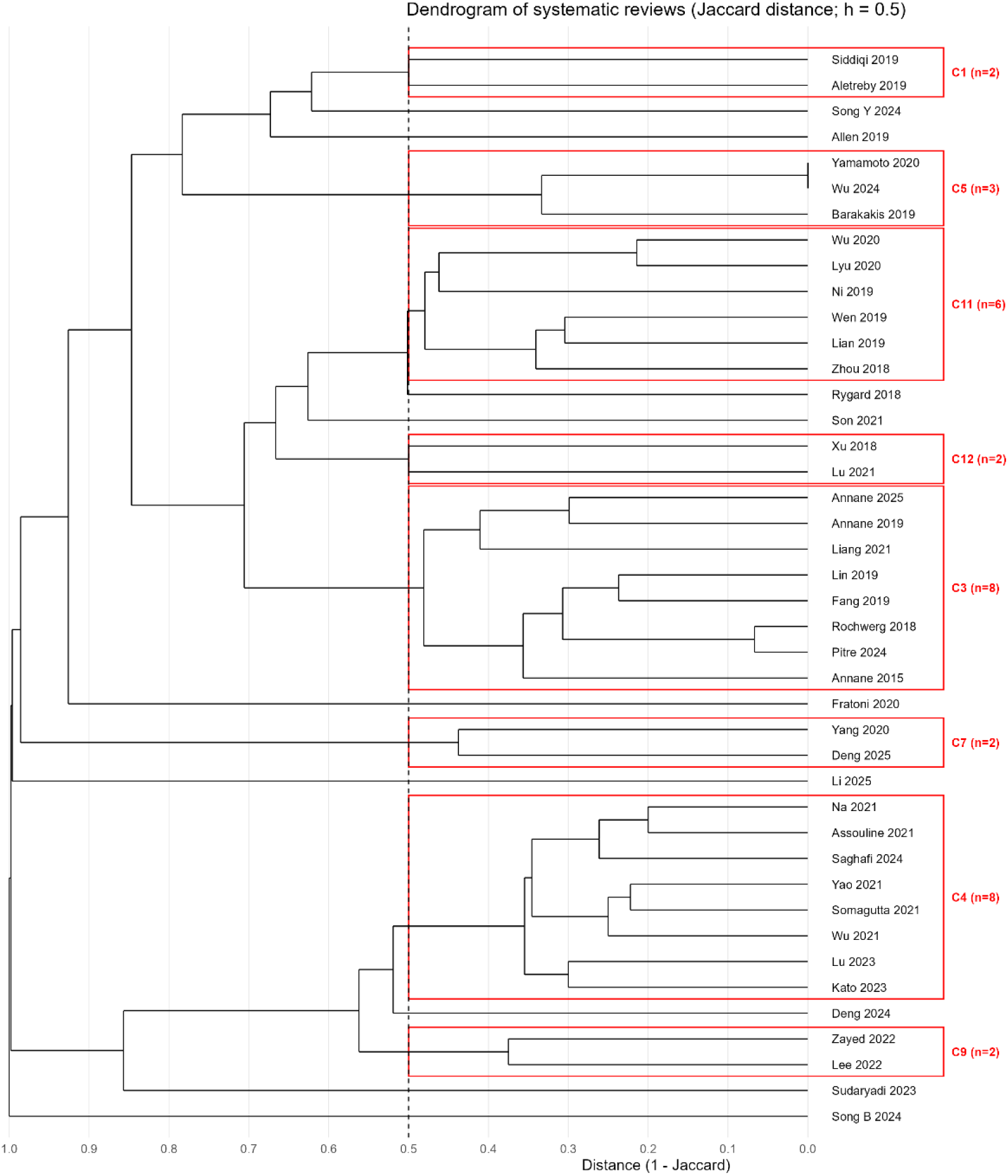
Hierarchical clustering of systematic reviews based on study pool overlap. Hierarchical clustering of systematic reviews (SRs) based on the similarity of included randomized controlled trials (RCTs). Pairwise similarity between SRs was quantified using the Jaccard coefficient, and clustering was performed using average linkage based on Jaccard distance (1 − J). The horizontal axis represents Jaccard distance, with lower values indicating greater overlap in included RCTs. Clusters were defined using a cut-off at a Jaccard distance of 0.5 (h = 0.5), corresponding to moderate similarity in study pool composition. The dashed vertical line indicates the cut-off used to define clusters. Red rectangles highlight clusters containing two or more SRs; clusters with a single SR are not outlined. Cluster labels (C1– C17) indicate cluster identity and number of SRs per cluster. SR names are displayed on the right.

### PICO profiles of SRs and included RCTs

To place study pool heterogeneity in a clinical context, SRs were classified according to standardized PICO profiles, focusing on intervention strategy and target population based on the reported eligibility criteria (Supplementary Table S11). Three main intervention profiles were identified: HC+FC combination therapy (3/42, 7%), HAT combination therapy (11/42, 26%), and non-specific broad corticosteroid strategies (25/42, 60%); one SR investigated dexamethasone only and two others investigated different timing or schedule of interventions. Population definitions also varied across groups. Within the non-specific broad group, SRs were split between populations including sepsis ± shock (15/25) and septic shock-only (10/25). In contrast, HAT-focused SRs predominantly included sepsis ± shock populations (9/11), whereas all HC+FC SRs were restricted to septic shock.

Of the 121 unique RCTs included across the 42 SRs, 104 (86%) had an available English full-text publication and could be classified. Based on the originally reported trial eligibility criteria, 22 of these 104 RCTs (21%) investigated populations other than sepsis or septic shock (Supplementary Table S12). Post hoc classification by us according to contemporary consensus definitions identified 41 (39%) RCTs as compatible, 11 (11%) as likely compatible, 34 (33%) as unclear, 15 (14%) as likely incompatible, and three (3%) as incompatible (Supplementary Table S13-14). RCTs classified as likely incompatible or incompatible (18/104, 17%) were included in 14 of 42 SRs (33%) and were concentrated in study-pool cluster 3, where all eight SRs included at least one such RCT (Supplementary Table S15-S16). Annane et al. [7] included the largest number of likely incompatible or incompatible RCTs (16/18, 89%); seven of these RCTs were exclusive to the Annane reviews (2019/2025, [7, 26]) (Supplementary Table S16).

### Alignment between PICO profiles and study pool composition

To examine whether SRs addressing similar clinical questions relied on similar evidence bases, we linked clinically defined intervention and target population profiles with study-pool clusters (**Table 1**, Supplementary Table S17).

**Table 1.**
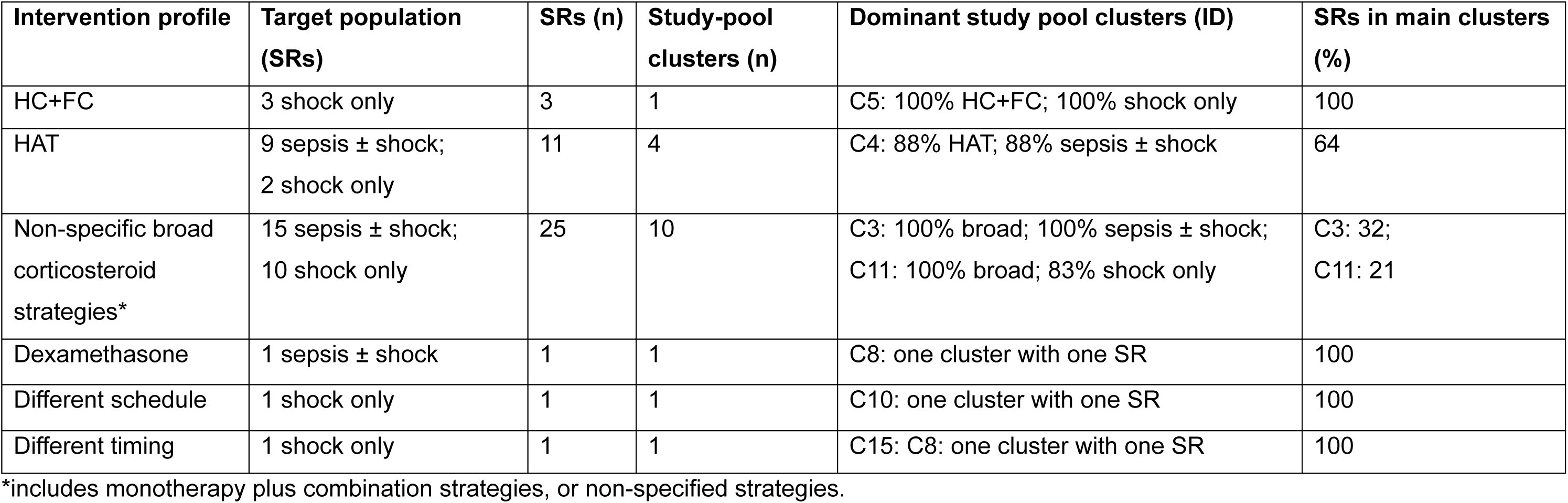
Distribution of systematic reviews across study-pool clusters according to intervention and target population profiles. Systematic reviews (SRs) were grouped according to corticosteroid intervention strategy. For each intervention profile, the table summarizes the target population profile of SRs, the number of SRs, the number of distinct study-pool clusters, the dominant study-pool cluster(s), and the proportion of SRs represented within these cluster(s). Percentages reported for the dominant study-pool cluster(s) indicate the proportion of SRs within each cluster sharing the same intervention profile and target population profile, respectively. Abbreviations: HC+FC, hydrocortisone plus fludrocortisone; HAT, hydrocortisone–ascorbic acid–thiamine; outcome; SR, systematic review.

| Intervention profile | Target population (SRs) | SRs (n) | Study-pool clusters (n) | Dominant study pool clusters (ID) | SRs in main clusters (%) |
| --- | --- | --- | --- | --- | --- |
| HC+FC | 3 shock only | 3 | 1 | C5: 100% HC+FC; 100% shock only | 100 |
| HAT | 9 sepsis ± shock;<br>2 shock only | 11 | 4 | C4: 88% HAT; 88% sepsis ± shock | 64 |
| Non-specific broad corticosteroid strategies* | 15 sepsis ± shock;<br>10 shock only | 25 | 10 | C3: 100% broad; 100% sepsis ± shock;<br>C11: 100% broad; 83% shock only | C3: 32;<br>C11: 21 |
| Dexamethasone | 1 sepsis ± shock | 1 | 1 | C8: one cluster with one SR | 100 |
| Different schedule | 1 shock only | 1 | 1 | C10: one cluster with one SR | 100 |
| Different timing | 1 shock only | 1 | 1 | C15: C8: one cluster with one SR | 100 |

\*includes monotherapy plus combination strategies, or non-specified strategies.

From a PICO-centred perspective, SRs evaluating non-specific broad corticosteroid strategies frequently relied on different sets of RCTs despite addressing similar clinical questions (**Table 1**, Supplementary Table S17). The 25 SRs evaluating non-specific broad corticosteroid strategies were distributed across 10 study-pool clusters, reflecting substantial fragmentation of the underlying RCT evidence. Two dominant clusters emerged, representing SRs of sepsis ± shock (C3; 8/25, 32%) and septic shock only (C11; 6/25, 24%), while the remaining SRs were dispersed across eight additional clusters. In contrast, all HC+FC SRs (3/3) were contained within a single study-pool cluster (C5), and most HAT SRs (7/11, 64%) were represented in one dominant cluster (C4), reflecting more similar study-pool composition among SRs addressing these defined intervention strategies.

From a cluster-centred perspective, the dominant clusters were clinically homogeneous with respect to both intervention strategy and target population (**Table 1**, Supplementary Table S17). Several smaller clusters reflected more specific clinical questions, including pediatric populations with the non-specific broad group (C7), dexamethasone-specific interventions (C8), or analyses restricted by corticosteroid timing or treatment schedule (C10 and C15). However, many other clusters addressed comparable intervention and target population profiles but remained separated from dominant clusters because they included different sets of RCTs (Supplementary Table S17), indicating substantial fragmentation of the underlying evidence base despite similar clinical questions.

### Discordance of mortality results among SRs

To assess discordance in mortality outcomes, we examined short-term mortality results across SRs. Among 42 SRs, 38 performed a meta-analysis of short-term mortality. Of these, 15 (39%) reported a mortality benefit and 23 (61%) reported no evidence of effect; no SR reported harm (**Figure 3**, Supplementary Table S18). Discordant conclusions occurred exclusively among SRs addressing non-specific broad corticosteroid strategies, whereas HAT-focused SRs consistently reported no evidence of effect and HC+FC-focused SRs consistently reported benefit (**Figure 3**). GRADE assessments were inconsistently reported and showed no clear pattern of evidence certainty across intervention strategies (Supplementary Table S18).

**Figure 3.**
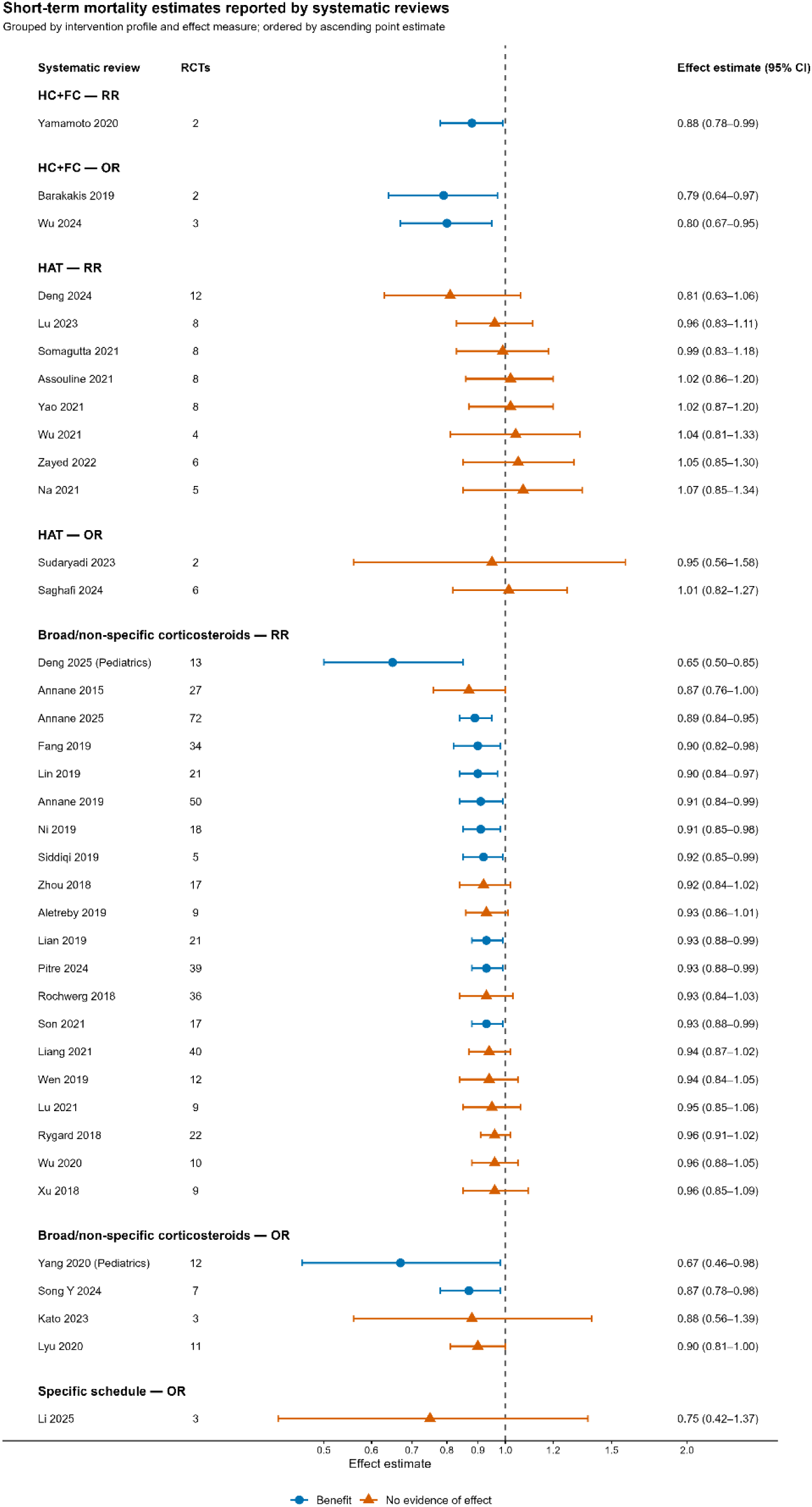
Short-term mortality effect estimates reported by systematic reviews of corticosteroids for sepsis. Systematic reviews (SRs) are grouped according to corticosteroid intervention strategy (hydrocortisone plus fludrocortisone (HC+FC), hydrocortisone, ascorbic acid, and thiamine (HAT), broad/non-specific corticosteroid strategies, and specific treatment schedule) and effect measure (risk ratio (RR) or odds ratio (OR)). Within each subgroup, SRs are ordered by ascending point estimate. Points represent the reported pooled effect estimates and horizontal lines the corresponding 95% confidence intervals (CI). Blue circles indicate a statistically significant mortality benefit, whereas orange triangles indicate no evidence of effect. Numbers on the left denote the number of randomized controlled trials (RCTs) contributing to each mortality meta-analysis. Dexamethasone-specific and treatment-timing-specific reviews did not report a short-term mortality meta-analysis and are therefore not shown.

### Sources of discordant mortality results

To explore sources for discordant mortality results, we analyzed all SRs addressing non-specific broad corticosteroid strategies in adults or mixed age populations excluding dexamethasone- and timing/schedule-specific SRs (n=23; 22 with meta-analysis). Mortality conclusions were nearly evenly split between benefit and no evidence of effect (10/22 vs 12/22) (**Figure 3**, Supplementary Table S18).

Study pool overlap across SRs was generally low (Supplementary Table S19-20). However, discordant conclusions were also observed among SRs with highly overlapping study pools. For example, Pitre 2024 [11] and Rochwerg 2018 [12] shared 36 of 39 RCTs (J=0.92) yet reached opposite conclusions regarding mortality (benefit vs no evidence of effect). No individual RCT was consistently associated with either direction of effect (Supplementary Table S21).

Exploratory analyses indicated that SRs reporting benefit included more RCTs compared to SRs reporting no evidence of effect (median 21, IQR 17–38 vs 12, 9–23). SRs including sepsis ± shock populations more frequently reported benefit than SRs restricted to septic shock (62% vs 22%), although estimates were imprecise (OR 5.15, 95% CI 0.62–70.89; p = 0.10; Supplementary Table S22). SRs reporting benefit more often included at least one RCT classified as likely incompatible or incompatible (50.0% vs 33.3%), although this association was imprecise (OR 1.94, 95% CI 0.26–15.60; p = 0.67; Supplementary Table S23). Mortality meta-analyses including at least one such RCT also comprised more RCTs overall (median 36, IQR 27–40 vs 10, 9–17). No consistent associations were observed for mortality timepoint, meta-analytic model, effect measure, GRADE certainty, AMSTAR 2 rating, and search date (Supplementary Tables S24).

### Sensitivity analyses of the Annane 2025 and Pitre 2024 meta-analyses

To explore the influence of RCTs investigating populations not fully compatible with contemporary sepsis definitions, we repeated the short-term mortality meta-analyses of the two most influential SRs after progressively restricting the evidence base according to the post hoc trial classification ([7, 11], **Figure 4 and 5**, Supplementary Tables S25-26). In both SRs, exclusion of RCTs classified as likely incompatible or incompatible resulted in only modest attenuation of the pooled treatment effect. In Annane 2025, the pooled RR changed from 0.90 (95% CI 0.84–0.96) to 0.92 (95% CI 0.86–0.99), while in Pitre 2024 the estimate changed from 0.93 (95% CI 0.88–0.99) to 0.94 (95% CI 0.88–1.01). In contrast, RCTs classified as likely incompatible or incompatible showed a larger treatment effects in both datasets (Annane: RR 0.71, 95% CI 0.55–0.93; Pitre: RR 0.69, 95% CI 0.42–1.13), although the latter analysis was based on only four RCTs.

**Figure 4.**
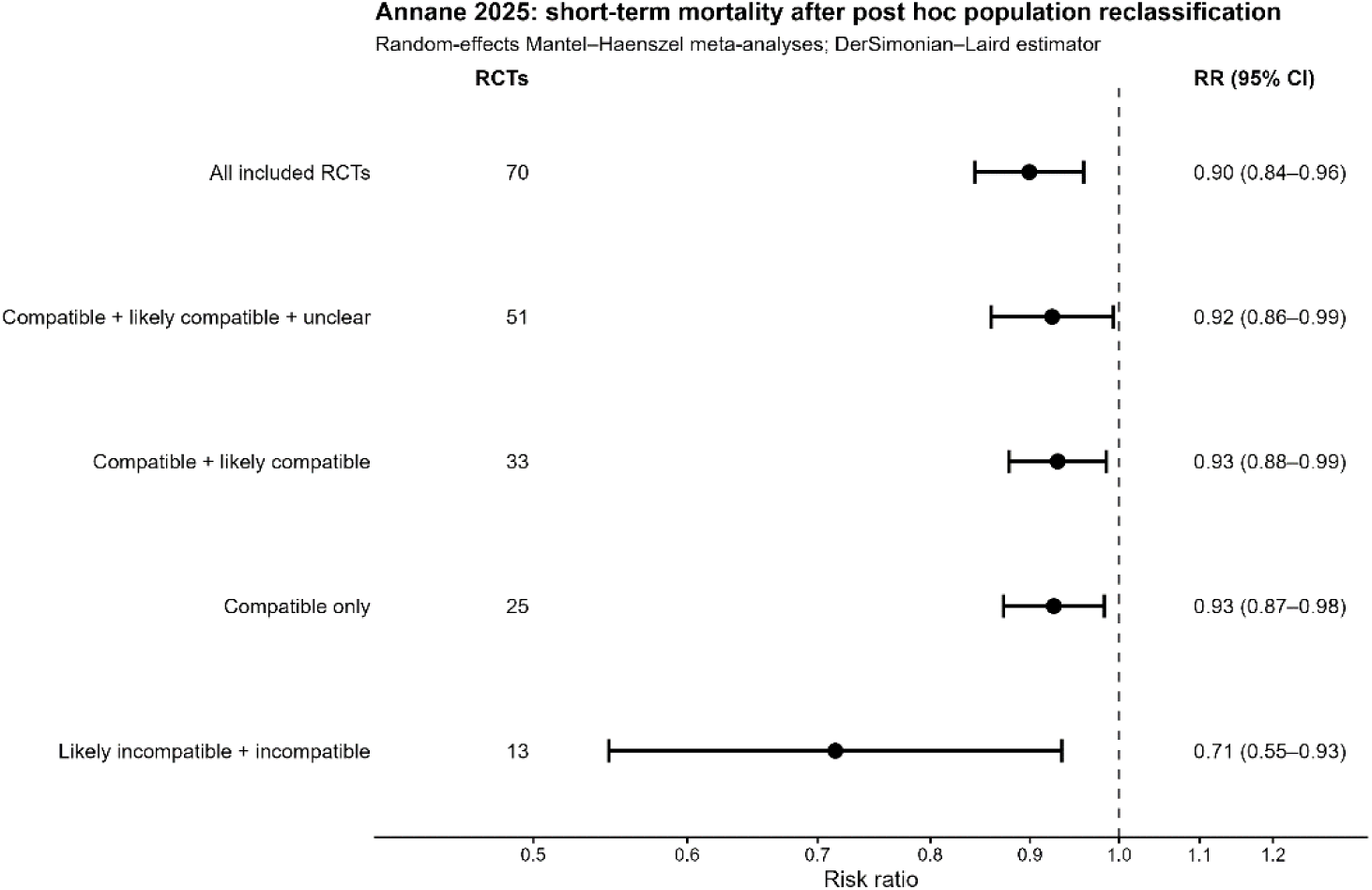
Sensitivity analysis of short-term mortality meta-analysis from Annane et al. Points represent pooled random-effects risk ratios (RR) and horizontal lines the corresponding 95% confidence intervals (CI). Sensitivity analyses progressively restricted the evidence base to randomized controlled trials (RCTs) classified as compatible, likely compatible, or unclear; compatible or likely compatible; and compatible only. The separate analysis of RCTs classified as likely incompatible or incompatible is also shown. Numbers indicate the RCTs contributing to each meta-analysis. Annane et al. included 71 RCTs in the short-term mortality analysis; one RCT with zero events in both treatment groups did not contribute to the pooled estimate. RCTs without an available full-text publication could not be included in analyses restricted by population compatibility.

**Figure 5.**
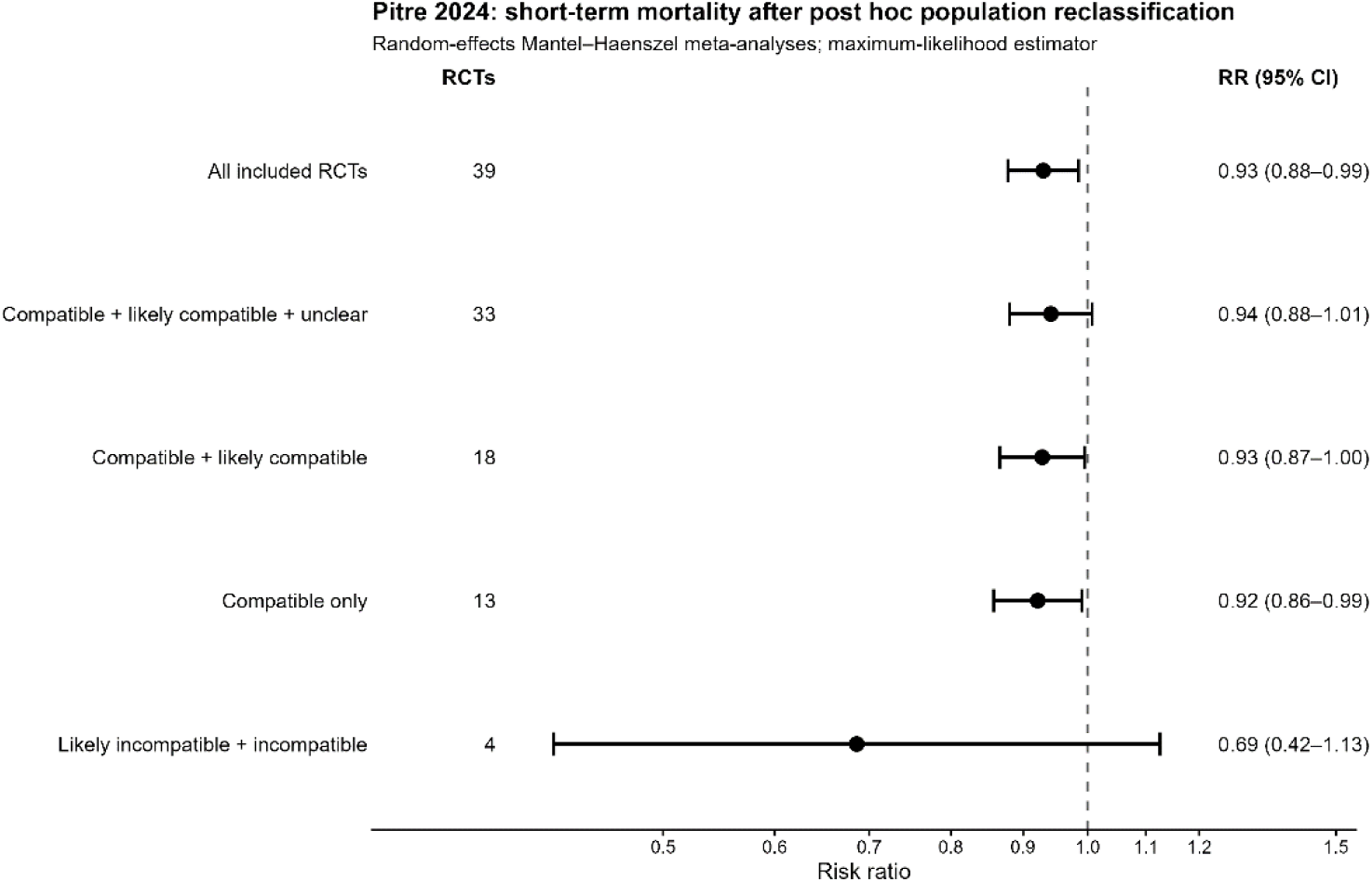
Sensitivity analysis of short-term mortality meta-analysis from Pitre et al. Points represent pooled random-effects risk ratios (RR) and horizontal lines the corresponding 95% confidence intervals (CI). Sensitivity analyses progressively restricted the evidence base to randomized controlled trails (RCTs) classified as compatible, likely compatible, or unclear; compatible or likely compatible; and compatible only. A separate analysis of RCTs classified as likely incompatible or incompatible is also shown. Numbers indicate the RCTs contributing to each meta-analysis. RCTs without an available full-text publication could not be included in analyses restricted by population compatibility.

## Discussion

### Principal findings

In this meta-research study, we found that SRs evaluating corticosteroids for sepsis frequently synthesized different underlying evidence bases despite addressing apparently similar clinical questions. Discordant mortality conclusions were confined to SRs evaluating non-specific broad corticosteroid strategies, whereas SRs of more narrowly defined interventions, such as HC+FC or HAT, consistently reached the same conclusions. Together, these findings suggest that the persistent uncertainty reflected in weak guideline recommendations may arise not only from clinical heterogeneity across populations and intervention strategies, but also from fragmentation of the underlying evidence base synthesized by apparently similar SRs.

### Implications for guideline development

The rapidly expanding number of overlapping SRs has been recognized as an increasing challenge for evidence users across medicine, raising concerns about redundancy, inconsistent conclusions, and difficulties in identifying the most appropriate evidence synthesis [17]. These findings have important implications for guideline development. Existing guidance recommends selecting the SR that most closely matches the clinical question while considering methodological quality, recency, comprehensiveness, and risk of bias [18, 48–50]. Our findings suggest that similarity of the reported clinical question alone may not be sufficient, as SRs with comparable PICO profiles can nevertheless synthesize substantially different underlying evidence bases. Consequently, selecting an appropriate SR should involve not only assessing its reported PICO, but also evaluating whether the RCTs included in that SR adequately represent the intended clinical question, particularly with respect to the target population and intervention strategy.

This distinction is important because apparently conflicting SRs may not represent competing summaries of the same evidence. Rather than asking which SR is methodologically superior, guideline developers should first determine whether candidate SRs synthesize comparable evidence bases. Our findings do not imply that current guideline recommendations are incorrect; however, they demonstrate that different choices of eligible SRs could reasonably lead to different interpretations of the available evidence. Explicitly documenting the alignment between the guideline question and the evidence base represented by the included RCTs could improve the transparency and reproducibility of evidence selection and help explain apparently conflicting SR conclusions.

### Clinical interpretation of discordance

Our exploratory analyses did not identify any single methodological or clinical factor that consistently explained discordant mortality results. Neither methodological quality, meta-analytic model, certainty of evidence, search date, nor any individual RCT accounted for the observed variation. Rather, discordance appeared to emerge from the cumulative effects of multiple decisions regarding intervention definitions, target populations, eligibility criteria, and study selection. Nevertheless, one clinically relevant observation emerged. Among SRs investigating non-specific broad corticosteroid strategies, SRs including populations with sepsis ± shock more frequently reported mortality benefit than those restricted to septic shock, although estimates were imprecise. Likewise, SRs including RCTs classified as likely incompatible or incompatible with contemporary sepsis definitions more often reported benefit and included more RCTs, but exclusion of these studies only modestly attenuated pooled treatment effects in sensitivity analyses. Together, these findings suggest that no single characteristic explains discordance, but they also indicate that variation in patient populations and intervention strategies across SRs may contribute to differences in pooled average treatment effects, consistent with previous concerns regarding clinical heterogeneity in sepsis research [6]. For clinicians and systematic reviewers, pooled average treatment effects may become increasingly difficult to interpret when they are derived from heterogeneous populations and intervention strategies, as they may obscure clinically meaningful differences in benefit or harm across specific patient groups [51–53].

### Implications for future evidence synthesis

Evidence synthesis traditionally aims to estimate an overall average treatment effect by combining trials considered sufficiently similar to address a common clinical question [54]. However, as patient populations, interventions, and disease definitions become increasingly heterogeneous, pooled average treatment effects may become progressively less informative for clinical decision-making because they can mask clinically relevant heterogeneity in treatment response [55]. Rather than continuing to produce increasingly broad pooled estimates, future SRs should place greater emphasis on clinically coherent populations and intervention strategies. Explicit and transparent characterization of the underlying evidence base, including which patient populations and treatment strategies are represented, may improve the interpretability of pooled estimates and facilitate more meaningful comparisons between SRs.

This perspective also aligns with the increasing interest in precision medicine in sepsis. Although our study was not designed to identify treatment-responsive subgroups, the observed differences between broader sepsis populations and septic shock populations illustrate how average treatment effects may differ depending on the populations included in evidence syntheses. Future SRs should therefore move beyond asking whether corticosteroids are effective on average and instead aim to characterize treatment effects within clinically coherent populations, providing a stronger foundation for identifying patients most likely to benefit.

### Strengths and limitations

The strengths of this study include the comprehensive characterization of all recent SRs evaluating corticosteroids in sepsis, systematic mapping of their underlying RCT evidence base, standardized PICO classification of both SRs and RCTs, and exploratory reclassification of RCTs according to contemporary sepsis definitions. By combining study-pool similarity analyses with clinical PICO assessment, we were able to characterize not only whether SRs differed, but also how differences in their clinical scope related to the composition of their underlying evidence bases.

Several limitations should also be acknowledged. First, the analyses exploring sources of discordance were exploratory and based on a relatively small number of SRs. Consequently, several associations were imprecisely estimated and should not be interpreted as causal. Second, classification of historical RCTs according to contemporary sepsis definitions required post hoc judgement and was limited by incomplete reporting in older trials. Third, we focused on short-term mortality because it was the most consistently reported outcome across SRs; findings may not be generalizable to other outcomes. Finally, our analyses were restricted to corticosteroids in sepsis, although the methodological framework may be applicable to other clinical fields with multiple overlapping SRs.

### Conclusion

SRs addressing apparently similar clinical questions frequently synthesize different underlying evidence bases and report discordant conclusions despite similar reported PICO profiles. Future guideline development should therefore evaluate not only methodological quality and certainty of evidence, but also whether the included RCTs adequately represent the intended clinical question. As the number of SRs continues to grow, selecting the SR that best reflects the clinical question and its underlying evidence base may become as important as assessing its methodological quality. More clinically coherent and transparent evidence syntheses may improve the interpretation of conflicting findings and help move beyond increasingly heterogeneous average treatment effects towards more targeted corticosteroid therapy in sepsis.

## Data Availability

All data extracted, generated or analysed during this study are included in this published article and supplementary information files.

## List of abbreviations

AMSTAR: A MeaSurement Tool to Assess systematic Reviews
ARDS: Acute Respiratory Distress Syndrome
GRADE: Grading of Recommendations, Assessment, Development and Evaluation
HAT: Hydrocortisone, Ascorbic acid, and Thiamine
HC + FC: Hydrocortisone and Fludrocortisone
IQR: Interquartile range
J: Jaccard
PICO: Participant, Intervention, Comparator, Outcome
PRISMA: Preferred Reporting Items for Systematic reviews and Meta-Analyses
PSS: Phoenix Sepsis Score
RCT: Randomized controlled trial
RoB: Risk of Bias
SOFA: Sequential Organ Failure Assessment
SR: Systematic Review
UPGMA: Unweighted Pair Group Method with Arithmetic mean

## Declarations

## Ethics approval and consent to participate

Not applicable.

## Consent for publication

Not applicable.

## Authors’ contributions

Conceptualization: SW, TP, MK, PM; Data extraction and curation: SW, HD, TP; Critical appraisal: HD, TP; Data analysis: SW; Interpretation: SW, HD, TP, MK, PM; Methodology: SW, HD, TP; Funding acquisition (departmental funding only): PM; Project administration: SW; Supervision: SW; Writing - original draft: SW; Writing - review and editing: SW, HD, TP, MK, PM.

The manuscript has been read and approved by all co-authors.

## Acknowledgements

We would like to thank Dr Maria-Inti Metzendorf for proof-reading the literature search strings and Elisabeth Friedrich-Würstlein for support in literature retrieval.

## Declaration of generative AI and AI-assisted technologies in the writing process

During the preparation of this work, the authors used generative artificial intelligence (ChatGPT) to assist with code generation (R scripts) and language editing. All AI-generated content was reviewed, critically evaluated, and edited by the authors as necessary. The authors take full responsibility for the accuracy, integrity, and originality of the content of this publication. No data analysis, interpretation of results, or scientific conclusions were generated using AI.

