## Supplementary Methods, Supplementary Table, and Supplementary Figure for "Discordant Evidence on Corticosteroids in Sepsis: A Meta-Research Study"

### Supplementary Material

Supplementary Table S19: SRxRCT mortality matrix showing randomized controlled trials (n=121) included, meta-analyzed, and not included across systematic reviews (n = 42) 203

### Supplementary Methods

#### Eligibility criteria and selection of SRs

Eligibility criteria were defined a priori and described in detail in the study protocol (https://osf.io/7d9fu). Briefly, we included interventional systematic reviews (SRs) with or without meta-analysis investigating randomized controlled trials (RCTs) of patients with suspected or confirmed sepsis or septic shock, irrespective of age or severity of sepsis. SRs were required to include a systematic literature search and a risk of bias assessment. SRs focusing on corticosteroid therapy as a primary intervention, either alone or in combination with other agents, were eligible for inclusion in the present analysis (pool #2 in the protocol). If update versions of the same SR were available, all published updates were retained for analysis.

We systematically searched MEDLINE and Epistemonikos from January 2015 to 20 June 2025 for eligible SRs.

*MEDLINE via PubMed*

#1: "sepsis"[MeSH Terms] OR "sepsis"[Title/Abstract] OR "endotoxin shock*"[Title/Abstract] OR "septic shock*"[Title/Abstract] OR "septicemia*"[Title/Abstract] OR "distributive shock*"[Title/Abstract] OR "toxic shock*"[Title/Abstract]

#2: ("systematic*"[Title] AND "review"[Title]) OR "systematic overview*"[Title] OR "cochrane review*"[Title] OR "systemic review*"[Title] OR "systematic literature search"[Title] OR "systematic literature research"[Title] OR "meta-analyses"[Title] OR "metaanalyses"[Title] OR "metaanalysis"[Title] OR "meta-analysis"[Title] OR "meta analytic review"[Title] OR "meta analytical review"[Title] OR "meta-analysis"[Publication Type] OR "The Cochrane database of systematic reviews"[Journal] OR (("search*"[Title/Abstract] OR "medline"[Title/Abstract] OR "pubmed"[Title/Abstract] OR "embase"[Title/Abstract] OR "Cochrane"[Title/Abstract] OR "scopus"[Title/Abstract] OR "web of science"[Title/Abstract] OR "sources of information"[Title/Abstract] OR "data sources"[Title/Abstract] OR "following databases"[Title/Abstract]) AND ("study selection"[Title/Abstract] OR "selection criteria"[Title/Abstract] OR "eligibility criteria"[Title/Abstract] OR "inclusion criteria"[Title/Abstract] OR "exclusion criteria"[Title/Abstract]))

#3: systematic review [pt]

#4: LETTER [PT] OR EDITORIAL [PT] OR COMMENT [PT] OR CASE REPORTS [PT] OR HISTORICAL ARTICLE [PT]

#5: (#2 OR #3) NOT #4

#6: #1 AND #5

*Epistemonikos*

(title:(sepsis) OR abstract:(sepsis)) OR (title:(septicemia*) OR abstract:(septicemia*)) OR (title:(septic shock*) OR abstract:(septic shock*)) OR (title:(endotoxic shock*) OR abstract:(endotoxic shock*)) OR (title:(endotoxin shock*) OR abstract:(endotoxin shock*)) OR (title:(distributive shock*) OR abstract:(distributive shock*)) OR (title:(toxic shock*) OR abstract:(toxic shock*))

Two reviewers independently screened titles/abstracts and full texts, with disagreements resolved by discussion with a third reviewer. Study selection was documented in a PRISMA flow diagram, and excluded full-text articles were listed with reasons.

**Data extraction and quality assessment**

For each included SR, we extracted bibliographic and methodological characteristics, eligibility criteria, mortality outcomes and corresponding meta-analytic results, certainty-of-evidence assessments, and all included randomized controlled trials (RCTs). We additionally recorded whether each RCT contributed to the respective short-term mortality meta-analysis.

For each unique RCT with an available full-text publication, detailed information was extracted on the original eligibility criteria and characteristics of the enrolled population relevant to the assessment of sepsis status. These included the infection status, reported sepsis or septic shock definitions, organ dysfunction, vasopressor or inotropic requirements, lactate concentrations, respiratory and renal dysfunction, and reported severity scores such as the Sequential Organ Failure Assessment (SOFA) score or Phoenix Sepsis Score (PSS). One reviewer performed the primary extraction, and a second reviewer independently cross-checked all extracted data for accuracy.

Reporting quality and methodological rigor of the included SRs were assessed using PRISMA 2020 [21] and AMSTAR 2 [22; see main article], respectively, as prespecified in the study protocol [20; see main article]. Overall confidence according to AMSTAR 2 was categorized as high, moderate, low, or critically low. Assessments were performed by one reviewer and verified by a second reviewer.

**Construction of the SR×RCT inclusion matrix and study-pool similarity analysis**

To characterize the structure and overlap of the evidence base, we constructed a binary SR×RCT inclusion matrix in which rows represented individual SRs and columns represented unique RCTs. An RCT was coded as present when it was reported as included in the respective SR and absent otherwise.

Pairwise study-pool similarity between all SRs was quantified using the Jaccard similarity coefficient (J), defined as the number of RCTs shared by two SRs divided by the total number of unique RCTs included across both SRs. Thus, $J = 0$ indicated no shared RCTs and $J = 1$ indicated identical study pools. The resulting pairwise similarity matrix was visualized as a heatmap.

To identify groups of SRs with similar underlying study pools, hierarchical agglomerative clustering was performed using Jaccard distance ($1 - J$) and average linkage (unweighted pair group method with arithmetic mean [UPGMA]). Clusters were defined from the resulting dendrogram using a Jaccard distance cut-off of 0.5, corresponding to a Jaccard similarity of 0.5.

**Classification of systematic reviews according to PICO profiles**

To enable standardized comparison of the clinical questions addressed by the included systematic reviews (SRs), we derived post hoc PICO profiles from the eligibility criteria reported in each SR. Classification was based on the clinical scope defined by the SR authors rather than on the characteristics of the RCTs ultimately included in the SR. Thus, the PICO profile represents the intended clinical question of each SR, whereas the composition of the underlying RCT study pool was assessed separately.

Eligibility criteria were translated into a multidimensional binary coding framework covering population, intervention, and comparator characteristics. Each category was coded as present or absent according to whether it was permitted by the reported eligibility criteria. Categories within a domain were not considered mutually exclusive; multiple categories were retained when the eligibility criteria encompassed more than one category. This approach preserved the breadth of the original review question rather than assigning each SR to a single mutually exclusive PICO category.

**Population** was characterized according to age and target population. Age was classified as adults (PA1), children (PA2), mixed-age populations (PA3), or age not defined (PA4). The target population was classified as sepsis irrespective of shock status (PD1), sepsis without shock (PD2), septic shock only (PD3), or other populations (PD4). Classification reflected the population explicitly eligible according to the SR rather than a retrospective assessment of whether individual included RCT populations fulfilled contemporary sepsis definitions.

**Intervention** was characterized across four dimensions: corticosteroid type, dose, timing/schedule, and combination strategy. Corticosteroid type distinguished between any or multiple corticosteroids without restriction (IT1), hydrocortisone (IT2), dexamethasone (IT3), methylprednisolone (IT4), and other corticosteroids (IT5). Dose was classified as not defined (ID0), low-dose (ID1), or high-dose (ID2). Timing and schedule were classified as not defined (ITM0), standard administration without a specific timing or schedule restriction (ITM1), a specific timing-focused intervention (ITM2), or a specific treatment-schedule intervention (ITM3). Combination strategy distinguished between SRs in which monotherapy versus combination therapy was not defined (IC0), corticosteroid monotherapy (IC1), hydrocortisone plus fludrocortisone (HC+FC; IC2), hydrocortisone, ascorbic acid, and thiamine (HAT; IC3), and other predefined combination strategies (IC4).

**Comparator** eligibility was classified as standard care, placebo, or no corticosteroid treatment (C1); an active corticosteroid comparator (C2); or another, unclear, or unspecified comparator (C3).

Because the full multidimensional coding framework generated highly granular PICO profiles, the primary analyses focused on the dimensions considered most directly relevant to the clinical question and to differences in study-pool composition: intervention combination strategy and target population. SRs were therefore clinically grouped according to their intervention strategy, including HC+FC, HAT, non-specific broad corticosteroid strategies, and other specific intervention strategies, and according to their target population, particularly sepsis irrespective of shock status (sepsis ± shock) versus septic shock only. Age, corticosteroid type, dose, timing/schedule, and comparator remained part of the full PICO characterization and were retained for descriptive analyses.

For structured presentation of the complete PICO profiles, SRs were ordered hierarchically according to (1) intervention combination strategy, (2) corticosteroid type, (3) target population, (4) age group, and (5) intervention timing and schedule. This ordering was used for visualization and descriptive comparison only and did not affect the study-pool similarity or clustering analyses.

Importantly, PICO classification and study-pool classification represented two distinct analytical levels. The PICO profile characterized the clinical question specified by the SR, whereas the SR×RCT inclusion matrix characterized the RCT evidence actually included in that SR. Comparing these two levels allowed us to assess whether SRs with similar clinical questions also relied on similar underlying RCT evidence bases.

#### Post hoc classification of RCT populations according to contemporary sepsis definitions

Because the RCTs included across systematic reviews (SRs) were conducted over several decades and used heterogeneous definitions of sepsis, septic shock, infection, and organ dysfunction, we performed an exploratory post hoc classification of RCT populations according to their compatibility with contemporary consensus definitions. For adult populations, compatibility was assessed with reference to Sepsis-3 criteria [23; see main article]; for pediatric populations, the Phoenix Sepsis Score (PSS) criteria were used [24; see main article]. The purpose of this classification was to characterize the clinical populations represented in the evidence base using a common contemporary framework, rather than to reassess the original eligibility or validity of individual RCTs.

Only RCTs for which a full-text publication was available were classified. Classification was based exclusively on information reported in the original trial publication and did not depend on the diagnostic terminology used by the original investigators. Thus, studies described as enrolling patients with sepsis, severe sepsis, septic shock, severe infection, pneumonia, acute respiratory distress syndrome (ARDS), COVID-19, or other conditions were assessed according to the clinical information reported for the enrolled population rather than according to the diagnostic label alone. RCTs without an available full-text publication were retained as a separate category and were not assigned a compatibility classification.

**Step 1**: Classification of the original target population

Each RCT was first classified according to the population targeted by its original eligibility criteria. Target populations were categorized as sepsis irrespective of shock status (sepsis ± shock), sepsis without shock, septic shock only, or another population not specifically defined by sepsis or septic shock. This classification described the population intended for enrollment in the original RCT and was kept separate from the subsequent assessment of compatibility with contemporary sepsis definitions.

**Step 2**: Assessment of infection and organ dysfunction

Compatibility with contemporary sepsis definitions was assessed using both the original eligibility criteria and reported baseline characteristics of the enrolled population. For adult RCTs, we evaluated whether the available information supported the presence of infection together with acute organ dysfunction consistent with Sepsis-3. For pediatric RCTs, corresponding information was evaluated with reference to the PSS framework.

The assessment considered all relevant information available in the publication, including the presence or suspicion of infection, reported sepsis or septic shock criteria, organ dysfunction, vasopressor or inotropic requirements, lactate concentrations, mechanical ventilation or respiratory dysfunction, renal dysfunction, and reported severity scores such as the Sequential Organ Failure Assessment (SOFA) score or PSS.

When a total SOFA score was explicitly reported, this information was used directly. Absence of a reported SOFA score did not automatically result in classification as unclear. Where sufficient information was available, reported clinical and laboratory characteristics corresponding to individual SOFA domains were considered to determine whether organ dysfunction compatible with Sepsis-3 was supported by the published data. These included, in particular, respiratory, cardiovascular, and renal dysfunction. Importantly, these assessments were made at the study-population level from aggregate published information; individual participants were not retrospectively assigned SOFA or PSS scores.

**Step 3**: Overall compatibility classification

Based on the combined evidence from eligibility criteria and baseline characteristics, each RCT with an available full text was assigned to one of five categories:

**Compatible:** The reported characteristics (and eligibility criteria not necessarily) of the enrolled population provided sufficient evidence that the target population fulfilled the relevant contemporary consensus definition.

**Likely compatible:** The available information strongly suggested compatibility with the contemporary definition, but the required criteria could not be established completely or explicitly from the published aggregate data.

**Unclear:** The available information was insufficient to determine whether the population was compatible or incompatible with the contemporary definition.

**Likely incompatible:** The reported eligibility criteria and/or population characteristics suggested that a substantial part or the intended target population would probably not fulfill the contemporary definition, although incompatibility could not be established unequivocally from the available aggregate data.

**Incompatible:** The reported eligibility criteria clearly permitted or defined a population that did not fulfill the relevant contemporary sepsis definition.

The distinction between likely compatible, unclear, and likely incompatible was therefore based on the direction and strength of the available evidence rather than simply on the amount of reporting. Missing information alone supported an unclear classification, whereas likely incompatible required reported information suggesting that the study population did not meet the contemporary definition.

The classification reflects compatibility of the RCT population as reported at the aggregate study level with contemporary consensus definitions and should not be interpreted as a retrospective diagnosis of individual trial participants. In particular, historical RCTs could be classified as compatible even when Sepsis-3 or PSS terminology was not used, provided that the reported eligibility criteria and population characteristics supported the corresponding contemporary criteria. Conversely, use of the terms sepsis or septic shock in the original publication was not considered sufficient by itself to establish compatibility.

The classification was performed solely to enable a standardized exploratory characterization of populations represented across the RCT evidence base. Because contemporary definitions were applied retrospectively to studies conducted under different historical definitions and because individual-patient data were unavailable, some classifications necessarily remained uncertain. Furthermore, this analysis was not prespecified in the study protocol. All analyses based on the compatibility classification were therefore considered exploratory.

**Linking PICO profiles and study-pool clusters**

To assess whether SRs addressing similar clinical questions relied on similar underlying evidence bases, we linked the standardized PICO profiles to clusters derived independently from study-pool similarity.

Two complementary descriptive perspectives were used. First, from a PICO-centred perspective, we examined how SRs sharing similar intervention and target-population profiles were distributed across study-pool clusters. Dispersion of clinically similar SRs across multiple clusters indicated that similar reported clinical questions were represented by different underlying study pools.

Second, from a cluster-centred perspective, we characterized the clinical composition of each study-pool cluster according to the PICO profiles of its constituent SRs. This allowed us to assess whether SRs grouped together because of similar RCT evidence bases also addressed clinically similar questions.

The PICO variables were not used to generate the study-pool clusters; clustering was based exclusively on overlap in included RCTs.

**Analysis of discordance in mortality outcomes**

Discordance was assessed for short-term mortality because this was the mortality outcome most consistently reported across SRs. Mortality conclusions were categorized as benefit when the pooled estimate indicated a statistically significant reduction in mortality, no evidence of effect when the pooled estimate was not statistically significant, and harm when the pooled estimate indicated a statistically significant increase in mortality.

Analyses of potential sources of discordance focused on clinically comparable SRs addressing non-specific broad corticosteroid strategies in adult or mixed-age populations, excluding SRs restricted to dexamethasone or specifically investigating corticosteroid timing or treatment schedule.

Potential sources of discordance included target population (sepsis ± shock vs septic shock only), study-pool composition and overlap, individual RCT inclusion, inclusion of RCTs classified as likely incompatible or incompatible with contemporary sepsis definitions, number of RCTs contributing to the mortality meta-analysis, mortality timepoint, meta-analytic model, effect measure, certainty of evidence according to GRADE, methodological confidence according to AMSTAR 2, and recency of the evidence according to search date.

Study-pool overlap between mortality meta-analyses was quantified using pairwise Jaccard similarity. We additionally examined whether inclusion of individual RCTs was consistently associated with mortality conclusions favoring corticosteroids or with no evidence of effect.

Categorical variables were compared using Fisher's exact test and continuous variables using the Wilcoxon rank-sum test. Given the limited number of eligible SRs and the exploratory nature of these comparisons, estimates were interpreted descriptively and were not considered evidence of causal associations.

**Sensitivity analyses of RCT population compatibility**

To explore whether RCT populations with varying compatibility with contemporary sepsis definitions influenced pooled treatment effects, we repeated the short-term mortality meta-analyses from Annane et al. [7; see main article] and Pitre et al. [11; see main article]. Annane et al. was selected because it contained the largest RCT study pool, whereas Pitre et al. informed current international guideline recommendations [15,16; see main article].

For each SR, the evidence base was progressively restricted according to the post hoc compatibility classification. Meta-analyses were performed including:

1. RCTs classified as compatible only;
2. RCTs classified as compatible or likely compatible;
3. RCTs classified as compatible, likely compatible, or unclear.

The third analysis therefore excluded RCTs classified as likely incompatible or incompatible and RCTs without an available full-text publication. In addition, RCTs classified as likely incompatible or incompatible were analyzed separately to characterize the pooled treatment effect within this subgroup.

RCTs without an available full-text publication were retained when reproducing the original meta-analysis but could not be assigned to a compatibility category and were therefore not included in classification-based restricted analyses.

For each SR, the effect measure and meta-analytic methods followed those used in the original publication. For Annane et al. [7], risk ratios (RRs) were pooled using the Mantel–Haenszel method under a random-effects model, with between-study variance ($\tau^2$) estimated using the DerSimonian–Laird method and without Hartung–Knapp adjustment. For Pitre et al. [11], RRs were pooled using the Mantel–Haenszel method under a random-effects model, with $\tau^2$ estimated using maximum likelihood and without Hartung–Knapp adjustment. This approach preserved the analytical framework of each original meta-analysis while varying only the composition of the underlying RCT evidence base according to the post hoc population classification.

**Statistical analysis**

All data processing and statistical analyses were performed in R. Heatmaps and clustering visualizations were generated using the pheatmap and ggplot2 packages. Pairwise similarities were calculated using the Jaccard coefficient, and hierarchical clustering was based on Jaccard distance with average linkage.

Given the exploratory nature of the analyses and the limited number of SRs available for several comparisons, statistical testing was used primarily to quantify observed associations rather than for confirmatory inference. Results were therefore interpreted with emphasis on effect estimates, uncertainty, and consistency of observed patterns rather than statistical significance alone.

**Supplementary Figure S1: PRISMA Flow diagram**

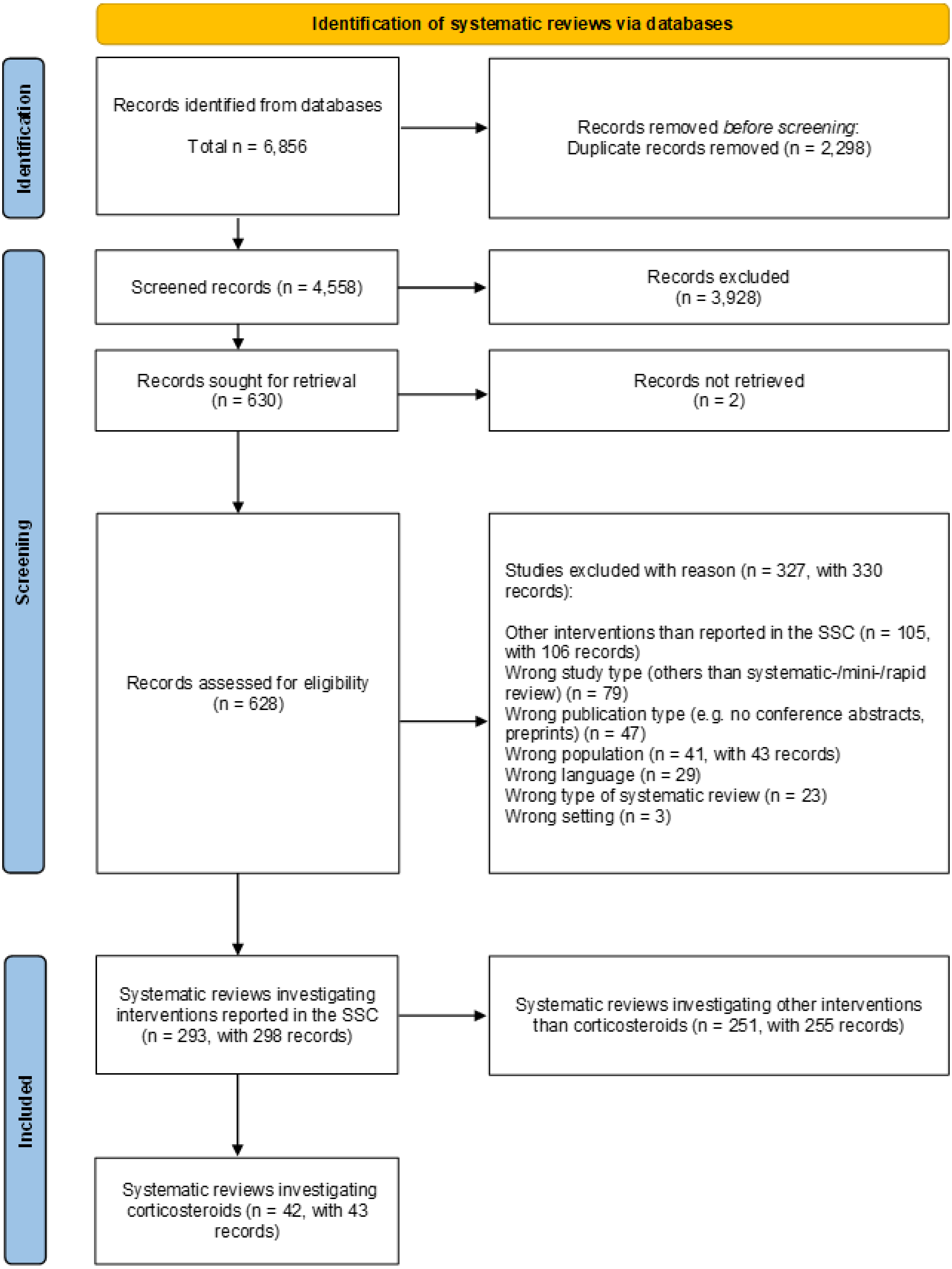

### Supplementary Figure S2: PRISMA 2020 item-level reporting across systematic reviews

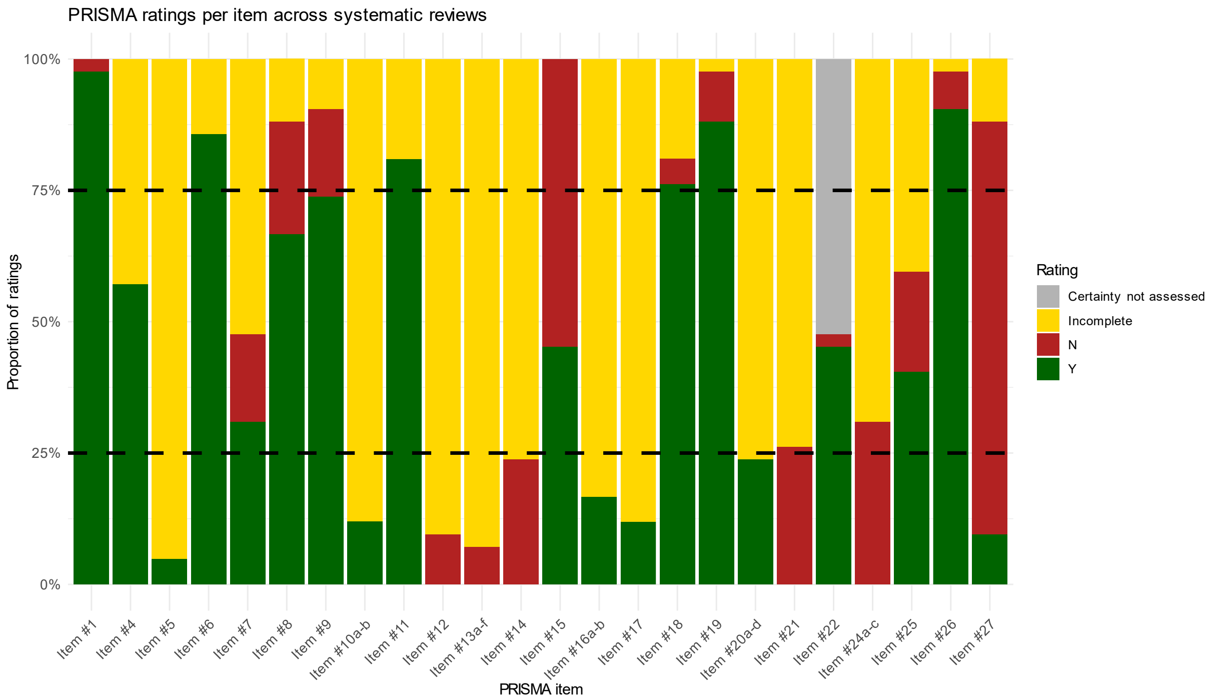

Reporting quality of systematic reviews (SRs) was assessed using the PRISMA 2020 checklist. Each item was operationalized in question format and rated as ‘yes’ (all reporting requirements fully met, Y), ‘incomplete’ (key elements missing), or ‘no’ (no relevant information identified, N). ‘Certainty not assessed’ indicated that certainty of evidence was not assessed (item # 22). Bars represent the proportion of ratings per item across included SRs. Dashed horizontal lines indicate thresholds at 25% and 75% of reviews rated ‘yes’.

**Supplementary Figure S3: AMSTAR 2 item-level appraisal across systematic reviews**

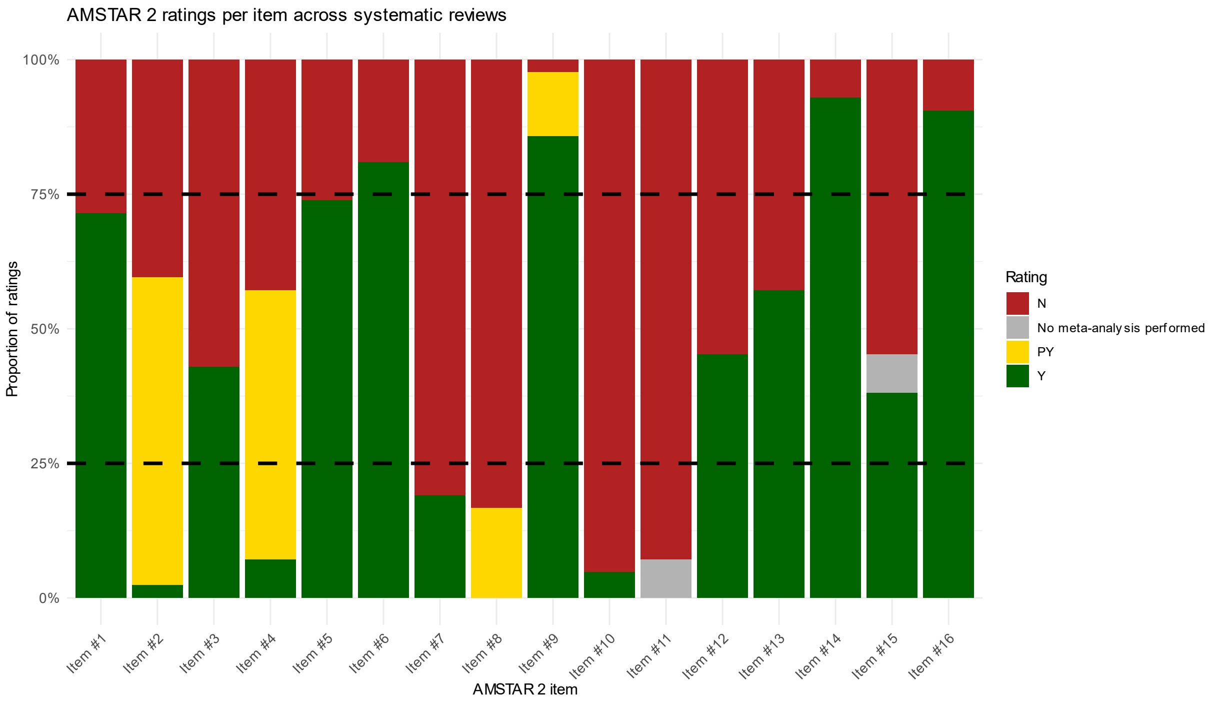

Methodological quality of systematic reviews (SRs) was assessed using the AMSTAR 2 instrument. Each of the 16 items (including critical and non-critical domains) was operationalized and rated as ‘yes’ (criterion fully met, Y), ‘partial yes’ (criterion partially met, PY), or ‘no’ (criterion not met, N). ‘No meta-analysis performed’ indicated that no quantitative synthesis was conducted in a systematic review (item #11 and #15). Bars represent the proportion of ratings per item across included SRs. Dashed horizontal lines indicate thresholds at 25% and 75% of reviews rated ‘yes’.

### Supplementary Figure S4: Inclusion matrix of randomized controlled trials across systematic reviews – heat-map

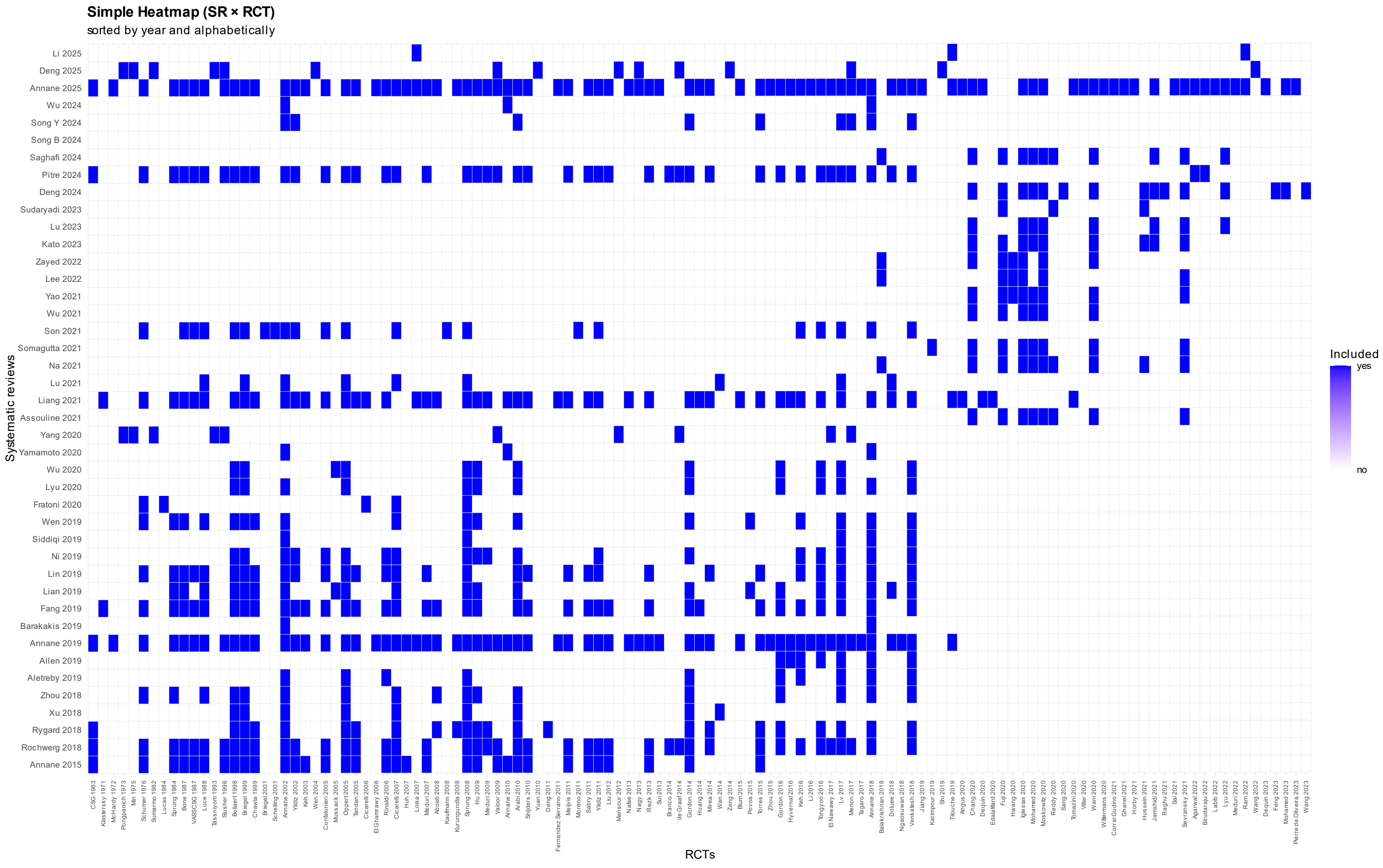

Heatmap showing the binary inclusion of randomized controlled trials (RCTs, n=121) across systematic reviews (SRs, n=42). Rows represent individual SRs and columns represent RCTs identified in the evidence base. Blue cells indicate that a given RCT was included in the corresponding SR, whereas white cells indicate absence of inclusion.

Both SRs and RCTs are ordered chronologically by publication year and alphabetically within the same year. This representation visualizes the distribution and overlap of primary studies across systematic reviews and highlights differences in study pools among reviews addressing corticosteroid therapy in sepsis.

#### Supplementary Table S1: References to excluded studies (n = 327) with reason for exclusion

| Study ID | Title | Authors | Published Year | Journal | Volume | Issue | Pages | DOI | Exclusion reason |
|---|---|---|---|---|---|---|---|---|---|
| Ding 2021 | Association Between Prior Calcium Channel Blocker Use and Mortality in Septic Patients: A Meta-Analysis of Cohort Studies | Ding, X.; Cui, Y.; Liang, H.; Wang, D.; Li, L.; Kan, Q.; Wang, L.; Sun, T. | 2021 | Frontiers in pharmacology | 12 | | 628825 | 10.3389/fphar.2021.628825 | Exclusion reason: Other intervention than reported in the SSCs |
| Diz 2025 | Effect of Treatment With Balanced Crystalloids Versus Normal Saline on the Mortality of Critically Ill Patients With and Without Traumatic Brain Injury: A Systematic Review and Meta-Analysis | Diz, J. C.; Luna-Rojas, P.; Díaz-Vidal, P.; Fernández-Vázquez, U.; Gil-Casado, C.; Diz-Ferreira, E. | 2025 | Anesthesia and analgesia | | | | 10.1213/ANE.0000000000007368 | Exclusion reason: Wrong population (e.g. animals and humans pooled together, sepsis only as subgroup,...) |
| Donà 2019 | Strategic Trials to Define the Best Available Treatment for Neonatal and Pediatric Sepsis Caused by Carbapenem-resistant Organisms | Donà, D.; Sharland, M.; Heath, P. T.; Folgori, L. | 2019 | Pediatr Infect Dis J | 38 | 8 | 825-827 | 10.1097/inf.0000000000002381 | Exclusion reason: Wrong study type (no systematic-/mini-/rapid review) |
| Akselsen 2022 | Efficacy of empiric antibiotic treatment of late-onset neonatal sepsis caused by Enterobacteriaceae: A Systematic Review | Akselsen, A. B.; Sheth, C. C.; Veses, V. | 2022 | Letters in applied microbiology | 75 | 3 | 500-510 | 10.1111/lam.13640 | Exclusion reason: Wrong study type (no systematic-/mini-/rapid review) |
| Draeger 2025 | Barriers and facilitators to optimal sepsis care - a systematized review of healthcare | Draeger, L.; Fleischmann-Struzek, C.; Gehrke- | 2025 | BMC health services research | 25 | 1 | 591 | 10.1186/s12913-025-12777-8 | Exclusion reason: Wrong study type (no systematic-/mini-/rapid review) |

| | | | | | | | | | |
|---|---|---|---|---|---|---|---|---|---|
| | professionals' perspectives | Beck, S.; Heintze, C.; Thomas-Rueddel, D. O.; Schmidt, K. | | | | | | | |
| AlSulaiman 2024 | The concomitant use of ultra short beta-blockers with vasopressors and inotropes in critically ill patients with septic shock: A systematic review and meta-analysis of randomized controlled trials | Al Sulaiman, K.; Alkofide, H. A.; AlFaifi, M. E.; Aljohani, S. S.; Al Harthi, A. F.; Alqahtani, R. A.; Alanazi, A. M.; Nazer, L. H.; Al Shaya, A. I.; Aljuhani, O. | 2024 | Saudi pharmaceutical journal : SPJ : the official publication of the Saudi Pharmaceutical Society | 32 | 6 | 102094 | 10.1016/j.jsps.2024.102094 | Exclusion reason: Other intervention than reported in the SSCs |
| Edding 2020 | SAT-006 Efficacy and Safety of High-volume Hemofiltration (HVHF) in patients with Septic Shock and Acute Kidney Injury: A Systematic Review and Meta-Analysis of Randomized Controlled Trials | Edding, S.; Cabral, B. M. | 2020 | Kidney Intl. Rep. | 5 | 3 | S2-S3 | 10.1016/j.ekir.2020.02.010 | Exclusion reason: Wrong publication type (e.g. conference abstract, preprints,...) |
| Edding 2020 | Efficacy and safety of high volume hemofiltration (HVHF) in patients with septic shock: A meta analysis of randomized controlled trials | Edding, S. N. | 2020 | Hemodial. Int. | 24 | 1 | A3-None | 10.1111/hdi.12814 | Exclusion reason: Wrong publication type (e.g. conference abstract, preprints,...) |
| Edding 2019 | Efficacy and safety of high-volume hemofiltration (HVHF) | Edding, S. N.; Cabral, B. M. I. | 2019 | Journal of the American | 30 | | 555 | | Exclusion reason: Wrong publication type |

| | | | | | | | | | |
|---|---|---|---|---|---|---|---|---|---|
| | in patients with septic shock and aki: A systematic review and meta-analysis of randomized controlled trials | | | Society of Nephrology | | | | | (e.g. conference abstract, preprints,...) |
| Al-Kader 2022 | Systematic Review on the Effects of Prompt Antibiotic Treatment on Survival in Septic Shock and Sepsis Patients in Different Hospital Settings | Al-Kader, D. A.; Anwar, S.; Hussaini, H.; Jones Amaowei, E. E.; Rasuli, S. F.; Hussain, N.; Kaddo, S.; Memon, A. | 2022 | Cureus | 14 | 12 | e32405 | 10.7759/cureus.32405 | Exclusion reason: Wrong study type (no systematic-/mini-/rapid review) |
| Alam 2017 | Long-term health related quality of life in patients with sepsis after intensive care stay: A systematic review | Alam, N.; Nannan Panday, R. S.; Heijnen, J. R.; Van Galen, L. S.; Kramer, M. H. H.; Nanayakkara, P. W. B. | 2017 | Acute Medicine | 16 | 4 | 164-169 | | Exclusion reason: Wrong type of systematic review (e.g. diagnostic (test accuracy), experiential (qualitative), prognostic, etiology and/or risk, prevalence and/or incidence reviews) |
| Evelyn 2020 | Should anticoagulation be used in the treatment of severe COVID-19? | Evelyn, O. Salido; Jaime Alfonso, M. Aherrera; Patricia Pauline, M. Remalante | 2020 | | 54 | | | 10.47895/AMP.V54I0.1717 | Exclusion reason: Wrong study type (no systematic-/mini-/rapid review) |
| Fan 2016 | Ulinastatin and Thymosin a1 Therapy in Adult Patients with Severe Sepsis: A Meta-analysis with Trial Sequential | Fan, H.; Zhao, Y.; Zhu, J. H.; Ye, J. H.; Yao, X. Q. | 2016 | Iranian journal of public health | 45 | 9 | 1234-1235 | | Exclusion reason: Other intervention than reported in the SSCs |

| | | | | | | | | | |
|---|---|---|---|---|---|---|---|---|---|
| | Analysis of Randomized Controlled Trials | | | | | | | | |
| Farrow 2015 | A systematic review and meta-analysis regarding the use of corticosteroids in septic arthritis | Farrow, L. | 2015 | BMC musculoskeletal disorders | 16 | 1 | 241 | 10.1186/s12891-015-0702-3 | Exclusion reason: Wrong population (e.g. animals and humans pooled together, sepsis only as subgroup,...) |
| Fedyaeva 2015 | Meta-analysis of mortality in adults, newborns and older children with bacterial infections and sepsis when treated by IGM-enriched intravenous immunoglobulins and standard schemes | Fedyaeva, V. K.; Rebrova, O. Y. | 2015 | Value in Health | | | A578 | | Exclusion reason: Wrong publication type (e.g. conference abstract, preprints,...) |
| Feng 2016 | Ulinastatin and/or Thymosin Alpha1 for Severe Sepsis: A Systematic Review and Meta-analysis | Feng, Z.; Shi, Q.; Fan, Y.; Wang, Q.; Yin, W. | 2016 | The journal of trauma and acute care surgery | 80 | 2 | 335-40 | 10.1097/TA.0000000000000909 | Exclusion reason: Other intervention than reported in the SSCs |
| Feng 2016 | Ulinastatin and/or thymosin α1 for severe sepsis: A systematic review and meta-analysis | Feng, Z.; Shi, Q.; Fan, Y.; Wang, Q.; Yin, W. | 2016 | J Trauma Acute Care Surg | 80 | 2 | 335-40 | 10.1097/ta.0000000000000909 | Exclusion reason: Other intervention than reported in the SSCs |
| Fernández-Sarmiento 2021 | Latin American Consensus on the Management of Sepsis in Children: Sociedad Latinoamericana de Cuidados Intensivos Pediátricos [Latin American Pediatric Intensive Care Society] (SLACIP) Task Force: Executive Summary | Fernández-Sarmiento, J.; De Souza, D. C.; Martinez, A.; Nieto, V.; López-Herce, J.; Soares Lanziotti, V.; Arias López, M. D. P.; De Carvalho, W. | 2021 | Journal of intensive care medicine | | | | 10.1177/08850666211054444 | Exclusion reason: Wrong study type (no systematic-/mini-/rapid review) |

| | | | | | | | | | |
|---|---|---|---|---|---|---|---|---|---|
| | | B.; Oliveira, C. F.; Jaramillo-Bustamante, J. C.; Díaz, F.; Yock-Corrales, A.; Ruvinsky, S.; Munaico, M.; Pavlicich, V.; Iramain, R.; Márquez, M. P.; González, G.; Yunge, M.; Tonial, C.; Cruces, P.; Palacio, G.; Grela, C.; Slöcker-Barrio, M.; Campos-Miño, S.; González-Dambrauskas, S.; Sánchez-Pinto, N. L.; Celiny García, P.; Jabornisky, R. | | | | | | | |
| Fernández-Sarmiento 2020 | The Endothelial Glycocalyx: A Fundamental Determinant of Vascular Permeability in Sepsis | Fernández-Sarmiento, J.; Salazar-Peláez, L. M.; Carcillo, J. A. | 2020 | Pediatr Crit Care Med | 21 | 5 | e291-e300 | 10.1097/pcc.0000000000002266 | Exclusion reason: Wrong study type (no systematic-/mini-/rapid review) |

| | | | | | | | | | |
|---|---|---|---|---|---|---|---|---|---|
| Fernando 2024 | Methylene Blue in Septic Shock: A Systematic Review and Meta-Analysis | Fernando, S. M.; Tran, A.; Soliman, K.; Flynn, B.; Oommen, T.; Wenzhe, L.; Adhikari, N. K. J.; Kanji, S.; Seely, A. J. E.; Fox-Robichaud, A. E.; Wax, R. S.; Cook, D. J.; Lamontagne, F.; Rochwerg, B. | 2024 | Critical care explorations | 6 | 7 | e1110 | 10.1097/CCE.0000000000001110 | Exclusion reason: Other intervention than reported in the SSCs |
| Flanders 2019 | The effect of dexmedetomidine and clonidine on the inflammatory response in critical illness: a systematic review of animal and human studies | Flanders, C. A.; Rocke, A. S.; Edwardson, S. A.; Baillie, J. K.; Walsh, T. S. | 2019 | Critical care (London, England) | 23 | 1 | 402 | 10.1186/s13054-019-2690-4 | Exclusion reason: Wrong population (e.g. animals and humans pooled together, sepsis only as subgroup,...) |
| Alemu 2021 | Perioperative management of adult surgical patients with septic shock in resource limiting setting, systematic review | Alemu, E. T.; Agegnehu, A. F.; Temesgen, M. M. | 2021 | Intl. J. Surg. | 29 | | 66-75 | 10.1016/j.ijso.2020.11.016 | Exclusion reason: Wrong type of systematic review (e.g. diagnostic (test accuracy), experiential (qualitative), prognostic, etiology and/or risk, prevalence and/or incidence reviews) |
| Gage-Brown 2022 | Is Piperacillin-Tazobactam an Appropriate Empirical Agent for Hospital- | Gage-Brown, A.; George, C.; Maleki, | 2022 | Healthcare (Basel, Switzerland) | 10 | 5 | | 10.3390/healthcare10050851 | Exclusion reason: Wrong study type (no systematic-/mini-/rapid review) |

| | | | | | | | | | |
|---|---|---|---|---|---|---|---|---|---|
| | Acquired Sepsis and Community-Acquired Septic Shock of Unknown Origin in Australia? | J.; Singh, K. P.; Muhi, S. | | | | | | | |
| Gale 2020 | The Use of Patient Monitoring Systems to Improve Sepsis Recognition and Outcomes: A Systematic Review | Gale, Bryan M.; Hall, Kendall K. | 2020 | Journal of Patient Safety | | | 8-11 | | Exclusion reason: Wrong study type (no systematic-/mini-/rapid review) |
| Alexandru 2024 | Mortality in septic patients treated with short-acting betablockers: a comprehensive meta-analysis of randomized controlled trials | Alexandru, M. G.; Niewald, P.; Krüger, S.; Borgstedt, R.; Whitehouse, T.; Singer, M.; Rehberg, S.; Scholz, S. S. | 2024 | Critical care (London, England) | 28 | 1 | 392 | 10.1186/s13054-024-05174-w | Exclusion reason: Other intervention than reported in the SSCs |
| Gelbart 2015 | Fluid Bolus Therapy-Based Resuscitation for Severe Sepsis in Hospitalized Children: A Systematic Review | Gelbart, B.; Glassford, N. J.; Bellomo, R. | 2015 | Pediatric critical care medicine : a journal of the Society of Critical Care Medicine and the World Federation of Pediatric Intensive and Critical Care Societies | 16 | 8 | e297-307 | 10.1097/PCC.0000000000000507 | Exclusion reason: Wrong study type (no systematic-/mini-/rapid review) |
| Gendreau 2024 | Geo-economic Influence on the Effect of Fluid Volume for Sepsis Resuscitation A Meta-Analysis | Gendreau, S.; Frapard, T.; Carteaux, G.; Kwizera, A.; Adhikari, | 2024 | American journal of respiratory and critical care medicine | 209 | 5 | 517-528 | 10.1164/rccm.202309-1617OC | Exclusion reason: Wrong type of systematic review (e.g. diagnostic (test accuracy), experiential |

| | | | | | | | | | |
|---|---|---|---|---|---|---|---|---|---|
| | | N. K.; Mer, M.; Hernandez, G.; Mekontso Dessap, A. | | | | | | | (qualitative), prognostic, etiology and/or risk, prevalence and/or incidence reviews) |
| Gerth 2019 | Changes in health-related quality of life after discharge from an intensive care unit: a systematic review | Gerth, A. M. J.; Hatch, R. A.; Young, J. D.; Watkinson, P. J. | 2019 | Anaesthesia | 74 | 1 | 100-108 | 10.1111/anae.14444 | Exclusion reason: Wrong type of systematic review (e.g. diagnostic (test accuracy), experiential (qualitative), prognostic, etiology and/or risk, prevalence and/or incidence reviews) |
| Girardis 2024 | Adjunctive immunotherapeutic agents in patients with sepsis and septic shock: a multidisciplinary consensus of 23 | Girardis, M.; Coloretti, I.; Antonelli, M.; Berlot, G.; Busani, S.; Cortegiani, A.; De Pascale, G.; De Rosa, F. G.; De Rosa, S.; Donadello, K.; Donati, A.; Forfori, F.; Giannella, M.; Grasselli, G.; Montrucchio, G.; Oliva, A.; Pasero, D.; Piazza, O.; Romagnoli, S.; Tascini, C.; Viaggi, | 2024 | Journal of anesthesia, analgesia and critical care | 4 | 1 | 28 | 10.1186/s44158-024-00165-3 | Exclusion reason: Wrong study type (no systematic-/mini-/rapid review) |

| | | | | | | | | | |
|---|---|---|---|---|---|---|---|---|---|
| | | B.; Tumbarello, M.; Viale, P. | | | | | | | |
| Gong 2025 | Effects of probiotic treatment on the prognosis of patients with sepsis: a systematic review | Gong, C.; Xu, S.; Pan, Y.; Guo, S.; Walline, J. H.; Wang, X.; Lu, X.; Yu, S.; Qin, M.; Zhu, H.; Gao, Y.; Li, Y. | 2025 | World journal of emergency medicine | 16 | 1 | 18-27 | 10.5847/wjem.j.1920-8642.2025.018 | Exclusion reason: Other intervention than reported in the SSCs |
| Gordon 2021 | A meta-analysis of early administration of vasopressor in septic shock: Is there mortality benefit? | Gordon, D.; Chan, C.; Tran, Q.; Bzhilyanskaya, V.; Bracey, A.; Yazdanpanah, F.; Downton, K.; Rao, T.; Schwartz, B.; Milzman, D. | 2021 | Crit. Care Med. | 49 | 1 SUPPL 1 | 641-None | 10.1097/01.ccm.0000730972.05471.29 | Exclusion reason: Wrong publication type (e.g. conference abstract, preprints,...) |
| Grazielle 2020 | Granulocyte colony stimulating factor in improving outcomes of neonatal sepsis: A meta-analysis | Grazielle, V. | 2020 | Curr. Paediatr. Res. | 24 | 6 | 4-None | | Exclusion reason: Other intervention than reported in the SSCs |
| Alkazemi 2024 | Methylene Blue for Septic Shock: A Systematic Review and Meta-analysis of Randomized and Prospective Observational Studies | Alkazemi, A.; Almoosawy, S. A.; Murad, A.; Alfares, A. | 2024 | Journal of intensive care medicine | | | | 10.1177/08850666241300312 | Exclusion reason: Other intervention than reported in the SSCs |
| Gu 2016 | Relationship between Annualized Case Volume and Mortality | Gu, W. J.; Wu, X. D.; Zhou, Q.; | 2016 | Anesthesiology | 125 | 1 | 168-79 | 10.1097/aln.0000000000001133 | Exclusion reason: Wrong type of systematic review (e.g. |

| | | | | | | | | | |
|---|---|---|---|---|---|---|---|---|---|
| | in Sepsis: A Dose-Response Meta-analysis | Zhang, J.; Wang, F.; Ma, Z. L.; Gu, X. P. | | | | | | | diagnostic (test accuracy), experiential (qualitative), prognostic, etiology and/or risk, prevalence and/or incidence reviews) |
| Guedes 2022 | The supplementation of L-carnitine in septic shock patients: Systematic review and meta-analysis | Guedes, G. V.; Minicucci, M. F.; Tanni, S. E. | 2022 | Clinics (Sao Paulo, Brazil) | 77 | | 100124 | 10.1016/j.clinsp.2022.100124 | Exclusion reason: Other intervention than reported in the SSCs |
| Wang 2022 | [Randomized controlled trial outcome indicators of traditional Chinese medicine for gastrointestinal dysfunction in sepsis in recent two years] | Wang, G. Q.; Zheng, H. J.; Cao, Y.; Peng, W.; Yao, W. H. | 2022 | Zhongguo Zhong yao za zhi = Zhongguo zhongyao zazhi = China journal of Chinese materia medica | 47 | 3 | 819-828 | 10.19540/j.cnki.cjcmm.20211013.501 | Exclusion reason: Wrong language |
| Guo-Min 2024 | Effect of vitamin C intravenous injection on prognosis of patients with sep-sis or septic shock:A Meta-analysis of randomized controlled trial | Guo-Min, Zhao; Wei-Shuai, Bian; Jie, Zhen; Wei, Chen | 2024 | Chinese Journal of Infection Control | | 4 | 32-41 | | Exclusion reason: Wrong language |
| Hammond 2020 | Balanced Crystalloids Versus Saline in Critically Ill Adults: A Systematic Review and Meta-analysis | Hammond, D. A.; Lam, S. W.; Rech, M. A.; Smith, M. N.; Westrick, J.; Trivedi, A. P.; Balk, R. A. | 2020 | The Annals of pharmacotherapy | 54 | 1 | | 10.1177/1060028019866420 | Exclusion reason: Wrong population (e.g. animals and humans pooled together, sepsis only as subgroup,...) |
| Han 2015 | Ulinastatin- and thymosin α1-based immunomodulatory | Han, D.; Shang, W.; Wang, G.; Sun, L.; | 2015 | International immunopharmacology | 29 | 2 | 377-82 | 10.1016/j.intimp.2015.10.026 | Exclusion reason: Other intervention than reported in the SSCs |

| | | | | | | | | | |
|---|---|---|---|---|---|---|---|---|---|
| | strategy for sepsis: A meta-analysis | Zhang, Y.; Wen, H.; Xu, L. | | | | | | | |
| Harris 2020 | Long-term Renal Outcomes in Adults With Sepsis-Induced Acute Kidney Injury: A Systematic Review | Harris, P. L.; Umberger, R. A. | 2020 | Dimensions of critical care nursing : DCCN | 39 | 5 | 259-268 | 10.1097/DCC.0000000000000432 | Exclusion reason: Wrong study type (no systematic-/mini-/rapid review) |
| Hasegawa 2022 | Premorbid angiotensin converting enzyme inhibitors or angiotensin II receptor blockers in patients with sepsis | Hasegawa, D.; Lee, Y. I.; Prasitlumkum, N.; Chopra, L.; Nishida, K.; Smith, R. L.; Sato, R. | 2022 | The American journal of emergency medicine | 62 | | 69-77 | 10.1016/j.ajem.2022.10.006 | Exclusion reason: Other intervention than reported in the SSCs |
| Hasegawa 2022 | Effect of Premorbid Beta-Blockers on Mortality in Patients With Sepsis: A Systematic Review and Meta-Analysis | Hasegawa, D.; Sato, R.; Prasitlumkum, N.; Nishida, K. | 2022 | Journal of intensive care medicine | 37 | 7 | | 10.1177/08850666211052926 | Exclusion reason: Other intervention than reported in the SSCs |
| Hasegawa 2021 | Effect of Ultrashort-Acting β-Blockers on Mortality in Patients With Sepsis With Persistent Tachycardia Despite Initial Resuscitation: A Systematic Review and Meta-analysis of Randomized Controlled Trials | Hasegawa, D.; Sato, R.; Prasitlumkum, N.; Nishida, K.; Takahashi, K.; Yatabe, T.; Nishida, O. | 2021 | Chest | 159 | 6 | 2289-2300 | 10.1016/j.chest.2021.01.009 | Exclusion reason: Other intervention than reported in the SSCs |
| Hazzard 2015 | Coupled plasma haemofiltration filtration in severe sepsis: Systematic review and meta-analysis | Hazzard, I.; Jones, S.; Quinn, T. | 2015 | Journal of the Royal Army Medical Corps | 161 Suppl 1 | | i17-i22 | 10.1136/jramc-2015-000552 | Exclusion reason: Other intervention than reported in the SSCs |

| | | | | | | | | | |
|---|---|---|---|---|---|---|---|---|---|
| He 2023 | Effect of midodrine on the prognosis of patients with septic shock: a systematic review and meta-analysis | He, Q. F.; Xing, X. K.; Wang, T. Q.; Jiang, Z. X.; Zhang, G. | 2023 | European review for medical and pharmacological sciences | 27 | 9 | 4211-4220 | 10.26355/eurrev_202305_32331 | Exclusion reason: Other intervention than reported in the SSCs |
| Helmbrecht 2019 | Systematic Review of the Effectiveness of the Neonatal Early-Onset Sepsis Calculator | Helmbrecht, A. R.; Marfurt, S.; Chaaban, H. | 2019 | The Journal of perinatal & neonatal nursing | 33 | 1 | 82-88 | 10.1097/JPN.0000000000000360 | Exclusion reason: Wrong type of systematic review (e.g. diagnostic (test accuracy), experiential (qualitative), prognostic, etiology and/or risk, prevalence and/or incidence reviews) |
| Helwani 2023 | Is venoarterial extracorporeal membrane oxygenation an option for managing septic shock | Helwani, M. A.; Lim, A. | 2023 | Current opinion in anaesthesiology | 36 | 1 | 45-49 | 10.1097/ACO.0000000000001206 | Exclusion reason: Wrong study type (no systematic-/mini-/rapid review) |
| Henderson 2018 | Use of melatonin as adjunctive therapy in neonatal sepsis: A systematic review and meta-analysis | Henderson, R.; Kim, S.; Lee, E. | 2018 | Complementary therapies in medicine | 39 | | 131-136 | 10.1016/j.ctim.2018.06.002 | Exclusion reason: Other intervention than reported in the SSCs |
| Heybati 2022 | Outcomes of dexmedetomidine versus propofol sedation in critically ill adults requiring mechanical ventilation: a systematic review and meta-analysis of randomised controlled trials | Heybati, K.; Zhou, F.; Ali, S.; Deng, J.; Mohananey, D.; Villablanca, P.; Ramakrishna, H. | 2022 | British journal of anaesthesia | 129 | 4 | 515-526 | 10.1016/j.bja.2022.06.020 | Exclusion reason: Wrong population (e.g. animals and humans pooled together, sepsis only as subgroup,...) |

| | | | | | | | | | |
|---|---|---|---|---|---|---|---|---|---|
| HidalgoAcosta 2023 | Mortality from sepsis in intensive care unit | Hidalgo Acosta, Javier Aquiles; Cobeña Vera, Jonathan Gabriel; Guzñay Muñoz, Daniela Abigail; Rivero Moreira, Lili Stephania; Defaz Freire, Paulina Gioconda; Zúñiga Arreaga, Leonel Amador; López Nazareno, Ingrid Lorena; Burgos Uyaguari, Karen Vanessa; Vera Espinoza, Yuridia Lisbeth | 2023 | Cambios rev. méd | 22 | 1 | 865-865 | 10.36015/cambios.v22.n1.2023.865 | Exclusion reason: Wrong language |
| Hilditch 2015 | Can pre-hospital recognition and intervention improve outcome for patients with severe sepsis? | Hilditch, Marina | 2015 | Journal of Paramedic Practice | 7 | 4 | 168-175 | | Exclusion reason: Wrong study type (no systematic-/mini-/rapid review) |

| | | | | | | | | | |
|---|---|---|---|---|---|---|---|---|---|
| Ho 2016 | The involvement of regulatory non-coding RNAs in sepsis: a systematic review | Ho, J.; Chan, H.; Wong, S. H.; Wang, M. H.; Yu, J.; Xiao, Z.; Liu, X.; Choi, G.; Leung, C. C.; Wong, W. T.; Li, Z.; Gin, T.; Chan, M. T.; Wu, W. K. | 2016 | Critical care (London, England) | 20 | 1 | 383 | 10.1186/s13054-016-1555-3 | Exclusion reason: Wrong type of systematic review (e.g. diagnostic (test accuracy), experiential (qualitative), prognostic, etiology and/or risk, prevalence and/or incidence reviews) |
| Hou 2015 | Efficacy of Xuebijing for coagulopathy in patients with sepsis | Hou, S. Y.; Feng, X. H.; Lin, C. L.; Tan, Y. F. | 2015 | Saudi Medical Journal | 36 | 2 | 164-169 | 10.15537/smj.2015.2.9895 | Exclusion reason: Other intervention than reported in the SSCs |
| Alshammari 2023 | Risk and diagnostic factors and therapy outcome of neonatal early onset sepsis in ICU patients of Saudi Arabia: a systematic review and meta analysis | Alshammari, M. K.; Alsanad, A. H.; Alnusayri, R. J.; Alanazi, A. S.; Shamakhi, F. Q.; Alshahrani, K. M.; Alshahrani, A. M.; Yahya, G.; Alshahrani, A. A.; Alshahrani, T. S.; Sultan, H. S.; Alshahrani, F. M.; Alreshidi, F. A.; Alnigaidan, | 2023 | Frontiers in pediatrics | 11 | | 1206389 | 10.3389/fped.2023.1206389 | Exclusion reason: Wrong type of systematic review (e.g. diagnostic (test accuracy), experiential (qualitative), prognostic, etiology and/or risk, prevalence and/or incidence reviews) |

| | | | | | | | | | |
|---|---|---|---|---|---|---|---|---|---|
| | | R. A.; Almazyad, A. A. | | | | | | | |
| Huang 2020 | Continuous Infusion Compared with Intermittent Intravenous Infusion of Beta-Lactam Antibiotics for Critically Ill Patients: A Systematic Review and Meta-Analysis of Randomized Trials | Huang, C.; Shih, C.; Tsay, M.; Hsuan, S.; Tseng, Y.; Tsai, W.; Cheng, C. | 2020 | ResearchSquare | | | | 10.21203/rs.3.rs-83282/v1 | Exclusion reason: Wrong publication type (e.g. conference abstract, preprints,...) |
| Huang 2019 | Efficacy and safety of Shenfu injection for septic shock: A systematic review and meta-analysis of randomized controlled trials | Huang, P.; Guo, Y.; Feng, S.; Zhao, G.; Li, B.; Liu, Q. | 2019 | The American journal of emergency medicine | 37 | 12 | 2197-2204 | 10.1016/j.ajem.2019.03.032 | Exclusion reason: Other intervention than reported in the SSCs |
| Huang 2024 | Effect of ultrashort-acting β-blockers on 28-day mortality in patients with sepsis with persistent tachycardia despite initial resuscitation: a meta-analysis of randomized controlled trials and trial sequential analysis | Huang, P.; Liu, F.; Hu, X.; Li, B.; Xu, X.; Liu, Q. | 2024 | Frontiers in pharmacology | 15 | | 1380175 | 10.3389/fphar.2024.1380175 | Exclusion reason: Other intervention than reported in the SSCs |
| Huang 2021 | The Efficacy and Safety of Esmolol for Septic Shock: A Systematic Review and Meta-analysis of Randomized Controlled Trials | Huang, P.; Zheng, X.; Liu, Z.; Fang, X. | 2021 | Frontiers in pharmacology | 12 | | 682232 | 10.3389/fphar.2021.682232 | Exclusion reason: Other intervention than reported in the SSCs |

| | | | | | | | | | |
|---|---|---|---|---|---|---|---|---|---|
| Hui 2022 | Linezolid Administration to Critically Ill Patients: Intermittent or Continuous Infusion? A Systematic Literature Search and Review | Hui, L. A.; Bodolea, C.; Vlase, L.; Hiriscau, E. I.; Popa, A. | 2022 | Antibiotics (Basel, Switzerland) | 11 | 4 | | 10.3390/antibiotics11040436 | Exclusion reason: Wrong study type (no systematic-/mini-/rapid review) |
| Iardino 2020 | Use of sepsis bundle in low and middle income countries: metanalysis and systematic review of literature | Iardino, A.; Ciezki, R.; Modi, K.; Dy, R. V.; Garner, O.; Barragan, A. M.; Dominguez, C.; Bilgrami, M.; Oud, L. | 2020 | Am. J. Respir. Crit. Care Med. | 201 | 1 | | | Exclusion reason: Wrong publication type (e.g. conference abstract, preprints,...) |
| IsmailHassan 2020 | A Review on The Protective Effects of Metformin in Sepsis-Induced Organ Failure | Ismail Hassan, F.; Didari, T.; Khan, F.; Niaz, K.; Mojtahedzadeh, M.; Abdollahi, M. | 2020 | Cell journal | 21 | 4 | 363-370 | 10.22074/cellj.2020.6286 | Exclusion reason: Wrong study type (no systematic-/mini-/rapid review) |
| Jaiswal 2016 | Feasibility and efficacy of gentamicin for treating neonatal sepsis in community-based settings: a systematic review | Jaiswal, N.; Singh, M.; Kondel, R.; Kaur, N.; Thumburu, K. K.; Kumar, A.; Kaur, H.; Chadha, N.; Gupta, N.; Agarwal, A.; Malhotra, S.; Shafiq, N. | 2016 | World journal of pediatrics : WJP | 12 | 4 | 408-414 | 10.1007/s12519-016-0005-2 | Exclusion reason: Wrong population (e.g. animals and humans pooled together, sepsis only as subgroup,...) |

| | | | | | | | | | |
|---|---|---|---|---|---|---|---|---|---|
| Janssens 2016 | [Early goal-directed therapy for septic shock : Systematic review and meta-analysis] | Janssens, U. | 2016 | Medizinische Klinik, Intensivmedizin und Notfallmedizin | 111 | 6 | 551-555 | 10.1007/s00063-015-0108-x | Exclusion reason: Wrong study type (no systematic-/mini-/rapid review) |
| Jianfei 2018 | Xuebijing combined with ulinastation benefits patients with sepsis: A meta-analysis | Jianfei, Zheng; Xudong, Xiang; Bing, Xiao; Huixia, Li; Xun, Gong; Shuo, Yao; Ting, Yuan | 2018 | American Journal of Emergency Medicine | 36 | 3 | 480-487 | 10.1016/j.ajem.2017.12.007 | Exclusion reason: Other intervention than reported in the SSCs |
| Jiang 2021 | Is restrictive fluid resuscitation beneficial not only for hemorrhagic shock but also for septic shock?: A meta-analysis | Jiang, S.; Wu, M.; Lu, X.; Zhong, Y.; Kang, X.; Song, Y.; Fan, Z. | 2021 | Medicine | 100 | 12 | e25143 | 10.1097/MD.0000000000025143 | Exclusion reason: Wrong population (e.g. animals and humans pooled together, sepsis only as subgroup,...) |
| Jiesisibieke 2020 | The relationship between different dialysis methods and septicemia: a systematic review and meta-analysis | Jiesisibieke, Z. L.; Zhang, S.; Chien, C. W.; Tung, T. H. | 2020 | Renal Fail. | 42 | 1 | 567-569 | 10.1080/0886022X.2020.1776733 | Exclusion reason: Wrong population (e.g. animals and humans pooled together, sepsis only as subgroup,...) |
| Jin 2021 | Efficacy of beta-blockers in the treatment of sepsis | Jin, P.; Zhao, T.; Wei, Y.; Zhao, F. | 2021 | Banladesh J. Pharm. | 16 | 1 | 1-18 | 10.3329/bjp.v16i1.46001 | Exclusion reason: Other intervention than reported in the SSCs |
| Julián-Jiménez 2024 | Early source control of infection in patients seen in the emergency department: a systematic review | Julián-Jiménez, A.; Lorenzo Álvarez, R.; Gutiérrez Bueno, V.; Sánchez Trujillo, M.; Eduardo García, D. | 2024 | Revista espanola de quimioterapia : publicacion oficial de la Sociedad Espanola de Quimioterapia | 37 | 4 | 323-333 | 10.37201/req/027.2024 | Exclusion reason: Wrong language |

| | | | | | | | | | |
|---|---|---|---|---|---|---|---|---|---|
| Kabil 2022 | Early fluid bolus in adults with sepsis in the emergency department: a systematic review, meta-analysis and narrative synthesis | Kabil, G.; Frost, S. A.; Hatcher, D.; Shetty, A.; Foster, J.; McNally, S. | 2022 | BMC emergency medicine | 22 | 1 | 3 | 10.1186/s12873-021-00558-5 | Exclusion reason: Wrong population (e.g. animals and humans pooled together, sepsis only as subgroup,...) |
| Karimi 2021 | A comprehensive systematic review of the therapeutic effects and mechanisms of action of quercetin in sepsis | Karimi, A.; Naeini, F.; Asghari Azar, V.; Hasanzadeh, M.; Ostadrahimi, A.; Niazkar, H. R.; Mobasseri, M.; Tutunchi, H. | 2021 | Phytomedicine : international journal of phytotherapy and phytopharmacology | 86 | | 153567 | 10.1016/j.phymed.2021.153567 | Exclusion reason: Wrong study type (no systematic-/mini-/rapid review) |
| Kato 2023 | Efficacy and safety of recombinant human soluble thrombomodulin in patients with sepsis-induced disseminated intravascular coagulation - A meta-analysis | Kato, H.; Hagihara, M.; Asai, N.; Umemura, T.; Hirai, J.; Mori, N.; Yamagishi, Y.; Iwamoto, T.; Mikamo, H. | 2023 | Thromb Res | 226 | | 165-172 | 10.1016/j.thromres.2023.05.009 | Exclusion reason: Other intervention than reported in the SSCs |
| Khalifa 2024 | Surgical Versus Conservative Management of Delayed Presentation of Acute Biliary Disease: A Systematic Literature Review | Khalifa, A.; Allami, S. J.; Tahhan, O.; Alhaj, S. S.; Al Tahan, M. A.; Elnogoomi, I. | 2024 | Cureus | 16 | 11 | e74237 | 10.7759/cureus.74237 | Exclusion reason: Other intervention than reported in the SSCs |
| Khalil 2020 | Neonatal early onset sepsis in Middle | Khalil, N.; Blunt, H. B.; | 2020 | Archives of disease in childhood | 105 | 7 | 639-647 | 10.1136/archdischild- | Exclusion reason: Wrong type of systematic review (e.g. |

| | | | | | | | | | |
|---|---|---|---|---|---|---|---|---|---|
| | Eastern countries: a systematic review | Li, Z.; Hartman, T. | | | | | | 2019-317110 | diagnostic (test accuracy), experiential (qualitative), prognostic, etiology and/or risk, prevalence and/or incidence reviews) |
| Khattar 2024 | Unravelling the Fluid Paradox in End Stage Kidney Disease and Sepsis: A Meta-analysis | Khattar, G.; El Gharib, K.; Ngowari, P.; Kotys, J.; Kandala, V.; Salman, K.; El Hage, H.; Flamenbaum, M.; Dany, E.; Chalhoub, M. N.; Suzanne, E. | 2024 | Am. J. Respir. Crit. Care Med. | 209 | | | | Exclusion reason: Wrong publication type (e.g. conference abstract, preprints,...) |
| Khokher 2021 | THE ROLE OF ASCORBIC ACID IN REDUCING THE INCIDENCE OF STAGE 3 ACUTE KIDNEY INJURY IN PATIENTS WITH SEPTIC SHOCK: A SYSTEMATIC REVIEW AND META-ANALYSIS | Khokher, W.; Kesireddy, N.; herath mudiyanselage, P.; Iftikhar, S.; Beran Beran, A.; Alom, M.; Rashid, R.; Ali, H.; Awad, M. | 2021 | Chest | 160 | 4 | A1057 | 10.1016/j.chest.2021.07.979 | Exclusion reason: Wrong publication type (e.g. conference abstract, preprints,...) |
| Ang 2024 | ADJUNCTIVE USE OF MIDODRINE IN PATIENTS WITH SEPTIC SHOCK: A META-ANALYSIS | Ang, S. P.; Chia, J. E.; Yosry, N.; Chajkowski, A.; Valladares, C.; Jaiswal, V.; Qaiser, | 2024 | Chest | 166 | 4 | A2325-A2326 | 10.1016/j.chest.2024.06.1426 | Exclusion reason: Wrong publication type (e.g. conference abstract, preprints,...) |

| | | | | | | | | | |
|---|---|---|---|---|---|---|---|---|---|
| | | S.; Iglesias, J. I. | | | | | | | |
| Killu 2025 | The association between integrating echocardiography use in the management of septic shock patients and outcomes in the intensive care unit: a systematic review and meta-analysis | Killu, K.; Patino-Sutton, C.; Kysh, L.; Castriotta, R.; Oropello, J.; Huerta, L.; Engracia, D.; Merchant, K.; Wee, C. P.; Cortessis, V. K. | 2025 | J Ultrasound | 28 | 2 | 281-294 | 10.1007/s40477-024-00958-w | Exclusion reason: Other intervention than reported in the SSCs |
| Kochkin 2020 | Modern vasopressor therapy of septic shock (Review) | Kochkin, A. A.; Berikashvili, L. B.; Likhvantsev, V. V.; Yavorovskiy, A. G. | 2020 | Obshchaya Reanimatol. | 16 | 2 | 77-93 | 10.15360/1813-9779-2020-2-77-93 | Exclusion reason: Wrong study type (no systematic-/mini-/rapid review) |
| Kruger 2017 | Statins in patients with sepsis and ARDS: is it over? No | Kruger, Peter; Terblanche, Marius; Kruger, Peter S. | 2017 | Intensive Care Medicine | 43 | 5 | 675-676 | 10.1007/s00134-016-4564-4 | Exclusion reason: Wrong study type (no systematic-/mini-/rapid review) |
| Kubo 2022 | Short- versus prolonged-course antibiotic therapy for sepsis or infectious diseases in critically ill adults: a systematic review and meta-analysis | Kubo, K.; Kondo, Y.; Yoshimura, J.; Kikutani, K.; Shime, N. | 2022 | Infectious diseases (London, England) | 54 | 3 | 1-11 | 10.1080/23744235.2021.2001046 | Exclusion reason: Wrong population (e.g. animals and humans pooled together, sepsis only as subgroup,...) |
| Kyeremanteng 2018 | Cost analysis of Omega-3 supplementation in | Kyeremanteng, K.; Shen, J.; Thavorn, | 2018 | Clinical nutrition ESPEN | 25 | | 63-67 | 10.1016/j.clnesp.2018.04.003 | Exclusion reason: Wrong study type (no |

| | | | | | | | | | |
|---|---|---|---|---|---|---|---|---|---|
| | critically ill patients with sepsis | K.; Fernando, S. M.; Herritt, B.; Chaudhuri, D.; Tanuseputro, P. | | | | | | | systematic-/mini-/rapid review) |
| Lai 2020 | Acupuncture at Zusanli (ST36) for Experimental Sepsis: A Systematic Review | Lai, F.; Ren, Y.; Lai, C.; Chen, R.; Yin, X.; Tan, C.; Li, J.; Yang, C.; Liang, G.; Zeng, R. | 2020 | Evidence-based complementary and alternative medicine : eCAM | 2020 | | 3620741 | 10.1155/2020/3620741 | Exclusion reason: Wrong population (e.g. animals and humans pooled together, sepsis only as subgroup,...) |
| Lai 2015 | A Systematic Review of Rhubarb (a Traditional Chinese Medicine) Used for the Treatment of Experimental Sepsis | Lai, F.; Zhang, Y.; Xie, D. P.; Mai, S. T.; Weng, Y. N.; Du, J. D.; Wu, G. P.; Zheng, J. X.; Han, Y. | 2015 | Evidence-based complementary and alternative medicine : eCAM | 2015 | no pagination | 131283 | 10.1155/2015/131283 | Exclusion reason: Wrong population (e.g. animals and humans pooled together, sepsis only as subgroup,...) |
| Lat 2021 | The Surviving Sepsis Campaign: Fluid Resuscitation and Vasopressor Therapy Research Priorities in Adult Patients | Lat, I.; Coopersmith, C. M.; De Backer, D. | 2021 | Crit Care Med | 49 | 4 | 623-635 | 10.1097/ccm.0000000000004864 | Exclusion reason: Wrong study type (no systematic-/mini-/rapid review) |
| Lat 2021 | The surviving sepsis campaign: fluid resuscitation and vasopressor therapy research priorities in adult patients | Lat, I.; Coopersmith, C. M.; De Backer, D. | 2021 | Intensive Care Med Exp | 9 | 1 | 10 | 10.1186/s40635-021-00369-9 | Exclusion reason: Wrong study type (no systematic-/mini-/rapid review) |

| | | | | | | | | | |
|---|---|---|---|---|---|---|---|---|---|
| Lee 2015 | The effect of protocol-based resuscitation for septic shock: A meta-analysis | Lee, W. K.; Kim, J.; Na, S. | 2015 | Critical Care Medicine | 43 | 12 | 272 | 10.1097/01.ccm.0000474911.93364.ab | Exclusion reason: Wrong publication type (e.g. conference abstract, preprints,...) |
| Lee 2019 | Benefits of Beta-Blockade in Sepsis and Septic Shock: A Systematic Review | Lee, Y. R.; Seth, M. S.; Soney, D.; Dai, H. | 2019 | Clinical drug investigation | 39 | 5 | 429-440 | 10.1007/s40261-019-00762-z | Exclusion reason: Other intervention than reported in the SSCs |
| Abdollahi 2021 | The effect of L-carnitine on mortality rate in septic patients: A systematic review and meta-analysis on randomized clinical trials | Abdollahi, H.; Abdolahi, M.; Sedighiyan, M.; Jafarieh, A. | 2021 | Endocrine, metabolic & immune disorders drug targets | 21 | 4 | 673-681 | 10.2174/187153032066620072715045 0 | Exclusion reason: Other intervention than reported in the SSCs |
| Leone 2016 | Intravenous immunoglobins in sepsis: the potential risk of overinterpreting a well-done meta-analysis | Leone, M.; Duclos, G.; Coopersmith, C. M. | 2016 | Minerva anestesiologica | 82 | 5 | 505-7 | | Exclusion reason: Wrong study type (no systematic-/mini-/rapid review) |
| Li 2015 | Thymosin alpha1 based immunomodulatory therapy for sepsis: a systematic review and meta-analysis | Li, C.; Bo, L.; Liu, Q.; Jin, F. | 2015 | Int J Infect Dis | 33 | | 90-6 | 10.1016/j.ijid.2014.12.032 | Exclusion reason: Other intervention than reported in the SSCs |
| Li 2018 | Efficacy and safety of Xuebijing injection (a Chinese patent) for sepsis: A meta-analysis of randomized controlled trials | Li, C.; Wang, P.; Zhang, L.; Li, M.; Lei, X.; Liu, S.; Feng, Z.; Yao, Y.; Chang, B.; Liu, B.; Shang, H. | 2018 | Journal of ethnopharmacology | 224 | | 512-521 | 10.1016/j.jep.2018.05.043 | Exclusion reason: Other intervention than reported in the SSCs |
| Li 2020 | Prognosis of β-adrenergic blockade therapy on septic | Li, J.; Sun, W.; Guo, Y.; | 2020 | Cytokine | 126 | | 154916 | 10.1016/j.cyto.2019.154916 | Exclusion reason: Other intervention than reported in the SSCs |

| | | | | | | | | | |
|---|---|---|---|---|---|---|---|---|---|
| | shock and sepsis: A systematic review and meta-analysis of randomized controlled studies | Ren, Y.; Li, Y.; Yang, Z. | | | | | | | |
| Li 2022 | Resveratrol: Potential Application in Sepsis | Li, J.; Zeng, X.; Yang, F.; Wang, L.; Luo, X.; Liu, R.; Zeng, F.; Lu, S.; Huang, X.; Lei, Y.; Lan, Y. | 2022 | Frontiers in pharmacology | 13 | | 821358 | 10.3389/fphar.2022.821358 | Exclusion reason: Wrong study type (no systematic-/mini-/rapid review) |
| Li 2020 | [A meta-analysis to evaluate the effects of corticosteroids on the outcomes of patients with septic shock] | Li, Q.; Xie, J. F.; Liu, L. | 2020 | Zhonghua nei ke za zhi | 59 | 6 | 451-459 | 10.3760/cma.j.cn112138-20190802-00536 | Exclusion reason: Wrong language |
| Li 2025 | Oral switch vs. continued intravenous antibiotic therapy in patients with bacteraemia and sepsis: a systematic review and meta-analysis | Li, Q.; Zhou, Q.; Fan, J.; Huang, S.; Chen, Y.; Song, F.; Fu, Z.; Liu, E.; Tang, D.; Zeng, L.; Luo, Z. | 2025 | Clin Microbiol Infect | 31 | 4 | 551-559 | 10.1016/j.cmi.2024.11.035 | Exclusion reason: Wrong population (e.g. animals and humans pooled together, sepsis only as subgroup,...) |
| Li 2019 | A meta-analysis of randomized controlled trials: Efficacy of selenium treatment for sepsis | Li, S.; Tang, T.; Guo, P.; Zou, Q.; Ao, X.; Hu, L.; Tan, L. | 2019 | Medicine | 98 | 9 | e14733 | 10.1097/MD.0000000000014733 | Exclusion reason: Wrong population (e.g. animals and humans pooled together, sepsis only as subgroup,...) |
| Li 2020 | [Effect of terlipressin on prognosis of adult septic shock patients: a Meta-analysis] | Li, W.; Pan, P.; Wang, Y.; Du, X.; Yu, X. | 2020 | Zhonghua wei zhong bing ji jiu yi xue | 32 | 2 | 134-139 | 10.3760/cma.j.cn121430-20200210-00025 | Exclusion reason: Wrong language |

| | | | | | | | | | |
|---|---|---|---|---|---|---|---|---|---|
| Li 2023 | Ameliorating effects of berberine on sepsis-associated lung inflammation induced by lipopolysaccharide: molecular mechanisms and preclinical evidence | Li, X.; Bai, Y.; Ma, Y.; Li, Y. | 2023 | Pharmacological reports : PR | | | | 10.1007/s43440-023-00492-2 | Exclusion reason: Wrong study type (no systematic-/mini-/rapid review) |
| Li 2021 | Association of Preadmission Metformin Use and Prognosis in Patients With Sepsis and Diabetes Mellitus: A Systematic Review and Meta-Analysis | Li, Y.; Zhao, H.; Guo, Y.; Duan, Y.; Ding, X. | 2021 | Frontiers in endocrinology | 12 | | 811776 | 10.3389/fendo.2021.811776 | Exclusion reason: Other intervention than reported in the SSCs |
| Li-juan 2018 | Meta analysis of clinical efficacy and safety of acerbic therapy for sepsis | Li-juan, Wang; Jian, L. I.; Xiao-juan, L. I. | 2018 | 中医药导报 | 24 | 23 | 86-90 | | Exclusion reason: Wrong language |
| Liang 2019 | Association of preadmission metformin use and mortality in patients with sepsis and diabetes mellitus: a systematic review and meta-analysis of cohort studies | Liang, H.; Ding, X.; Li, L.; Wang, T.; Kan, Q.; Wang, L.; Sun, T. | 2019 | Critical care (London, England) | 23 | 1 | 50 | 10.1186/s13054-019-2346-4 | Exclusion reason: Other intervention than reported in the SSCs |
| Liang 2015 | Efficacy of traditional Chinese medicine on sepsis: a systematic review and Meta-Analysis | Liang, X.; Zhou, M.; Ge, X. Y.; Li, C. B.; Fang, S. P.; Tang, L.; Shao, D. H.; Xu, G. | 2015 | International journal of clinical and experimental medicine | 8 | 11 | 20024-34 | | Exclusion reason: Other intervention than reported in the SSCs |
| Liao 2024 | Effect of shenfu injection in patients with septic shock: A | Liao, J.; Qin, C.; Wang, Z.; Gao, L.; | 2024 | J Ethnopharmacol | 320 | | 117431 | 10.1016/j.jep.2023.117431 | Exclusion reason: Other intervention than reported in the SSCs |

| | | | | | | | | | |
|---|---|---|---|---|---|---|---|---|---|
| | systemic review and meta-analysis for randomized clinical trials | Zhang, S.; Feng, Y.; Liu, J.; Tao, L. | | | | | | | |
| Lieberman 2022 | Plasma and Platelet Transfusion Strategies in Critically Ill Children With Malignancy, Acute Liver Failure and/or Liver Transplantation, or Sepsis: From the Transfusion and Anemia EXpertise Initiative-Control/Avoidance of Bleeding | Lieberman, L.; Karam, O.; Stanworth, S. J.; Goobie, S. M.; Crighton, G.; Goel, R.; Lacroix, J.; Nellis, M. E.; Parker, R. I.; Steffen, K.; Stricker, P.; Valentine, S. L.; Steiner, M. E. | 2022 | Pediatr Crit Care Med | 23 | 13 Suppl 1 1S | e37-e49 | 10.1097/pcc.0000000000002857 | Exclusion reason: Wrong study type (no systematic-/mini-/rapid review) |
| Lin 2023 | Efficacy and Safety of Citric Acid and Heparin Anticoagulation in Patients with Septic Acute Kidney Injury Undergoing Continuous Renal Replacement Therapy: A Meta-Analysis | Lin, S.; Xiao, M.; Cai, Q.; Lin, Y.; Yao, J.; Sun, X.; Huang, J. | 2023 | Alternative therapies in health and medicine | | | | | Exclusion reason: Other intervention than reported in the SSCs |
| Liu 2017 | Effect of ulinastatin combined with thymosin alpha1 on sepsis: A systematic review and meta-analysis of Chinese and Indian patients | Liu, D.; Yu, Z.; Yin, J.; Chen, Y.; Zhang, H.; Fan, X.; Fu, H.; Wan, B. | 2017 | Journal of Critical Care | 39 | | 285-287 | 10.1016/j.jcrc.2017.02.005 | Exclusion reason: Wrong study type (no systematic-/mini-/rapid review) |
| Liu 2017 | Effect of ulinastatin combined with thymosin alpha1 on | Liu, D.; Yu, Z.; Yin, J.; Chen, Y.; | 2017 | Journal of Critical Care | 39 | | 259-266 | 10.1016/j.jcrc.2016.12.013 | Exclusion reason: Other intervention than reported in the SSCs |

| | | | | | | | | | |
|---|---|---|---|---|---|---|---|---|---|
| | sepsis: A systematic review and meta-analysis of Chinese and Indian patients | Zhang, H.; Xin, F.; Fu, H.; Wan, B. | | | | | | | |
| Liu 2016 | The efficacy of thymosin α1 as immunomodulatory treatment for sepsis: a systematic review of randomized controlled trials | Liu, F.; Wang, H. M.; Wang, T.; Zhang, Y. M.; Zhu, X. | 2016 | BMC infectious diseases | 16 | 1 | 488 | 10.1186/s12879-016-1823-5 | Exclusion reason: Other intervention than reported in the SSCs |
| Liu 2017 | Early actate levelsfor prediction of mortality in patients with sepsis or septic shock: A meta-analysis | Liu, G.; Lv, H.; An, Y.; Wei, X.; Yi, X.; Yi, H. | 2017 | International Journal of Clinical and Experimental Medicine | 10 | 1 | 37-47 | | Exclusion reason: Wrong type of systematic review (e.g. diagnostic (test accuracy), experiential (qualitative), prognostic, etiology and/or risk, prevalence and/or incidence reviews) |
| Liu 2018 | The influence of esmolol on septic shock and sepsis: A meta-analysis of randomized controlled studies | Liu, P.; Wu, Q.; Tang, Y.; Zhou, Z.; Feng, M. | 2018 | The American journal of emergency medicine | 36 | 3 | 470-474 | 10.1016/j.ajem.2017.11.013 | Exclusion reason: Other intervention than reported in the SSCs |
| Lu 2015 | Effect of early goal-directed therapy on mortality in patients with severe sepsis or septic shock: A Meta analysis | Lu, X.; Li, T.; Li, J.; Gao, X.; Xu, L. | 2015 | Chinese Critical Care Medicine | 27 | 9 | 735-738 | 10.3760/cma.j.issn.2095-4352.2015.09.007 | Exclusion reason: Wrong language |
| Xing 2015 | [Effect of early goal-directed therapy on mortality in patients with severe sepsis or septic shock: a Meta analysis] | Xing, L.; Tong, L.; Jun, L.; Xinjing, G.; Lei, X. | 2015 | Zhonghua Wei Zhong Bing Ji Jiu Yi Xue | 27 | 9 | 735-8 | | Exclusion reason: Wrong language |

| | | | | | | | | | |
|---|---|---|---|---|---|---|---|---|---|
| Asner 2021 | Impact of the Timeliness of Antibiotic Therapy on the Outcome of Patients with Sepsis and Septic Shock | Asner, S. A.; Desgranges, F.; Schrijver, I. T.; Calandra, T. | 2021 | The Journal of infection | | | | 10.1016/j.jinf.2021.03.003 | Exclusion reason: Wrong study type (no systematic-/mini-/rapid review) |
| Assiut 2023 | Treatment Outcome With Antibiotic Use and Its Resistance Pattern Among Patient With Neonatal Sepsis | Assiut, University | 2023 | clinicaltrials.gov | | | | | Exclusion reason: Wrong publication type (e.g. conference abstract, preprints,...) |
| Marchetto 2022 | OUTCOMES OF PEDIATRIC FLUIDREFRACTORY SEPTIC SHOCK ACCORDING TO DIFFERENT INOTROPIC OR VASOACTIVE STRATEGIES: A SYSTEMATIC REVIEW AND META-ANALYSIS | Marchetto, L.; Zanetto, L.; Comoretto, R.; Amigoni, A.; Daverio, M. | 2022 | Pediatric Critical Care Medicine | 23 | 11 | | 10.1097/01.pcc.0000900396.77115.25 | Exclusion reason: Wrong publication type (e.g. conference abstract, preprints,...) |
| Marks 2020 | Is it time to change the standard duration of 48 hours antibiotic cover for term and late preterm infants with suspected early onset sepsis? | Marks, L.; De Waal, K.; Ferguson, J. K. | 2020 | J. Paediatr. Child Health | 56 | SUPPL 1 | 13-14 | 10.1111/jpc.14831 | Exclusion reason: Wrong publication type (e.g. conference abstract, preprints,...) |
| Martin 2019 | Crystalloids vs. colloids for fluid resuscitation in the Intensive Care Unit: A systematic review and meta-analysis | Martin, G. S.; Bassett, P. | 2019 | Journal of critical care | 50 | | 144-154 | 10.1016/j.jcrc.2018.11.031 | Exclusion reason: Wrong population (e.g. animals and humans pooled together, sepsis only as subgroup,...) |
| McChesney 2025 | Impact of Short-Acting Beta-Blockers on the Outcomes of Patients | McChesney, C.; Orozco, N.; Fiorini, | 2025 | Critical care medicine | 53 | 5 | 1125-1139 | 10.1097/CCM.0000000000006604 | Exclusion reason: Other intervention than reported in the SSCs |

| | | | | | | | | | |
|---|---|---|---|---|---|---|---|---|---|
| | with Septic Shock: A Systematic Review and Meta-Analysis | K.; Wong, M. Y. S.; Slessarev, M.; Prager, R.; Kao, R.; Leligdowicz, A.; Sharif, S.; Lewis, K.; Rochwerg, B.; Honarmand, K.; Ball, I. M.; Arntfield, R.; Houlton, R.; VanNynatten, L.; Basmaji, J. | | | | | | | |
| Mehta 2024 | Systematic review with expert consensus on use of extracorporeal hemoadsorption in septic shock: An Indian perspective | Mehta, Y.; Ansari, A. S.; Mandal, A. K.; Chatterjee, D.; Sharma, G. S.; Sathe, P.; Umraniya, P. V.; Paul, R.; Gupta, S.; Singh, V.; Singh, Y. P. | 2024 | World journal of critical care medicine | 13 | 1 | 89026 | 10.5492/wjccm.v13.i1.89026 | Exclusion reason: Wrong study type (no systematic-/mini-/rapid review) |
| Meng 2024 | The supplementation of l-carnitine in critically ill patients with sepsis: a systematic review and meta-analysis of randomized controlled trials | Meng, C.; Ma, Y.; Fu, N.; Li, J.; Sun, B.; Li, Z.; Wang, Q.; Liu, P. | 2024 | European journal of medical research | 29 | 1 | 488 | 10.1186/s40001-024-02087-w | Exclusion reason: Other intervention than reported in the SSCs |

| | | | | | | | | | |
|---|---|---|---|---|---|---|---|---|---|
| Messmer 2023 | Active fluid de-resuscitation in critically ill patients with septic shock: A systematic review and meta-analysis | Messmer, A. S.; Dill, T.; Müller, M.; Pfortmueller, C. A. | 2023 | European journal of internal medicine | 109 | | 89-96 | 10.1016/j.ejim.2023.01.009 | Exclusion reason: Other intervention than reported in the SSCs |
| Author 2019 | Effect of Dexmedetomidine on Duration of Mechanical Ventilation in Septic Patients: A Systematic Review and Meta-Analysis | Author, p; Jiang, J.; Zhang, Y.; Li, G.; Qiu, Z.; Levy, M.; Hu, B. | 2019 | ResearchSquare | | | | 10.21203/rs.2.10571/v2 | Exclusion reason: Wrong publication type (e.g. conference abstract, preprints,...) |
| Miskulin 2020 | The impact of prior statin therapy on the outcomes of community-acquired sepsis | Miskulin, I.; Krizic, M.; Santini, M.; Barsic, B. | 2020 | Infektol. Glas. | 40 | 1 | 20-34 | 10.37797/ig.40.1.4 | Exclusion reason: Wrong language |
| Moon 2023 | The Effectiveness of Early Enteral Nutrition on Clinical Outcomes in Critically Ill Sepsis Patients: A Systematic Review | Moon, S. J.; Ko, R. E.; Park, C. M.; Suh, G. Y.; Hwang, J.; Chung, C. R. | 2023 | Nutrients | 15 | 14 | | 10.3390/nu15143201 | Exclusion reason: Wrong population (e.g. animals and humans pooled together, sepsis only as subgroup,...) |
| Mostaghim 2020 | Evaluation for evidence of unrecognized heterogeneity in treatment effects of corticosteroids in septic shock: A meta-analysis | Mostaghim, A.; Aberegg, S. K.; Walkey, A. J. | 2020 | Am. J. Respir. Crit. Care Med. | 201 | 1 | | | Exclusion reason: Wrong publication type (e.g. conference abstract, preprints,...) |
| Mou 2015 | Clinical Effect of Shenfu Injection in Patients with Septic Shock: A Meta-Analysis and Systematic Review | Mou, Z.; Lv, Z.; Li, Y.; Wang, M.; Xu, Q.; Yu, X. | 2015 | Evidence-based complementary and alternative medicine : eCAM | 2015 | no pagination | 863149 | 10.1155/2015/863149 | Exclusion reason: Other intervention than reported in the SSCs |

| | | | | | | | | | |
|---|---|---|---|---|---|---|---|---|---|
| Mousavi 2016 | New horizon in the treatment of sepsis: a systematic review of alternative medicine | Mousavi, S.; Ghannadi, A.; Meidani, M. | 2016 | Journal of complementary & integrative medicine | 13 | 4 | 317-332 | 10.1515/jcim-2016-0003 | Exclusion reason: Wrong study type (no systematic-/mini-/rapid review) |
| Nagaraju 2024 | Steroids in the Management of Ionotropic-Resistant Septic Shock: A Comprehensive Review of Efficacy and Outcomes | Nagaraju, N.; Varma, A.; Meshram, R. J. | 2024 | Cureus | 16 | 8 | e67795 | 10.7759/cureus.67795 | Exclusion reason: Wrong study type (no systematic-/mini-/rapid review) |
| Nasir 2015 | Serum Procalcitonin Assay for Investigations and Clinical Management of Neonatal Sepsis: A Review | Nasir, I. A.; Mele, H. U.; Babayo, A.; Yahaya, F. | 2015 | Journal of Pediatric Infectious Diseases | 1 | 1 | 3-11 | 10.1055/s-0035-1554969 | Exclusion reason: Wrong study type (no systematic-/mini-/rapid review) |
| Navalha 2024 | EFFICACY AND SAFETY OF FLUDROCORTISONE PLUS HYDROCORTISONE VS HYDROCORTISONE ALONE OR PLACEBO IN PATIENTS WITH SEPTIC SHOCK: A SYSTEMATIC REVIEW AND META-ANALYSIS | Navalha, D.; Aljazeeri, J.; Villegas Roberson, M.; Pareja-Plata, J. O. S. E.; Guillen, C.; Advani, K. | 2024 | Chest | 166 | 4 | A2337-None | 10.1016/j.chest.2024.06.1432 | Exclusion reason: Wrong publication type (e.g. conference abstract, preprints,...) |
| Ndomba 2023 | Management of sepsis in a cirrhotic patient admitted to the intensive care unit: A systematic literature review | Ndomba, N.; Soldera, J. | 2023 | World journal of hepatology | 15 | 6 | 850-866 | 10.4254/wjh.v15.i6.850 | Exclusion reason: Wrong study type (no systematic-/mini-/rapid review) |
| Nellis 2022 | Executive Summary of Recommendations and Expert Consensus | Nellis, M. E.; Karam, O.; Valentine, S. | 2022 | Pediatric critical care medicine : a journal of the | 23 | 1 | 34-51 | 10.1097/PCC.0000000000002851 | Exclusion reason: Wrong study type (no |

| | | | | | | | | | |
|---|---|---|---|---|---|---|---|---|---|
| | for Plasma and Platelet Transfusion Practice in Critically Ill Children: From the Transfusion and Anemia EXpertise Initiative-Control/Avoidance of Bleeding (TAXI-CAB) | L.; Bateman, S. T.; Remy, K. E.; Lacroix, J.; Cholette, J. M.; Bembea, M. M.; Russell, R. T.; Steiner, M. E.; Goobie, S. M.; Tucci, M.; Stricker, P. A.; Stanworth, S. J.; Delaney, M.; Lieberman, L.; Muszynski, J. A.; Bauer, D. F.; Steffen, K.; Nishijima, D.; Ibla, J.; Emani, S.; Vogel, A. M.; Haas, T.; Goel, R.; Crighton, G.; Delgado, D.; Demetres, M.; Parker, R. I.; Pediatric Critical Care, Transfusion; Anemia Expertise Initiative— | | Society of Critical Care Medicine and the World Federation of Pediatric Intensive and Critical Care Societies | | | | | systematic-/mini-/rapid review) |

| | | | | | | | | | |
|---|---|---|---|---|---|---|---|---|---|
| | | Control/Avoidance of Bleeding, in collaboration with the Pediatric Critical Care Blood Research Network; the Pediatric Acute Lung, Injury; Sepsis Investigators, Network | | | | | | | |
| Ng 2024 | The use of methylene blue in adult patients with septic shock: a systematic review and meta-analysis | Ng, K. T.; Kwok, P. E.; Lim, W. E.; Teoh, W. Y.; Hasan, M. S.; Abidin, M. F. Z. | 2024 | Brazilian journal of anesthesiology (Elsevier) | 75 | 1 | 844580 | 10.1016/j.bjane.2024.844580 | Exclusion reason: Other intervention than reported in the SSCs |
| North 2024 | Efficacy of Antibiotic Regimens for Sepsis or Possible Serious Bacterial Infection in Young Infants Aged 0 to 59 Days: A Systematic Review and Meta-analysis | North, K.; Mathias, S.; Schmeck, N.; Kim, Y.; Kehoe, T.; Folger, L. V.; Hoey, A.; Wade, C.; Driker, S.; Chou, R.; Edmond, K. M.; Lee, A. C. | 2024 | Pediatrics | 154 | Suppl 1 | | 10.1542/peds.2024-066588F | Exclusion reason: Wrong setting (e.g. out-of-hospital settings) |
| Nugent 2022 | Does Fluid Administration Based on Fluid Responsiveness Tests | Nugent, K.; Berdine, G.; Pena, C. | 2022 | Curr Cardiol Rev | 18 | 5 | 18-23 | 10.2174/1573403x18666220304202556 | Exclusion reason: Wrong study type (no systematic-/mini-/rapid review) |

| | | | | | | | | | |
|---|---|---|---|---|---|---|---|---|---|
| | such as Passive Leg Raising Improve Outcomes in Sepsis? | | | | | | | | |
| Ouyang 2019 | Effects of antiplatelet therapy on the mortality rate of patients with sepsis: A meta-analysis | Ouyang, Y.; Wang, Y.; Liu, B.; Ma, X.; Ding, R. | 2019 | Journal of critical care | 50 | | 162-168 | 10.1016/j.jcrc.2018.12.004 | Exclusion reason: Other intervention than reported in the SSCs |
| Pammi 2019 | Enteral lactoferrin for the treatment of sepsis and necrotizing enterocolitis in neonates | Pammi, M.; Abrams, S. A. | 2019 | The Cochrane database of systematic reviews | 5 | 5 | CD007138 | 10.1002/14651858.CD007138.pub4 | Exclusion reason: Other intervention than reported in the SSCs |
| Pammi 2015 | Pentoxifylline for treatment of sepsis and necrotizing enterocolitis in neonates | Pammi, M.; Haque, K. N. | 2015 | Cochrane Database Syst Rev | | 3 | Cd004205 | 10.1002/14651858.CD004205.pub3 | Exclusion reason: Other intervention than reported in the SSCs |
| Pammi 2023 | Pentoxifylline for treatment of sepsis and necrotising enterocolitis in neonates | Pammi, M.; Haque, K. N. | 2023 | The Cochrane database of systematic reviews | 6 | 6 | CD004205 | 10.1002/14651858.CD004205.pub4 | Exclusion reason: Other intervention than reported in the SSCs |
| Papp 2023 | Procalcitonin-guided antibiotic therapy may shorten length of treatment and may improve survival—a systematic review and meta-analysis | Papp, M.; Kiss, N.; Baka, M.; Trásy, D.; Zubek, L.; Fehérvári, P.; Harnos, A.; Turan, C.; Hegyi, P.; Molnár, Z. | 2023 | Critical care (London, England) | 27 | 1 | 394 | 10.1186/s13054-023-04677-2 | Exclusion reason: Wrong population (e.g. animals and humans pooled together, sepsis only as subgroup,...) |
| Peng 2019 | Ineffectiveness of Procalcitonin-guided Antibiotic Therapy in Severely Critically Ill Patients: A Meta-Analysis | Peng, F.; Chang, W.; Xie, J. F.; Sun, Q.; Qiu, H. B.; Yang, Y. | 2019 | International journal of infectious diseases : IJID : official publication of | 85 | | 158-166 | 10.1016/j.ijid.2019.05.034 | Exclusion reason: Wrong population (e.g. animals and humans pooled together, sepsis only as subgroup,...) |

| | | | | | | | | | |
|---|---|---|---|---|---|---|---|---|---|
| | | | | the International Society for Infectious Diseases | | | | | |
| Peng 2019 | Influency of pentoxifylline treatment for neonatal sepsis: A meta-analysis of randomized controlled studies | Peng, P.; Xia, Y. | 2019 | Hong Kong Journal of Emergency Medicine | 29 | 2 | 121-128 | 10.1177/1024907919864248 | Exclusion reason: Other intervention than reported in the SSCs |
| Bajgai 2024 | Naringin: A flavanone with a multifaceted target against sepsis-associated organ injuries | Bajgai, B.; Suri, M.; Singh, H.; Hanifa, M.; Bhatti, J. S.; Randhawa, P. K.; Bali, A. | 2024 | Phytomedicine : international journal of phytotherapy and phytopharmacology | 130 | | 155707 | 10.1016/j.phymed.2024.155707 | Exclusion reason: Wrong study type (no systematic-/mini-/rapid review) |
| Perala 2024 | Efficacy of β-Blockers in Decreasing Mortality in Sepsis and Septic Shock Patients: A Systematic Review | Perala, A.; Wishart, A. V.; Hamouda, R. K.; Elsaady, E.; Aslam, M. R.; Khan, S. | 2024 | Cureus | 16 | 8 | e66888 | 10.7759/cureus.66888 | Exclusion reason: Other intervention than reported in the SSCs |
| Pertzov 2019 | Hydroxymethylglutaryl-CoA reductase inhibitors (statins) for the treatment of sepsis in adults - a systematic review and meta-analysis | Pertzov, B.; Eliakim-Raz, N.; Atamna, H.; Trestioreanu, A. Z.; Yahav, D.; Leibovici, L. | 2019 | Clinical microbiology and infection : the official publication of the European Society of Clinical Microbiology and Infectious Diseases | 25 | 3 | 280-289 | 10.1016/j.cmi.2018.11.003 | Exclusion reason: Other intervention than reported in the SSCs |
| Pfortmueller 2025 | Correction to: Fluid accumulation syndrome in sepsis and septic shock: | Pfortmueller, C. A.; Dabrowski, W.; Malbrain, | 2025 | Ann. Intensive Care | 15 | 1 | | 10.1186/s13613-024-01403-1 | Exclusion reason: Wrong study type (no systematic-/mini-/rapid review); Correction to |

| | | | | | | | | | |
|---|---|---|---|---|---|---|---|---|---|
| | pathophysiology, relevance and treatment—a comprehensive review (Annals of Intensive Care, (2024), 14, 1, (115), 10.1186/s13613-024-01336-9) | M. L. N. G.; Wise, R.; van Regenmortel, N. | | | | | | | the original publication: https://pubmed.ncbi.nlm.nih.gov/39033219/; our PubMed search did neither identify the correction nor the original publication. In Epistemonikos the correction is listed, but not the original publication. The original publication is not a systematic review and was therefore, correctly not identified with our PubMed search. |
| Po-Yu 2024 | Effect of Ultrashort-Acting B-Blocker on the Mortality of Patients with Sepsis or Septic Shock: A Systematic Review and Trial Sequential Meta-Analysis of Randomized Controlled Trials | Po-Yu, Huang; Ting-Hui, Liu; Jheng-Yan, Wu; Ya-Wen, Tsai; Wan-Hsuan, Hsu; Min-Hsiang, Chuang; Hung-Jen, Tang; Chih-Cheng, Lai | 2024 | SSRN | | | | 10.2139/ssrn.4819264 | Exclusion reason: Wrong publication type (e.g. conference abstract, preprints,...) |
| Poggi 2023 | New Antimicrobials for the Treatment of Neonatal Sepsis Caused by Multi-Drug-Resistant Bacteria: A Systematic Review | Poggi, C.; Dani, C. | 2023 | Antibiotics (Basel, Switzerland) | 12 | 6 | | 10.3390/antibiotics12060956 | Exclusion reason: Wrong study type (no systematic-/mini-/rapid review) |
| Ballarin 2024 | Methylene blue in sepsis and septic shock: a systematic | Ballarin, R. S.; Lazzarin, T.; Zornoff, L.; Azevedo, | 2024 | Frontiers in medicine | 11 | | 1366062 | 10.3389/fmed.2024.1366062 | Exclusion reason: Other intervention than reported in the SSCs |

| | | | | | | | | | |
|---|---|---|---|---|---|---|---|---|---|
| | review and meta-analysis | P. S.; Pereira, F. W. L.; Tanni, S. E.; Minicucci, M. F. | | | | | | | |
| Pongsuwun 2024 | Factor Impacting Quality of Life Among Sepsis Survivors During and After Hospitalization: A Systematic Review of Current Empirical Evidence | Pongsuwun, K.; Puwarawuttipanit, W.; Nguantad, S.; Samart, B.; Saikaew, K.; Ruksakulpiwat, S. | 2024 | Journal of multidisciplinary healthcare | 17 | | 3791-3802 | 10.2147/JMDH.S477873 | Exclusion reason: Wrong type of systematic review (e.g. diagnostic (test accuracy), experiential (qualitative), prognostic, etiology and/or risk, prevalence and/or incidence reviews) |
| Poon 2020 | Extracorporeal membrane oxygenation (ECMO) in adult septic shock: A systematic review and meta-analysis | Poon, W. H.; Ling, R. R.; Lin, Y. L.; Khai, N. Y. J.; MacLaren, G.; Ramanathan, K. | 2020 | ASAIO J. | 66 | SUPPL 3 | 7-None | | Exclusion reason: Wrong publication type (e.g. conference abstract, preprints,...) |
| Poon 2020 | Extracorporeal membrane oxygenation (ECMO) for adult septic shock: A systematic review and meta-analysis | Poon, W. H.; Ling, R. R.; Yeo, N. J. K.; Maclaren, G.; Ramanathan, K. | 2020 | Perfusion | 35 | 1 SUPPL | 216-None | 10.1177/0267659120909723 | Exclusion reason: Wrong publication type (e.g. conference abstract, preprints,...) |
| Pourmand 2019 | The controversial role of corticosteroids in septic shock | Pourmand, A.; Whiteside, T.; Yamane, D.; Rashed, A.; Mazer-Amirshahi, M. | 2019 | The American journal of emergency medicine | 37 | 7 | 1353-1361 | 10.1016/j.ajem.2019.04.045 | Exclusion reason: Wrong study type (no systematic-/mini-/rapid review) |

| | | | | | | | | | |
|---|---|---|---|---|---|---|---|---|---|
| Procianoy 2020 | The challenges of neonatal sepsis management | Procianoy, R. S.; Silveira, R. C. | 2020 | Jornal de pediatria | 96 | | 80-86 | 10.1016/j.jped.2019.10.004 | Exclusion reason: Wrong study type (no systematic-/mini-/rapid review) |
| Pudpong 2025 | Evaluation of Standardized Treatment Regimens for Patients with Documented Melioidosis Septicemia at Tertiary Care Hospital in Thailand | Pudpong, K.; Gatechan, T. | 2025 | Open Forum Infect. Dis. | 12 | | S929-S930 | 10.1093/ofid/ofae631.1692 | Exclusion reason: Wrong publication type (e.g. conference abstract, preprints,...) |
| Qi 2023 | Effect of Anticoagulant Versus Non-Anticoagulant Therapy on Mortality of Sepsis-Induced Disseminated Intravascular Coagulation: A Systematic Review and Meta-Analysis | Qi, W.; Liu, J.; Li, A. | 2023 | Clinical and applied thrombosis/hemostasis : official journal of the International Academy of Clinical and Applied Thrombosis/Hemostasis | 29 | | | 10.1177/10760296231157766 | Exclusion reason: Other intervention than reported in the SSCs |
| Qi 2024 | Efficacy and Safety of Anticoagulant Therapy in Patients with Sepsis: A Meta-Analysis of Randomized Controlled Trials | Qi, Wang; QiuPing, Zheng; HuanHuan, Liu; Hong, Zhang | 2024 | SSRN | | | | 10.2139/ssrn.4781964 | Exclusion reason: Wrong publication type (e.g. conference abstract, preprints,...) |
| Qin 2024 | Prebiotics and sepsis in infants: An updated systematic review and meta-analysis | Qin, Y.; Pan, L. | 2024 | Advances in clinical and experimental medicine : official organ Wroclaw Medical University | 33 | 9 | 999-1005 | 10.17219/acem/174307 | Exclusion reason: Wrong type of systematic review (e.g. diagnostic (test accuracy), experiential (qualitative), prognostic, etiology and/or risk, prevalence |

| | | | | | | | | | |
|---|---|---|---|---|---|---|---|---|---|
| | | | | | | | | | and/or incidence reviews) |
| Qing 2016 | Efficacy of intravenous immunoglobulin in the adjuvant treatment of patients with severe sepsis: A Meta analysis | Qing, Q.; Luo, Y.; Yan, S.; Zhu, H.; Huang, P.; Zhou, Z.; Shen, H.; Xie, B.; He, W.; Liu, X.; Li, Y.; Xu, Y. | 2016 | Zhonghua Wei Zhong Bing Ji Jiu Yi Xue | 28 | 5 | 407-412 | 10.3760/cma.j.issn.2095-4352.2016.05.006 | Exclusion reason: Wrong language |
| Qing 2016 | [Efficacy of intravenous immunoglobulin in the adjuvant treatment of patients with severe sepsis: a Meta analysis] | Qing, Z.; Luo, Y.; Yan, S.; Zhu, H.; Huang, P.; Zhou, Z.; Shen, H.; Xie, B.; He, W.; Liu, X.; Li, Y.; Xu, Y. | 2016 | Zhonghua wei zhong bing ji jiu yi xue | 28 | 5 | 407-12 | | Exclusion reason: Wrong language |
| Quinn 2018 | Recommendations for active correction of hypernatremia in volume-resuscitated shock or sepsis patients should be taken with a grain of salt: A systematic review | Quinn, J. W.; Sewell, K.; Simmons, D. E. | 2018 | SAGE open medicine | 6 | | | 10.1177/20503121187620 43 | Exclusion reason: Wrong study type (no systematic-/mini-/rapid review) |
| Quinn 2016 | Systematic review of statins in sepsis: There is no evidence of dose response | Quinn, M.; Moody, C.; Tunnicliffe, B.; Khan, Z.; Manji, M.; Gudibande, S.; Murphy, N.; Whitehouse, T.; Snelson, | 2016 | Indian Journal of Critical Care Medicine | 20 | 9 | 534-541 | 10.4103/0972-5229.190366 | Exclusion reason: Wrong population (e.g. animals and humans pooled together, sepsis only as subgroup,...) |

| | | | | | | | | | |
|---|---|---|---|---|---|---|---|---|---|
| | | C.; Veenith, T. | | | | | | | |
| Rababa 2022 | Sepsis assessment and management in critically Ill adults: A systematic review | Rababa, M.; Bani Hamad, D.; Hayajneh, A. A. | 2022 | PloS one | 17 | 7 | e0270711 | 10.1371/journal.pone.0270711 | Exclusion reason: Wrong population (e.g. animals and humans pooled together, sepsis only as subgroup,...) |
| Ragoonanan 2022 | Unanswered questions on use of hydrocortisone, ascorbic acid, and thiamine therapy in sepsis and septic shock | Ragoonanan, D.; Tran, N.; Modi, V.; Morgan Nickelsen, P. | 2022 | American journal of health-system pharmacy : AJHP : official journal of the American Society of Health-System Pharmacists | 79 | 19 | 1626-1633 | 10.1093/ajhp/zxac169 | Exclusion reason: Wrong study type (no systematic-/mini-/rapid review) |
| Rao 2016 | Probiotic supplementation and late-onset sepsis in preterm infants: A meta-analysis | Rao, S.; Athayle-Jape, G.; Deshpande, G.; Simmer, K.; Patole, S. | 2016 | Journal of Paediatrics and Child Health | | | 56-57 | 10.1111/jpc.13194 | Exclusion reason: Wrong publication type (e.g. conference abstract, preprints,...) |
| Rao 2016 | Probiotic Supplementation and Late-Onset Sepsis in Preterm Infants: A Meta-analysis | Rao, S. C.; Athalye-Jape, G. K.; Deshpande, G. C.; Simmer, K. N.; Patole, S. K. | 2016 | Pediatrics | 137 | 3 | e20153684 | 10.1542/peds.2015-3684 | Exclusion reason: Wrong population (e.g. animals and humans pooled together, sepsis only as subgroup,...) |
| Reddy 2023 | Pediatric Sepsis Requiring Intensive Care Admission: Potential Structured Follow-Up Protocols to Identify and Manage New or Exacerbated Medical Conditions | Reddy, A. R.; Stinson, H. R.; Alcamo, A. M.; Pinto, N. P.; Fitzgerald, J. C. | 2023 | Risk management and healthcare policy | 16 | | 1881-1891 | 10.2147/RMHP.S394458 | Exclusion reason: Other intervention than reported in the SSCs |

| | | | | | | | | | |
|---|---|---|---|---|---|---|---|---|---|
| Reinert 2025 | Thiamine and Ascorbic Acid in Sepsis and Septic Shock: A Review of Evidence for their Role in Practice | Reinert, J. P.; Becker, K.; Ohlinger, M. J. | 2025 | J Pharm Technol | | | | 10.1177/8755122525132 0873 | Exclusion reason: Wrong study type (no systematic-/mini-/rapid review) |
| ReintamBlaser 2023 | Intravenous vitamin C therapy in adult patients with sepsis: A rapid practice guideline | Reintam Blaser, A.; Alhazzani, W.; Belley-Cote, E.; Møller, M. H.; Adhikari, N. K. J.; Burry, L.; Coopersmith, C. M.; Al Duhailib, Z.; Fujii, T.; Granholm, A.; Gunst, J.; Hammond, N.; Ke, L.; Lamontagne, F.; Loudet, C.; Morgan, M.; Ostermann, M.; Reinikainen, M.; Rosenfeld, R.; Spies, C.; Oczkowski, S. | 2023 | Acta anaesthesiologica Scandinavica | | | | 10.1111/aas.14311 | Exclusion reason: Wrong study type (no systematic-/mini-/rapid review) |
| ReyesZamora 2021 | Biomodulina T® as an alternative to the treatment of septic shock in pediatrics | Reyes Zamora, M. C.; Rodríguez, Y. | 2021 | Revista Habanera de Ciencias Medicas | 20 | 6 | | | Exclusion reason: Wrong language |

| | | | | | | | | | |
|---|---|---|---|---|---|---|---|---|---|
| | | A.; Chávez, S. R. | | | | | | | |
| Robey 2024 | Immunomodulatory drugs in sepsis: a systematic review and meta-analysis | Robey, R. C.; Logue, C.; Caird, C. A.; Hansel, J.; Hellyer, T. P.; Simpson, J.; Dark, P.; Mathioudakis, A. G.; Felton, T. | 2024 | Anaesthesia | 79 | 8 | 869-879 | 10.1111/anae.16263 | Exclusion reason: Other intervention than reported in the SSCs |
| Rothrock 2020 | Outcome of immediate vs early antibiotics in severe sepsis and septic shock: A meta-analysis | Rothrock, S. G.; Nguyen, L.; Earwood, R.; Cassidy, D. D.; Briscoe, J. G.; Barneck, M.; Guetschow, B.; Myburg, C.; Schinkel, M.; Nanayakkara, P.; Panday, R. N. | 2020 | Acad. Emerg. Med. | 27 | | S21-None | 10.1111/acem.13961 | Exclusion reason: Wrong publication type (e.g. conference abstract, preprints,...) |
| Sæterdal 2015 | NIPH Systematic Reviews | Sæterdal, I.; Holte, H. H.; Harboe, I.; Klemp, M. | 2015 | Effect of Using Aminoglycosides for Treatment of Sepsis | | | | | Exclusion reason: Wrong publication type (e.g. conference abstract, preprints,...) |
| Sahebnasagh 2022 | EFFECT OF ASCORBIC ACID, CORTICOSTEROIDS, AND THIAMINE (HAT THERAPY) FOR THE TREATMENT OF SEPSIS OR SEPTIC SHOCK: A | Sahebnasagh, A.; Boostani Moghadam, Z.; Salehi-Abargouei, A.; Saghafi, F. | 2022 | Chest | 161 | 1 | A213 | 10.1016/j.chest.2021.12.244 | Exclusion reason: Wrong publication type (e.g. conference abstract, preprints,...) |

| | | | | | | | | | |
|---|---|---|---|---|---|---|---|---|---|
| | SYSTEMATIC REVIEW AND META-ANALYSIS | | | | | | | | |
| Sanfilippo 2015 | Beta-blockers use in severe sepsis and septic shock: a systematic review | Sanfilippo, F.; Santonocito, C.; Morelli, A.; Foex, P. | 2015 | Current medical research and opinion | 31 | 10 | 1-24 | 10.1185/03007995.2015.1062357 | Exclusion reason: Other intervention than reported in the SSCs |
| Sangli 2020 | A Systematic Review of Preexisting Sepsis and Extracorporeal Membrane Oxygenation | Sangli, S. S.; Noronha, S. F.; Mourad, B.; Jean, R.; Bohman, J. K.; Seelhammer, T. G. | 2020 | ASAIO journal (American Society for Artificial Internal Organs : 1992) | 66 | 1 | 1-7 | 10.1097/MAT.0000000000000934 | Exclusion reason: Wrong study type (no systematic-/mini-/rapid review) |
| Saran 2023 | Role of Mycobacterium indicus pranii in clinical outcomes in patients with sepsis: A systematic review and meta-analysis of randomised controlled trials | Saran, K.; Iyengar, S. S.; Sinha, N.; Abhishek, K.; Kumar, A.; Prakash, J. | 2023 | Indian journal of anaesthesia | 67 | 12 | 1029-1035 | 10.4103/ija.ija_726_23 | Exclusion reason: Other intervention than reported in the SSCs |
| Sartini 2024 | Beyond the Surviving Sepsis Campaign Guidelines: a systematic review of interventions affecting mortality in sepsis | Sartini, C.; Landoni, G.; Belletti, A.; Kotani, Y.; Maimeri, N.; Umbrello, M.; Yavorovskiy, A.; Jabaudon, M. | 2024 | Panminerva Med | 66 | 1 | 55-62 | 10.23736/s0031-0808.23.04986-8 | Exclusion reason: Wrong study type (no systematic-/mini-/rapid review) |
| Sato 2025 | Mortality in Patients With Sepsis Treated With Esmolol or Landiolol: A Systematic Review and Meta-Analysis of | Sato, R.; Messina, S.; Hasegawa, D.; Santonocito, C.; | 2025 | Chest | 167 | 1 | 121-138 | 10.1016/j.chest.2024.08.020 | Exclusion reason: Other intervention than reported in the SSCs |

| | | | | | | | | | |
|---|---|---|---|---|---|---|---|---|---|
| | Randomized Controlled Trials With Trial Sequential Analysis | Scimonello, G.; Sanfilippo, G.; Morelli, A.; Dugar, S.; Sanfilippo, F. | | | | | | | |
| Basu 2024 | PCR197 Health-related Quality of Life (HRQoL) Among Patients Who Have Survived an Episode of Sepsis in the United States: A Systematic Review | Basu, S.; Wei, Z. J.; Laor, A.; Bennetts, L.; Khoury, Ac E.; Neary, M.; Geurtsen, J. | 2024 | Value Health | 27 | 6 | S333-None | 10.1016/j.jval.2024.03.2076 | Exclusion reason: Wrong publication type (e.g. conference abstract, preprints,...) |
| Shafique 2024 | EFFECT OF B-ADRENERGIC BLOCKADE THERAPY ON HEMODYNAMIC AND CLINICAL OUTCOMES IN PATIENTS WITH SEPTIC SHOCK: AN UPDATED SYSTEMIC REVIEW AND META-ANALYSIS | Shafique, N.; Ehsan, A. M. N. A.; Aleem, M.; Muhibullah, F. N. U.; Tariq, S. A.; Haider, A. L. I.; Shahid, M. A.; Lohana, A. B. H. I.; Ali, F.; Shafique, A.; Khan, M. A.; Adhikari, S.; Rasool, M. W.; Akbar, U.; Vohra, A.; Khan, S. A. N. A. | 2024 | Chest | 166 | 4 | A2081-A2082 | 10.1016/j.chest.2024.06.1289 | Exclusion reason: Other intervention than reported in the SSCs |
| Shan 2021 | Traditional Chinese medicine for septic patients undergoing ulinastatin therapy: A meta-analysis | Shan, R. F.; Zhu, Y. A.; Qin, J.; Chen, J. P. | 2021 | Medicine (Baltimore) | 100 | 38 | e27151 | 10.1097/md.0000000000027151 | Exclusion reason: Other intervention than reported in the SSCs |

| | | | | | | | | | |
|---|---|---|---|---|---|---|---|---|---|
| Bauer 2024 | Association Between Adjunctive Vasopressors and Short-term Mortality in Adults With Septic Shock: A Systematic Review and Meta-analysis | Bauer, S.; Sacha, G. L.; Wieruszewski, P. M.; Bissell Turpin, B. D.; Dugar, S.; Siuba, M.; Vachharajani, V.; Sato, R.; Schleicher, M.; Falck-Ytter, Y.; Morgan, R. L. | 2024 | Am. J. Respir. Crit. Care Med. | 209 | | | | Exclusion reason: Wrong publication type (e.g. conference abstract, preprints,...) |
| Shi 2017 | Xuebijing in the treatment of patients with sepsis | Shi, Heng; Hong, Yun; Qian, Jianfang; Cai, Xiaofang; Chen, Shanwen | 2017 | American Journal of Emergency Medicine | 35 | 2 | 285-291 | 10.1016/j.ajem.2016.11.007 | Exclusion reason: Other intervention than reported in the SSCs |
| Shi 2018 | Efficacy of esmolol for septic shock and sepsis: A meta-analysis of randomized controlled studies | Shi, K.; Hu, Y.; Huang, J.; Chen, Y.; Shen, Q. | 2018 | International Journal of Clinical and Experimental Medicine | 11 | 11 | 11458-11464 | | Exclusion reason: Other intervention than reported in the SSCs |
| Shrestha 2024 | Prior Use of Angiotensin-converting Enzyme Inhibitors or Angiotensin II Receptor Blockers and Clinical Outcomes of Sepsis and Septic Shock: A Systematic Review and Meta-analysis | Shrestha, D. B.; Sedhai, Y. R.; Oli, P. R.; Proskuriakova, E.; Adelkhanova, A.; Shtembari, J.; Khan, T. M. A.; Singh, K.; Ahmed, | 2024 | J Cardiovasc Pharmacol | 83 | 1 | 16-22 | 10.1097/fjc.0000000000001491 | Exclusion reason: Other intervention than reported in the SSCs |

| | | | | | | | | | |
|---|---|---|---|---|---|---|---|---|---|
| | | M. A.; Waheed, I.; Kazimuddin, N.; Steff, R.; Acharya, R.; Patel, N. K. | | | | | | | |
| Silversides 2016 | Conservative fluid management or deresuscitation for patients with sepsis or acute respiratory distress syndrome: A systematic review and meta-analysis | Silversides, J. A.; Major, E.; Mann, E. E.; Ferguson, A. J.; McAuley, D. F.; Marshall, J. C.; Blackwood, B.; Fan, E. | 2016 | Intensive Care Medicine Experimental | | | | 10.1186/s40635-016-0098-x | Exclusion reason: Wrong publication type (e.g. conference abstract, preprints,...) |
| Silverstein 2015 | Controversies regarding choice of vasopressor therapy for management of septic shock in animals | Silverstein, D. C.; Beer, K. A. | 2015 | Journal of veterinary emergency and critical care (San Antonio, Tex. : 2001) | 25 | 1 | 48-54 | 10.1111/vec.12282 | Exclusion reason: Wrong study type (no systematic-/mini-/rapid review) |
| Simonetti 2024 | VITAMIN B12 AND SEPTIC SHOCK MORTALITY: A SYSTEMATIC REVIEW AND META-ANALYSIS | Simonetti, G. R. E. G.; Desai, K.; Robinson, R. J.; Nguyen-Luu, T.; Sharma, N.; S, F. Naidoo; Shariff, R.; Sharma, R. | 2024 | Chest | 166 | 4 | A2610-None | 10.1016/j.chest.2024.06.1588 | Exclusion reason: Wrong publication type (e.g. conference abstract, preprints,...) |
| Smith 2025 | From Theory to Therapy: Methylene Blue's Emerging Role in the Management of Septic Shock | Smith, B. A.; Robinson, R.; Most, A. K. | 2025 | Journal of pharmacy practice | | | | 10.1177/08971900251350554 | Exclusion reason: Wrong study type (no systematic-/mini-/rapid review) |

| | | | | | | | | | |
|---|---|---|---|---|---|---|---|---|---|
| Smyth 2016 | Impact of Prehospital Care on Outcomes in Sepsis: A Systematic Review | Smyth, M. A.; Brace-McDonnell, S. J.; Perkins, G. D. | 2016 | The western journal of emergency medicine | 17 | 4 | 427-437 | 10.5811/westjem.2016.5.30172 | Exclusion reason: Wrong setting (e.g. out-of-hospital settings) |
| Song 2021 | The controversy and value of the combination of hydrocortisone, ascorbic acid and thiamine in treating sepsis | Song, W.; Wu, J. | 2021 | Nat. Med. J. China | 101 | 17 | 1206-1209 | 10.3760/cma.j.cn112137-20210107-00045 | Exclusion reason: Wrong language |
| Stahl 2021 | [Extracorporeal Strategies in Sepsis Treatment: Role of Therapeutic Plasma Exchange] | Stahl, K.; Bode, C.; David, S. | 2021 | Anasthesiologie, Intensivmedizin, Notfallmedizin, Schmerztherapie : AINS | 56 | 2 | 101-110 | 10.1055/a-1105-0572 | Exclusion reason: Wrong study type (no systematic-/mini-/rapid review) |
| Stefanos 2023 | RESTRICTIVE FLUIDS IN SEPSIS: A SYSTEMATIC REVIEW AND META-ANALYSIS | Stefanos, S. S.; Reynolds, P.; MacLaren, R. | 2023 | Critical Care Medicine | 51 | 1 | 32 | 10.1097/01.ccm.0000906128.52363.ea | Exclusion reason: Wrong publication type (e.g. conference abstract, preprints,...) |
| Sun 2022 | Adjuvant Application of Shenmai Injection for Sepsis: A Systematic Review and Meta-Analysis | Sun, Y.; Liu, Y.; Li, L.; Xue, B.; Cao, Y. | 2022 | Evidence-based complementary and alternative medicine : eCAM | 2022 | | 3710672 | 10.1155/2022/3710672 | Exclusion reason: Other intervention than reported in the SSCs |
| Sun 2025 | Efficacy of melatonin as adjunctive therapy for sepsis: A meta-analysis of randomized controlled trials | Sun, Z.; Yu, C.; Zhang, Z.; Hu, C.; Li, X.; Dong, X.; Zhang, R.; Zhu, T.; Su, X.; Guo, J. | 2025 | Complementary therapies in medicine | 89 | | 103147 | 10.1016/j.ctim.2025.103147 | Exclusion reason: Other intervention than reported in the SSCs |
| Sungkar 2018 | Implementation of guidelines for sepsis management in emergency | Sungkar, Y.; Considine, J.; | 2018 | Australasian emergency care | 21 | 4 | 111-120 | 10.1016/j.auec.2018.10.003 | Exclusion reason: Other intervention than reported in the SSCs |

| | | | | | | | | | |
|---|---|---|---|---|---|---|---|---|---|
| | departments: A systematic review | Hutchinson, A. | | | | | | | |
| Tadel 2024 | AI Algorithms for Modeling the Risk, Progression, and Treatment of Sepsis, Including Early-Onset Sepsis-A Systematic Review | Tadel, K.; Bil-Lula, I.; Dudek, A. | 2024 | J. Clin. Med. | 13 | 19 | | 10.3390/jcm13195959 | Exclusion reason: Wrong study type (no systematic-/mini-/rapid review) |
| Taito 2018 | Rehabilitation for patients with sepsis: A systematic review and meta-analysis | Taito, S.; Taito, M.; Banno, M.; Tsujimoto, H.; Kataoka, Y.; Tsujimoto, Y. | 2018 | PloS one | 13 | 7 | e0201292 | 10.1371/journal.pone.0201292 | Exclusion reason: Wrong population (e.g. animals and humans pooled together, sepsis only as subgroup,...) |
| | Correction: Rehabilitation for patients with sepsis: A systematic review and meta-analysis | Taito, S.; Taito, M.; Banno, M.; Tsujimoto, H.; Kataoka, Y.; Tsujimoto, Y. | 2019 | PLoS One | 14 | 8 | e0221224 | 10.1371/journal.pone.0221224 | |
| Tan 2019 | The association between premorbid beta blocker exposure and mortality in sepsis-a systematic review | Tan, K.; Harazim, M.; Tang, B.; McLean, A.; Nalos, M. | 2019 | Crit Care | 23 | 1 | 298 | 10.1186/s13054-019-2562-y | Exclusion reason: Other intervention than reported in the SSCs |
| | Correction to: The association between premorbid beta blocker exposure and mortality in sepsis-a systematic review | Tan, K.; Harazim, M.; Tang, B.; McLean, A.; Nalos, M. | 2020 | Crit Care | 24 | 1 | 3 | 10.1186/s13054-019-2699-8 | |
| Tan 2019 | The Association of Premorbid Metformin Exposure With Mortality and Organ Dysfunction in Sepsis: | Tan, K.; Simpson, A.; Huang, S.; Tang, B.; | 2019 | Critical care explorations | 1 | 4 | e0009 | 10.1097/CCE.0000000000000009 | Exclusion reason: Wrong type of systematic review (e.g. diagnostic (test accuracy), experiential |

| | | | | | | | | | |
|---|---|---|---|---|---|---|---|---|---|
| | A Systematic Review and Meta-Analysis | McLean, A.; Nalos, M. | | | | | | | (qualitative), prognostic, etiology and/or risk, prevalence and/or incidence reviews) |
| Tang 2020 | Effect of alkaline phosphatase on sepsis-associated acute kidney injury patients: A systematic review and meta-analysis | Tang, W.; Huang, J.; Huang, X.; Han, X.; Ke, G.; Xu, Q. | 2020 | Medicine (Baltimore) | 99 | 4 | e18788 | 10.1097/md.0000000000018788 | Exclusion reason: Other intervention than reported in the SSCs |
| Tang 2020 | Efficacy and safety of acupuncture on sepsis: A systematic review | Tang, Y.; Xu, M.; Wang, R.; Yang, J.; Liu, J. | 2020 | Chin. J. Evid.-Based Med. | 20 | 2 | 174-180 | 10.7507/1672-2531.201910006 | Exclusion reason: Wrong language |
| Tariq 2022 | Meta-analysis assessing the effectiveness of intravenous vitamin C in patients with sepsis and septic shock | Tariq, M. A.; Amin, H.; Ali, U. | 2022 | Acute and critical care | | | | 10.4266/acc.2022.01088 | Exclusion reason: Wrong study type (no systematic-/mini-/rapid review) |
| Teja 2020 | Vasopressor Dosing in Septic Shock Clinical Trials: A Systematic Review and Ecologic Study | Teja, Bijan; Bosch, Nicholas A.; Walkey, Allan J.; Pinto, Ruxandra; Wunsch, Hannah | 2020 | Annals of the American Thoracic Society | 17 | 6 | 773-776 | | Exclusion reason: Wrong study type (no systematic-/mini-/rapid review) |
| Thomas 2015 | Statin therapy in critically-ill patients with severe sepsis: a review and meta-analysis of randomized clinical trials | Thomas, G.; Hraiech, S.; Loundou, A.; Truwit, J.; Kruger, P.; McAuley, D. F.; Papazian, L.; Roch, A. | 2015 | Minerva anestesiologica | 81 | 8 | 921-30 | | Exclusion reason: Other intervention than reported in the SSCs |

| | | | | | | | | | |
|---|---|---|---|---|---|---|---|---|---|
| Tian 2019 | Efficacy of pentoxifylline treatment for neonatal sepsis: a meta-analysis of randomized controlled studies | Tian, J.; Shen, P.; Pan, K.; Zhou, Q. | 2019 | Italian journal of pediatrics | 45 | 1 | 108 | 10.1186/s13052-019-0697-8 | Exclusion reason: Other intervention than reported in the SSCs |
| Tian 2021 | Retraction Note: Eficacy of pentoxifylline treatment for neonatal sepsis: a meta-analysis of randomized controlled studies | Tian, J.; Shen, P.; Pan, K.; Zhou, Q. | 2021 | Ital J Pediatr | 47 | 1 | 30 | 10.1186/s13052-021-00979-9 | Exclusion reason: Other intervention than reported in the SSCs |
| Tibi 2023 | Role of the Renin Angiotensin Aldosterone System in the Pathogenesis of Sepsis-Induced Acute Kidney Injury: A Systematic Review | Tibi, S.; Zeynalvand, G.; Mohsin, H. | 2023 | Journal of clinical medicine | 12 | 14 | | 10.3390/jcm12144566 | Exclusion reason: Other intervention than reported in the SSCs |
| Totoki 2024 | Effects of combination therapy of antithrombin and thrombomodulin for sepsis-associated disseminated intravascular coagulation: a systematic review and meta-analysis | Totoki, T.; Makino, Y.; Yamakawa, K.; Koami, H.; Wada, T.; Ito, T.; Iba, T. | 2024 | Thromb J | 22 | 1 | 10 | 10.1186/s12959-023-00579-z | Exclusion reason: Other intervention than reported in the SSCs |
| Triana 2021 | Randomized clinical trials targeting lps/tlr4/tnfa driven vasodilation in sepsis and septic shock: A systematic review and metanalysis | Triana, A.; Meza, J.; Del Rio-Pertuz, G.; Leiper, J. | 2021 | Am. J. Respir. Crit. Care Med. | 203 | 9 | | 10.1164/ajrccm-conference.2021.203.1_MeetingAbstracts.A2743 | Exclusion reason: Wrong publication type (e.g. conference abstract, preprints,...) |
| Tsuchida 2024 | Efficacy of antithrombin administration for | Tsuchida, T.; Makino, Y.; Wada, T.; | 2024 | Acute medicine & surgery | 11 | 1 | e950 | 10.1002/ams2.950 | Exclusion reason: Other intervention than reported in the SSCs |

| | | | | | | | | | |
|---|---|---|---|---|---|---|---|---|---|
| | patients with sepsis: A systematic review, meta-analysis, and meta-regression | Ushio, N.; Totoki, T.; Fujie, N.; Yasuo, S.; Matsuoka, T.; Koami, H.; Yamakawa, K.; Iba, T. | | | | | | | |
| Tu 2025 | The impact of telehealth in sepsis care: A systematic review | Tu, K. J.; Wymore, C.; Tchangalova, N.; Fuller, B. M.; Mohr, N. M. | 2025 | J Telemed Telecare | 31 | 1 | 3-13 | 10.1177/1357633x231170038 | Exclusion reason: Other intervention than reported in the SSCs |
| Umar 2024 | EVALUATION OF FLUID RESPONSIVENESS USING LEFT VENTRICULAR OUTFLOW TRACT VELOCITY TIME INTEGRAL IN PATIENTS WITH SEPSIS AND SEPTIC SHOCK: A SYSTEMATIC REVIEW | Umar, Z.; Haseeb Ul Rasool, M.; Mergen, D.; Parikh, A.; Hosna, A. U.; Patrizi, S.; Rauf, F.; Miller, D.; Iqbal, H.; Trandafirescu, T. D. | 2024 | Chest | 166 | 4 | A1798-A1799 | 10.1016/j.chest.2024.06.1130 | Exclusion reason: Wrong publication type (e.g. conference abstract, preprints,...) |
| Umemura 2016 | Efficacy and safety of anticoagulant therapy in three specific populations with sepsis: a meta-analysis of randomized controlled trials | Umemura, Y.; Yamakawa, K.; Ogura, H.; Yuhara, H.; Fujimi, S. | 2016 | Journal of thrombosis and haemostasis : JTH | 14 | 3 | 518-30 | 10.1111/jth.13230 | Exclusion reason: Other intervention than reported in the SSCs |
| Vagionas 2022 | Thromboinflammation in Sepsis and Heparin: A Review of Literature and Pathophysiology | Vagionas, D.; Papadakis, D. D.; Politou, M.; Koutsoukou, | 2022 | In vivo (Athens, Greece) | 36 | 6 | 2542-2557 | 10.21873/invivo.12991 | Exclusion reason: Wrong study type (no systematic-/mini-/rapid review) |

| | | | | | | | | | |
|---|---|---|---|---|---|---|---|---|---|
| | | A.; Vasileiadis, I. | | | | | | | |
| Vajdi 2023 | Effects of luteolin on sepsis: A comprehensive systematic review | Vajdi, M.; Karimi, A.; Karimi, M.; Abbasalizad Farhangi, M.; Askari, G. | 2023 | Phytomedicine : international journal of phytotherapy and phytopharmacology | 113 | | 154734 | 10.1016/j.phymed.2023.154734 | Exclusion reason: Wrong population (e.g. animals and humans pooled together, sepsis only as subgroup,...) |
| Vajdi 2023 | A comprehensive insight into the molecular and cellular mechanisms of action of resveratrol on complications of sepsis a systematic review | Vajdi, M.; Sefidmooye Azar, P.; Mahmoodpoor, A.; Dashti, F.; Sanaie, S.; Kiani Chalmardi, F.; Karimi, A. | 2023 | Phytotherapy research : PTR | 37 | 9 | 3780-3808 | 10.1002/ptr.7917 | Exclusion reason: Wrong population (e.g. animals and humans pooled together, sepsis only as subgroup,...) |
| Valeriani 2020 | Efficacy and safety of recombinant human soluble thrombomodulin in patients with sepsis-associated coagulopathy: a systematic review and meta-analysis | Valeriani, E.; Squizzato, A.; Gallo, A.; Porreca, E.; Vincent, J. L.; Iba, T.; Hagiwara, A.; Di Nisio, M. | 2020 | Journal of thrombosis and haemostasis : JTH | 18 | 7 | 1618-1625 | 10.1111/jth.14812 | Exclusion reason: Other intervention than reported in the SSCs |
| Varney 2022 | Prehospital administration of broad-spectrum antibiotics for sepsis patients: A systematic review and meta-analysis | Varney, J.; Motawea, K. R.; Kandil, O. A.; Hashim, H. T.; Murry, K.; Shah, J.; Shaheen, A.; Akwari, J.; Awad, A. K.; Rivera, A.; Mostafa, M. | 2022 | Health science reports | 5 | 3 | e582 | 10.1002/hsr2.582 | Exclusion reason: Wrong setting (e.g. out-of-hospital settings) |

| | | | | | | | | | |
|---|---|---|---|---|---|---|---|---|---|
| | | R.; Swed, S.; Awad, D. M. | | | | | | | |
| Vásquez-Tirado 2024 | Association of beta blockers and mortality in adults with septic shock: systematic review and meta-analysis of randomized clinical trial | Vásquez-Tirado, G. A.; Quispe-Castañeda, C. V.; Meregildo-Rodríguez, E. D.; Cuadra-Campos, M.; Segura-Plasencia, N. M.; Arbayza-Avalos, Y. K.; Alva-Guarniz, H. N.; Guzmán-Aguilar, W. M.; Zavaleta-Alaya, P. | 2024 | Frontiers in medicine | 11 | | 1448573 | 10.3389/fmed.2024.1448573 | Exclusion reason: Other intervention than reported in the SSCs |
| Velayati 2023 | Therapeutic effects and mechanisms of action of ginger and its bioactive components on inflammatory response, oxidative stress, the immune system, and organ failure in sepsis: a comprehensive systematic review | Velayati, A.; Vafa, M. R.; Sani'ee, N.; Darabi, Z. | 2023 | Nutrition reviews | 82 | 12 | 1800-1819 | 10.1093/nutrit/nuad156 | Exclusion reason: Wrong population (e.g. animals and humans pooled together, sepsis only as subgroup,...) |
| Verzosa 2019 | Granulocyte colony stimulating factor in improving outcomes of neonatal sepsis: A meta-analysis | Verzosa, G. S.; Yu, M. M. C. N.; Abat-Senen, K. A.; Quilendrino, M. I. O. | 2019 | Pediatrics | 144 | 2 | | 10.1542/peds.144.2-MeetingAbstract.699 | Exclusion reason: Wrong publication type (e.g. conference abstract, preprints,...) |

| | | | | | | | | | |
|---|---|---|---|---|---|---|---|---|---|
| Volbeda 2015 | Glucocorticosteroids for sepsis: systematic review with meta-analysis and trial sequential analysis | Volbeda, M.; Wetterslev, J.; Gluud, C.; Zijlstra, J. G.; van der Horst, I. C.; Keus, F. | 2015 | Intensive care medicine | 41 | 7 | 1220-34 | 10.1007/s00134-015-3899-6 | Exclusion reason: Wrong population (e.g. animals and humans pooled together, sepsis only as subgroup,...) |
| Wang 2016 | The Efficacy and Immunomodulatory Effects of Ulinastatin and Thymosin α1 for Sepsis: A Systematic Review and Meta-Analysis | Wang, F. Y.; Fang, B.; Qiang, X. H.; Yu, T. O.; Zhong, J. R.; Cao, J.; Zhou, L. X. | 2016 | BioMed research international | 2016 | no pagination | | 10.1155/2016/9508493 | Exclusion reason: Other intervention than reported in the SSCs |
| Wang 2019 | Improvement of sepsis prognosis by ulinastatin: A systematic review and meta-analysis of randomized controlled trials | Wang, H.; Liu, B.; Tang, Y.; Chang, P.; Yao, L.; Huang, B.; Lodato, R. F.; Liu, Z. | 2019 | Frontiers in pharmacology | 10 | | 1370 | 10.3389/fphar.2019.01370 | Exclusion reason: Other intervention than reported in the SSCs |
| Wang 2019 | Corrigendum: Improvement of Sepsis Prognosis by Ulinastatin: A Systematic Review and Meta-Analysis of Randomized Controlled Trials | Wang, H.; Liu, B.; Tang, Y.; Chang, P.; Yao, L.; Huang, B.; Lodato, R. F.; Liu, Z. | 2019 | Front Pharmacol | 10 | | 1697 | 10.3389/fphar.2019.01697 | Exclusion reason: Other intervention than reported in the SSCs |
| Wang 2023 | Protein Phosphorylation Mechanism of Mesenchymal Stem Cells in the Treatment of Sepsis: A Systematic Review and Meta-analysis | Wang, H.; Luo, J.; Zhong, Y.; Ma, L. | 2023 | Curr Mol Med | 23 | 10 | 1087-1094 | 10.2174/1566524023666221020124204 | Exclusion reason: Wrong study type (no systematic-/mini-/rapid review) |

| | | | | | | | | | |
|---|---|---|---|---|---|---|---|---|---|
| Wang 2023 | Blood gas analysis as a surrogate for microhemodynamic monitoring in sepsis | Wang, J.; Weng, L.; Xu, J.; Du, B. | 2023 | World journal of emergency medicine | 14 | 6 | 421-427 | 10.5847/wjem.j.1920-8642.2023.093 | Exclusion reason: Wrong study type (no systematic-/mini-/rapid review) |
| Wang 2022 | The Use of Hydrocortisone, Ascorbic Acid and Thiamine in Patients with Sepsis and Septic Shock - A Systematic Review | Wang, K.; Yin, L.; Song, Y.; Zhang, M.; Lu, Y.; Wang, S. | 2022 | Journal of pharmacy practice | | | | 10.1177/08971900221097193 | Exclusion reason: Wrong study type (no systematic-/mini-/rapid review) |
| Wang 2021 | Analysis of structure indicators influencing 3-h and 6-h compliance with the surviving sepsis campaign guidelines in China: a systematic review | Wang, L.; Ma, X.; He, H.; Su, L.; Guo, Y.; Shan, G.; Zhou, X.; Liu, D.; Long, Y. | 2021 | Eur J Med Res | 26 | 1 | 27 | 10.1186/s40001-021-00498-7 | Exclusion reason: Wrong study type (no systematic-/mini-/rapid review) |
| Bezzio 2015 | Transperineal ultrasound for perianal sepsis. A systematic review | Bezzio, C.; Furfaro, F.; Dell'Era, A.; Massari, A.; Cassinotti, A.; De Franchis, R.; Maconi, G. | 2015 | Digestive and Liver Disease | | | e171 | | Exclusion reason: Wrong publication type (e.g. conference abstract, preprints,...) |
| Wang 2021 | Blood Glucose Levels and Mortality in Patients With Sepsis: Dose-Response Analysis of Observational Studies | Wang, W.; Chen, W.; Liu, Y.; Li, L.; Li, S.; Tan, J.; Sun, X. | 2021 | Journal of intensive care medicine | 36 | 2 | | 10.1177/0885066619889322 | Exclusion reason: Wrong type of systematic review (e.g. diagnostic (test accuracy), experiential (qualitative), prognostic, etiology and/or risk, prevalence and/or incidence reviews) |

| | | | | | | | | | |
|---|---|---|---|---|---|---|---|---|---|
| Wang 2025 | Impact of antiplatelet therapy on outcomes of sepsis: A systematic review and meta-analysis | Wang, X.; Zhou, H. | 2025 | PloS one | 20 | 4 | e0322293 | 10.1371/journal.pone.0322293 | Exclusion reason: Other intervention than reported in the SSCs |
| Wang 2023 | Unraveling the impact of nitric oxide, almitrine, and their combination in COVID-19 (at the edge of sepsis) patients: a systematic review | Wang, Y.; Yu, Q.; Tian, Y.; Ren, S.; Liu, L.; Wei, C.; Liu, R.; Wang, J.; Li, D.; Zhu, K. | 2023 | Frontiers in pharmacology | 14 | | 1172447 | 10.3389/fphar.2023.1172447 | Exclusion reason: Wrong population (e.g. animals and humans pooled together, sepsis only as subgroup,...) |
| Ward 2024 | The Effect of Early Fluid Resuscitation on Mortality in Sepsis: A Systematic Review | Ward, M.; Kuttab, H.; Badgett, R. | 2024 | Ann. Emerg. Med. | 84 | 4 | S11-S12 | 10.1016/j.annemergmed.2024.08.034 | Exclusion reason: Wrong publication type (e.g. conference abstract, preprints,...) |
| Wei 2024 | The role of esmolol in sepsis: a meta-analysis based on randomized controlled trials | Wei, Y.; Bo, F.; Wang, J.; Fu, J.; Qiu, Y.; Bi, H.; He, D.; Liu, X. | 2024 | BMC anesthesiology | 24 | 1 | 326 | 10.1186/s12871-024-02714-3 | Exclusion reason: Other intervention than reported in the SSCs |
| Wei 2024 | Circulating micronutrient levels and their association with sepsis susceptibility and severity: a Mendelian randomization study | Wei, Z.; Liu, Y.; Mei, X.; Zhong, J.; Huang, F. | 2024 | Frontiers in genetics | 15 | | 1353118 | 10.3389/fgene.2024.1353118 | Exclusion reason: Wrong study type (no systematic-/mini-/rapid review) |
| Wen 2021 | Effect of traditional Chinese medicine on serum inflammation and efficacy in patients with sepsis: a systematic review and meta-analysis | Wen, Y.; Feng, C.; Chen, W.; Chen, C.; Kuang, S.; Liu, F.; Tang, Q.; Chen, M. | 2021 | Annals of palliative medicine | 10 | 12 | 12456-12466 | 10.21037/apm-21-3179 | Exclusion reason: Other intervention than reported in the SSCs |
| Wirz 2018 | Effect of procalcitonin-guided antibiotic treatment on clinical outcomes in intensive | Wirz, Y.; Meier, M. A.; Bouadma, L.; Luyt, C. E.; | 2018 | Critical care (London, England) | 22 | 1 | 191 | 10.1186/s13054-018-2125-7 | Exclusion reason: Wrong type of systematic review (e.g. diagnostic (test |

| | | | | | | | | | |
|---|---|---|---|---|---|---|---|---|---|
| | care unit patients with infection and sepsis patients: a patient-level meta-analysis of randomized trials | Wolff, M.; Chastre, J.; Tubach, F.; Schroeder, S.; Nobre, V.; Annane, D.; Reinhart, K.; Damas, P.; Nijsten, M.; Shajiei, A.; deLange, D. W.; Deliberato, R. O.; Oliveira, C. F.; Shehabi, Y.; van Oers, J. A. H.; Beishuizen, A.; Girbes, A. R. J.; de Jong, E.; Mueller, B.; Schuetz, P. | | | | | | | accuracy), experiential (qualitative), prognostic, etiology and/or risk, prevalence and/or incidence reviews) |
| Wood 2021 | The Use of Near-Infrared Spectroscopy and/or Transcranial Doppler as Non-Invasive Markers of Cerebral Perfusion in Adult Sepsis Patients With Delirium: A Systematic Review | Wood, M. D.; Boyd, J. G.; Wood, N.; Frank, J.; Girard, T. D.; Ross-White, A.; Chopra, A.; Foster, D.; Griesdale, D. E. G. | 2021 | Journal of intensive care medicine | | | | 10.1177/0885066621997090 | Exclusion reason: Wrong type of systematic review (e.g. diagnostic (test accuracy), experiential (qualitative), prognostic, etiology and/or risk, prevalence and/or incidence reviews) |
| Worley 2018 | Management of early pouch-related septic complications in | Worley, G. H. T.; Segal, J. P.; Warusavitarn | 2018 | Colorectal Dis | 20 | 8 | O181-o189 | 10.1111/codi.14266 | Exclusion reason: Wrong study type (no systematic-/mini-/rapid review) |

| | | | | | | | | | |
|---|---|---|---|---|---|---|---|---|---|
| | ulcerative colitis: a systematic review | e, J.; Clark, S. K.; Faiz, O. D. | | | | | | | |
| Wu 2020 | [Clinical efficacy and safety of Xuebijing injection on sepsis: a Meta-analysis] | Wu, Y.; Zhang, J.; Qi, L. | 2020 | Zhonghua wei zhong bing ji jiu yi xue | 32 | 6 | 691-695 | 10.3760/cma.j.cn121430-20200427-00475 | Exclusion reason: Wrong language |
| Wu 2021 | Intravenous corticosteroid treatment in adult patients with sepsis defined by the Sepsis-3 criteria: a systematic review and meta-analysis | Wu, Y. P.; Lin, C. K.; Hamaya, R.; Huang, F. Y.; Chien, Y. S.; Hsu, Y. T.; Chen, S. T.; Papatheodorou, S. | 2021 | medRxiv | | | | 10.1101/2021.10.17.21265100 | Exclusion reason: Wrong publication type (e.g. conference abstract, preprints,...) |
| Xian 2023 | Efficacy and safety of acupuncture as a complementary therapy for sepsis: a systematic review and meta-analysis | Xian, J.; Wang, L.; Zhang, C.; Wang, J.; Zhu, Y.; Yu, H.; Zhang, X.; Tan, Q. | 2023 | Acupuncture in medicine : journal of the British Medical Acupuncture Society | 41 | 1 | | 10.1177/09645284221086288 | Exclusion reason: Other intervention than reported in the SSCs |
| Xiao 2018 | Curative efficacy and safety of traditional Chinese medicine xuebijing injections combined with ulinastatin for treating sepsis in the Chinese population: A meta-analysis | Xiao, S. H.; Luo, L.; Liu, X. H.; Zhou, Y. M.; Liu, H. M.; Huang, Z. F. | 2018 | Medicine | 97 | 23 | e10971 | 10.1097/MD.0000000000010971 | Exclusion reason: Other intervention than reported in the SSCs |
| Xie 2019 | The Clinical Significance of Thrombocytopenia Complicating Sepsis: A meta-analysis | Xie, Y.; Tian, R.; Xie, H.; Jin, W.; Du, J.; Huang, P.; Zhou, Z.; Wang, R. | 2019 | The Journal of infection | 78 | 4 | 323-337 | 10.1016/j.jinf.2018.12.002 | Exclusion reason: Wrong type of systematic review (e.g. diagnostic (test accuracy), experiential (qualitative), |

| | | | | | | | | | |
|---|---|---|---|---|---|---|---|---|---|
| | | | | | | | | | prognostic, etiology and/or risk, prevalence and/or incidence reviews) |
| Bilg 2020 | Comparison of procalcitonin and C-reactive protein (CRP) in neonatal bacterial sepsis | Bilg, A.; Choi, M.; Tewari, S.; Chan, B.; Anis, S.; Luu, J.; Afghani, B. | 2020 | J. Invest. Med. | 68 | 1 | A18-A19 | 10.1136/jim-2019-WMRC.45 | Exclusion reason: Wrong publication type (e.g. conference abstract, preprints,...) |
| Xingji 2018 | Reevaluating the Role of Corticosteroids in Septic Shock: An Updated Meta-Analysis of Randomized Controlled Trials | Xingji, Lian; Daozheng, Huang; Yinshu, Cao; Yanxing, Wei; Zhiwen, Lian; Shouhong, Wang; Tiehe, Qin; Pengcheng, He; Yuan-hui, Liu | 2018 | SSRN | | | | 10.2139/ssrn.3244808 | Exclusion reason: Wrong publication type (e.g. conference abstract, preprints,...) |
| Xu 2015 | The effect of early goal-directed therapy on outcome in adult severe sepsis and septic shock patients: A meta-analysis of randomised clinical trials | Xu, J.; Yang, Y.; Qiu, H. | 2015 | Intensive Care Medicine Experimental | 3 | no pagination | | 10.1186/2197-425X-3-S1-A47 | Exclusion reason: Wrong publication type (e.g. conference abstract, preprints,...) |
| Yamakawa 2015 | Recombinant human soluble thrombomodulin in severe sepsis: a systematic review and meta-analysis | Yamakawa, K.; Aihara, M.; Ogura, H.; Yuhara, H.; Hamasaki, T.; Shimazu, T. | 2015 | Journal of thrombosis and haemostasis : JTH | 13 | (Yamakawa K.,; | 508-19 | 10.1111/jth.12841 | Exclusion reason: Other intervention than reported in the SSCs |

| | | | | | | | | | |
|---|---|---|---|---|---|---|---|---|---|
| | | | | | | Ogura H.; Shimazu T.) Department of Traumatology and Acute Critical Medicine Osaka University Graduate School of Medicine Osaka Japan | | | |
| Yamakawa 2019 | Recombinant Human Soluble Thrombomodulin in Sepsis-Induced Coagulopathy: An Updated Systematic Review and Meta-Analysis | Yamakawa, K.; Murao, S.; Aihara, M. | 2019 | Thrombosis and haemostasis | 119 | 1 | 56-65 | 10.1055/s-0038-1676345 | Exclusion reason: Other intervention than reported in the SSCs |
| Yan 2023 | Clinical Efficacy of Blood Purification in the Treatment of Sepsis: a Meta-Analysis of the last 5 Years | Yan, J.; Zhang, Y.; Zhang, J. | 2023 | Clinical laboratory | 69 | 6 | | 10.7754/Clin.Lab.2022.220931 | Exclusion reason: Wrong study type (no systematic-/mini-/rapid review) |

| | | | | | | | | | |
|---|---|---|---|---|---|---|---|---|---|
| Yang 2023 | Impact of angiotensin-converting enzyme inhibitors or angiotensin receptor blockers on the mortality in sepsis: A meta-analysis | Yang, D. C.; Xu, J.; Jian, L.; Yu, Y. | 2023 | World J Clin Cases | 11 | 36 | 8498-8506 | 10.12998/wjcc.v11.i36.8498 | Exclusion reason: Other intervention than reported in the SSCs |
| Yang 2021 | Role of Extracorporeal Membrane Oxygenation in Adults and Children With Refractory Septic Shock: A Systematic Review and Meta-Analysis | Yang, Y.; Xiao, Z.; Huang, J.; Gong, L.; Lu, X. | 2021 | Frontiers in pediatrics | 9 | | 791781 | 10.3389/fped.2021.791781 | Exclusion reason: Wrong type of systematic review (e.g. diagnostic (test accuracy), experiential (qualitative), prognostic, etiology and/or risk, prevalence and/or incidence reviews) |
| Yao 2020 | Is haemoglobin below 7.0 g/dL an optimal trigger for allogenic red blood cell transfusion in patients admitted to intensive care units? A meta-analysis and systematic review | Yao, R. Q.; Ren, C.; Zhang, Z. C.; Zhu, Y. B.; Xia, Z. F.; Yao, Y. M. | 2020 | BMJ open | 10 | 2 | e030854 | 10.1136/bmjopen-2019-030854 | Exclusion reason: Wrong population (e.g. animals and humans pooled together, sepsis only as subgroup,...) |
| Yao 2019 | Are Corticosteroids Beneficial for Sepsis and Septic Shock? Based on Pooling Analysis of 16 Studies | Yao, Y. Y.; Lin, L. L.; Gu, H. Y.; Wu, J. Y.; Niu, Y. M.; Zhang, C. | 2019 | Frontiers in pharmacology | 10 | 101548923 | 714 | 10.3389/fphar.2019.00714 | Exclusion reason: Wrong study type (no systematic-/mini-/rapid review) |
| Yeo 2020 | Extracorporeal membrane oxygenation in pediatric septic shock: A systematic review of literature and metanalysis | Yeo, N.; Maclaren, G.; Kollengode, R.; Alexander, P.; Barbarro, R.; Raman, L. | 2020 | Perfusion | 35 | 1 SUPPL | 268-None | 10.1177/0267659120909723 | Exclusion reason: Wrong publication type (e.g. conference abstract, preprints,...) |

| | | | | | | | | | |
|---|---|---|---|---|---|---|---|---|---|
| Yidan 2020 | Systematic evaluation of acupuncture efficacy and safety of sepsis | Yidan, Tang; Min, X. U.; Rui, Wang; Mingzhe, X. U.; Jing, Yang; Jin, L. I. U. | 2020 | 中国循证医学杂志 | 20 | 2 | 174-180 | 10.7507/1672-2531.201910006 | Exclusion reason: Wrong language |
| Yin 2018 | Efficacy of norepinephrine, dopamine or vasopressor in the management of septic shock and severe sepsis: A meta-analysis | Yin, L. B.; Hou, L.; Liu, R. Y.; Wang, J. L.; Hu, Y. Q.; Hu, S. Y.; Zhu, Z. J.; Zhu, J. L.; Zhang, W. J.; Guo, G. L. | 2018 | International Journal of Clinical and Experimental Medicine | 11 | 11 | 11383-11395 | | Exclusion reason: Wrong study type (no systematic-/mini-/rapid review) |
| Yoshimoto 2022 | Optimal target blood pressure in critically ill adult patients with vasodilatory shock: A systematic review and meta-analysis | Yoshimoto, H.; Fukui, S.; Higashio, K.; Endo, A.; Takasu, A.; Yamakawa, K. | 2022 | Frontiers in physiology | 13 | | 962670 | 10.3389/fphys.2022.962670 | Exclusion reason: Wrong population (e.g. animals and humans pooled together, sepsis only as subgroup,...) |
| Yu 2019 | [Evaluation and treatment of sepsis-induced myocardial dysfunction: a systematic study] | Yu, L.; Sun, R. | 2019 | Zhonghua Wei Zhong Bing Ji Jiu Yi Xue | 31 | 3 | 378-380 | 10.3760/cma.j.issn.2095-4352.2019.03.025 | Exclusion reason: Wrong language |
| Yu 2021 | Pharmacokinetic Drug-drug Interaction of Antibiotics Used in Sepsis Care in China | Yu, X.; Chu, Z.; Li, J.; He, R.; Wang, Y.; Cheng, C. | 2021 | Curr Drug Metab | 22 | 1 | 5-23 | 10.2174/1389200221666200929115117 | Exclusion reason: Wrong study type (no systematic-/mini-/rapid review) |
| Yu 2016 | [Effects of β-blockers in patients with septic shock: a meta analysis] | Yu, Y. W.; Sun, T. W.; Wan, Y. D.; Liu, Z. Q.; Kan, Q. C. | 2016 | Zhonghua yi xue za zhi | 96 | 7 | 570-4 | 10.3760/cma.j.issn.0376-2491.2016.07.017 | Exclusion reason: Wrong language |

| | | | | | | | | | |
|---|---|---|---|---|---|---|---|---|---|
| Yuan 2020 | Systematic review of ultrasound-guided fluid resuscitation vs. early goal-directed therapy in patients with septic shock | Yuan, J.; Yang, X.; Yuan, Q.; Li, M.; Chen, Y.; Dong, C. | 2020 | Zhonghua wei zhong bing ji jiu yi xue | 32 | 1 | 56-61 | 10.3760/cma.j.cn121430-20191114-00010 | Exclusion reason: Wrong language |
| Yuan 2024 | [Analysis of clinical research features and outcome indexes of traditional Chinese medicine intervention in septic kidney injury] | Yuan, Y.; Wang, J. J.; Li, J. S.; Lei, S. Y. | 2024 | Zhongguo Zhong yao za zhi = Zhongguo zhongyao zazhi = China journal of Chinese materia medica | 49 | 5 | 1388-1396 | 10.19540/j.cnki.cjcmm.20231119.501 | Exclusion reason: Wrong language |
| Yue 2020 | Childhood Mortality After Fluid Bolus with Septic or Severe Infection Shock: A Systematic Review And Meta-Analysis | Yue, J.; Zheng, R.; Wei, H.; Li, J.; Wu, J.; Wang, P.; Zhao, H. | 2020 | Shock (Augusta, Ga.) | | | | 10.1097/SHK.0000000000001657 | Exclusion reason: Wrong population (e.g. animals and humans pooled together, sepsis only as subgroup,...) |
| yushan 2024 | Is Restriction of Intravenous Fluid Beneficial for Septic Shock in Icu Patients? A Meta-Analysis of Randomized Controlled Trial | yushan, duan; Yue, Yin; Jinxi, Yue; Weihua, Zhu; Xiumei, Yang; Yiming, Ma; Yuan, Yang; Xiaohong, Wan | 2024 | SSRN | | | | 10.2139/ssrn.4895304 | Exclusion reason: Wrong publication type (e.g. conference abstract, preprints,...) |
| Zamora 2020 | Vasopressor discontinuation order in the recovery phase of septic shock: A systematic review and meta-analysis | Zamora, M. K.; Barbon, C. E.; Angeles, R. R.; Guevarra, D.; Albay, A. | 2020 | Philipp. J. Intern. Med. | 58 | 4 | 141-145 | | Exclusion reason: Wrong language |
| Zarychanski 2015 | The efficacy and safety of heparin in patients with sepsis: a | Zarychanski, R.; Abou-Setta, A. M.; Kanji, S.; | 2015 | Crit Care Med | 43 | 3 | 511-8 | 10.1097/ccm.0000000000000763 | Exclusion reason: Wrong population (e.g. animals and humans |

| | | | | | | | | | |
|---|---|---|---|---|---|---|---|---|---|
| | systematic review and metaanalysis | Turgeon, A. F.; Kumar, A.; Houston, D. S.; Rimmer, E.; Houston, B. L.; McIntyre, L.; Fox-Robichaud, A. E.; Hébert, P.; Cook, D. J.; Fergusson, D. A. | | | | | | | pooled together, sepsis only as subgroup,...) |
| Zhang 2017 | Variable efficacy of drotrecogin alfa (activated) in severe sepsis and septic shock: A meta-analysis | Zhang, D. L.; Zhu, H. Y.; Zhang, S.; Wu, Q. Q.; Zhang, W. J. | 2017 | International Journal of Clinical and Experimental Medicine | 10 | 9 | 12976-12985 | | Exclusion reason: Other intervention than reported in the SSCs |
| Zhang 2016 | Probiotics for Preventing Late-Onset Sepsis in Preterm Neonates: A PRISMA-Compliant Systematic Review and Meta-Analysis of Randomized Controlled Trials | Zhang, G. Q.; Hu, H. J.; Liu, C. Y.; Shakya, S.; Li, Z. Y. | 2016 | Medicine | 95 | 8 | e2581 | 10.1097/MD.0000000000002581 | Exclusion reason: Wrong type of systematic review (e.g. diagnostic (test accuracy), experiential (qualitative), prognostic, etiology and/or risk, prevalence and/or incidence reviews) |
| Zhang 2022 | Benefits of esmolol in adults with sepsis and septic shock: An updated meta-analysis of randomized controlled trials | Zhang, J.; Chen, C.; Liu, Y.; Yang, Y.; Yang, X.; Yang, J. | 2022 | Medicine | 101 | 27 | e29820 | 10.1097/MD.0000000000029820 | Exclusion reason: Other intervention than reported in the SSCs |
| Zhang 2020 | Efficacy and safety of recombinant human thrombopoietin on sepsis patients with | Zhang, J.; Lu, Z.; Xiao, W.; Hua, T.; | 2020 | Frontiers in pharmacology | 11 | | 940 | 10.3389/fphar.2020.00940 | Exclusion reason: Other intervention than reported in the SSCs |

| | | | | | | | | | |
|---|---|---|---|---|---|---|---|---|---|
| | thrombocytopenia: A systematic review and meta-analysis | Zheng, Y.; Yang, M. | | | | | | | |
| Zhang 2024 | A meta-analysis of the efficacy and safety of aspirin in the treatment of elderly patients with sepsis | Zhang, J.; Wu, J. | 2024 | Pak. J. Pharma. Sci. | 37 | 6 | 1375-1389 | 10.36721/PJPS.2024.37.6.REG.1375-1389.1 | Exclusion reason: Other intervention than reported in the SSCs |
| Zhang 2023 | Progress of research on organ protection of acupuncture and moxibustion in the prevention and treatment of sepsis | Zhang, J. Y.; Chen, B.; Hu, X. Y.; Li, N. C.; Chen, Y.; Yang, K. J.; Liu, Y. Y.; Chen, Z. L.; Guo, Y. | 2023 | Zhen ci yan jiu = Acupuncture research | 48 | 11 | 1159-1167 | 10.13702/j.1000-0607.20221026 | Exclusion reason: Wrong language |
| Zhang 2020 | Xuebijing combined with ulinastation for sepsis: A few noteworthy additions | Zhang, K. | 2020 | The American journal of emergency medicine | | | | 10.1016/j.ajem.2020.02.008 | Exclusion reason: Wrong study type (no systematic-/mini-/rapid review) |
| Zhang 2015 | Adjuvant treatment with crude rhubarb for patients with systemic inflammation reaction syndrome/sepsis: A meta-analysis of randomized controlled trials | Zhang, L.; Chen, J.; Jiang, D.; Zhang, P. | 2015 | Journal of Critical Care | 30 | 2 | 282-289 | 10.1016/j.jcrc.2014.11.008 | Exclusion reason: Other intervention than reported in the SSCs |
| Zhang 2021 | Effect of Tongfu traditional Chinese medicine preparation on patients with septic gastrointestinal dysfunction: a systematic review and meta-analysis | Zhang, L.; Wu, Q.; Chen, P.; Xu, Y.; Zheng, Y.; Ma, S.; Zheng, J. | 2021 | Annals of palliative medicine | 10 | 12 | 12072-12085 | 10.21037/apm-21-2461 | Exclusion reason: Other intervention than reported in the SSCs |
| Zhang 2016 | Additional traditional Chinese medicine on gastrointestinal | Zhang, Y.; Zhao, Z.; | 2016 | Pakistan journal of | 29 | 2 Suppl | 663-9 | | Exclusion reason: Other intervention than reported in the SSCs |

| | | | | | | | | | |
|---|---|---|---|---|---|---|---|---|---|
| | dysfunction in patients with sepsis: A systematic review and meta-analysis | Tang, E.; Hu, Y.; Mao, J. | | pharmaceutical sciences | | | | | |
| Zhang 2021 | Signaling pathways and intervention therapies in sepsis | Zhang, Y. Y.; Ning, B. T. | 2021 | Signal transduction and targeted therapy | 6 | 1 | 407 | 10.1038/s41392-021-00816-9 | Exclusion reason: Wrong study type (no systematic-/mini-/rapid review) |
| Zhang 2015 | The efficacy of activated protein C for the treatment of sepsis: incorporating observational evidence with a Bayesian approach | Zhang, Z. | 2015 | BMJ open | 5 | 1 | e006524 | 10.1136/bmjopen-2014-006524 | Exclusion reason: Other intervention than reported in the SSCs |
| Zhang 2022 | Effectiveness of automated alerting system compared to usual care for the management of sepsis | Zhang, Z.; Chen, L.; Xu, P.; Wang, Q.; Zhang, J.; Chen, K.; Clements, C. M.; Celi, L. A.; Herasevich, V.; Hong, Y. | 2022 | NPJ digital medicine | 5 | 1 | 101 | 10.1038/s41746-022-00650-5 | Exclusion reason: Other intervention than reported in the SSCs |
| Zhao 2025 | Effect of Thiamine Supplementation on Mortality in Septic Patients: A Meta-Analysis of Randomized Controlled Trials | Zhao, C.; Du, Q.; Sun, C.; Du, S.; Zhang, X.; Zu, G. | 2025 | ResearchSquare | | | | 10.21203/rs.3.rs-6639223/v1 | Exclusion reason: Wrong publication type (e.g. conference abstract, preprints,...) |
| Zhou 2023 | Should we initiate vasopressors earlier in patients with septic shock: A mini systemic review | Zhou, H. X.; Yang, C. F.; Wang, H. Y.; Teng, Y.; He, H. Y. | 2023 | World journal of critical care medicine | 12 | 4 | 204-216 | 10.5492/wjcm.v12.i4.204 | Exclusion reason: Wrong study type (no systematic-/mini-/rapid review) |

| | | | | | | | | | |
|---|---|---|---|---|---|---|---|---|---|
| Zhou 2021 | Extended or continuous infusion of carbapenems in children with severe infections: A systematic review and narrative synthesis | Zhou, P.; Zhang, Y.; Wang, Z.; Ying, Y.; Xing, Y.; Tong, X.; Zhai, S. | 2021 | Antibiotics (Basel, Switzerland) | 10 | 9 | | 10.3390/antibiotics10091088 | Exclusion reason: Wrong population (e.g. animals and humans pooled together, sepsis only as subgroup,...) |
| Zhou 2019 | Rules of anti-infection therapy for sepsis and septic shock | Zhou, X.; Su, L. X.; Zhang, J. H.; Liu, D. W.; Long, Y. | 2019 | Chin Med J (Engl) | 132 | 5 | 589-596 | 10.1097/cm9.0000000000000101 | Exclusion reason: Wrong study type (no systematic-/mini-/rapid review) |
| Bond 2020 | Sepsis alerts in emergency departments: A systematic review of accuracy and quality measure impact | Bond, W. F.; Powell, E. S.; Hwang, M. | 2020 | Acad. Emerg. Med. | 27 | | S209-None | 10.1111/acem.13961 | Exclusion reason: Wrong publication type (e.g. conference abstract, preprints,...) |
| Zhu 2018 | [Effects of different administration methods of hydrocortisone on blood glucose in patients with septic shock: a Meta-analysis] | Zhu, L.; Li, X.; Liu, Y.; Yang, K.; Jing, G.; Chen, Y.; Dou, Z.; Chen, Q.; Liu, J. | 2018 | Zhonghua wei zhong bing ji jiu yi xue | 30 | 10 | 915-919 | 10.3760/cma.j.issn.2095-4352.2018.010.001 | Exclusion reason: Wrong language |
| Zochios 2015 | Raised cardiac troponin in intensive care patients with sepsis, in the absence of angiographically documented coronary artery disease: A systematic review | Zochios, V.; Valchanov, K. | 2015 | Journal of the Intensive Care Society | 16 | 1 | 52-57 | 10.1177/1751143714555303 | Exclusion reason: Wrong study type (no systematic-/mini-/rapid review) |
| Zou 2023 | Efficacy and safety of glucocorticoid in the treatment of sepsis: A systematic review and meta-analysis | Zou, H.; Yi, F.; Wang, Q. | 2023 | Asian journal of surgery | | | | 10.1016/j.asjsur.2023.02.004 | Exclusion reason: Wrong study type (no systematic-/mini-/rapid review) |

| | | | | | | | | | |
|---|---|---|---|---|---|---|---|---|---|
| Borouchaki 2022 | Immunotherapy to treat sepsis induced-immunosuppression: Immune eligibility or outcome criteria, a systematic review | Borouchaki, A.; de Roquetaillade, C.; Barthélémy, R.; Mebazaa, A.; Chousterman, B. G. | 2022 | Journal of critical care | 72 | | 154137 | 10.1016/j.jcrc.2022.154137 | Exclusion reason: Other intervention than reported in the SSCs |
| BroksoHolst 2016 | Benefits and harms of red blood cell transfusions in patients with septic shock in the intensive care unit | Brokso Holst, L. | 2016 | Danish Medical Journal | 63 | 2 | | | Exclusion reason: Wrong population (e.g. animals and humans pooled together, sepsis only as subgroup,...) |
| Holst 2016 | Benefits and harms of red blood cell transfusions in patients with septic shock in the intensive care unit | Holst, L. B. | 2016 | Danish medical journal | 63 | 2 | | | Exclusion reason: Wrong study type (no systematic-/mini-/rapid review) |
| Brown 2022 | A Systematic Review and Meta-Analysis on Possible Role of Vitamin C in Sepsis | Brown, J.; Robertson, C.; Sevilla, L.; Garza, J.; Rashid, H.; Benitez, A. C.; Shipotko, M.; Ali, Z. | 2022 | Cureus | 14 | 12 | e32886 | 10.7759/cureus.32886 | Exclusion reason: Wrong study type (no systematic-/mini-/rapid review) |
| Buckman 2018 | Empiric Antibiotics for Sepsis | Buckman, S. A.; Turnbull, I. R.; Mazuski, J. E. | 2018 | Surgical infections | 19 | 2 | 147-154 | 10.1089/sur.2017.282 | Exclusion reason: Wrong study type (no systematic-/mini-/rapid review) |
| Cai 2015 | The effects of early goal-directed therapy on mortality rate in patients with severe sepsis and septic shock: A systematic | Cai, G.; Tong, H.; Hao, X.; Hu, C.; Yan, M.; Chen, J.; Yan, J. | 2015 | Zhonghua wei zhong bing ji jiu yi xue | 27 | 6 | 439-42 | 10.3760/cma.j.issn.2095-4352.2015.06.005 | Exclusion reason: Wrong language |

| | | | | | | | | | |
|---|---|---|---|---|---|---|---|---|---|
| | literature review and Meta-analysis | | | | | | | | |
| Calabrò 2025 | Ivabradine use in critical care: a systematic review and metanalysis of cardiogenic and septic shock patients | Calabrò, L. A.; Pasetto, M.; Scolletta, S.; Annoni, F.; Demailly, Z.; Halenarova, K.; Donadello, K.; Taccone, F. S. | 2025 | BMC Anesthesiol | 25 | 1 | 276 | 10.1186/s12871-025-03121-y | Exclusion reason: Wrong population (e.g. animals and humans pooled together, sepsis only as subgroup,...) |
| Carsetti 2021 | Good clinical practice for the use of vasopressor and inotropic drugs in critically ill patients: state-of-the-art and expert consensus | Carsetti, A.; Bignami, E.; Cortegiani, A.; Donadello, K.; Donati, A.; Foti, G.; Grasselli, G.; Romagnoli, S.; Antonelli, M.; E, D. E. Blasio; Forfori, F.; Guarracino, F.; Scolletta, S.; Tritapepe, L.; Scudeller, L.; Cecconi, M.; Girardis, M. | 2021 | Minerva Anestesiol | 87 | 6 | 714-732 | 10.23736/s0375-9393.20.14866-1 | Exclusion reason: Wrong study type (no systematic-/mini-/rapid review) |
| Catalanotto 2023 | Hyperoxia in critically ill patients with sepsis and septic shock: a systematic review | Catalanotto, F. R.; Ippolito, M.; Mirasola, A.; Catalisano, G.; Milazzo, | 2023 | Journal of Anesthesia, Analgesia and Critical Care (Online) | 3 | 1 | 12 | 10.1186/s44158-023-00096-5 | Exclusion reason: Wrong study type (no systematic-/mini-/rapid review) |

| | | | | | | | | | |
|---|---|---|---|---|---|---|---|---|---|
| | | M.; Giarratano, A.; Cortegiani, A. | | | | | | | |
| Chacko 2015 | Systematic review of use of beta-blockers in sepsis | Chacko, C. J.; Gopal, S. | 2015 | Journal of Anaesthesiology Clinical Pharmacology | 31 | 4 | 460-465 | 10.4103/0970-9185.169063 | Exclusion reason: Wrong study type (no systematic-/mini-/rapid review) |
| Chander 2024 | Effect of low vs. high vancomycin trough level on the clinical outcomes of adult patients with sepsis or gram-positive bacterial infections: a systematic review and meta-analysis | Chander, S.; Kumari, R.; Wang, H. Y.; Mohammed, Y. N.; Parkash, O.; Lohana, S.; Sorath, F.; Lohana, A. C.; Sadarat, F.; Shiwlani, S. | 2024 | BMC Infect Dis | 24 | 1 | 1114 | 10.1186/s12879-024-09927-4 | Exclusion reason: Wrong population (e.g. animals and humans pooled together, sepsis only as subgroup,...) |
| | Correction: Effect of low vs. high Vancomycin trough level on the clinical outcomes of adult patients with sepsis or gram-positive bacterial infections: a systematic review and meta-analysis | Chander, S.; Kumari, R.; Yu Wang, H.; Nadeem Mohammed, Y.; Parkash, O.; Lohana, S.; Sorath, F.; Chand Lohana, A.; Sadarat, F.; Shiwlani, S. | 2025 | BMC Infect Dis | 25 | 1 | 371 | 10.1186/s12879-025-10745-5 | |
| Chang 2021 | [Therapeutic effect of hydrocortisone combined with vitamin C and vitamin B1 on patients with sepsis: a Meta-analysis] | Chang, C.; Luo, K.; Zhu, H.; Deng, G.; Gao, Q. | 2021 | Zhonghua wei zhong bing ji jiu yi xue | 33 | 9 | 1040-1046 | 10.3760/cma.j.cn121430-20210728-01101 | Exclusion reason: Wrong language |

| | | | | | | | | | |
|---|---|---|---|---|---|---|---|---|---|
| Chao 2018 | Impact of Corticosteroids on Short- and Long-Term Mortality in Patients with Sepsis and Septic Shock: A Meta-Analysis with Trial Sequential Analysis | Chao, Zhang | 2018 | SSRN | | | | 10.2139/ssrn.3223121 | Exclusion reason: Wrong publication type (e.g. conference abstract, preprints,...) |
| Chaudhuri 2024 | Adverse Effects Related to Corticosteroid Use in Sepsis, Acute Respiratory Distress Syndrome, and Community-Acquired Pneumonia: A Systematic Review and Meta-Analysis | Chaudhuri, D.; Israelian, L.; Putowski, Z.; Prakash, J.; Pitre, T.; Nei, A. M.; Spencer-Segal, J. L.; Gershengorn, H. B.; Annane, D.; Pastores, S. M.; Rochwerg, B. | 2024 | Critical care explorations | 6 | 4 | e1071 | 10.1097/CCE.0000000000001071 | Exclusion reason: Wrong population (e.g. animals and humans pooled together, sepsis only as subgroup,...) |
| Chemtob 2016 | Systematic review of the association of venous oxygenation and outcome in adult hospitalized patients | Chemtob, R. A.; Eskesen, T. G.; Moeller-Soerensen, H.; Perner, A.; Ravn, H. B. | 2016 | Acta anaesthesiologica Scandinavica | 60 | 10 | 1367-1378 | 10.1111/aas.12810 | Exclusion reason: Wrong population (e.g. animals and humans pooled together, sepsis only as subgroup,...) |
| Adegbite 2023 | Clinical features, treatment outcomes and mortality risk of tuberculosis sepsis in HIV-negative patients: a systematic review and meta-analysis of case reports | Adegbite, B. R.; Elegbede-Adegbite, N. O. M.; Edoa, J. R.; Honkpehedji, Y. J.; Zinsou, J. F.; Dejon- | 2023 | Infection | 51 | 3 | 609-621 | 10.1007/s15010-022-01950-4 | Exclusion reason: Wrong population (e.g. animals and humans pooled together, sepsis only as subgroup,...) |

| | | | | | | | | | |
|---|---|---|---|---|---|---|---|---|---|
| | | Agobé, J. C.; Adegnika, A. A.; Grobusch, M. P. | | | | | | | |
| Chen 2018 | Efficacy and safety of Xuebijing injection combined with ulinastatin as adjunctive therapy on sepsis: A systematic review and meta-analysis | Chen, G.; Gao, Y.; Jiang, Y.; Yang, F.; Li, S.; Tan, D.; Ma, Q. | 2018 | Frontiers in Pharmacology | 9 | JUL | | 10.3389/fphar.2018.00743 | Exclusion reason: Other intervention than reported in the SSCs |
| Chen 2018 | The effects of statin therapy on mortality in patients with sepsis: A meta-analysis of randomized trials | Chen, M.; Ji, M.; Si, X. | 2018 | Medicine | 97 | 31 | e11578 | 10.1097/MD.0000000000011578 | Exclusion reason: Other intervention than reported in the SSCs |
| Chen 2023 | Comparison of Balanced Crystalloids versus Normal Saline in Critically Ill Patients: A Systematic Review with Meta-Analysis and Trial Sequential Analysis of Randomized Controlled Trials | Chen, Y.; Gao, Y. | 2023 | Therapeutics and clinical risk management | 19 | | 783-799 | 10.2147/TCRM.S416785 | Exclusion reason: Wrong population (e.g. animals and humans pooled together, sepsis only as subgroup,...) |
| Cheng 2019 | A meta-analysis of Qingre Jiedu and Liangxue Sanyu method in the treatment of sepsis | Cheng, L.; Jiang, H.; Chen, M.; Chen, Q.; Wang, X.; Lu, J. | 2019 | Zhonghua wei zhong bing ji jiu yi xue | 31 | 1 | 73-80 | 10.3760/cma.j.issn.2095-4352.2019.01.015 | Exclusion reason: Wrong language |
| ChineseSocietyofCriticalCare 2015 | [Chinese guidelines for management of severe sepsis and septic shock 2014] | Chinese Society of Critical Care, Medicine | 2015 | Zhonghua wei zhong bing ji jiu yi xue | 27 | 6 | 401-26 | 10.3760/cma.j.issn.2095-4352.2015.06.001 | Exclusion reason: Wrong language |

| | | | | | | | | | |
|---|---|---|---|---|---|---|---|---|---|
| Choudhary 2022 | Sepsis Management, Controversies, and Advancement in Nanotechnology: A Systematic Review | Choudhary, R. | 2022 | Cureus | 14 | 2 | e22112 | 10.7759/cureus.22112 | Exclusion reason: Wrong type of systematic review (e.g. diagnostic (test accuracy), experiential (qualitative), prognostic, etiology and/or risk, prevalence and/or incidence reviews) |
| Choy 2022 | Impact of sepsis education for healthcare professionals and students on learner and patient outcomes: A systematic review | Choy, C. L.; Liaw, S. Y.; Goh, E. L.; See, K. C.; Chua, W. L. | 2022 | The Journal of hospital infection | 122 | | 84-95 | 10.1016/j.jhin.2022.01.004 | Exclusion reason: Wrong population (e.g. animals and humans pooled together, sepsis only as subgroup,...) |
| Chu 2024 | Impact of premorbid use of beta-blockers on survival outcomes of patients with sepsis: A systematic review and meta-analysis | Chu, H.; Fei, F.; Su, Y.; Zhou, H. | 2024 | Experimental and therapeutic medicine | 28 | 1 | 300 | 10.3892/etm.2024.12589 | Exclusion reason: Other intervention than reported in the SSCs |
| Cioni 2021 | Procalcitonin in clinical practice: From diagnosis of sepsis to antibiotic therapy | Cioni, G.; Canini, J.; Pieralli, F. | 2021 | Ital. J. Med. | 15 | 1 | 7-16 | 10.4081/ITJM.2021.1438 | Exclusion reason: Wrong study type (no systematic-/mini-/rapid review) |
| Ahc 2019 | Corticosteroid Administration in Sepsis May Be Associated With Lower 28-Day Mortality | Ahc, Media | 2019 | Infectious Disease Alert | 38 | 8 | 0-0 | | Exclusion reason: Wrong study type (no systematic-/mini-/rapid review) |
| Cortegiani 2022 | Empiric treatment of patients with sepsis and septic shock and place in therapy of cefiderocol: a systematic review and | Cortegiani, A.; Ingoglia, G.; Ippolito, M.; Girardis, M.; Falcone, M.; Pea, F.; Pugliese, F.; | 2022 | Journal of Anesthesia, Analgesia and Critical Care (Online) | 2 | 1 | 34 | 10.1186/s44158-022-00062-7 | Exclusion reason: Wrong study type (no systematic-/mini-/rapid review) |

| | | | | | | | | | |
|---|---|---|---|---|---|---|---|---|---|
| | expert opinion statement | Stefani, S.; Viale, P.; Giarratano, A. | | | | | | | |
| Datta 2024 | The Clinical Utility of Targeted Heart Rate Control in Septic Shock: A Systematic Review and Meta-analysis of Randomized Controlled Trials with Trial Sequential Analysis | Datta, P. K.; Sathe, P.; Roy, A.; Baronia, T.; Bhattacharje e, A.; Kundu, R. | 2024 | Indian journal of critical care medicine : peer-reviewed, official publication of Indian Society of Critical Care Medicine | 28 | 12 | 1170-1179 | 10.5005/jp-journals-10071-24849 | Exclusion reason: Other intervention than reported in the SSCs |
| DeSilva 2025 | Health-Related Quality of Life of Adult Sepsis Survivors Following Critical Illness: A Systematic Review | De Silva, S.; Urwin, A.; Grimwood, C.; Goh, E. K. E.; Higgins, A. M.; Hodgson, C. L. | 2025 | Critical care explorations | 7 | 4 | e1238 | 10.1097/CCE.0000000000001238 | Exclusion reason: Wrong type of systematic review (e.g. diagnostic (test accuracy), experiential (qualitative), prognostic, etiology and/or risk, prevalence and/or incidence reviews) |
| Deng 2024 | Assessing the Efficacy and Safety of Corticosteroids for the Management of Pediatric Patients With Sepsis and Septic Shock: A Systematic Review and Meta-Analysis of Randomized Controlled Trials | Deng, J.; Azab, M.; Bedri, N.; Venugopal, K.; Zuo, Q. K.; Hung, J.; Heybati, K.; Zhou, F. | 2024 | Am. J. Respir. Crit. Care Med. | 209 | | | | Exclusion reason: Wrong publication type (e.g. conference abstract, preprints,...) |
| Deng 2024 | Assessing the Efficacy and Safety of Hydrocortisone, Ascorbic Acid, and | Deng, J.; Zubair, A.; Zuo, Q. K.; Blais, S.; | 2024 | Am. J. Respir. Crit. Care Med. | 209 | | | | Exclusion reason: Wrong publication type (e.g. conference abstract, preprints,...) |

| | | | | | | | | | |
|---|---|---|---|---|---|---|---|---|---|
| | Thiamine Combination Therapy for the Management of Sepsis and Septic Shock: A Systematic Review and Meta-Analysis of Randomized Controlled Trials | Porter, V.; Venugopal, K.; Hung, J.; Heybati, K. | | | | | | | |
| Deshmukh 2021 | Sepsis calculator for neonatal early onset sepsis - A Systematic Reviews and meta-analysis | Deshmukh, M.; Mehta, S.; Patole, S. | 2021 | The journal of maternal-fetal & neonatal medicine : the official journal of the European Association of Perinatal Medicine, the Federation of Asia and Oceania Perinatal Societies, the International Society of Perinatal Obstetricians | 34 | 11 | 1-11 | 10.1080/14767058.2019.1649650 | Exclusion reason: Wrong population (e.g. animals and humans pooled together, sepsis only as subgroup,...) |
| Deshpande 2015 | Statin Therapy and Mortality from Sepsis A Meta-Analysis of Randomized Trials | Deshpande, A.; Pasupuleti, V.; Rothberg, M. B. | 2015 | The American journal of medicine | 128 | 4 | 410-7.e1 | 10.1016/j.amjmed.2014.10.057 | Exclusion reason: Other intervention than reported in the SSCs |
| Despins 2017 | Automated Detection of Sepsis Using Electronic Medical Record Data: A Systematic Review | Despins, L. A. | 2017 | Journal for healthcare quality : official publication of the National Association for Healthcare Quality | 39 | 6 | 322-333 | 10.1097/JHQ.0000000000000066 | Exclusion reason: Wrong type of systematic review (e.g. diagnostic (test accuracy), experiential (qualitative), prognostic, etiology and/or risk, prevalence |

| | | | | | | | | | |
|---|---|---|---|---|---|---|---|---|---|
| | | | | | | | | | and/or incidence reviews) |
| Diaztagle Fernández 2024 | Assessment of the shock index in septic shock: A systematic review | Diaztagle Fernández, J. J.; Castañeda-González, J. P.; Trujillo Zambrano, J. I.; Duarte Martínez, F. E.; Saavedra Ortiz, MÁ | 2024 | Medicina intensiva | | | | 10.1016/j.medine.2024.07.006 | Exclusion reason: Wrong type of systematic review (e.g. diagnostic (test accuracy), experiential (qualitative), prognostic, etiology and/or risk, prevalence and/or incidence reviews) |
| Diaztagle Fernández 2017 | Venous-to-arterial carbon dioxide difference in the resuscitation of patients with severe sepsis and septic shock: A systematic review | Diaztagle Fernández, J. J.; Rodríguez Murcia, J. C.; Sprockel Díaz, J. J. | 2017 | Medicina intensiva | 41 | 7 | 401-410 | 10.1016/j.medin.2017.03.008 | Exclusion reason: Wrong language |

| **Covidence #** | **Study ID** | **Title** | **Authors** |
|---|---|---|---|
| #1010 | Ding 2021 | Association Between Prior Calcium Channel Blocker Use and Mortality in Septic Patients: A Meta-Analysis of Cohort Studies | Ding, X.; Cui, Y.; Liang, H.; Wang, D.; [illegible] Kan, Q.; Wang, L.; Sun, T. |
| #1017 | Diz 2025 | Effect of Treatment With Balanced Crystalloids Versus Normal Saline on the Mortality of Critically Ill Patients With and Without Traumatic Brain Injury: A Systematic Review and Meta-Analysis | Diz, J. C.; Luna-Rojas, P.; Díaz-Vidal, [illegible] Fernández-Vázquez, U.; Gil-Casado, [illegible] Ferreira, E. |

### Supplementary Table S2: References to systematic reviews (n = 251) investigating other interventions than corticosteroids

| Study ID | Title | Authors | Published Year | Journal | Volume | Issue | Pages | DOI |
|---|---|---|---|---|---|---|---|---|
| Abdelazeem 2022 | Meta-analysis comparing dexmedetomidine versus standard of care for sedation in patients with sepsis | Abdelazeem, B.; Malik, B.; Mohamed, M. M. G.; Savarapu, P.; Isa, S.; Kheiri, B.; Hassan, M. | 2022 | Proceedings (Baylor University. Medical Center) | 35 | 2 | 184-189 | 10.1080/08998280.2021.1997063 |
| Abdelbaky 2023 | Restrictive Versus Liberal Fluid Regimen in Refractory Sepsis and Septic Shock: A Systematic Review and Meta-Analysis | Abdelbaky, A. M.; Elmasry, W. G.; Awad, A. H. | 2023 | Cureus | 15 | 10 | e47783 | 10.7759/cureus.47783 |
| Abdul-Aziz 2024 | Prolonged vs Intermittent Infusions of β-Lactam Antibiotics in Adults with Sepsis or Septic Shock: A Systematic Review and Meta-Analysis | Abdul-Aziz, M. H.; Hammond, N. E.; Brett, S. J.; Cotta, M. O.; De Waele, J. J.; Devaux, A.; Di Tanna, G. L.; Dulhunty, J. M.; Elkady, H.; Eriksson, L.; Hasan, M. S.; Khan, A. B.; Lipman, J.; Liu, X.; Monti, G.; Myburgh, J.; Novy, E.; Omar, S.; Rajbhandari, D.; Roger, C.; Sjövall, F.; Zaghi, I.; Zangrillo, A.; Delaney, A.; Roberts, J. A. | 2024 | JAMA | 332 | 8 | 638-648 | 10.1001/jama.2024.9803 |
| Acharya 2024 | Efficacy of Early Goal-Directed Therapy in Septic Shock Management: A Systematic Review | Acharya, P.; Virani, S.; Afreen, S.; Perthiani, A.; Sangster, E.; Lanka, N.; Malasevskaia, I. | 2024 | Cureus | 16 | 11 | e74358 | 10.7759/cureus.74358 |
| Achten 2019 | Association of Use of the Neonatal Early-Onset Sepsis Calculator With Reduction in Antibiotic Therapy and Safety: A | Achten, N. B.; Klingenberg, C.; Benitz, W. E.; Stocker, M.; Schlapbach, L. J.; Giannoni, E.; Bokelaar, R.; Driessen, G. J. A.; Brodin, P.; | 2019 | JAMA pediatrics | 173 | 11 | 1032-1040 | 10.1001/jamapediatrics.2019.2825 |

| | | | | | | | | |
|---|---|---|---|---|---|---|---|---|
| | Systematic Review and Meta-analysis | Uthaya, S.; van Rossum, A. M. C.; Plötz, F. B. | | | | | | |
| Ahn 2024 | Comparison of Early and Late Norepinephrine Administration in Patients With Septic Shock: A Systematic Review and Meta-Analysis | Ahn, C.; Yu, G.; Shin, T. G.; Cho, Y.; Park, S.; Suh, G. Y. | 2024 | Chest | 166 | 6 | 1417-1430 | 10.1016/j.chest.2024.05.042 |
| Aljarbou 2024 | Short course of intravenous antibiotics in the treatment of uncomplicated proven neonatal bacterial sepsis: A systematic review | Aljarbou, A.; Cuello, C.; Leslie, Atfs | 2024 | Acta Paediatr | 113 | 1 | 56-66 | 10.1111/apa.16972 |
| Ammar 2021 | Vitamin C for Sepsis and Septic Shock | Ammar, M. A.; Ammar, A. A.; Condeni, M. S.; Bell, C. M. | 2021 | American journal of therapeutics | 28 | 6 | e649-e679 | 10.1097/MJT.0000000000001423 |
| Andriolo 2017 | Effectiveness and safety of procalcitonin evaluation for reducing mortality in adults with sepsis, severe sepsis or septic shock | Andriolo, B. N.; Andriolo, R. B.; Salomão, R.; Atallah Á, N. | 2017 | Cochrane Database Syst Rev | 1 | 1 | Cd010959 | 10.1002/14651858.CD010959.pub2 |
| Angus 2015 | A systematic review and meta-analysis of early goal-directed therapy for septic shock: the ARISE, ProCESS and ProMISe Investigators | Angus, D. C.; Barnato, A. E.; Bell, D.; Bellomo, R.; Chong, C. R.; Coats, T. J.; Davies, A.; Delaney, A.; Harrison, D. A.; Holdgate, A.; Howe, B.; Huang, D. T.; Iwashyna, T.; Kellum, J. A.; Peake, S. L.; Pike, F.; Reade, M. C.; Rowan, K. M.; Singer, M.; Webb, S. A.; Weissfeld, L. A.; Yealy, D. M.; Young, J. D. | 2015 | Intensive care medicine | 41 | 9 | 1549-60 | 10.1007/s00134-015-3822-1 |
| | Early goal-directed therapy versus usual care in the management of septic shock | Gottlieb, M. | 2017 | Cjem | 19 | 1 | 65-67 | 10.1017/cem.2016.2 |

| | | | | | | | | |
|---|---|---|---|---|---|---|---|---|
| Arango-Granados 2021 | Impact of red blood cell transfusion on oxygen transport and metabolism in patients with sepsis and septic shock: a systematic review and meta-analysis | Arango-Granados, M. C.; Umaña, M.; Sánchez Á, I.; García, A. F.; Granados, M.; Ospina-Tascón, G. A. | 2021 | Revista Brasileira de terapia intensiva | 33 | 1 | 154-166 | 10.5935/0103-507x.20210017 |
| Arulkumaran 2020 | Effect of Antibiotic Discontinuation Strategies on Mortality and Infectious Complications in Critically Ill Septic Patients: A Meta-Analysis and Trial Sequential Analysis | Arulkumaran, N.; Khpal, M.; Tam, K.; Baheerathan, A.; Corredor, C.; Singer, M. | 2020 | Critical care medicine | 48 | 5 | 757-764 | 10.1097/CCM.0000000000004267 |
| Atkins 2023 | Pharmacist Involvement in Sepsis Response and Time to Antibiotics: A Systematic Review | Atkins, P. E.; Bastin, M. L. T.; Morgan, R. J.; Laine, M. E.; Flannery, A. H. | 2023 | Journal of the American College of Clinical Pharmacy : JACCP | 6 | 8 | 942-953 | 10.1002/jac5.1723 |
| Avni 2015 | Vasopressors for the treatment of septic shock: Systematic review and meta-analysis | Avni, T.; Lador, A.; Lev, S.; Leibovici, L.; Paul, M.; Grossman, A. | 2015 | PloS one | 10 | 8 | e0129305 | 10.1371/journal.pone.0129305 |
| Azadian 2022 | Mortality Benefit From the Passive Leg Raise Maneuver in Guiding Resuscitation of Septic Shock Patients: A Systematic Review and Meta-Analysis of Randomized Trials | Azadian, M.; Win, S.; Abdipour, A.; Kim, C. K.; Nguyen, H. B. | 2022 | Journal of intensive care medicine | 37 | 5 | | 10.1177/08850666211019713 |
| Bai 2024 | Human albumin for adults with sepsis: An updated systematic review and meta-analysis of randomized controlled trials | Bai, Z.; Lai, Y.; Han, K.; Shi, L.; Guan, X.; Xu, Y. | 2024 | Medicine (Baltimore) | 103 | 52 | e40983 | 10.1097/md.0000000000040983 |

| | | | | | | | | |
|---|---|---|---|---|---|---|---|---|
| Bannard-Smith 2024 | The efficacy, safety and effectiveness of hyperoncotic albumin solutions in patients with sepsis: A systematic review and meta-analysis | Bannard-Smith, J.; Elrakhawy, M.; Norman, G.; Owen, R.; Felton, T.; Dark, P. | 2024 | J Intensive Care Soc | 25 | 3 | 308-318 | 10.1177/17511437241259437 |
| Bauer 2025 | ADJUNCTIVE VASOPRESSORS AND SHORT-TERM MORTALITY IN ADULTS WITH SEPTIC SHOCK: A SYSTEMATIC REVIEW AND META-ANALYSIS | Bauer, S. R.; Wieruszewski, P. M.; Bissell Turpin, B. D.; Dugar, S.; Sacha, G. L.; Sato, R.; Siuba, M. T.; Schleicher, M.; Vachharajani, V.; Falck-Ytter, Y.; Morgan, R. L. | 2025 | Shock (Augusta, Ga.) | 63 | 5 | 668-676 | 10.1097/SHK.0000000000002558 |
| Beran 2022 | Balanced Crystalloids versus Normal Saline in Adults with Sepsis: A Comprehensive Systematic Review and Meta-Analysis | Beran, A.; Altorok, N.; Srour, O.; Malhas, S. E.; Khokher, W.; Mhanna, M.; Ayesh, H.; Aladamat, N.; Abuhelwa, Z.; Srour, K.; Mahmood, A.; Taleb, M.; Assaly, R. | 2022 | Journal of clinical medicine | 11 | 7 | | 10.3390/jcm11071971 |
| Bharwani 2025 | Conservative versus liberal fluid resuscitation for septic patients at risk for fluid overload: A systematic review with meta-analysis | Bharwani, A.; Dionne, J. C.; Pérez, M. L.; Englesakis, M.; Meyhoff, T. S.; Sivapalan, P.; Zampieri, F. G.; Wilcox, M. E. | 2025 | J Crit Care | 87 | | 155045 | 10.1016/j.jcrc.2025.155045 |
| Bhatt 2022 | Literature Review of the Efficacy of High-Volume Hemofiltration in Critically Ill Pediatric Patients | Bhatt, G. C.; Sethi, S. K.; Mehta, I.; Nair, N.; Chakraborty, R.; Sharma, B.; Singh, S.; Kumar, N.; Gulati, K.; Raina, R. | 2022 | Blood purification | 51 | 8 | 1-11 | 10.1159/000520519 |
| Bhattacharjee 2017 | Levosimendan does not provide mortality benefit over dobutamine in adult patients with septic shock: A meta-analysis of randomized controlled trials | Bhattacharjee, S.; Soni, K. D.; Maitra, S.; Baidya, D. K. | 2017 | Journal of Clinical Anesthesia | 39 | | 67-72 | 10.1016/j.jclinane.2017.03.011 |

| | | | | | | | | |
|---|---|---|---|---|---|---|---|---|
| Borthwick 2017 | High-volume haemofiltration for sepsis in adults | Borthwick, E. M.; Hill, C. J.; Rabindranath, K. S.; Maxwell, A. P.; McAuley, D. F.; Blackwood, B. | 2017 | Cochrane Database of Systematic Reviews | 1 | 1 | CD008075 | 10.1002/14651858.CD008075.pub3 |
| Busani 2016 | Intravenous immunoglobulin in septic shock: review of the mechanisms of action and meta-analysis of the clinical effectiveness | Busani, S.; Damiani, E.; Cavazzuti, I.; Donati, A.; Girardis, M. | 2016 | Minerva anestesiologica | 82 | 5 | 559-72 | |
| Cai 2022 | Clinical efficacy and safety of vitamin C in the treatment of septic shock patients: systematic review and meta-analysis | Cai, B.; Lv, X.; Lin, M.; Feng, C.; Chen, C. | 2022 | Annals of palliative medicine | 11 | 4 | 1369-1380 | 10.21037/apm-22-225 |
| Cai 2025 | Effect of restrictive fluid resuscitation on severe acute kidney injury in septic shock: a systematic review and meta-analysis | Cai, X. E.; Ling, W. T.; Cai, X. T.; Yan, M. K.; Zhang, Y. J.; Xu, J. Y. | 2025 | BMJ open | 15 | 2 | e086367 | 10.1136/bmjopen-2024-086367 |
| Casserly 2015 | Meta-analysis based on limited data shows no evidence to support the guideline recommendation for early administration of antibiotics in severe sepsis and septic shock | Casserly, Brian; Hannigan, Ailish | 2015 | Evidence Based Medicine | 20 | 6 | 214-215 | 10.1136/ebmed-2015-110269 |
| Chandelia 2025 | Balanced Salt Solution Versus Normal Saline as Resuscitation Fluid in Pediatric Septic Shock: A Systematic Review and Meta-Analysis of Randomized Controlled Trials | Chandelia, S.; Angurana, S. K.; Prasad, S.; Tiwari, M. K.; Jayashree, M.; Nallasamy, K.; Bansal, A. | 2025 | Journal of intensive care medicine | | | | 10.1177/08850666251315705 |

| | | | | | | | | |
|---|---|---|---|---|---|---|---|---|
| Chang 2017 | Effects of Polymyxin B Hemoperfusion on Mortality in Patients With Severe Sepsis and Septic Shock: A Systemic Review, Meta-Analysis Update, and Disease Severity Subgroup Meta-Analysis | Chang, T.; Tu, Y. K.; Lee, C. T.; Chao, A.; Huang, C. H.; Wang, M. J.; Yeh, Y. C. | 2017 | Crit Care Med | 45 | 8 | e858-e864 | 10.1097/ccm.0000000000002362 |
| Chang 2018 | Effect of levosimendan on mortality in severe sepsis and septic shock: a meta-analysis of randomised trials | Chang, W.; Xie, J. F.; Xu, J. Y.; Yang, Y. | 2018 | BMJ open | 8 | 3 | e019338 | 10.1136/bmjopen-2017-019338 |
| Chelkeba 2015 | Early goal-directed therapy reduces mortality in adult patients with severe sepsis and septic shock: Systematic review and meta-analysis | Chelkeba, L.; Ahmadi, A.; Abdollahi, M.; Najafi, A.; Mojtahedzadeh, M. | 2015 | Indian journal of critical care medicine : peer-reviewed, official publication of Indian Society of Critical Care Medicine | 19 | 7 | 401-11 | 10.4103/0972-5229.160281 |
| Chen 2022 | The impact of vitamin C-containing treatment on the mortality of patients with sepsis: A systematic review and meta-analysis of randomized controlled trials | Chen, C. Y.; Chiu, C. T.; Lee, H. S.; Lai, C. C. | 2022 | Journal of infection and public health | 15 | 12 | 1514-1520 | 10.1016/j.jiph.2022.11.015 |
| Chen 2018 | Correlation analysis of omega-3 fatty acids and mortality of sepsis and sepsis-induced ARDS in adults: data from previous randomized controlled trials | Chen, H.; Wang, S.; Zhao, Y.; Luo, Y.; Tong, H.; Su, L. | 2018 | Nutrition journal | 17 | 1 | 57 | 10.1186/s12937-018-0356-8 |
| Chen 2020 | Clinical outcomes of continuous vs intermittent meropenem infusion for | Chen, P.; Chen, F.; Lei, J.; Zhou, B. | 2020 | Advances in clinical and experimental | 29 | 8 | | 10.17219/acem/121934 |

| | | | | | | | | |
|---|---|---|---|---|---|---|---|---|
| | the treatment of sepsis: A systematic review and meta-analysis | | | medicine : official organ Wroclaw Medical University | | | | |
| Chen 2020 | Effect of Dexmedetomidine on duration of mechanical ventilation in septic patients: a systematic review and meta-analysis | Chen, P.; Jiang, J.; Zhang, Y.; Li, G.; Qiu, Z.; Levy, M. M.; Hu, B. | 2020 | BMC pulmonary medicine | 20 | 1 | 42 | 10.1186/s12890-020-1065-6 |
| Chen 2017 | Early outcome of early-goal directed therapy for patients with sepsis or septic shock: A systematic review and meta-analysis of randomized controlled trials | Chen, X.; Zhu, W.; Tan, J.; Nie, H.; Liu, L.; Yan, D.; Zhou, X.; Sun, X. | 2017 | Oncotarget | 8 | 16 | 27510-27519 | 10.18632/oncotarget.15550 |
| Chidambaram 2019 | Vasopressin vs noradrenaline: Have we found the perfect recipe to improve outcome in septic shock? | Chidambaram, S.; Goh, E. L.; Rey, V. G.; Khan, M. A. | 2019 | Journal of critical care | 49 | | 99-104 | 10.1016/j.jcrc.2018.10.029 |
| Claxton 2017 | Safety and efficacy of procalcitonin guidance in patients with suspected or confirmed sepsis: A systematic review and meta-analysis | Claxton, A.; Thompson-Leduc, P.; Kirson, N.; Rice, B.; Hey, J.; Iankova, I.; Krause, A.; Schonfeld, S.; DeBrase, C.; Bozzette, S.; Schuetz, P. | 2017 | American Journal of Respiratory and Critical Care Medicine | | | | 10.1164/ajrccm-conference.2017.A51 |
| | Efficacy and Safety of Procalcitonin Guidance in Patients With Suspected or Confirmed Sepsis: A Systematic Review and Meta-Analysis: Author Correction | Claxton, A. J.; Thompson-Leduc, P.; Kirson, N. Y.; Rice, B.; Hey, J.; Iankova, I.; Krause, A.; Schonfeld, S. A.; DeBrase, C. R.; Bozzette, S.; Schuetz, P. | 2018 | Crit Care Med | 46 | 9 | 1560 | 10.1097/ccm.0000000000003308 |

| | | | | | | | | |
|---|---|---|---|---|---|---|---|---|
| Coccolini 2016 | Early goal-directed treatment versus standard care in management of early septic shock: Meta-analysis of randomized trials | Coccolini, F.; Sartelli, M.; Catena, F.; Ceresoli, M.; Montori, G.; Ansaloni, L. | 2016 | The journal of trauma and acute care surgery | 81 | 5 | 971-978 | 10.1097/TA.0000000000001246 |
| Cui 2019 | The clinical efficacy of intravenous IgM-enriched immunoglobulin (pentaglobin) in sepsis or septic shock: a meta-analysis with trial sequential analysis | Cui, J.; Wei, X.; Lv, H.; Li, Y.; Li, P.; Chen, Z.; Liu, G. | 2019 | Annals of intensive care | 9 | 1 | 27 | 10.1186/s13613-019-0501-3 |
| Cusack 2022 | Effects of Fluids on the Sublingual Microcirculation in Sepsis | Cusack, R.; O'Neill, S.; Martin-Loeches, I. | 2022 | Journal of clinical medicine | 11 | 24 | | 10.3390/jcm11247277 |
| D'Aragon 2015 | Blood pressure targets for vasopressor therapy: a systematic review | D'Aragon, F.; Belley-Cote, E. P.; Meade, M. O.; Lauzier, F.; Adhikari, N. K.; Briel, M.; Lalu, M.; Kanji, S.; Asfar, P.; Turgeon, A. F.; Fox-Robichaud, A.; Marshall, J. C.; Lamontagne, F. | 2015 | Shock | 43 | 6 | 530-9 | 10.1097/shk.0000000000000348 |
| Damiani 2015 | Effect of performance improvement programs on compliance with sepsis bundles and mortality: a systematic review and meta-analysis of observational studies | Damiani, E.; Donati, A.; Serafini, G.; Rinaldi, L.; Adrario, E.; Pelaia, P.; Busani, S.; Girardis, M. | 2015 | PloS one | 10 | 5 | e0125827 | 10.1371/journal.pone.0125827 |
| Dari 2024 | Comparison of High-Normal Versus Low-Normal Mean Arterial Pressure at Target on Outcomes in Sepsis or Shock Patients: A Meta-Analysis of Randomized Control Trials | Dari, M. A.; Fayaz, A.; Sharif, S.; Hernandez Galaviz, S.; Hernandez Galaviz, E.; Bataineh, S. M.; Wei, C. R.; Allahwala, D. | 2024 | Cureus | 16 | 1 | e52258 | 10.7759/cureus.52258 |

| | | | | | | | | |
|---|---|---|---|---|---|---|---|---|
| Devi 2024 | Efficacy and safety of short- vs. standard-course antibiotics for culture-negative neonatal sepsis: a systematic review and meta-analysis | Devi, R.; Priyadarshi, M.; Singh, P.; Chaurasia, S.; Basu, S. | 2024 | Journal of tropical pediatrics | 70 | 2 | | 10.1093/tropej/fmae002 |
| Ding 2022 | Comparison of Dexmedetomidine Versus Propofol in Mechanically Ventilated Patients With Sepsis: A Meta-Analysis of Randomized Controlled Trials | Ding, H. Z.; Dong, Y. L.; Zhang, K. Y.; Bai, J. Y. | 2022 | Frontiers in pharmacology | 13 | | 901898 | 10.3389/fphar.2022.901898 |
| Ding 2018 | Early goal-directed and lactate-guided therapy in adult patients with severe sepsis and septic shock: a meta-analysis of randomized controlled trials | Ding, Xian-Fei; Yang, Zi-Yue; Xu, Zhen-Tao; Li, Li-Feng; Yuan, Bo; Guo, Li-Na; Wang, Le-Xin; Zhu, Xi; Sun, Tong-Wen | 2018 | Journal of Translational Medicine | 16 | 1 | 331 | 10.1186/s12967-018-1700-7 |
| Dinleyici 2023 | Clinical efficacy of IgM-enriched immunoglobulin as adjunctive therapy in neonatal and pediatric sepsis: a systematic review and meta-analysis | Dinleyici, E. C.; Frey, G.; Kola, E.; Wippermann, U.; Bauhofer, A.; Staus, A.; Griffiths, P.; Azharry, M.; Rohsiswatmo, R. | 2023 | Frontiers in pediatrics | 11 | | 1239014 | 10.3389/fped.2023.1239014 |
| Do-Wan 2024 | The Effect of Polymyxin B-Immobilized Fiber Column Hemoperfusion for Sepsis: A Systemic Review and Meta-Analysis | Do-Wan, K. I. M.; Jeong-Min, K. I. M.; Yong-Sang, Seong; Reverien, Habimana; Hwa-Jin, C. H. O.; In-Seok, Jeong | 2024 | Journal of Acute Care Surgery | | 2 | 45-51 | |
| Drewry 2017 | Antipyretic Therapy in Critically Ill Septic Patients: A Systematic Review and Meta-Analysis | Drewry, A. M.; Ablordeppey, E. A.; Murray, E. T.; Stoll, C. R.; Izadi, S. R.; Dalton, C. M.; Hardi, A. C.; Fowler, S. A.; Fuller, B. M.; Colditz, G. A. | 2017 | Critical care medicine | 45 | 5 | 806-813 | 10.1097/CCM.0000000000002285 |

| | | | | | | | | |
|---|---|---|---|---|---|---|---|---|
| Duclos 2019 | Effects of the discontinuation sequence of norepinephrine and vasopressin on hypotension incidence in patients with septic shock: A meta-analysis | Duclos, G.; Baumstarck, K.; Dünser, M.; Zieleskiewicz, L.; Leone, M. | 2019 | Heart & lung : the journal of critical care | 48 | 6 | 560-565 | 10.1016/j.hrtlng.2019.05.007 |
| Dupuis 2017 | Impact of transfusion on patients with sepsis admitted in intensive care unit: a systematic review and meta-analysis | Dupuis, C.; Sonneville, R.; Adrie, C.; Gros, A.; Darmon, M.; Bouadma, L.; Timsit, J. F. | 2017 | Annals of intensive care | 7 | 1 | 5 | 10.1186/s13613-016-0226-5 |
| Ehrman 2019 | Resuscitation Guided by Volume Responsiveness Does Not Reduce Mortality in Sepsis: A Meta-Analysis | Ehrman, R. R.; Gallien, J. Z.; Smith, R. K.; Akers, K. G.; Malik, A. N.; Harrison, N. E.; Welch, R. D.; Levy, P. D.; Sherwin, R. L. | 2019 | Critical care explorations | 1 | 5 | e0015 | 10.1097/CCE.0000000000000015 |
| EinAlshaeba 2025 | The Effect of Early Warning Systems for Sepsis on Mortality: A Systematic Review and Meta-analysis | Ein Alshaeba, S.; Marhoffer, E. A.; Holleck, J. L.; Theisen-Toupal, J.; Grimshaw, A. A.; Gunderson, C. G. | 2025 | Journal of general internal medicine | | | | 10.1007/s11606-025-09569-5 |
| Fan 2016 | Efficacy and safety of low-molecular-weight heparin in patients with sepsis: a meta-analysis of randomized controlled trials | Fan, Y.; Jiang, M.; Gong, D.; Zou, C. | 2016 | Scientific reports | 6 | | 25984 | 10.1038/srep25984 |
| Feng 2019 | Levosimendan does not reduce the mortality of critically ill adult patients with sepsis and septic shock: a meta-analysis | Feng, F.; Chen, Y.; Li, M.; Yuan, J. J.; Chang, X. N.; Dong, C. M. | 2019 | Chinese medical journal | 132 | 10 | 1212-1217 | 10.1097/CM9.0000000000000197 |
| Feng 2021 | Effect of vitamin C in critically ill patients with sepsis and septic shock: A meta-analysis | Feng, F.; Yang, H.; Yang, W.; Li, M.; Chang, X.; Chen, Y. | 2021 | Science progress | 104 | 1 | | 10.1177/0036850421998175 |
| Fernández- | Association between fluid overload and mortality in | Fernández-Sarmiento, J.; Sierra-Zuñiga, M. F.; Salazar | 2023 | BMJ paediatrics open | 7 | 1 | | 10.1136/bmjpo-2023-002094 |

| | | | | | | | | |
|---|---|---|---|---|---|---|---|---|
| Sarmiento 2023 | children with sepsis: a systematic review and meta-analysis | González, M. P.; Lucena, N.; Soares Lanziotti, V.; Agudelo, S. | | | | | | |
| Ford 2025 | The Effect of Severe Sepsis and Septic Shock Management Bundle (SEP-1) Compliance and Implementation on Mortality Among Patients With Sepsis A Systematic Review | Ford, J. S.; Morrison, J. C.; Kyaw, M.; Hewlett, M.; Tahir, P.; Jain, S.; Nemati, S.; Malhotra, A.; Wardi, G. | 2025 | Annals of internal medicine | 178 | 4 | 543-557 | 10.7326/ANNALS-24-02426 |
| Fu 2022 | Unfractionated heparin improves the clinical efficacy in adult sepsis patients: a systematic review and meta-analysis | Fu, S.; Yu, S.; Wang, L.; Ma, X.; Li, X. | 2022 | BMC anesthesiology | 22 | 1 | 28 | 10.1186/s12871-021-01545-w |
| Fujii 2018 | Polymyxin B-immobilized hemoperfusion and mortality in critically ill adult patients with sepsis/septic shock: a systematic review with meta-analysis and trial sequential analysis | Fujii, T.; Ganeko, R.; Kataoka, Y.; Furukawa, T. A.; Featherstone, R.; Doi, K.; Vincent, J. L.; Pasero, D.; Robert, R.; Ronco, C.; Bagshaw, S. M. | 2018 | Intensive care medicine | 44 | 2 | 1-12 | 10.1007/s00134-017-5004-9 |
| | Correction to: Polymyxin B-immobilized hemoperfusion and mortality in critically ill adult patients with sepsis/septic shock: a systematic review with meta-analysis and trial sequential analysis | Fujii, T.; Ganeko, R.; Kataoka, Y.; Furukawa, T. A.; Featherstone, R.; Doi, K.; Vincent, J. L.; Pasero, D.; Robert, R.; Ronco, C.; Bagshaw, S. M. | 2018 | Intensive care medicine | 44 | 2 | 1-2 | 10.1007/s00134-018-5055-6 |
| Ge 2024 | The association between levosimendan and mortality in patients with sepsis or septic shock: a systematic review and meta-analysis | Ge, Z.; Gao, Y.; Lu, X.; Yu, S.; Qin, M.; Gong, C.; Walline, J. H.; Zhu, H.; Li, Y. | 2024 | Eur J Emerg Med | 31 | 2 | 90-97 | 10.1097/mej.0000000000001105 |

| | | | | | | | | |
|---|---|---|---|---|---|---|---|---|
| Ge 2021 | Thiamine combined with vitamin C in sepsis or septic shock: a systematic review and meta-analysis | Ge, Z.; Huang, J.; Liu, Y.; Xiang, J.; Gao, Y.; Walline, J. H.; Lu, X.; Yu, S.; Zhao, L.; Li, Y. | 2021 | European journal of emergency medicine : official journal of the European Society for Emergency Medicine | 28 | 3 | 189-195 | 10.1097/MEJ.0000000000000812 |
| Geng 2023 | Different Concentrations of Albumin Versus Crystalloid in Patients with Sepsis and Septic Shock: A Meta-Analysis of Randomized Clinical Trials | Geng, L.; Tian, X.; Gao, Z.; Mao, A.; Feng, L.; He, C. | 2023 | Journal of intensive care medicine | 38 | 8 | | 10.1177/08850666231170778 |
| Goetz 2021 | Extracorporeal Cytokine Adsorption Therapy As a Preventive Measure in Cardiac Surgery and As a Therapeutic Add-On Treatment in Sepsis: An Updated Systematic Review of Comparative Efficacy and Safety | Goetz, G.; Hawlik, K.; Wild, C. | 2021 | Crit Care Med | 49 | 8 | 1347-1357 | 10.1097/ccm.0000000000005023 |
| Grillo-Ardila 2024 | Early Enteral Nutrition (within 48 h) for Patients with Sepsis or Septic Shock: A Systematic Review and Meta-Analysis | Grillo-Ardila, C. F.; Tibavizco-Palacios, D.; Triana, L. C.; Rugeles, S. J.; Vallejo-Ortega, M. T.; Calderón-Franco, C. H.; Ramírez-Mosquera, J. J. | 2024 | Nutrients | 16 | 11 | | 10.3390/nu16111560 |
| Gu 2015 | Single-Dose Etomidate Does Not Increase Mortality in Patients with Sepsis: A Systematic Review and Meta-Analysis of Randomized Controlled Trials and Observational Studies | Gu, W. J.; Wang, F.; Tang, L.; Liu, J. C. | 2015 | Chest | 147 | 2 | 335-46 | 10.1378/chest.14-1012 |

| | | | | | | | | |
|---|---|---|---|---|---|---|---|---|
| Gu 2015 | Early lactate clearance-guided therapy in patients with sepsis: a meta-analysis with trial sequential analysis of randomized controlled trials | Gu, W. J.; Zhang, Z.; Bakker, J. | 2015 | Intensive care medicine | 41 | 10 | 1862-3 | 10.1007/s00134-015-3955-2 |
| Guan 2023 | Meta-Analysis of the Efficacy of Levosimendan in the Treatment of Severe Sepsis Complicated with Septic Cardiomyopathy | Guan, Q.; Zhang, C.; Li, B.; Huang, D.; Li, A.; Qin, J.; Zhang, X. | 2023 | The heart surgery forum | 26 | 5 | E609-E620 | 10.59958/hsf.6439 |
| Guo 2016 | De-escalation of empiric antibiotics in patients with severe sepsis or septic shock: A meta-analysis | Guo, Y.; Gao, W.; Yang, H.; Ma, C.; Sui, S. | 2016 | Heart and Lung: Journal of Acute and Critical Care | 45 | 5 | 454-459 | 10.1016/j.hrtlng.2016.06.001 |
| Gutiérrez-Pizarraya 2022 | Clinical impact of procalcitonin-based algorithms for duration of antibiotic treatment in critically ill adult patients with sepsis: a meta-analysis of randomized clinical trials | Gutiérrez-Pizarraya, A.; León-García, M. D. C.; De Juan-Idígoras, R.; Garnacho-Montero, J. | 2022 | Expert review of anti-infective therapy | 20 | 1 | 103-112 | 10.1080/14787210.2021.1932462 |
| Hajjar 2019 | Vasopressin Versus Norepinephrine for the Management of Septic Shock in Cancer Patients: The VANCS II Randomized Clinical Trial* | Hajjar, L. A.; Zambolim, C.; Belletti, A.; de Almeida, J. P.; Gordon, A. C.; Oliveira, G.; Park, C. H. L.; Fukushima, J. T.; Rizk, S. I.; Szeles, T. F.; Dos Santos Neto, N. C.; Filho, R. K.; Galas, Frbg; Landoni, G. | 2019 | Critical care medicine | 47 | 12 | 1743-1750 | 10.1097/CCM.0000000000004023 |
| He 2020 | Parenteral selenium for the clinical effect of sepsis: A meta-analysis of randomized controlled trials | He, S.; Cao, B.; Yu, H.; Zhen, J.; Wang, F. | 2020 | Pak. J. Pharma. Sci. | 33 | 6 | 2679-2686 | 10.36721/PJPS.2020.33.6.REG.2687-2696.1 |

| | | | | | | | | |
|---|---|---|---|---|---|---|---|---|
| Hernandez 2024 | Enhancing Survival in Septic Shock: A Systematic Review and Meta-Analysis of the Efficacy of Plasma Exchange Therapy | Hernandez, G. N.; Francis, A. J.; Hamid, P. | 2024 | Cureus | 16 | 5 | e60947 | 10.7759/cureus.60947 |
| Higgins 2020 | Health economic evaluations of sepsis interventions in critically ill adult patients: A systematic review | Higgins, A. M.; Brooker, J. E.; Mackie, M.; Cooper, D. J.; Harris, A. H. | 2020 | Journal of intensive care | 8 | 1 | 5 | 10.1186/s40560-019-0412-2 |
| Hirano 2019 | Liberal versus restrictive red blood cell transfusion strategy in sepsis or septic shock: a systematic review and meta-analysis of randomized trials | Hirano, Y.; Miyoshi, Y.; Kondo, Y.; Okamoto, K.; Tanaka, H. | 2019 | Critical care (London, England) | 23 | 1 | 262 | 10.1186/s13054-019-2543-1 |
| Hu 2025 | The Effect of Different Temperature Management Strategies in Adult Sepsis Patients: A Meta-Analysis of Randomized Controlled Trials | Hu, Y.; Jiang, J.; Wei, M.; Dong, T.; Zhang, Y.; Qin, Y. | 2025 | Therapeutic hypothermia and temperature management | | | | 10.1089/ther.2025.0017 |
| Huang 2021 | The effect of early vasopressin use on patients with septic shock: A systematic review and meta-analysis | Huang, H.; Wu, C.; Shen, Q.; Xu, H.; Fang, Y.; Mao, W. | 2021 | The American journal of emergency medicine | 48 | | 203-208 | 10.1016/j.ajem.2021.05.007 |
| Huang 2023 | The association between mortality and door-to-antibiotic time: a systematic review and meta-analysis | Huang, J.; Yang, J. T.; Liu, J. C. | 2023 | Postgraduate medical journal | | | | 10.1093/postmj/qgad024 |
| Huang 2020 | Terlipressin for the treatment of septic shock in adults: a systematic review and meta-analysis | Huang, L.; Zhang, S.; Chang, W.; Xia, F.; Liu, S.; Yang, Y.; Qiu, H. | 2020 | BMC anesthesiology | 20 | 1 | 58 | 10.1186/s12871-020-00965-4 |

| | | | | | | | | |
|---|---|---|---|---|---|---|---|---|
| Huang 2019 | Terlipressin versus norepinephrine for septic shock: A systematic review and meta-analysis | Huang, P.; Guo, Y.; Li, B.; Liu, Q. | 2019 | Frontiers in pharmacology | 10 | | 1492 | 10.3389/fphar.2019.01492 |
| Huang 2021 | Dexmedetomidine Versus Propofol for Patients With Sepsis Requiring Mechanical Ventilation: A Systematic Review and Meta-Analysis | Huang, P.; Zheng, X.; Liu, Z.; Fang, X. | 2021 | Frontiers in pharmacology | 12 | | 717023 | 10.3389/fphar.2021.717023 |
| Huang 2023 | The efficacy of dexmedetomidine for septic shock: A meta-analysis of randomized controlled trials | Huang, X.; He, C. | 2023 | Medicine | 102 | 35 | e34414 | 10.1097/MD.0000000000034414 |
| Hwang 2020 | Sepsis Alerts in Emergency Departments: A Systematic Review of Accuracy and Quality Measure Impact | Hwang, M. I.; Bond, W. F.; Powell, E. S. | 2020 | The western journal of emergency medicine | 21 | 5 | 1201-1210 | 10.5811/westjem.2020.5.46010 |
| Iankova 2018 | Efficacy and Safety of Procalcitonin Guidance in Patients With Suspected or Confirmed Sepsis: A Systematic Review and Meta-Analysis | Iankova, I.; Thompson-Leduc, P.; Kirson, N. Y.; Rice, B.; Hey, J.; Krause, A.; Schonfeld, S. A.; DeBrase, C. R.; Bozzette, S.; Schuetz, P. | 2018 | Critical care medicine | 46 | 5 | 691-698 | 10.1097/CCM.0000000000002928 |
| Irfan 2022 | Zinc Supplementation and the Prevention and Treatment of Sepsis in Young Infants: A Systematic Review and Meta-Analysis | Irfan, O.; Black, R. E.; Lassi, Z. S.; Bhutta, Z. A. | 2022 | Neonatology | 119 | 2 | 1-12 | 10.1159/000521275 |
| Jiang 2019 | The effects and safety of vasopressin receptor agonists in patients with septic shock: a meta-analysis and trial sequential analysis | Jiang, L.; Sheng, Y.; Feng, X.; Wu, J. | 2019 | Critical care (London, England) | 23 | 1 | 91 | 10.1186/s13054-019-2362-4 |

| | | | | | | | | |
|---|---|---|---|---|---|---|---|---|
| Jiang 2016 | Early goal-directed resuscitation for patients with severe sepsis and septic shock: a meta-analysis and trial sequential analysis | Jiang, L. B.; Zhang, M.; Jiang, S. Y.; Ma, Y. F. | 2016 | Scandinavian journal of trauma, resuscitation and emergency medicine | 24 | | 23 | 10.1186/s13049-016-0214-7 |
| Johansen 2021 | Use of hydroxyethyl starch in sepsis research: A systematic review with meta-analysis | Johansen, J. R.; Perner, A.; Brodtkorb, J. H.; Møller, M. H. | 2021 | Acta anaesthesiologica Scandinavica | 65 | 10 | 1355-1364 | 10.1111/aas.13954 |
| Johnston 2017 | Effect of Immediate Administration of Antibiotics in Patients With Sepsis in Tertiary Care: A Systematic Review and Meta-analysis | Johnston, A. N.; Park, J.; Doi, S. A.; Sharman, V.; Clark, J.; Robinson, J.; Crilly, J. | 2017 | Clinical therapeutics | 39 | 1 | 190-190 | 10.1016/j.clinthera.2016.12.003 |
| Joshi 2019 | Digital alerting and outcomes in patients with sepsis: Systematic review and meta-analysis | Joshi, M.; Ashrafian, H.; Arora, S.; Khan, S.; Cooke, G.; Darzi, A. | 2019 | Journal of medical Internet research | 21 | 12 | e15166 | 10.2196/15166 |
| Kalil 2017 | Early Goal-Directed Therapy for Sepsis: A Novel Solution for Discordant Survival Outcomes in Clinical Trials | Kalil, Andre C.; Johnson, Daniel W.; Lisco, Steven J.; Junfeng, Sun; Sun, Junfeng | 2017 | Critical Care Medicine | 45 | 4 | 607-614 | 10.1097/CCM.0000000000002235 |
| Kanchanasurakit 2021 | Effectiveness of thiamine therapy in mortality rate in patients with septic shock: A systematic review and meta-analysis | Kanchanasurakit, S.; Suthumpoung, P.; Santimaleeworagun, W.; Nakaranurack, C.; Huynh, N. S.; Srisawat, C.; Nunta, M.; Chirakan, V.; Saokaew, S. | 2021 | International journal of critical illness and injury science | 11 | 2 | 86-94 | 10.4103/IJCIIS.IJCIIS_159_20 |
| Khan 2024 | Extracorporeal Membrane Oxygenation as Circulatory Support in Adult Patients with Septic Shock: A Systematic Review | Khan, M. F.; Nazir, M.; Khan, M. K.; Rajendram, R. K.; Shamim, F. | 2024 | Journal of critical care medicine (Universitatea de Medicina si | 10 | 2 | 119-129 | 10.2478/jccm-2024-0017 |

| | | | | | | | | |
|---|---|---|---|---|---|---|---|---|
| | | | | Farmacie din Targu-Mures) | | | | |
| Khattar 2024 | Fluid Resuscitation Dilemma in End-stage Renal Disease Patients Presenting with Sepsis: A Systematic Review and Meta-analysis | Khattar, G.; El Gharib, K.; Pokima, N.; Kotys, J.; Kandala, V.; Mina, J.; Haddadin, F.; Abu Baker, S.; Asmar, S.; Rizvi, T.; Flamenbaum, M.; Elsayegh, D.; Chalhoub, M.; El Hage, H.; El Sayegh, S. | 2024 | Journal of intensive care medicine | | | | 10.1177/08850666241261673 |
| Kim 2024 | Sepsis Alert Systems, Mortality, and Adherence in Emergency Departments: A Systematic Review and Meta-Analysis | Kim, H. J.; Ko, R. E.; Lim, S. Y.; Park, S.; Suh, G. Y.; Lee, Y. J. | 2024 | JAMA network open | 7 | 7 | e2422823 | 10.1001/jamanetworkopen.2024.22823 |
| Kondo 2020 | Prolonged versus intermittent β-lactam antibiotics intravenous infusion strategy in sepsis or septic shock patients: a systematic review with meta-analysis and trial sequential analysis of randomized trials | Kondo, Y.; Ota, K.; Imura, H.; Hara, N.; Shime, N. | 2020 | J Intensive Care | 8 | | 77 | 10.1186/s40560-020-00490-z |
| Kong 2021 | The impact of selenium administration on severe sepsis or septic shock: a meta-analysis of randomized controlled trials | Kong, L.; Wu, Q.; Liu, B. | 2021 | Afr Health Sci | 21 | 1 | 277-285 | 10.4314/ahs.v21i1.36 |
| Korang 2021 | The effects of adding quinolones to beta-lactam antibiotics for sepsis | Korang, S. K.; Maagaard, M.; Feinberg, J. B.; Perner, A.; Gluud, C.; Jakobsen, J. C. | 2021 | Acta anaesthesiologica Scandinavica | 65 | 8 | 1023-1032 | 10.1111/aas.13831 |
| Korang 2021 | Antibiotic regimens for late-onset neonatal sepsis | Korang, S. K.; Safi, S.; Nava, C.; Greisen, G.; Gupta, M.; Lausten-Thomsen, U.; Jakobsen, J. C. | 2021 | The Cochrane database of systematic reviews | 5 | 5 | CD013836 | 10.1002/14651858.CD013836.pub2 |

| | | | | | | | | |
|---|---|---|---|---|---|---|---|---|
| Kotani 2024 | Non-adrenergic vasopressors for vasodilatory shock or perioperative vasoplegia: a meta-analysis of randomized controlled trials | Kotani, Y.; Belletti, A.; D'Amico, F.; Bonaccorso, A.; Wieruszewski, P. M.; Fujii, T.; Khanna, A. K.; Landoni, G.; Bellomo, R. | 2024 | Critical care (London, England) | 28 | 1 | 439 | 10.1186/s13054-024-05212-7 |
| Kuklin 2024 | Influence of therapeutic plasma exchange treatment on short-term mortality of critically ill adult patients with sepsis-induced organ dysfunction: a systematic review and meta-analysis | Kuklin, V.; Sovershaev, M.; Bjerner, J.; Keith, P.; Scott, L. K.; Thomas, O. M. T.; Szpirt, W.; Rock, G.; Stegmayr, B. | 2024 | Critical care (London, England) | 28 | 1 | 12 | 10.1186/s13054-023-04795-x |
| Kuriyama 2018 | Impact of polymyxin B hemoperfusion in the treatment of patients with sepsis and septic shock: a meta-analysis of randomized controlled trials | Kuriyama, A.; Katsura, M.; Urushidani, S.; Takada, T. | 2018 | Annals of translational medicine | 6 | 11 | 206 | 10.21037/atm.2018.05.41 |
| Lang 2017 | Effects of goal directed therapy for adult patients in sepsis: A systemic review and meta-analysis of randomized controlled trials | Lang, X.; Yang, Y.; Zhang, P.; Lei, W.; Chen, J. | 2017 | International Journal of Clinical and Experimental Medicine | 10 | 3 | 4287-4296 | |
| Lee 2023 | Survival Benefits of Therapeutic Plasma Exchange in Severe Sepsis and Septic Shock: A Systematic Review and Meta-analysis | Lee, O. P. E.; Kanesan, N.; Leow, E. H.; Sultana, R.; Chor, Y. K.; Gan, C. S.; Lee, J. H. | 2023 | J Intensive Care Med | 38 | 7 | 598-611 | 10.1177/08850666231170775 |
| Lee 2016 | Protocol-Based Resuscitation for Septic Shock: A Meta-Analysis of Randomized Trials and Observational Studies | Lee, W. K.; Kim, H. Y.; Lee, J.; Koh, S. O.; Kim, J. M.; Na, S. | 2016 | Yonsei medical journal | 57 | 5 | 1260-70 | 10.3349/ymj.2016.57.5.1260 |

| | | | | | | | | |
|---|---|---|---|---|---|---|---|---|
| Leung 2024 | Door-to-antibiotic time and mortality in patients with sepsis: Systematic review and meta-analysis | Leung, L. Y.; Huang, H. L.; Hung, K. K.; Leung, C. Y.; Lam, C. C.; Lo, R. S.; Yeung, C. Y.; Tsoi, P. J.; Lai, M.; Brabrand, M.; Walline, J. H.; Graham, C. A. | 2024 | European journal of internal medicine | | | | 10.1016/j.ejim.2024.06.015 |
| Levy 2019 | Ischemic limb necrosis in septic shock: What is the role of high-dose vasopressor therapy? | Levy, J. H.; Ghadimi, K.; Faraoni, D.; van Diepen, S.; Levy, B.; Hotchkiss, R.; Connors, J. M.; Iba, T.; Warkentin, T. E. | 2019 | Journal of thrombosis and haemostasis : JTH | 17 | 11 | 1973-1978 | 10.1111/jth.14566 |
| Li 2024 | The role of polymyxin B-immobilized hemoperfusion in reducing mortality and enhancing hemodynamics in patients with sepsis and septic shock: A systematic review and meta-analysis | Li, C.; Zhang, J.; Yang, P.; Wang, R.; Chen, T.; Li, L. | 2024 | Heliyon | 10 | 13 | e33735 | 10.1016/j.heliyon.2024.e33735 |
| Li 2018 | Liberal versus conservative fluid therapy in adults and children with sepsis or septic shock | Li, D.; Li, X.; Cui, W.; Shen, H.; Zhu, H.; Xia, Y. | 2018 | The Cochrane database of systematic reviews | 12 | 12 | CD010593 | 10.1002/14651858.CD010593.pub2 |
| Li 2021 | Efficacy of intravenous vitamin C intervention for septic patients: A systematic review and meta-analysis based on randomized controlled trials | Li, T.; Zeng, J.; Li, D. H.; Yang, G. Y.; Wang, K.; Deng, H. F.; Jiang, H. | 2021 | The American journal of emergency medicine | 50 | | 242-250 | 10.1016/j.ajem.2021.08.012 |
| Li 2021 | Effectiveness of polymyxin B-immobilized hemoperfusion against sepsis and septic shock: A systematic review and meta-analysis | Li, X.; Liu, C.; Mao, Z.; Qi, S.; Song, R.; Zhou, F. | 2021 | J Crit Care | 63 | | 187-195 | 10.1016/j.jcrc.2020.09.007 |

| | | | | | | | | |
|---|---|---|---|---|---|---|---|---|
| Li 2021 | Therapeutic effect of low-molecular-weight heparin on adult sepsis: a meta-analysis | Li, X.; Liu, Z.; Luo, M.; Xi, Y.; Li, C.; Wang, S.; Yang, R. | 2021 | Annals of palliative medicine | 10 | 3 | 3115-3127 | 10.21037/apm-21-169 |
| Li 2023 | Prolonged vs intermittent intravenous infusion of β-lactam antibiotics for patients with sepsis: a systematic review of randomized clinical trials with meta-analysis and trial sequential analysis | Li, X.; Long, Y.; Wu, G.; Li, R.; Zhou, M.; He, A.; Jiang, Z. | 2023 | Annals of intensive care | 13 | 1 | 121 | 10.1186/s13613-023-01222-w |
| Li 2022 | Coupled plasma filtration adsorption for the treatment of sepsis or septic shock: a systematic review and meta-analysis | Li, Y.; Li, H.; Guo, J.; Wang, Y.; Zhang, D. | 2022 | BMC infectious diseases | 22 | 1 | 714 | 10.1186/s12879-022-07689-5 |
| Li 2019 | Timing of continuous renal replacement therapy in patients with septic AKI: A systematic review and meta-analysis | Li, Y.; Li, H.; Zhang, D. | 2019 | Medicine (Baltimore) | 98 | 33 | e16800 | 10.1097/md.0000000000016800 |
| Li 2020 | Timing of norepinephrine initiation in patients with septic shock: a systematic review and meta-analysis | Li, Y.; Li, H.; Zhang, D. | 2020 | Critical care (London, England) | 24 | 1 | 488 | 10.1186/s13054-020-03204-x |
| Liang 2023 | The outcome of IV vitamin C therapy in patients with sepsis or septic shock: a meta-analysis of randomized controlled trials | Liang, B.; Su, J.; Shao, H.; Chen, H.; Xie, B. | 2023 | Critical care (London, England) | 27 | 1 | 109 | 10.1186/s13054-023-04392-y |
| Liang 2023 | Effect of intravenous vitamin C on adult septic patients: a systematic review and meta-analysis | Liang, H.; Mu, Q.; Sun, W.; Liu, L.; Qiu, S.; Xu, Z.; Cui, Y.; Yan, Y.; Sun, T. | 2023 | Frontiers in nutrition | 10 | | 1211194 | 10.3389/fnut.2023.1211194 |

| | | | | | | | | |
|---|---|---|---|---|---|---|---|---|
| Liu 2016 | Effect of early goal directed therapy in the treatment of severe sepsis and/or septic shock: A meta-Analysis | Liu, B.; Ding, X.; Yang, J. | 2016 | Current medical research and opinion | 32 | 11 | 1-28 | 10.1080/03007995.2016.1206872 |
| Liu 2021 | Levosimendan versus dobutamine for sepsis-induced cardiac dysfunction: a systematic review and meta-analysis | Liu, D. H.; Ning, Y. L.; Lei, Y. Y.; Chen, J.; Liu, Y. Y.; Lin, X. F.; Yang, Z. Q.; Xian, S. X.; Chen, W. T. | 2021 | Sci Rep | 11 | 1 | 20333 | 10.1038/s41598-021-99716-9 |
| Liu 2022 | Effects of Albumin Supplements on In-Hospital Mortality in Patients with Sepsis or Septic Shock: A Systemic Review and Meta-Analysis | Liu, P.; Zhi, D.; Wang, Y.; Lin, J.; Zhang, M.; Duan, M. | 2022 | Evidence-based complementary and alternative medicine : eCAM | 2022 | | 2384730 | 10.1155/2022/2384730 |
| | Retracted: Effects of Albumin Supplements on In-Hospital Mortality in Patients with Sepsis or Septic Shock: A Systemic Review and Meta-Analysis | And Alternative Medicine, E. C. | 2023 | Evid Based Complement Alternat Med | 2023 | | 9821574 | 10.1155/2023/9821574 |
| Liu 2022 | Efficacy and safety of dexmedetomidine in sepsis patients requiring mechanical ventilation: a systematic review and meta-analysis | Liu, Z.; Zeng, Y.; Yang, B.; Liao, P. | 2022 | Journal of clinical pharmacy and therapeutics | 47 | 3 | 298-305 | 10.1111/jcpt.13548 |
| Lokhandwala 2023 | Comparison of the Effectiveness of Prolonged Infusion and Intermittent Infusion of Meropenem in Patients With Sepsis: A Meta-Analysis | Lokhandwala, A.; Patel, P.; Isaak, A. K.; Faizan Yousaf, R.; Maslamani, A. N. J.; Khalil, S. K.; Riaz, E.; Hirani, S. | 2023 | Cureus | 15 | 10 | e46990 | 10.7759/cureus.46990 |
| Lu 2017 | Omega-3 supplementation in | Lu, C.; Sharma, S.; McIntyre, L.; Rhodes, A.; Evans, L.; | 2017 | Annals of intensive care | 7 | 1 | 58 | 10.1186/s13613-017-0282-5 |

| | | | | | | | | |
|---|---|---|---|---|---|---|---|---|
| | patients with sepsis: a systematic review and meta-analysis of randomized trials | Almenawer, S.; Leduc, L.; Angus, D. C.; Alhazzani, W. | | | | | | |
| Lu 2016 | The effect of early goal-directed therapy on mortality in patients with severe sepsis and septic shock: A meta-analysis | Lu, J.; Wang, X.; Chen, Q.; Chen, M.; Cheng, L.; Dai, L.; Jiang, H.; Sun, Z. | 2016 | Journal of Surgical Research | 202 | 2 | 389-397 | 10.1016/j.jss.2015.12.048 |
| Lu 2021 | Norepinephrine was superior in death risk reducing and hemodynamics compared to dopamine in treatment of patients with septic shock | Lu, X.; Xu, X.; Wu, Y. | 2021 | Pteridines | 32 | 1 | 5-10 | 10.1515/pteridines-2021-0002 |
| Lu 2018 | Early Goal-Directed Therapy in Severe Sepsis and Septic Shock: A Meta-Analysis and Trial Sequential Analysis of Randomized Controlled Trials | Lu, Y.; Zhang, H.; Teng, F.; Xia, W. J.; Sun, G. X.; Wen, A. Q. | 2018 | Journal of intensive care medicine | 33 | 5 | 296-309 | 10.1177/0885066616671710 |
| Luo 2023 | The effect of vitamin C in adults with sepsis: a meta-analysis of randomized controlled trials | Luo, X.; Zhu, Y.; Zhang, R.; Zhu, J.; Kuang, H.; Shao, Y.; Guo, X.; Ning, B. | 2023 | Frontiers in medicine | 10 | | 1244484 | 10.3389/fmed.2023.1244484 |
| Ma 2025 | Extremely early initiation of vasopressors might not decrease short-term mortality for adults with septic shock: a systematic review and meta-analysis | Ma, C. H.; Healy, J.; Kinteh, E.; Ma, C. C.; Tzeng, C. T.; Chou, E. H.; Wu, C. C.; Shao, S. C.; Chen, K. F. | 2025 | Annals of intensive care | 15 | 1 | 18 | 10.1186/s13613-025-01428-0 |
| Ma 2015 | Sources of Heterogeneity in Trials Reporting Hydroxyethyl Starch 130/0.4 or 0.42 | Ma, P. L.; Peng, X. X.; Du, B.; Hu, X. L.; Gong, Y. C.; Wang, Y.; Xi, X. M. | 2015 | Chinese medical journal | 128 | 17 | 2374-82 | 10.4103/0366-6999.163387 |

| | | | | | | | | |
|---|---|---|---|---|---|---|---|---|
| | Associated Excess Mortality in Septic Patients: A Systematic Review and Meta-regression | | | | | | | |
| Ma 2023 | A Meta-Analysis of the Effects of Levosimendan on Cardiac Function and Outcomes in Patients with Sepsis | Ma, Z.; Jin, H.; Liu, F.; Wang, S.; Huang, P.; Fang, X. | 2023 | Alternative therapies in health and medicine | | | | |
| Mahdizad eAri 2024 | The effect of polyclonal and monoclonal based antibodies as promising potential therapy for treatment of sepsis: A systematic review | Mahdizade Ari, M.; Amini, M. E.; Sholeh, M.; Zahedi Bialvaei, A. | 2024 | New microbes and new infections | 60-61 | | 101435 | 10.1016/j.nmni.2024.101435 |
| Makam 2015 | Diagnostic accuracy and effectiveness of automated electronic sepsis alert systems: A systematic review | Makam, A. N.; Nguyen, O. K.; Auerbach, A. D. | 2015 | Journal of Hospital Medicine | 10 | (Makam A.N.,; Nguyen O.K.) Division of General Internal MedicineUniversity Texas Southwester | 396-402 | 10.1002/jhm.2347 |

| | | | | | | | | |
|---|---|---|---|---|---|---|---|---|
| | | | | | | n Medic al Center Dallas, Texas | | |
| Mandal 2022 | Efficacy and Safety of Vasopressin Alone or in Combination With Catecholamines in the Treatment of Septic Shock: A Systematic Review | Mandal, N.; Kham, N. I.; Shahid, R.; Naik, S. S.; Ramphall, S.; Rijal, S.; Prakash, V.; Ekladios, H.; Mulayamkuzhiyil Saju, J.; Venugopal, S. | 2022 | Cureus | 14 | 9 | e29143 | 10.7759/cureus.29 143 |
| Mangalore 2022 | Beta-lactam antibiotic therapeutic drug monitoring in critically ill patients: a systematic review and meta-analysis et al | Mangalore, R. P.; Ashok, A.; Lee, S. J.; Romero, L.; Peel, T. N.; Udy, A. A.; Peleg, A. Y. | 2022 | Clinical infectious diseases : an official publication of the Infectious Diseases Society of America | 75 | 10 | 1848-1860 | 10.1093/cid/ciac50 6 |
| Mao 2023 | Terlipressin combined with norepinephrine in the treatment of septic shock: a systematic review | Mao, F.; Liang, D.; Tang, Z.; Xu, Y.; Lin, L. | 2023 | Shock (Augusta, Ga.) | | | | 10.1097/SHK.0000 000000002204 |
| Marchetto 2024 | OUTCOMES OF PEDIATRIC FLUID-REFRACTORY SEPTIC SHOCK ACCORDING TO DIFFERENT VASOACTIVE STRATEGIES: A SYSTEMATIC REVIEW AND META-ANALYSIS | Marchetto, L.; Zanetto, L.; Comoretto, R. I.; Padrin, D.; Menon, K.; Amigoni, A.; Daverio, M. | 2024 | Shock (Augusta, Ga.) | 62 | 5 | 599-611 | 10.1097/SHK.0000 000000002427 |
| Martimbia nco 2022 | Vitamin C-based regimens for sepsis and septic shock: Systematic review and meta-analysis | Martimbianco, A. L. C.; Pacheco, R. L.; Bagattini Â, M.; de Fátima Carreira | 2022 | Journal of critical care | 71 | | 154099 | 10.1016/j.jcrc.2022 .154099 |

| | | | | | | | | |
|---|---|---|---|---|---|---|---|---|
| | of randomized clinical trials | Moreira Padovez, R.; Azevedo, L. C. P.; Riera, R. | | | | | | |
| Mathias 2022 | The effect of exchange transfusion on mortality in neonatal sepsis: a meta-analysis | Mathias, S.; Balachander, B.; Bosco, A.; Britto, C.; Rao, S. | 2022 | European journal of pediatrics | 181 | 1 | 369-381 | 10.1007/s00431-021-04194-w |
| Mavrothalassitis 2023 | Impact of Vasodilator Administration on Survival in Patients with Sepsis A Systematic Review and Meta-Analysis | Mavrothalassitis, O. Y.; Allen, I. E.; Lazzareschi, D. V.; Tahir, P.; Legrand, M. | 2023 | Annals of the American Thoracic Society | 20 | 9 | 1345-1352 | 10.1513/AnnalsATS.202303-205OC |
| Medeiros 2015 | Colloids for the Initial Management of Severe Sepsis and Septic Shock in Pediatric Patients: A Systematic Review | Medeiros, D. N.; Ferranti, J. F.; Delgado, A. F.; de Carvalho, W. B. | 2015 | Pediatric emergency care | 31 | 11 | e11-6 | 10.1097/PEC.0000000000000601 |
| Meng 2024 | Intensive or liberal glucose control in intensive care units for septic patients? A meta-analysis of randomized controlled trials | Meng, J.; Li, X.; Xiao, Y.; Tang, H.; Liu, P.; Wu, Y.; Xiong, Y.; Gao, S. | 2024 | Diabetes Metab Syndr | 18 | 5 | 103045 | 10.1016/j.dsx.2024.103045 |
| Meyhoff 2020 | Lower versus higher fluid volumes during initial management of sepsis - a systematic review with meta-analysis and trial sequential analysis | Meyhoff, T. S.; Møller, M. H.; Hjortrup, P. B.; Cronhjort, M.; Perner, A.; Wetterslev, J. | 2020 | Chest | 157 | 6 | 1478-1496 | 10.1016/j.chest.2019.11.050 |
| Mhanna 2024 | Balanced crystalloids versus isotonic saline in pediatric sepsis: a comprehensive systematic review and meta-analysis | Mhanna, A.; Beran, A.; Srour, O.; Mhanna, M.; Assaly, A.; Elsayed, A.; Horen, N. G.; Assaly, R. | 2024 | Proceedings (Baylor University. Medical Center) | 37 | 2 | 295-302 | 10.1080/08998280.2024.2301904 |
| Morton 2018 | The Early Recognition and Management of Sepsis in Sub-Saharan African Adults: A | Morton, B.; Stolbrink, M.; Kagima, W.; Rylance, J.; Mortimer, K. | 2018 | International journal of environmental | 15 | 9 | | 10.3390/ijerph15092017 |

| | | | | | | | | |
|---|---|---|---|---|---|---|---|---|
| | Systematic Review and Meta-Analysis | | | research and public health | | | | |
| Muhammad 2022 | The Role and Efficacy of Vitamin C in Sepsis: A Systematic Review and Meta-Analysis | Muhammad, M.; Jahangir, A.; Kassem, A.; Sattar, S. B. A.; Sahra, S.; Niazi, M. R. K.; Mustafa, A.; Zia, Z.; Siddiqui, F. S.; Sadiq, W.; Mishiyev, D.; Sammar, A.; Dahabra, L.; Irshad, A.; Elsayegh, D.; Chalhoub, M. | 2022 | Advances in respiratory medicine | 90 | 4 | 281-299 | 10.3390/arm90040038 |
| Nadeem 2017 | Impact of Dobutamine in Patients With Septic Shock: A Meta-Regression Analysis | Nadeem, Rashid; Sockanathan, Shivani; Singh, Mukesh; Hussain, Tamseela; Kent, Patrick; AbuAlreesh, Sarah | 2017 | American Journal of Therapeutics | 24 | 3 | 333-345 | |
| Orban 2025 | To Hemoadsorb or Not to Hemoadsorb—Do We Have the Answer Yet? An Updated Meta-Analysis on the Use of CytoSorb in Sepsis and Septic Shock | Orban, C.; Bratu, A.; Agapie, M.; Borjog, T.; Jafal, M.; Sima, R. M.; Dumitrașcu, O. C.; Popescu, M. | 2025 | Biomedicines | 13 | 1 | | 10.3390/biomedicines13010180 |
| Orbegozo 2019 | Hypertonic Saline in Human Sepsis: A Systematic Review of Randomized Controlled Trials | Orbegozo, D.; Vincent, J. L.; Creteur, J.; Su, F. | 2019 | Anesthesia and analgesia | 128 | 6 | 1175-1184 | 10.1213/ANE.0000000000003955 |
| Pan 2023 | Efficacy of IVIG therapy for patients with sepsis: a systematic review and meta-analysis | Pan, B.; Sun, P.; Pei, R.; Lin, F.; Cao, H. | 2023 | Journal of translational medicine | 21 | 1 | 765 | 10.1186/s12967-023-04592-8 |
| Pan 2019 | Relative efficacy and safety of early lactate clearance-guided therapy resuscitation in patients with sepsis: A meta-analysis | Pan, J.; Peng, M.; Liao, C.; Hu, X.; Wang, A.; Li, X. | 2019 | Medicine | 98 | 8 | e14453 | 10.1097/MD.0000000000014453 |
| Park 2017 | The effect of early goal-directed therapy for treatment of severe | Park, S. K.; Shin, S. R.; Hur, M.; Kim, W. H.; Oh, E. A.; Lee, S. H. | 2017 | Journal of Critical Care | 38 | | 115-122 | 10.1016/j.jcrc.2016.10.019 |

| | | | | | | | | |
|---|---|---|---|---|---|---|---|---|
| | sepsis or septic shock: A systemic review and meta-analysis | | | | | | | |
| Pascual-Ramirez 2019 | The nitric oxide pathway antagonists in septic shock: Meta-analysis of controlled clinical trials | Pascual-Ramirez, J.; Koutrouvelis, A. | 2019 | Journal of critical care | 51 | | 34-38 | 10.1016/j.jcrc.2019.01.013 |
| Pei 2024 | The effects of antioxidant supplementation on short-term mortality in sepsis patients | Pei, H.; Qu, J.; Chen, J. M.; Zhang, Y. L.; Zhang, M.; Zhao, G. J.; Lu, Z. Q. | 2024 | Heliyon | 10 | 8 | e29156 | 10.1016/j.heliyon.2024.e29156 |
| Pence 2022 | Outcomes of CMS-mandated fluid administration among fluid-overloaded patients with sepsis: A systematic review and meta-analysis | Pence, M.; Tran, Q. K.; Shesser, R.; Payette, C.; Pourmand, A. | 2022 | The American journal of emergency medicine | 55 | | 157-166 | 10.1016/j.ajem.2022.03.004 |
| Pepper 2018 | Evidence Underpinning the U.S. Government-Mandated Hemodynamic Interventions for Sepsis: A Systematic Review | Pepper, D. J.; Jaswal, D.; Sun, J.; Welsh, J.; Natanson, C.; Eichacker, P. Q. | 2018 | Annals of internal medicine | 168 | 4f | | 10.7326/M17-2947 |
| Pepper 2019 | Antibiotic- and Fluid-Focused Bundles Potentially Improve Sepsis Management, but High-Quality Evidence Is Lacking for the Specificity Required in the Centers for Medicare and Medicaid Service's Sepsis Bundle (SEP-1) | Pepper, D. J.; Sun, J.; Cui, X.; Welsh, J.; Natanson, C.; Eichacker, P. Q. | 2019 | Critical care medicine | 47 | 10 | 1290-1300 | 10.1097/CCM.0000000000003892 |
| Potter 2019 | Manipulating the Microcirculation in Sepsis - the Impact of Vasoactive Medications on Microcirculatory Blood Flow. A Systematic Review | Potter, E. K.; Hodgson, L.; Creagh-Brown, B.; Forni, L. G. | 2019 | Shock (Augusta, Ga.) | 52 | 1 | 5-12 | 10.1097/SHK.0000000000001239 |

| | | | | | | | | |
|---|---|---|---|---|---|---|---|---|
| Putzu 2017 | Blood purification with continuous veno-venous hemofiltration in patients with sepsis or ARDS: a systematic review and meta-analysis | Putzu, A.; Fang, M. X.; Boscolo Berto, M.; Belletti, A.; Cabrini, L.; Cassina, T.; Landoni, G. | 2017 | Minerva anestesiologica | 83 | 8 | 867-877 | 10.23736/S0375-9393.17.11946-2 |
| Putzu 2019 | Blood Purification and Mortality in Sepsis and Septic Shock: A Systematic Review and Meta-analysis of Randomized Trials | Putzu, A.; Schorer, R.; Lopez-Delgado, J. C.; Cassina, T.; Landoni, G. | 2019 | Anesthesiology | 131 | 3 | 580-593 | 10.1097/ALN.0000000000002820 |
| Qayyum 2023 | Fluid Resuscitation in Septic Patients | Qayyum, S.; Shahid, K. | 2023 | Cureus | 15 | 8 | e44317 | 10.7759/cureus.44317 |
| Qian 2020 | Intravenous thiamine for septic shock: A meta-analysis of randomized controlled trials | Qian, X.; Zhang, Z.; Li, F.; Wu, L. | 2020 | Am. J. Emerg. Med. | 38 | 12 | 2718-2722 | 10.1016/j.ajem.2020.08.050 |
| Ramanathan 2020 | Role of extracorporeal membrane oxygenation in children with sepsis: a systematic review and meta-analysis | Ramanathan, K.; Yeo, N.; Alexander, P.; Raman, L.; Barbaro, R.; Tan, C. S.; Schlapbach, L. J.; MacLaren, G. | 2020 | Critical care (London, England) | 24 | 1 | 684 | 10.1186/s13054-020-03418-z |
| Rao 2016 | One dose per day compared to multiple doses per day of gentamicin for treatment of suspected or proven sepsis in neonates | Rao, Shripada C.; Srinivasjois, Ravisha; Moon, Kwi | 2016 | Cochrane Database of Systematic Reviews | 12 | 12 | CD005091 | 10.1002/14651858.CD005091.pub4 |
| Reynolds 2023 | Restrictive resuscitation in patients with sepsis and mortality: A systematic review and meta-analysis with trial sequential analysis | Reynolds, P. M.; Stefanos, S.; MacLaren, R. | 2023 | Pharmacotherapy | 43 | 2 | 104-114 | 10.1002/phar.2764 |
| Reynolds 2020 | Establishing the Therapeutic Index of Fluid Resuscitation in the Septic Patient: A | Reynolds, P. M.; Wells, L.; MacLaren, R.; Scoular, S. K. | 2020 | Pharmacotherapy | 40 | 3 | 256-269 | 10.1002/phar.2371 |

| | | | | | | | | |
|---|---|---|---|---|---|---|---|---|
| | Narrative Review and Meta-Analysis | | | | | | | |
| Rodríguez 2025 | Early empirical antibiotherapy in patients attended for suspected sepsis in emergency departments: a systematic review | Rodríguez, M. R.; Llopis Roca, F.; Rubio Díaz, R.; García, D. E.; Julián-Jiménez, A. | 2025 | Emergencias : revista de la Sociedad Espanola de Medicina de Emergencias | 37 | 1 | 44-55 | 10.55633/s3me/092.2024 |
| Rothrock 2020 | Outcome of Immediate Versus Early Antibiotics in Severe Sepsis and Septic Shock: A Systematic Review and Meta-analysis | Rothrock, S. G.; Cassidy, D. D.; Barneck, M.; Guetschow, B.; Myburgh, C.; Briscoe, J. G.; Nguyen, L.; Earwood, R.; Schinkel, M.; Nanayakkara, P. W. B.; Nannan Panday, R. S. | 2020 | Ann. Emerg. Med. | 76 | 4 | 427-441 | 10.1016/j.annemergmed.2020.04.042 |
| Rusconi 2015 | Early goal-directed therapy vs usual care in the treatment of severe sepsis and septic shock: a systematic review and meta-analysis | Rusconi, A. M.; Bossi, I.; Lampard, J. G.; Szava-Kovats, M.; Bellone, A.; Lang, E. | 2015 | Internal and emergency medicine | 10 | 6 | 731-43 | 10.1007/s11739-015-1248-y |
| Ruslan 2021 | Norepinephrine in septic shock: A systematic review and meta-analysis | Ruslan, M. A.; Baharuddin, K. A.; Noor, N. M.; Yazid, M. B.; Noh, A. Y. M.; Rahman, A. | 2021 | The western journal of emergency medicine | 22 | 2 | 196-203 | 10.5811/WESTJEM.2020.10.47825 |
| Sæterdal 2015 | Effect of Using Aminoglycosides for Treatment of Sepsis | Sæterdal, I.; Holte, H. H.; Harboe, I.; Klemp, M. | 2015 | NIPH Systematic Reviews | | | | |
| Saldaña-Gastulo 2023 | Cytokine hemoadsorption with CytoSorb® in patients with sepsis: a systematic review and meta-analysis | Saldaña-Gastulo, J. J. C.; Llamas-Barbarán, M. D. R.; Coronel-Chucos, L. G.; Hurtado-Roca, Y. | 2023 | Critical care science | 35 | 2 | 217-225 | 10.5935/2965-2774.20230289-en |
| Sangla 2023 | Thiamine as a metabolic resuscitator in septic shock: a meta-analysis of randomized controlled trials with trial sequential analysis | Sangla, F.; Verissimo, T.; Faivre, A.; Glauser, T.; Cheah, S. K.; Assouline, B.; Sgardello, S.; Legouis, D. | 2023 | Front Med (Lausanne) | 10 | | 1223862 | 10.3389/fmed.2023.1223862 |

| | | | | | | | | |
|---|---|---|---|---|---|---|---|---|
| Sankar 2024 | Fluid resuscitation in children with severe infection and septic shock: a systematic review and meta-analysis | Sankar, J.; Das, R. R.; Banothu, K. K. | 2024 | European journal of pediatrics | 183 | 9 | 3925-3932 | 10.1007/s00431-024-05653-w |
| Sankar 2022 | Comparison of Intermittent versus Continuous Superior Venal Caval Oxygen Saturation Monitoring in Early Goal Directed Therapy in Septic Shock: A Systematic Review | Sankar, J. M.; Das, R. R.; Kumar, U. V. | 2022 | J Pediatr Intensive Care | 11 | 4 | 267-274 | 10.1055/s-0041-1729742 |
| Sato 2021 | Effect of IV High-Dose Vitamin C on Mortality in Patients With Sepsis: A Systematic Review and Meta-Analysis of Randomized Controlled Trials | Sato, R.; Hasegawa, D.; Prasitlumkum, N.; Ueoka, M.; Nishida, K.; Takahashi, K.; Nasu, M.; Dugar, S. | 2021 | Critical care medicine | 49 | 12 | 2121-2130 | 10.1097/CCM.0000000000005263 |
| Sato 2020 | Venoarterial Extracorporeal Membranous Oxygenation: Treatment Option for Sepsis-Induced Cardiogenic Shock? A Systematic Review | Sato, R.; Kuriyama, A. | 2020 | Critical care medicine | 48 | 8 | e722-e729 | 10.1097/CCM.0000000000004432 |
| Scholz 2021 | Mortality in septic patients treated with vitamin C: a systematic meta-analysis | Scholz, S. S.; Borgstedt, R.; Ebeling, N.; Menzel, L. C.; Jansen, G.; Rehberg, S. | 2021 | Critical care (London, England) | 25 | 1 | 17 | 10.1186/s13054-020-03438-9 |
| Scully 2020 | The effects of static and dynamic measurements using transpulmonary thermodilution devices on fluid therapy in septic shock: A systematic review | Scully, T. G.; Huang, Y.; Huang, S.; McLean, A. S.; Orde, S. R. | 2020 | Anaesthesia and intensive care | | | 310057X19893703 | 10.1177/0310057X19893703 |

| | | | | | | | | |
|---|---|---|---|---|---|---|---|---|
| Seale 2018 | Does antibiotic choice for the treatment of suspected late-onset sepsis in premature infants determine the risk of developing necrotising enterocolitis? A systematic review | Seale, J. V.; Hutchinson, R. A.; Fleming, P. F.; Sinha, A.; Kempley, S. T.; Husain, S. M.; Millar, M. R. | 2018 | Early human development | 123 | | 6-10 | 10.1016/j.earlhumdev.2018.06.009 |
| Sedhai 2022 | Vasopressin versus norepinephrine as the first-line vasopressor in septic shock: A systematic review and meta-analysis | Sedhai, Y. R.; Shrestha, D. B.; Budhathoki, P.; Memon, W.; Acharya, R.; Gaire, S.; Pokharel, N.; Maharjan, S.; Jasaraj, R.; Sodhi, A.; Kadariya, D.; Asija, A.; Kashiouris, M. G. | 2022 | Journal of clinical and translational research | 8 | 3 | 185-199 | |
| Shahnoor 2023 | The Effects of Restrictive Fluid Resuscitation on the Clinical Outcomes in Patients with Sepsis or Septic Shock: A Meta-Analysis of Randomized-Controlled Trials | Shahnoor, H.; Divi, R.; Addi Palle, L. R.; Sharma, A.; Contractor, B.; Krupanagaram, S.; Batool, S.; Ali, N. | 2023 | Cureus | 15 | 9 | e45620 | 10.7759/cureus.45620 |
| Sherwin 2017 | Does Early and Appropriate Antibiotic Administration Improve Mortality in Emergency Department Patients with Severe Sepsis or Septic Shock? | Sherwin, R.; Winters, M. E.; Vilke, G. M.; Wardi, G. | 2017 | The Journal of emergency medicine | 53 | 4 | 588-595 | 10.1016/j.jemermed.2016.12.009 |
| Shi 2025 | Early norepinephrine for patients with septic shock: an updated systematic review and meta-analysis with trial sequential analysis | Shi, R.; Braïk, R.; Monnet, X.; Gu, W. J.; Ospina-Tascon, G.; Permpikul, C.; Djebbour, M.; Soumare, A.; Agaleridis, V.; Lai, C. | 2025 | Critical care (London, England) | 29 | 1 | 182 | 10.1186/s13054-025-05400-z |
| Shiber 2015 | β-Lactam/β-lactamase inhibitors versus carbapenems for the treatment of sepsis: | Shiber, S.; Yahav, D.; Avni, T.; Leibovici, L.; Paul, M. | 2015 | The Journal of antimicrobial chemotherapy | 70 | 1 | 41-47 | 10.1093/jac/dku351 |

| | | | | | | | | |
|---|---|---|---|---|---|---|---|---|
| | Systematic review and meta-analysis of randomized controlled trials | | | | | | | |
| Silva 2018 | Inadequate use of antibiotics and increase in neonatal sepsis caused by resistant bacteria related to health care assistance: a systematic review | Silva, A. C. B.; Anchieta, L. M.; Lopes, M. F. P.; Romanelli, R. M. C. | 2018 | The Brazilian journal of infectious diseases : an official publication of the Brazilian Society of Infectious Diseases | 22 | 4 | 328-337 | 10.1016/j.bjid.2018.07.009 |
| Silversides 2017 | Conservative fluid management or deresuscitation for patients with sepsis or acute respiratory distress syndrome following the resuscitation phase of critical illness: a systematic review and meta-analysis | Silversides, J. A.; Major, E.; Ferguson, A. J.; Mann, E. E.; McAuley, D. F.; Marshall, J. C.; Blackwood, B.; Fan, E. | 2017 | Intensive care medicine | 43 | 2 | 1-16 | 10.1007/s00134-016-4573-3 |
| Simpson 2016 | Early goal-directed therapy for severe sepsis and septic shock: A living systematic review | Simpson, S. Q.; Gaines, M.; Hussein, Y.; Badgett, R. G. | 2016 | Journal of critical care | 36 | | 43-48 | 10.1016/j.jcrc.2016.06.017 |
| Singh 2024 | Efficacy of short-course antibiotics for culture-positive neonatal sepsis: A systematic review and meta-analysis | Singh, P.; Priyadarshi, M.; Chaurasia, S.; Basu, S. | 2024 | Journal of paediatrics and child health | 60 | 11 | 630-639 | 10.1111/jpc.16647 |
| Sivapalan 2023 | Lower vs Higher Fluid Volumes in Adult Patients with Sepsis - An Updated Systematic Review with Meta-Analysis and Trial Sequential Analysis | Sivapalan, P.; Ellekjaer, K. L.; Jessen, M. K.; Meyhoff, T. S.; Cronhjort, M.; Hjortrup, P. B.; Wetterslev, J.; Granholm, A.; Møller, M. H.; Perner, A. | 2023 | Chest | 164 | 4 | 892-912 | 10.1016/j.chest.2023.04.036 |

| | | | | | | | | |
|---|---|---|---|---|---|---|---|---|
| Sjövall 2017 | Empirical mono- versus combination antibiotic therapy in adult intensive care patients with severe sepsis - A systematic review with meta-analysis and trial sequential analysis | Sjövall, F.; Perner, A.; Hylander Møller, M. | 2017 | J Infect | 74 | 4 | 331-344 | 10.1016/j.jinf.2016.11.013 |
| Snow 2021 | Effect of Extracorporeal Blood Purification on Mortality in Sepsis: A Meta-Analysis and Trial Sequential Analysis | Snow, T. A. C.; Littlewood, S.; Corredor, C.; Singer, M.; Arulkumaran, N. | 2021 | Blood purification | 50 | 4-5 | 1-11 | 10.1159/000510982 |
| Somaili 2022 | Early versus Delayed Strategies for Renal Replacement Therapy Initiation in Adult Patients with Severe Acute Kidney Injury Complicating Septic Shock: A Systematic Review and Meta-analysis | Somaili, M. | 2022 | Saudi J Kidney Dis Transpl | 33 | 3 | 449-486 | 10.4103/1319-2442.385969 |
| Song 2020 | Incidence of hypotension after discontinuation of norepinephrine or arginine vasopressin in patients with septic shock: A systematic review and meta-analysis | Song, J. U.; Lee, J.; Park, H. K.; Suh, G. Y.; Jeon, K. | 2020 | Journal of Korean medical science | 35 | 1 | e8 | 10.3346/jkms.2020.35.e8 |
| Steindl 2025 | Hemoadsorption in the Management of Septic Shock: A Systematic Review and Meta-Analysis | Steindl, D.; Schroeder, T.; Krannich, A.; Nee, J. | 2025 | Journal of clinical medicine | 14 | 7 | | 10.3390/jcm14072285 |
| Sterling 2015 | The Impact of Timing of Antibiotics on Outcomes in Severe Sepsis and Septic Shock: A | Sterling, S. A.; Miller, W. R.; Pryor, J.; Puskarich, M. A.; Jones, A. E. | 2015 | Critical care medicine | 43 | 9 | 1907-15 | 10.1097/CCM.0000000000001142 |

| | | | | | | | | |
|---|---|---|---|---|---|---|---|---|
| | Systematic Review and Meta-Analysis | | | | | | | |
| Sukudom 2024 | Association between intravenous fluid administration and endothelial glycocalyx shedding in humans: a systematic review | Sukudom, S.; Smart, L.; Macdonald, S. | 2024 | Intensive care medicine experimental | 12 | 1 | 16 | 10.1186/s40635-024-00602-1 |
| Taj 2022 | Sepsis protocols to reduce mortality in resource-restricted settings: A systematic review | Taj, M.; Brenner, M.; Sulaiman, Z.; Pandian, V. | 2022 | Intensive & critical care nursing | | | 103255 | 10.1016/j.iccn.2022.103255 |
| Tan 2016 | Vasopressin and its analog terlipressin versus norepinephrine in the treatment of septic shock: A meta-analysis | Tan, J.; Chen, H.; Chen, X.; Zhang, D.; He, F. | 2016 | International Journal of Clinical and Experimental Medicine | 9 | 7 | 14183-14190 | |
| Tang 2024 | Effect of delayed antibiotic use on mortality outcomes in patients with sepsis or septic shock: A systematic review and meta-analysis | Tang, F.; Yuan, H.; Li, X.; Qiao, L. | 2024 | International immunopharmacology | 129 | | 111616 | 10.1016/j.intimp.2024.111616 |
| Tang 2019 | Efficacy of zinc supplementation for neonatal sepsis: a systematic review and meta-analysis | Tang, Z.; Wei, Z.; Wen, F.; Wu, Y. | 2019 | The journal of maternal-fetal & neonatal medicine : the official journal of the European Association of Perinatal Medicine, the Federation of Asia and Oceania Perinatal Societies, the International | 32 | 7 | 1-131 | 10.1080/14767058.2017.1402001 |

| | | | | | | | | |
|---|---|---|---|---|---|---|---|---|
| | | | | Society of Perinatal Obstetricians | | | | |
| Tao 2016 | Effects of omega-3 fatty acid nutrition on mortality in septic patients: a meta-analysis of randomized controlled trials | Tao, W.; Li, P. S.; Shen, Z.; Shu, Y. S.; Liu, S. | 2016 | BMC Anesthesiol | 16 | 1 | 39 | 10.1186/s12871-016-0200-7 |
| Terayama 2017 | Polymyxin B Hemoperfusion for Sepsis and Septic Shock: A Systematic Review and Meta-Analysis | Terayama, T.; Yamakawa, K.; Umemura, Y.; Aihara, M.; Fujimi, S. | 2017 | Surgical infections | 18 | 3 | 225-233 | 10.1089/sur.2016.168 |
| Tigabu 2018 | Fluid volume, fluid balance and patient outcome in severe sepsis and septic shock: A systematic review | Tigabu, B. M.; Davari, M.; Kebriaeezadeh, A.; Mojtahedzadeh, M. | 2018 | Journal of critical care | 48 | | 153-159 | 10.1016/j.jcrc.2018.08.018 |
| Totoki 2024 | Heparin therapy in sepsis and sepsis-associated disseminated intravascular coagulation: a systematic review and meta-analysis | Totoki, T.; Koami, H.; Makino, Y.; Wada, T.; Ito, T.; Yamakawa, K.; Iba, T. | 2024 | Thrombosis journal | 22 | 1 | 84 | 10.1186/s12959-024-00653-0 |
| VaeliZadeh 2023 | Guideline-based and restricted fluid resuscitation strategy in sepsis patients with heart failure: A systematic review and meta-analysis | Vaeli Zadeh, A.; Wong, A.; Crawford, A. C.; Collado, E.; Larned, J. M. | 2023 | The American journal of emergency medicine | 73 | | 34-39 | 10.1016/j.ajem.2023.08.006 |
| Vardakas 2018 | Prolonged versus short-term intravenous infusion of antipseudomonal β-lactams for patients with sepsis: a systematic review and meta-analysis of randomised trials | Vardakas, K. Z.; Voulgaris, G. L.; Maliaros, A.; Samonis, G.; Falagas, M. E. | 2018 | The Lancet. Infectious diseases | 18 | 1 | 108-120 | 10.1016/S1473-3099(17)30615-1 |
| Vijendra 2025 | Balanced crystalloid versus saline for | Vijendra, B.; Bertol, A. B.; de Almeida, M. M. G.; de | 2025 | BMC pediatrics | 25 | 1 | 81 | 10.1186/s12887-025-05442-w |

| | | | | | | | | |
|---|---|---|---|---|---|---|---|---|
| | resuscitation in pediatric septic shock: a systematic review and meta-analysis | Freitas, Phag; Simão Á, M. S.; de Faria, B. L. | | | | | | |
| Villa 2016 | Effects of continuous renal replacement therapy on linezolid pharmacokinetic/pharmacodynamics: a systematic review | Villa, G.; Di Maggio, P.; De Gaudio, A. R.; Novelli, A.; Antoniotti, R.; Fiaccadori, E.; Adembri, C. | 2016 | Crit Care | 20 | 1 | 374 | 10.1186/s13054-016-1551-7 |
| Wang 2022 | Effect of Pulse Indicator Continuous Cardiac Output Monitoring on Septic Shock Patients: A Meta-Analysis | Wang, B.; Cai, L.; Lin, B.; He, Q.; Ding, X. | 2022 | Computational and mathematical methods in medicine | 2022 | | 8604322 | 10.1155/2022/8604322 |
| Wang 2017 | Effects of levosimendan on mortality in patients with septic shock: Systematic review with meta-analysis and trial sequential analysis | Wang, B.; Chen, R.; Guo, X.; Zhang, W.; Hu, J.; Gong, Y.; Cheng, B. | 2017 | Oncotarget | 8 | 59 | 100524-100532 | 10.18632/oncotarget.20123 |
| Wang 2021 | The Effect of Dexmedetomidine as a Sedative Agent for Mechanically Ventilated Patients With Sepsis: A Systematic Review and Meta-Analysis | Wang, C.; Chen, Q.; Wang, P.; Jin, W.; Zhong, C.; Ge, Z.; Xu, K. | 2021 | Frontiers in medicine | 8 | | 776882 | 10.3389/fmed.2021.776882 |
| Wang 2020 | Omega-3 fatty acid supplementation is associated with favorable outcomes in patients with sepsis: an updated meta-analysis | Wang, C.; Han, D.; Feng, X.; Wu, J. | 2020 | The Journal of international medical research | 48 | 12 | | 10.1177/0300060520953684 |
| Wang 2023 | Continuous renal replacement therapy with the adsorptive oXiris filter may be associated with the lower 28-day mortality | Wang, G.; He, Y.; Guo, Q.; Zhao, Y.; He, J.; Chen, Y.; Chen, W.; Zhou, Y.; Peng, Z.; Deng, K.; Guan, J.; Xie, W.; Chang, P.; Liu, Z. | 2023 | Critical care (London, England) | 27 | 1 | 275 | 10.1186/s13054-023-04555-x |

| | | | | | | | | |
|---|---|---|---|---|---|---|---|---|
| | in sepsis: a systematic review and meta-analysis | | | | | | | |
| Wang 2022 | Effects of fish oil-containing nutrition supplementation in adult sepsis patients: a systematic review and meta-analysis | Wang, H.; Su, S.; Wang, C.; Hu, J.; Dan, W.; Peng, X. | 2022 | Burns & trauma | 10 | | tkac012 | 10.1093/burnst/tkac012 |
| Wang 2024 | Timing of renal replacement therapy in patients with sepsis-associated acute kidney injury: A systematic review and meta-analysis | Wang, Q.; Liu, F.; Tao, W.; Qian, K. | 2024 | Aust Crit Care | 37 | 2 | 369-379 | 10.1016/j.aucc.2023.06.011 |
| Wang 2024 | Clinical efficacy of various resuscitation fluids in the management of sepsis in postoperative surgical and trauma patients: a systematic review and meta--analysis | Wang, Y.; Chen, K.; Li, X.; Guan, J. | 2024 | Wideochir Inne Tech Maloinwazyjne | 19 | 3 | 275-288 | 10.20452/wiitm.2024.17900 |
| Ward 2022 | The Effect of Fluid Initiation Timing on Sepsis Mortality: A Meta-Analysis | Ward, M. A.; Kuttab, H. I.; Tuck, N.; Taleb, A.; Okut, H.; Badgett, R. G. | 2022 | Journal of intensive care medicine | 37 | 11 | | 10.1177/08850666221118513 |
| Wei 2020 | Efficacy of vitamin C in patients with sepsis: A updated meta-analysis | Wei, X. B.; Wang, Z. H.; Liao, X. L.; Guo, W. X.; Wen, J. Y.; Qin, T. H.; Wang, S. H. | 2020 | European journal of pharmacology | 868 | | 172889 | 10.1016/j.ejphar.2019.172889 |
| Wen 2023 | IV Vitamin C in Sepsis: A Latest Systematic Review and Meta-Analysis | Wen, C.; Li, Y.; Hu, Q.; Liu, H.; Xu, X.; Lü, M. | 2023 | International journal of clinical practice | 2023 | | 6733465 | 10.1155/2023/6733465 |
| Wen 2020 | The efficacy of dopamine versus epinephrine for pediatric or neonatal septic shock: A meta-analysis of randomized controlled studies | Wen, L.; Xu, L. | 2020 | Italian journal of pediatrics | 46 | 1 | 6 | 10.1186/s13052-019-0768-x |

| | | | | | | | | |
|---|---|---|---|---|---|---|---|---|
| Winters 2017 | Does Early Goal-Directed Therapy Decrease Mortality Compared with Standard Care in Patients with Septic Shock? | Winters, M. E.; Sherwin, R.; Vilke, G. M.; Wardi, G. | 2017 | The Journal of emergency medicine | 52 | 3 | 379-384 | 10.1016/j.jemermed.2016.10.028 |
| Wolbrink 2020 | Are omega-3 fatty acids safe and effective in acute pancreatitis or sepsis? A systematic review and meta-analysis | Wolbrink, D. R. J.; Grundsell, J. R.; Witteman, B.; Poll, M. V.; Santvoort, H. C. V.; Issa, E.; Dennison, A.; Goor, H. V.; Besselink, M. G.; Bouwense, S. A. W. | 2020 | Clin Nutr | 39 | 9 | 2686-2694 | 10.1016/j.clnu.2019.12.006 |
| Wu 2020 | Norepinephrine vs vasopressin: which vasopressor should be discontinued first in septic shock? a meta-analysis | Wu, Z.; Zhang, S.; Xu, J.; Xie, J.; Huang, L.; Huang, Y.; Yang, Y.; Qiu, H. | 2020 | Shock (Augusta, Ga.) | 53 | 1 | 50-57 | 10.1097/SHK.0000000000001345 |
| Xantus 2020 | Antibiotics administered within 1 hour to adult emergency department patients screened positive for sepsis: a systematic review | Xantus, G.; Allen, P.; Norman, S.; Kanizsai, P. | 2020 | European journal of emergency medicine : official journal of the European Society for Emergency Medicine | 27 | 4 | 260-267 | 10.1097/MEJ.0000000000000654 |
| Xantus 2021 | Mortality benefit of crystalloids administered in 1-6 hours in septic adults in the ED systematic review with narrative synthesis | Xantus, G. Z.; Allen, P.; Norman, S.; Kanizsai, P. L. | 2021 | Emergency medicine journal : EMJ | | | | 10.1136/emermed-2020-210298 |
| Xu 2016 | The Effect of Early Goal-Directed Therapy on Outcome in Adult Severe Sepsis and Septic Shock Patients: A Meta-Analysis of Randomized Clinical Trials | Xu, J. Y.; Chen, Q. H.; Liu, S. Q.; Pan, C.; Xu, X. P.; Han, J. B.; Xie, J. F.; Huang, Y. Z.; Guo, F. M.; Yang, Y.; Qiu, H. B. | 2016 | Anesthesia and analgesia | 123 | 2 | 371-81 | 10.1213/ANE.0000000000001278 |

| | | | | | | | | |
|---|---|---|---|---|---|---|---|---|
| Xu 2025 | Application of norepinephrine in the treatment of septic shock: a meta-analysis | Xu, Q. Y.; Jin, Y. H.; Fu, L.; Li, Y. Y. | 2025 | Ir J Med Sci | 194 | 1 | 361-369 | 10.1007/s11845-024-03827-x |
| Yang 2025 | Prognosis of dexmedetomidine sedation in patients with sepsis: a systematic review and meta-analysis | Yang, Q.; Wang, M.; Zhang, S.; Fan, C.; Xue, F. | 2025 | Minerva Anestesiol | 91 | 5 | 450-461 | 10.23736/s0375-9393.25.18560-x |
| Yao 2020 | Clinical efficiency of vasopressin or its analogs in comparison with catecholamines alone on patients with septic shock: A systematic review and meta-analysis | Yao, R. Q.; Xia, D. M.; Wang, L. X.; Wu, G. S.; Zhu, Y. B.; Zhao, H. Q.; Liu, Q.; Xia, Z. F.; Ren, C.; Yao, Y. M. | 2020 | Frontiers in pharmacology | 11 | | 563 | 10.3389/fphar.2020.00563 |
| Ye 2023 | Initiation timing of vasopressor in patients with septic shock: a systematic review and meta-analysis | Ye, E.; Ye, H.; Wang, S.; Fang, X. | 2023 | Shock (Augusta, Ga.) | 60 | 5 | 627-636 | 10.1097/SHK.0000000000002214 |
| Yeo 2020 | Role of Extracorporeal Membrane Oxygenation in pediatric sepsis: Systematic Review and metanalysis | Yeo, N.; Kollengode, R.; MacLaren, G.; Tan, C. S.; Alexander, P.; Rama, L.; Barbaro, R.; Schlapbach, L. J. | 2020 | ASAIO J. | 66 | SUPPL 3 | 47-None | |
| Yin 2020 | The therapeutic effect of high-volume hemofiltration on sepsis: A systematic review and meta-analysis | Yin, F.; Zhang, F.; Liu, S.; Ning, B. | 2020 | Annals of translational medicine | 8 | 7 | 488 | 10.21037/atm.2020.03.48 |
| Yu 2016 | Effect of early goal-directed therapy on mortality in patients with severe sepsis or septic shock: a meta-analysis of randomised controlled trials | Yu, H.; Chi, D.; Wang, S.; Liu, B. | 2016 | BMJ open | 6 | 3 | e008330 | 10.1136/bmjopen-2015-008330 |

| | | | | | | | | |
|---|---|---|---|---|---|---|---|---|
| Yuan 2022 | The Effect of Dexmedetomidine on the Prognosis of Mechanically Ventilated Patients with Sepsis: A Meta-Analysis of Randomized Controlled Trials | Yuan, J.; Lu, S. F.; Xu, P.; Niu, Y. L. | 2022 | Iranian journal of public health | 51 | 10 | 2171-2180 | 10.18502/ijph.v51i10.10976 |
| Zamani 2016 | Survival benefits of dexmedetomidine used for sedating septic patients in intensive care setting: A systematic review | Zamani, M. M.; Keshavarz-Fathi, M.; Fakhri-Bafghi, M. S.; Hirbod-Mobarakeh, A.; Rezaei, N.; Bahrami, A.; Nader, N. D. | 2016 | Journal of critical care | 32 | | 93-100 | 10.1016/j.jcrc.2015.11.013 |
| Zangrillo 2015 | Levosimendan reduces mortality in patients with severe sepsis and septic shock: A meta-analysis of randomized trials | Zangrillo, Alberto; Putzu, Alessandro; Monaco, Fabrizio; Oriani, Alessandro; Frau, Giovanna; De Luca, Monica; Di Tomasso, Nora; Bignami, Elena; Lomivorotov, Vladimir; Likhvantsev, Valery; Landoni, Giovanni | 2015 | Journal of Critical Care | 30 | 5 | 908-913 | 10.1016/j.jcrc.2015.05.017 |
| Zeng 2023 | Intravenous high-dose vitamin C monotherapy for sepsis and septic shock: A meta-analysis of randomized controlled trials | Zeng, Y.; Liu, Z.; Xu, F.; Tang, Z. | 2023 | Medicine | 102 | 42 | e35648 | 10.1097/MD.0000000000035648 |
| Zhang 2023 | An inquiry into the treatment of sepsis using plasma exchange therapy: A systematic review and meta-analysis | Zhang, L.; Zhao, X. Y.; Guo, S. Y.; Jiang, J.; Wang, G.; Weng, Y. B. | 2023 | International wound journal | 20 | 6 | 1979-1986 | 10.1111/iwj.14059 |
| Zhang 2015 | Early goal-directed therapy in the management of severe sepsis or septic shock in adults: a meta-analysis of | Zhang, L.; Zhu, G.; Han, L.; Fu, P. | 2015 | BMC medicine | 13 | 1 | 71 | 10.1186/s12916-015-0312-9 |

| | | | | | | | | |
|---|---|---|---|---|---|---|---|---|
| | randomized controlled trials | | | | | | | |
| Zhang 2024 | The therapeutic effect of continuous blood purification on sepsis in children: A systematic review and meta-analysis | Zhang, M.; Ling, Z.; Zhang, W.; Huang, Q. | 2024 | Journal of infection and chemotherapy : official journal of the Japan Society of Chemotherapy | 30 | 12 | 1201-1214 | 10.1016/j.jiac.2024.07.016 |
| Zhang 2022 | Use of dexmedetomidine in patients with sepsis: a systematic review and meta-analysis of randomized-controlled trials | Zhang, T.; Mei, Q.; Dai, S.; Liu, Y.; Zhu, H. | 2022 | Annals of intensive care | 12 | 1 | 81 | 10.1186/s13613-022-01052-2 |
| Zhang 2019 | Efficacy of dexmedetomidine for treatment of patients with sepsis: A meta-analysis of randomized controlled trials | Zhang, W. Q.; Xu, P.; Zhan, X. H.; Zheng, P.; Yang, W. | 2019 | Medicine | 98 | 18 | e15469 | 10.1097/MD.0000000000015469 |
| Zhang 2023 | Effect of Fluid Resuscitation Strategies for Obese Patients with Sepsis and Septic Shock: A Systematic Review | Zhang, Y.; Wang, M.; Lu, Z.; Yang, M. | 2023 | Intensive Care Res | 3 | 1 | 61-68 | 10.1007/s44231-022-00019-y |
| Zhang 2015 | Antipyretic Therapy in Critically Ill Patients with Established Sepsis: A Trial Sequential Analysis | Zhang, Z. | 2015 | PloS one | 10 | 2 | e0117279 | 10.1371/journal.pone.0117279 |
| Zhao 2024 | Prolonged versus intermittent β-lactam infusion in sepsis: a systematic review and meta-analysis of randomized controlled trials | Zhao, Y.; Zang, B.; Wang, Q. | 2024 | Annals of intensive care | 14 | 1 | 30 | 10.1186/s13613-024-01263-9 |

| | | | | | | | | |
|---|---|---|---|---|---|---|---|---|
| Zhong 2020 | Non-catecholamine vasopressors in the treatment of adult patients with septic shock-evidence from meta-analysis and trial sequential analysis of randomized clinical trials | Zhong, L.; Ji, X. W.; Wang, H. L.; Zhao, G. M.; Zhou, Q.; Xie, B. | 2020 | J Intensive Care | 8 | 1 | 83 | 10.1186/s40560-020-00500-0 |
| Zhu 2022 | The effect of intravenous vitamin C on clinical outcomes in patients with sepsis or septic shock: A meta-analysis of randomized controlled trials | Zhu, H.; Xu, X.; Zhang, K.; Ye, Q. | 2022 | Frontiers in nutrition | 9 | | 964484 | 10.3389/fnut.2022.964484 |
| Zhu 2019 | Terlipressin for septic shock patients: A meta-analysis of randomized controlled study | Zhu, Y.; Huang, H.; Xi, X.; Du, B. | 2019 | Journal of intensive care | 7 | 1 | 16 | 10.1186/s40560-019-0369-1 |
| Zou 2018 | Comparison of the effects of albumin and crystalloid on mortality among patients with septic shock: systematic review with meta-analysis and trial sequential analysis | Zou, Y.; Ma, K.; Xiong, J. B.; Xi, C. H.; Deng, X. J. | 2018 | Sao Paulo medical journal = Revista paulista de medicina | 136 | 5 | 421-432 | 10.1590/1516-3180.2017.0285281017 |

### Supplementary Table S3: References to included systematic reviews (n = 42)

| Study ID | Title | Authors | Published Year | Journal | Volume | Issue | Pages | DOI |
|---|---|---|---|---|---|---|---|---|
| Aletreby 2019 | Impact on Efficacy and Safety of Hydrocortisone in Sepsis and Septic Shock - A Systematic Literature Review and Meta-analysis | Aletreby, W. T.; Alharthy, A. M.; Madi, A. F.; Soliman, I. R.; Hamido, H. M.; Ramadan, O. E.; Alzayer, W.; Huwait, B. M.; Alodat, M. A.; Mumtaz, S. A.; Mahmood, N. N.; Al Kurdi, M. H.; Farrag, H. A.; Karakitsos, D. | 2019 | Archives of Iranian medicine | 22 | 7 | 394-402 | none |
| Allen 2019 | Recent Updates in the Pharmacological Management of Sepsis and Septic Shock: A Systematic Review Focused on Fluid Resuscitation, Vasopressors, and Corticosteroids | Allen, J. M.; Feild, C.; Shoulders, B. R.; Voils, S. A. | 2019 | The Annals of pharmacotherapy | 53 | 4 | 385-395 | 10.1177/1060028018812940 |
| Annane 2015 | Corticosteroids for treating sepsis | Annane, D.; Bellissant, E.; Bollaert, P. E.; Briegel, J.; Keh, D.; Kupfer, Y. | 2015 | Cochrane Database Syst Rev | 2015 | 12 | Cd002243 | 10.1002/14651858.CD002243.pub3 |
| Annane 2019 | Corticosteroids for treating sepsis in children and adults | Annane, D.; Bellissant, E.; Bollaert, P. E.; Briegel, J.; Keh, D.; Kupfer, Y.; Pirracchio, R.; Rochwerg, B. | 2019 | Cochrane Database Syst Rev | 12 | 12 | Cd002243 | 10.1002/14651858.CD002243.pub4 |
| Annane 2025 | Corticosteroids for treating sepsis in children and adults | Annane, D.; Briegel, J.; Granton, D.; Bellissant, E.; Bollaert, P. E.; Keh, D.; Kupfer, Y.; Pirracchio, R.; Rochwerg, B. | 2025 | The Cochrane database of systematic reviews | 6 | 6 | CD002243 | 10.1002/14651858.CD002243.pub5 |
| Barakakis 2019 | Combination of low-dose glucocorticosteroids and mineralocorticoids as adjunct therapy for adult patients with septic shock: A systematic review and meta-analysis of randomized trials and observational studies | Archontakis Barakakis, P.; Palaiodimos, L.; Fleitas Sosa, D.; Benes, L.; Gulani, P.; Fein, D. | 2019 | Avicenna journal of medicine | 9 | 4 | 134-142 | 10.4103/ajm.AJM_97_19 |

| | | | | | | | | |
|---|---|---|---|---|---|---|---|---|
| Assouline 2021 | Thiamine, Ascorbic Acid, and Hydrocortisone As a Metabolic Resuscitation Cocktail in Sepsis: A Meta-Analysis of Randomized Controlled Trials With Trial Sequential Analysis | Assouline, B.; Faivre, A.; Verissimo, T.; Sangla, F.; Berchtold, L.; Giraud, R.; Bendjelid, K.; Sgardello, S.; Elia, N.; Pugin, J.; de Seigneux, S.; Legouis, D. | 2021 | Critical care medicine | 49 | 12 | 2112-2120 | 10.1097/CCM.0000000000005262 |
| Deng 2025 | Corticosteroids for Managing Pediatric Sepsis and Septic Shock: A Systematic Review and Meta-analysis | Deng, J.; Bedri, N.; Zuo, Q. K.; Azab, M.; Chang, O.; Virdi, R.; Hung, J.; Venugopal, K.; Tahir, U.; Heybati, K. | 2025 | Pediatr Infect Dis J | 44 | 1 | 74-82 | 10.1097/inf.0000000000004551 |
| Deng 2024 | Efficacy and Safety of Hydrocortisone, Ascorbic Acid, and Thiamine Combination Therapy for the Management of Sepsis and Septic Shock: A Systematic Review and Meta-Analysis of Randomised Controlled Trials | Deng, J.; Zuo, Q. K.; Venugopal, K.; Hung, J.; Zubair, A.; Blais, S.; Porter, V.; Moskalyk, M.; Heybati, K. | 2024 | International archives of allergy and immunology | 185 | 10 | 1-21 | 10.1159/000538959 |
| Fang 2019 | Association of Corticosteroid Treatment With Outcomes in Adult Patients With Sepsis: A Systematic Review and Meta-analysis | Fang, F.; Zhang, Y.; Tang, J.; Lunsford, L. D.; Li, T.; Tang, R.; He, J.; Xu, P.; Faramand, A.; Xu, J.; You, C. | 2019 | JAMA internal medicine | 179 | 2 | 213-223 | 10.1001/jamainternmed.2018.5849 |
| Fratoni 2020 | Effect of Dexamethasone on Mortality in Adult and Elderly Patients with Sepsis: a Systematic Review | Fratoni, E.; dos Santos Nascimento, M. V. P.; Bramorski Mohr, E. T.; Monguilhott Dalmarco, E.; da Silva Fontoura, E.; de Moraes Trindade, E. B. S. | 2020 | SN Compr. Clin. Med. | 2 | 7 | 886-892 | 10.1007/s42399-020-00334-9 |
| Kato 2023 | Efficacy of Ascorbic Acid, Thiamine, and Hydrocortisone Combination Therapy: Meta-analysis of Randomized Controlled Trials | Kato, T.; Mizuno, T.; Nakanishi, M.; Lee, J. K.; Yamada, S.; Tsuboi, N.; Takahashi, K. | 2023 | In vivo (Athens, Greece) | 37 | 3 | 1236-1245 | 10.21873/invivo.13200 |
| Lee 2022 | Benefits of combination therapy of hydrocortisone, ascorbic acid and thiamine in sepsis and septic shock: A systematic review | Lee, Y. R.; Vo, K.; Varughese, J. T. | 2022 | Nutrition and health | 28 | 1 | 77-93 | 10.1177/02601060211018371 |
| Li 2025 | Influence of hydrocortisone infusion method on the clinical outcome of patients with septic shock: A | Li, Y.; Wang, Y.; Guo, J.; Zhang, D. | 2025 | J. Intensive Med. | 5 | 1 | 100-107 | 10.1016/j.jointm.2024.05.001 |

| | | | | | | | | |
|---|---|---|---|---|---|---|---|---|
| | systematic review and meta-analysis | | | | | | | |
| Lian 2019 | Reevaluating the Role of Corticosteroids in Septic Shock: An Updated Meta-Analysis of Randomized Controlled Trials | Lian, X. J.; Huang, D. Z.; Cao, Y. S.; Wei, Y. X.; Lian, Z. Z.; Qin, T. H.; He, P. C.; Liu, Y. H.; Wang, S. H. | 2019 | BioMed Research International | 2019 | | 1-14 | 10.1155/2019/3175047 |
| Liang 2021 | Corticosteroids for Treating Sepsis in Adult Patients: A Systematic Review and Meta-Analysis | Liang, H.; Song, H.; Zhai, R.; Song, G.; Li, H.; Ding, X.; Kan, Q.; Sun, T. | 2021 | Frontiers in immunology | 12 | | 709155 | 10.3389/fimmu.2021.709155 |
| | Corrigendum: Corticosteroids for Treating Sepsis in Adult Patients: A Systematic Review and Meta-Analysis | Liang, H.; Song, H.; Zhai, R.; Song, G.; Li, H.; Ding, X.; Kan, Q.; Sun, T. | 2021 | Front Immunol | 12 | | 771779 | 10.3389/fimmu.2021.771779 |
| Lin 2019 | Impact and beneficial critical points of clinical outcome in corticosteroid management of adult patients with sepsis: Meta-analysis and grade assessment | Lin, L. L.; Gu, H. Y.; Luo, J.; Wang, L.; Zhang, C.; Niu, Y. M.; Zuo, H. X. | 2019 | Frontiers in pharmacology | 10 | SEP | 1101 | 10.3389/fphar.2019.01101 |
| Lu 2023 | Hydrocortisone Combined with Vitamin C and Thiamine in the Treatment of Sepsis/Septic Shock: A Systematic Review with Meta-Analysis and Trial Sequential Analysis | Lu, D.; Mao, W. | 2023 | Clinical and investigative medicine. Medecine clinique et experimentale | 46 | 1 | E36-49 | 10.25011/cim.v46i1.40275 |
| Lu 2021 | Effi cacy and safety of corticosteroids in immunocompetent patients with septic shock | Lu, X.; Han, W.; Gao, Y. X.; Guo, S. G.; Yu, S. Y.; Yu, X. Z.; Zhu, H. D.; Li, Y. | 2021 | World journal of emergency medicine | 12 | 2 | 124-130 | 10.5847/WJEM.J.1920-8642.2021.02.007 |
| Lyu 2020 | Effect of Low-Dose Hydrocortisone Therapy in Adult Patients With Septic Shock: A Meta-Analysis With Trial Sequential Analysis of Randomized Controlled Trials | Lyu, Q. Q.; Chen, Q. H.; Zheng, R. Q.; Yu, J. Q.; Gu, X. H. | 2020 | Journal of intensive care medicine | 35 | 10 | 971-983 | 10.1177/0885066618803062 |
| Na 2021 | Hydrocortisone, ascorbic acid, and thiamine (HAT) for sepsis and septic shock: a meta-analysis with sequential trial analysis | Na, W.; Shen, H.; Li, Y.; Qu, D. | 2021 | Journal of intensive care | 9 | 1 | 75 | 10.1186/s40560-021-00589-x |

| | | | | | | | | |
|---|---|---|---|---|---|---|---|---|
| Ni 2019 | Can corticosteroids reduce the mortality of patients with severe sepsis? A systematic review and meta-analysis | Ni, Y. N.; Liu, Y. M.; Wang, Y. W.; Liang, B. M.; Liang, Z. A. | 2019 | The American journal of emergency medicine | 37 | 9 | 1657-1664 | 10.1016/j.ajem.2018.11.040 |
| Pitre 2024 | Corticosteroids in Sepsis and Septic Shock: A Systematic Review, Pairwise, and Dose-Response Meta-Analysis | Pitre, T.; Drover, K.; Chaudhuri, D.; Zeraaktkar, D.; Menon, K.; Gershengorn, H. B.; Jayaprakash, N.; Spencer-Segal, J. L.; Pastores, S. M.; Nei, A. M.; Annane, D.; Rochwerg, B. | 2024 | Critical care explorations | 6 | 1 | e1000 | 10.1097/CCE.0000000000001000 |
| Rochwerg 2018 | Corticosteroids in Sepsis: An Updated Systematic Review and Meta-Analysis | Rochwerg, B.; Oczkowski, S. J.; Siemieniuk, R. A. C.; Agoritsas, T.; Belley-Cote, E.; D'Aragon, F.; Duan, E.; English, S.; Gossack-Keenan, K.; Alghuroba, M.; Szczeklik, W.; Menon, K.; Alhazzani, W.; Sevransky, J.; Vandvik, P. O.; Annane, D.; Guyatt, G. | 2018 | Critical care medicine | 46 | 9 | 1411-1420 | 10.1097/CCM.0000000000003262 |
| Rygård 2018 | Low-dose corticosteroids for adult patients with septic shock: a systematic review with meta-analysis and trial sequential analysis | Rygård, S. L.; Butler, E.; Granholm, A.; Møller, M. H.; Cohen, J.; Finfer, S.; Perner, A.; Myburgh, J.; Venkatesh, B.; Delaney, A. | 2018 | Intensive care medicine | 44 | 7 | 1003-1016 | 10.1007/s00134-018-5197-6 |
| Saghafi 2024 | Therapeutic Role of HAT Therapy in Sepsis: A Systematic Review and Meta-Analysis | Saghafi, F.; Boostani Moghadam, Z.; Salehi-Abargouei, A.; Beigrezaei, S.; Sohrevardi, S. M.; Jamialahmadi, T.; Sahebnasagh, A.; Sahebkar, A. | 2024 | Current medicinal chemistry | | | | 10.2174/0109298673245464231121094448 |
| Siddiqui 2019 | Hydrocortisone Reduces 28-day Mortality in Septic Patients: A Systemic Review and Meta-analysis | Siddiqui, W. J.; Iyer, P.; Aftab, G.; Zafrullah, F.; Zain, M. A.; Jethwani, K.; Mazhar, R.; Abdulsalam, U.; Raza, A.; Hanif, M. O.; Sharma, E.; Aggarwal, S. | 2019 | Cureus | 11 | 6 | e4914 | 10.7759/cureus.4914 |

| | | | | | | | | |
|---|---|---|---|---|---|---|---|---|
| Somagutta 2021 | The Efficacy of vitamin C, thiamine, and corticosteroid therapy in adult sepsis patients: a systematic review and meta-analysis | Somagutta, M. K. R.; Pormento, M. K. L.; Khan, M. A.; Hamdan, A.; Hange, N.; Kc, M.; Pagad, S.; Jain, M. S.; Lingarajah, S.; Sharma, V.; Kaur, J.; Emuze, B.; Batti, E.; Iloeje, O. J. | 2021 | Acute and critical care | 36 | 3 | 185-200 | 10.4266/acc.2021.00108 |
| Son 2021 | New evidence of potential benefits of dexamethasone and added on therapy of fludrocortisone on clinical outcomes of corticosteroid in sepsis patients: A systematic review and meta-analysis | Son, J. Y.; Shin, S.; Choi, Y. J. | 2021 | Journal of personalized medicine | 11 | 6 | | 10.3390/jpm11060544 |
| Song 2024 | Early use of low-dose hydrocortisone can reduce in-hospital mortality in patients with septic shock: A systematic review and meta-analysis | Song, B.; Zheng, X.; Fu, K.; Liu, C. | 2024 | Medicine | 103 | 48 | e40635 | 10.1097/MD.0000000000040635 |
| Song 2024 | Role of corticosteroids in the treatment of critically ill sepsis patients: a meta-analysis review | Song, Y.; Hao, J.; Liu, Y. | 2024 | Inflammopharmacology | | | | 10.1007/s10787-023-01426-3 |
| Sudaryadi 2023 | Comparison of Hydrocortisone with Combined Hydrocortisone, Ascorbic Acid, and Thiamine as an Adjuvant Therapy on Septic Shock Patients on Mortality: A Systematic Review and Meta-analysis | Sudaryadi, S.; Harijanto, E.; Mulyono, I. | 2023 | Open Access Macedonian Journal of Medical Sciences | 11 | | 171-179 | 10.3889/oamjms.2023.9952 |
| Wen 2019 | The Effectiveness and Safety of Corticosteroids Therapy in Adult Critical Ill Patients With Septic Shock: A Meta-Analysis of Randomized Controlled Trials | Wen, Y.; Zhu, Y.; Jiang, Q.; Guo, N.; Cai, Y.; Shen, X. | 2019 | Shock | 52 | 2 | 198-207 | 10.1097/shk.0000000000001202 |
| Wu 2020 | Effects and safety of separate low-dose hydrocortisone use in patients with septic shock: A meta-analysis | Wu, J.; Wang, Q.; Ma, Y.; Jiang, L.; Huang, M. | 2020 | Hong Kong J. Emerg. Med. | 27 | 1 | 39-50 | 10.1177/1024907919833205 |
| Wu 2024 | Effect of hydrocortisone-fludrocortisone combination on mortality in septic shock: a | Wu, J. Y.; Liu, M. Y.; Liu, T. H.; Huang, P. Y.; Restinia, M.; Hsu, W. H.; Tsai, Y. W.; | 2024 | Infection | 53 | 2 | 553-560 | 10.1007/s15010-024-02381-z |

| | | | | | | | | |
|---|---|---|---|---|---|---|---|---|
| | systematic review and meta-analysis | Chuang, M. H.; Hung, K. C.; Lai, C. C. | | | | | | |
| Wu 2021 | Effect of Combined Hydrocortisone, Ascorbic Acid and Thiamine for Patients with Sepsis and Septic Shock: A Systematic Review and Meta-Analysis | Wu, T.; Hu, C.; Huang, W.; Xu, Q.; Hu, B.; Li, J. | 2021 | Shock (Augusta, Ga.) | 56 | 6 | 880-889 | 10.1097/SHK.0000000000001781 |
| Xu 2018 | Do low-dose corticosteroids improve survival or shock reversal from septic shock in adults? Meta-analysis with trial sequential analysis | Xu, R.; Wang, Q.; Huang, Y.; Wu, L.; Liu, Q.; Hu, W.; Zhou, C.; Du, Q. | 2018 | The Journal of international medical research | 46 | 7 | 2513-2524 | 10.1177/0300060518774985 |
| Yamamoto 2020 | Hydrocortisone with fludrocortisone for septic shock: a systematic review and meta-analysis | Yamamoto, R.; Nahara, I.; Toyosaki, M.; Fukuda, T.; Masuda, Y.; Fujishima, S. | 2020 | Acute medicine & surgery | 7 | 1 | e563 | 10.1002/ams2.563 |
| Yang 2020 | Controversies in the application of corticosteroids for pediatric septic shock treatment: a preferred reporting items for systematic reviews and meta-analysis-compliant updated meta-analysis | Yang, J.; Sun, S. | 2020 | Medicine | 99 | 30 | e20762 | 10.1097/MD.0000000000020762 |
| Yao 2021 | Combination therapy of thiamine, vitamin C and hydrocortisone in treating patients with sepsis and septic shock: a meta-analysis and trial sequential analysis | Yao, R.; Zhu, Y.; Yu, Y.; Li, Z.; Wang, L.; Zheng, L.; Li, J.; Huang, H.; Wu, G.; Zhu, F.; Xia, Z.; Ren, C.; Yao, Y. | 2021 | Burns & trauma | 9 | | tkab040 | 10.1093/burnst/tkab040 |
| Zayed 2022 | Vitamin C, Thiamine, and Hydrocortisone in the Treatment of Sepsis: A Meta-Analysis and Trial Sequential Analysis of Randomized Controlled Trials | Zayed, Y.; Alzghoul, B. N.; Banifadel, M.; Venigandla, H.; Hyde, R.; Sutchu, S.; Khasawneh, M.; Borok, Z.; Urbine, D.; Jantz, M.; Reddy, R. | 2022 | Journal of intensive care medicine | 37 | 3 | 327-336 | 10.1177/0885066620987809 |
| Zhou 2018 | Effect of adjunctive corticosteroids on clinical outcomes in adult patients with septic shock - a meta-analysis of randomized controlled trials and trial sequential analysis | Zhou, X.; Hu, C.; Yao, L.; Fan, Z.; Sun, L.; Wang, Y.; Xu, Z. | 2018 | Journal of critical care | 48 | | 296-306 | 10.1016/j.jcrc.2018.09.013 |

**Supplementary Table S4: Characteristics of included systematic reviews (n = 42)**

| SR ID | Date of publication | Cochrane Review | Country (according to the first author of the study) | Protocol published on/registered at | Protocol registration number OR DOI | Study design(s) included | Search date | Number of databases searched | Number of registers searched | Number of included RCTs | Type of analysis method |
|---|---|---|---|---|---|---|---|---|---|---|---|
| Aletreby 2019 | 01.07.2019 | Non-Cochrane review | Asia | PROSPERO | CRD42018100112 | RCTs | n.r. | 2 | 0 | 10 | With meta-analysis |
| Allen 2019 | 08.11.2018 | Non-Cochrane review | North America | no protocol available | NA | RCTs | 31/08/2018 | 1 | 1 | 7 | Without meta-analysis |
| Annane 2015 | 03.12.2015 | Cochrane review | Europe | Cochrane | 10.1002/14651858.CD002243/full | RCTs | Okt 14 | 4 | 1 | 33 | With meta-analysis |
| Annane 2019 | 06.12.2019 | Cochrane review | Europe | Cochrane | 10.1002/14651858.CD002243/full | RCTs | 25/07/2019 | 4 | 3 | 61 | With meta-analysis |
| Annane 2025 | 05.06.2025 | Cochrane review | Europe | Cochrane | 10.1002/14651858.CD002243/full | RCTs | 31/12/2023 | 4 | 3 | 87 | With meta-analysis |
| Assouline 2021 | 01.12.2021 | Non-Cochrane review | Europe | PROSPERO | CRD4202022013 | RCTs | 30/03/2021 | 3 | 0 | 8 | With meta-analysis |
| Barakakis 2019 | 03.10.2019 | Non-Cochrane review | North America | no protocol available | NA | RCTs; NRSIs | n.r. | 3 | 0 | 2 | With meta-analysis |
| Deng 2024 | 13.06.2024 | Non-Cochrane review | North America | PROSPERO | CRD42023459882 | RCTs | 19/08/2023 | 5 | 2 | 15 | With meta-analysis |
| Deng 2025 | 01.01.2025 | Non-Cochrane review | North America | PROSPERO | CRD42023487819 | RCTs | 03/09/2023 | 5 | 2 | 15 | With meta-analysis |
| Fang 2019 | 21.12.2018 | Non-Cochrane review | Asia | PROSPERO | CRD42018095867 | RCTs | 10/08/2018 | 3 | 1 | 37 | With meta-analysis |

| | | | | | | | | | | | |
|---|---|---|---|---|---|---|---|---|---|---|---|
| Fratoni 2020 | 19.06.2020 | Non-Cochrane review | South America | PROSPERO | CRD42018088150 | RCTs | 01/11/2019 | 4 | none | 5 | Without meta-analysis |
| Kato 2023 | 01.05.2023 | Non-Cochrane review | Asia | no protocol available | NA | RCTs | 05/2022 | 5 | 0 | 9 | With meta-analysis |
| Lee 2022 | 26.05.2021 | Non-Cochrane review | North America | no protocol available | NA | RCTs; NRSIs | 04/2021 | 5 | 1 | 6 | Without meta-analysis |
| Li 2025 | 25.06.2024 | Non-Cochrane review | Asia | PROSPERO | CRD42023392160 | RCTs; NRSIs | 01/01/2023 | 3 | 0 | 3 | With meta-analysis |
| Lian 2019 | 10.06.2019 | Non-Cochrane review | Asia | PROSPERO | CRD4201809253 | RCTs | 04/2018 | 4 | 1 | 21 | With meta-analysis |
| Liang 2021 | 16.08.2021 | Non-Cochrane review | Asia | INPLASY | INPLASY2020110122 | RCTs | 05/07/2021 | 3 | 0 | 50 | With meta-analysis |
| Lin 2019 | 24.09.2019 | Non-Cochrane review | Asia | PROSPERO | CRD42018092849 | RCTs | 22/03/2019 | 4 | 0 | 30 | With meta-analysis |
| Lu 2021 | 01.04.2021 | Non-Cochrane review | Asia | no protocol available | NA | RCTs | 12/03/2020 | 3 | 0 | 9 | With meta-analysis |
| Lu 2023 | 26.03.2023 | Non-Cochrane review | Asia | no protocol available | NA | RCTs | 31/10/2022 | 3 | 0 | 8 | With meta-analysis |
| Lyu 2020 | 01.10.2018 | Non-Cochrane review | Asia | no protocol available | NA | RCTs | 07/03/2018 | 4 | 1 | 13 | With meta-analysis |
| Na 2021 | 18.12.2021 | Non-Cochrane review | Asia | PROSPERO | CRD42020203166 | RCTs | 14/08/2021 | 3 | 0 | 10 | With meta-analysis |
| Ni 2019 | 26.11.2018 | Non-Cochrane review | Asia | Protocol in supplement | NA | RCTs | 03/2018 | 4 | 0 | 19 | With meta-analysis |

| | | | | | | | | | | | |
|---|---|---|---|---|---|---|---|---|---|---|---|
| Pitre 2024 | 19.01.2024 | Non-Cochrane review | North America | OSF | https://osf.io/v5qrz | RCTs | 01/01/2023 | 4 | 0 | 45 | With meta-analysis |
| Rochwerg 2018 | 01.09.2018 | Non-Cochrane review | North America | PROSPERO | CRD42017058537 | RCTs | 10/01/2018 | 4 | 0 | 42 | With meta-analysis |
| Rygard 2018 | 14.05.2018 | Non-Cochrane review | Europe | PROSPERO | CRD42017084037 | RCTs | 03/03/2018 | 3 | 4 | 23 | With meta-analysis |
| Saghafi 2024 | 22.01.2024 | Non-Cochrane review | Asia | PROSPERO | CRD42020170523 | RCTs; Other: cohort studies | 06/03/2021 | 5 | 0 | 11 | With meta-analysis |
| Siddiqui 2019 | 17.06.2019 | Non-Cochrane review | North America | no protocol available | NA | RCTs | 01/03/2018 | 2 | 0 | 5 | With meta-analysis |
| Somagutta 2021 | 30.06.2021 | Non-Cochrane review | North America | PROSPERO | CRD42020209086 | RCTs; NRSIs | 10/2020 | 8 | 1 | 8 | With meta-analysis |
| Son 2021 | 11.06.2021 | Non-Cochrane review | Asia | no protocol available | NA | RCTs | 10/2020 | 3 | 0 | 22 | With meta-analysis |
| Song B 2024 | 29.11.2024 | Non-Cochrane review | Asia | INPLASY | INPLASY202480070 | RCTs; NRSIs | 01/08/2024 | 3 | 0 | 0 | With meta-analysis |
| Song Y 2024 | 12.02.2024 | Non-Cochrane review | Asia | no protocol available | NA | RCTs; NRSIs | n.r. | 4 | 0 | 9 | With meta-analysis |
| Sudaryadi 2023 | 02.01.2023 | Non-Cochrane review | Asia | PROSPERO | CRD42022296055 | RCTs; NRSIs | 12/2021 | 4 | 0 | 3 | With meta-analysis |
| Wen 2019 | 01.08.2019 | Non-Cochrane review | Asia | no protocol available | NA | RCTs | 24/03/2018 | 4 | 0 | 18 | With meta-analysis |
| Wu 2020 | 15.04.2019 | Non-Cochrane review | Asia | PROSPERO | CRD42018090792 | RCTs | 01/08/2018 | 3 | 3 | 12 | With meta-analysis |

| | | | | | | | | | | | |
|---|---|---|---|---|---|---|---|---|---|---|---|
| Wu 2021 | 01.12.2021 | Non-Cochrane review | Asia | PROSPERO | CRD42020170648 | RCTs; NRSIs | 11/11/2020 | 3 | 0 | 6 | With meta-analysis |
| Wu 2024 | 27.08.2024 | Non-Cochrane review | Asia | PROSPERO | CRD42023472376 | RCTs; NRSIs | 01/10/2023 | 3 | 1 | 3 | With meta-analysis |
| Xu 2018 | 18.06.2018 | Non-Cochrane review | Asia | PROSPERO | CRD42017077531 | RCTs | 23/10/2017 | 4 | 0 | 9 | With meta-analysis |
| Yamamoto 2020 | 01.09.2020 | Non-Cochrane review | Asia | PROSPERO | CRD42019139069 | RCTs | 04/2019 | 3 | 0 | 3 | With meta-analysis |
| Yang 2020 | 24.07.2020 | Non-Cochrane review | Asia | no protocol available | NA | RCTs | 08/06/2019 | 3 | 0 | 10 | With meta-analysis |
| Yao 2021 | 06.12.2021 | Non-Cochrane review | Asia | PROSPERO | CRD42020206950 | RCTs | 01/03/2021 | 3 | n.r. | 8 | With meta-analysis |
| Zayed 2022 | 29.01.2021 | Non-Cochrane review | North America | no protocol available | NA | RCTs | n.r. | 2 | 1 | 7 | With meta-analysis |
| Zhou 2018 | 01.12.2018 | Non-Cochrane review | Asia | no protocol available | NA | RCTs | 12/03/2018 | 4 | 0 | 17 | With meta-analysis |

### Supplementary Table S5: PRISMA 2020 reporting quality of systematic reviews (n = 42)

PRISMA 2020 items were operationalized and rated as “yes”, “incomplete”, “no”, or not applicable (i.e. no meta-analysis performed, certainty not assessed), and frequency of fulfillment was counted in PRISMA overall rating categories resulting in very poor (up to 25% of items), poor (25-50%), moderate (50-75%), and high (75-100%) quality of reporting.

#### Assessment of all PRISMA items per systematic review:

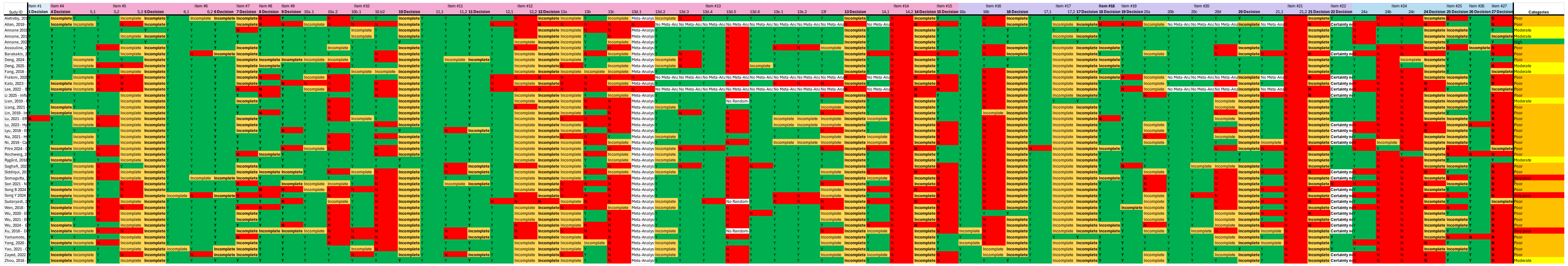

#### PRISMA overall rating category per systematic review:

| Sudy ID | PRISMA overall rating categories |
|---|---|
| Aletreby, 2019 - Impact on Efficacy and Safety of Hydrocortisone in Sepsis and Septic Shock – A Systematic Literature Review and Meta-analysis | Poor |
| Allen, 2019 - Recent Updates in the Pharmacological Management of Sepsis and Septic Shock: A Systematic Review Focused on Fluid Resuscitation, Vasopressors, and Corticosteroids | Poor |
| Annane 2019 - Corticosteroids for treating sepsis in children and adults (Review) | Moderate |
| Annane, 2015 - Corticosteroids for treating sepsis | Moderate |
| Annane, 2025 - Corticosteroids for treating sepsis in children and adults (Review) | High |
| Assouline, 2021 - Thiamine, Ascorbic Acid, and Hydrocortisone As a Metabolic Resuscitation Cocktail in Sepsis: A Meta-Analysis of Randomized Controlled Trials With Trial Sequential Analysis | Poor |
| Barakakis, 2019 - Combination of low-dose glucocorticosteroids and mineralocorticoids as adjunct therapy for adult patients with septic shock: a systematic review and meta-analysis of randomized trials and observational studies | Poor |
| Deng, 2024 - Efficacy and Safety of Hydrocortisone, Ascorbic Acid, and Thiamine Combination Therapy for the Management of Sepsis and Septic Shock: A Systematic Review and Meta-Analysis of Randomised Controlled Trials | Poor |

| | |
|---|---|
| Deng, 2025 - Corticosteroids for Managing Pediatric Sepsis and Septic Shock - A Systematic Review and Meta-analysis | Moderate |
| Fang, 2018 - Association of Corticosteroid Treatment With Outcomes in Adult Patients With Sepsis A Systematic Review and Meta-analysis | Moderate |
| Fratoni, 2020 - Effect of Dexamethasone on Mortality in Adult and Elderly Patients with Sepsis: a Systematic Review | Poor |
| Kato, 2023 - Efficacy of Ascorbic Acid, Thiamine, and Hydrocortisone Combination Therapy: Meta-analysis of Randomized Controlled Trials | Poor |
| Lee, 2022 - Benefits of combination therapy of hydrocortisone, ascorbic acid and thiamine in sepsis and septic shock: A systematic review | Poor |
| Li 2025 - Influence of hydrocortisone infusion method on the clinical outcome of patients with septic shock: A systematic review and meta-analysis | Poor |
| Lian, 2019 - Reevaluating the Role of Corticosteroids in Septic Shock: An Updated Meta-Analysis of Randomized Controlled Trials | Moderate |
| Liang, 2021 - Corticosteroids for Treating Sepsis in Adult Patients: A Systematic Review and Meta-Analysis | Poor |
| Lin, 2019 - Impact and Beneficial Critical Points of Clinical Outcome in Corticosteroid Management of Adult Patients With Sepsis: Meta-Analysis and GRADE Assessment | Poor |
| Lu, 2021 - Efficacy and safety of corticosteroids in immunocompetent patients with septic shock | Poor |
| Lu, 2023 - Hydrocortisone combined with Vitamin C and Thiamine in the Treatment of Sepsis/ Septic Shock: a Systematic Review with Meta-Analysis and Trial Sequential Analysis | Poor |
| Lyu, 2018 - Effect of Low-Dose Hydrocortisone Therapy in Adult Patients With Septic Shock: A Meta-Analysis With Trial Sequential Analysis of Randomized Controlled Trials | Poor |
| Na, 2021 - Hydrocortisone, ascorbic acid, and thiamine (HAT) for sepsis and septic shock: a meta-analysis with sequential trial analysis | Poor |
| Ni, 2019 - Can corticosteroids reduce the mortality of patients with severe sepsis? A systematic review and meta-analysis | Poor |
| Pitre 2024 - Corticosteroids in Sepsis and Septic Shock: A Systematic Review, Pairwise, and Dose-Response Meta-Analysis | Poor |
| Rochwerg, 2018 - Corticosteroids in Sepsis: An Updated Systematic Review and Meta-Analysis | Poor |
| Rygård, 2018 - Low-dose corticosteroids for adult patients with septic shock: a systematic review with meta-analysis and trial sequential analysis | Moderate |
| Saghafi, 2023 - Therapeutic Role of HAT Therapy in Sepsis: A Systematic Review and Meta-Analysis | Poor |
| Siddiqui, 2019 - Hydrocortisone Reduces 28-day Mortality in Septic Patients: A Systemic Review and Meta-analysis | Poor |
| Somagutta, 2021 - The efficacy of vitamin C, thiamine, and corticosteroid therapy in adult sepsis patients: a systematic review and meta-analysis | Very poor |
| Son 2021 - New Evidence of Potential Benefits of Dexamethasone and Added on Therapy of Fludrocortisone on Clinical Outcomes of Corticosteroid in Sepsis Patients: A Systematic Review and Meta-Analysis | Poor |

| | |
|---|---|
| Song B 2024 - Early use of low-dose hydrocortisone can reduce in-hospital mortality in patients with septic shock A systematic review and meta-analysis | Poor |
| Song Y 2024 - Role of corticosteroids in the treatment of critically ill sepsis patients: a meta-analysis review | Very poor |
| Sudaryadi, 2023 - Comparison of Hydrocortisone with Combined Hydrocortisone, Ascorbic Acid, and Thiamine as an Adjuvant Therapy on Septic Shock Patients on Mortality: A Systematic Review and Meta-analysis | Poor |
| Wen, 2018 - The Effectiveness and Safety of Corticosteroids therapy in adult critical ill patients with Septic Shock: a meta-analysis of randomized controlled trials | Poor |
| Wu, 2020 - Effects and safety of separate low-dose hydrocortisone use in patients with septic shock: A meta-analysis | Poor |
| Wu, 2021 - Effect of combined hydrocortisone, ascorbic acid and thiamine for patients with sepsis and septic shock: a systematic review and meta-analysis | Poor |
| Wu, 2024 - Effect of hydrocortisone-fludrocortisone combination on mortality in septic shock: a systematic review and meta-analysis | Poor |
| Xu, 2018 - Do low-dose corticosteroids improve survival or shock reversal from septic shock in adults? Meta-analysis with trial sequential analysis | Very poor |
| Yamamoto, 2020 - Hydrocortisone with fludrocortisone for septic shock: a systematic review and meta-analysis | Poor |
| Yang, 2020 - Controversies in the application of corticosteroids for pediatric septic shock treatment: a preferred reporting items for systematic reviews and meta-analysis-compliant updated meta-analysis | Poor |
| Yao, 2021 - Combination therapy of thiamine, vitamin C and hydrocortisone in treating patients with sepsis and septic shock: a meta-analysis and trial sequential analysis | Poor |
| Zayed, 2022 - Vitamin C, Thiamine, and Hydrocortisone in the Treatment of Sepsis: A Meta-Analysis and Trial Sequential Analysis of Randomized Controlled Trials | Poor |
| Zhou, 2018 - Effect of adjunctive corticosteroids on clinical outcomes in adult patients with septic shock- a meta-analysis of randomized controlled trials and trial sequential analysis | Moderate |

### Supplementary Table S6: AMSTAR 2 overall confidence of systematic reviews (n = 42)

AMSTAR 2 was applied according to original guidance (with further specification reported in the protocol) using critical and non-critical domains, with overall confidence categorized as high, moderate, low, or critically low. Deviating from the protocol, assessments were performed by one reviewer and independently verified by a second reviewer due to time constraints.

#### Assessment of all AMSTAR 2 items per systematic review:

| | | | | | AMSTAR 2 - Summary | | | | | | | | | | | | | | | | | | |
|---|---|---|---|---|---|---|---|---|---|---|---|---|---|---|---|---|---|---|---|---|---|---|---|
| | | | | | METHODS | | | | | | | | | | RESULTS | | | | | | OTHER INFORMATION | | |
| | | | | | Review question/ Eligibilit | | Protocol/ re | Search strat | Selection pr | Data extract | Risk of bias | Data synthesis | | Reporting bi | Study select | Study characteristics | | Presentation/Interpretation/Discussi | | | Conflict of interest, fund | | Overall confi |
| | | | | | Item #1 | Item #3 | Item #2 | Item #4 | Item #5 | Item #6 | Item #9 | Item #11 | Item #12 | Item #15.1 | Item #7 | Item #8 | Item #10 | Item #13 | Item #14 | Item #15.2 | Item #16 | Item #15 (#15.1 +#15.2) | |
| Study ID | Assessor | Assessed date | Revisor | Revised date | | | | | | | | | | | | | | | | | | | |
| Aletreby, 201 | HD | 12.06.2025 | TP | 04.08.2025 | Y | N | PY | N | N | Y | Y | N | Y | N | N | PY | N | Y | Y | Y | Y | N | Critically low |
| Allen, 2019 - | HD | 28.08.2025 | TP | 24.09.2025 | N | N | N | N | N | N | Y | No Meta-Ana | N | No Meta-Ana | N | N | N | N | N | No Meta-Ana | Y | Y | Critically low |
| Annane 2019 | HD | 06.06.2025 | TP | 11.08.2025 | Y | N | PY | Y | Y | Y | Y | N | Y | Y | Y | PY | Y | Y | Y | Y | Y | Y | Low |
| Annane, 201 | HD | 26.08.2025 | TP | 01.10.2025 | Y | N | PY | Y | Y | Y | Y | N | Y | Y | Y | PY | N | Y | Y | Y | Y | Y | Low |
| Annane, 202 | HD | 12.06.2025 | TP | 04.08.2025 | Y | Y | PY | Y | Y | Y | Y | N | Y | Y | Y | PY | Y | Y | Y | Y | Y | Y | Low |
| Assouline, 2( | HD | 09.10.2025 | TP | 25.07.2025 | Y | Y | PY | PY | Y | Y | Y | N | N | N | N | N | N | Y | Y | Y | N | N | Critically low |
| Barakakis, 2 | HD | 13.06.2025 | TP | 01.08.2025 | Y | Y | N | PY | Y | Y | Y | N | Y | N | N | N | N | Y | Y | N | Y | N | Critically low |
| Deng, 2024 - | HD | 18.06.2025 | TP | 24.07.2025 | Y | Y | PY | PY | N | Y | Y | N | Y | Y | Y | N | N | Y | Y | Y | Y | Y | Low |
| Deng, 2025 - | HD | 20.08.2025 | TP | 24.09.2025 | N | Y | PY | PY | Y | Y | Y | N | Y | N | Y | N | N | Y | Y | Y | Y | N | Critically low |
| Fang, 2018 - | HD | 13.06.2025 | TP | 01.08.2025 | Y | N | PY | PY | Y | Y | Y | N | Y | Y | N | N | N | Y | Y | Y | Y | Y | Critically low |
| Fratoni, 2020 | HD | 20.08.2025 | TP | 25.09.2025 | N | N | PY | N | Y | Y | Y | No Meta-Ana | N | No Meta-Ana | N | N | N | Y | N | No Meta-Ana | Y | Y | Critically low |
| Kato, 2023 - I | HD | 21.08.2025 | TP | 25.09.2025 | N | N | N | N | Y | N | Y | N | Y | Y | N | N | N | Y | Y | N | Y | N | Critically low |
| Lee, 2022 - B | HD | 18.06.2025 | TP | 23.07.2025 | N | N | N | N | N | Y | N | No Meta-Ana | N | No Meta-Ana | N | N | N | N | N | No Meta-Ana | Y | Y | Critically low |
| Li 2025 - Influ | HD | 10.06.2025 | TP | 07.08.2025 | Y | N | N | N | N | Y | Y | N | N | Y | N | N | N | N | Y | N | Y | N | Critically low |
| Lian, 2019 - F | HD | 13.06.2025 | TP | 31.07.2025 | Y | Y | PY | PY | Y | Y | Y | N | N | Y | N | N | N | Y | Y | Y | Y | Y | Critically low |
| Liang, 2021 - | HD | 21.08.2025 | TP | 26.09.2025 | Y | N | N | N | Y | Y | Y | N | N | Y | N | N | N | N | Y | Y | Y | Y | Critically low |
| Lin, 2019 - Im | HD | 26.08.2025 | TP | 26.09.2025 | Y | N | PY | N | N | Y | Y | N | N | Y | Y | N | N | N | Y | Y | Y | Y | Critically low |
| Lu, 2021 - Effi | HD | 19.08.2025 | TP | 26.09.2025 | Y | N | N | PY | Y | Y | Y | N | Y | Y | N | N | N | Y | Y | Y | Y | Y | Critically low |
| Lu, 2023 - Hy | HD | 09.10.2025 | TP | 22.07.2025 | N | N | N | PY | Y | Y | Y | N | Y | N | N | PY | N | Y | Y | N | N | N | Critically low |
| Lyu, 2018 - Ef | HD | 16.06.2025 | TP | 31.07.2025 | Y | Y | N | N | Y | N | Y | N | N | Y | N | N | N | N | Y | Y | Y | Y | Critically low |
| Na, 2021 - Hy | HD | 09.10.2025 | TP | 17.07.2025 | Y | N | N | PY | Y | Y | Y | N | N | N | N | PY | N | N | Y | N | Y | N | Critically low |
| Ni, 2019 - Ca | HD | 19.08.2025 | TP | 01.10.2025 | Y | N | PY | PY | Y | Y | Y | N | N | Y | N | N | N | N | Y | N | Y | N | Critically low |
| Pitre 2024 - C | HD | 05.05.2025 | TP | 11.08.2025 | Y | Y | PY | PY | Y | N | Y | N | N | N | N | N | N | Y | Y | Y | Y | N | Critically low |
| Rochwerg, 2( | HD | 11.06.2025 | TP | 04.08.2025 | Y | Y | Y | PY | N | Y | Y | N | Y | Y | N | N | N | Y | Y | Y | N | Y | Critically low |
| Rygård, 2018 | HD | 16.06.2025 | TP | 30.07.2025 | Y | Y | PY | PY | Y | Y | Y | N | Y | Y | N | N | N | Y | Y | Y | Y | Y | Critically low |
| Saghafi, 2023 | HD | 24.06.2025 | TP | 15.07.2025 | Y | Y | PY | PY | Y | Y | PY | N | N | N | N | N | N | Y | Y | Y | Y | N | Critically low |
| Siddiqui, 201 | HD | 21.08.2025 | TP | 29.09.2025 | Y | Y | N | N | N | N | Y | N | N | Y | N | N | N | N | Y | N | Y | N | Critically low |
| Somagutta, 2 | HD | 12.10.2025 | TP | 14.07.2025 | Y | N | PY | N | Y | Y | PY | N | N | N | N | PY | N | N | Y | N | Y | N | Critically low |
| Son 2021 - N | HD | 06.05.2025 | TP | 11.08.2025 | N | Y | N | N | Y | N | Y | N | N | Y | N | N | N | N | Y | N | Y | N | Critically low |
| Song B 2024 - | HD | 10.06.2025 | TP | 07.08.2025 | N | N | PY | N | N | N | PY | N | Y | N | N | N | N | Y | Y | N | Y | N | Critically low |
| Song Y 2024 - | HD | 06.06.2025 | TP | 07.08.2025 | N | N | N | PY | N | Y | Y | N | N | N | N | N | N | N | Y | N | Y | N | Critically low |
| Sudaryadi, 2 | HD | 22.08.2025 | TP | 29.06.2025 | Y | N | PY | N | Y | N | PY | N | Y | Y | N | N | N | Y | Y | N | Y | N | Critically low |
| Wen, 2018 - 1 | HD | 16.06.2025 | TP | 29.07.2025 | N | Y | N | PY | N | Y | Y | N | N | Y | N | N | N | N | Y | N | Y | N | Critically low |
| Wu, 2020 - Ef | HD | 20.08.2025 | TP | 29.09.2025 | Y | Y | PY | PY | Y | Y | Y | N | N | N | Y | N | N | N | Y | N | Y | N | Critically low |
| Wu, 2021 - Ef | HD | 22.08.2025 | TP | 30.09.2025 | Y | N | PY | N | Y | Y | PY | N | N | Y | N | N | N | N | Y | Y | Y | Y | Critically low |
| Wu, 2024 - E | HD | 09.10.2025 | TP | 29.07.2025 | Y | N | PY | N | Y | Y | Y | N | N | Y | N | N | N | N | Y | N | Y | N | Critically low |
| Xu, 2018 - D | HD | 17.06.2025 | TP | 28.07.2025 | N | N | PY | PY | Y | Y | Y | N | N | N | N | N | N | N | Y | N | Y | N | Critically low |
| Yamamoto, 2 | HD | 11.06.2025 | TP | 05.08.2025 | Y | N | PY | N | Y | Y | Y | N | Y | Y | N | N | N | Y | Y | Y | N | Y | Critically low |
| Yang, 2020 - | HD | 25.08.2025 | TP | 30.09.2025 | Y | Y | N | PY | Y | Y | Y | N | Y | Y | N | N | N | Y | Y | Y | Y | Y | Critically low |
| Yao, 2021 - C | HD | 26.08.2025 | TP | 30.09.2025 | Y | N | PY | PY | Y | Y | Y | N | Y | N | N | N | N | Y | Y | Y | Y | N | Critically low |
| Zayed, 2022 - | HD | 25.08.2025 | TP | 01.10.2025 | N | Y | N | N | Y | Y | Y | N | N | N | N | N | N | N | Y | Y | Y | N | Critically low |
| Zhou, 2018 - | HD | 25.08.2025 | TP | 01.10.2025 | Y | Y | N | PY | Y | Y | Y | N | Y | Y | Y | N | N | Y | Y | Y | Y | Y | Critically low |

**AMSTAR 2 overall confidence in the results of the systematic review:**

| Study ID | AMSTAR 2 overall confidence in results |
|---|---|
| Aletreby, 2019 - Impact on Efficacy and Safety of Hydrocortisone in Sepsis and Septic Shock – A Systematic Literature Review and Meta-analysis | Critically low |
| Allen, 2019 - Recent Updates in the Pharmacological Management of Sepsis and Septic Shock: A Systematic Review Focused on Fluid Resuscitation, Vasopressors, and Corticosteroids | Critically low |
| Annane 2019 - Corticosteroids for treating sepsis in children and adults (Review) | Low |
| Annane, 2015 - Corticosteroids for treating sepsis | Low |
| Annane, 2025 - Corticosteroids for treating sepsis in children and adults (Review) | Low |
| Assouline, 2021 - Thiamine, Ascorbic Acid, and Hydrocortisone As a Metabolic Resuscitation Cocktail in Sepsis: A Meta-Analysis of Randomized Controlled Trials With Trial Sequential Analysis | Critically low |
| Barakakis, 2019 - Combination of low-dose glucocorticosteroids and mineralocorticoids as adjunct therapy for adult patients with septic shock: a systematic review and meta-analysis of randomized trials and observational studies | Critically low |
| Deng, 2024 - Efficacy and Safety of Hydrocortisone, Ascorbic Acid, and Thiamine Combination Therapy for the Management of Sepsis and Septic Shock: A Systematic Review and Meta-Analysis of Randomised Controlled Trials | Low |
| Deng, 2025 - Corticosteroids for Managing Pediatric Sepsis and Septic Shock - A Systematic Review and Meta-analysis | Critically low |
| Fang, 2018 - Association of Corticosteroid Treatment With Outcomes in Adult Patients With Sepsis A Systematic Review and Meta-analysis | Critically low |
| Fratoni, 2020 - Effect of Dexamethasone on Mortality in Adult and Elderly Patients with Sepsis: a Systematic Review | Critically low |
| Kato, 2023 - Efficacy of Ascorbic Acid, Thiamine, and Hydrocortisone Combination Therapy: Meta-analysis of Randomized Controlled Trials | Critically low |
| Lee, 2022 - Benefits of combination therapy of hydrocortisone, ascorbic acid and thiamine in sepsis and septic shock: A systematic review | Critically low |
| Li 2025 - Influence of hydrocortisone infusion method on the clinical outcome of patients with septic shock: A systematic review and meta-analysis | Critically low |
| Lian, 2019 - Reevaluating the Role of Corticosteroids in Septic Shock: An Updated Meta-Analysis of Randomized Controlled Trials | Critically low |
| Liang, 2021 - Corticosteroids for Treating Sepsis in Adult Patients: A Systematic Review and Meta-Analysis | Critically low |
| Lin, 2019 - Impact and Beneficial Critical Points of Clinical Outcome in Corticosteroid Management of Adult Patients With Sepsis: Meta-Analysis and GRADE Assessment | Critically low |
| Lu, 2021 - Efficacy and safety of corticosteroids in immunocompetent patients with septic shock | Critically low |
| Lu, 2023 - Hydrocortisone combined with Vitamin C and Thiamine in the Treatment of Sepsis/ Septic Shock: a Systematic Review with Meta-Analysis and Trial Sequential Analysis | Critically low |

| | |
|---|---|
| Lyu, 2018 - Effect of Low-Dose Hydrocortisone Therapy in Adult Patients With Septic Shock: A Meta-Analysis With Trial Sequential Analysis of Randomized Controlled Trials | Critically low |
| Na, 2021 - Hydrocortisone, ascorbic acid, and thiamine (HAT) for sepsis and septic shock: a meta-analysis with sequential trial analysis | Critically low |
| Ni, 2019 - Can corticosteroids reduce the mortality of patients with severe sepsis? A systematic review and meta-analysis | Critically low |
| Pitre 2024 - Corticosteroids in Sepsis and Septic Shock: A Systematic Review, Pairwise, and Dose-Response Meta-Analysis | Critically low |
| Rochwerg, 2018 - Corticosteroids in Sepsis: An Updated Systematic Review and Meta-Analysis | Critically low |
| Rygård, 2018 - Low-dose corticosteroids for adult patients with septic shock: a systematic review with meta-analysis and trial sequential analysis | Critically low |
| Saghafi, 2023 - Therapeutic Role of HAT Therapy in Sepsis: A Systematic Review and Meta-Analysis | Critically low |
| Siddiqui, 2019 - Hydrocortisone Reduces 28-day Mortality in Septic Patients: A Systemic Review and Meta-analysis | Critically low |
| Somagutta, 2021 - The efficacy of vitamin C, thiamine, and corticosteroid therapy in adult sepsis patients: a systematic review and meta-analysis | Critically low |
| Son 2021 - New Evidence of Potential Benefits of Dexamethasone and Added on Therapy of Fludrocortisone on Clinical Outcomes of Corticosteroid in Sepsis Patients: A Systematic Review and Meta-Analysis | Critically low |
| Song B 2024 - Early use of low-dose hydrocortisone can reduce in-hospital mortality in patients with septic shock: A systematic review and meta-analysis | Critically low |
| Song Y 2024 - Role of corticosteroids in the treatment of critically ill sepsis patients: a meta-analysis review | Critically low |
| Sudaryadi, 2023 - Comparison of Hydrocortisone with Combined Hydrocortisone, Ascorbic Acid, and Thiamine as an Adjuvant Therapy on Septic Shock Patients on Mortality: A Systematic Review and Meta-analysis | Critically low |
| Wen, 2018 - The Effectiveness and Safety of Corticosteroids therapy in adult critical ill patients with Septic Shock: a meta-analysis of randomized controlled trials | Critically low |
| Wu, 2020 - Effects and safety of separate low-dose hydrocortisone use in patients with septic shock: A meta-analysis | Critically low |
| Wu, 2021 - Effect of combined hydrocortisone, ascorbic acid and thiamine for patients with sepsis and septic shock: a systematic review and meta-analysis | Critically low |
| Wu, 2024 - Effect of hydrocortisone-fludrocortisone combination on mortality in septic shock: a systematic review and meta-analysis | Critically low |
| Xu, 2018 - Do low-dose corticosteroids improve survival or shock reversal from septic shock in adults? Meta-analysis with trial sequential analysis | Critically low |
| Yamamoto, 2020 - Hydrocortisone with fludrocortisone for septic shock: a systematic review and meta-analysis | Critically low |
| Yang, 2020 - Controversies in the application of corticosteroids for pediatric septic shock treatment: a preferred reporting items for systematic reviews and meta-analysis-compliant updated meta-analysis | Critically low |
| Yao, 2021 - Combination therapy of thiamine, vitamin C and hydrocortisone in treating patients with sepsis and septic shock: a meta-analysis and trial sequential analysis | Critically low |
| Zayed, 2022 - Vitamin C, Thiamine, and Hydrocortisone in the Treatment of Sepsis: A Meta-Analysis and Trial Sequential Analysis of Randomized Controlled Trials | Critically low |

| | |
|---|---|
| Zhou, 2018 - Effect of adjunctive corticosteroids on clinical outcomes in adult patients with septic shock- a meta-analysis of randomized controlled trials and trial sequential analysis | Critically low |

### Supplementary Table S7: References to randomized controlled trials included in the 42 systematic reviews on corticosteroids (n = 121)

| RCT ID | Primary reference |
|---|---|
| Aboab 2008 | Aboab J, Polito A, Orlikowski D, Sharshar T, Castel M, Annane D. Hydrocortisone effects on cardiovascular variability in septic shock: a spectral analysis approach. Critical Care Medicine 2008;36(5):1481-6. PMID: 18434902 |
| Agarwal 2022 | Agarwal M, Dhar M, Agarwal D, Murlidharan A. Early initiation of low-dose hydrocortisone therapy for septic shock in geriatric patients: a randomized control trial. Journal of the Association of Physicians of India 2022;70(2):11-2. |
| Angus 2020 | Angus DC, Derde L, Al-Beidh F, Annane D, Arabi Y, Beane A, et al. Effect of hydrocortisone on mortality and organ support in patients with severe COVID-19: the REMAP-CAP COVID-19 corticosteroid domain randomized clinical trial. JAMA 2020;324(13):1317-29. DOI: 10.1001/jama.2020.17022 |
| Annane 2002 | Annane D, Sebille V, Charpentier C, Bollaert PE, François B, Korach JM, et al. Effect of treatment with low doses of hydrocortisone and fludrocortisone on mortality in patients with septic shock. JAMA 2002;288(7):862-71. PMID: 12186604 |
| Annane 2010 | COIITSS Study Investigators, Annane D, Cariou A, Maxime V, Azoulay E, D'honneur G, et al. Corticosteroid treatment and intensive insulin therapy for septic shock in adults: a randomized controlled trial. JAMA 2010;303(4):341-8. PMID:20103758 |
| Annane 2018 | Annane D, Renault A, Brun-Buisson C, Megarbane B, Quenot JP, Siami S, et al. Hydrocortisone plus fludrocortisone for adults with septic shock. New England Journal of Medicine 2018;378(9):809-18. PMID: 29490185 |
| Arabi 2010 | Arabi YM, Aljumah A, Dabbagh O, Tamim HM, Rishu AH, Al- Abdulkareem A, et al. Low-dose hydrocortisone in patients with cirrhosis and septic shock: a randomized controlled trial. Canadian Medical Association Journal 2010;182(18):1971-7. PMID: 21059778 |
| Balakrishnan 2018 | Gandhi, H.; Balakrishnan, M.; Shah, K.; Pandya, H.; Patel, R.; Keshwani, S.; Yadav, N. Hydrocortisone, Vitamin C and thiamine for the treatment of sepsis and septic shock following cardiac surgery. Indian J. Anaesth., 2018, 62(12), 934-939. PMID: 30636793 |
| Birudaraju 2022 | Birudaraju D, Hamal S, Tayek JA. Solumedrol treatment for severe sepsis in humans with a blunted adrenocorticotropic hormone-cortisol response: a prospective randomized double-blind placebo-controlled pilot clinical trial.Journal of Intensive Care Medicine 2022;37(5):693-7. DOI: 10.1177/08850666211038883 |
| Blum 2015 | Blum CA, Nigro N, Briel M, Schuetz P, Ullmer E, Suter-Widmer, et al. Adjunct prednisone therapy for patients with communityacquired pneumonia: a multicentre, double-blind, randomised, placebo-controlled trial. Lancet 2015;385(9977):1511-8. PMID:25608756 |
| Bollaert 1998 | Bollaert PE, Charpentier C, Levy B, Debouverie M, Audibert G, Larcan A. Reversal of late septic shock with supraphysiologic doses of hydrocortisone. Critical Care Medicine 1998;26(4):645-50. PMID: 9559600 |
| Bone 1987 | Bone RG, Fisher CJ, Clemmer TP, Slotman GJ, Metz CA, Balk RA. A controlled clinical trial of high-dose methylprednisolone in the treatment of severe sepsis and septic shock. New England Journal of Medicine 1987;317(11):653-8. PMID: 3306374 |
| Branco 2014 | Branco RG, Amoretti CF, Garcia PCR, et al: Corticosteroid replacement in sepsis induces lymphopenia: Results of a randomized controlled trial. Pediatr Crit Care Med 2014; 15(4 Suppl):12 |
| Briegel 1999 | Briegel J, Forst H, Haller M, Schelling G, Kilger E, Kuprat G, et al. Stress doses of hydrocortisone reverse hyperdynamic septic shock: a prospective, randomized, double-blind, single-center study. Critical Care Medicine 1999;27(4):723-32. PMID: 10321661 |

| | |
|---|---|
| Briegel 2001 | Briegel, J.; Jochum, M.; Gippner-Steppert, C.; Thiel, M. Immunomodulation in septic shock: Hydrocortisone differentially regulates cytokine responses. J. Am. Soc. Nephrol. 2001, 12 (Suppl. 17), S70–S74. |
| Chang 2020 | Chang P, Liao Y, Guan J, Guo Y, Zhao M, Hu J, et al. Combined treatment with hydrocortisone, vitamin c, and thiamine for sepsis and septic shock (HYVCTTSSS): a randomized controlled clinical trial. Chest 2020;158:174-82. DOI: 10.1016/j.chest.2020.02.065 |
| Chawla 1999 | Chawla K, Kupfer Y, Tessler S. Hydrocortisone reverses refractory septic shock. Critical Care Medicine 1999;27(1):A33. |
| Cicarelli 2007 | Cicarelli DD, Vieira JE, Benseñor FE. Early dexamethasone treatment for septic shock patients: a prospective randomized clinical trial. São Paulo Medical Journal 2007;125(4):237-41. PMID: 17992396 |
| Cicarelli 2006 | Cicarelli DD, Benseñor FE, Vieira JE. Effects of Single Dose of Dexamethasone on Patients With Systemic Inflammatory Response. Sao Paulo Med J = Rev paulista medicina (2007) 124(2):90–5. DOI: 10.1590/S1516-31802006000200008 |
| Confalonieri 2005 | Confalonieri M, Urbino R, Potena A, Piatella M, Parigi P, Giacomo P, et al. Hydrocortisone infusion for severe community acquired pneumonia: a preliminary randomized study. American Journal of Respiratory and Critical Care Medicine 2005;171(3):242-8. PMID: 15557131 |
| Corral-Gudino 2021 | Corral-Gudino L, Bahamonde A, Arnaiz-Revillas F, Gómez-Barquero J, Abadía-Otero J, García-Ibarbia C, et al. Methylprednisolone in adults hospitalized with COVID-19 pneumonia: an open-label randomized trial (GLUCOCOVID). Wiener Klinische Wochenschrift 2021;133:303-11. DOI: 10.1007/s00508-020-01805-8 |
| CSG 1963 | Cooperative Study Group. The effectiveness of hydrocortisone in the management of severe infections. JAMA 1963;183(6):462-5. DOI: 10.1001/jama.1963.63700060029012 |
| de Graaf 2014 | de Graaf H, Ramakrishnan KA, Pappachan J, et al. Abstract 621: evaluation of corticosteroid replacement therapy in children with severe septic shock – a randomised intervention trial. Pediatr Crit Care Med. 2014;15:141. |
| Deng 2011 | Deng QM, Shang D, Wan XY (2011) Efects of low-dose glucocorticoid on renal function and prognosis in patients with sepsis. Zhongguo Wei Zhong Bing Ji Jiu Yi Xue 23:183–184 |
| Dequin 2020 | Dequin PF, Heming N, Meziani F, Plantefève G, Voiriot G, Badié J, et al. Effect of hydrocortisone on 21-day mortality or respiratory support among critically ill patients with COVID-19: a randomized clinical trial. JAMA 2020;324(13):1298-306. DOI:10.1001/jama.2020.16761 |
| Dequin 2023 | Dequin P-F, Meziani F, Quenot J-P, Kamel T, Ricard J-D, Badie J, et al. Hydrocortisone in severe community-acquired pneumonia. New England Journal of Medicine 2023;388(21):1931-41. DOI: 10.1056/NEJMoa2215145 |
| Doluee 2018 | Talebi Doluee M, Salehi M, Mahmoudi Gharaee A, Jalalyazdi M, Reihani H. The effect of physiologic dose of intravenous hydrocortisone in patients with refractory septic shock: a randomized control trial. Journal of Emergency Practice and Trauma 2018;4:29-33. DOI: 10.15171/JEPT.2017.25 |
| Edalatifard 2020 | Edalatifard M, Akhtari M. Intravenous Methylprednisolone Pulse as a Treatment for Hospitalised Severe COVID-19 Patients: Results From a Randomised Controlled Clinical Trial. Eur Respir J (2020) 56(6):2002808. DOI: 10.1183/13993003.02808-2020 |
| El Ghamrawy 2006 | El-Ghamraway AH, Shokeir MH, Esmat AA. Effects of low dose hydrocortisone in ICU patients with severe community acquired pneumonia. Egyptian Journal of Chest 2006;55:91-9. |
| El-Nawawy 2017 | El-Nawawy A, Khater D, Omar H, Wali Y. Evaluation of early corticosteroid therapy in management of paediatric septic shock in paediatric intensive care patients: a randomized clinical study. Pediatric Infectious Disease Journal 2017;36(2):155-9. PMID: 27798546 |

| | |
|---|---|
| Feng 2023 | Feng F, Yang H, Yang W, Chen Y. Metabolic resuscitation therapy in critically ill patients with sepsis and septic shock: a pilot prospective randomized controlled trial. Open Med. 2023;18(1):20230637. DOI: 10.1515/med-2023-0637 |
| Fernández-Serrano 2011 | Fernández-Serrano S, Dorca J, Garcia-Vidal C, Fernández-Sabé N, Carratalà J, Fernández-Agüera A, et al. Effect of corticosteroids on the clinical course of community-acquired pneumonia: a randomized controlled trial. Critical Care 2011;15(2):R96. PMID: 21406101 |
| Fuji 2020 | Fujii T, Luethi N, Young PJ, Frei DR, Eastwood GM, French CJ, et al. Effect of vitamin C, hydrocortisone, and thiamine vs hydrocortisone alone on time alive and free of vasopressor support among patients with septic shock: the VITAMINS randomized clinical trial. JAMA. 2020; 323(5):423–31. DOI: 10.1001/jama.2019.22176 |
| Ghanei 2021 | Ghanei M, Solaymani-Dodaran M, Qazvini A, Ghazale AH, Setarehdan SA, Saadat SH, et al. The efficacy of corticosteroids therapy in patients with moderate to severe SARS-Cov-2 infection: a multicenter, randomized, open-label trial. Respiratory Research 2021;22(1):245. DOI: 10.1186/s12931-021-01833-6 |
| Gordon 2014 | Gordon AC, Mason AJ, Perkins GD, Stotz M, Terblanche M, Ashby D, et al. The interaction of vasopressin and corticosteroids in septic shock: a pilot randomized controlled trial. Critical Care Medicine 2014;42(6):1325-33. PMID: 24557425 |
| Gordon 2016 | Gordon AC, Mason AJ, Thirunavukkarasu N, Perkins GD, Cecconi M, Cepkova M, et al. Effect of early vasopressin vs norepinephrine on kidney failure in patients with septic shock: the VANISH randomized clinical trial. JAMA 2016;316(5):509-18. PMID: 27483065 |
| Horby 2021 | Horby P, Lim WS, Emberson JR, Mafham M, Bell JL, Linsell L, et al. Dexamethasone in hospitalized patients with Covid-19. New England Journal of Medicine 2021;384(8):693-704. DOI: 10.1056/NEJMoa2021436 |
| Hu 2009 | Hu B, Li JG, Liang H, Zhou Q, Yu Z, Li L, et al. The effect of low-dose hydrocortisone on requirement of norepinephrine and lactate clearance in patients with refractory septic shock. Zhongguo Wei Zhong Bing Ji Jiu Yi Xue 2009;21(9):529-31. PMID:19751560 |
| Huang 2014 | Huang R, Zhang Z, Xu M, Chang X, Qiao Q, Wang L, et al. Effect of Sini decoction on function of hypothalamic-pituitary-adrenal axis in patients with sepsis. Zhonghua Wei Zhong Bing Ji Jiu Yi Xue 2014;26(3):184-7. PMID: 24598293 |
| Huh 2007 | Huh JW, Lim CM, Koh Y, Hong SB. Effect of low doses of hydrocortisone in patient with septic shock and relative adrenal insufficiency: 3 days versus 7 days treatment. Critical Care Medicine 2006;34(12 Suppl):A101. |
| Hussein 2021 | Hussein AA, Sabry NA, Abdalla MS, Farid SF. A prospective, randomised clinical study comparing triple therapy regimen to hydrocortisone monotherapy in reducing mortality in septic shock patients. Int J Clin Pract. 2021;75(9):e14376. DOI: 10.1111/ijcp.14376 |
| Hwang 2020 | Hwang SY, et al. (2020) Combination therapy of vitamin C and thiamine for septic shock: a multi-centre, double-blinded randomized, controlled study. Intensive Care Medicine 46(11): 2015–2025. |
| Hyvernat 2016 | Hyvernat H, Barel R, Gentilhomme A, Césari-Giordani JF, Freche A, Kaidomar M, et al. Effects of increasing hydrocortisone to 300 mg per day in the treatment of septic shock: a pilot study. Shock 2016;46:498-505. PMID: 27405061 |
| Iglesias 2020 | Iglesias J, Vassallo AV, Patel VV, Sullivan JB, Cavanaugh J, Elbaga Y. Outcomes of metabolic resuscitation using ascorbic acid, thiamine, and glucocorticoids in the early treatment of sepsis: the ORANGES trial. Chest 2020;158(1):164-73. DOI: 10.1016/j.chest.2020.02.049 |
| Jamshidi 2021 | Jamshidi M, Zeraati M, Forouzanfar B, Tahrekhani M, Motamed N. Effects of triple combination of hydrocortisone, thiamine, and vitamin C on clinical outcome in patients with septic shock: a single-center randomized controlled trial. Journal of Research in Medical Sciences 2021;26(1):47. DOI: 10.4103/jrms.JRMS_593_19 |

| | |
|---|---|
| Jiang 2019 | Jiang X, You D, Zhao H, Yang F, Yuan Z, Lu P, et al. Early low- dose glucocorticoid therapy effectively suppresses serum pro-inflammatory factors such as IL-6 and inhibits apoptosis of CD4+ cells in septic shock patients. International Journal of Clinical and Experimental Medicine 2019;12(1):718-23. |
| Karimpour 2019 | Karimpour H, Bahrami A, Amini S, Rezaei M, Amini-Saman J, Shahbazi F. Effects of a high dose of vitamin C along with thiamine in critically-ill patients with septic shock: a preliminary study. J Pharm Res Int 2019;29:1-7. |
| Kaufmann 2008 | Kaufmann, I.; Briegel, J.; Schliephake, F.; Hoelzl, A.; Chouker, A.; Hummel, T.; Schelling, G.; Thiel, M. Stress doses of hydrocortisone in septic shock: Beneficial effects on opsonization-dependent neutrophil functions. Intensive Care Med. 2008, 34, 344–349. |
| Keh 2003 | Keh D, Boehnke T, Weber-Cartens S, Schulz C, Ahlers O, Bercker S, et al. Immunologic and hemodynamic effects of "low-dose" hydrocortisone in septic shock: a double-blind, randomized, placebo-controlled, crossover study. American Journal of Respiratory and Critical Care Medicine 2003;167(4):512-20. PMID: 12426230 |
| Keh 2016 | Keh D, Trips E, Marx G, Wirtz SP, Abduljawwad E, Bercker S, et al. Effect of hydrocortisone on development of shock among patients with severe sepsis: the HYPRESS randomized clinical trial. JAMA 2016;316(17):1775-85. PMID: 27695824 |
| Klastersky 1971 | Klastersky J, Cappel R, Debusscher L. Effectiveness of Betamethasone in Management of Severe Infections. A Double-Blind Study. N Engl J Med (1971) 284(22):1248–50. DOI: 10.1056/NEJM197106032842206 |
| Kurungundla 2008 | Kurugundla N, Irugulapati L, Kilari D, Amchentsev A, Devakonda A, George L, et al. Effect of steroids in septic shock patients without relative adrenal insufficiency. American Thoracic Society 2008;116:NA. |
| Labib 2022 | Labib HA, Ali I, Hassan AI, Kamaly AM, Sherif S, Wahba SS, et al. Evaluation of the role of hydrocortisone either alone or combined with fludrocortisone in the outcome of septic shock in adults. Ain-Shams Journal of Anesthesiology 2022;14:60. DOI: 10.1186/s42077-022-00259-6] |
| Li 2016 | Li G, Gu C, Zhang S, Lian R, Zhang G. Value of glucocorticoid steroids in the treatment of patients with severe community acquired pneumonia complicated with septic shock. Zhonghua Wei Zhong Bing Ji Jiu Yi Xue 2016;28(9):780-4. DOI: 10.3760/cma.j.issn.2095-4352.2016.09.003 |
| Liu 2012 | Liu L, Li J, Huang YZ, Liu SQ, Yang CS, Guo FM, et al. The effect of stress dose glucocorticoid on patients with acute respiratory distress syndrome combined with critical illness- related corticosteroid insufficiency. Zhonghua Nei Ke Za Zhi 2012;51(8):599-603. PMID: 23158856 |
| Loisa 2007 | Loisa P, Parviainen I, Tenhunen J, Hovilehto S, Ruokonen E. Effect of mode of hydrocortisone administration on glycemic control in patients with septic shock: a prospective randomized trial. Critical Care 2007;11(1):R21. PMID: 17306016 |
| Lucas 1984 | Lucas CE, Ledgerwood AM. The cardiopulmonary response to massive doses of steroids in patients with septic shock. Arch Surg. 1984;119:537–41. |
| Luce 1988 | Luce JM, Montgomery AB, Marks JD, Turner J, Metz CA, Murray JF. Ineffectiveness of high-dose methylprednisolone in preventing parenchymal lung injury and improving mortality in patients with septic shock. American Review of Respiratory Diseases 1988;138(1):62-8. PMID: 3202402 |
| Lv 2017 | Lv QQ, Gu XH, Chen QH, Yu JQ, Zheng RQ. Early initiation of low-dose hydrocortisone treatment for septic shock in adults: a randomized clinical trial. American Journal of Emergency Medicine 2017;35(12):1810-4. PMID: 28615145 |

| | |
|---|---|
| Lyu 2022 | Lyu QQ, Zheng RQ, Chen QH, Yu JQ, Shao J, Gu XH. Early administration of hydrocortisone, vitamin C, and thiamine in adult patients with septic shock: a randomized controlled clinical trial. Critical Care 2022;26(1):295. DOI: 10.1186/s13054-022-04175-x |
| Mansour 2012 | Mansour M, Tarek M, Zayton Y, et al. 1576 effect of dexamethasone therapy on the expression of macrophage migration inhibitory factor in critically ill children with septic shock. Arch Dis Child. 2012;97:A446–A446. |
| McHardy 1972 | McHardy VU, Schonell ME. Ampicillin dosage and use of prednisolone in treatment of pneumonia: co-operative controlled trial. British Medical Journal 1972;4(5840):569-73. PMID: 4404939 |
| Meduri 2007 | Meduri GU, Golden E, Freire AX, Taylor E, Zaman M, Carson SJ, et al. Methylprednisolone infusion in early severe ARDS: results of a randomized controlled trial. Chest 2007;131(4):954-63. PMID: 17426195 |
| Meduri 2009 | Meduri GU, Golden E, Umberger R. Prospective Double-Blind Randomized Clinical Trial on the Effects of Low-Dose Hydrocortisone Infusion in Patients With Severe Sepsis. Chest (2009) 136(4):45S. DOI: 10.1016/S0012-3692(16)48001-3 |
| Meduri 2022 | Meduri GU, Shih MC, Bridges L, Martin TJ, El-Solh A, Seam N, et al. Low-dose methylprednisolone treatment in critically ill patients with severe community-acquired pneumonia. Intensive Care Medicine 2022;48(8):1009-23. DOI: 10.1007/s00134-022-06684-3 |
| Meijvis 2011 | Meijvis SC, Hardeman H, Remmelts HH, Heijligenberg R, Rijkers GT, van Velzen-Blad H, et al. Dexamethasone and length of hospital stay in patients with community-acquired pneumonia: a randomised, double-blind, placebo-controlled trial. Lancet 2011;377(9782):2023-30. PMID: 21636122 |
| Menon 2017 | Menon K, McNally D, O'Hearn K, Acharya A, Wong HR, Lawson M, et al. A randomized controlled trial of corticosteroids in paediatric septic shock: a pilot feasibility study. Pediatric Critical Care Medicine 2017;18(6):505-12. PMID: 28406862 |
| Min 1975 | Min M, U T, Aye M, et al. Hydrocortisone in the management of dengue shock syndrome. Southeast Asian J Trop Med Public Health. 1975;6:573–579. |
| Mirea 2014 | Mirea L, Ungureanu R, Pavelescu D, Grintescu IC, Dumitrache C, Grintescu I, et al. Continuous administration of corticosteroids in septic shock can reduce risk of hypernatraemia. Critical Care 2014;18:S86. |
| Mohamed 2020 | Mohamed ZU, Prasannan P, Moni M, Edathadathil F, Prasanna P, Menon A, et al. Vitamin C therapy for routine care in septic shock (Victor) trial: effect of intravenous vitamin C, thiamine, and hydrocortisone administration on inpatient mortality among patients with septic shock. Indian Journal of Critical Care Medicine 2020;24(8):653-61. DOI: 10.5005/jp-journals-10071-23517 |
| Mohamed 2023 | Mohamed A, Abdelaty M, Saad MO, Shible A, Mitwally H, Akkari AR, et al. Evaluation of hydrocortisone, vitamin C, and thiamine for the treatment of septic shock: a randomized controlled trial (The HYVITS Trial). Shock 2023;59(5):697-701. DOI: 10.1097/SHK.0000000000002110 |
| Moreno 2011 | Moreno, R.; Sprung, C.L.; Annane, D.; Chevret, S.; Briegel, J.; Keh, D.; Singer, M.; Weiss, Y.G.; Payen, D.; Cuthbertson, B.H.; et al. Time course of organ failure in patients with septic shock treated with hydrocortisone: Results of the Corticus study. Intensive Care Med. 2011, 37, 1765–1772. |
| Moskowitz 2020 | Moskowitz A, Huang DT, Hou PC, Gong J, Doshi PB, Grossestreuer AV, et al. Effect of ascorbic acid, corticosteroids, and thiamine on organ injury in septic shock: the ACTS randomized clinical trial. JAMA 2020;324(7):642-50. DOI: 10.1001/jama.2020.11946 |
| Mussack 2005 | Mussack T, Briegel J, Schelling G, et al. Effect of stress doses of hydrocortisone on S-100B vs. interleukin-8 and polymorphonuclear elastase levels in human septic shock. Clin Chem Lab Med 2005; 43(3): 259–268. |
| Nafae 2013 | Nafae RM, Ragab MI, Amany FM, Rashed SB. Adjuvant role of corticosteroids in the treatment of community acquired pneumonia. Egyptian Journal of Chest Diseases and Tuberculosis 2013; 62:439-45. |

| | |
|---|---|
| Nagy 2013 | Nagy B, Gaspar I, Papp A, Bene Z, Nagy B Jr, Voko Z, et al. Efficacy of methylprednisolone in children with severe community acquired pneumonia. Pediatric Pulmonology 2013; 48:168-75. PMID: 22588852 |
| Ngaosuwan 2018 | Ngaosuwan K, Ounchokdee K, Chalermchai T. Clinical outcomes of minimized hydrocortisone dosage of 100mg/day on lower occurrence of hyperglycaemia in septic shock. Shock 2018;50(3):280-5. PMID: 29176402 |
| Oppert 2005 | Oppert M, Schindler R, Husung C, Offerman K, Graef KJ, Boenisch O, et al. Low-dose hydrocortisone improves shock reversal and reduces cytokine levels in early hyperdynamic septic shock. Critical Care Medicine 2005;33(11):2457-64. PMID: 16276166 |
| Pierre de Oliveira 2023 | Pierre de Oliveira, E., Marconi de Sousa, J., Jaoude, C.V.G., Malacize, V.Q., Lobato, L.C. & et al. Journal of Community Medicine and Public Health 2023; 369 DOI: 10.29011/2577-2228.100369 |
| Pongpanich 1973 | Pongpanich B, Bhanchet P, Phanichyakarn P, et al. Studies on dengue hemorrhagic fever. Clinical study: an evaluation of steroids as a treatment. J Med Assoc Thai. 1973;56:6–14. |
| Póvoa 2015 | P. Povoa, J. I. F. Salluh, M. L. Martinez et al., "Clinical impact of ´stress dose steroids in patients with septic shock: insights from the PROWESS-Shock trial," Critical Care, vol. 19, no. 193, 2015. |
| Raghu 2021 | Raghu K, Ramalingam K. Safety and Efficacy of Vitamin C, Vitamin B1, and Hydrocortisone in clinical outcome of septic shock receiving standard care: a quasi experimental randomized open label two arm parallel group study. Eur J Mol Clin Med. 2021;8(2):873–91. |
| Ram 2022 | Ram GK, Shekhar S, Singh RB, Anand R, De RR, Kumar N. Hyperglycemia risk evaluation of hydrocortisone intermittent boluses versus continuous infusion in septic shock: a prospective randomized trial. Anesthesia Essays and Researchers 2022;16(3):321-5. DOI: 10.4103/aer.aer115__22 |
| Reddy 2020 | Reddy PR, Samavedam S, Aluru N, Yelle S, Rajyalakshmi B. Metabolic resuscitation using hydrocortisone, ascorbic acid, and thiamine: Do individual components influence reversal of shock independently? Indian J Crit Care Med. 2020;24(8):649-52. DOI: 10.5005/jp-journals-10071-23515 |
| Rezk 2013 | Rezk NA, Ibrahim AM. Effects of methylprednisolone in early ARDS. Egyptian Journal of Chest Diseases and Tuberculosis 2013;62(1):167-72. DOI: 10.1016/j.ejcdt.2013.02.013 |
| Rinaldi 2006 | Rinaldi S, Adembri C, Grechi S, DeGaudio R. Low- dose hydrocortisone during severe sepsis: effects on microalbuminemia. Critical Care Medicine 2006;34(9):2334-9. PMID: 16850006 |
| Sabry 2011 | Sabry NA, El-Din Omar E. Corticosteroids and ICU course of community acquired pneumonia in Egyptian settings. Pharmacology and Pharmacy 2011;2:73-81. |
| Sai 2021 | Sai TA, Satyanarayana PVS, Gollapudi S. Effect of treatment with low-dose corticosteroids on mortality in patients with severe sepsis and septic shock. JP Journals 2021;25(Suppl 1):S104-5. DOI: 10.5005/jp-journals-10071-23711.183 |
| Sang 2020 | Sang ZZ, Wang SJ, Wang W, Feng SY, Wang SL, Li Y. Clinical efficacy of hydrocortisone combined with vitamin C and vitamin B1 in the treatment of septic shock in the elderly. Chin J Gerontol. 2020;40(20):4351–4. |
| Schelling 2001 | Schelling, G.; Briegel, J.; Roozendaal, B.; Stoll, C.; Rothenhäusler, H.-B.; Kapfhammer, H.-P. The effect of stress doses of hydrocortisone during septic shock on posttraumatic stress disorder in survivors. Biol. Psychiatry 2001, 50, 978–985. |
| Schumer 1976 | Schumer W. Steroids in the treatment of clinical septic shock. Annals of Surgery 1976;184(3):333-41. PMID: 786190 |
| Sevransky 2021 | Sevransky JE, Rothman RE, Hager DN, Bernard GR, Brown SM, Buchman TG, et al. Effect of vitamin C, thiamine, and hydrocortisone on ventilator- and vasopressor-free days in patients with sepsis: the VICTAS randomized clinical trial. JAMA 2021;325(8):742-50. DOI: 10.1001/jama.2020.24505 |

| | |
|---|---|
| Shi 2019 | Shi XN, Xu MX, Geng WJ, et al. [Therapeutic effects of early application of low-dose glucocorticoid on septic shock in children and its effects on immune function of patients]. Hebei Med J. 2019;41:1770–1774. |
| Slusher 1996 | Slusher T, Gbadero D, Howard C, Lewinson L, Giroir B, Toro L, et al. Randomized, placebo-controlled, double blinded trial of dexamethasone in African children with sepsis. Pediatric Infectious Diseases Journal 1996;15(7):579-83. PMID: 8823850 |
| Snijders 2010 | Snijders D, Daniels JMA, de Graaff CS, van der Werf TS, Boersma WG. Efficacy of corticosteroids in community-acquired pneumonia – a randomized double blinded clinical trial. American Journal of Respiratory and Critical Care Medicine 2010;181(9):975-82. PMID: 20133929 |
| Sprung 1984 | Sprung CL, Caralis PV, Marcial EH, Pierce M, Gelbard MA, Long WM, et al. The effects of high-dose corticosteroids in patients with septic shock. A prospective, controlled study. New England Journal of Medicine 1984;311(18):1137-43. PMID: 6384785 |
| Sprung 2008 | Sprung C, Annane D, Keh D, Moreno R, Singer M, Freivogel K, et al. Hydrocortisone therapy for patients with septic shock. New England Journal of Medicine 2008;358(2):111-24. PMID: 18184957 |
| Sui 2013 | Sui DJ, Zhang W, Li WS, Zhao HX, Wang ZY. Clinical efficacy in the treatment of severe community acquired pneumonia and its impact on CRP. Chinese Journal of Integrative Medicine 2013;18:1171-3. |
| Sumarmo 1982 | Sumarmo D, Talogo W, Asrin A, et al. Failure of hydrocortisone to affect outcome in dengue shock syndrome. Pediatrics. 1982;69:45–49. |
| Tagaro 2017 | Tagaro A, Otheo E, Baquero-Artigao F, Navarro ML, Velasco R, Ruiz M, et al. Dexamethasone for parapneumonic pleural effusion: a randomized, double-blind, clinical trial. Journal of Pediatrics 2017;185:117-23. PMID: 28363363 |
| Tandan 2005 | Tandan SM, Guleria R, Gupta N. Low dose steroids and adrenocortical insufficiency in septic shock: a double-blind randomised controlled trial from India. In: Proceedings of the American Thoracic Society Meeting. 2005:A24. |
| Tassniyom 1993 | Tassniyom S, Vasanawathana S, Chirawatkul A, et al. Failure of high-dose methylprednisolone in established dengue shock syndrome: a placebo-controlled, double-blind study. Pediatrics. 1993;92:111–115. |
| Tilouche 2019 | Tilouche N, Jaoued O, Ben Sik Ali H, Gharbi R, Fekih Hassen M, Elatrous S. Comparison between continuous and intermittent administration of hydrocortisone during septic shock: a randomized controlled clinical trial. Shock 2019;52:481-6. DOI: 10.1097/SHK.0000000000001316] [PMID: 30628950 |
| Tomazini 2020 | Tomazini BM, Maia IS, Cavalcanti AB, Berwanger O, Rosa RG, Veiga VC, et al. Effect of dexamethasone on days alive and ventilator-free in patients with moderate or severe acute respiratory distress syndrome and COVID-19: the CoDEX randomized clinical trial. JAMA 2020;324(13):1307-16. DOI: 10.1001/jama.2020.17021 |
| Tongyoo 2016 | Tongyoo S, Permpikul C, Mongkolpun W, Vattanavanit V, Udompanturak S, Kocak M, et al. Hydrocortisone treatment in early sepsis associated acute respiratory distress syndrome: results of a randomized controlled trial. Critical Care 2016;20(1):329. PMID: 27741949 |
| Torres 2015 | Torres A, Sibila O, Ferrer M, Polverino E, Menendez R, Mensa J, et al. Effect of corticosteroids on treatment failure among hospitalized patients with severe community-acquired pneumonia and high inflammatory response: a randomized clinical trial. JAMA 2015;313(7):677-86. PMID: 25688779 |
| Valoor 2009 | Valoor HT, Singhi S, Jayashree M. Low-dose hydrocortisone in paediatric septic shock: an exploratory study in a third world setting. Pediatric Critical Care Medicine 2009;10(1):121-5. PMID: 19057445 |
| VASSCSG 1987 | Veterans Administration Systemic Sepsis Cooperative Study Group. Effect of high-dose glucocorticoid therapy on mortality in patients with clinical signs of systemic sepsis. New England Journal of Medicine 1987;317(11):659-65. PMID: 2888017 |

| | |
|---|---|
| Venkatesh 2018 | Venkatesh B, Finfer S, Cohen J, Rajbhandari D, Arabi Y, Bellomo R, et al. Adjunctive glucocorticoid therapy in patients with septic shock. New England Journal of Medicine 2018;378:797-808. DOI: 10.1056/NEJMoa1705835 |
| Villar 2020 | Villar J, Ferrando C, Martínez D, Ambrós A, Muñoz T, Soler JA, et al. Dexamethasone treatment for the acute respiratory distress syndrome: a multicentre, randomised controlled trial. Lancet Respiratory Medicine 2020;8(3):267-76. DOI: 10.1016/S2213-2600(19)30417-5 |
| Wan 2014 | Wan X, Su M, Yue J, Huang Y, Zhu W, Wan L. Clinical trial of low-dose corticosteroid treatment for surgical postoperative patients with septic shock. Journal of Kunming Medical University. 2014;35:92-9. |
| Wang 2022 | Wang QZ, Lin XF, Zhu HL. [Influences of Dexamethasone combined with cefoperazone and sulbactam on efficacy of sepsis in children and serum PCT, Presepsin and NT-proBNP]. Drug Eval Res. 2022;45:118–123. |
| Wang 2023 | Wang J, Song Q, Yang S, Wang H, Meng S, Huang L, et al. Effects of hydrocortisone combined with vitamin C and vitamin B1 versus hydrocortisone alone on microcirculation in septic shock patients: a pilot study. Clin Hemorheol Microcirc. 2023;84(2):111–23. DOI: 10.3233/CH-221444 |
| Wani 2020 | Wani SJ, Mufti SA, Jan RA, Shah SU, Qadri SM, Khan UH, et al. Combination of vitamin C, thiamine and hydrocortisone added to standard treatment in the management of sepsis: results from an open label randomised controlled clinical trial and a review of the literature. Infectious Diseases 2020;52(4):271-78. DOI: 10.1080/23744235.2020.1718200 |
| Wen 2004 | Wen WY. [Observation on the treatment of septic shock in children with high-dose methylprednisolone]. J Pract Diagn Ther. 2004;18:404–405. |
| Wittermans 2020 | Wittermans E, Vestjens S, Spoorenberg S, Blok W, Grutters J, Janssen R, et al. Late breaking abstract - adjunctive treatment with oral dexamethasone in adults hospitalised with community-acquired pneumonia: a randomised clinical trial. European Respiratory Journal 2020;56:4132. DOI: 10.1183/13993003.congress-2020 |
| Yildiz 2002 | Yildiz O, Doganay M, Aygen B, Guven M, Keleutimur F, Tutuu A. Physiological-dose steroid therapy in sepsis. Critical Care 2002;6(3):251-9. PMID: 12133187 |
| Yildiz 2011 | Yildiz O, Tanriverdi F, Simsek S, Aygen B, Kelestimur F. The effects of moderate-dose steroid therapy in sepsis: a placebo-controlled, randomized study. Journal of Research in Medical Sciences 2011;16(11):1410-21. [PMID: 22973341] |
| Yuan 2010 | Yuan YH, Xu ZY, Lu XL. [Observation on the efficacy of hydrocortisone in the treatment of septic shock in children]. Chin J Misdiagn. 2010;10:61. |
| Zeng 2014 | Zeng R. [The clinical effects comparison of different doses of methylprednisolone in the treatment of children with severe sepsis]. J Chengdu Med Coll.2014;9:734–737. |
| Zhou 2015 | Zhou M. Application value of glucocorticoid for comprehensive treatment of acute respiratory distress syndrome induced by serious community acquired pneumonia. Clinical Medicine and Engineering 2015;22:57-8. DOI: 10.3969/j.issn.1674-4659.2015.01.0057 |

### Supplementary Table S8: Distribution of randomized controlled trials across systematic reviews - binary SRxRCT inclusion matrix (n = 42, n = 121)

| RCTs | | | Systematic reviews | | | | | | | | | | | | | | | | | | | | | | | | | | | | | | | | | | | | | | | | | |
|---|---|---|---|---|---|---|---|---|---|---|---|---|---|---|---|---|---|---|---|---|---|---|---|---|---|---|---|---|---|---|---|---|---|---|---|---|---|---|---|---|---|---|---|---|
| rct_id | first_author | year | Aletreby 2019 | Allen 2019 | Annane 2015 | Annane 2019 | Annane 2025 | Assouline 2021 | Barakakis 2019 | Deng 2024 | Deng 2025 | Fang 2019 | Fratoni 2020 | Kato 2023 | Lee 2022 | Li 2025 | Lian 2019 | Liang 2021 | Lin 2019 | Lu 2021 | Lu 2023 | Lyu 2020 | Na 2021 | Ni 2019 | Pitre 2024 | Rochwerg 2018 | Rygard 2018 | Saghafi 2024 | Siddiqi 2019 | Somagutta 2021 | Son 2021 | Song B 2024 | Song Y 2024 | Sudaryadi 2023 | Wen 2019 | Wu 2020 | Wu 2021 | Wu 2024 | Xu 2018 | Yamamoto 2020 | Yang 2020 | Yao 2021 | Zayed 2022 | Zhou 2018 | Number of SRs including the RCT |
| Aboab 2008 | Aboab | 2008 | 0 | 0 | 0 | 1 | 1 | 0 | 0 | 0 | 0 | 1 | 0 | 0 | 0 | 0 | 0 | 1 | 0 | 0 | 0 | 0 | 0 | 0 | 0 | 0 | 1 | 0 | 0 | 0 | 0 | 0 | 0 | 0 | 0 | 0 | 0 | 0 | 0 | 0 | 0 | 0 | 0 | 1 | 6 |
| Agarwal 2022 | Agarwal | 2022 | 0 | 0 | 0 | 0 | 1 | 0 | 0 | 0 | 0 | 0 | 0 | 0 | 0 | 0 | 0 | 0 | 0 | 0 | 0 | 0 | 0 | 0 | 1 | 0 | 0 | 0 | 0 | 0 | 0 | 0 | 0 | 0 | 0 | 0 | 0 | 0 | 0 | 0 | 0 | 0 | 0 | 0 | 2 |
| Angus 2020 | Angus | 2020 | 0 | 0 | 0 | 0 | 1 | 0 | 0 | 0 | 0 | 0 | 0 | 0 | 0 | 0 | 0 | 1 | 0 | 0 | 0 | 0 | 0 | 0 | 0 | 0 | 0 | 0 | 0 | 0 | 0 | 0 | 0 | 0 | 0 | 0 | 0 | 0 | 0 | 0 | 0 | 0 | 0 | 0 | 2 |
| Annane 2002 | Annane | 2002 | 1 | 0 | 1 | 1 | 1 | 0 | 1 | 0 | 0 | 1 | 0 | 0 | 0 | 0 | 1 | 1 | 1 | 1 | 0 | 1 | 0 | 1 | 1 | 1 | 1 | 0 | 1 | 0 | 1 | 0 | 1 | 0 | 1 | 0 | 0 | 1 | 1 | 1 | 0 | 0 | 0 | 1 | 23 |
| Annane 2010 | Annane | 2010 | 0 | 0 | 1 | 1 | 1 | 0 | 0 | 0 | 0 | 0 | 0 | 0 | 0 | 0 | 0 | 1 | 0 | 0 | 0 | 0 | 0 | 0 | 0 | 0 | 0 | 0 | 0 | 0 | 0 | 0 | 0 | 0 | 0 | 0 | 0 | 1 | 0 | 1 | 0 | 0 | 0 | 0 | 6 |
| Annane 2018 | Annane | 2018 | 1 | 1 | 0 | 1 | 1 | 0 | 1 | 0 | 0 | 1 | 0 | 0 | 0 | 0 | 1 | 1 | 1 | 0 | 0 | 1 | 0 | 1 | 1 | 1 | 1 | 0 | 1 | 0 | 1 | 0 | 1 | 0 | 1 | 0 | 0 | 1 | 0 | 1 | 0 | 0 | 0 | 1 | 21 |
| Arabi 2010 | Arabi | 2010 | 0 | 0 | 1 | 1 | 1 | 0 | 0 | 0 | 0 | 1 | 0 | 0 | 0 | 0 | 1 | 1 | 1 | 0 | 0 | 1 | 0 | 1 | 1 | 1 | 1 | 0 | 0 | 0 | 0 | 0 | 1 | 0 | 1 | 1 | 0 | 0 | 1 | 0 | 0 | 0 | 0 | 1 | 17 |
| Balakrishnan 2018 | Balakrishnan | 2018 | 0 | 0 | 0 | 0 | 0 | 0 | 0 | 0 | 0 | 0 | 0 | 0 | 1 | 0 | 0 | 0 | 0 | 0 | 0 | 0 | 1 | 0 | 0 | 0 | 0 | 1 | 0 | 0 | 0 | 0 | 0 | 0 | 0 | 0 | 0 | 0 | 0 | 0 | 0 | 0 | 1 | 0 | 4 |
| Birudaraju 2022 | Birudaraju | 2022 | 0 | 0 | 0 | 0 | 1 | 0 | 0 | 0 | 0 | 0 | 0 | 0 | 0 | 0 | 0 | 0 | 0 | 0 | 0 | 0 | 0 | 0 | 1 | 0 | 0 | 0 | 0 | 0 | 0 | 0 | 0 | 0 | 0 | 0 | 0 | 0 | 0 | 0 | 0 | 0 | 0 | 0 | 2 |
| Blum 2015 | Blum | 2015 | 0 | 0 | 0 | 1 | 1 | 0 | 0 | 0 | 0 | 0 | 0 | 0 | 0 | 0 | 0 | 1 | 0 | 0 | 0 | 0 | 0 | 0 | 0 | 0 | 0 | 0 | 0 | 0 | 0 | 0 | 0 | 0 | 0 | 0 | 0 | 0 | 0 | 0 | 0 | 0 | 0 | 0 | 3 |
| Bollaert 1998 | Bollaert | 1998 | 0 | 0 | 1 | 1 | 1 | 0 | 0 | 0 | 0 | 1 | 0 | 0 | 0 | 0 | 1 | 1 | 1 | 0 | 0 | 1 | 0 | 1 | 1 | 1 | 1 | 0 | 0 | 0 | 1 | 0 | 0 | 0 | 1 | 1 | 0 | 0 | 1 | 0 | 0 | 0 | 0 | 1 | 17 |
| Bone 1987 | Bone | 1987 | 0 | 0 | 1 | 1 | 1 | 0 | 0 | 0 | 0 | 1 | 0 | 0 | 0 | 0 | 1 | 1 | 1 | 0 | 0 | 0 | 0 | 0 | 1 | 1 | 0 | 0 | 0 | 0 | 1 | 0 | 0 | 0 | 1 | 0 | 0 | 0 | 0 | 0 | 0 | 0 | 0 | 0 | 11 |
| Branco 2014 | Branco | 2014 | 0 | 0 | 0 | 0 | 0 | 0 | 0 | 0 | 0 | 0 | 0 | 0 | 0 | 0 | 0 | 0 | 0 | 0 | 0 | 0 | 0 | 0 | 1 | 1 | 0 | 0 | 0 | 0 | 0 | 0 | 0 | 0 | 0 | 0 | 0 | 0 | 0 | 0 | 0 | 0 | 0 | 0 | 2 |
| Briegel 1999 | Briegel | 1999 | 0 | 0 | 1 | 1 | 1 | 0 | 0 | 0 | 0 | 1 | 0 | 0 | 0 | 0 | 1 | 1 | 1 | 1 | 0 | 1 | 0 | 1 | 1 | 1 | 1 | 0 | 0 | 0 | 1 | 0 | 0 | 0 | 1 | 1 | 0 | 0 | 1 | 0 | 0 | 0 | 0 | 1 | 18 |
| Briegel 2001 | Briegel | 2001 | 0 | 0 | 0 | 0 | 0 | 0 | 0 | 0 | 0 | 0 | 0 | 0 | 0 | 0 | 0 | 0 | 0 | 0 | 0 | 0 | 0 | 0 | 0 | 0 | 0 | 0 | 0 | 0 | 1 | 0 | 0 | 0 | 0 | 0 | 0 | 0 | 0 | 0 | 0 | 0 | 0 | 0 | 1 |
| Chang 2020 | Chang | 2020 | 0 | 0 | 0 | 0 | 1 | 1 | 0 | 1 | 0 | 0 | 0 | 1 | 0 | 0 | 0 | 0 | 0 | 0 | 1 | 0 | 1 | 0 | 0 | 0 | 0 | 1 | 0 | 1 | 0 | 0 | 0 | 0 | 0 | 0 | 1 | 0 | 0 | 0 | 0 | 1 | 1 | 0 | 11 |
| Chawla 1999 | Chawla | 1999 | 0 | 0 | 1 | 1 | 1 | 0 | 0 | 0 | 0 | 1 | 0 | 0 | 0 | 0 | 1 | 1 | 1 | 0 | 0 | 0 | 0 | 0 | 1 | 1 | 1 | 0 | 0 | 0 | 0 | 0 | 0 | 0 | 1 | 0 | 0 | 0 | 0 | 0 | 0 | 0 | 0 | 0 | 11 |
| Cicarelli 2007 | Cicarelli | 2007 | 0 | 0 | 1 | 1 | 1 | 0 | 0 | 0 | 0 | 1 | 1 | 0 | 0 | 0 | 1 | 0 | 1 | 1 | 0 | 0 | 0 | 1 | 1 | 1 | 1 | 0 | 0 | 0 | 1 | 0 | 0 | 0 | 1 | 0 | 0 | 0 | 1 | 0 | 0 | 0 | 0 | 1 | 16 |
| Cicarelli 2006 | Cicarelli | 2006 | 0 | 0 | 0 | 0 | 0 | 0 | 0 | 0 | 0 | 0 | 1 | 0 | 0 | 0 | 0 | 1 | 0 | 0 | 0 | 0 | 0 | 0 | 0 | 0 | 0 | 0 | 0 | 0 | 0 | 0 | 0 | 0 | 0 | 0 | 0 | 0 | 0 | 0 | 0 | 0 | 0 | 0 | 2 |
| Confalonieri 2005 | Confalonieri | 2005 | 0 | 0 | 1 | 1 | 1 | 0 | 0 | 0 | 0 | 1 | 0 | 0 | 0 | 0 | 0 | 1 | 1 | 0 | 0 | 0 | 0 | 1 | 1 | 1 | 0 | 0 | 0 | 0 | 1 | 0 | 0 | 0 | 0 | 0 | 0 | 0 | 0 | 0 | 0 | 0 | 0 | 0 | 10 |
| Corral Gudino 2021 | Corral-Gudino | 2021 | 0 | 0 | 0 | 0 | 1 | 0 | 0 | 0 | 0 | 0 | 0 | 0 | 0 | 0 | 0 | 0 | 0 | 0 | 0 | 0 | 0 | 0 | 0 | 0 | 0 | 0 | 0 | 0 | 0 | 0 | 0 | 0 | 0 | 0 | 0 | 0 | 0 | 0 | 0 | 0 | 0 | 0 | 1 |
| CSG 1963 | CSG | 1963 | 0 | 0 | 1 | 1 | 1 | 0 | 0 | 0 | 0 | 0 | 0 | 0 | 0 | 0 | 0 | 0 | 0 | 0 | 0 | 0 | 0 | 0 | 1 | 1 | 1 | 0 | 0 | 0 | 0 | 0 | 0 | 0 | 0 | 0 | 0 | 0 | 0 | 0 | 0 | 0 | 0 | 0 | 6 |
| de Graaf 2014 | de Graaf | 2014 | 0 | 0 | 0 | 0 | 0 | 0 | 0 | 0 | 1 | 0 | 0 | 0 | 0 | 0 | 0 | 0 | 0 | 0 | 0 | 0 | 0 | 0 | 1 | 1 | 0 | 0 | 0 | 0 | 0 | 0 | 0 | 0 | 0 | 0 | 0 | 0 | 0 | 0 | 1 | 0 | 0 | 0 | 4 |
| Deng 2011 | Deng | 2011 | 0 | 0 | 0 | 0 | 0 | 0 | 0 | 0 | 0 | 0 | 0 | 0 | 0 | 0 | 0 | 0 | 0 | 0 | 0 | 0 | 0 | 0 | 0 | 0 | 1 | 0 | 0 | 0 | 0 | 0 | 0 | 0 | 0 | 0 | 0 | 0 | 0 | 0 | 0 | 0 | 0 | 0 | 1 |
| Dequin 2020 | Dequin | 2020 | 0 | 0 | 0 | 0 | 1 | 0 | 0 | 0 | 0 | 0 | 0 | 0 | 0 | 0 | 0 | 1 | 0 | 0 | 0 | 0 | 0 | 0 | 0 | 0 | 0 | 0 | 0 | 0 | 0 | 0 | 0 | 0 | 0 | 0 | 0 | 0 | 0 | 0 | 0 | 0 | 0 | 0 | 2 |
| Dequin 2023 | Dequin | 2023 | 0 | 0 | 0 | 0 | 1 | 0 | 0 | 0 | 0 | 0 | 0 | 0 | 0 | 0 | 0 | 0 | 0 | 0 | 0 | 0 | 0 | 0 | 0 | 0 | 0 | 0 | 0 | 0 | 0 | 0 | 0 | 0 | 0 | 0 | 0 | 0 | 0 | 0 | 0 | 0 | 0 | 0 | 1 |
| Doluee 2018 | Doluee | 2018 | 0 | 0 | 0 | 1 | 1 | 0 | 0 | 0 | 0 | 0 | 0 | 0 | 0 | 0 | 1 | 1 | 0 | 1 | 0 | 0 | 0 | 0 | 1 | 0 | 0 | 0 | 0 | 0 | 0 | 0 | 0 | 0 | 0 | 0 | 0 | 0 | 0 | 0 | 0 | 0 | 0 | 0 | 6 |
| Edalatifard 2020 | Edalatifard | 2020 | 0 | 0 | 0 | 0 | 0 | 0 | 0 | 0 | 0 | 0 | 0 | 0 | 0 | 0 | 0 | 1 | 0 | 0 | 0 | 0 | 0 | 0 | 0 | 0 | 0 | 0 | 0 | 0 | 0 | 0 | 0 | 0 | 0 | 0 | 0 | 0 | 0 | 0 | 0 | 0 | 0 | 0 | 1 |
| El Ghamrawy 2006 | El Ghamrawy | 2006 | 0 | 0 | 0 | 1 | 1 | 0 | 0 | 0 | 0 | 0 | 0 | 0 | 0 | 0 | 0 | 0 | 0 | 0 | 0 | 0 | 0 | 0 | 0 | 0 | 0 | 0 | 0 | 0 | 0 | 0 | 0 | 0 | 0 | 0 | 0 | 0 | 0 | 0 | 0 | 0 | 0 | 0 | 2 |
| El Nawawy 2017 | El-Nawawy | 2017 | 0 | 0 | 0 | 1 | 1 | 0 | 0 | 0 | 0 | 0 | 0 | 0 | 0 | 0 | 0 | 0 | 0 | 0 | 0 | 0 | 0 | 0 | 1 | 1 | 0 | 0 | 0 | 0 | 0 | 0 | 0 | 0 | 0 | 0 | 0 | 0 | 0 | 0 | 1 | 0 | 0 | 0 | 5 |
| Feng 2023 | Feng | 2023 | 0 | 0 | 0 | 0 | 0 | 0 | 0 | 1 | 0 | 0 | 0 | 0 | 0 | 0 | 0 | 0 | 0 | 0 | 0 | 0 | 0 | 0 | 0 | 0 | 0 | 0 | 0 | 0 | 0 | 0 | 0 | 0 | 0 | 0 | 0 | 0 | 0 | 0 | 0 | 0 | 0 | 0 | 1 |
| Fernandez Serrano | Fernández-S | 2011 | 0 | 0 | 0 | 1 | 1 | 0 | 0 | 0 | 0 | 0 | 0 | 0 | 0 | 0 | 0 | 1 | 0 | 0 | 0 | 0 | 0 | 0 | 0 | 0 | 0 | 0 | 0 | 0 | 0 | 0 | 0 | 0 | 0 | 0 | 0 | 0 | 0 | 0 | 0 | 0 | 0 | 0 | 3 |
| Fuji 2020 | Fuji | 2020 | 0 | 0 | 0 | 0 | 0 | 1 | 0 | 1 | 0 | 0 | 0 | 1 | 1 | 0 | 0 | 0 | 0 | 0 | 0 | 0 | 1 | 0 | 0 | 0 | 0 | 1 | 0 | 1 | 0 | 0 | 0 | 1 | 0 | 0 | 1 | 0 | 0 | 0 | 0 | 1 | 1 | 0 | 11 |
| Ghanei 2021 | Ghanei | 2021 | 0 | 0 | 0 | 0 | 1 | 0 | 0 | 0 | 0 | 0 | 0 | 0 | 0 | 0 | 0 | 0 | 0 | 0 | 0 | 0 | 0 | 0 | 0 | 0 | 0 | 0 | 0 | 0 | 0 | 0 | 0 | 0 | 0 | 0 | 0 | 0 | 0 | 0 | 0 | 0 | 0 | 0 | 1 |
| Gordon 2014 | Gordon | 2014 | 1 | 0 | 1 | 1 | 1 | 0 | 0 | 0 | 0 | 1 | 0 | 0 | 0 | 0 | 1 | 1 | 0 | 0 | 0 | 1 | 0 | 1 | 1 | 1 | 1 | 0 | 0 | 0 | 0 | 0 | 1 | 0 | 1 | 1 | 0 | 0 | 1 | 0 | 0 | 0 | 0 | 1 | 17 |
| Gordon 2016 | Gordon | 2016 | 1 | 1 | 0 | 1 | 1 | 0 | 0 | 0 | 0 | 1 | 0 | 0 | 0 | 0 | 1 | 1 | 0 | 0 | 0 | 1 | 0 | 0 | 1 | 1 | 1 | 0 | 0 | 0 | 0 | 0 | 0 | 0 | 0 | 1 | 0 | 0 | 0 | 0 | 0 | 0 | 0 | 1 | 13 |
| Horby 2021 | Horby | 2021 | 0 | 0 | 0 | 0 | 1 | 0 | 0 | 0 | 0 | 0 | 0 | 0 | 0 | 0 | 0 | 0 | 0 | 0 | 0 | 0 | 0 | 0 | 0 | 0 | 0 | 0 | 0 | 0 | 0 | 0 | 0 | 0 | 0 | 0 | 0 | 0 | 0 | 0 | 0 | 0 | 0 | 0 | 1 |
| Hu 2009 | Hu | 2009 | 0 | 0 | 1 | 1 | 1 | 0 | 0 | 0 | 0 | 1 | 0 | 0 | 0 | 0 | 1 | 1 | 0 | 0 | 0 | 1 | 0 | 1 | 1 | 1 | 1 | 0 | 0 | 0 | 0 | 0 | 0 | 0 | 1 | 1 | 0 | 0 | 0 | 0 | 0 | 0 | 0 | 1 | 14 |
| Huang 2014 | Huang | 2014 | 0 | 0 | 0 | 1 | 1 | 0 | 0 | 0 | 0 | 1 | 0 | 0 | 0 | 0 | 0 | 1 | 0 | 0 | 0 | 0 | 0 | 0 | 0 | 0 | 0 | 0 | 0 | 0 | 0 | 0 | 0 | 0 | 0 | 0 | 0 | 0 | 0 | 0 | 0 | 0 | 0 | 0 | 4 |
| Huh 2007 | Huh | 2007 | 0 | 0 | 1 | 1 | 1 | 0 | 0 | 0 | 0 | 0 | 0 | 0 | 0 | 0 | 0 | 0 | 0 | 0 | 0 | 0 | 0 | 0 | 0 | 0 | 0 | 0 | 0 | 0 | 0 | 0 | 0 | 0 | 0 | 0 | 0 | 0 | 0 | 0 | 0 | 0 | 0 | 0 | 3 |
| Hussein 2021 | Hussein | 2021 | 0 | 0 | 0 | 0 | 0 | 0 | 0 | 1 | 0 | 0 | 0 | 1 | 0 | 0 | 0 | 0 | 0 | 0 | 0 | 0 | 1 | 0 | 0 | 0 | 0 | 0 | 0 | 0 | 0 | 0 | 0 | 1 | 0 | 0 | 0 | 0 | 0 | 0 | 0 | 0 | 0 | 0 | 4 |
| Hwang 2020 | Hwang | 2020 | 0 | 0 | 0 | 0 | 0 | 0 | 0 | 0 | 0 | 0 | 0 | 0 | 1 | 0 | 0 | 0 | 0 | 0 | 0 | 0 | 0 | 0 | 0 | 0 | 0 | 0 | 0 | 0 | 0 | 0 | 0 | 0 | 0 | 0 | 0 | 0 | 0 | 0 | 0 | 1 | 1 | 0 | 3 |
| Hyvernat 2016 | Hyvernat | 2016 | 0 | 1 | 0 | 1 | 1 | 0 | 0 | 0 | 0 | 0 | 0 | 0 | 0 | 0 | 0 | 1 | 0 | 0 | 0 | 0 | 0 | 0 | 0 | 0 | 0 | 0 | 0 | 0 | 0 | 0 | 0 | 0 | 0 | 0 | 0 | 0 | 0 | 0 | 0 | 0 | 0 | 0 | 4 |
| Iglesias 2020 | Iglesias | 2020 | 0 | 0 | 0 | 0 | 1 | 1 | 0 | 1 | 0 | 0 | 0 | 1 | 1 | 0 | 0 | 0 | 0 | 0 | 1 | 0 | 1 | 0 | 0 | 0 | 0 | 1 | 0 | 1 | 0 | 0 | 0 | 0 | 0 | 0 | 1 | 0 | 0 | 0 | 0 | 1 | 1 | 0 | 12 |
| Jamshidi 2021 | Jamshidi | 2021 | 0 | 0 | 0 | 0 | 1 | 0 | 0 | 1 | 0 | 0 | 0 | 1 | 0 | 0 | 0 | 0 | 0 | 0 | 1 | 0 | 0 | 0 | 0 | 0 | 0 | 1 | 0 | 0 | 0 | 0 | 0 | 0 | 0 | 0 | 0 | 0 | 0 | 0 | 0 | 0 | 0 | 0 | 5 |
| Jiang 2019 | Jiang | 2019 | 0 | 0 | 0 | 0 | 1 | 0 | 0 | 0 | 0 | 0 | 0 | 0 | 0 | 0 | 0 | 0 | 0 | 0 | 0 | 0 | 0 | 0 | 0 | 0 | 0 | 0 | 0 | 0 | 0 | 0 | 0 | 0 | 0 | 0 | 0 | 0 | 0 | 0 | 0 | 0 | 0 | 0 | 1 |
| Karimpour 2019 | Karimpour | 2019 | 0 | 0 | 0 | 0 | 0 | 0 | 0 | 0 | 0 | 0 | 0 | 0 | 0 | 0 | 0 | 0 | 0 | 0 | 0 | 0 | 0 | 0 | 0 | 0 | 0 | 0 | 0 | 1 | 0 | 0 | 0 | 0 | 0 | 0 | 0 | 0 | 0 | 0 | 0 | 0 | 0 | 0 | 1 |
| Kaufmann 2008 | Kaufmann | 2008 | 0 | 0 | 0 | 0 | 0 | 0 | 0 | 0 | 0 | 0 | 0 | 0 | 0 | 0 | 0 | 0 | 0 | 0 | 0 | 0 | 0 | 0 | 0 | 0 | 0 | 0 | 0 | 0 | 1 | 0 | 0 | 0 | 0 | 0 | 0 | 0 | 0 | 0 | 0 | 0 | 0 | 0 | 1 |
| Keh 2003 | Keh | 2003 | 0 | 0 | 1 | 1 | 1 | 0 | 0 | 0 | 0 | 1 | 0 | 0 | 0 | 0 | 0 | 1 | 0 | 0 | 0 | 0 | 0 | 0 | 0 | 0 | 0 | 0 | 0 | 0 | 0 | 0 | 0 | 0 | 0 | 0 | 0 | 0 | 0 | 0 | 0 | 0 | 0 | 0 | 5 |
| Keh 2016 | Keh | 2016 | 1 | 1 | 0 | 1 | 1 | 0 | 0 | 0 | 0 | 1 | 0 | 0 | 0 | 0 | 0 | 1 | 1 | 0 | 0 | 0 | 0 | 0 | 1 | 1 | 1 | 0 | 0 | 0 | 1 | 0 | 0 | 0 | 1 | 0 | 0 | 0 | 0 | 0 | 0 | 0 | 0 | 0 | 12 |
| Klastersky 1971 | Klastersky | 1971 | 0 | 0 | 0 | 0 | 0 | 0 | 0 | 0 | 0 | 1 | 0 | 0 | 0 | 0 | 0 | 1 | 0 | 0 | 0 | 0 | 0 | 0 | 0 | 0 | 0 | 0 | 0 | 0 | 0 | 0 | 0 | 0 | 0 | 0 | 0 | 0 | 0 | 0 | 0 | 0 | 0 | 0 | 2 |
| Kurungundla 2008 | Kurungundla | 2008 | 0 | 0 | 0 | 1 | 1 | 0 | 0 | 0 | 0 | 0 | 0 | 0 | 0 | 0 | 0 | 0 | 0 | 0 | 0 | 0 | 0 | 0 | 0 | 0 | 1 | 0 | 0 | 0 | 0 | 0 | 0 | 0 | 0 | 0 | 0 | 0 | 0 | 0 | 0 | 0 | 0 | 0 | 3 |
| Labib 2022 | Labib | 2022 | 0 | 0 | 0 | 0 | 1 | 0 | 0 | 0 | 0 | 0 | 0 | 0 | 0 | 0 | 0 | 0 | 0 | 0 | 0 | 0 | 0 | 0 | 0 | 0 | 0 | 0 | 0 | 0 | 0 | 0 | 0 | 0 | 0 | 0 | 0 | 0 | 0 | 0 | 0 | 0 | 0 | 0 | 1 |
| Li 2016 | Li | 2016 | 0 | 0 | 0 | 1 | 1 | 0 | 0 | 0 | 0 | 0 | 0 | 0 | 0 | 0 | 0 | 0 | 0 | 0 | 0 | 0 | 0 | 0 | 0 | 0 | 0 | 0 | 0 | 0 | 0 | 0 | 0 | 0 | 0 | 0 | 0 | 0 | 0 | 0 | 0 | 0 | 0 | 0 | 2 |
| Liu 2012 | Liu | 2012 | 0 | 0 | 1 | 1 | 1 | 0 | 0 | 0 | 0 | 1 | 0 | 0 | 0 | 0 | 0 | 0 | 0 | 0 | 0 | 0 | 0 | 0 | 1 | 1 | 0 | 0 | 0 | 0 | 0 | 0 | 0 | 0 | 0 | 0 | 0 | 0 | 0 | 0 | 0 | 0 | 0 | 0 | 6 |
| Loisa 2007 | Loisa | 2007 | 0 | 0 | 0 | 1 | 1 | 0 | 0 | 0 | 0 | 0 | 0 | 0 | 0 | 1 | 0 | 1 | 0 | 0 | 0 | 0 | 0 | 0 | 0 | 0 | 0 | 0 | 0 | 0 | 0 | 0 | 0 | 0 | 0 | 0 | 0 | 0 | 0 | 0 | 0 | 0 | 0 | 0 | 4 |
| Lucas 1984 | Lucas | 1984 | 0 | 0 | 0 | 0 | 0 | 0 | 0 | 0 | 0 | 0 | 1 | 0 | 0 | 0 | 0 | 0 | 0 | 0 | 0 | 0 | 0 | 0 | 0 | 0 | 0 | 0 | 0 | 0 | 0 | 0 | 0 | 0 | 0 | 0 | 0 | 0 | 0 | 0 | 0 | 0 | 0 | 0 | 1 |
| Luce 1988 | Luce | 1988 | 0 | 0 | 1 | 1 | 1 | 0 | 0 | 0 | 0 | 1 | 0 | 0 | 0 | 0 | 1 | 1 | 1 | 1 | 0 | 0 | 0 | 0 | 1 | 1 | 0 | 0 | 0 | 0 | 1 | 0 | 0 | 0 | 1 | 0 | 0 | 0 | 0 | 0 | 0 | 0 | 0 | 1 | 13 |
| Lv 2017 | Lv | 2017 | 1 | 1 | 0 | 1 | 1 | 0 | 0 | 0 | 0 | 1 | 0 | 0 | 0 | 0 | 1 | 1 | 1 | 1 | 0 | 1 | 0 | 1 | 1 | 1 | 1 | 0 | 1 | 0 | 1 | 0 | 1 | 0 | 1 | 1 | 0 | 0 | 0 | 0 | 0 | 0 | 0 | 1 | 20 |
| Lyu 2022 | Lyu | 2022 | 0 | 0 | 0 | 0 | 1 | 0 | 0 | 1 | 0 | 0 | 0 | 0 | 0 | 0 | 0 | 0 | 0 | 0 | 1 | 0 | 0 | 0 | 0 | 0 | 0 | 1 | 0 | 0 | 0 | 0 | 0 | 0 | 0 | 0 | 0 | 0 | 0 | 0 | 0 | 0 | 0 | 0 | 4 |
| Mansour 2012 | Mansour | 2012 | 0 | 0 | 0 | 0 | 0 | 0 | 0 | 0 | 1 | 0 | 0 | 0 | 0 | 0 | 0 | 0 | 0 | 0 | 0 | 0 | 0 | 0 | 0 | 0 | 0 | 0 | 0 | 0 | 0 | 0 | 0 | 0 | 0 | 0 | 0 | 0 | 0 | 0 | 1 | 0 | 0 | 0 | 2 |
| McHardy 1972 | McHardy | 1972 | 0 | 0 | 0 | 1 | 1 | 0 | 0 | 0 | 0 | 0 | 0 | 0 | 0 | 0 | 0 | 0 | 0 | 0 | 0 | 0 | 0 | 0 | 0 | 0 | 0 | 0 | 0 | 0 | 0 | 0 | 0 | 0 | 0 | 0 | 0 | 0 | 0 | 0 | 0 | 0 | 0 | 0 | 2 |
| Meduri 2007 | Meduri | 2007 | 0 | 0 | 1 | 1 | 1 | 0 | 0 | 0 | 0 | 1 | 0 | 0 | 0 | 0 | 0 | 1 | 1 | 0 | 0 | 0 | 0 | 0 | 1 | 1 | 0 | 0 | 0 | 0 | 0 | 0 | 0 | 0 | 0 | 0 | 0 | 0 | 0 | 0 | 0 | 0 | 0 | 0 | 8 |
| Meduri 2009 | Meduri | 2009 | 0 | 0 | 0 | 1 | 1 | 0 | 0 | 0 | 0 | 0 | 0 | 0 | 0 | 0 | 0 | 1 | 0 | 0 | 0 | 0 | 0 | 1 | 1 | 1 | 1 | 0 | 0 | 0 | 0 | 0 | 0 | 0 | 0 | 0 | 0 | 0 | 0 | 0 | 0 | 0 | 0 | 0 | 7 |
| Meduri 2022 | Meduri | 2022 | 0 | 0 | 0 | 0 | 1 | 0 | 0 | 0 | 0 | 0 | 0 | 0 | 0 | 0 | 0 | 0 | 0 | 0 | 0 | 0 | 0 | 0 | 0 | 0 | 0 | 0 | 0 | 0 | 0 | 0 | 0 | 0 | 0 | 0 | 0 | 0 | 0 | 0 | 0 | 0 | 0 | 0 | 1 |
| Meijvis 2011 | Meijvis | 2011 | 0 | 0 | 1 | 1 | 1 | 0 | 0 | 0 | 0 | 1 | 0 | 0 | 0 | 0 | 0 | 1 | 1 | 0 | 0 | 0 | 0 | 0 | 1 | 1 | 0 | 0 | 0 | 0 | 0 | 0 | 0 | 0 | 0 | 0 | 0 | 0 | 0 | 0 | 0 | 0 | 0 | 0 | 8 |
| Menon 2017 | Menon | 2017 | 0 | 0 | 0 | 1 | 1 | 0 | 0 | 0 | 1 | 0 | 0 | 0 | 0 | 0 | 0 | 0 | 0 | 0 | 0 | 0 | 0 | 0 | 1 | 1 | 0 | 0 | 0 | 0 | 0 | 0 | 1 | 0 | 0 | 0 | 0 | 0 | 0 | 0 | 1 | 0 | 0 | 0 | 7 |
| Min 1975 | Min | 1975 | 0 | 0 | 0 | 0 | 0 | 0 | 0 | 0 | 1 | 0 | 0 | 0 | 0 | 0 | 0 | 0 | 0 | 0 | 0 | 0 | 0 | 0 | 0 | 0 | 0 | 0 | 0 | 0 | 0 | 0 | 0 | 0 | 0 | 0 | 0 | 0 | 0 | 0 | 1 | 0 | 0 | 0 | 2 |
| Mirea 2014 | Mirea | 2014 | 0 | 0 | 0 | 1 | 1 | 0 | 0 | 0 | 0 | 0 | 0 | 0 | 0 | 0 | 0 | 1 | 1 | 0 | 0 | 0 | 0 | 0 | 1 | 1 | 1 | 0 | 0 | 0 | 0 | 0 | 0 | 0 | 0 | 0 | 0 | 0 | 0 | 0 | 0 | 0 | 0 | 0 | 7 |
| Mohamed 2020 | Mohamed | 2020 | 0 | 0 | 0 | 0 | 1 | 1 | 0 | 1 | 0 | 0 | 0 | 1 | 0 | 0 | 0 | 0 | 0 | 0 | 1 | 0 | 1 | 0 | 0 | 0 | 0 | 1 | 0 | 1 | 0 | 0 | 0 | 0 | 0 | 0 | 1 | 0 | 0 | 0 | 0 | 1 | 0 | 0 | 10 |
| Mohamed 2023 | Mohamed | 2023 | 0 | 0 | 0 | 0 | 1 | 0 | 0 | 1 | 0 | 0 | 0 | 0 | 0 | 0 | 0 | 0 | 0 | 0 | 0 | 0 | 0 | 0 | 0 | 0 | 0 | 0 | 0 | 0 | 0 | 0 | 0 | 0 | 0 | 0 | 0 | 0 | 0 | 0 | 0 | 0 | 0 | 0 | 2 |
| Moreno 2011 | Moreno | 2011 | 0 | 0 | 0 | 0 | 0 | 0 | 0 | 0 | 0 | 0 | 0 | 0 | 0 | 0 | 0 | 0 | 0 | 0 | 0 | 0 | 0 | 0 | 0 | 0 | 0 | 0 | 0 | 0 | 1 | 0 | 0 | 0 | 0 | 0 | 0 | 0 | 0 | 0 | 0 | 0 | 0 | 0 | 1 |
| Moskowitz 2020 | Moskowitz | 2020 | 0 | 0 | 0 | 0 | 1 | 1 | 0 | 1 | 0 | 0 | 0 | 1 | 1 | 0 | 0 | 0 | 0 | 0 | 1 | 0 | 1 | 0 | 0 | 0 | 0 | 1 | 0 | 1 | 0 | 0 | 0 | 0 | 0 | 0 | 1 | 0 | 0 | 0 | 0 | 1 | 1 | 0 | 12 |
| Mussack 2005 | Mussack | 2005 | 0 | 0 | 0 | 0 | 0 | 0 | 0 | 0 | 0 | 0 | 0 | 0 | 0 | 0 | 1 | 0 | 0 | 0 | 0 | 0 | 0 | 0 | 0 | 0 | 0 | 0 | 0 | 0 | 0 | 0 | 0 | 0 | 0 | 1 | 0 | 0 | 0 | 0 | 0 | 0 | 0 | 0 | 2 |
| Nafae 2013 | Nafae | 2013 | 0 | 0 | 0 | 1 | 1 | 0 | 0 | 0 | 0 | 0 | 0 | 0 | 0 | 0 | 0 | 1 | 0 | 0 | 0 | 0 | 0 | 0 | 0 | 0 | 0 | 0 | 0 | 0 | 0 | 0 | 0 | 0 | 0 | 0 | 0 | 0 | 0 | 0 | 0 | 0 | 0 | 0 | 3 |
| Nagy 2013 | Nagy | 2013 | 0 | 0 | 0 | 1 | 1 | 0 | 0 | 0 | 1 | 0 | 0 | 0 | 0 | 0 | 0 | 0 | 0 | 0 | 0 | 0 | 0 | 0 | 0 | 0 | 0 | 0 | 0 | 0 | 0 | 0 | 0 | 0 | 0 | 0 | 0 | 0 | 0 | 0 | 0 | 0 | 0 | 0 | 3 |
| Ngaosuwan 2018 | Ngaosuwan | 2018 | 0 | 0 | 0 | 1 | 1 | 0 | 0 | 0 | 0 | 0 | 0 | 0 | 0 | 0 | 0 | 0 | 0 | 0 | 0 | 0 | 0 | 0 | 0 | 0 | 0 | 0 | 0 | 0 | 0 | 0 | 0 | 0 | 0 | 0 | 0 | 0 | 0 | 0 | 0 | 0 | 0 | 0 | 2 |
| Oppert 2005 | Oppert | 2005 | 1 | 0 | 1 | 1 | 1 | 0 | 0 | 0 | 0 | 1 | 0 | 0 | 0 | 0 | 1 | 1 | 1 | 1 | 0 | 1 | 0 | 1 | 1 | 1 | 1 | 0 | 0 | 0 | 1 | 0 | 0 | 0 | 0 | 1 | 0 | 0 | 1 | 0 | 0 | 0 | 0 | 1 | 18 |
| Pierre de Oliveira 20 | Pierre de Oli | 2023 | 0 | 0 | 0 | 0 | 1 | 0 | 0 | 0 | 0 | 0 | 0 | 0 | 0 | 0 | 0 | 0 | 0 | 0 | 0 | 0 | 0 | 0 | 0 | 0 | 0 | 0 | 0 | 0 | 0 | 0 | 0 | 0 | 0 | 0 | 0 | 0 | 0 | 0 | 0 | 0 | 0 | 0 | 1 |
| Pongpanich 1973 | Pongpanich | 1973 | 0 | 0 | 0 | 0 | 0 | 0 | 0 | 0 | 1 | 0 | 0 | 0 | 0 | 0 | 0 | 0 | 0 | 0 | 0 | 0 | 0 | 0 | 0 | 0 | 0 | 0 | 0 | 0 | 0 | 0 | 0 | 0 | 0 | 0 | 0 | 0 | 0 | 0 | 1 | 0 | 0 | 0 | 2 |
| Povoa 2015 | Póvoa | 2015 | 0 | 0 | 0 | 0 | 0 | 0 | 0 | 0 | 0 | 0 | 0 | 0 | 0 | 0 | 1 | 0 | 0 | 0 | 0 | 0 | 0 | 0 | 0 | 0 | 0 | 0 | 0 | 0 | 0 | 0 | 0 | 0 | 1 | 0 | 0 | 0 | 0 | 0 | 0 | 0 | 0 | 0 | 2 |
| Raghu 2021 | Raghu | 2021 | 0 | 0 | 0 | 0 | 0 | 0 | 0 | 1 | 0 | 0 | 0 | 0 | 0 | 0 | 0 | 0 | 0 | 0 | 0 | 0 | 0 | 0 | 0 | 0 | 0 | 0 | 0 | 0 | 0 | 0 | 0 | 0 | 0 | 0 | 0 | 0 | 0 | 0 | 0 | 0 | 0 | 0 | 1 |
| Ram 2022 | Ram | 2022 | 0 | 0 | 0 | 0 | 1 | 0 | 0 | 0 | 0 | 0 | 0 | 0 | 0 | 1 | 0 | 0 | 0 | 0 | 0 | 0 | 0 | 0 | 0 | 0 | 0 | 0 | 0 | 0 | 0 | 0 | 0 | 0 | 0 | 0 | 0 | 0 | 0 | 0 | 0 | 0 | 0 | 0 | 2 |
| Reddy 2020 | Reddy | 2020 | 0 | 0 | 0 | 0 | 0 | 1 | 0 | 0 | 0 | 0 | 0 | 0 | 0 | 0 | 0 | 0 | 0 | 0 | 0 | 0 | 1 | 0 | 0 | 0 | 0 | 1 | 0 | 0 | 0 | 0 | 0 | 1 | 0 | 0 | 0 | 0 | 0 | 0 | 0 | 0 | 0 | 0 | 4 |
| Rezk 2013 | Rezk | 2013 | 0 | 0 | 1 | 1 | 1 | 0 | 0 | 0 | 0 | 1 | 0 | 0 | 0 | 0 | 0 | 1 | 1 | 0 | 0 | 0 | 0 | 0 | 1 | 1 | 0 | 0 | 0 | 0 | 0 | 0 | 0 | 0 | 0 | 0 | 0 | 0 | 0 | 0 | 0 | 0 | 0 | 0 | 8 |
| Rinaldi 2006 | Rinaldi | 2006 | 1 | 0 | 1 | 1 | 1 | 0 | 0 | 0 | 0 | 1 | 0 | 0 | 0 | 0 | 0 | 1 | 1 | 0 | 0 | 0 | 0 | 1 | 1 | 1 | 0 | 0 | 0 | 0 | 0 | 0 | 0 | 0 | 0 | 0 | 0 | 0 | 0 | 0 | 0 | 0 | 0 | 0 | 10 |
| Sabry 2011 | Sabry | 2011 | 0 | 0 | 1 | 1 | 1 | 0 | 0 | 0 | 0 | 1 | 0 | 0 | 0 | 0 | 0 | 1 | 1 | 0 | 0 | 0 | 0 | 0 | 1 | 1 | 0 | 0 | 0 | 0 | 0 | 0 | 0 | 0 | 0 | 0 | 0 | 0 | 0 | 0 | 0 | 0 | 0 | 0 | 8 |
| Sai 2021 | Sai | 2021 | 0 | 0 | 0 | 0 | 1 | 0 | 0 | 0 | 0 | 0 | 0 | 0 | 0 | 0 | 0 | 0 | 0 | 0 | 0 | 0 | 0 | 0 | 0 | 0 | 0 | 0 | 0 | 0 | 0 | 0 | 0 | 0 | 0 | 0 | 0 | 0 | 0 | 0 | 0 | 0 | 0 | 0 | 1 |
| Sang 2020 | Sang | 2020 | 0 | 0 | 0 | 0 | 0 | 0 | 0 | 1 | 0 | 0 | 0 | 0 | 0 | 0 | 0 | 0 | 0 | 0 | 0 | 0 | 0 | 0 | 0 | 0 | 0 | 0 | 0 | 0 | 0 | 0 | 0 | 0 | 0 | 0 | 0 | 0 | 0 | 0 | 0 | 0 | 0 | 0 | 1 |
| Schelling 2001 | Schelling | 2001 | 0 | 0 | 0 | 0 | 0 | 0 | 0 | 0 | 0 | 0 | 0 | 0 | 0 | 0 | 0 | 0 | 0 | 0 | 0 | 0 | 0 | 0 | 0 | 0 | 0 | 0 | 0 | 0 | 1 | 0 | 0 | 0 | 0 | 0 | 0 | 0 | 0 | 0 | 0 | 0 | 0 | 0 | 1 |
| Schumer 1976 | Schumer | 1976 | 0 | 0 | 1 | 1 | 1 | 0 | 0 | 0 | 0 | 1 | 1 | 0 | 0 | 0 | 0 | 1 | 1 | 0 | 0 | 0 | 0 | 0 | 1 | 1 | 0 | 0 | 0 | 0 | 1 | 0 | 0 | 0 | 1 | 0 | 0 | 0 | 0 | 0 | 0 | 0 | 0 | 1 | 12 |
| Sevransky 2021 | Sevransky | 2021 | 0 | 0 | 0 | 0 | 1 | 1 | 0 | 1 | 0 | 0 | 0 | 1 | 1 | 0 | 0 | 0 | 0 | 0 | 1 | 0 | 1 | 0 | 0 | 0 | 0 | 1 | 0 | 1 | 0 | 0 | 0 | 0 | 0 | 0 | 0 | 0 | 0 | 0 | 0 | 1 | 0 | 0 | 10 |
| Shi 2019 | Shi | 2019 | 0 | 0 | 0 | 0 | 0 | 0 | 0 | 0 | 1 | 0 | 0 | 0 | 0 | 0 | 0 | 0 | 0 | 0 | 0 | 0 | 0 | 0 | 0 | 0 | 0 | 0 | 0 | 0 | 0 | 0 | 0 | 0 | 0 | 0 | 0 | 0 | 0 | 0 | 0 | 0 | 0 | 0 | 1 |
| Slusher 1996 | Slusher | 1996 | 0 | 0 | 1 | 1 | 1 | 0 | 0 | 0 | 1 | 0 | 0 | 0 | 0 | 0 | 0 | 0 | 0 | 0 | 0 | 0 | 0 | 0 | 1 | 1 | 0 | 0 | 0 | 0 | 0 | 0 | 0 | 0 | 0 | 0 | 0 | 0 | 0 | 0 | 1 | 0 | 0 | 0 | 7 |
| Snijders 2010 | Snijders | 2010 | 0 | 0 | 1 | 1 | 1 | 0 | 0 | 0 | 0 | 1 | 0 | 0 | 0 | 0 | 0 | 1 | 1 | 0 | 0 | 0 | 0 | 0 | 1 | 1 | 0 | 0 | 0 | 0 | 0 | 0 | 0 | 0 | 0 | 0 | 0 | 0 | 0 | 0 | 0 | 0 | 0 | 0 | 8 |
| Sprung 1984 | Sprung | 1984 | 0 | 0 | 1 | 1 | 1 | 0 | 0 | 0 | 0 | 1 | 0 | 0 | 0 | 0 | 1 | 1 | 1 | 0 | 0 | 0 | 0 | 0 | 1 | 1 | 0 | 0 | 0 | 0 | 0 | 0 | 0 | 0 | 1 | 0 | 0 | 0 | 0 | 0 | 0 | 0 | 0 | 1 | 11 |
| Sprung 2008 | Sprung | 2008 | 1 | 0 | 1 | 1 | 1 | 0 | 0 | 0 | 0 | 1 | 1 | 0 | 0 | 0 | 1 | 1 | 1 | 1 | 0 | 1 | 0 | 1 | 1 | 1 | 1 | 0 | 1 | 0 | 1 | 0 | 0 | 0 | 1 | 1 | 0 | 0 | 1 | 0 | 0 | 0 | 0 | 1 | 21 |
| Sui 2013 | Sui | 2013 | 0 | 0 | 0 | 1 | 1 | 0 | 0 | 0 | 0 | 0 | 0 | 0 | 0 | 0 | 0 | 0 | 0 | 0 | 0 | 0 | 0 | 0 | 0 | 0 | 0 | 0 | 0 | 0 | 0 | 0 | 0 | 0 | 0 | 0 | 0 | 0 | 0 | 0 | 0 | 0 | 0 | 0 | 2 |
| Sumarmo 1982 | Sumarmo | 1982 | 0 | 0 | 0 | 0 | 0 | 0 | 0 | 0 | 1 | 0 | 0 | 0 | 0 | 0 | 0 | 0 | 0 | 0 | 0 | 0 | 0 | 0 | 0 | 0 | 0 | 0 | 0 | 0 | 0 | 0 | 0 | 0 | 0 | 0 | 0 | 0 | 0 | 0 | 1 | 0 | 0 | 0 | 2 |
| Tagaro 2017 | Tagaro | 2017 | 0 | 0 | 0 | 1 | 1 | 0 | 0 | 0 | 0 | 0 | 0 | 0 | 0 | 0 | 0 | 0 | 0 | 0 | 0 | 0 | 0 | 0 | 0 | 0 | 0 | 0 | 0 | 0 | 0 | 0 | 0 | 0 | 0 | 0 | 0 | 0 | 0 | 0 | 0 | 0 | 0 | 0 | 2 |
| Tandan 2005 | Tandan | 2005 | 0 | 0 | 1 | 1 | 1 | 0 | 0 | 0 | 0 | 1 | 0 | 0 | 0 | 0 | 0 | 1 | 1 | 0 | 0 | 0 | 0 | 0 | 1 | 1 | 1 | 0 | 0 | 0 | 0 | 0 | 0 | 0 | 0 | 0 | 0 | 0 | 0 | 0 | 0 | 0 | 0 | 0 | 9 |
| Tassniyom 1993 | Tassniyom | 1993 | 0 | 0 | 0 | 0 | 0 | 0 | 0 | 0 | 1 | 0 | 0 | 0 | 0 | 0 | 0 | 0 | 0 | 0 | 0 | 0 | 0 | 0 | 0 | 0 | 0 | 0 | 0 | 0 | 0 | 0 | 0 | 0 | 0 | 0 | 0 | 0 | 0 | 0 | 1 | 0 | 0 | 0 | 2 |
| Tilouche 2019 | Tilouche | 2019 | 0 | 0 | 0 | 1 | 1 | 0 | 0 | 0 | 0 | 0 | 0 | 0 | 0 | 1 | 0 | 1 | 0 | 0 | 0 | 0 | 0 | 0 | 0 | 0 | 0 | 0 | 0 | 0 | 0 | 0 | 0 | 0 | 0 | 0 | 0 | 0 | 0 | 0 | 0 | 0 | 0 | 0 | 4 |
| Tomazini 2020 | Tomazini | 2020 | 0 | 0 | 0 | 0 | 1 | 0 | 0 | 0 | 0 | 0 | 0 | 0 | 0 | 0 | 0 | 1 | 0 | 0 | 0 | 0 | 0 | 0 | 0 | 0 | 0 | 0 | 0 | 0 | 0 | 0 | 0 | 0 | 0 | 0 | 0 | 0 | 0 | 0 | 0 | 0 | 0 | 0 | 2 |
| Tongyoo 2016 | Tongyoo | 2016 | 0 | 1 | 0 | 1 | 1 | 0 | 0 | 0 | 0 | 1 | 0 | 0 | 0 | 0 | 1 | 1 | 1 | 0 | 0 | 1 | 0 | 1 | 1 | 1 | 1 | 0 | 0 | 0 | 1 | 0 | 0 | 0 | 0 | 1 | 0 | 0 | 0 | 0 | 0 | 0 | 0 | 0 | 14 |
| Torres 2015 | Torres | 2015 | 0 | 0 | 1 | 1 | 1 | 0 | 0 | 0 | 0 | 1 | 0 | 0 | 0 | 0 | 0 | 1 | 1 | 0 | 0 | 0 | 0 | 0 | 1 | 1 | 1 | 0 | 0 | 0 | 0 | 0 | 1 | 0 | 0 | 0 | 0 | 0 | 0 | 0 | 0 | 0 | 0 | 0 | 10 |
| Valoor 2009 | Valoor | 2009 | 0 | 0 | 1 | 1 | 1 | 0 | 0 | 0 | 1 | 0 | 0 | 0 | 0 | 0 | 0 | 0 | 0 | 0 | 0 | 0 | 0 | 0 | 1 | 1 | 0 | 0 | 0 | 0 | 0 | 0 | 0 | 0 | 0 | 0 | 0 | 0 | 0 | 0 | 1 | 0 | 0 | 0 | 7 |
| VASSCSG 1987 | VASSCSG | 1987 | 0 | 0 | 1 | 1 | 1 | 0 | 0 | 0 | 0 | 1 | 0 | 0 | 0 | 0 | 0 | 1 | 1 | 0 | 0 | 0 | 0 | 0 | 1 | 1 | 0 | 0 | 0 | 0 | 1 | 0 | 0 | 0 | 0 | 0 | 0 | 0 | 0 | 0 | 0 | 0 | 0 | 0 | 9 |
| Venkatesh 2018 | Venkatesh | 2018 | 1 | 1 | 0 | 1 | 1 | 0 | 0 | 0 | 0 | 1 | 0 | 0 | 0 | 0 | 1 | 1 | 1 | 0 | 0 | 1 | 0 | 1 | 1 | 1 | 1 | 0 | 1 | 0 | 1 | 0 | 1 | 0 | 1 | 1 | 0 | 0 | 0 | 0 | 0 | 0 | 0 | 1 | 19 |
| Villar 2020 | Villar | 2020 | 0 | 0 | 0 | 0 | 1 | 0 | 0 | 0 | 0 | 0 | 0 | 0 | 0 | 0 | 0 | 0 | 0 | 0 | 0 | 0 | 0 | 0 | 0 | 0 | 0 | 0 | 0 | 0 | 0 | 0 | 0 | 0 | 0 | 0 | 0 | 0 | 0 | 0 | 0 | 0 | 0 | 0 | 1 |
| Wan 2014 | Wan | 2014 | 0 | 0 | 0 | 0 | 0 | 0 | 0 | 0 | 0 | 0 | 0 | 0 | 0 | 0 | 0 | 0 | 0 | 1 | 0 | 0 | 0 | 0 | 0 | 0 | 0 | 0 | 0 | 0 | 0 | 0 | 0 | 0 | 0 | 0 | 0 | 0 | 1 | 0 | 0 | 0 | 0 | 0 | 2 |
| Wang 2022 | Wang | 2022 | 0 | 0 | 0 | 0 | 0 | 0 | 0 | 0 | 1 | 0 | 0 | 0 | 0 | 0 | 0 | 0 | 0 | 0 | 0 | 0 | 0 | 0 | 0 | 0 | 0 | 0 | 0 | 0 | 0 | 0 | 0 | 0 | 0 | 0 | 0 | 0 | 0 | 0 | 0 | 0 | 0 | 0 | 1 |
| Wang 2023 | Wang | 2023 | 0 | 0 | 0 | 0 | 0 | 0 | 0 | 1 | 0 | 0 | 0 | 0 | 0 | 0 | 0 | 0 | 0 | 0 | 0 | 0 | 0 | 0 | 0 | 0 | 0 | 0 | 0 | 0 | 0 | 0 | 0 | 0 | 0 | 0 | 0 | 0 | 0 | 0 | 0 | 0 | 0 | 0 | 1 |
| Wani 2020 | Wani | 2020 | 0 | 0 | 0 | 0 | 1 | 1 | 0 | 1 | 0 | 0 | 0 | 1 | 0 | 0 | 0 | 0 | 0 | 0 | 1 | 0 | 1 | 0 | 0 | 0 | 0 | 1 | 0 | 1 | 0 | 0 | 0 | 0 | 0 | 0 | 1 | 0 | 0 | 0 | 0 | 1 | 1 | 0 | 11 |
| Wen 2004 | Wen | 2004 | 0 | 0 | 0 | 0 | 0 | 0 | 0 | 0 | 1 | 0 | 0 | 0 | 0 | 0 | 0 | 0 | 0 | 0 | 0 | 0 | 0 | 0 | 0 | 0 | 0 | 0 | 0 | 0 | 0 | 0 | 0 | 0 | 0 | 0 | 0 | 0 | 0 | 0 | 0 | 0 | 0 | 0 | 1 |
| Wittermans 2020 | Wittermans | 2020 | 0 | 0 | 0 | 0 | 1 | 0 | 0 | 0 | 0 | 0 | 0 | 0 | 0 | 0 | 0 | 0 | 0 | 0 | 0 | 0 | 0 | 0 | 0 | 0 | 0 | 0 | 0 | 0 | 0 | 0 | 0 | 0 | 0 | 0 | 0 | 0 | 0 | 0 | 0 | 0 | 0 | 0 | 1 |
| Yildiz 2002 | Yildiz | 2002 | 0 | 0 | 1 | 1 | 1 | 0 | 0 | 0 | 0 | 1 | 0 | 0 | 0 | 0 | 0 | 1 | 1 | 0 | 0 | 0 | 0 | 1 | 1 | 1 | 0 | 0 | 0 | 0 | 1 | 0 | 1 | 0 | 0 | 0 | 0 | 0 | 0 | 0 | 0 | 0 | 0 | 0 | 11 |
| Yildiz 2011 | Yildiz | 2011 | 0 | 0 | 1 | 1 | 1 | 0 | 0 | 0 | 0 | 1 | 0 | 0 | 0 | 0 | 0 | 1 | 1 | 0 | 0 | 0 | 0 | 1 | 1 | 1 | 0 | 0 | 0 | 0 | 1 | 0 | 0 | 0 | 0 | 0 | 0 | 0 | 0 | 0 | 0 | 0 | 0 | 0 | 10 |
| Yuan 2010 | Yuan | 2010 | 0 | 0 | 0 | 0 | 0 | 0 | 0 | 0 | 1 | 0 | 0 | 0 | 0 | 0 | 0 | 0 | 0 | 0 | 0 | 0 | 0 | 0 | 0 | 0 | 0 | 0 | 0 | 0 | 0 | 0 | 0 | 0 | 0 | 0 | 0 | 0 | 0 | 0 | 0 | 0 | 0 | 0 | 1 |
| Zeng 2014 | Zeng | 2014 | 0 | 0 | 0 | 0 | 0 | 0 | 0 | 0 | 1 | 0 | 0 | 0 | 0 | 0 | 0 | 0 | 0 | 0 | 0 | 0 | 0 | 0 | 0 | 0 | 0 | 0 | 0 | 0 | 0 | 0 | 0 | 0 | 0 | 0 | 0 | 0 | 0 | 0 | 0 | 0 | 0 | 0 | 1 |
| Zhou 2015 | Zhou | 2015 | 0 | 0 | 0 | 1 | 1 | 0 | 0 | 0 | 0 | 0 | 0 | 0 | 0 | 0 | 0 | 0 | 0 | 0 | 0 | 0 | 0 | 0 | 0 | 0 | 0 | 0 | 0 | 0 | 0 | 0 | 0 | 0 | 0 | 0 | 0 | 0 | 0 | 0 | 0 | 0 | 0 | 0 | 2 |
| Number of included RCTs per SR | | | 10 | 7 | 33 | 61 | 87 | 8 | 2 | 15 | 15 | 37 | 5 | 9 | 6 | 3 | 21 | 50 | 30 | 9 | 8 | 13 | 10 | 19 | 45 | 42 | 23 | 11 | 5 | 8 | 22 | 0 | 9 | 3 | 18 | 12 | 6 | 3 | 9 | 3 | 10 | 8 | 7 | 17 | |

#### Supplementary Table S9: Jaccard similarity and distance: pairwise comparison of study pools

| SR_1 | SR_2 | Intersect RCTs | Unio RCTs | Jaccard (J) | Distance (1-J) |
|---|---|---|---|---|---|
| Wu 2024 | Yamamoto 2020 | 3 | 3 | 1 | 0 |
| Pitre 2024 | Rochwerg 2018 | 42 | 45 | 0,933333333 | 0,066666667 |
| Assouline 2021 | Na 2021 | 8 | 10 | 0,8 | 0,2 |
| Lyu 2020 | Wu 2020 | 11 | 14 | 0,785714286 | 0,214285714 |
| Assouline 2021 | Somagutta 2021 | 7 | 9 | 0,777777778 | 0,222222222 |
| Assouline 2021 | Yao 2021 | 7 | 9 | 0,777777778 | 0,222222222 |
| Somagutta 2021 | Yao 2021 | 7 | 9 | 0,777777778 | 0,222222222 |
| Fang 2019 | Lin 2019 | 29 | 38 | 0,763157895 | 0,236842105 |
| Na 2021 | Saghafi 2024 | 9 | 12 | 0,75 | 0,25 |
| Assouline 2021 | Wu 2021 | 6 | 8 | 0,75 | 0,25 |
| Somagutta 2021 | Wu 2021 | 6 | 8 | 0,75 | 0,25 |
| Wu 2021 | Yao 2021 | 6 | 8 | 0,75 | 0,25 |
| Kato 2023 | Na 2021 | 8 | 11 | 0,727272727 | 0,272727273 |
| Assouline 2021 | Saghafi 2024 | 8 | 11 | 0,727272727 | 0,272727273 |
| Lu 2023 | Saghafi 2024 | 8 | 11 | 0,727272727 | 0,272727273 |
| Fang 2019 | Rochwerg 2018 | 33 | 46 | 0,717391304 | 0,282608696 |
| Lin 2019 | Rochwerg 2018 | 30 | 42 | 0,714285714 | 0,285714286 |
| Annane 2019 | Annane 2025 | 61 | 87 | 0,701149425 | 0,298850575 |
| Assouline 2021 | Kato 2023 | 7 | 10 | 0,7 | 0,3 |
| Kato 2023 | Lu 2023 | 7 | 10 | 0,7 | 0,3 |
| Kato 2023 | Somagutta 2021 | 7 | 10 | 0,7 | 0,3 |
| Kato 2023 | Yao 2021 | 7 | 10 | 0,7 | 0,3 |
| Lian 2019 | Wen 2019 | 16 | 23 | 0,695652174 | 0,304347826 |
| Fang 2019 | Pitre 2024 | 33 | 49 | 0,673469388 | 0,326530612 |
| Fang 2019 | Liang 2021 | 35 | 52 | 0,673076923 | 0,326923077 |
| Annane 2015 | Fang 2019 | 28 | 42 | 0,666666667 | 0,333333333 |
| Lin 2019 | Pitre 2024 | 30 | 45 | 0,666666667 | 0,333333333 |
| Annane 2015 | Rochwerg 2018 | 30 | 45 | 0,666666667 | 0,333333333 |
| Kato 2023 | Saghafi 2024 | 8 | 12 | 0,666666667 | 0,333333333 |
| Kato 2023 | Wu 2021 | 6 | 9 | 0,666666667 | 0,333333333 |
| Barakakis 2019 | Wu 2024 | 2 | 3 | 0,666666667 | 0,333333333 |
| Barakakis 2019 | Yamamoto 2020 | 2 | 3 | 0,666666667 | 0,333333333 |
| Yao 2021 | Zayed 2022 | 6 | 9 | 0,666666667 | 0,333333333 |
| Lyu 2020 | Zhou 2018 | 12 | 18 | 0,666666667 | 0,333333333 |
| Wen 2019 | Zhou 2018 | 14 | 21 | 0,666666667 | 0,333333333 |
| Annane 2019 | Liang 2021 | 44 | 67 | 0,656716418 | 0,343283582 |
| Lian 2019 | Zhou 2018 | 15 | 23 | 0,652173913 | 0,347826087 |
| Na 2021 | Somagutta 2021 | 7 | 11 | 0,636363636 | 0,363636364 |
| Na 2021 | Yao 2021 | 7 | 11 | 0,636363636 | 0,363636364 |
| Annane 2019 | Rochwerg 2018 | 40 | 63 | 0,634920635 | 0,365079365 |
| Annane 2019 | Pitre 2024 | 41 | 65 | 0,630769231 | 0,369230769 |

| | | | | | |
|---|---|---|---|---|---|
| Annane 2015 | Pitre 2024 | 30 | 48 | 0,625 | 0,375 |
| Lee 2022 | Zayed 2022 | 5 | 8 | 0,625 | 0,375 |
| Wu 2021 | Zayed 2022 | 5 | 8 | 0,625 | 0,375 |
| Lian 2019 | Lyu 2020 | 13 | 21 | 0,619047619 | 0,380952381 |
| Annane 2015 | Lin 2019 | 24 | 39 | 0,615384615 | 0,384615385 |
| Deng 2024 | Kato 2023 | 9 | 15 | 0,6 | 0,4 |
| Assouline 2021 | Lu 2023 | 6 | 10 | 0,6 | 0,4 |
| Lyu 2020 | Ni 2019 | 12 | 20 | 0,6 | 0,4 |
| Lu 2023 | Somagutta 2021 | 6 | 10 | 0,6 | 0,4 |
| Na 2021 | Wu 2021 | 6 | 10 | 0,6 | 0,4 |
| Lu 2023 | Yao 2021 | 6 | 10 | 0,6 | 0,4 |
| Saghafi 2024 | Somagutta 2021 | 7 | 12 | 0,583333333 | 0,416666667 |
| Saghafi 2024 | Yao 2021 | 7 | 12 | 0,583333333 | 0,416666667 |
| Annane 2019 | Fang 2019 | 36 | 62 | 0,580645161 | 0,419354839 |
| Lian 2019 | Wu 2020 | 12 | 21 | 0,571428571 | 0,428571429 |
| Liang 2021 | Lin 2019 | 29 | 51 | 0,568627451 | 0,431372549 |
| Lyu 2020 | Rygard 2018 | 13 | 23 | 0,565217391 | 0,434782609 |
| Deng 2025 | Yang 2020 | 9 | 16 | 0,5625 | 0,4375 |
| Liang 2021 | Rochwerg 2018 | 33 | 59 | 0,559322034 | 0,440677966 |
| Liang 2021 | Pitre 2024 | 34 | 61 | 0,557377049 | 0,442622951 |
| Lu 2023 | Wu 2021 | 5 | 9 | 0,555555556 | 0,444444444 |
| Lee 2022 | Yao 2021 | 5 | 9 | 0,555555556 | 0,444444444 |
| Saghafi 2024 | Wu 2021 | 6 | 11 | 0,545454545 | 0,454545455 |
| Na 2021 | Zayed 2022 | 6 | 11 | 0,545454545 | 0,454545455 |
| Annane 2015 | Annane 2019 | 33 | 61 | 0,540983607 | 0,459016393 |
| Rygard 2018 | Zhou 2018 | 14 | 26 | 0,538461538 | 0,461538462 |
| Deng 2024 | Lu 2023 | 8 | 15 | 0,533333333 | 0,466666667 |
| Aletreby 2019 | Lyu 2020 | 8 | 15 | 0,533333333 | 0,466666667 |
| Deng 2024 | Saghafi 2024 | 9 | 17 | 0,529411765 | 0,470588235 |
| Lin 2019 | Son 2021 | 18 | 34 | 0,529411765 | 0,470588235 |
| Wu 2020 | Zhou 2018 | 10 | 19 | 0,526315789 | 0,473684211 |
| Annane 2025 | Liang 2021 | 47 | 90 | 0,522222222 | 0,477777778 |
| Ni 2019 | Son 2021 | 14 | 27 | 0,518518519 | 0,481481481 |
| Lian 2019 | Rygard 2018 | 15 | 29 | 0,517241379 | 0,482758621 |
| Lu 2023 | Na 2021 | 6 | 12 | 0,5 | 0,5 |
| Ni 2019 | Rygard 2018 | 14 | 28 | 0,5 | 0,5 |
| Aletreby 2019 | Siddiqi 2019 | 5 | 10 | 0,5 | 0,5 |
| Lu 2021 | Xu 2018 | 6 | 12 | 0,5 | 0,5 |
| Assouline 2021 | Zayed 2022 | 5 | 10 | 0,5 | 0,5 |
| Saghafi 2024 | Zayed 2022 | 6 | 12 | 0,5 | 0,5 |
| Somagutta 2021 | Zayed 2022 | 5 | 10 | 0,5 | 0,5 |
| Ni 2019 | Zhou 2018 | 12 | 24 | 0,5 | 0,5 |
| Annane 2019 | Lin 2019 | 30 | 61 | 0,491803279 | 0,508196721 |
| Lin 2019 | Ni 2019 | 16 | 33 | 0,484848485 | 0,515151515 |
| Annane 2025 | Pitre 2024 | 43 | 89 | 0,483146067 | 0,516853933 |

| | | | | | |
|---|---|---|---|---|---|
| Annane 2015 | Liang 2021 | 27 | 56 | 0,482142857 | 0,517857143 |
| Lian 2019 | Ni 2019 | 13 | 27 | 0,481481481 | 0,518518519 |
| Ni 2019 | Wen 2019 | 12 | 25 | 0,48 | 0,52 |
| Lyu 2020 | Wen 2019 | 10 | 21 | 0,476190476 | 0,523809524 |
| Ni 2019 | Wu 2020 | 10 | 21 | 0,476190476 | 0,523809524 |
| Fang 2019 | Ni 2019 | 18 | 38 | 0,473684211 | 0,526315789 |
| Deng 2024 | Na 2021 | 8 | 17 | 0,470588235 | 0,529411765 |
| Lyu 2020 | Xu 2018 | 7 | 15 | 0,466666667 | 0,533333333 |
| Fang 2019 | Zhou 2018 | 17 | 37 | 0,459459459 | 0,540540541 |
| Rygard 2018 | Wu 2020 | 11 | 24 | 0,458333333 | 0,541666667 |
| Lee 2022 | Na 2021 | 5 | 11 | 0,454545455 | 0,545454545 |
| Lin 2019 | Wen 2019 | 15 | 33 | 0,454545455 | 0,545454545 |
| Kato 2023 | Zayed 2022 | 5 | 11 | 0,454545455 | 0,545454545 |
| Ni 2019 | Rochwerg 2018 | 19 | 42 | 0,452380952 | 0,547619048 |
| Fang 2019 | Lian 2019 | 18 | 40 | 0,45 | 0,55 |
| Aletreby 2019 | Ni 2019 | 9 | 20 | 0,45 | 0,55 |
| Annane 2025 | Rochwerg 2018 | 40 | 89 | 0,449438202 | 0,550561798 |
| Fang 2019 | Wen 2019 | 17 | 38 | 0,447368421 | 0,552631579 |
| Rochwerg 2018 | Rygard 2018 | 20 | 45 | 0,444444444 | 0,555555556 |
| Xu 2018 | Zhou 2018 | 8 | 18 | 0,444444444 | 0,555555556 |
| Fang 2019 | Son 2021 | 18 | 41 | 0,43902439 | 0,56097561 |
| Assouline 2021 | Deng 2024 | 7 | 16 | 0,4375 | 0,5625 |
| Deng 2024 | Somagutta 2021 | 7 | 16 | 0,4375 | 0,5625 |
| Deng 2024 | Yao 2021 | 7 | 16 | 0,4375 | 0,5625 |
| Fang 2019 | Rygard 2018 | 18 | 42 | 0,428571429 | 0,571428571 |
| Son 2021 | Wen 2019 | 12 | 28 | 0,428571429 | 0,571428571 |
| Ni 2019 | Pitre 2024 | 19 | 45 | 0,422222222 | 0,577777778 |
| Aletreby 2019 | Zhou 2018 | 8 | 19 | 0,421052632 | 0,578947368 |
| Aletreby 2019 | Allen 2019 | 5 | 12 | 0,416666667 | 0,583333333 |
| Lian 2019 | Lin 2019 | 15 | 36 | 0,416666667 | 0,583333333 |
| Pitre 2024 | Rygard 2018 | 20 | 48 | 0,416666667 | 0,583333333 |
| Lee 2022 | Saghafi 2024 | 5 | 12 | 0,416666667 | 0,583333333 |
| Rygard 2018 | Wen 2019 | 12 | 29 | 0,413793103 | 0,586206897 |
| Annane 2025 | Fang 2019 | 36 | 88 | 0,409090909 | 0,590909091 |
| Lian 2019 | Pitre 2024 | 19 | 47 | 0,404255319 | 0,595744681 |
| Assouline 2021 | Lee 2022 | 4 | 10 | 0,4 | 0,6 |
| Lian 2019 | Rochwerg 2018 | 18 | 45 | 0,4 | 0,6 |
| Barakakis 2019 | Siddiqi 2019 | 2 | 5 | 0,4 | 0,6 |
| Lee 2022 | Somagutta 2021 | 4 | 10 | 0,4 | 0,6 |
| Siddiqi 2019 | Song Y 2024 | 4 | 10 | 0,4 | 0,6 |
| Deng 2024 | Wu 2021 | 6 | 15 | 0,4 | 0,6 |
| Ni 2019 | Xu 2018 | 8 | 20 | 0,4 | 0,6 |
| Wu 2020 | Xu 2018 | 6 | 15 | 0,4 | 0,6 |
| Rochwerg 2018 | Wen 2019 | 17 | 43 | 0,395348837 | 0,604651163 |
| Lin 2019 | Rygard 2018 | 15 | 38 | 0,394736842 | 0,605263158 |

| | | | | | |
|---|---|---|---|---|---|
| Son 2021 | Zhou 2018 | 11 | 28 | 0,392857143 | 0,607142857 |
| Rochwerg 2018 | Son 2021 | 18 | 46 | 0,391304348 | 0,608695652 |
| Lian 2019 | Son 2021 | 12 | 31 | 0,387096774 | 0,612903226 |
| Lyu 2020 | Siddiqi 2019 | 5 | 13 | 0,384615385 | 0,615384615 |
| Lin 2019 | Zhou 2018 | 13 | 34 | 0,382352941 | 0,617647059 |
| Annane 2015 | Annane 2025 | 33 | 87 | 0,379310345 | 0,620689655 |
| Lyu 2020 | Song Y 2024 | 6 | 16 | 0,375 | 0,625 |
| Aletreby 2019 | Wu 2020 | 6 | 16 | 0,375 | 0,625 |
| Rochwerg 2018 | Zhou 2018 | 16 | 43 | 0,372093023 | 0,627906977 |
| Pitre 2024 | Wen 2019 | 17 | 46 | 0,369565217 | 0,630434783 |
| Lu 2021 | Zhou 2018 | 7 | 19 | 0,368421053 | 0,631578947 |
| Pitre 2024 | Son 2021 | 18 | 49 | 0,367346939 | 0,632653061 |
| Kato 2023 | Lee 2022 | 4 | 11 | 0,363636364 | 0,636363636 |
| Lian 2019 | Lu 2021 | 8 | 22 | 0,363636364 | 0,636363636 |
| Wen 2019 | Wu 2020 | 8 | 22 | 0,363636364 | 0,636363636 |
| Lian 2019 | Xu 2018 | 8 | 22 | 0,363636364 | 0,636363636 |
| Lu 2023 | Zayed 2022 | 4 | 11 | 0,363636364 | 0,636363636 |
| Aletreby 2019 | Song Y 2024 | 5 | 14 | 0,357142857 | 0,642857143 |
| Annane 2019 | Rygard 2018 | 22 | 62 | 0,35483871 | 0,64516129 |
| Liang 2021 | Ni 2019 | 18 | 51 | 0,352941176 | 0,647058824 |
| Liang 2021 | Rygard 2018 | 19 | 54 | 0,351851852 | 0,648148148 |
| Fang 2019 | Lyu 2020 | 13 | 37 | 0,351351351 | 0,648648649 |
| Wen 2019 | Xu 2018 | 7 | 20 | 0,35 | 0,65 |
| Aletreby 2019 | Lian 2019 | 8 | 23 | 0,347826087 | 0,652173913 |
| Pitre 2024 | Zhou 2018 | 16 | 46 | 0,347826087 | 0,652173913 |
| Lyu 2020 | Son 2021 | 9 | 26 | 0,346153846 | 0,653846154 |
| Annane 2025 | Lin 2019 | 30 | 87 | 0,344827586 | 0,655172414 |
| Annane 2015 | Wen 2019 | 13 | 38 | 0,342105263 | 0,657894737 |
| Lian 2019 | Liang 2021 | 18 | 53 | 0,339622642 | 0,660377358 |
| Allen 2019 | Lyu 2020 | 5 | 15 | 0,333333333 | 0,666666667 |
| Annane 2015 | Ni 2019 | 13 | 39 | 0,333333333 | 0,666666667 |
| Allen 2019 | Siddiqi 2019 | 3 | 9 | 0,333333333 | 0,666666667 |
| Ni 2019 | Song Y 2024 | 7 | 21 | 0,333333333 | 0,666666667 |
| Aletreby 2019 | Wen 2019 | 7 | 21 | 0,333333333 | 0,666666667 |
| Lee 2022 | Wu 2021 | 3 | 9 | 0,333333333 | 0,666666667 |
| Siddiqi 2019 | Wu 2024 | 2 | 6 | 0,333333333 | 0,666666667 |
| Rygard 2018 | Xu 2018 | 8 | 24 | 0,333333333 | 0,666666667 |
| Siddiqi 2019 | Yamamoto 2020 | 2 | 6 | 0,333333333 | 0,666666667 |
| Aletreby 2019 | Rygard 2018 | 8 | 25 | 0,32 | 0,68 |
| Annane 2015 | Lian 2019 | 13 | 41 | 0,317073171 | 0,682926829 |
| Annane 2015 | Zhou 2018 | 12 | 38 | 0,315789474 | 0,684210526 |
| Liang 2021 | Zhou 2018 | 16 | 51 | 0,31372549 | 0,68627451 |
| Annane 2019 | Ni 2019 | 19 | 61 | 0,31147541 | 0,68852459 |
| Lyu 2020 | Rochwerg 2018 | 13 | 42 | 0,30952381 | 0,69047619 |
| Annane 2015 | Son 2021 | 13 | 42 | 0,30952381 | 0,69047619 |

| | | | | | |
|---|---|---|---|---|---|
| Liang 2021 | Son 2021 | 17 | 55 | 0,309090909 | 0,690909091 |
| Liang 2021 | Wen 2019 | 16 | 52 | 0,307692308 | 0,692307692 |
| Lin 2019 | Lyu 2020 | 10 | 33 | 0,303030303 | 0,696969697 |
| Annane 2015 | Rygard 2018 | 13 | 43 | 0,302325581 | 0,697674419 |
| Annane 2019 | Lian 2019 | 19 | 63 | 0,301587302 | 0,698412698 |
| Na 2021 | Sudaryadi 2023 | 3 | 10 | 0,3 | 0,7 |
| Song Y 2024 | Zhou 2018 | 6 | 20 | 0,3 | 0,7 |
| Lu 2021 | Lyu 2020 | 5 | 17 | 0,294117647 | 0,705882353 |
| Deng 2024 | Zayed 2022 | 5 | 17 | 0,294117647 | 0,705882353 |
| Siddiqi 2019 | Zhou 2018 | 5 | 17 | 0,294117647 | 0,705882353 |
| Lu 2021 | Son 2021 | 7 | 24 | 0,291666667 | 0,708333333 |
| Fang 2019 | Wu 2020 | 11 | 38 | 0,289473684 | 0,710526316 |
| Lyu 2020 | Pitre 2024 | 13 | 45 | 0,288888889 | 0,711111111 |
| Rygard 2018 | Son 2021 | 10 | 35 | 0,285714286 | 0,714285714 |
| Lu 2021 | Wen 2019 | 6 | 21 | 0,285714286 | 0,714285714 |
| Song Y 2024 | Wen 2019 | 6 | 21 | 0,285714286 | 0,714285714 |
| Aletreby 2019 | Son 2021 | 7 | 25 | 0,28 | 0,72 |
| Rygard 2018 | Song Y 2024 | 7 | 25 | 0,28 | 0,72 |
| Annane 2019 | Zhou 2018 | 17 | 61 | 0,278688525 | 0,721311475 |
| Siddiqi 2019 | Wen 2019 | 5 | 18 | 0,277777778 | 0,722222222 |
| Annane 2019 | Son 2021 | 18 | 65 | 0,276923077 | 0,723076923 |
| Annane 2019 | Wen 2019 | 17 | 62 | 0,274193548 | 0,725806452 |
| Lee 2022 | Lu 2023 | 3 | 11 | 0,272727273 | 0,727272727 |
| Lu 2021 | Ni 2019 | 6 | 22 | 0,272727273 | 0,727272727 |
| Lu 2021 | Siddiqi 2019 | 3 | 11 | 0,272727273 | 0,727272727 |
| Aletreby 2019 | Fang 2019 | 10 | 37 | 0,27027027 | 0,72972973 |
| Aletreby 2019 | Lu 2021 | 4 | 15 | 0,266666667 | 0,733333333 |
| Allen 2019 | Wu 2020 | 4 | 15 | 0,266666667 | 0,733333333 |
| Aletreby 2019 | Xu 2018 | 4 | 15 | 0,266666667 | 0,733333333 |
| Ni 2019 | Siddiqi 2019 | 5 | 19 | 0,263157895 | 0,736842105 |
| Liang 2021 | Lyu 2020 | 13 | 50 | 0,26 | 0,74 |
| Son 2021 | Wu 2020 | 7 | 27 | 0,259259259 | 0,740740741 |
| Rochwerg 2018 | Wu 2020 | 11 | 43 | 0,255813953 | 0,744186047 |
| Aletreby 2019 | Lin 2019 | 8 | 32 | 0,25 | 0,75 |
| Annane 2025 | Rygard 2018 | 22 | 88 | 0,25 | 0,75 |
| Lian 2019 | Song Y 2024 | 6 | 24 | 0,25 | 0,75 |
| Son 2021 | Xu 2018 | 6 | 25 | 0,24 | 0,76 |
| Pitre 2024 | Wu 2020 | 11 | 46 | 0,239130435 | 0,760869565 |
| Allen 2019 | Ni 2019 | 5 | 21 | 0,238095238 | 0,761904762 |
| Aletreby 2019 | Rochwerg 2018 | 10 | 42 | 0,238095238 | 0,761904762 |
| Lian 2019 | Siddiqi 2019 | 5 | 21 | 0,238095238 | 0,761904762 |
| Deng 2024 | Lee 2022 | 4 | 17 | 0,235294118 | 0,764705882 |
| Lin 2019 | Wu 2020 | 8 | 34 | 0,235294118 | 0,764705882 |
| Lu 2021 | Wu 2020 | 4 | 17 | 0,235294118 | 0,764705882 |
| Song Y 2024 | Wu 2020 | 4 | 17 | 0,235294118 | 0,764705882 |

| | | | | | |
|---|---|---|---|---|---|
| Annane 2015 | Xu 2018 | 8 | 34 | 0,235294118 | 0,764705882 |
| Lu 2021 | Rygard 2018 | 6 | 26 | 0,230769231 | 0,769230769 |
| Allen 2019 | Song Y 2024 | 3 | 13 | 0,230769231 | 0,769230769 |
| Siddiqi 2019 | Son 2021 | 5 | 22 | 0,227272727 | 0,772727273 |
| Aletreby 2019 | Pitre 2024 | 10 | 45 | 0,222222222 | 0,777777778 |
| Barakakis 2019 | Song Y 2024 | 2 | 9 | 0,222222222 | 0,777777778 |
| Assouline 2021 | Sudaryadi 2023 | 2 | 9 | 0,222222222 | 0,777777778 |
| Lin 2019 | Lu 2021 | 7 | 32 | 0,21875 | 0,78125 |
| Lin 2019 | Song Y 2024 | 7 | 32 | 0,21875 | 0,78125 |
| Lin 2019 | Xu 2018 | 7 | 32 | 0,21875 | 0,78125 |
| Annane 2025 | Ni 2019 | 19 | 87 | 0,218390805 | 0,781609195 |
| Allen 2019 | Lian 2019 | 5 | 23 | 0,217391304 | 0,782608696 |
| Rygard 2018 | Siddiqi 2019 | 5 | 23 | 0,217391304 | 0,782608696 |
| Liang 2021 | Wu 2020 | 11 | 51 | 0,215686275 | 0,784313725 |
| Rochwerg 2018 | Song Y 2024 | 9 | 42 | 0,214285714 | 0,785714286 |
| Siddiqi 2019 | Wu 2020 | 3 | 14 | 0,214285714 | 0,785714286 |
| Annane 2025 | Lian 2019 | 19 | 89 | 0,213483146 | 0,786516854 |
| Annane 2019 | Lyu 2020 | 13 | 61 | 0,213114754 | 0,786885246 |
| Annane 2015 | Lyu 2020 | 8 | 38 | 0,210526316 | 0,789473684 |
| Fang 2019 | Song Y 2024 | 8 | 38 | 0,210526316 | 0,789473684 |
| Fang 2019 | Xu 2018 | 8 | 38 | 0,210526316 | 0,789473684 |
| Allen 2019 | Son 2021 | 5 | 24 | 0,208333333 | 0,791666667 |
| Aletreby 2019 | Barakakis 2019 | 2 | 10 | 0,2 | 0,8 |
| Aletreby 2019 | Liang 2021 | 10 | 50 | 0,2 | 0,8 |
| Allen 2019 | Rygard 2018 | 5 | 25 | 0,2 | 0,8 |
| Pitre 2024 | Song Y 2024 | 9 | 45 | 0,2 | 0,8 |
| Kato 2023 | Sudaryadi 2023 | 2 | 10 | 0,2 | 0,8 |
| Song Y 2024 | Wu 2024 | 2 | 10 | 0,2 | 0,8 |
| Song Y 2024 | Xu 2018 | 3 | 15 | 0,2 | 0,8 |
| Song Y 2024 | Yamamoto 2020 | 2 | 10 | 0,2 | 0,8 |
| Allen 2019 | Zhou 2018 | 4 | 20 | 0,2 | 0,8 |
| Annane 2025 | Son 2021 | 18 | 91 | 0,197802198 | 0,802197802 |
| Annane 2025 | Zhou 2018 | 17 | 87 | 0,195402299 | 0,804597701 |
| Annane 2025 | Wen 2019 | 17 | 88 | 0,193181818 | 0,806818182 |
| Son 2021 | Song Y 2024 | 5 | 26 | 0,192307692 | 0,807692308 |
| Allen 2019 | Wen 2019 | 4 | 21 | 0,19047619 | 0,80952381 |
| Rochwerg 2018 | Xu 2018 | 8 | 43 | 0,186046512 | 0,813953488 |
| Annane 2015 | Wu 2020 | 7 | 38 | 0,184210526 | 0,815789474 |
| Aletreby 2019 | Wu 2024 | 2 | 11 | 0,181818182 | 0,818181818 |
| Aletreby 2019 | Yamamoto 2020 | 2 | 11 | 0,181818182 | 0,818181818 |
| Fang 2019 | Lu 2021 | 7 | 39 | 0,179487179 | 0,820512821 |
| Annane 2019 | Wu 2020 | 11 | 62 | 0,177419355 | 0,822580645 |
| Lu 2021 | Pitre 2024 | 8 | 46 | 0,173913043 | 0,826086957 |
| Pitre 2024 | Xu 2018 | 8 | 46 | 0,173913043 | 0,826086957 |
| Annane 2015 | Lu 2021 | 6 | 36 | 0,166666667 | 0,833333333 |

| | | | | | |
|---|---|---|---|---|---|
| Fratoni 2020 | Lu 2021 | 2 | 12 | 0,166666667 | 0,833333333 |
| Lin 2019 | Siddiqi 2019 | 5 | 30 | 0,166666667 | 0,833333333 |
| Saghafi 2024 | Sudaryadi 2023 | 2 | 12 | 0,166666667 | 0,833333333 |
| Fratoni 2020 | Xu 2018 | 2 | 12 | 0,166666667 | 0,833333333 |
| Siddiqi 2019 | Xu 2018 | 2 | 12 | 0,166666667 | 0,833333333 |
| Aletreby 2019 | Annane 2019 | 10 | 61 | 0,163934426 | 0,836065574 |
| Lu 2021 | Rochwerg 2018 | 7 | 44 | 0,159090909 | 0,840909091 |
| Allen 2019 | Fang 2019 | 6 | 38 | 0,157894737 | 0,842105263 |
| Fratoni 2020 | Zhou 2018 | 3 | 19 | 0,157894737 | 0,842105263 |
| Liang 2021 | Song Y 2024 | 8 | 51 | 0,156862745 | 0,843137255 |
| Allen 2019 | Lin 2019 | 5 | 32 | 0,15625 | 0,84375 |
| Barakakis 2019 | Lyu 2020 | 2 | 13 | 0,153846154 | 0,846153846 |
| Fratoni 2020 | Wen 2019 | 3 | 20 | 0,15 | 0,85 |
| Annane 2025 | Lyu 2020 | 13 | 87 | 0,149425287 | 0,850574713 |
| Annane 2019 | Song Y 2024 | 9 | 61 | 0,147540984 | 0,852459016 |
| Lyu 2020 | Wu 2024 | 2 | 14 | 0,142857143 | 0,857142857 |
| Lyu 2020 | Yamamoto 2020 | 2 | 14 | 0,142857143 | 0,857142857 |
| Allen 2019 | Liang 2021 | 7 | 50 | 0,14 | 0,86 |
| Allen 2019 | Rochwerg 2018 | 6 | 43 | 0,139534884 | 0,860465116 |
| Fang 2019 | Siddiqi 2019 | 5 | 37 | 0,135135135 | 0,864864865 |
| Annane 2015 | Song Y 2024 | 5 | 37 | 0,135135135 | 0,864864865 |
| Liang 2021 | Lu 2021 | 7 | 52 | 0,134615385 | 0,865384615 |
| Liang 2021 | Xu 2018 | 7 | 52 | 0,134615385 | 0,865384615 |
| Aletreby 2019 | Annane 2015 | 5 | 38 | 0,131578947 | 0,868421053 |
| Allen 2019 | Pitre 2024 | 6 | 46 | 0,130434783 | 0,869565217 |
| Annane 2019 | Lu 2021 | 8 | 62 | 0,129032258 | 0,870967742 |
| Annane 2019 | Xu 2018 | 8 | 62 | 0,129032258 | 0,870967742 |
| Allen 2019 | Barakakis 2019 | 1 | 8 | 0,125 | 0,875 |
| Fratoni 2020 | Son 2021 | 3 | 24 | 0,125 | 0,875 |
| Lu 2021 | Song Y 2024 | 2 | 16 | 0,125 | 0,875 |
| Deng 2024 | Sudaryadi 2023 | 2 | 16 | 0,125 | 0,875 |
| Lee 2022 | Sudaryadi 2023 | 1 | 8 | 0,125 | 0,875 |
| Annane 2025 | Wu 2020 | 11 | 88 | 0,125 | 0,875 |
| Sudaryadi 2023 | Wu 2021 | 1 | 8 | 0,125 | 0,875 |
| Rochwerg 2018 | Siddiqi 2019 | 5 | 42 | 0,119047619 | 0,880952381 |
| Barakakis 2019 | Zhou 2018 | 2 | 17 | 0,117647059 | 0,882352941 |
| Aletreby 2019 | Annane 2025 | 10 | 87 | 0,114942529 | 0,885057471 |
| Allen 2019 | Annane 2019 | 7 | 61 | 0,114754098 | 0,885245902 |
| Fratoni 2020 | Siddiqi 2019 | 1 | 9 | 0,111111111 | 0,888888889 |
| Pitre 2024 | Siddiqi 2019 | 5 | 45 | 0,111111111 | 0,888888889 |
| Barakakis 2019 | Wen 2019 | 2 | 18 | 0,111111111 | 0,888888889 |
| Allen 2019 | Wu 2024 | 1 | 9 | 0,111111111 | 0,888888889 |
| Allen 2019 | Yamamoto 2020 | 1 | 9 | 0,111111111 | 0,888888889 |
| Sudaryadi 2023 | Zayed 2022 | 1 | 9 | 0,111111111 | 0,888888889 |
| Wu 2024 | Zhou 2018 | 2 | 18 | 0,111111111 | 0,888888889 |

| | | | | | |
|---|---|---|---|---|---|
| Yamamoto 2020 | Zhou 2018 | 2 | 18 | 0,111111111 | 0,888888889 |
| Rochwerg 2018 | Yang 2020 | 5 | 47 | 0,106382979 | 0,893617021 |
| Barakakis 2019 | Ni 2019 | 2 | 19 | 0,105263158 | 0,894736842 |
| Wen 2019 | Wu 2024 | 2 | 19 | 0,105263158 | 0,894736842 |
| Wen 2019 | Yamamoto 2020 | 2 | 19 | 0,105263158 | 0,894736842 |
| Annane 2025 | Song Y 2024 | 9 | 87 | 0,103448276 | 0,896551724 |
| Barakakis 2019 | Lu 2021 | 1 | 10 | 0,1 | 0,9 |
| Liang 2021 | Siddiqi 2019 | 5 | 50 | 0,1 | 0,9 |
| Somagutta 2021 | Sudaryadi 2023 | 1 | 10 | 0,1 | 0,9 |
| Ni 2019 | Wu 2024 | 2 | 20 | 0,1 | 0,9 |
| Barakakis 2019 | Xu 2018 | 1 | 10 | 0,1 | 0,9 |
| Ni 2019 | Yamamoto 2020 | 2 | 20 | 0,1 | 0,9 |
| Pitre 2024 | Yang 2020 | 5 | 50 | 0,1 | 0,9 |
| Sudaryadi 2023 | Yao 2021 | 1 | 10 | 0,1 | 0,9 |
| Annane 2025 | Deng 2024 | 9 | 93 | 0,096774194 | 0,903225806 |
| Barakakis 2019 | Lian 2019 | 2 | 21 | 0,095238095 | 0,904761905 |
| Fratoni 2020 | Lin 2019 | 3 | 32 | 0,09375 | 0,90625 |
| Annane 2025 | Lu 2023 | 8 | 87 | 0,091954023 | 0,908045977 |
| Annane 2025 | Lu 2021 | 8 | 88 | 0,090909091 | 0,909090909 |
| Fratoni 2020 | Ni 2019 | 2 | 22 | 0,090909091 | 0,909090909 |
| Barakakis 2019 | Son 2021 | 2 | 22 | 0,090909091 | 0,909090909 |
| Lian 2019 | Wu 2024 | 2 | 22 | 0,090909091 | 0,909090909 |
| Lu 2021 | Wu 2024 | 1 | 11 | 0,090909091 | 0,909090909 |
| Annane 2025 | Xu 2018 | 8 | 88 | 0,090909091 | 0,909090909 |
| Wu 2024 | Xu 2018 | 1 | 11 | 0,090909091 | 0,909090909 |
| Lian 2019 | Yamamoto 2020 | 2 | 22 | 0,090909091 | 0,909090909 |
| Lu 2021 | Yamamoto 2020 | 1 | 11 | 0,090909091 | 0,909090909 |
| Xu 2018 | Yamamoto 2020 | 1 | 11 | 0,090909091 | 0,909090909 |
| Annane 2025 | Saghafi 2024 | 8 | 90 | 0,088888889 | 0,911111111 |
| Barakakis 2019 | Rygard 2018 | 2 | 23 | 0,086956522 | 0,913043478 |
| Son 2021 | Wu 2024 | 2 | 23 | 0,086956522 | 0,913043478 |
| Son 2021 | Yamamoto 2020 | 2 | 23 | 0,086956522 | 0,913043478 |
| Annane 2015 | Fratoni 2020 | 3 | 35 | 0,085714286 | 0,914285714 |
| Fratoni 2020 | Lian 2019 | 2 | 24 | 0,083333333 | 0,916666667 |
| Rygard 2018 | Wu 2024 | 2 | 24 | 0,083333333 | 0,916666667 |
| Rygard 2018 | Yamamoto 2020 | 2 | 24 | 0,083333333 | 0,916666667 |
| Annane 2019 | Siddiqi 2019 | 5 | 61 | 0,081967213 | 0,918032787 |
| Allen 2019 | Annane 2025 | 7 | 87 | 0,08045977 | 0,91954023 |
| Annane 2025 | Kato 2023 | 7 | 89 | 0,078651685 | 0,921348315 |
| Fang 2019 | Fratoni 2020 | 3 | 39 | 0,076923077 | 0,923076923 |
| Fratoni 2020 | Rygard 2018 | 2 | 26 | 0,076923077 | 0,923076923 |
| Deng 2025 | Rochwerg 2018 | 4 | 53 | 0,075471698 | 0,924528302 |
| Aletreby 2019 | Fratoni 2020 | 1 | 14 | 0,071428571 | 0,928571429 |
| Deng 2025 | Pitre 2024 | 4 | 56 | 0,071428571 | 0,928571429 |
| Fratoni 2020 | Rochwerg 2018 | 3 | 44 | 0,068181818 | 0,931818182 |

| | | | | | |
|---|---|---|---|---|---|
| Annane 2025 | Assouline 2021 | 6 | 89 | 0,06741573 | 0,93258427 |
| Annane 2025 | Somagutta 2021 | 6 | 89 | 0,06741573 | 0,93258427 |
| Annane 2025 | Yao 2021 | 6 | 89 | 0,06741573 | 0,93258427 |
| Barakakis 2019 | Lin 2019 | 2 | 30 | 0,066666667 | 0,933333333 |
| Allen 2019 | Lu 2021 | 1 | 15 | 0,066666667 | 0,933333333 |
| Annane 2025 | Na 2021 | 6 | 91 | 0,065934066 | 0,934065934 |
| Lin 2019 | Wu 2024 | 2 | 31 | 0,064516129 | 0,935483871 |
| Lin 2019 | Yamamoto 2020 | 2 | 31 | 0,064516129 | 0,935483871 |
| Fratoni 2020 | Pitre 2024 | 3 | 47 | 0,063829787 | 0,936170213 |
| Fratoni 2020 | Wu 2020 | 1 | 16 | 0,0625 | 0,9375 |
| Liang 2021 | Wu 2024 | 3 | 50 | 0,06 | 0,94 |
| Liang 2021 | Yamamoto 2020 | 3 | 50 | 0,06 | 0,94 |
| Annane 2019 | Yang 2020 | 4 | 67 | 0,059701493 | 0,940298507 |
| Fratoni 2020 | Lyu 2020 | 1 | 17 | 0,058823529 | 0,941176471 |
| Annane 2015 | Wu 2024 | 2 | 34 | 0,058823529 | 0,941176471 |
| Annane 2015 | Yamamoto 2020 | 2 | 34 | 0,058823529 | 0,941176471 |
| Fratoni 2020 | Liang 2021 | 3 | 52 | 0,057692308 | 0,942307692 |
| Annane 2025 | Siddiqi 2019 | 5 | 87 | 0,057471264 | 0,942528736 |
| Annane 2025 | Wu 2021 | 5 | 88 | 0,056818182 | 0,943181818 |
| Annane 2019 | Deng 2025 | 4 | 72 | 0,055555556 | 0,944444444 |
| Annane 2015 | Siddiqi 2019 | 2 | 36 | 0,055555556 | 0,944444444 |
| Song Y 2024 | Yang 2020 | 1 | 18 | 0,055555556 | 0,944444444 |
| Barakakis 2019 | Fang 2019 | 2 | 37 | 0,054054054 | 0,945945946 |
| Fang 2019 | Wu 2024 | 2 | 38 | 0,052631579 | 0,947368421 |
| Fang 2019 | Yamamoto 2020 | 2 | 38 | 0,052631579 | 0,947368421 |
| Annane 2019 | Wu 2024 | 3 | 61 | 0,049180328 | 0,950819672 |
| Annane 2019 | Yamamoto 2020 | 3 | 61 | 0,049180328 | 0,950819672 |
| Annane 2015 | Yang 2020 | 2 | 41 | 0,048780488 | 0,951219512 |
| Annane 2019 | Fratoni 2020 | 3 | 63 | 0,047619048 | 0,952380952 |
| Barakakis 2019 | Rochwerg 2018 | 2 | 42 | 0,047619048 | 0,952380952 |
| Rochwerg 2018 | Wu 2024 | 2 | 43 | 0,046511628 | 0,953488372 |
| Rochwerg 2018 | Yamamoto 2020 | 2 | 43 | 0,046511628 | 0,953488372 |
| Barakakis 2019 | Pitre 2024 | 2 | 45 | 0,044444444 | 0,955555556 |
| Annane 2025 | Zayed 2022 | 4 | 90 | 0,044444444 | 0,955555556 |
| Annane 2015 | Deng 2025 | 2 | 46 | 0,043478261 | 0,956521739 |
| Deng 2025 | Song Y 2024 | 1 | 23 | 0,043478261 | 0,956521739 |
| Pitre 2024 | Wu 2024 | 2 | 46 | 0,043478261 | 0,956521739 |
| Pitre 2024 | Yamamoto 2020 | 2 | 46 | 0,043478261 | 0,956521739 |
| Annane 2025 | Yang 2020 | 4 | 93 | 0,043010753 | 0,956989247 |
| Annane 2025 | Deng 2025 | 4 | 98 | 0,040816327 | 0,959183673 |
| Barakakis 2019 | Liang 2021 | 2 | 50 | 0,04 | 0,96 |
| Li 2025 | Liang 2021 | 2 | 51 | 0,039215686 | 0,960784314 |
| Annane 2025 | Li 2025 | 3 | 87 | 0,034482759 | 0,965517241 |
| Annane 2025 | Wu 2024 | 3 | 87 | 0,034482759 | 0,965517241 |
| Annane 2025 | Yamamoto 2020 | 3 | 87 | 0,034482759 | 0,965517241 |

| | | | | | |
|---|---|---|---|---|---|
| Annane 2025 | Fratoni 2020 | 3 | 89 | 0,033707865 | 0,966292135 |
| Annane 2025 | Lee 2022 | 3 | 90 | 0,033333333 | 0,966666667 |
| Annane 2019 | Barakakis 2019 | 2 | 61 | 0,032786885 | 0,967213115 |
| Annane 2019 | Li 2025 | 2 | 62 | 0,032258065 | 0,967741935 |
| Annane 2015 | Barakakis 2019 | 1 | 34 | 0,029411765 | 0,970588235 |
| Annane 2025 | Barakakis 2019 | 2 | 87 | 0,022988506 | 0,977011494 |
| Allen 2019 | Annane 2015 | 0 | 40 | 0 | 1 |
| Aletreby 2019 | Assouline 2021 | 0 | 18 | 0 | 1 |
| Allen 2019 | Assouline 2021 | 0 | 15 | 0 | 1 |
| Annane 2015 | Assouline 2021 | 0 | 41 | 0 | 1 |
| Annane 2019 | Assouline 2021 | 0 | 69 | 0 | 1 |
| Assouline 2021 | Barakakis 2019 | 0 | 10 | 0 | 1 |
| Aletreby 2019 | Deng 2024 | 0 | 25 | 0 | 1 |
| Allen 2019 | Deng 2024 | 0 | 22 | 0 | 1 |
| Annane 2015 | Deng 2024 | 0 | 48 | 0 | 1 |
| Annane 2019 | Deng 2024 | 0 | 76 | 0 | 1 |
| Barakakis 2019 | Deng 2024 | 0 | 17 | 0 | 1 |
| Aletreby 2019 | Deng 2025 | 0 | 25 | 0 | 1 |
| Allen 2019 | Deng 2025 | 0 | 22 | 0 | 1 |
| Assouline 2021 | Deng 2025 | 0 | 23 | 0 | 1 |
| Barakakis 2019 | Deng 2025 | 0 | 17 | 0 | 1 |
| Deng 2024 | Deng 2025 | 0 | 30 | 0 | 1 |
| Assouline 2021 | Fang 2019 | 0 | 45 | 0 | 1 |
| Deng 2024 | Fang 2019 | 0 | 52 | 0 | 1 |
| Deng 2025 | Fang 2019 | 0 | 52 | 0 | 1 |
| Allen 2019 | Fratoni 2020 | 0 | 12 | 0 | 1 |
| Assouline 2021 | Fratoni 2020 | 0 | 13 | 0 | 1 |
| Barakakis 2019 | Fratoni 2020 | 0 | 7 | 0 | 1 |
| Deng 2024 | Fratoni 2020 | 0 | 20 | 0 | 1 |
| Deng 2025 | Fratoni 2020 | 0 | 20 | 0 | 1 |
| Aletreby 2019 | Kato 2023 | 0 | 19 | 0 | 1 |
| Allen 2019 | Kato 2023 | 0 | 16 | 0 | 1 |
| Annane 2015 | Kato 2023 | 0 | 42 | 0 | 1 |
| Annane 2019 | Kato 2023 | 0 | 70 | 0 | 1 |
| Barakakis 2019 | Kato 2023 | 0 | 11 | 0 | 1 |
| Deng 2025 | Kato 2023 | 0 | 24 | 0 | 1 |
| Fang 2019 | Kato 2023 | 0 | 46 | 0 | 1 |
| Fratoni 2020 | Kato 2023 | 0 | 14 | 0 | 1 |
| Aletreby 2019 | Lee 2022 | 0 | 16 | 0 | 1 |
| Allen 2019 | Lee 2022 | 0 | 13 | 0 | 1 |
| Annane 2015 | Lee 2022 | 0 | 39 | 0 | 1 |
| Annane 2019 | Lee 2022 | 0 | 67 | 0 | 1 |
| Barakakis 2019 | Lee 2022 | 0 | 8 | 0 | 1 |
| Deng 2025 | Lee 2022 | 0 | 21 | 0 | 1 |
| Fang 2019 | Lee 2022 | 0 | 43 | 0 | 1 |

| | | | | | |
|---|---|---|---|---|---|
| Fratoni 2020 | Lee 2022 | 0 | 11 | 0 | 1 |
| Aletreby 2019 | Li 2025 | 0 | 13 | 0 | 1 |
| Allen 2019 | Li 2025 | 0 | 10 | 0 | 1 |
| Annane 2015 | Li 2025 | 0 | 36 | 0 | 1 |
| Assouline 2021 | Li 2025 | 0 | 11 | 0 | 1 |
| Barakakis 2019 | Li 2025 | 0 | 5 | 0 | 1 |
| Deng 2024 | Li 2025 | 0 | 18 | 0 | 1 |
| Deng 2025 | Li 2025 | 0 | 18 | 0 | 1 |
| Fang 2019 | Li 2025 | 0 | 40 | 0 | 1 |
| Fratoni 2020 | Li 2025 | 0 | 8 | 0 | 1 |
| Kato 2023 | Li 2025 | 0 | 12 | 0 | 1 |
| Lee 2022 | Li 2025 | 0 | 9 | 0 | 1 |
| Assouline 2021 | Lian 2019 | 0 | 29 | 0 | 1 |
| Deng 2024 | Lian 2019 | 0 | 36 | 0 | 1 |
| Deng 2025 | Lian 2019 | 0 | 36 | 0 | 1 |
| Kato 2023 | Lian 2019 | 0 | 30 | 0 | 1 |
| Lee 2022 | Lian 2019 | 0 | 27 | 0 | 1 |
| Li 2025 | Lian 2019 | 0 | 24 | 0 | 1 |
| Assouline 2021 | Liang 2021 | 0 | 58 | 0 | 1 |
| Deng 2024 | Liang 2021 | 0 | 65 | 0 | 1 |
| Deng 2025 | Liang 2021 | 0 | 65 | 0 | 1 |
| Kato 2023 | Liang 2021 | 0 | 59 | 0 | 1 |
| Lee 2022 | Liang 2021 | 0 | 56 | 0 | 1 |
| Assouline 2021 | Lin 2019 | 0 | 38 | 0 | 1 |
| Deng 2024 | Lin 2019 | 0 | 45 | 0 | 1 |
| Deng 2025 | Lin 2019 | 0 | 45 | 0 | 1 |
| Kato 2023 | Lin 2019 | 0 | 39 | 0 | 1 |
| Lee 2022 | Lin 2019 | 0 | 36 | 0 | 1 |
| Li 2025 | Lin 2019 | 0 | 33 | 0 | 1 |
| Assouline 2021 | Lu 2021 | 0 | 17 | 0 | 1 |
| Deng 2024 | Lu 2021 | 0 | 24 | 0 | 1 |
| Deng 2025 | Lu 2021 | 0 | 24 | 0 | 1 |
| Kato 2023 | Lu 2021 | 0 | 18 | 0 | 1 |
| Lee 2022 | Lu 2021 | 0 | 15 | 0 | 1 |
| Li 2025 | Lu 2021 | 0 | 12 | 0 | 1 |
| Aletreby 2019 | Lu 2023 | 0 | 18 | 0 | 1 |
| Allen 2019 | Lu 2023 | 0 | 15 | 0 | 1 |
| Annane 2015 | Lu 2023 | 0 | 41 | 0 | 1 |
| Annane 2019 | Lu 2023 | 0 | 69 | 0 | 1 |
| Barakakis 2019 | Lu 2023 | 0 | 10 | 0 | 1 |
| Deng 2025 | Lu 2023 | 0 | 23 | 0 | 1 |
| Fang 2019 | Lu 2023 | 0 | 45 | 0 | 1 |
| Fratoni 2020 | Lu 2023 | 0 | 13 | 0 | 1 |
| Li 2025 | Lu 2023 | 0 | 11 | 0 | 1 |
| Lian 2019 | Lu 2023 | 0 | 29 | 0 | 1 |

| | | | | | |
|---|---|---|---|---|---|
| Liang 2021 | Lu 2023 | 0 | 58 | 0 | 1 |
| Lin 2019 | Lu 2023 | 0 | 38 | 0 | 1 |
| Lu 2021 | Lu 2023 | 0 | 17 | 0 | 1 |
| Assouline 2021 | Lyu 2020 | 0 | 21 | 0 | 1 |
| Deng 2024 | Lyu 2020 | 0 | 28 | 0 | 1 |
| Deng 2025 | Lyu 2020 | 0 | 28 | 0 | 1 |
| Kato 2023 | Lyu 2020 | 0 | 22 | 0 | 1 |
| Lee 2022 | Lyu 2020 | 0 | 19 | 0 | 1 |
| Li 2025 | Lyu 2020 | 0 | 16 | 0 | 1 |
| Lu 2023 | Lyu 2020 | 0 | 21 | 0 | 1 |
| Aletreby 2019 | Na 2021 | 0 | 20 | 0 | 1 |
| Allen 2019 | Na 2021 | 0 | 17 | 0 | 1 |
| Annane 2015 | Na 2021 | 0 | 43 | 0 | 1 |
| Annane 2019 | Na 2021 | 0 | 71 | 0 | 1 |
| Barakakis 2019 | Na 2021 | 0 | 12 | 0 | 1 |
| Deng 2025 | Na 2021 | 0 | 25 | 0 | 1 |
| Fang 2019 | Na 2021 | 0 | 47 | 0 | 1 |
| Fratoni 2020 | Na 2021 | 0 | 15 | 0 | 1 |
| Li 2025 | Na 2021 | 0 | 13 | 0 | 1 |
| Lian 2019 | Na 2021 | 0 | 31 | 0 | 1 |
| Liang 2021 | Na 2021 | 0 | 60 | 0 | 1 |
| Lin 2019 | Na 2021 | 0 | 40 | 0 | 1 |
| Lu 2021 | Na 2021 | 0 | 19 | 0 | 1 |
| Lyu 2020 | Na 2021 | 0 | 23 | 0 | 1 |
| Assouline 2021 | Ni 2019 | 0 | 27 | 0 | 1 |
| Deng 2024 | Ni 2019 | 0 | 34 | 0 | 1 |
| Deng 2025 | Ni 2019 | 0 | 34 | 0 | 1 |
| Kato 2023 | Ni 2019 | 0 | 28 | 0 | 1 |
| Lee 2022 | Ni 2019 | 0 | 25 | 0 | 1 |
| Li 2025 | Ni 2019 | 0 | 22 | 0 | 1 |
| Lu 2023 | Ni 2019 | 0 | 27 | 0 | 1 |
| Na 2021 | Ni 2019 | 0 | 29 | 0 | 1 |
| Assouline 2021 | Pitre 2024 | 0 | 53 | 0 | 1 |
| Deng 2024 | Pitre 2024 | 0 | 60 | 0 | 1 |
| Kato 2023 | Pitre 2024 | 0 | 54 | 0 | 1 |
| Lee 2022 | Pitre 2024 | 0 | 51 | 0 | 1 |
| Li 2025 | Pitre 2024 | 0 | 48 | 0 | 1 |
| Lu 2023 | Pitre 2024 | 0 | 53 | 0 | 1 |
| Na 2021 | Pitre 2024 | 0 | 55 | 0 | 1 |
| Assouline 2021 | Rochwerg 2018 | 0 | 50 | 0 | 1 |
| Deng 2024 | Rochwerg 2018 | 0 | 57 | 0 | 1 |
| Kato 2023 | Rochwerg 2018 | 0 | 51 | 0 | 1 |
| Lee 2022 | Rochwerg 2018 | 0 | 48 | 0 | 1 |
| Li 2025 | Rochwerg 2018 | 0 | 45 | 0 | 1 |
| Lu 2023 | Rochwerg 2018 | 0 | 50 | 0 | 1 |

| | | | | | |
|---|---|---|---|---|---|
| Na 2021 | Rochwerg 2018 | 0 | 52 | 0 | 1 |
| Assouline 2021 | Rygard 2018 | 0 | 31 | 0 | 1 |
| Deng 2024 | Rygard 2018 | 0 | 38 | 0 | 1 |
| Deng 2025 | Rygard 2018 | 0 | 38 | 0 | 1 |
| Kato 2023 | Rygard 2018 | 0 | 32 | 0 | 1 |
| Lee 2022 | Rygard 2018 | 0 | 29 | 0 | 1 |
| Li 2025 | Rygard 2018 | 0 | 26 | 0 | 1 |
| Lu 2023 | Rygard 2018 | 0 | 31 | 0 | 1 |
| Na 2021 | Rygard 2018 | 0 | 33 | 0 | 1 |
| Aletreby 2019 | Saghafi 2024 | 0 | 21 | 0 | 1 |
| Allen 2019 | Saghafi 2024 | 0 | 18 | 0 | 1 |
| Annane 2015 | Saghafi 2024 | 0 | 44 | 0 | 1 |
| Annane 2019 | Saghafi 2024 | 0 | 72 | 0 | 1 |
| Barakakis 2019 | Saghafi 2024 | 0 | 13 | 0 | 1 |
| Deng 2025 | Saghafi 2024 | 0 | 26 | 0 | 1 |
| Fang 2019 | Saghafi 2024 | 0 | 48 | 0 | 1 |
| Fratoni 2020 | Saghafi 2024 | 0 | 16 | 0 | 1 |
| Li 2025 | Saghafi 2024 | 0 | 14 | 0 | 1 |
| Lian 2019 | Saghafi 2024 | 0 | 32 | 0 | 1 |
| Liang 2021 | Saghafi 2024 | 0 | 61 | 0 | 1 |
| Lin 2019 | Saghafi 2024 | 0 | 41 | 0 | 1 |
| Lu 2021 | Saghafi 2024 | 0 | 20 | 0 | 1 |
| Lyu 2020 | Saghafi 2024 | 0 | 24 | 0 | 1 |
| Ni 2019 | Saghafi 2024 | 0 | 30 | 0 | 1 |
| Pitre 2024 | Saghafi 2024 | 0 | 56 | 0 | 1 |
| Rochwerg 2018 | Saghafi 2024 | 0 | 53 | 0 | 1 |
| Rygard 2018 | Saghafi 2024 | 0 | 34 | 0 | 1 |
| Assouline 2021 | Siddiqi 2019 | 0 | 13 | 0 | 1 |
| Deng 2024 | Siddiqi 2019 | 0 | 20 | 0 | 1 |
| Deng 2025 | Siddiqi 2019 | 0 | 20 | 0 | 1 |
| Kato 2023 | Siddiqi 2019 | 0 | 14 | 0 | 1 |
| Lee 2022 | Siddiqi 2019 | 0 | 11 | 0 | 1 |
| Li 2025 | Siddiqi 2019 | 0 | 8 | 0 | 1 |
| Lu 2023 | Siddiqi 2019 | 0 | 13 | 0 | 1 |
| Na 2021 | Siddiqi 2019 | 0 | 15 | 0 | 1 |
| Saghafi 2024 | Siddiqi 2019 | 0 | 16 | 0 | 1 |
| Aletreby 2019 | Somagutta 2021 | 0 | 18 | 0 | 1 |
| Allen 2019 | Somagutta 2021 | 0 | 15 | 0 | 1 |
| Annane 2015 | Somagutta 2021 | 0 | 41 | 0 | 1 |
| Annane 2019 | Somagutta 2021 | 0 | 69 | 0 | 1 |
| Barakakis 2019 | Somagutta 2021 | 0 | 10 | 0 | 1 |
| Deng 2025 | Somagutta 2021 | 0 | 23 | 0 | 1 |
| Fang 2019 | Somagutta 2021 | 0 | 45 | 0 | 1 |
| Fratoni 2020 | Somagutta 2021 | 0 | 13 | 0 | 1 |
| Li 2025 | Somagutta 2021 | 0 | 11 | 0 | 1 |

| | | | | | |
|---|---|---|---|---|---|
| Lian 2019 | Somagutta 2021 | 0 | 29 | 0 | 1 |
| Liang 2021 | Somagutta 2021 | 0 | 58 | 0 | 1 |
| Lin 2019 | Somagutta 2021 | 0 | 38 | 0 | 1 |
| Lu 2021 | Somagutta 2021 | 0 | 17 | 0 | 1 |
| Lyu 2020 | Somagutta 2021 | 0 | 21 | 0 | 1 |
| Ni 2019 | Somagutta 2021 | 0 | 27 | 0 | 1 |
| Pitre 2024 | Somagutta 2021 | 0 | 53 | 0 | 1 |
| Rochwerg 2018 | Somagutta 2021 | 0 | 50 | 0 | 1 |
| Rygard 2018 | Somagutta 2021 | 0 | 31 | 0 | 1 |
| Siddiqi 2019 | Somagutta 2021 | 0 | 13 | 0 | 1 |
| Assouline 2021 | Son 2021 | 0 | 30 | 0 | 1 |
| Deng 2024 | Son 2021 | 0 | 37 | 0 | 1 |
| Deng 2025 | Son 2021 | 0 | 37 | 0 | 1 |
| Kato 2023 | Son 2021 | 0 | 31 | 0 | 1 |
| Lee 2022 | Son 2021 | 0 | 28 | 0 | 1 |
| Li 2025 | Son 2021 | 0 | 25 | 0 | 1 |
| Lu 2023 | Son 2021 | 0 | 30 | 0 | 1 |
| Na 2021 | Son 2021 | 0 | 32 | 0 | 1 |
| Saghafi 2024 | Son 2021 | 0 | 33 | 0 | 1 |
| Somagutta 2021 | Son 2021 | 0 | 30 | 0 | 1 |
| Aletreby 2019 | Song B 2024 | 0 | 10 | 0 | 1 |
| Allen 2019 | Song B 2024 | 0 | 7 | 0 | 1 |
| Annane 2015 | Song B 2024 | 0 | 33 | 0 | 1 |
| Annane 2019 | Song B 2024 | 0 | 61 | 0 | 1 |
| Annane 2025 | Song B 2024 | 0 | 87 | 0 | 1 |
| Assouline 2021 | Song B 2024 | 0 | 8 | 0 | 1 |
| Barakakis 2019 | Song B 2024 | 0 | 2 | 0 | 1 |
| Deng 2024 | Song B 2024 | 0 | 15 | 0 | 1 |
| Deng 2025 | Song B 2024 | 0 | 15 | 0 | 1 |
| Fang 2019 | Song B 2024 | 0 | 37 | 0 | 1 |
| Fratoni 2020 | Song B 2024 | 0 | 5 | 0 | 1 |
| Kato 2023 | Song B 2024 | 0 | 9 | 0 | 1 |
| Lee 2022 | Song B 2024 | 0 | 6 | 0 | 1 |
| Li 2025 | Song B 2024 | 0 | 3 | 0 | 1 |
| Lian 2019 | Song B 2024 | 0 | 21 | 0 | 1 |
| Liang 2021 | Song B 2024 | 0 | 50 | 0 | 1 |
| Lin 2019 | Song B 2024 | 0 | 30 | 0 | 1 |
| Lu 2021 | Song B 2024 | 0 | 9 | 0 | 1 |
| Lu 2023 | Song B 2024 | 0 | 8 | 0 | 1 |
| Lyu 2020 | Song B 2024 | 0 | 13 | 0 | 1 |
| Na 2021 | Song B 2024 | 0 | 10 | 0 | 1 |
| Ni 2019 | Song B 2024 | 0 | 19 | 0 | 1 |
| Pitre 2024 | Song B 2024 | 0 | 45 | 0 | 1 |
| Rochwerg 2018 | Song B 2024 | 0 | 42 | 0 | 1 |
| Rygard 2018 | Song B 2024 | 0 | 23 | 0 | 1 |

| | | | | | |
|---|---|---|---|---|---|
| Saghafi 2024 | Song B 2024 | 0 | 11 | 0 | 1 |
| Siddiqi 2019 | Song B 2024 | 0 | 5 | 0 | 1 |
| Somagutta 2021 | Song B 2024 | 0 | 8 | 0 | 1 |
| Son 2021 | Song B 2024 | 0 | 22 | 0 | 1 |
| Assouline 2021 | Song Y 2024 | 0 | 17 | 0 | 1 |
| Deng 2024 | Song Y 2024 | 0 | 24 | 0 | 1 |
| Fratoni 2020 | Song Y 2024 | 0 | 14 | 0 | 1 |
| Kato 2023 | Song Y 2024 | 0 | 18 | 0 | 1 |
| Lee 2022 | Song Y 2024 | 0 | 15 | 0 | 1 |
| Li 2025 | Song Y 2024 | 0 | 12 | 0 | 1 |
| Lu 2023 | Song Y 2024 | 0 | 17 | 0 | 1 |
| Na 2021 | Song Y 2024 | 0 | 19 | 0 | 1 |
| Saghafi 2024 | Song Y 2024 | 0 | 20 | 0 | 1 |
| Somagutta 2021 | Song Y 2024 | 0 | 17 | 0 | 1 |
| Song B 2024 | Song Y 2024 | 0 | 9 | 0 | 1 |
| Aletreby 2019 | Sudaryadi 2023 | 0 | 13 | 0 | 1 |
| Allen 2019 | Sudaryadi 2023 | 0 | 10 | 0 | 1 |
| Annane 2015 | Sudaryadi 2023 | 0 | 36 | 0 | 1 |
| Annane 2019 | Sudaryadi 2023 | 0 | 64 | 0 | 1 |
| Annane 2025 | Sudaryadi 2023 | 0 | 90 | 0 | 1 |
| Barakakis 2019 | Sudaryadi 2023 | 0 | 5 | 0 | 1 |
| Deng 2025 | Sudaryadi 2023 | 0 | 18 | 0 | 1 |
| Fang 2019 | Sudaryadi 2023 | 0 | 40 | 0 | 1 |
| Fratoni 2020 | Sudaryadi 2023 | 0 | 8 | 0 | 1 |
| Li 2025 | Sudaryadi 2023 | 0 | 6 | 0 | 1 |
| Lian 2019 | Sudaryadi 2023 | 0 | 24 | 0 | 1 |
| Liang 2021 | Sudaryadi 2023 | 0 | 53 | 0 | 1 |
| Lin 2019 | Sudaryadi 2023 | 0 | 33 | 0 | 1 |
| Lu 2021 | Sudaryadi 2023 | 0 | 12 | 0 | 1 |
| Lu 2023 | Sudaryadi 2023 | 0 | 11 | 0 | 1 |
| Lyu 2020 | Sudaryadi 2023 | 0 | 16 | 0 | 1 |
| Ni 2019 | Sudaryadi 2023 | 0 | 22 | 0 | 1 |
| Pitre 2024 | Sudaryadi 2023 | 0 | 48 | 0 | 1 |
| Rochwerg 2018 | Sudaryadi 2023 | 0 | 45 | 0 | 1 |
| Rygard 2018 | Sudaryadi 2023 | 0 | 26 | 0 | 1 |
| Siddiqi 2019 | Sudaryadi 2023 | 0 | 8 | 0 | 1 |
| Son 2021 | Sudaryadi 2023 | 0 | 25 | 0 | 1 |
| Song B 2024 | Sudaryadi 2023 | 0 | 3 | 0 | 1 |
| Song Y 2024 | Sudaryadi 2023 | 0 | 12 | 0 | 1 |
| Assouline 2021 | Wen 2019 | 0 | 26 | 0 | 1 |
| Deng 2024 | Wen 2019 | 0 | 33 | 0 | 1 |
| Deng 2025 | Wen 2019 | 0 | 33 | 0 | 1 |
| Kato 2023 | Wen 2019 | 0 | 27 | 0 | 1 |
| Lee 2022 | Wen 2019 | 0 | 24 | 0 | 1 |
| Li 2025 | Wen 2019 | 0 | 21 | 0 | 1 |

| | | | | | |
|---|---|---|---|---|---|
| Lu 2023 | Wen 2019 | 0 | 26 | 0 | 1 |
| Na 2021 | Wen 2019 | 0 | 28 | 0 | 1 |
| Saghafi 2024 | Wen 2019 | 0 | 29 | 0 | 1 |
| Somagutta 2021 | Wen 2019 | 0 | 26 | 0 | 1 |
| Song B 2024 | Wen 2019 | 0 | 18 | 0 | 1 |
| Sudaryadi 2023 | Wen 2019 | 0 | 21 | 0 | 1 |
| Assouline 2021 | Wu 2020 | 0 | 20 | 0 | 1 |
| Barakakis 2019 | Wu 2020 | 0 | 14 | 0 | 1 |
| Deng 2024 | Wu 2020 | 0 | 27 | 0 | 1 |
| Deng 2025 | Wu 2020 | 0 | 27 | 0 | 1 |
| Kato 2023 | Wu 2020 | 0 | 21 | 0 | 1 |
| Lee 2022 | Wu 2020 | 0 | 18 | 0 | 1 |
| Li 2025 | Wu 2020 | 0 | 15 | 0 | 1 |
| Lu 2023 | Wu 2020 | 0 | 20 | 0 | 1 |
| Na 2021 | Wu 2020 | 0 | 22 | 0 | 1 |
| Saghafi 2024 | Wu 2020 | 0 | 23 | 0 | 1 |
| Somagutta 2021 | Wu 2020 | 0 | 20 | 0 | 1 |
| Song B 2024 | Wu 2020 | 0 | 12 | 0 | 1 |
| Sudaryadi 2023 | Wu 2020 | 0 | 15 | 0 | 1 |
| Aletreby 2019 | Wu 2021 | 0 | 16 | 0 | 1 |
| Allen 2019 | Wu 2021 | 0 | 13 | 0 | 1 |
| Annane 2015 | Wu 2021 | 0 | 39 | 0 | 1 |
| Annane 2019 | Wu 2021 | 0 | 67 | 0 | 1 |
| Barakakis 2019 | Wu 2021 | 0 | 8 | 0 | 1 |
| Deng 2025 | Wu 2021 | 0 | 21 | 0 | 1 |
| Fang 2019 | Wu 2021 | 0 | 43 | 0 | 1 |
| Fratoni 2020 | Wu 2021 | 0 | 11 | 0 | 1 |
| Li 2025 | Wu 2021 | 0 | 9 | 0 | 1 |
| Lian 2019 | Wu 2021 | 0 | 27 | 0 | 1 |
| Liang 2021 | Wu 2021 | 0 | 56 | 0 | 1 |
| Lin 2019 | Wu 2021 | 0 | 36 | 0 | 1 |
| Lu 2021 | Wu 2021 | 0 | 15 | 0 | 1 |
| Lyu 2020 | Wu 2021 | 0 | 19 | 0 | 1 |
| Ni 2019 | Wu 2021 | 0 | 25 | 0 | 1 |
| Pitre 2024 | Wu 2021 | 0 | 51 | 0 | 1 |
| Rochwerg 2018 | Wu 2021 | 0 | 48 | 0 | 1 |
| Rygard 2018 | Wu 2021 | 0 | 29 | 0 | 1 |
| Siddiqi 2019 | Wu 2021 | 0 | 11 | 0 | 1 |
| Son 2021 | Wu 2021 | 0 | 28 | 0 | 1 |
| Song B 2024 | Wu 2021 | 0 | 6 | 0 | 1 |
| Song Y 2024 | Wu 2021 | 0 | 15 | 0 | 1 |
| Wen 2019 | Wu 2021 | 0 | 24 | 0 | 1 |
| Wu 2020 | Wu 2021 | 0 | 18 | 0 | 1 |
| Assouline 2021 | Wu 2024 | 0 | 11 | 0 | 1 |
| Deng 2024 | Wu 2024 | 0 | 18 | 0 | 1 |

| | | | | | |
|---|---|---|---|---|---|
| Deng 2025 | Wu 2024 | 0 | 18 | 0 | 1 |
| Fratoni 2020 | Wu 2024 | 0 | 8 | 0 | 1 |
| Kato 2023 | Wu 2024 | 0 | 12 | 0 | 1 |
| Lee 2022 | Wu 2024 | 0 | 9 | 0 | 1 |
| Li 2025 | Wu 2024 | 0 | 6 | 0 | 1 |
| Lu 2023 | Wu 2024 | 0 | 11 | 0 | 1 |
| Na 2021 | Wu 2024 | 0 | 13 | 0 | 1 |
| Saghafi 2024 | Wu 2024 | 0 | 14 | 0 | 1 |
| Somagutta 2021 | Wu 2024 | 0 | 11 | 0 | 1 |
| Song B 2024 | Wu 2024 | 0 | 3 | 0 | 1 |
| Sudaryadi 2023 | Wu 2024 | 0 | 6 | 0 | 1 |
| Wu 2020 | Wu 2024 | 0 | 15 | 0 | 1 |
| Wu 2021 | Wu 2024 | 0 | 9 | 0 | 1 |
| Allen 2019 | Xu 2018 | 0 | 16 | 0 | 1 |
| Assouline 2021 | Xu 2018 | 0 | 17 | 0 | 1 |
| Deng 2024 | Xu 2018 | 0 | 24 | 0 | 1 |
| Deng 2025 | Xu 2018 | 0 | 24 | 0 | 1 |
| Kato 2023 | Xu 2018 | 0 | 18 | 0 | 1 |
| Lee 2022 | Xu 2018 | 0 | 15 | 0 | 1 |
| Li 2025 | Xu 2018 | 0 | 12 | 0 | 1 |
| Lu 2023 | Xu 2018 | 0 | 17 | 0 | 1 |
| Na 2021 | Xu 2018 | 0 | 19 | 0 | 1 |
| Saghafi 2024 | Xu 2018 | 0 | 20 | 0 | 1 |
| Somagutta 2021 | Xu 2018 | 0 | 17 | 0 | 1 |
| Song B 2024 | Xu 2018 | 0 | 9 | 0 | 1 |
| Sudaryadi 2023 | Xu 2018 | 0 | 12 | 0 | 1 |
| Wu 2021 | Xu 2018 | 0 | 15 | 0 | 1 |
| Assouline 2021 | Yamamoto 2020 | 0 | 11 | 0 | 1 |
| Deng 2024 | Yamamoto 2020 | 0 | 18 | 0 | 1 |
| Deng 2025 | Yamamoto 2020 | 0 | 18 | 0 | 1 |
| Fratoni 2020 | Yamamoto 2020 | 0 | 8 | 0 | 1 |
| Kato 2023 | Yamamoto 2020 | 0 | 12 | 0 | 1 |
| Lee 2022 | Yamamoto 2020 | 0 | 9 | 0 | 1 |
| Li 2025 | Yamamoto 2020 | 0 | 6 | 0 | 1 |
| Lu 2023 | Yamamoto 2020 | 0 | 11 | 0 | 1 |
| Na 2021 | Yamamoto 2020 | 0 | 13 | 0 | 1 |
| Saghafi 2024 | Yamamoto 2020 | 0 | 14 | 0 | 1 |
| Somagutta 2021 | Yamamoto 2020 | 0 | 11 | 0 | 1 |
| Song B 2024 | Yamamoto 2020 | 0 | 3 | 0 | 1 |
| Sudaryadi 2023 | Yamamoto 2020 | 0 | 6 | 0 | 1 |
| Wu 2020 | Yamamoto 2020 | 0 | 15 | 0 | 1 |
| Wu 2021 | Yamamoto 2020 | 0 | 9 | 0 | 1 |
| Aletreby 2019 | Yang 2020 | 0 | 20 | 0 | 1 |
| Allen 2019 | Yang 2020 | 0 | 17 | 0 | 1 |
| Assouline 2021 | Yang 2020 | 0 | 18 | 0 | 1 |

| | | | | | |
|---|---|---|---|---|---|
| Barakakis 2019 | Yang 2020 | 0 | 12 | 0 | 1 |
| Deng 2024 | Yang 2020 | 0 | 25 | 0 | 1 |
| Fang 2019 | Yang 2020 | 0 | 47 | 0 | 1 |
| Fratoni 2020 | Yang 2020 | 0 | 15 | 0 | 1 |
| Kato 2023 | Yang 2020 | 0 | 19 | 0 | 1 |
| Lee 2022 | Yang 2020 | 0 | 16 | 0 | 1 |
| Li 2025 | Yang 2020 | 0 | 13 | 0 | 1 |
| Lian 2019 | Yang 2020 | 0 | 31 | 0 | 1 |
| Liang 2021 | Yang 2020 | 0 | 60 | 0 | 1 |
| Lin 2019 | Yang 2020 | 0 | 40 | 0 | 1 |
| Lu 2021 | Yang 2020 | 0 | 19 | 0 | 1 |
| Lu 2023 | Yang 2020 | 0 | 18 | 0 | 1 |
| Lyu 2020 | Yang 2020 | 0 | 23 | 0 | 1 |
| Na 2021 | Yang 2020 | 0 | 20 | 0 | 1 |
| Ni 2019 | Yang 2020 | 0 | 29 | 0 | 1 |
| Rygard 2018 | Yang 2020 | 0 | 33 | 0 | 1 |
| Saghafi 2024 | Yang 2020 | 0 | 21 | 0 | 1 |
| Siddiqi 2019 | Yang 2020 | 0 | 15 | 0 | 1 |
| Somagutta 2021 | Yang 2020 | 0 | 18 | 0 | 1 |
| Son 2021 | Yang 2020 | 0 | 32 | 0 | 1 |
| Song B 2024 | Yang 2020 | 0 | 10 | 0 | 1 |
| Sudaryadi 2023 | Yang 2020 | 0 | 13 | 0 | 1 |
| Wen 2019 | Yang 2020 | 0 | 28 | 0 | 1 |
| Wu 2020 | Yang 2020 | 0 | 22 | 0 | 1 |
| Wu 2021 | Yang 2020 | 0 | 16 | 0 | 1 |
| Wu 2024 | Yang 2020 | 0 | 13 | 0 | 1 |
| Xu 2018 | Yang 2020 | 0 | 19 | 0 | 1 |
| Yamamoto 2020 | Yang 2020 | 0 | 13 | 0 | 1 |
| Aletreby 2019 | Yao 2021 | 0 | 18 | 0 | 1 |
| Allen 2019 | Yao 2021 | 0 | 15 | 0 | 1 |
| Annane 2015 | Yao 2021 | 0 | 41 | 0 | 1 |
| Annane 2019 | Yao 2021 | 0 | 69 | 0 | 1 |
| Barakakis 2019 | Yao 2021 | 0 | 10 | 0 | 1 |
| Deng 2025 | Yao 2021 | 0 | 23 | 0 | 1 |
| Fang 2019 | Yao 2021 | 0 | 45 | 0 | 1 |
| Fratoni 2020 | Yao 2021 | 0 | 13 | 0 | 1 |
| Li 2025 | Yao 2021 | 0 | 11 | 0 | 1 |
| Lian 2019 | Yao 2021 | 0 | 29 | 0 | 1 |
| Liang 2021 | Yao 2021 | 0 | 58 | 0 | 1 |
| Lin 2019 | Yao 2021 | 0 | 38 | 0 | 1 |
| Lu 2021 | Yao 2021 | 0 | 17 | 0 | 1 |
| Lyu 2020 | Yao 2021 | 0 | 21 | 0 | 1 |
| Ni 2019 | Yao 2021 | 0 | 27 | 0 | 1 |
| Pitre 2024 | Yao 2021 | 0 | 53 | 0 | 1 |
| Rochwerg 2018 | Yao 2021 | 0 | 50 | 0 | 1 |

| | | | | | |
|---|---|---|---|---|---|
| Rygard 2018 | Yao 2021 | 0 | 31 | 0 | 1 |
| Siddiqi 2019 | Yao 2021 | 0 | 13 | 0 | 1 |
| Son 2021 | Yao 2021 | 0 | 30 | 0 | 1 |
| Song B 2024 | Yao 2021 | 0 | 8 | 0 | 1 |
| Song Y 2024 | Yao 2021 | 0 | 17 | 0 | 1 |
| Wen 2019 | Yao 2021 | 0 | 26 | 0 | 1 |
| Wu 2020 | Yao 2021 | 0 | 20 | 0 | 1 |
| Wu 2024 | Yao 2021 | 0 | 11 | 0 | 1 |
| Xu 2018 | Yao 2021 | 0 | 17 | 0 | 1 |
| Yamamoto 2020 | Yao 2021 | 0 | 11 | 0 | 1 |
| Yang 2020 | Yao 2021 | 0 | 18 | 0 | 1 |
| Aletreby 2019 | Zayed 2022 | 0 | 17 | 0 | 1 |
| Allen 2019 | Zayed 2022 | 0 | 14 | 0 | 1 |
| Annane 2015 | Zayed 2022 | 0 | 40 | 0 | 1 |
| Annane 2019 | Zayed 2022 | 0 | 68 | 0 | 1 |
| Barakakis 2019 | Zayed 2022 | 0 | 9 | 0 | 1 |
| Deng 2025 | Zayed 2022 | 0 | 22 | 0 | 1 |
| Fang 2019 | Zayed 2022 | 0 | 44 | 0 | 1 |
| Fratoni 2020 | Zayed 2022 | 0 | 12 | 0 | 1 |
| Li 2025 | Zayed 2022 | 0 | 10 | 0 | 1 |
| Lian 2019 | Zayed 2022 | 0 | 28 | 0 | 1 |
| Liang 2021 | Zayed 2022 | 0 | 57 | 0 | 1 |
| Lin 2019 | Zayed 2022 | 0 | 37 | 0 | 1 |
| Lu 2021 | Zayed 2022 | 0 | 16 | 0 | 1 |
| Lyu 2020 | Zayed 2022 | 0 | 20 | 0 | 1 |
| Ni 2019 | Zayed 2022 | 0 | 26 | 0 | 1 |
| Pitre 2024 | Zayed 2022 | 0 | 52 | 0 | 1 |
| Rochwerg 2018 | Zayed 2022 | 0 | 49 | 0 | 1 |
| Rygard 2018 | Zayed 2022 | 0 | 30 | 0 | 1 |
| Siddiqi 2019 | Zayed 2022 | 0 | 12 | 0 | 1 |
| Son 2021 | Zayed 2022 | 0 | 29 | 0 | 1 |
| Song B 2024 | Zayed 2022 | 0 | 7 | 0 | 1 |
| Song Y 2024 | Zayed 2022 | 0 | 16 | 0 | 1 |
| Wen 2019 | Zayed 2022 | 0 | 25 | 0 | 1 |
| Wu 2020 | Zayed 2022 | 0 | 19 | 0 | 1 |
| Wu 2024 | Zayed 2022 | 0 | 10 | 0 | 1 |
| Xu 2018 | Zayed 2022 | 0 | 16 | 0 | 1 |
| Yamamoto 2020 | Zayed 2022 | 0 | 10 | 0 | 1 |
| Yang 2020 | Zayed 2022 | 0 | 17 | 0 | 1 |
| Assouline 2021 | Zhou 2018 | 0 | 25 | 0 | 1 |
| Deng 2024 | Zhou 2018 | 0 | 32 | 0 | 1 |
| Deng 2025 | Zhou 2018 | 0 | 32 | 0 | 1 |
| Kato 2023 | Zhou 2018 | 0 | 26 | 0 | 1 |
| Lee 2022 | Zhou 2018 | 0 | 23 | 0 | 1 |
| Li 2025 | Zhou 2018 | 0 | 20 | 0 | 1 |

| | | | | | |
|---|---|---|---|---|---|
| Lu 2023 | Zhou 2018 | 0 | 25 | 0 | 1 |
| Na 2021 | Zhou 2018 | 0 | 27 | 0 | 1 |
| Saghafi 2024 | Zhou 2018 | 0 | 28 | 0 | 1 |
| Somagutta 2021 | Zhou 2018 | 0 | 25 | 0 | 1 |
| Song B 2024 | Zhou 2018 | 0 | 17 | 0 | 1 |
| Sudaryadi 2023 | Zhou 2018 | 0 | 20 | 0 | 1 |
| Wu 2021 | Zhou 2018 | 0 | 23 | 0 | 1 |
| Yang 2020 | Zhou 2018 | 0 | 27 | 0 | 1 |
| Yao 2021 | Zhou 2018 | 0 | 25 | 0 | 1 |
| Zayed 2022 | Zhou 2018 | 0 | 24 | 0 | 1 |

#### Supplementary Table S10: Hierarchical average-linkage clustering of the Jaccard distance matrix (1 − J)

| SR | Cluster rank | Cluster number | Order dendrogram |
|---|---|---|---|
| Annane 2015 | 1 | 3 | 18 |
| Pitre 2024 | 1 | 3 | 19 |
| Rochwerg 2018 | 1 | 3 | 20 |
| Fang 2019 | 1 | 3 | 21 |
| Lin 2019 | 1 | 3 | 22 |
| Liang 2021 | 1 | 3 | 23 |
| Annane 2019 | 1 | 3 | 24 |
| Annane 2025 | 1 | 3 | 25 |
| Kato 2023 | 2 | 4 | 6 |
| Lu 2023 | 2 | 4 | 7 |
| Wu 2021 | 2 | 4 | 8 |
| Somagutta 2021 | 2 | 4 | 9 |
| Yao 2021 | 2 | 4 | 10 |
| Saghafi 2024 | 2 | 4 | 11 |
| Assouline 2021 | 2 | 4 | 12 |
| Na 2021 | 2 | 4 | 13 |
| Zhou 2018 | 3 | 11 | 30 |
| Lian 2019 | 3 | 11 | 31 |
| Wen 2019 | 3 | 11 | 32 |
| Ni 2019 | 3 | 11 | 33 |
| Lyu 2020 | 3 | 11 | 34 |
| Wu 2020 | 3 | 11 | 35 |
| Barakakis 2019 | 4 | 5 | 36 |
| Wu 2024 | 4 | 5 | 37 |
| Yamamoto 2020 | 4 | 5 | 38 |
| Aletreby 2019 | 5 | 1 | 41 |
| Siddiqi 2019 | 5 | 1 | 42 |
| Deng 2025 | 6 | 7 | 15 |
| Yang 2020 | 6 | 7 | 16 |
| Lee 2022 | 7 | 9 | 3 |
| Zayed 2022 | 7 | 9 | 4 |
| Lu 2021 | 8 | 12 | 26 |
| Xu 2018 | 8 | 12 | 27 |
| Allen 2019 | 9 | 2 | 39 |
| Deng 2024 | 10 | 6 | 5 |
| Fratoni 2020 | 11 | 8 | 17 |
| Li 2025 | 12 | 10 | 14 |
| Rygard 2018 | 13 | 13 | 29 |
| Son 2021 | 14 | 14 | 28 |
| Song B 2024 | 15 | 15 | 1 |

| | | | |
|---|---|---|---|
| Song Y 2024 | 16 | 16 | 40 |
| Sudaryadi 2023 | 17 | 17 | 2 |

### Supplementary Table S11: PICO profiling of systematic reviews as defined by the study (n = 42)

PICO domains with categories:

**Population – Age (PA1-4):** Adults (PA1); Children (PA2); Mixed-age populations (PA3); Age not defined (PA4).

**Population – Target (PD1-4):** Sepsis irrespective of shock status (PD1); Sepsis without shock (PD2); Septic shock only (PD3); Other (PD4)

**Intervention – Corticosteroid Type (IT1-5):** Any/multiple corticosteroids without restriction (IT1); Hydrocortisone (IT2); Dexamethasone (IT3); Methylprednisolone (IT4); Other corticosteroids (IT5).

**Intervention – Dose (ID0-2):** Dose not defined (ID0); Low-dose regimen (ID1); High-dose regimen (ID2).

**Intervention – Timing and Schedule (ITM0-3):** Timing or schedule not defined (ITM0); Standard administration schedule (ITM1); Specific timing focus (ITM2); Specific schedule focus (ITM3).

**Intervention – Combination Strategy (IC0-4):** Monotherapy or combination not defined (IC0); Monotherapy (IC1); Hydrocortisone plus fludrocortisone (HC+FC) (IC2); hydrocortisone, ascorbic acid, and thiamine (HAT) (IC3); Other predefined combinations (IC4).

**Comparator (C1-3):** Standard care, placebo, or no corticosteroid (C1); Active corticosteroid comparator (C2); Other, unclear, or not defined comparator (C3).

**PICO profiles of systematic reviews summarized according to PICO domains and with structured clinical ordering: 1. Intervention combination strategy, 2. Corticosteroid type, 3. Population target, 4. Age group, 5. Intervention timing and schedule**

| **SR ID** | **PICO profiles (summarized)** | | | | | | | |
|---|---|---|---|---|---|---|---|---|
| | I_combination_strategy (IC0-IC4) | I_type (IT1-5) | P_target (PD1-PD4) | P_age (PA1-PA4) | I_timing (ITM0-3) | I_dose (ID0-2) | Comparator (C1-3) | Clinical rank |
| Assouline 2021 | HAT | HC | Sepsis±Shock | Adults | TS_ND | DoseND | C1+C2 | 1 |

| | | | | | | | | |
|---|---|---|---|---|---|---|---|---|
| Deng 2024 | HAT | HC | Sepsis±Shock | Adults | TS_ND | DoseND | C1+C2 | 2 |
| Lu 2023 | HAT | HC | Sepsis±Shock | Adults | TS_Standard | DoseND | C1 | 3 |
| Na 2021 | HAT | HC | Sepsis±Shock | Adults | TS_ND | DoseND | C1+C2 | 4 |
| Saghafi 2024 | HAT | HC | Sepsis±Shock | Adults | TS_ND | DoseND | C3 | 5 |
| Wu 2021 | HAT | HC | Sepsis±Shock | Adults | TS_ND | DoseND | C1+C2 | 6 |
| Zayed 2022 | HAT | HC | Sepsis±Shock | Adults | TS_ND | DoseND | C3 | 7 |
| Lee 2022 | HAT | HC | Sepsis±Shock | AgeND | TS_ND | DoseND | C1 | 8 |
| Yao 2021 | HAT | HC | Sepsis±Shock | AgeND | TS_ND | DoseND | C1 | 9 |
| Somagutta 2021 | HAT | HC | ShockOnly | Adults | TS_ND | DoseND | C1 | 10 |
| Sudaryadi 2023 | HAT | HC | ShockOnly | Adults | TS_ND | DoseND | C2 | 11 |
| Barakakis 2019 | HC+FC | Any/Multiple | ShockOnly | Adults | TS_ND | Low | C1 | 12 |
| Yamamoto 2020 | HC+FC | HC | ShockOnly | Adults | TS_ND | DoseND | C1+C2 | 13 |
| Wu 2024 | HC+FC | HC | ShockOnly | AgeND | TS_ND | DoseND | C1+C2 | 14 |
| Annane 2025 | Mono+HAT | Any/Multiple | Sepsis±Shock | AnyAge(Mixed) | TS_Standard+ScheduleFocus | Low+High | C1+C2 | 15 |
| Kato 2023 | Mono+HAT | HC | Sepsis±Shock | AgeND | TS_ND | DoseND | C3 | 16 |
| Siddiqi 2019 | Mono+HC+FC | HC | ShockOnly | Adults | TS_ND | DoseND | C1 | 17 |
| Pitre 2024 | Mono+HC+FC+HAT | Any/Multiple | Sepsis±Shock | AnyAge(Mixed) | TS_ND | DoseND | C1 | 18 |
| Zhou 2018 | Mono+HC+FC+HAT+OtherCombo | Any/Multiple | ShockOnly | Adults | TS_Standard | Low+High | C1 | 19 |
| Aletreby 2019 | Mono+HC+FC+OtherCombo | HC | Sepsis±Shock | Adults | TS_Standard | Low+High | C1 | 20 |

| | | | | | | | | |
|---|---|---|---|---|---|---|---|---|
| Allen 2019 | Mono/ComboND | Any/Multiple | Sepsis±Shock | Adults | TS_ND | DoseND | C3 | 21 |
| Fang 2019 | Mono/ComboND | Any/Multiple | Sepsis±Shock | Adults | TS_ND | DoseND | C1 | 22 |
| Liang 2021 | Mono/ComboND | Any/Multiple | Sepsis±Shock | Adults | TS_ND | DoseND | C1 | 23 |
| Lin 2019 | Mono/ComboND | Any/Multiple | Sepsis±Shock | Adults | TS_ND | DoseND | C1 | 24 |
| Ni 2019 | Mono/ComboND | Any/Multiple | Sepsis±Shock | Adults | TS_ND | DoseND | C1 | 25 |
| Son 2021 | Mono/ComboND | Any/Multiple | Sepsis±Shock | Adults | TS_ND | DoseND | C1 | 26 |
| Song Y 2024 | Mono/ComboND | Any/Multiple | Sepsis±Shock | AgeND | TS_ND | DoseND | C1 | 27 |
| Annane 2015 | Mono/ComboND | Any/Multiple | Sepsis±Shock | AnyAge(Mixed) | TS_Standard | Low+High | C1 | 28 |
| Annane 2019 | Mono/ComboND | Any/Multiple | Sepsis±Shock | AnyAge(Mixed) | TS_Standard+ScheduleFocus | Low+High | C1+C2 | 29 |
| Rochwerg 2018 | Mono/ComboND | Any/Multiple | Sepsis±Shock | AnyAge(Mixed) | TS_ND | DoseND | C1 | 30 |
| Deng 2025 | Mono/ComboND | Any/Multiple | Sepsis±Shock+Other | Children | TS_ND | DoseND | C1 | 31 |
| Lian 2019 | Mono/ComboND | Any/Multiple | ShockOnly | Adults | TS_ND | DoseND | C1 | 32 |
| Lu 2021 | Mono/ComboND | Any/Multiple | ShockOnly | Adults | TS_Standard | Low+High | C1 | 33 |
| Rygard 2018 | Mono/ComboND | Any/Multiple | ShockOnly | Adults | TS_ND | Low+High | C1 | 34 |
| Wen 2019 | Mono/ComboND | Any/Multiple | ShockOnly | Adults | TS_ND | DoseND | C3 | 35 |
| Xu 2018 | Mono/ComboND | Any/Multiple | ShockOnly | Adults | TS_Standard | Low | C3 | 36 |
| Yang 2020 | Mono/ComboND | Any/Multiple | ShockOnly | Children | TS_ND | DoseND | C3 | 37 |
| Fratoni 2020 | Mono/ComboND | Dex | Sepsis±Shock | Adults | TS_Standard | Low+High | C1 | 38 |
| Li 2025 | Mono/ComboND | HC | ShockOnly | Adults | ScheduleFocus | DoseND | C2 | 39 |

| | | | | | | | | |
|---|---|---|---|---|---|---|---|---|
| Lyu 2020 | Mono/ComboND | HC | ShockOnly | Adults | TS_ND | Low | C1 | 40 |
| Song B 2024 | Mono/ComboND | HC | ShockOnly | Adults | TimingFocus | Low | C2 | 41 |
| Wu 2020 | Mono/ComboND | HC | ShockOnly | AgeND | TS_ND | Low | C1 | 42 |

**PICO profiles of systematic reviews: Binary coding variables (0/1)**

| SR ID | PICO Domains - Binary PICO coding variables (0/1) | | | | | | | | | | | | | | | | | | | | | | | | | | | | |
|---|---|---|---|---|---|---|---|---|---|---|---|---|---|---|---|---|---|---|---|---|---|---|---|---|---|---|---|---|---|
| | PA1_Adults | PA2_Children | PA3_Mixed | PA4_AgeNotDefined | PD1_Sepsis_any_severity | PD2_Sepsis_without_shock | PD3_Septic_shock | PD4_Other | IT1_AnyMixed_corticosteroid | IT2_Hydrocortisone | IT3_Dexamethasone | IT4_Methylprednisolone | IT5_Other_corticosteroid | ID0_DoseNotDefined | ID1_Low_dose | ID2_High_dose | ITM0_TimingScheduleNotDefined | ITM1_StandardTimingSchedule | ITM2_Specific_timing | ITM3_Specific_schedule | IC0_MonoCombiNotdefined | IC1_Monotherapy | IC2_Hydrocortisone_Fludrocortisone | IC3_HAT | IC4_Other_combination | C1_StandardCare_Placebo_NoCorticosteroid | C2_Active_corticosteroid | C3_Other_UnclearNotDefined | O_Mortality_reported |
| Assouline 2021 | 1 | 0 | 0 | 0 | 1 | 0 | 0 | 0 | 0 | 1 | 0 | 0 | 0 | 1 | 0 | 0 | 1 | 0 | 0 | 0 | 0 | 0 | 0 | 1 | 0 | 1 | 1 | 0 | 1 |
| Deng 2024 | 1 | 0 | 0 | 0 | 1 | 0 | 0 | 0 | 0 | 1 | 0 | 0 | 0 | 1 | 0 | 0 | 1 | 0 | 0 | 0 | 0 | 0 | 0 | 1 | 0 | 1 | 1 | 0 | 1 |
| Lu 2023 | 1 | 0 | 0 | 0 | 1 | 0 | 0 | 0 | 0 | 1 | 0 | 0 | 0 | 1 | 0 | 0 | 0 | 1 | 0 | 0 | 0 | 0 | 0 | 1 | 0 | 1 | 0 | 0 | 1 |
| Na 2021 | 1 | 0 | 0 | 0 | 1 | 0 | 0 | 0 | 0 | 1 | 0 | 0 | 0 | 1 | 0 | 0 | 1 | 0 | 0 | 0 | 0 | 0 | 0 | 1 | 0 | 1 | 1 | 0 | 1 |
| Saghafi 2024 | 1 | 0 | 0 | 0 | 1 | 0 | 0 | 0 | 0 | 1 | 0 | 0 | 0 | 1 | 0 | 0 | 1 | 0 | 0 | 0 | 0 | 0 | 0 | 1 | 0 | 0 | 0 | 1 | 1 |
| Wu 2021 | 1 | 0 | 0 | 0 | 1 | 0 | 0 | 0 | 0 | 1 | 0 | 0 | 0 | 1 | 0 | 0 | 1 | 0 | 0 | 0 | 0 | 0 | 0 | 1 | 0 | 1 | 1 | 0 | 1 |
| Zayed 2022 | 1 | 0 | 0 | 0 | 1 | 0 | 0 | 0 | 0 | 1 | 0 | 0 | 0 | 1 | 0 | 0 | 1 | 0 | 0 | 0 | 0 | 0 | 0 | 1 | 0 | 0 | 0 | 1 | 1 |
| Lee 2022 | 0 | 0 | 0 | 1 | 1 | 0 | 0 | 0 | 0 | 1 | 0 | 0 | 0 | 1 | 0 | 0 | 1 | 0 | 0 | 0 | 0 | 0 | 0 | 1 | 0 | 1 | 0 | 0 | 1 |
| Yao 2021 | 0 | 0 | 0 | 1 | 1 | 0 | 0 | 0 | 0 | 1 | 0 | 0 | 0 | 1 | 0 | 0 | 1 | 0 | 0 | 0 | 0 | 0 | 0 | 1 | 0 | 1 | 0 | 0 | 1 |
| Somagutta 2021 | 1 | 0 | 0 | 0 | 0 | 0 | 1 | 0 | 0 | 1 | 0 | 0 | 0 | 1 | 0 | 0 | 1 | 0 | 0 | 0 | 0 | 0 | 0 | 1 | 0 | 1 | 0 | 0 | 1 |
| Sudaryadi 2023 | 1 | 0 | 0 | 0 | 0 | 0 | 1 | 0 | 0 | 1 | 0 | 0 | 0 | 1 | 0 | 0 | 1 | 0 | 0 | 0 | 0 | 0 | 0 | 1 | 0 | 0 | 1 | 0 | 1 |
| Barakakis 2019 | 1 | 0 | 0 | 0 | 0 | 0 | 1 | 0 | 1 | 0 | 0 | 0 | 0 | 0 | 1 | 0 | 1 | 0 | 0 | 0 | 0 | 0 | 1 | 0 | 0 | 1 | 0 | 0 | 1 |
| Yamamoto 2020 | 1 | 0 | 0 | 0 | 0 | 0 | 1 | 0 | 0 | 1 | 0 | 0 | 0 | 1 | 0 | 0 | 1 | 0 | 0 | 0 | 0 | 0 | 1 | 0 | 0 | 1 | 1 | 0 | 1 |
| Wu 2024 | 0 | 0 | 0 | 1 | 0 | 0 | 1 | 0 | 0 | 1 | 0 | 0 | 0 | 1 | 0 | 0 | 1 | 0 | 0 | 0 | 0 | 0 | 1 | 0 | 0 | 1 | 1 | 0 | 1 |
| Annane 2025 | 0 | 0 | 1 | 0 | 1 | 0 | 0 | 0 | 1 | 0 | 0 | 0 | 0 | 0 | 1 | 1 | 0 | 1 | 0 | 1 | 0 | 1 | 0 | 1 | 0 | 1 | 1 | 0 | 1 |
| Kato 2023 | 0 | 0 | 0 | 1 | 1 | 0 | 0 | 0 | 0 | 1 | 0 | 0 | 0 | 1 | 0 | 0 | 1 | 0 | 0 | 0 | 0 | 1 | 0 | 1 | 0 | 0 | 0 | 1 | 1 |
| Siddiqi 2019 | 1 | 0 | 0 | 0 | 0 | 0 | 1 | 0 | 0 | 1 | 0 | 0 | 0 | 1 | 0 | 0 | 1 | 0 | 0 | 0 | 0 | 1 | 1 | 0 | 0 | 1 | 0 | 0 | 1 |
| Pitre 2024 | 0 | 0 | 1 | 0 | 1 | 0 | 0 | 0 | 1 | 0 | 0 | 0 | 0 | 1 | 0 | 0 | 1 | 0 | 0 | 0 | 0 | 1 | 1 | 1 | 0 | 1 | 0 | 0 | 1 |
| Zhou 2018 | 1 | 0 | 0 | 0 | 0 | 0 | 1 | 0 | 1 | 0 | 0 | 0 | 0 | 0 | 1 | 1 | 0 | 1 | 0 | 0 | 0 | 1 | 1 | 1 | 1 | 1 | 0 | 0 | 1 |
| Aletreby 2019 | 1 | 0 | 0 | 0 | 1 | 0 | 0 | 0 | 0 | 1 | 0 | 0 | 0 | 0 | 1 | 1 | 0 | 1 | 0 | 0 | 0 | 1 | 1 | 0 | 1 | 1 | 0 | 0 | 1 |
| Allen 2019 | 1 | 0 | 0 | 0 | 1 | 0 | 0 | 0 | 1 | 0 | 0 | 0 | 0 | 1 | 0 | 0 | 1 | 0 | 0 | 0 | 1 | 0 | 0 | 0 | 0 | 0 | 0 | 1 | 1 |
| Fang 2019 | 1 | 0 | 0 | 0 | 1 | 0 | 0 | 0 | 1 | 0 | 0 | 0 | 0 | 1 | 0 | 0 | 1 | 0 | 0 | 0 | 1 | 0 | 0 | 0 | 0 | 1 | 0 | 0 | 1 |
| Liang 2021 | 1 | 0 | 0 | 0 | 1 | 0 | 0 | 0 | 1 | 0 | 0 | 0 | 0 | 1 | 0 | 0 | 1 | 0 | 0 | 0 | 1 | 0 | 0 | 0 | 0 | 1 | 0 | 0 | 1 |
| Lin 2019 | 1 | 0 | 0 | 0 | 1 | 0 | 0 | 0 | 1 | 0 | 0 | 0 | 0 | 1 | 0 | 0 | 1 | 0 | 0 | 0 | 1 | 0 | 0 | 0 | 0 | 1 | 0 | 0 | 1 |
| Ni 2019 | 1 | 0 | 0 | 0 | 1 | 0 | 0 | 0 | 1 | 0 | 0 | 0 | 0 | 1 | 0 | 0 | 1 | 0 | 0 | 0 | 1 | 0 | 0 | 0 | 0 | 1 | 0 | 0 | 1 |
| Son 2021 | 1 | 0 | 0 | 0 | 1 | 0 | 0 | 0 | 1 | 0 | 0 | 0 | 0 | 1 | 0 | 0 | 1 | 0 | 0 | 0 | 1 | 0 | 0 | 0 | 0 | 1 | 0 | 0 | 1 |
| Song Y 2024 | 0 | 0 | 0 | 1 | 1 | 0 | 0 | 0 | 1 | 0 | 0 | 0 | 0 | 1 | 0 | 0 | 1 | 0 | 0 | 0 | 1 | 0 | 0 | 0 | 0 | 1 | 0 | 0 | 1 |
| Annane 2015 | 0 | 0 | 1 | 0 | 1 | 0 | 0 | 0 | 1 | 0 | 0 | 0 | 0 | 0 | 1 | 1 | 0 | 1 | 0 | 0 | 1 | 0 | 0 | 0 | 0 | 1 | 0 | 0 | 1 |
| Annane 2019 | 0 | 0 | 1 | 0 | 1 | 0 | 0 | 0 | 1 | 0 | 0 | 0 | 0 | 0 | 1 | 1 | 0 | 1 | 0 | 1 | 1 | 0 | 0 | 0 | 0 | 1 | 1 | 0 | 1 |
| Rochwerg 2018 | 0 | 0 | 1 | 0 | 1 | 0 | 0 | 0 | 1 | 0 | 0 | 0 | 0 | 1 | 0 | 0 | 1 | 0 | 0 | 0 | 1 | 0 | 0 | 0 | 0 | 1 | 0 | 0 | 1 |
| Deng 2025 | 0 | 1 | 0 | 0 | 1 | 0 | 0 | 1 | 1 | 0 | 0 | 0 | 0 | 1 | 0 | 0 | 1 | 0 | 0 | 0 | 1 | 0 | 0 | 0 | 0 | 1 | 0 | 0 | 1 |
| Lian 2019 | 1 | 0 | 0 | 0 | 0 | 0 | 1 | 0 | 1 | 0 | 0 | 0 | 0 | 1 | 0 | 0 | 1 | 0 | 0 | 0 | 1 | 0 | 0 | 0 | 0 | 1 | 0 | 0 | 1 |
| Lu 2021 | 1 | 0 | 0 | 0 | 0 | 0 | 1 | 0 | 1 | 0 | 0 | 0 | 0 | 0 | 1 | 1 | 0 | 1 | 0 | 0 | 1 | 0 | 0 | 0 | 0 | 1 | 0 | 0 | 1 |
| Rygard 2018 | 1 | 0 | 0 | 0 | 0 | 0 | 1 | 0 | 1 | 0 | 0 | 0 | 0 | 0 | 1 | 1 | 1 | 0 | 0 | 0 | 1 | 0 | 0 | 0 | 0 | 1 | 0 | 0 | 1 |
| Wen 2019 | 1 | 0 | 0 | 0 | 0 | 0 | 1 | 0 | 1 | 0 | 0 | 0 | 0 | 1 | 0 | 0 | 1 | 0 | 0 | 0 | 1 | 0 | 0 | 0 | 0 | 0 | 0 | 1 | 1 |
| Xu 2018 | 1 | 0 | 0 | 0 | 0 | 0 | 1 | 0 | 1 | 0 | 0 | 0 | 0 | 0 | 1 | 0 | 0 | 1 | 0 | 0 | 1 | 0 | 0 | 0 | 0 | 0 | 0 | 1 | 1 |
| Yang 2020 | 0 | 1 | 0 | 0 | 0 | 0 | 1 | 0 | 1 | 0 | 0 | 0 | 0 | 1 | 0 | 0 | 1 | 0 | 0 | 0 | 1 | 0 | 0 | 0 | 0 | 0 | 0 | 1 | 1 |
| Fratoni 2020 | 1 | 0 | 0 | 0 | 1 | 0 | 0 | 0 | 0 | 0 | 1 | 0 | 0 | 0 | 1 | 1 | 0 | 1 | 0 | 0 | 1 | 0 | 0 | 0 | 0 | 1 | 0 | 0 | 1 |
| Li 2025 | 1 | 0 | 0 | 0 | 0 | 0 | 1 | 0 | 0 | 1 | 0 | 0 | 0 | 1 | 0 | 0 | 0 | 0 | 0 | 1 | 1 | 0 | 0 | 0 | 0 | 0 | 1 | 0 | 1 |
| Lyu 2020 | 1 | 0 | 0 | 0 | 0 | 0 | 1 | 0 | 0 | 1 | 0 | 0 | 0 | 0 | 1 | 0 | 1 | 0 | 0 | 0 | 1 | 0 | 0 | 0 | 0 | 1 | 0 | 0 | 1 |
| Song B 2024 | 1 | 0 | 0 | 0 | 0 | 0 | 1 | 0 | 0 | 1 | 0 | 0 | 0 | 0 | 1 | 0 | 0 | 0 | 1 | 0 | 1 | 0 | 0 | 0 | 0 | 0 | 1 | 0 | 1 |
| Wu 2020 | 0 | 0 | 0 | 1 | 0 | 0 | 1 | 0 | 0 | 1 | 0 | 0 | 0 | 0 | 1 | 0 | 1 | 0 | 0 | 0 | 1 | 0 | 0 | 0 | 0 | 1 | 0 | 0 | 1 |

### Supplementary Table S12: PICO profiling of randomized controlled trials as defined by the study (n = 121)

PICO domains with categories:

**Population – Age (PA1-4):** Adults (PA1); Children (PA2); Mixed-age populations (PA3); Age not defined (PA4).

**Population – Target (PD1-4):** Sepsis irrespective of shock status (PD1); Sepsis without shock (PD2); Septic shock only (PD3); Other (PD4)

**Intervention – Corticosteroid Type (IT1-5):** Any/multiple corticosteroids without restriction (IT1); Hydrocortisone (IT2); Dexamethasone (IT3); Methylprednisolone (IT4); Other corticosteroids (IT5).

**Intervention – Dose (ID0-2):** Dose not defined (ID0); Low-dose regimen (ID1); High-dose regimen (ID2).

**Intervention – Timing and Schedule (ITM0-3):** Timing or schedule not defined (ITM0); Standard administration schedule (ITM1); Specific timing focus (ITM2); Specific schedule focus (ITM3).

**Intervention – Combination Strategy (IC0-4):** Monotherapy or combination not defined (IC0); Monotherapy (IC1); Hydrocortisone plus fludrocortisone (HC+FC) (IC2); hydrocortisone, ascorbic acid, and thiamine (HAT) (IC3); Other predefined combinations (IC4).

**Comparator (C1-3):** Standard care, placebo, or no corticosteroid (C1); Active corticosteroid comparator (C2); Other, unclear, or not defined comparator (C3).

**PICO profiles of randomized controlled trails: Binary coding variables (0/1)**

22 RCTs investigated populations other than sepsis: Angus 2020, Blum 2015, Cicarelli 2006, Confalonieri 2005, Corral Gudino 2021, CSG 1963, Edalatifard 2020, Fernandez Serrano 2011, Ghanei 2021, Horby 2021, Klastersky 1971, McHardy 1972, Meduri 2007, Meijvis 2011, Nafae 2013, Nagy 2013, Rezk 2013, Snijders 2010, Tagaro 2017, Torres 2015, Villar 2020, Wittermans 2020

RCTs

| rct_id | first_author | year | full text | P (target as described in the study) | P (age) | I | C | design | PA1_Adults | PA2_Children | PA3_Mixed | PA4_AgeNotDefined | PD1_Sepsis_any_severity | PD2_Sepsis_without_shock | PD3_Septic_shock | PD4_Other | IT1_AnyMixed_corticosteroid | IT2_Hydrocortisone | IT3_Dexamethasone | IT4_Methylprednisolone | IT5_Other_corticosteroid | ID0_DoseNotDefined | ID1_Low_dose | ID2_High_dose | ITM0_TimingScheduleNotDefined | ITM1_StandardTimingSchedule | ITM2_Specific_timing | ITM3_Specific_schedule | IC0_MonoCombiNotdefined | IC1_Monotherapy | IC2_Hydrocortisone_Fludrocortisone | IC3_HAT | IC4_Other_combination | C1_StandardCare_Placebo_NoCorticosteroid | C2_Active_corticosteroid | C3_Other_UnclearNotDefined | O_Mortality_reported |
|---|---|---|---|---|---|---|---|---|---|---|---|---|---|---|---|---|---|---|---|---|---|---|---|---|---|---|---|---|---|---|---|---|---|---|---|---|---|
| Aboab 2008 | Aboab | 2008 | y | septic shock | adults | hydrocortiso | placebo | RCT | 1 | 0 | 0 | 0 | 0 | 0 | 1 | 0 | 0 | 1 | 0 | 0 | 0 | 0 | 1 | 0 | 0 | 1 | 0 | 0 | 0 | 0 | 1 | 0 | 0 | 1 | 0 | 0 | 1 |
| Agarwal 2022 | Agarwal | 2022 | y | septic shock | geriatric | early low-do | SoC with hyd | RCT | 1 | 0 | 0 | 0 | 0 | 0 | 1 | 0 | 0 | 1 | 0 | 0 | 0 | 0 | 1 | 0 | 0 | 0 | 1 | 0 | 0 | 1 | 0 | 0 | 0 | 0 | 1 | 0 | 1 |
| Angus 2020 | Angus | 2020 | y | COVID-19 (severe) | adults | hydrocortiso | no treatment | RCT | 1 | 0 | 0 | 0 | 0 | 0 | 0 | 1 | 0 | 1 | 0 | 0 | 0 | 0 | 1 | 0 | 0 | 1 | 0 | 1 | 0 | 1 | 0 | 0 | 0 | 1 | 1 | 0 | 1 |
| Annane 2002 | Annane | 2002 | y | septic shock | adults | hydrocortiso | placebo | RCT | 1 | 0 | 0 | 0 | 0 | 0 | 1 | 0 | 0 | 1 | 0 | 0 | 0 | 0 | 1 | 0 | 0 | 1 | 0 | 0 | 0 | 0 | 1 | 0 | 0 | 1 | 0 | 0 | 1 |
| Annane 2010 | Annane | 2010 | y | septic shock | adults | fludrocortiso | hydrocortiso | RCT | 1 | 0 | 0 | 0 | 0 | 0 | 1 | 0 | 0 | 1 | 0 | 0 | 0 | 0 | 1 | 0 | 0 | 1 | 0 | 0 | 0 | 0 | 1 | 0 | 0 | 0 | 1 | 0 | 1 |
| Annane 2018 | Annane | 2018 | y | septic shock | adults | hydrocortiso | placebo | RCT | 1 | 0 | 0 | 0 | 0 | 0 | 1 | 0 | 0 | 1 | 0 | 0 | 0 | 0 | 1 | 0 | 0 | 1 | 0 | 0 | 0 | 0 | 1 | 0 | 0 | 1 | 0 | 0 | 1 |
| Arabi 2010 | Arabi | 2010 | y | septic shock (cirrhosis | adults | hydrocortiso | placebo | RCT | 1 | 0 | 0 | 0 | 0 | 0 | 1 | 0 | 0 | 1 | 0 | 0 | 0 | 0 | 1 | 0 | 0 | 1 | 0 | 0 | 0 | 1 | 0 | 0 | 0 | 1 | 0 | 0 | 1 |
| Balakrishnan 2018 | Balakrishnan | 2018 | y | septic shock | adults | HAT | placebo | RCT | 1 | 0 | 0 | 0 | 0 | 0 | 1 | 0 | 0 | 1 | 0 | 0 | 0 | 0 | 1 | 0 | 0 | 1 | 0 | 0 | 0 | 0 | 0 | 1 | 0 | 1 | 0 | 0 | 1 |
| Birudaraju 2022 | Birudaraju | 2022 | y | severe sepsis and sept | adults | solumedrol | placebo | RCT | 1 | 0 | 0 | 0 | 1 | 0 | 0 | 0 | 0 | 0 | 0 | 1 | 0 | 0 | 1 | 0 | 0 | 1 | 0 | 0 | 0 | 1 | 0 | 0 | 0 | 1 | 0 | 0 | 1 |
| Blum 2015 | Blum | 2015 | y | CAP (all severity classe | adults | prednisone | placebo | RCT | 1 | 0 | 0 | 0 | 0 | 0 | 0 | 1 | 0 | 0 | 0 | 0 | 1 | 0 | 1 | 0 | 0 | 1 | 0 | 0 | 0 | 1 | 0 | 0 | 0 | 1 | 0 | 0 | 1 |
| Bollaert 1998 | Bollaert | 1998 | y | septic shock | adults | hydrocortiso | placebo | RCT | 1 | 0 | 0 | 0 | 0 | 0 | 1 | 0 | 0 | 1 | 0 | 0 | 0 | 0 | 1 | 0 | 0 | 1 | 0 | 0 | 1 | 0 | 0 | 0 | 0 | 1 | 0 | 0 | 1 |
| Bone 1987 | Bone | 1987 | y | severe sepsis and sept | adults | methylpredn | placebo | RCT | 1 | 0 | 0 | 0 | 1 | 0 | 0 | 0 | 0 | 0 | 0 | 1 | 0 | 0 | 0 | 1 | 0 | 1 | 0 | 0 | 1 | 0 | 0 | 0 | 0 | 1 | 0 | 0 | 1 |
| Branco 2014 | Branco | 2014 | y | septic shock | children | hydrocortiso | placebo | RCT | 0 | 1 | 0 | 0 | 0 | 0 | 1 | 0 | 0 | 1 | 0 | 0 | 0 | 0 | 1 | 0 | 0 | 1 | 0 | 0 | 0 | 1 | 0 | 0 | 0 | 1 | 0 | 0 | 1 |
| Briegel 1999 | Briegel | 1999 | y | septic shock | NI | hydrocortiso | placebo | RCT | 0 | 0 | 0 | 1 | 0 | 0 | 1 | 0 | 0 | 1 | 0 | 0 | 0 | 0 | 1 | 0 | 0 | 1 | 0 | 0 | 1 | 0 | 0 | 0 | 0 | 1 | 0 | 0 | 1 |
| Briegel 2001 | Briegel | 2001 | y | septic shock | NI | hydrocortiso | placebo | RCT | 0 | 0 | 0 | 1 | 0 | 0 | 1 | 0 | 0 | 1 | 0 | 0 | 0 | 0 | 1 | 0 | 0 | 1 | 0 | 0 | 0 | 1 | 0 | 0 | 0 | 1 | 0 | 0 | 1 |
| Chang 2020 | Chang | 2020 | y | sepsis and septic shoc | adults | HAT | saline | RCT | 1 | 0 | 0 | 0 | 1 | 0 | 0 | 0 | 0 | 1 | 0 | 0 | 0 | 0 | 1 | 0 | 0 | 1 | 0 | 0 | 0 | 0 | 0 | 1 | 0 | 1 | 0 | 0 | 1 |
| Chawla 1999 | Chawla | 1999 | y | septic shock | NI | hydrocortiso | placebo | RCT | 0 | 0 | 0 | 1 | 0 | 0 | 1 | 0 | 0 | 1 | 0 | 0 | 0 | 0 | 1 | 0 | 0 | 1 | 0 | 0 | 1 | 0 | 0 | 0 | 0 | 1 | 0 | 0 | 1 |
| Cicarelli 2007 | Cicarelli | 2007 | y | septic shock | adults | dexamethas | placebo | RCT | 1 | 0 | 0 | 0 | 0 | 0 | 1 | 0 | 0 | 0 | 1 | 0 | 0 | 0 | 0 | 1 | 0 | 1 | 0 | 0 | 0 | 1 | 0 | 0 | 0 | 1 | 0 | 0 | 1 |
| Cicarelli 2006 | Cicarelli | 2006 | y | SIRS | adults | dexamethas | placebo | RCT | 1 | 0 | 0 | 0 | 0 | 0 | 0 | 1 | 0 | 0 | 1 | 0 | 0 | 0 | 1 | 0 | 0 | 1 | 0 | 0 | 0 | 1 | 0 | 0 | 0 | 1 | 0 | 0 | 1 |
| Confalonieri 2005 | Confalonieri | 2005 | y | CAP (severe) | NI | hydrocortiso | placebo | RCT | 0 | 0 | 0 | 1 | 0 | 0 | 0 | 1 | 0 | 1 | 0 | 0 | 0 | 0 | 1 | 0 | 0 | 1 | 0 | 0 | 0 | 1 | 0 | 0 | 0 | 1 | 0 | 0 | 1 |
| Corral Gudino 2021 | Corral-Gudir | 2021 | y | COVID-19 (moderate to | adults | methylpredn | SoC | RCT | 1 | 0 | 0 | 0 | 0 | 0 | 0 | 1 | 0 | 0 | 0 | 1 | 0 | 0 | 1 | 0 | 0 | 1 | 0 | 0 | 0 | 1 | 0 | 0 | 0 | 1 | 0 | 0 | 1 |
| CSG 1963 | CSG | 1963 | y | severe infection | children and adults | hydrocortiso | placebo | RCT | 0 | 0 | 1 | 0 | 0 | 0 | 0 | 1 | 0 | 1 | 0 | 0 | 0 | 0 | 1 | 0 | 0 | 1 | 0 | 0 | 1 | 0 | 0 | 0 | 0 | 1 | 0 | 0 | 1 |
| de Graaf 2014 | de Graaf | 2014 | y | septic shock | children | hydrocortiso | SoC | RCT | 0 | 1 | 0 | 0 | 0 | 0 | 1 | 0 | 0 | 1 | 0 | 0 | 0 | 0 | 1 | 0 | 0 | 1 | 0 | 0 | 0 | 1 | 0 | 0 | 0 | 1 | 0 | 0 | 1 |
| Deng 2011 | Deng | 2011 | no | NI (sepsis) | | NI | NI | NI | NI | NI | NI | NI | NI | NI | NI | NI | NI | NI | NI | NI | NI | NI | NI | NI | NI | NI | NI | NI | NI | NI | NI | NI | NI | NI | NI | NI | NI |
| Dequin 2020 | Dequin | 2020 | y | COVID-19 (severe, > 80 | adults | hydrocortiso | placebo | RCT | 1 | 0 | 0 | 0 | 1 | 0 | 0 | 0 | 0 | 1 | 0 | 0 | 0 | 0 | 1 | 0 | 0 | 1 | 0 | 0 | 0 | 1 | 0 | 0 | 0 | 1 | 0 | 0 | 1 |
| Dequin 2023 | Dequin | 2023 | y | CAP (severe, SOFA mea | adults | hydrocortiso | placebo | RCT | 1 | 0 | 0 | 0 | 0 | 1 | 0 | 0 | 0 | 1 | 0 | 0 | 0 | 0 | 1 | 0 | 0 | 1 | 0 | 0 | 0 | 1 | 0 | 0 | 0 | 1 | 0 | 0 | 1 |
| Doluee 2018 | Doluee | 2018 | y | septic shock | adults | hydrocortiso | placebo | RCT | 1 | 0 | 0 | 0 | 0 | 0 | 1 | 0 | 0 | 1 | 0 | 0 | 0 | 0 | 1 | 0 | 0 | 1 | 0 | 0 | 0 | 1 | 0 | 0 | 0 | 1 | 0 | 0 | 1 |
| Edalatifard 2020 | Edalatifard | 2020 | y | COVID-19 (moderate to | adults | methylpredn | placebo | RCT | 1 | 0 | 0 | 0 | 0 | 0 | 0 | 1 | 0 | 0 | 0 | 1 | 0 | 0 | 0 | 1 | 0 | 1 | 0 | 0 | 0 | 1 | 0 | 0 | 0 | 1 | 0 | 0 | 1 |
| El Ghamrawy 2006 | El Ghamrawy | 2006 | no | NI (CAP (severe)) | | NI | NI | NI | NI | NI | NI | NI | NI | NI | NI | NI | NI | NI | NI | NI | NI | NI | NI | NI | NI | NI | NI | NI | NI | NI | NI | NI | NI | NI | NI | NI | NI |
| El Nawawy 2017 | El-Nawawy | 2017 | y | septic shock | children | early hydroc | SoC with and | RCT | 0 | 1 | 0 | 0 | 0 | 0 | 1 | 0 | 0 | 1 | 0 | 0 | 0 | 0 | 1 | 0 | 0 | 1 | 1 | 0 | 0 | 1 | 0 | 0 | 0 | 1 | 1 | 0 | 1 |
| Feng 2023 | Feng | 2023 | y | sepsis and septic shoc | adults | HAT | SoC | RCT | 1 | 0 | 0 | 0 | 1 | 0 | 0 | 0 | 0 | 1 | 0 | 0 | 0 | 0 | 1 | 0 | 0 | 1 | 0 | 0 | 0 | 0 | 0 | 1 | 0 | 1 | 0 | 0 | 1 |
| Fernandez Serrano 2 | Fernández-S | 2011 | y | CAP (no information o | adults | methylpredn | placebo | RCT | 1 | 0 | 0 | 0 | 0 | 0 | 0 | 1 | 0 | 0 | 0 | 1 | 0 | 0 | 1 | 0 | 0 | 1 | 0 | 0 | 0 | 1 | 0 | 0 | 0 | 1 | 0 | 0 | 1 |
| Fuji 2020 | Fuji | 2020 | y | septic shock | adults | HAT | hydrocortiso | RCT | 1 | 0 | 0 | 0 | 0 | 0 | 1 | 0 | 0 | 1 | 0 | 0 | 0 | 0 | 1 | 0 | 0 | 1 | 0 | 0 | 0 | 0 | 0 | 1 | 0 | 0 | 1 | 0 | 1 |
| Ghanei 2021 | Ghanei | 2021 | y | COVID-19 (moderate to | adults (>= 16 years) | prednisolon | no prednisol | RCT | 1 | 0 | 0 | 0 | 0 | 0 | 0 | 1 | 0 | 0 | 0 | 0 | 1 | 0 | 1 | 0 | 0 | 1 | 0 | 0 | 0 | 1 | 0 | 0 | 0 | 1 | 0 | 0 | 1 |
| Gordon 2014 | Gordon | 2014 | y | septic shock | adults (>= 16 years) | hydrocortiso | placebo + va | RCT | 1 | 0 | 0 | 0 | 0 | 0 | 1 | 0 | 0 | 1 | 0 | 0 | 0 | 0 | 1 | 0 | 0 | 1 | 0 | 0 | 0 | 1 | 0 | 0 | 0 | 1 | 0 | 0 | 1 |
| Gordon 2016 | Gordon | 2016 | y | septic shock | adults (>= 16 years) | hydrocortiso | placebo + va | RCT | 1 | 0 | 0 | 0 | 0 | 0 | 1 | 0 | 0 | 1 | 0 | 0 | 0 | 0 | 1 | 0 | 0 | 1 | 0 | 0 | 0 | 1 | 0 | 0 | 0 | 1 | 0 | 0 | 1 |
| Horby 2021 | Horby | 2021 | y | COVID-19 (moderate to | Initially, recruitment w | dexamethas | SoC | RCT | 0 | 0 | 1 | 0 | 0 | 0 | 0 | 1 | 0 | 0 | 1 | 0 | 0 | 0 | 1 | 0 | 0 | 1 | 0 | 0 | 0 | 1 | 0 | 0 | 0 | 1 | 0 | 0 | 1 |
| Hu 2009 | Hu | 2009 | no | septic shock | | hydrocortiso | SoC | RCT | NI | NI | NI | NI | NI | NI | NI | NI | NI | NI | NI | NI | NI | NI | NI | NI | NI | NI | NI | NI | NI | NI | NI | NI | NI | NI | NI | NI | NI |
| Huang 2014 | Huang | 2014 | no | sepsis | | corticosteroi | SoC, Chinese | RCT | NI | NI | NI | NI | NI | NI | NI | NI | NI | NI | NI | NI | NI | NI | NI | NI | NI | NI | NI | NI | NI | NI | NI | NI | NI | NI | NI | NI | NI |
| Huh 2007 | Huh | 2007 | y | septic shock | NI | 3 days hydro | 7 days hydro | RCT | 0 | 0 | 0 | 1 | 0 | 0 | 1 | 0 | 0 | 1 | 0 | 0 | 0 | 0 | 1 | 0 | 0 | 0 | 1 | 0 | 0 | 1 | 0 | 0 | 0 | 0 | 1 | 0 | 1 |
| Hussein 2021 | Hussein | 2021 | y | septic shock | adults | HAT | hydrocortiso | RCT | 1 | 0 | 0 | 0 | 0 | 0 | 1 | 0 | 0 | 1 | 0 | 0 | 0 | 0 | 1 | 0 | 0 | 1 | 0 | 0 | 0 | 0 | 0 | 1 | 0 | 0 | 1 | 0 | 1 |
| Hwang 2020 | Hwang | 2020 | y | septic shock | adults | vitamin C + t | placebo (hyc | RCT | 1 | 0 | 0 | 0 | 0 | 0 | 1 | 0 | 0 | 1 | 0 | 0 | 0 | 0 | 1 | 0 | 0 | 0 | 0 | 1 | 0 | 0 | 0 | 1 | 0 | 0 | 1 | 0 | 1 |
| Hyvernat 2016 | Hyvernat | 2016 | y | septic shock | adults | bolus hydroc | continuous h | RCT | 1 | 0 | 0 | 0 | 0 | 0 | 1 | 0 | 0 | 1 | 0 | 0 | 0 | 0 | 1 | 0 | 0 | 0 | 0 | 1 | 0 | 1 | 0 | 0 | 0 | 0 | 1 | 0 | 1 |
| Iglesias 2020 | Iglesias | 2020 | y | sepsis and septic shoc | adults | HAT | placebo | RCT | 1 | 0 | 0 | 0 | 1 | 0 | 0 | 0 | 0 | 1 | 0 | 0 | 0 | 0 | 1 | 0 | 0 | 1 | 0 | 0 | 0 | 0 | 0 | 1 | 0 | 1 | 0 | 0 | 1 |
| Jamshidi 2021 | Jamshidi | 2021 | y | septic shock | adults | HAT | placebo | RCT | 1 | 0 | 0 | 0 | 0 | 0 | 1 | 0 | 0 | 1 | 0 | 0 | 0 | 0 | 1 | 0 | 0 | 1 | 0 | 0 | 0 | 0 | 0 | 1 | 0 | 1 | 0 | 0 | 1 |
| Jiang 2019 | Jiang | 2019 | y | septic shock | NI | early hydroc | late hydroco | RCT | 0 | 0 | 0 | 1 | 0 | 0 | 1 | 0 | 0 | 1 | 0 | 0 | 0 | 0 | 1 | 0 | 0 | 0 | 1 | 0 | 0 | 1 | 0 | 0 | 0 | 1 | 1 | 0 | 0 |
| Karimpour 2019 | Karimpour | 2019 | y | septic shock | NI | vitamin C + t | saline (plus | RCT | 0 | 0 | 0 | 1 | 0 | 0 | 1 | 0 | 0 | 1 | 0 | 0 | 0 | 0 | 1 | 0 | 0 | 1 | 0 | 0 | 0 | 0 | 0 | 1 | 0 | 0 | 1 | 0 | 1 |
| Kaufmann 2008 | Kaufmann | 2008 | y | septic shock | adults | hydrocortiso | placebo | RCT | 1 | 0 | 0 | 0 | 0 | 0 | 1 | 0 | 0 | 1 | 0 | 0 | 0 | 0 | 1 | 0 | 0 | 1 | 0 | 0 | 0 | 1 | 0 | 0 | 0 | 1 | 0 | 0 | 0 |
| Keh 2003 | Keh | 2003 | y | septic shock | adults | early (hydroc | late (placebo | RCT (cross ov | 1 | 0 | 0 | 0 | 0 | 0 | 1 | 0 | 0 | 1 | 0 | 0 | 0 | 0 | 1 | 0 | 0 | 0 | 1 | 0 | 0 | 1 | 0 | 0 | 0 | 0 | 1 | 0 | 1 |
| Keh 2016 | Keh | 2016 | y | severe sepsis (not sep | adults | hydrocortiso | placebo | RCT | 1 | 0 | 0 | 0 | 0 | 1 | 0 | 0 | 0 | 1 | 0 | 0 | 0 | 0 | 1 | 0 | 0 | 1 | 0 | 0 | 0 | 1 | 0 | 0 | 0 | 1 | 0 | 0 | 1 |
| Klastersky 1971 | Klastersky | 1971 | y | severe infection | children and adults | betamethas | placebo | RCT | 0 | 0 | 1 | 0 | 0 | 0 | 0 | 1 | 0 | 0 | 0 | 0 | 1 | 0 | 1 | 0 | 0 | 1 | 0 | 0 | 1 | 0 | 0 | 0 | 0 | 1 | 0 | 0 | 1 |
| Kurungundla 2008 | Kurungundla | 2008 | no | NI (septic shock) | | NI | NI | NI | NI | NI | NI | NI | NI | NI | NI | NI | NI | NI | NI | NI | NI | NI | NI | NI | NI | NI | NI | NI | NI | NI | NI | NI | NI | NI | NI | NI | NI |
| Labib 2022 | Labib | 2022 | y | septic shock | adults | hydrocortiso | SoC | RCT | 1 | 0 | 0 | 0 | 0 | 0 | 1 | 0 | 0 | 1 | 0 | 0 | 0 | 0 | 1 | 0 | 0 | 1 | 0 | 0 | 0 | 1 | 1 | 0 | 0 | 1 | 0 | 0 | 1 |
| Li 2016 | Li | 2016 | no | CAP with septic shock | | corticosteroi | SoC | RCT | NI | NI | NI | NI | NI | NI | NI | NI | NI | NI | NI | NI | NI | NI | NI | NI | NI | NI | NI | NI | NI | NI | NI | NI | NI | NI | NI | NI | NI |
| Liu 2012 | Liu | 2012 | y (chinesisch) | ARDS | | hydrocortiso | saline | RCT | NI | NI | NI | NI | NI | NI | NI | NI | NI | NI | NI | NI | NI | NI | NI | NI | NI | NI | NI | NI | NI | NI | NI | NI | NI | NI | NI | NI | NI |
| Loisa 2007 | Loisa | 2007 | y | septic shock | adults | continuous h | bolus hydroc | RCT | 1 | 0 | 0 | 0 | 0 | 0 | 1 | 0 | 0 | 1 | 0 | 0 | 0 | 0 | 1 | 0 | 0 | 0 | 1 | 0 | 0 | 1 | 0 | 0 | 0 | 0 | 1 | 0 | 1 |
| Lucas 1984 | Lucas | 1984 | y | septic shock (extensiv | NI | dexamethas | no steroid | RCT | 0 | 0 | 0 | 1 | 0 | 0 | 1 | 0 | 0 | 0 | 1 | 0 | 0 | 0 | 0 | 1 | 0 | 1 | 0 | 0 | 1 | 0 | 0 | 0 | 0 | 1 | 0 | 0 | 1 |
| Luce 1988 | Luce | 1988 | y | septic shock | adults | methylpredn | placebo | RCT | 1 | 0 | 0 | 0 | 0 | 0 | 1 | 0 | 0 | 0 | 0 | 1 | 0 | 0 | 0 | 1 | 0 | 1 | 0 | 0 | 1 | 0 | 0 | 0 | 0 | 1 | 0 | 0 | 1 |
| Lv 2017 | Lv | 2017 | y | septic shock | adults | early hydroc | SoC with hyd | RCT | 1 | 0 | 0 | 0 | 0 | 0 | 1 | 0 | 0 | 1 | 0 | 0 | 0 | 0 | 1 | 0 | 0 | 0 | 1 | 0 | 0 | 1 | 0 | 0 | 0 | 0 | 1 | 0 | 1 |
| Lyu 2022 | Lyu | 2022 | y | septic shock | adults | HAT | placebo | RCT | 1 | 0 | 0 | 0 | 0 | 0 | 1 | 0 | 0 | 1 | 0 | 0 | 0 | 0 | 1 | 0 | 0 | 1 | 0 | 0 | 0 | 0 | 0 | 1 | 0 | 1 | 0 | 0 | 1 |
| Mansour 2012 | Mansour | 2012 | y | septic shock | children | dexamethas | placebo | RCT | 0 | 1 | 0 | 0 | 0 | 0 | 1 | 0 | 0 | 0 | 1 | 0 | 0 | 0 | 1 | 0 | 0 | 1 | 0 | 0 | 0 | 1 | 0 | 0 | 0 | 1 | 0 | 0 | 1 |
| McHardy 1972 | McHardy | 1972 | y | CAP (all severity classe | There were no children | prednisolon | no prednisol | RCT | 0 | 0 | 1 | 0 | 0 | 0 | 0 | 1 | 0 | 0 | 0 | 0 | 1 | 0 | 1 | 0 | 0 | 1 | 0 | 0 | 0 | 1 | 0 | 0 | 0 | 1 | 0 | 0 | 1 |
| Meduri 2007 | Meduri | 2007 | y | ARDS, 66% with sepsis | adults | methylpredn | placebo | RCT | 1 | 0 | 0 | 0 | 0 | 0 | 0 | 1 | 0 | 0 | 0 | 1 | 0 | 0 | 1 | 0 | 0 | 1 | 0 | 0 | 0 | 1 | 0 | 0 | 0 | 1 | 0 | 0 | 1 |
| Meduri 2009 | Meduri | 2009 | y | severe sepsis (with an | NI | hydrocortiso | placebo | RCT | 0 | 0 | 0 | 1 | 1 | 0 | 0 | 0 | 0 | 1 | 0 | 0 | 0 | 0 | 1 | 0 | 0 | 1 | 0 | 0 | 0 | 1 | 0 | 0 | 0 | 1 | 0 | 0 | 1 |
| Meduri 2022 | Meduri | 2022 | y | CAP (severe, SOFA mea | adults | methylpredn | placebo | RCT | 1 | 0 | 0 | 0 | 1 | 0 | 0 | 0 | 0 | 0 | 0 | 1 | 0 | 0 | 1 | 0 | 0 | 1 | 0 | 0 | 0 | 1 | 0 | 0 | 0 | 1 | 0 | 0 | 1 |
| Meijvis 2011 | Meijvis | 2011 | y | CAP (all severity classe | adults | dexamethas | placebo | RCT | 1 | 0 | 0 | 0 | 0 | 0 | 0 | 1 | 0 | 0 | 1 | 0 | 0 | 0 | 1 | 0 | 0 | 1 | 0 | 0 | 0 | 1 | 0 | 0 | 0 | 1 | 0 | 0 | 1 |
| Menon 2017 | Menon | 2017 | y | septic shock | children | hydrocortiso | placebo | RCT | 0 | 1 | 0 | 0 | 0 | 0 | 1 | 0 | 0 | 1 | 0 | 0 | 0 | 0 | 1 | 0 | 0 | 1 | 0 | 0 | 0 | 1 | 0 | 0 | 0 | 1 | 0 | 0 | 1 |
| Min 1975 | Min | 1975 | y | dengue shock | children | hydrocortiso | no treatment | RCT | 0 | 1 | 0 | 0 | 0 | 0 | 1 | 0 | 0 | 1 | 0 | 0 | 0 | 0 | 0 | 1 | 0 | 1 | 0 | 0 | 1 | 0 | 0 | 0 | 0 | 1 | 0 | 0 | 1 |
| Mirea 2014 | Mirea | 2014 | y | septic shock | NI | bolus hydroc | continuous h | RCT | 0 | 0 | 0 | 1 | 0 | 0 | 1 | 0 | 0 | 1 | 0 | 0 | 0 | 0 | 1 | 0 | 0 | 0 | 1 | 0 | 0 | 1 | 0 | 0 | 0 | 1 | 1 | 0 | 0 |
| Mohamed 2020 | Mohamed | 2020 | y | septic shock | adults | HAT | SoC (hydroc | RCT | 1 | 0 | 0 | 0 | 0 | 0 | 1 | 0 | 0 | 1 | 0 | 0 | 0 | 0 | 1 | 0 | 0 | 1 | 0 | 0 | 0 | 0 | 0 | 1 | 0 | 0 | 1 | 0 | 1 |
| Mohamed 2023 | Mohamed | 2023 | y | septic shock | adults | HAT | SoC | RCT | 1 | 0 | 0 | 0 | 0 | 0 | 1 | 0 | 0 | 1 | 0 | 0 | 0 | 0 | 1 | 0 | 0 | 1 | 0 | 0 | 0 | 0 | 0 | 1 | 0 | 1 | 0 | 0 | 1 |
| Moreno 2011 | Moreno | 2011 | y | septic shock | adults | hydrocortiso | placebo | RCT | 1 | 0 | 0 | 0 | 0 | 0 | 1 | 0 | 0 | 1 | 0 | 0 | 0 | 0 | 1 | 0 | 0 | 1 | 0 | 0 | 0 | 1 | 0 | 0 | 0 | 1 | 0 | 0 | 1 |
| Moskowitz 2020 | Moskowitz | 2020 | y | septic shock | adults | HAT | placebo | RCT | 1 | 0 | 0 | 0 | 0 | 0 | 1 | 0 | 0 | 1 | 0 | 0 | 0 | 0 | 1 | 0 | 0 | 1 | 0 | 0 | 0 | 0 | 0 | 1 | 0 | 1 | 0 | 0 | 1 |
| Mussack 2005 | Mussack | 2005 | y | septic shock | NI | hydrocortiso | placebo | RCT | 0 | 0 | 0 | 1 | 0 | 0 | 1 | 0 | 0 | 1 | 0 | 0 | 0 | 0 | 1 | 0 | 0 | 1 | 0 | 0 | 0 | 1 | 0 | 0 | 0 | 1 | 0 | 0 | 1 |
| Nafae 2013 | Nafae | 2013 | y | CAP (mild, moderate, a | adults | hydrocortiso | placebo | RCT | 1 | 0 | 0 | 0 | 0 | 0 | 0 | 1 | 0 | 1 | 0 | 0 | 0 | 0 | 1 | 0 | 0 | 1 | 0 | 0 | 0 | 1 | 0 | 0 | 0 | 1 | 0 | 0 | 1 |
| Nagy 2013 | Nagy | 2013 | y | CAP (severe) | children | methylpredn | placebo | RCT | 0 | 1 | 0 | 0 | 0 | 0 | 0 | 1 | 0 | 0 | 0 | 1 | 0 | 0 | 1 | 0 | 0 | 1 | 0 | 0 | 0 | 1 | 0 | 0 | 0 | 1 | 0 | 0 | 0 |
| Ngaosuwan 2018 | Ngaosuwan | 2018 | y | septic shock | adults | low-dose hy | high-dose hy | RCT | 1 | 0 | 0 | 0 | 0 | 0 | 1 | 0 | 0 | 1 | 0 | 0 | 0 | 0 | 1 | 0 | 0 | 1 | 0 | 0 | 0 | 1 | 0 | 0 | 0 | 0 | 1 | 0 | 1 |
| Oppert 2005 | Oppert | 2005 | y | septic shock | adults | hydrocortiso | placebo | RCT | 1 | 0 | 0 | 0 | 0 | 0 | 1 | 0 | 0 | 1 | 0 | 0 | 0 | 0 | 1 | 0 | 0 | 1 | 0 | 0 | 0 | 1 | 0 | 0 | 0 | 1 | 0 | 0 | 1 |
| Pierre de Oliveira 20 | Pierre de Oliv | 2023 | y | COVID-19 with ARDS (S | adults | methylpredn | placebo | RCT | 1 | 0 | 0 | 0 | 1 | 0 | 0 | 0 | 0 | 0 | 0 | 1 | 0 | 0 | 1 | 0 | 0 | 1 | 0 | 0 | 0 | 1 | 0 | 0 | 0 | 1 | 0 | 0 | 1 |
| Pongpanich 1973 | Pongpanich | 1973 | y | dengue hemorrhagic fe | children | hydrocortiso | no steroid | RCT | 0 | 1 | 0 | 0 | 0 | 0 | 1 | 0 | 0 | 1 | 0 | 0 | 0 | 0 | 0 | 1 | 0 | 1 | 0 | 0 | 1 | 0 | 0 | 0 | 0 | 1 | 0 | 0 | 1 |
| Povoa 2015 | Póvoa | 2015 | y | septic shock | NI | corticosteroi | placebo (+/- | RCT | 0 | 0 | 0 | 1 | 0 | 0 | 1 | 0 | 1 | 0 | 0 | 0 | 0 | 1 | 0 | 0 | 1 | 0 | 0 | 0 | 0 | 1 | 0 | 0 | 0 | 1 | 0 | 0 | 1 |
| Raghu 2021 | Raghu | 2021 | y | sepsis and septic shoc | adults | HAT | SoC | RCT (study de | 1 | 0 | 0 | 0 | 1 | 0 | 0 | 0 | 0 | 1 | 0 | 0 | 0 | 1 | 0 | 0 | 1 | 0 | 0 | 0 | 0 | 0 | 0 | 1 | 0 | 0 | 0 | 1 | 1 |
| Ram 2022 | Ram | 2022 | y | septic shock | NI | bolus hydroc | continuous h | RCT | 0 | 0 | 0 | 1 | 0 | 0 | 1 | 0 | 0 | 1 | 0 | 0 | 0 | 0 | 1 | 0 | 0 | 0 | 1 | 0 | 0 | 1 | 0 | 0 | 0 | 0 | 1 | 0 | 1 |
| Reddy 2020 | Reddy | 2020 | y | septic shock | adults | HAT | HA, hydrocor | RCT | 1 | 0 | 0 | 0 | 0 | 0 | 1 | 0 | 0 | 1 | 0 | 0 | 0 | 0 | 1 | 0 | 0 | 1 | 0 | 0 | 0 | 0 | 0 | 1 | 0 | 0 | 1 | 0 | 1 |
| Rezk 2013 | Rezk | 2013 | y | ARDS (early) | NI | methylpredn | no methylpre | RCT | 0 | 0 | 0 | 1 | 0 | 0 | 0 | 1 | 0 | 0 | 0 | 1 | 0 | 0 | 1 | 0 | 0 | 1 | 0 | 0 | 0 | 1 | 0 | 0 | 0 | 1 | 0 | 0 | 1 |
| Rinaldi 2006 | Rinaldi | 2006 | y | severe sepsis | adults | hydrocortiso | SoC | RCT | 1 | 0 | 0 | 0 | 0 | 1 | 0 | 0 | 0 | 1 | 0 | 0 | 0 | 0 | 1 | 0 | 0 | 1 | 0 | 0 | 0 | 1 | 0 | 0 | 0 | 1 | 0 | 0 | 1 |
| Sabry 2011 | Sabry | 2011 | y | CAP (SOFA mean 8) | adults | hydrocortiso | placebo | RCT | 1 | 0 | 0 | 0 | 1 | 0 | 0 | 0 | 0 | 1 | 0 | 0 | 0 | 0 | 1 | 0 | 0 | 1 | 0 | 0 | 0 | 1 | 0 | 0 | 0 | 1 | 0 | 0 | 1 |
| Sai 2021 | Sai | 2021 | y | severe sepsis and sept | NI | hydrocortiso | placebo | RCT | 0 | 0 | 0 | 1 | 1 | 0 | 0 | 0 | 0 | 1 | 0 | 0 | 0 | 0 | 1 | 0 | 0 | 1 | 0 | 0 | 0 | 0 | 1 | 0 | 0 | 1 | 0 | 0 | 1 |
| Sang 2020 | Sang | 2020 | no | NI (septic shock) | | NI | NI | NI | NI | NI | NI | NI | NI | NI | NI | NI | NI | NI | NI | NI | NI | NI | NI | NI | NI | NI | NI | NI | NI | NI | NI | NI | NI | NI | NI | NI | NI |
| Schelling 2001 | Schelling | 2001 | y | septic shock | NI | hydrocortiso | placebo | RCT | 0 | 0 | 0 | 1 | 0 | 0 | 1 | 0 | 0 | 1 | 0 | 0 | 0 | 0 | 1 | 0 | 0 | 1 | 0 | 0 | 0 | 1 | 0 | 0 | 0 | 1 | 0 | 0 | 0 |
| Schumer 1976 | Schumer | 1976 | y | septic shock | adults | dexamethas | saline | RCT | 1 | 0 | 0 | 0 | 0 | 0 | 1 | 0 | 0 | 0 | 1 | 1 | 0 | 0 | 0 | 1 | 0 | 1 | 0 | 0 | 0 | 1 | 0 | 0 | 0 | 1 | 1 | 0 | 1 |
| Sevransky 2021 | Sevransky | 2021 | y | sepsis | adults | HAT | placebo | RCT | 1 | 0 | 0 | 0 | 1 | 0 | 0 | 0 | 0 | 1 | 0 | 0 | 0 | 0 | 1 | 0 | 0 | 1 | 0 | 0 | 0 | 0 | 0 | 1 | 0 | 1 | 0 | 0 | 1 |
| Shi 2019 | Shi | 2019 | no (could not be | NI (septic shock) | | NI | NI | NI | NI | NI | NI | NI | NI | NI | NI | NI | NI | NI | NI | NI | NI | NI | NI | NI | NI | NI | NI | NI | NI | NI | NI | NI | NI | NI | NI | NI | NI |
| Slusher 1996 | Slusher | 1996 | y | sepsis and septic shoc | children | dexamethas | placebo | RCT | 0 | 1 | 0 | 0 | 1 | 0 | 0 | 0 | 0 | 0 | 1 | 0 | 0 | 0 | 1 | 0 | 0 | 1 | 0 | 0 | 1 | 0 | 0 | 0 | 0 | 1 | 0 | 0 | 1 |
| Snijders 2010 | Snijders | 2010 | y | CAP (all severity classe | adults | prednisolon | placebo | RCT | 1 | 0 | 0 | 0 | 0 | 0 | 0 | 1 | 0 | 0 | 0 | 0 | 1 | 0 | 1 | 0 | 0 | 1 | 0 | 0 | 0 | 1 | 0 | 0 | 0 | 1 | 0 | 0 | 1 |
| Sprung 1984 | Sprung | 1984 | y | septic shock | NI | methylpredn | no steroid | RCT | 0 | 0 | 0 | 1 | 0 | 0 | 1 | 0 | 0 | 0 | 1 | 1 | 0 | 0 | 0 | 1 | 0 | 1 | 0 | 0 | 1 | 0 | 0 | 0 | 0 | 1 | 1 | 0 | 1 |
| Sprung 2008 | Sprung | 2008 | y | septic shock | adults | hydrocortiso | placebo | RCT | 1 | 0 | 0 | 0 | 0 | 0 | 1 | 0 | 0 | 1 | 0 | 0 | 0 | 0 | 1 | 0 | 0 | 1 | 0 | 0 | 0 | 1 | 0 | 0 | 0 | 1 | 0 | 0 | 1 |
| Sui 2013 | Sui | 2013 | no | NI (CAP) | | NI | NI | NI | NI | NI | NI | NI | NI | NI | NI | NI | NI | NI | NI | NI | NI | NI | NI | NI | NI | NI | NI | NI | NI | NI | NI | NI | NI | NI | NI | NI | NI |
| Sumarmo 1982 | Sumarmo | 1982 | y | dengue shock | children | hydrocortiso | placebo | RCT | 0 | 1 | 0 | 0 | 0 | 0 | 1 | 0 | 0 | 1 | 0 | 0 | 0 | 0 | 1 | 0 | 0 | 1 | 0 | 0 | 0 | 1 | 0 | 0 | 0 | 1 | 0 | 0 | 1 |
| Tagaro 2017 | Tagaro | 2017 | y | CAP (with pleural effus | children | dexamethas | placebo | RCT | 0 | 1 | 0 | 0 | 0 | 0 | 0 | 1 | 0 | 0 | 1 | 0 | 0 | 0 | 1 | 0 | 0 | 1 | 0 | 0 | 0 | 1 | 0 | 0 | 0 | 1 | 0 | 0 | 1 |
| Tandan 2005 | Tandan | 2005 | no | NI (septic shock) | | NI | NI | NI (RCT) | NI | NI | NI | NI | NI | NI | NI | NI | NI | NI | NI | NI | NI | NI | NI | NI | NI | NI | NI | NI | NI | NI | NI | NI | NI | NI | NI | NI | NI |
| Tassniyom 1993 | Tassniyom | 1993 | y | dengue shock | children | methylpredn | placebo | RCT | 0 | 1 | 0 | 0 | 0 | 0 | 1 | 0 | 0 | 0 | 0 | 1 | 0 | 0 | 0 | 1 | 0 | 1 | 0 | 0 | 1 | 0 | 0 | 0 | 0 | 1 | 0 | 0 | 1 |
| Tilouche 2019 | Tilouche | 2019 | y | septic shock | adults | continuous h | bolus hydroc | RCT | 1 | 0 | 0 | 0 | 0 | 0 | 1 | 0 | 0 | 1 | 0 | 0 | 0 | 0 | 1 | 0 | 0 | 0 | 1 | 0 | 0 | 1 | 0 | 0 | 0 | 0 | 1 | 0 | 1 |
| Tomazini 2020 | Tomazini | 2020 | y | COVID-19 with ARDS (S | adults | dexamethas | SoC | RCT | 1 | 0 | 0 | 0 | 1 | 0 | 0 | 0 | 0 | 0 | 1 | 0 | 0 | 0 | 0 | 1 | 0 | 1 | 0 | 0 | 0 | 1 | 0 | 0 | 0 | 1 | 0 | 0 | 1 |
| Tongyoo 2016 | Tongyoo | 2016 | y | ARDS (sepsis-associat | adults | hydrocortiso | placebo | RCT | 1 | 0 | 0 | 0 | 1 | 0 | 0 | 0 | 0 | 1 | 0 | 0 | 0 | 0 | 1 | 0 | 0 | 1 | 0 | 0 | 0 | 1 | 0 | 0 | 0 | 1 | 0 | 0 | 1 |
| Torres 2015 | Torres | 2015 | y | CAP (severe) | adults | methylpredn | placebo | RCT | 1 | 0 | 0 | 0 | 0 | 0 | 0 | 1 | 0 | 0 | 0 | 1 | 0 | 0 | 1 | 0 | 0 | 1 | 0 | 0 | 0 | 1 | 0 | 0 | 0 | 1 | 0 | 0 | 1 |
| Valoor 2009 | Valoor | 2009 | y | septic shock | children | hydrocortiso | saline | RCT | 0 | 1 | 0 | 0 | 0 | 0 | 1 | 0 | 0 | 1 | 0 | 0 | 0 | 0 | 1 | 0 | 0 | 1 | 0 | 0 | 0 | 1 | 0 | 0 | 0 | 1 | 0 | 0 | 1 |
| VASSCSG 1987 | VASSCSG | 1987 | y | sepsis | NI | methylpredn | placebo | RCT | 0 | 0 | 0 | 1 | 1 | 0 | 0 | 0 | 0 | 0 | 0 | 1 | 0 | 0 | 0 | 1 | 0 | 1 | 0 | 0 | 1 | 0 | 0 | 0 | 0 | 1 | 0 | 0 | 1 |
| Venkatesh 2018 | Venkatesh | 2018 | y | septic shock | adults | hydrocortiso | placebo | RCT | 1 | 0 | 0 | 0 | 0 | 0 | 1 | 0 | 0 | 1 | 0 | 0 | 0 | 0 | 1 | 0 | 0 | 1 | 0 | 0 | 0 | 1 | 0 | 0 | 0 | 1 | 0 | 0 | 1 |
| Villar 2020 | Villar | 2020 | y | ARDS (moderate to sev | adults | dexamethas | SoC | RCT | 1 | 0 | 0 | 0 | 0 | 0 | 0 | 1 | 0 | 0 | 1 | 0 | 0 | 0 | 0 | 1 | 0 | 1 | 0 | 0 | 0 | 1 | 0 | 0 | 0 | 1 | 0 | 0 | 1 |
| Wan 2014 | Wan | 2014 | no (could not be | NI (septic shock) | | NI | NI | NI | NI | NI | NI | NI | NI | NI | NI | NI | NI | NI | NI | NI | NI | NI | NI | NI | NI | NI | NI | NI | NI | NI | NI | NI | NI | NI | NI | NI | NI |
| Wang 2022 | Wang | 2022 | chinesisch | NI (sepsis) | | NI | NI | NI | NI | NI | NI | NI | NI | NI | NI | NI | NI | NI | NI | NI | NI | NI | NI | NI | NI | NI | NI | NI | NI | NI | NI | NI | NI | NI | NI | NI | NI |
| Wang 2023 | Wang | 2023 | y | septic shock | adults | HAT | hydrocortiso | NI | 1 | 0 | 0 | 0 | 0 | 0 | 1 | 0 | 0 | 1 | 0 | 0 | 0 | 0 | 1 | 0 | 0 | 1 | 0 | 0 | 0 | 0 | 0 | 1 | 0 | 0 | 1 | 0 | 1 |
| Wani 2020 | Wani | 2020 | y | sepsis and septic shoc | adults | HAT | SoC | RCT | 1 | 0 | 0 | 0 | 1 | 0 | 0 | 0 | 0 | 1 | 0 | 0 | 0 | 0 | 1 | 0 | 0 | 1 | 0 | 0 | 0 | 0 | 0 | 1 | 0 | 1 | 0 | 0 | 1 |
| Wen 2004 | Wen | 2004 | chinesisch | NI (septic shock) | | NI | NI | NI | NI | NI | NI | NI | NI | NI | NI | NI | NI | NI | NI | NI | NI | NI | NI | NI | NI | NI | NI | NI | NI | NI | NI | NI | NI | NI | NI | NI | NI |
| Wittermans 2020 | Wittermans | 2020 | y | CAP (all severity classe | adults | dexamethas | placebo | RCT | 1 | 0 | 0 | 0 | 0 | 0 | 0 | 1 | 0 | 0 | 1 | 0 | 0 | 0 | 1 | 0 | 0 | 1 | 0 | 0 | 0 | 1 | 0 | 0 | 0 | 1 | 0 | 0 | 1 |
| Yildiz 2002 | Yildiz | 2002 | y | sepsis | adults | prednisolon | placebo | RCT | 1 | 0 | 0 | 0 | 1 | 0 | 0 | 0 | 0 | 0 | 0 | 0 | 1 | 0 | 1 | 0 | 0 | 1 | 0 | 0 | 0 | 1 | 0 | 0 | 0 | 1 | 0 | 0 | 1 |
| Yildiz 2011 | Yildiz | 2011 | y | sepsis | adults | prednisolon | placebo | RCT | 1 | 0 | 0 | 0 | 1 | 0 | 0 | 0 | 0 | 0 | 0 | 0 | 1 | 0 | 1 | 0 | 0 | 1 | 0 | 0 | 0 | 1 | 0 | 0 | 0 | 1 | 0 | 0 | 1 |
| Yuan 2010 | Yuan | 2010 | chinesisch | NI (septic shock) | | NI | NI | NI | NI | NI | NI | NI | NI | NI | NI | NI | NI | NI | NI | NI | NI | NI | NI | NI | NI | NI | NI | NI | NI | NI | NI | NI | NI | NI | NI | NI | NI |
| Zeng 2014 | Zeng | 2014 | no | NI (severe sepsis) | | NI | NI | NI | NI | NI | NI | NI | NI | NI | NI | NI | NI | NI | NI | NI | NI | NI | NI | NI | NI | NI | NI | NI | NI | NI | NI | NI | NI | NI | NI | NI | NI |
| Zhou 2015 | Zhou | 2015 | chinesisch | NI (CAP (severe)) | | NI | NI | NI | NI | NI | NI | NI | NI | NI | NI | NI | NI | NI | NI | NI | NI | NI | NI | NI | NI | NI | NI | NI | NI | NI | NI | NI | NI | NI | NI | NI | NI |

#### Supplementary Table S13: Post hoc classification of adult RCTs (n = 91)

Post hoc patient population classification according to Sepsis-3 consensus definition based on the reported eligibility criteria and baseline characteristics, including infection status, organ dysfunction, vasopressor requirement, lactate concentrations, and severity scores (e.g. SOFA); each RCT was then classified as compatible, likely compatible, unclear, likely incompatible, or incompatible with the respective consensus sepsis definition

16 RCTs were classified as likely incompatible or incompatible: Blum-2015, Corral-Gudino-2021, CSG-1963, Edalatifard-2020, Ghanei-2021, Horby 2021, Jamshidi-2021, Klastersky-1971, McHardy-1972, Meduri-2007, Meijvis-2011, Nafae-2013, Rezk-2013, Snijders-2010, Villar-2020, Wittermans-2021

| STUDY IDENTIFICATION | | | | | | | INTERVENTION | | ELIGIBILITY CRITERIA – ORIGINAL POPULATION | | | | | | | | | | | |
|---|---|---|---|---|---|---|---|---|---|---|---|---|---|---|---|---|---|---|---|---|
| ial_ID | First_author | Publication_year | Journal | Country | Multicenter | Population_type | Steroid_strategy | Comparator | Population_wording_verbatim | Sepsis_term_used | Sepsis_definition_used | If Sepsis-3 not explicitly reported, complete columns O-U | Infection_required_for_inclusion | Organ_dysfunction_required_for_inclusion | Vasopressor_required | Lactate_required | Mechanical_ventilation_required | AKI_required | Other_organ_failure_required | Eligibility_population_assessment_sepsis-3 |
| Short study identifier | First author | Year of publication | Journal name | Country, region | yes;no; not reported | adults;mixed; not reported | HC;HC+FC;dexamethasone;methylprednisolone; HAT;mixed | placebo;standard care; HC; other | Original wording of inclusion criteria | sepsis;severe sepsis;septic shock;infection+SIRS;ARDS;pneumonia;critical illness;COVID-19; other | Sepsis-1;Sepsis-2;Sepsis-3;pre-Sepsis-1;non-sepsis infection; non-sepsis disease; unclear | reported; not reported | yes;no;unclear | yes;no;unclear | yes;no | yes;no | yes;no | yes;no | Free text | compatible;likely compatible;unclear;likely incompatible; incompatible |
| Aboab 2008 | Aboab | 2008 | Critical Care | France | no | adults | HC+FC | placebo | Intensive car | septic shock | Sepsis-1 | not reported | yes | unclear | no | no | no | no | septic shock | unclear |
| Agarwal 2022 | Agarwal | 2022 | Journal of th | India | no | adults | HC | standard ca | Geriatric pat | septic shock | Sepsis-3 | reported | | | | | | | | compatible |
| Angus-2020 | Angus | 2020 | JAMA | multinationa | yes | adults | HC | no hydrocort | Patients age | COVID-19 | non-sepsis i | not reported | yes | yes | no | no | no | no | ICU admissic | likely compatible |
| Annane 2002 | Annane | 2002 | JAMA | France | yes | adults | HC+FC | placebo | (1) documen | septic shock | Sepsis-1 | not reported | yes | yes | yes | yes | no | no | Documented | likely compatible |
| Annane 2018 | Annane | 2018 | New Englanc | France | yes | adults | HC+FC | placebo | ICU patients | septic shock | Sepsis-3 | reported | | | | | | | | compatible |
| Annane-2010 | Annane | 2010 | JAMA | France | yes | adults | HC+FC | HC | Adults with s | septic shock | Sepsis-1 | not reported | yes | yes | yes | no | no | no | multiple orga | likely compatible |
| Arabi 2010 | Arabi | 2010 | CMAJ | Saudi Arabia | no | adults | HC | placebo | Patients age | septic shock | unclear | not reported | unclear | unclear | no | no | no | no | septic shock | unclear |
| Balakrishnan-2018 | Balakrishnai | 2018 | Indian Journ | India | no | adults | HAT | placebo | Patients diag | septic shock | unclear | not reported | yes | yes | yes | no | no | no | septic shock | likely compatible |
| Birudaraju-2022 | Birudaraju | 2022 | Journal of In | USA | no | adults | methylpredn | placebo | Entry criteria | severe sepsi | unclear | not reported | unclear | unclear | no | no | no | no | severe sepsi | unclear |
| Blum-2015 | Blum | 2015 | The Lancet | Switzerland | yes | adults | prednisolon | placebo | Patients age | pneumonia | non-sepsis i | not reported | yes | no | no | no | no | no | not reported | likely incompatible |
| Bollaert 1998 | Bollaert | 1998 | Critical Care | France | yes | adults | HC | placebo | Eligible patie | septic shock | Sepsis-1 | not reported | yes | yes | yes | no | yes | no | mechanical | likely compatible |
| Bone 1987 | Bone | 1987 | New Englanc | USA | yes | adults | methylpredn | placebo | Inclusion (al | sepsis; septi | pre-Sepsis-1 | not reported | yes | yes | no | yes | no | no | altered ment | likely compatible |
| Briegel 1999 | Briegel | 1999 | Critical Care | Germany | no | adults | HC | placebo | Patients suff | septic shock | Sepsis-1 | not reported | yes | yes | yes | no | no | no | organ dysfun | likely compatible |
| Briegel-2001 | Briegel | 2001 | Journal of th | Germany | no | adults | HC | placebo | Patients with | septic shock | Sepsis-1 | not reported | yes | yes | yes | no | no | no | evidence of o | likely compatible |
| Chang 2020 | Chang | 2020 | CHEST | China | no | adults | HAT | placebo | meeting the | sepsis; septi | Sepsis-3 | reported | | | | | | | | compatible |
| Chawla 1999 | Chawla | 1999 | Critical Care | USA | not reported | not reported | HC | placebo | All patients v | septic shock | unclear | not reported | unclear | unclear | yes | no | no | no | not reported | unclear |
| Cicarelli 2007 | Cicarelli | 2007 | Sao Paulo M | Brazil | no | adults | dexamethas | placebo | Patients with | septic shock | unclear | not reported | unclear | unclear | no | no | no | no | not reported | unclear |
| Cicarelli-2006 | Cicarelli | 2006 | Sao Paulo M | Brazil | no | adults | dexamethas | placebo | Patients with | infection+SIF | mixed | not reported | no | no | no | no | no | no | SIRS with or | incompatible |
| Confalonieri 2005 | Confalonieri | 2005 | American Jo | Italy | yes | not reported | HC | placebo | patients with | pneumonia | non-sepsis i | not reported | yes | yes | no | no | no | no | not for all pts | likely compatible |
| Corral-Gudino-2021 | Corral-Gudir | 2021 | Wien Klinisc | Spain | yes | adults | methylpredn | standard ca | Eligible patie | COVID-19 | non-sepsis i | not reported | yes | no | no | no | no | no | abnormal ga | likely incompatible |
| CSG-1963 | Cooperative | 1963 | JAMA | USA | yes | mixed | HC | placebo | According to | other | pre-Sepsis-1 | not reported | yes | no | no | no | no | no | life-threateni | likely incompatible |
| Dequin-2020 | Dequin | 2020 | JAMA | France | yes | adults | HC | placebo | Patients age | COVID-19 | non-sepsis i | not reported | yes | unclear | no | no | no | no | acute respira | unclear |
| Dequin-2023 | Dequin | 2023 | New Englanc | France | yes | adults | HC | placebo | Adult patient | pneumonia | non-sepsis i | not reported | yes | unclear | no | no | no | no | Severe comm | unclear |
| Doluee 2018 | Doluee | 2018 | Journal of En | Iran | no | adults | HC | placebo | Patients >18 | septic shock | unclear | not reported | unclear | unclear | yes | no | no | no | not reported | unclear |
| Edalatifard-2020 | Edalatifard | 2020 | European Re | Iran | yes | adults | methylpredn | standard ca | Inclusion cri | COVID-19 | non-sepsis i | not reported | yes | no | no | no | no | no | severe COVID | likely incompatible |
| Feng-2023 | Feng | 2023 | Open Medici | China | no | adults | HAT | standard ca | Consistent w | sepsis | unclear | not reported | yes | yes | no | no | no | no | definite or su | likely compatible |
| Fernández-Serrano-2 | Fernández-S | 2011 | Critical Care | Spain | no | adults | methylpredn | placebo | Patients adn | pneumonia | non-sepsis i | not reported | yes | yes | no | no | no | no | Respiratory f | likely compatible |
| Fujii-2020 | Fujii | 2020 | JAMA | Australia/Ne | yes | adults | HAT | HC | Patients adn | septic shock | Sepsis-3 | reported | | | | | | | | compatible |
| Ghanei-2021 | Ghanei | 2021 | Respiratory I | Iran | yes | adults | HC | other | Hospitalized | COVID-19 | non-sepsis i | not reported | yes | no | no | no | no | no | SpO2 ≤94% r | likely incompatible |
| Gordon 2014 | Gordon | 2014 | Critical Care | United Kingd | yes | adults | HC | placebo | Adult patient | sepsis | Sepsis-1 | not reported | yes | unclear | yes | no | no | no | not reported | unclear |
| Gordon 2016 | Gordon | 2016 | JAMA | United Kingd | yes | adults | HC | placebo | Adult patient | sepsis | Sepsis-1 | not reported | yes | unclear | yes | no | no | no | not reported | unclear |
| Horby 2021 | Horby | 2021 | New Englanc | United Kingd | yes | adults | dexamethas | standard ca | hospitalized | COVID-19 | non-sepsis i | not reported | yes | no | no | no | no | no | not reported | likely incompatible |
| Huh-2007 | Huh | 2007 | Critical Care | South Korea | not reported | adults | HC | HC | Patients met | septic shock | unclear | not reported | unclear | unclear | no | no | no | no | septic shock | unclear |
| Hussein-2021 | Hussein | 2021 | Internationa | Egypt | no | adults | HAT | HC | The study su | septic shock | Sepsis-3 | reported | | | | | | | | compatible |
| Hwang-2020 | Hwang | 2020 | Intensive Ca | South Korea | yes | adults | HC+FC | placebo | Adult patient | septic shock | Sepsis-3 | reported | | | | | | | | compatible |
| Hyvernat-2016 | Hyvernat | 2016 | Shock | France | yes | adults | HC | HC | Patients over | septic shock | Sepsis-2 | not reported | yes | yes | yes | no | no | yes | persisting hy | likely compatible |
| Iglesias 2020 | Iglesias | 2020 | CHEST | USA | yes | adults | HAT | placebo | adults with a | sepsis; septi | Sepsis-3 | reported | | | | | | | | compatible |
| Jamshidi-2021 | Jamshidi | 2021 | Journal of Re | Iran | no | adults | HAT | placebo | consecutive | septic shock | unclear | not reported | unclear | unclear | no | no | yes | no | septic shock | unclear |
| Jiang-2019 | Jiang | 2019 | Int J Clin Exp | China | no | adults | HC | standard ca | Sixty patients | septic shock | unclear | not reported | unclear | unclear | no | no | no | no | septic shock | unclear |
| Karimpour-2019 | Karimpour | 2019 | Journal of Pl | Iran | no | adults | HAT | placebo | Quick sofa w | septic shock | unclear | not reported | yes | yes | no | no | no | no | Eligibility reli | likely compatible |
| Kaufmann-2008 | Kaufmann | 2008 | Intensive Ca | Germany | no | adults | HC | placebo | Patients adn | septic shock | Sepsis-1 | not reported | yes | yes | no | no | no | no | septic shock | likely compatible |
| Keh 2016 | Keh | 2016 | JAMA | Germany | yes | adults | HC | placebo | Patients wer | severe sepsi | Sepsis-2 | not reported | yes | yes | no | no | no | no | SIRS plus org | likely compatible |
| Keh-2003 | Keh | 2003 | American Jo | Germany | no | adults | HC | placebo | Patients who | septic shock | Sepsis-1 | not reported | yes | yes | yes | no | no | no | septic shock | likely compatible |
| Klastersky-1971 | Klastersky | 1971 | New Englanc | Belgium | no | adults | betamethas | placebo | Only patients | other | pre-Sepsis-1 | not reported | yes | no | no | no | no | no | life-threateni | likely incompatible |
| Labib-2022 | Labib | 2022 | Ain-Shams J | Egypt | no | adults | HC+FC | standard ca | Patients incl | septic shock | unclear | not reported | unclear | unclear | no | no | no | no | eligibility crit | unclear |
| Loisa-2007 | Loisa | 2007 | Critical Care | Finland | yes | adults | HC | HC | Patients wer | septic shock | Sepsis-1 | not reported | yes | yes | yes | no | no | no | documented | likely compatible |
| Lucas-1984 | Lucas | 1984 | Archives of S | USA | no | adults | dexamethas | placebo | From August | septic shock | pre-Sepsis-1 | not reported | unclear | unclear | no | no | no | no | severe sepsi | unclear |
| Luce 1988 | Luce | 1988 | American Re | USA | no | adults | methylpredn | placebo | patients with | septic shock | pre-Sepsis-1 | not reported | yes | unclear | no | no | no | no | systolic bloo | unclear |
| Lv-2017 | Lv | 2017 | American Jo | China | no | adults | HC | placebo | Age 18 years | septic shock | Sepsis-2 | not reported | yes | unclear | no | no | no | no | not reported | unclear |
| Lyu-2022 | Lyu | 2022 | Critical Care | China | no | adults | HAT | placebo | age 18 years | septic shock | Sepsis-3 | reported | | | | | | | | compatible |
| McHardy-1972 | McHardy | 1972 | British Medic | United Kingd | no | mixed | prednisolon | other | Patients adn | pneumonia | non-sepsis i | not reported | yes | no | no | no | no | no | not reported | likely incompatible |
| Meduri 2009 | Meduri | 2009 | CHEST | USA | not reported | not reported | HC | placebo | patients with | severe sepsi | unclear | not reported | unclear | yes | no | no | no | no | stratified by | unclear |
| Meduri-2007 | Meduri | 2007 | Chest | United States | yes | adults | methylpredn | placebo | Adult intuba | ARDS | non-sepsis c | not reported | no | yes | no | no | yes | no | ARDS (Ameri | likely incompatible |
| Meduri-2022 | Meduri | 2022 | Intensive Ca | USA | yes | adults | methylpredn | placebo | Adult patient | pneumonia | non-sepsis i | not reported | yes | unclear | no | no | no | no | severe CAP/H | unclear |
| Meijvis-2011 | Meijvis | 2011 | The Lancet | Netherlands | yes | adults | dexamethas | placebo | Adults aged | pneumonia | non-sepsis i | not reported | yes | no | no | no | no | no | not reported | likely incompatible |
| Mirea-2014 | Mirea | 2014 | Critical Care | Romania | not reported | not reported | HC | standard ca | Patients with | septic shock | unclear | not reported | unclear | unclear | no | no | no | no | not reported | unclear |
| Mohamed-2020 | Mohamed | 2020 | Indian Journ | India | no | adults | HAT | standard ca | Adult non-pr | septic shock | Sepsis-3 | reported | | | | | | | | compatible |
| Mohamed-2023 | Mohamed | 2023 | Shock | Qatar | yes | adults | HAT | standard ca | Adult patient | septic shock | Sepsis-3 | reported | | | | | | | | compatible |
| Moreno-2011 | Moreno | 2011 | Intensive Ca | Multinationa | yes | adults | HC | placebo | All patients 1 | septic shock | unclear | not reported | yes | yes | no | no | no | yes | hypoperfusic | likely compatible |
| Moskowitz-2020 | Moskowitz | 2020 | JAMA | United States | yes | adults | HAT | placebo | Adult patient | septic shock | unclear | not reported | yes | unclear | yes | no | no | no | not reported | unclear |
| Mussack-2005 | Mussack | 2005 | Clinical Cher | Germany | yes | adults | HC | placebo | Patients suff | septic shock | Sepsis-1 | not reported | yes | yes | no | no | no | no | evidence of o | likely compatible |
| Nafae-2013 | Nafae | 2013 | Egyptian Jou | Egypt | no | adults | HC | placebo | Patients with | pneumonia | non-sepsis i | not reported | yes | no | no | no | no | no | not reported | likely incompatible |
| Ngaosuwan-2018 | Ngaosuwan | 2018 | Shock | Thailand | yes | adults | HC | HC | Patients with | septic shock | Sepsis-2 | not reported | yes | yes | yes | no | no | no | MAP <65 mm | likely compatible |
| Oppert-2005 | Oppert | 2005 | Critical Care | Germany | no | adults | HC | placebo | Adult patient | septic shock | unclear | not reported | yes | yes | yes | no | no | no | Cardiac inde | likely compatible |
| PierreDeOliveira-202 | Pierre de Oli | 2023 | Journal of Cc | Brazil | no | adults | methylpredn | placebo | Patients over | COVID-19 | non-sepsis i | not reported | yes | yes | no | no | yes | no | moderate or | likely compatible |
| Povoa-2015 | Póvoa | 2015 | Critical Care | Multinationa | yes | adults | HC | placebo | Patients diag | septic shock | unclear | not reported | unclear | unclear | yes | no | no | no | septic shock | unclear |
| Raghu-2021 | Raghu | 2021 | European Jo | India | no | adults | HAT | standard ca | Diagnosis of | sepsis; septi | unclear | not reported | unclear | unclear | no | no | no | no | sepsis or sep | unclear |
| Ram-2022 | Ram | 2022 | Anesthesia: | India | no | adults | HC | HC | All patients v | septic shock | unclear | not reported | unclear | unclear | yes | no | no | no | septic shock | unclear |
| Reddy-2020 | Reddy | 2020 | Indian Journ | India | no | adults | HAT | HC | Patients adn | septic shock | unclear | not reported | unclear | yes | yes | no | no | no | SOFA increas | unclear |
| Rezk-2013 | Rezk | 2013 | Egyptian Jou | Egypt | no | adults | methylpredn | placebo | All patients r | ARDS | non-sepsis c | not reported | no | yes | no | no | yes | no | ARDS (PaO2/ | likely incompatible |
| Rinaldi 2006 | Rinaldi | 2006 | Critical Care | Italy | no | adults | HC | standard ca | 1) A clinicall | severe sepsi | Sepsis-1 | not reported | yes | yes | no | no | no | no | cardiovascul | likely compatible |
| Sabry-2011 | Sabry | 2011 | Pharmacolo | Egypt | no | adults | HC | placebo | Patients to b | pneumonia | non-sepsis i | not reported | yes | yes | no | no | no | no | severe pneur | likely compatible |
| Sai 2021 | Sai | 2021 | Indian Journ | India | not reported | adults | HC+FC | placebo | patients with | severe sepsi | unclear | not reported | unclear | unclear | no | no | no | no | not reported | unclear |
| Schelling-2001 | Schelling | 2001 | Biological Ps | Germany | no | adults | HC | placebo | Forty patient | septic shock | Sepsis-1 | not reported | yes | yes | yes | no | no | no | Patients had | likely compatible |
| Schumer-1976 | Schumer | 1976 | Annals of Su | United States | no | adults | dexamethas | placebo | Septic shock | septic shock | pre-Sepsis-1 | not reported | yes | unclear | no | no | no | no | not reported | unclear |
| Sevransky-2021 | Sevransky | 2021 | JAMA | United States | yes | adults | HAT | placebo | Patients age | sepsis | Sepsis-3 | reported | | | | | | | | compatible |
| Snijders-2010 | Snijders | 2010 | American Jo | Netherlands | no | adults | prednisolon | placebo | Patients wer | pneumonia | non-sepsis i | not reported | yes | no | no | no | no | no | not reported | likely incompatible |
| Sprung-1984 | Sprung | 1984 | New Englanc | United States | no | adults | mixed | standard ca | Patients with | septic shock | pre-Sepsis-1 | not reported | yes | yes | no | no | no | no | Altered ment | likely compatible |
| Sprung-2008 | Sprung | 2008 | New Englanc | multinationa | yes | adults | HC | placebo | Patients who | septic shock | unclear | not reported | yes | yes | yes | no | no | no | Hypoperfusic | likely compatible |
| Tilouche-2019 | Tilouche | 2019 | Shock | Tunisia | no | adults | HC | HC | Patients with | septic shock | unclear | not reported | unclear | unclear | yes | no | no | no | persistent hy | unclear |
| Tomazini-2020 | Tomazini | 2020 | JAMA | Brazil | yes | adults | dexamethas | standard ca | Patients wer | COVID-19 | non-sepsis i | not reported | yes | yes | no | no | yes | no | moderate to | likely compatible |
| Tongyoo-2016 | Tongyoo | 2016 | Critical Care | Thailand | no | adults | HC | placebo | We prospect | severe sepsi | Sepsis-1 | not reported | yes | yes | no | no | yes | no | ARDS accord | likely compatible |
| Torres-2015 | Torres | 2015 | JAMA | Spain | yes | adults | methylpredn | placebo | Patients wer | pneumonia | non-sepsis i | not reported | yes | unclear | no | no | no | no | severe comm | unclear |
| VASSCSG 1987 | Hinshaw | 1987 | New Englanc | USA | yes | not reported | methylpredn | placebo | patients with | sepsis | pre-Sepsis-1 | not reported | unclear | unclear | no | no | no | no | clinical susp | unclear |
| Venkatesh-2018 | Venkatesh | 2018 | New Englanc | multinationa | yes | adults | HC | placebo | Eligible parti | septic shock | Sepsis-2 | not reported | yes | unclear | yes | no | yes | no | not reported | likely compatible |
| Villar-2020 | Villar | 2020 | The Lancet R | Spain | yes | adults | dexamethas | standard ca | Patients with | ARDS | non-sepsis c | not reported | no | yes | no | no | yes | no | ARDS accord | likely incompatible |
| Wang-2023 | Wang | 2023 | Clinical Hem | China | no | adults | HAT | HC | From Februa | septic shock | Sepsis-3 | reported | | | | | | | | compatible |
| Wani 2020 | Wani | 2020 | Infectious Di | India | no | adults | HAT | standard ca | patients with | sepsis; septi | Sepsis-3 | reported | | | | | | | | compatible |
| Wittermans-2021 | Wittermans | 2021 | European Re | Netherlands | yes | adults | dexamethas | placebo | Inclusion cri | pneumonia | non-sepsis i | not reported | yes | no | no | no | no | no | not reported | likely incompatible |
| Yildiz 2002 | Yildiz | 2002 | Critical Care | Turkey | no | adults | prednisolon | placebo | Patients over | sepsis; sever | Sepsis-1 | not reported | yes | unclear | no | no | no | no | sepsis define | unclear |
| Yildiz 2011 | Yildiz | 2011 | Journal of Re | Turkey | no | adults | prednisolon | placebo | Patients over | sepsis; sever | Sepsis-1 | not reported | yes | unclear | no | no | no | no | sepsis define | unclear |

| | BASELINE – INFECTION | | | | | BASELINE – SEVERITY SCORES | | | | | |
|---|---|---|---|---|---|---|---|---|---|---|---|
| ial_ID | Infection_described_at_baseline | Infection_confirmation | Infection_source | Infection_type | Positive_microbiology_reported | SOFA_reported | SOFA_mean_median | APACHE_II | SAPS_II | qSOFA_reported | If SOFA not reported, please fill in the following columns (AH-BC) |
| Short study identifier | yes;no; not reported | suspected;confirmed;mixed;not reported | pulmonary; abdominal; urinary;bloodstream;mixed;not reported | bacterial;viral;fungal;parasitic;mixed;not reported | yes;no; yes (but not for all patients); not reported | yes;no | Numeric value | Numeric value | Numeric value | yes;no | reported; not reported |
| Aboab 2008 | not reported | not reported | not reported | not reported | not reported | yes | 7 ± 2 / 9 ± 2 | NR | 43 ± 24 / 48 ± | no | reported |
| Agarwal 2022 | yes | mixed | mixed | mixed | yes (but not f | yes | 9.6±2.6 / 10.4 | NR | NR | no | reported |
| Angus-2020 | yes | mixed | pulmonary | viral | yes (but not f | no | NR | 18 (10–23) / 1 | NR | no | not reported |
| Annane 2002 | yes | mixed | mixed | mixed | yes (but not f | no | NR | NR | 57 ± 19 / 60 ± | no | not reported |
| Annane 2018 | yes | mixed | mixed | mixed | yes (but not f | yes | 11 ± 3 / 12 ± 3 | NR | 56 ± 19 / 56 ± | no | reported |
| Annane-2010 | yes | mixed | mixed | mixed | yes (but not f | yes | 10.6 (10.2–11 | NR | 58.9 (56.7–61 | no | reported |
| Arabi 2010 | yes | mixed | mixed | mixed | yes (but not f | yes | 14.6 ± 3.7 / 1 | 30.0 ± 7.4 / 2 | NR | no | reported |
| Balakrishnan-2018 | yes | not reported | not reported | not reported | yes (but not f | no | NR | 51.81 ± 10.76 | NR | no | not reported |
| Birudaraju-2022 | yes | mixed | not reported | not reported | yes (but not f | no | NR | 59±6 / 59±6 ( | NR | no | not reported |
| Blum-2015 | yes | mixed | pulmonary | not reported | yes (but not f | no | NR | NR | NR | no | not reported |
| Bollaert 1998 | yes | mixed | mixed | mixed | yes (but not f | no | NR | NR | 14 ± 3 / 14 ± 3 | no | not reported |
| Bone 1987 | yes | confirmed | mixed | mixed | yes (but not f | no | NR | NR | NR | no | not reported |
| Briegel 1999 | yes | confirmed | mixed | mixed | yes | yes | 12.8 ± 0.6 / 1 | 26 ± 1 / 27 ± 1 | 55 ± 4 / 55 ± 5 | no | reported |
| Briegel-2001 | yes | confirmed | mixed | mixed | yes | yes | 14 and 12 (m | 25 ± 1 / 27 ± 1 | 57 ± 4 / 60 ± 5 | no | reported |
| Chang 2020 | yes | mixed | mixed | not reported | yes (but not f | yes | 9.6 ± 4.5; 10. | 22.1 ± 8.4; 23 | NR | no | reported |
| Chawla 1999 | not reported | not reported | not reported | not reported | not reported | no | NR | NR | NR | no | not reported |
| Cicarelli 2007 | not reported | not reported | not reported | not reported | not reported | yes | 10 ± 2 / 9 ± 3 | 19 ± 4 / 20 ± | NR | no | reported |
| Cicarelli-2006 | yes | mixed | mixed | not reported | yes (but not f | yes | 7.1 / 6.9 | 15 ± 5 / 16 ± | NR | no | reported |
| Confalonieri 2005 | yes | mixed | pulmonary | bacterial | yes (but not f | no | NR | 18.2 ± 4.0; 17 | NR | no | not reported |
| Corral-Gudino-2021 | yes | confirmed | pulmonary | viral | yes | no | NR | NR | NR | yes | not reported |
| CSG-1963 | yes | mixed | mixed | mixed | yes (but not f | no | NR | NR | NR | no | not reported |
| Dequin-2020 | yes | mixed | pulmonary | viral | yes (but not f | yes | 6.0 (4.0–8.0) | NR | 32.5 (25.0–38 | no | reported |
| Dequin-2023 | yes | mixed | pulmonary | not reported | yes (but not f | yes | 4 (3–6) / 4 (3– | NR | 37 (30–45) / 3 | no | reported |
| Doluee 2018 | not reported | not reported | not reported | not reported | not reported | no | NR | NR | NR | no | not reported |
| Edalatifard-2020 | yes | confirmed | pulmonary | viral | no | no | NR | NR | NR | no | not reported |
| Feng-2023 | yes | mixed | mixed | mixed | yes (but not f | yes | 9.1 ± 3.2 / 8.9 | 11.9 ± 5.6 / 1 | NR | yes | reported |
| Fernández-Serrano-2 | yes | mixed | pulmonary | bacterial | yes (but not f | no | NR | NR | 7 (6–12) / 8 ( | no | not reported |
| Fujii-2020 | yes | mixed | mixed | not reported | not reported | yes | 8.6 ± 2.7 / 8.4 | 77.4 ± 29.7 / | NR | no | reported |
| Ghanei-2021 | yes | confirmed | pulmonary | viral | not reported | no | NR | NR | NR | no | not reported |
| Gordon 2014 | yes | mixed | mixed | not reported | not reported | no | NR | 19 (14,22) / 2 | NR | no | not reported |
| Gordon 2016 | yes | mixed | mixed | not reported | not reported | no | NR | 24 (19-30) | NR | no | not reported |
| Horby 2021 | yes | mixed | pulmonary | viral | yes (but not f | no | NR | NR | NR | no | not reported |
| Huh-2007 | not reported | not reported | not reported | not reported | not reported | no | NR | NR | NR | no | not reported |
| Hussein-2021 | not reported | not reported | not reported | not reported | not reported | yes | 8.72 ± 2.08 / | 19.66 ± 5.62 / | NR | no | reported |
| Hwang-2020 | yes | mixed | mixed | not reported | yes (but not f | yes | 8 (6–10) / 8 ( | 22 (14–32) / 2 | NR | no | reported |
| Hyvernat-2016 | yes | mixed | mixed | mixed | yes (but not f | yes | 10.4 ± 3.1 / 1 | NR | 57.9 ± 17.8 / | no | reported |
| Iglesias 2020 | yes | mixed | mixed | mixed | yes (but not f | yes | 8.3 ± 3; 7.9 ± | 24 ± 7.6; 24.9 | NR | no | reported |
| Jamshidi-2021 | no | not reported | not reported | not reported | not reported | yes | 4.5 ± 1.4 / 5.3 | 18.79 ± 1.54 / | NR | no | reported |
| Jiang-2019 | no | not reported | not reported | not reported | not reported | yes | 17.11 ± 2.11 / | 32.21 ± 6.41 / | NR | no | reported |
| Karimpour-2019 | yes | mixed | mixed | not reported | not reported | yes | 10.2 ± 2.5 / 9. | NR | NR | yes | reported |
| Kaufmann-2008 | yes | confirmed | mixed | mixed | yes (but not f | yes | 13.0 ± 1.9 / 1 | 26.8 ± 4.9 / 2 | NR | no | reported |
| Keh 2016 | yes | mixed | mixed | not reported | not reported | yes | 6.3 ± 2.5 | 19.0 ± 6.5 | 54.1 ± 11.8 | no | reported |
| Keh-2003 | yes | mixed | mixed | mixed | yes (but not f | yes | 9.7 (8.5–10.9 | NR | 42 (35–49) / 4 | no | reported |
| Klastersky-1971 | yes | mixed | mixed | mixed | yes (but not f | no | NR | NR | NR | no | not reported |
| Labib-2022 | no | not reported | not reported | not reported | not reported | yes | 9.14 ± 1.98 / | NR | NR | no | reported |
| Loisa-2007 | not reported | confirmed | not reported | not reported | not reported | yes | 10.0 ± 2.1 / 1 | 22.7 ± 5.6 / 2 | 51.5 ± 11.3 / | no | reported |
| Lucas-1984 | yes | mixed | mixed | not reported | yes (but not f | no | NR | NR | NR | no | not reported |
| Luce 1988 | yes | mixed | mixed | bacterial | yes (but not f | no | NR | NR | NR | no | not reported |
| Lv-2017 | yes | mixed | mixed | bacterial | yes (but not f | yes | 11.9 ± 3.3 / 9. | 25.5 ± 9.5 / 2 | NR | no | reported |
| Lyu-2022 | yes | not reported | mixed | not reported | yes (but not f | yes | 10.0 (7.0-12.0 | 20.0 (14.5-24 | NR | no | reported |
| McHardy-1972 | yes | mixed | pulmonary | mixed | yes (but not f | no | NR | NR | NR | no | not reported |
| Meduri 2009 | not reported | not reported | not reported | not reported | not reported | no | NR | NR | NR | no | not reported |
| Meduri-2007 | yes | mixed | mixed | not reported | yes (but not f | no | NR | NR | NR | no | not reported |
| Meduri-2022 | yes | mixed | pulmonary | mixed | yes (but not f | yes | 6.68 ± 3.0 / 6. | 54.3 ± 29.4 / | 59.4 ± 10.7 / | no | reported |
| Meijvis-2011 | yes | mixed | pulmonary | mixed | yes (but not f | no | NR | NR | NR | no | not reported |
| Mirea-2014 | not reported | not reported | not reported | not reported | not reported | no | NR | NR | NR | no | not reported |
| Mohamed-2020 | yes | not reported | mixed | not reported | yes (but not f | yes | 11.22 ± 2.99 / | NR | NR | no | reported |
| Mohamed-2023 | yes | not reported | mixed | not reported | yes (but not f | yes | 10 (8–12) / 10 | 25 (7) / 25 (7) | NR | no | reported |
| Moreno-2011 | yes | not reported | not reported | not reported | not reported | yes | 10.85 ± 3.2 / | NR | NR | no | reported |
| Moskowitz-2020 | yes | mixed | mixed | not reported | not reported | yes | 9.1 (3.5) / 9.2 | NR | NR | no | reported |
| Mussack-2005 | yes | confirmed | mixed | not reported | yes | no | NR | 25 (20–28) / 2 | 53 (46–60) / 4 | no | not reported |
| Nafae-2013 | yes | mixed | pulmonary | mixed | not reported | no | NR | NR | NR | no | not reported |
| Ngaosuwan-2018 | yes | confirmed | mixed | mixed | yes | no | NR | NR | 54.5 ± 16.5 / | no | not reported |
| Oppert-2005 | yes | mixed | mixed | not reported | yes (but not f | yes | 11 (10–13) / 1 | 25 (19–30) / 2 | NR | no | reported |
| PierreDeOliveira-202 | yes | confirmed | pulmonary | viral | not reported | yes | 7.24 ± 2.41 / | NR | 61.24 ± 12.31 | no | reported |
| Povoa-2015 | yes | mixed | mixed | mixed | yes (but not f | yes | 11 (9–13) / 10 | 25 (20–31) / 2 | NR | no | reported |
| Raghu-2021 | yes | mixed | mixed | mixed | yes | yes | 9.899 ± 4.289 | 22.0 ± 9.70 / | NR | no | reported |
| Ram-2022 | no | not reported | not reported | not reported | not reported | no | NR | NR | NR | no | not reported |
| Reddy-2020 | yes | not reported | mixed | not reported | not reported | yes | 8.8 ± 2.9 / 11. | 21.2 ± 6.5 / 1 | NR | no | reported |
| Rezk-2013 | yes | not reported | mixed | not reported | not reported | no | NR | NR | NR | no | not reported |
| Rinaldi 2006 | yes | confirmed | mixed | bacterial | yes | yes | 5 (11–1) / 4 ( | NR | 54 (41–58) / 5 | no | reported |
| Sabry-2011 | yes | mixed | pulmonary | bacterial | yes (but not f | yes | 8.2 ± 1.46 / 8. | NR | NR | no | reported |
| Sai 2021 | not reported | not reported | not reported | not reported | not reported | yes | 16 / 16 | NR | NR | no | reported |
| Schelling-2001 | yes | not reported | mixed | not reported | not reported | no | NR | 22 (14–35) / 2 | NR | no | not reported |
| Schumer-1976 | yes | confirmed | mixed | bacterial | yes | no | NR | NR | NR | no | not reported |
| Sevransky-2021 | yes | suspected | mixed | not reported | not reported | yes | 9 (7–12) / 9 ( | 27 (22–33) / 2 | NR | no | reported |
| Snijders-2010 | yes | mixed | pulmonary | mixed | yes (but not f | no | NR | NR | NR | no | not reported |
| Sprung-1984 | yes | mixed | not reported | not reported | yes (but not f | no | NR | NR | NR | no | not reported |
| Sprung-2008 | yes | mixed | mixed | mixed | yes | yes | 10.6 ± 3.4 / 1 | NR | 49.5 ± 17.8 / | no | reported |
| Tilouche-2019 | yes | not reported | mixed | not reported | not reported | yes | 12 (10–13) / 1 | NR | 48 (33–61) / 4 | no | reported |
| Tomazini-2020 | yes | mixed | pulmonary | viral | not reported | yes | 9 (7–10.5) / 8 | NR | 69.4 ± 12.6 / | no | reported |
| Tongyoo-2016 | yes | mixed | mixed | not reported | yes (but not f | yes | 10.9 ± 3.5 / 1 | 21.7 ± 5.7 / 2 | NR | no | reported |
| Torres-2015 | yes | mixed | pulmonary | mixed | yes (but not f | no | NR | NR | NR | no | not reported |
| VASSCSG 1987 | yes | mixed | mixed | mixed | yes | no | NR | NR | NR | no | not reported |
| Venkatesh-2018 | yes | mixed | mixed | not reported | not reported | no | NR | 24 (19–29) / 2 | NR | no | not reported |
| Villar-2020 | no | not reported | not reported | not reported | not reported | yes | 8.7 ± 3.1 / 8.6 | NR | NR | no | reported |
| Wang-2023 | yes | suspected | mixed | not reported | not reported | yes | 8.3 ± 2.9 / 8.8 | 20.4 ± 6.1 / 2 | NR | no | reported |
| Wani 2020 | yes | mixed | mixed | not reported | not reported | yes | 9.36 ± 3.66 / | 20 (15–24) / 1 | NR | no | reported |
| Wittermans-2021 | yes | mixed | pulmonary | mixed | yes (but not f | no | NR | NR | NR | no | not reported |
| Yildiz 2002 | yes | mixed | not reported | mixed | yes (but not f | no | NR | 16.6 ± 6.9 | NR | no | not reported |
| Yildiz 2011 | yes | confirmed | mixed | mixed | yes (but not f | yes | 8.9 ± 3.8 | 20.7 ± 7.3 | NR | no | reported |

| | BASELINE – RESPIRATORY SOFA DOMAIN | | | BASELINE – CARDIOVASCULAR SOFA DOMAIN | | | | | BASELINE – RENAL SOFA DOMAIN | | | BASELINE – LIVER SOFA DOMAIN | BASELINE – COAGULATION SOFA DOMAIN | BASELINE – NEUROLOGIC SOFA DOMAIN | | SOFA RECONSTRUCTION | | | SEPTIC SHOCK RECONSTRUCTION | | | | OVERALL RECLASSIFICATION |
|---|---|---|---|---|---|---|---|---|---|---|---|---|---|---|---|---|---|---|---|---|---|---|---|
| ial_ID | PaO2_FiO2 | Mechanical_ventilation_baseline | ARDS_present | MAP | Vasopressor_use_baseline | Vasopressor_type | Vasopressor_dose_reported | Lactate_baseline | Creatinine | Renal_replacement_therapy | AKI_present_baseline | Bilirubin | Platelets | GCS | Sedation_reported | SOFA_reconstructed_possible | Reconstructed_SOFA_minimum | SOFA_>=2_compatible | Vasopressor_needed | Lactate_>2 | Fluids_before_vasopressor | Septic_shock_Sepsis3_compatible | Final_population_classification (whole population) |
| Short study identifier | Numeric value | yes;no; yes (but not for all patients); not reported | yes;no; not reported | Numeric value | yes;no; yes (but not all patients); not reported | norepinephrine;dopamine;other; not reported | yes;no | Numeric value | Numeric value | yes;no; not reported | yes;no; not reported | Numeric value | Numeric value | Numeric value | yes;no | yes; partial; no | Numeric value (if reconstruction possible) | yes;likely;unclear;no | yes;no; yes (but not all patients); not reported | yes;no; yes (but not all patients); not reported | yes;no; not reported | yes;partial; unclear;no | Sepsis-3 compatible;likely compatible;unclear;likely incompatible; incompatible |
| Aboab 2008 | | | | | | | | | | | | | | | | | | | | | | | unclear |
| Agarwal 2022 | | | | | | | | | | | | | | | | | | | | | | | Sepsis-3 compatible |
| Angus-2020 | 149 ± 83 / 13 | yes (but not f | not reported | NR | yes (but not a | not reported | no | 1.2 (0.9-1.5) / | 0.9 (0.7-1.2) / | not reported | not reported | 0.1 (0.1-0.2) / | 254 (117) / 2 | 13 (4) / 13 (4) | yes | yes | ≥2 | likely | yes (but not a | no | not reported | partial | Sepsis-3 compatible |
| Annane 2002 | 171 ± 124 / 1 | yes | not reported | 55 ± 10 / 54 ± | yes | dopamine; d | yes | 4.3 ± 4.3 / 4.6 | NR | not reported | not reported | NR | 175 ± 129 / 1 | NR | no | yes | ≥2 | yes | yes | yes | yes | yes | Sepsis-3 compatible |
| Annane 2018 | | | | | | | | | | | | | | | | | | | | | | | Sepsis-3 compatible |
| Annane-2010 | | | | | | | | | | | | | | | | | | | | | | | Sepsis-3 compatible |
| Arabi 2010 | | | | | | | | | | | | | | | | | | | | | | | unclear |
| Balakrishnan-2018 | NR | not reported | not reported | <65 | yes | norepinephr | no | 5.57 ± 2.41 / | 1.52 ± 0.79 / | no | not reported | 4.8 ± 6.26 / 1. | NR | NR | yes | partial | ≥2 | likely | yes | yes | yes | yes | likely compatible |
| Birudaraju-2022 | NR | yes (but not f | not reported | NR | not reported | not reported | no | NR | 2.7 ± 0.6 / 1.9 | not reported | not reported | NR | NR | NR | no | no | | unclear | not reported | not reported | not reported | partial | unclear |
| Blum-2015 | NR | not reported | not reported | NR | no | not reported | no | NR | NR | not reported | not reported | NR | NR | NR | no | no | | unclear | not reported | not reported | not reported | unclear | likely incompatible |
| Bollaert 1998 | 179 ± 74 / 19 | yes | not reported | 83 ± 16 / 91 ± | yes | epinephrine; | yes | 2.0 ± 0.9 / 1.9 | 14 (6-61) / 17 | not reported | not reported | NR | NR | NR | no | yes | ≥2 | yes | yes | yes (but not a | yes | partial | Sepsis-3 compatible |
| Bone 1987 | NR | not reported | not reported | NR | not reported | not reported | no | NR | NR | not reported | not reported | NR | NR | NR | no | no | | unclear | not reported | not reported | not reported | unclear | unclear |
| Briegel 1999 | | | | | | | | | | | | | | | | | | | | | | | Sepsis-3 compatible |
| Briegel-2001 | | | | | | | | | | | | | | | | | | | | | | | Sepsis-3 compatible |
| Chang 2020 | | | | | | | | | | | | | | | | | | | | | | | Sepsis-3 compatible |
| Chawla 1999 | NR | not reported | not reported | NR | yes | not reported | no | NR | NR | not reported | not reported | NR | NR | NR | no | no | | unclear | yes | not reported | not reported | unclear | unclear |
| Cicarelli 2007 | | | | | | | | | | | | | | | | | | | | | | | unclear |
| Cicarelli-2006 | | | | | | | | | | | | | | | | | | | | | | | unclear |
| Confalonieri 2005 | 178 ± 58 / 14 | yes (but not f | not reported | NR | yes (but not a | not reported | no | NR | NR | not reported | not reported | NR | NR | NR | no | partial | ≥2 | likely | yes (but not a | not reported | not reported | unclear | likely compatible |
| Corral-Gudino-2021 | NR | no | not reported | NR | no | not reported | no | NR | 0.9 (0.7–1.1) | no | not reported | NR | 244 (215–359 | NR | no | partial | 1 | unclear | not reported | not reported | not reported | unclear | likely incompatible |
| CSG-1963 | NR | not reported | not reported | NR | not reported | not reported | no | NR | NR | not reported | not reported | NR | NR | NR | no | no | | unclear | no | not reported | not reported | no | incompatible |
| Dequin-2020 | | | | | | | | | | | | | | | | | | | | | | | Sepsis-3 compatible |
| Dequin-2023 | | | | | | | | | | | | | | | | | | | | | | | unclear |
| Doluee 2018 | NR | not reported | not reported | NR | not reported | not reported | no | NR | NR | not reported | not reported | NR | NR | NR | no | no | | unclear | yes | not reported | not reported | unclear | unclear |
| Edalatifard-2020 | NR | no | no | NR | no | not reported | no | NR | NR | no | not reported | NR | 219.9 ± 106.8 | NR | no | no | | unclear | not reported | not reported | not reported | unclear | incompatible |
| Feng-2023 | | | | | | | | | | | | | | | | | | | | | | | Sepsis-3 compatible |
| Fernández-Serrano-2 | 257 (209–276 | no | not reported | NR | no | not reported | no | NR | NR | not reported | not reported | NR | NR | NR | no | partial | ≥2 | likely | not reported | not reported | not reported | unclear | likely compatible |
| Fujii-2020 | | | | | | | | | | | | | | | | | | | | | | | Sepsis-3 compatible |
| Ghanei-2021 | NR | no | not reported | NR | no | not reported | no | NR | 1.41 ± 6.25 | no | not reported | 0.32 ± 0.41 | 197.56 ± 73.6 | NR | no | partial | 1 | unclear | not reported | not reported | not reported | unclear | likely incompatible |
| Gordon 2014 | 225 (146,344 | yes (but not f | not reported | 72 (63,80) / 6 | yes | norepinephr | yes | 2.1 (1.4,5.0) / | 85 (66,129) / | no | yes | 12 (10,32) / 1 | 162 (106,218 | 15 (15,15) / 1 | no | yes | ≥2 | yes | yes | yes (but not a | yes | partial | Sepsis-3 compatible |
| Gordon 2016 | 188 (121-302 | yes (but not f | not reported | 70 (62-77) | yes | norepinephr | yes | 2.3 (1.4-4.0) | 1.38 (0.84-2. | no | yes | 0.88 (0.47-1. | 188 (121-288 | 14 (4-15) | no | yes | ≥2 | yes | yes | yes (but not a | yes | partial | Sepsis-3 compatible |
| Horby 2021 | NR | yes (but not f | not reported | NR | not reported | not reported | no | NR | NR | not reported | not reported | NR | NR | NR | no | no | | unclear | not reported | not reported | not reported | unclear | likely incompatible |
| Huh-2007 | NR | not reported | not reported | NR | not reported | not reported | no | NR | NR | not reported | not reported | NR | NR | NR | no | no | | unclear | not reported | not reported | not reported | unclear | unclear |
| Hussein-2021 | | | | | | | | | | | | | | | | | | | | | | | likely compatible |
| Hwang-2020 | | | | | | | | | | | | | | | | | | | | | | | Sepsis-3 compatible |
| Hyvernat-2016 | | | | | | | | | | | | | | | | | | | | | | | Sepsis-3 compatible |
| Iglesias 2020 | | | | | | | | | | | | | | | | | | | | | | | Sepsis-3 compatible |
| Jamshidi-2021 | | | | | | | | | | | | | | | | | | | | | | | likely incompatible |
| Jiang-2019 | | | | | | | | | | | | | | | | | | | | | | | unclear |
| Karimpour-2019 | | | | | | | | | | | | | | | | | | | | | | | Sepsis-3 compatible |
| Kaufmann-2008 | | | | | | | | | | | | | | | | | | | | | | | Sepsis-3 compatible |
| Keh 2016 | | | | | | | | | | | | | | | | | | | | | | | Sepsis-3 compatible |
| Keh-2003 | | | | | | | | | | | | | | | | | | | | | | | Sepsis-3 compatible |
| Klastersky-1971 | NR | not reported | not reported | NR | not reported | not reported | no | NR | NR | not reported | not reported | NR | NR | NR | no | no | | unclear | not reported | not reported | not reported | unclear | incompatible |
| Labib-2022 | | | | | | | | | | | | | | | | | | | | | | | unclear |
| Loisa-2007 | | | | | | | | | | | | | | | | | | | | | | | Sepsis-3 compatible |
| Lucas-1984 | NR | yes | not reported | NR | unclear | not reported | no | NR | NR | not reported | not reported | NR | NR | NR | no | no | | unclear | not reported | not reported | not reported | unclear | unclear |
| Luce 1988 | NR | yes (but not f | not reported | NR | yes (but not a | dopamine | no | NR | 2.1 ± 0.2 / 1.8 | not reported | not reported | 2.1 ± 0.5 / 2.4 | NR | NR | no | partial | ≥2 | likely | yes (but not a | not reported | not reported | unclear | likely compatible |
| Lv-2017 | | | | | | | | | | | | | | | | | | | | | | | Sepsis-3 compatible |
| Lyu-2022 | | | | | | | | | | | | | | | | | | | | | | | Sepsis-3 compatible |
| McHardy-1972 | NR | no | not reported | NR | no | not reported | no | NR | NR | not reported | not reported | NR | NR | NR | no | no | | unclear | not reported | not reported | not reported | unclear | likely incompatible |
| Meduri 2009 | NR | yes (but not f | not reported | NR | not reported | not reported | no | NR | NR | not reported | not reported | NR | NR | NR | no | no | | unclear | not reported | not reported | not reported | unclear | unclear |
| Meduri-2007 | 118.4 ± 51.2 / | yes | yes | NR | yes (but not a | not reported | no | NR | NR | not reported | not reported | NR | NR | NR | no | partial | ≥2 | likely | yes (but not a | not reported | not reported | partial | likely incompatible |
| Meduri-2022 | | | | | | | | | | | | | | | | | | | | | | | likely compatible |
| Meijvis-2011 | NR | no | not reported | NR | no | not reported | no | NR | NR | not reported | not reported | NR | NR | NR | no | no | | unclear | not reported | not reported | not reported | unclear | likely incompatible |
| Mirea-2014 | NR | not reported | not reported | NR | not reported | not reported | no | NR | NR | not reported | not reported | NR | NR | NR | no | no | | unclear | not reported | not reported | not reported | unclear | unclear |
| Mohamed-2020 | | | | | | | | | | | | | | | | | | | | | | | Sepsis-3 compatible |
| Mohamed-2023 | | | | | | | | | | | | | | | | | | | | | | | Sepsis-3 compatible |
| Moreno-2011 | | | | | | | | | | | | | | | | | | | | | | | Sepsis-3 compatible |
| Moskowitz-2020 | | | | | | | | | | | | | | | | | | | | | | | Sepsis-3 compatible |
| Mussack-2005 | NR | not reported | not reported | NR | yes | not reported | no | NR | NR | no | not reported | NR | NR | NR | no | no | | unclear | not reported | not reported | not reported | unclear | unclear |
| Nafae-2013 | 343.9 ± 51.6 / | no | not reported | NR | yes (but not a | not reported | no | NR | 1.14 ± 0.5 / 1. | not reported | not reported | NR | 249.56 ± 120. | NR | no | partial | 1 | unclear | not reported | not reported | not reported | unclear | likely incompatible |
| Ngaosuwan-2018 | NR | yes (but not f | no | <65 | yes | other | no | 4.6 ± 2.9 / 5.8 | 2.16 ± 1.5 / 2. | no | not reported | NR | 162.6 ± 97.6 / | NR | no | partial | ≥2 | likely | yes | yes (but not a | yes | partial | likely compatible |
| Oppert-2005 | | | | | | | | | | | | | | | | | | | | | | | Sepsis-3 compatible |
| PierreDeOliveira-202 | | | | | | | | | | | | | | | | | | | | | | | Sepsis-3 compatible |
| Povoa-2015 | | | | | | | | | | | | | | | | | | | | | | | Sepsis-3 compatible |
| Raghu-2021 | | | | | | | | | | | | | | | | | | | | | | | Sepsis-3 compatible |
| Ram-2022 | NR | not reported | not reported | NR | yes | other | no | NR | NR | not reported | not reported | NR | NR | NR | no | no | | unclear | yes | not reported | not reported | unclear | unclear |
| Reddy-2020 | | | | | | | | | | | | | | | | | | | | | | | Sepsis-3 compatible |
| Rezk-2013 | estimated Pa | yes | yes | NR | no | not reported | no | 3.40 ± 2.11 / | 98.33 ± 54.29 | not reported | not reported | 35.12 ± 9.07 / | 178.00 ± 106. | NR | no | partial | ≥2 | likely | no | yes | not reported | no | likely incompatible |
| Rinaldi 2006 | | | | | | | | | | | | | | | | | | | | | | | likely compatible |
| Sabry-2011 | | | | | | | | | | | | | | | | | | | | | | | likely compatible |
| Sai 2021 | | | | | | | | | | | | | | | | | | | | | | | unclear |
| Schelling-2001 | NR | not reported | not reported | NR | yes | other | no | NR | NR | not reported | not reported | NR | NR | NR | yes | no | | unclear | yes | not reported | not reported | unclear | unclear |
| Schumer-1976 | NR | not reported | not reported | NR | not reported | not reported | no | NR | NR | not reported | not reported | NR | NR | NR | no | no | | unclear | not reported | not reported | not reported | unclear | unclear |
| Sevransky-2021 | | | | | | | | | | | | | | | | | | | | | | | Sepsis-3 compatible |
| Snijders-2010 | NR | no | not reported | NR | no | not reported | no | NR | NR | not reported | not reported | NR | NR | NR | no | no | | unclear | not reported | not reported | not reported | unclear | likely incompatible |
| Sprung-1984 | NR | not reported | not reported | 65 ± 4 / 64 ± 3 | yes (but not a | dopamine | yes | 6.7 ± 2.0 / 5.1 | NR | not reported | not reported | NR | NR | NR | no | partial | ≥2 | likely | yes (but not a | yes | yes | partial | likely compatible |
| Sprung-2008 | | | | | | | | | | | | | | | | | | | | | | | Sepsis-3 compatible |
| Tilouche-2019 | | | | | | | | | | | | | | | | | | | | | | | Sepsis-3 compatible |
| Tomazini-2020 | | | | | | | | | | | | | | | | | | | | | | | Sepsis-3 compatible |
| Tongyoo-2016 | | | | | | | | | | | | | | | | | | | | | | | Sepsis-3 compatible |
| Torres-2015 | 236 ± 75 / 23 | yes (but not f | not reported | NR | not reported | not reported | no | NR | 1.3 (0.9–1.8) | no | not reported | NR | 214 (176–282 | NR | no | partial | 1 to 2 | unclear | yes (but not a | not reported | not reported | partial | unclear |
| VASSCSG 1987 | NR | not reported | not reported | NR | not reported | not reported | no | 25.5 / 24.1 m | 2.2 / 2.2 | not reported | not reported | NR | NR | NR | no | partial | ≥2 | unclear | not reported | yes | not reported | unclear | unclear |
| Venkatesh-2018 | 164.6 ± 91.3 / | yes | not reported | 72.5 ± 8.2 / 7 | yes | norepinephr | yes | NR | 2.2 ± 2.0 / 2.1 | not reported | not reported | 1.7 ± 2.4 / 1.7 | NR | NR | no | yes | ≥2 | likely | yes | yes | not reported | partial | Sepsis-3 compatible |
| Villar-2020 | | | | | | | | | | | | | | | | | | | | | | | likely incompatible |
| Wang-2023 | | | | | | | | | | | | | | | | | | | | | | | Sepsis-3 compatible |
| Wani 2020 | | | | | | | | | | | | | | | | | | | | | | | Sepsis-3 compatible |
| Wittermans-2021 | NR | no | not reported | NR | no | not reported | no | NR | NR | not reported | not reported | NR | NR | NR | no | no | | unclear | not reported | not reported | not reported | unclear | likely incompatible |
| Yildiz 2002 | NR | not reported | not reported | NR | not reported | not reported | no | NR | NR | not reported | not reported | NR | 183,050 ± 13 | NR | no | no | | unclear | not reported | not reported | not reported | unclear | unclear |
| Yildiz 2011 | | | | | | | | | | | | | | | | | | | | | | | unclear |

### Supplementary Table S14: Post hoc classification of pediatric RCTs (n = 13)

Post hoc patient population classification according to Phoenix Sepsis consensus definition based on the reported eligibility criteria and baseline characteristics, including infection status, organ dysfunction, vasopressor requirement, lactate concentrations, and severity scores (e.g. Phoenix Sepsis Score (PSS)); each RCT was then classified as compatible, likely compatible, unclear, likely incompatible, or incompatible with the respective consensus sepsis definition

2 RCTs were classified as likely incompatible: Nagy-2013, Tagarro-2017

| STUDY IDENTIFICATION | | | | | | | INTERVENTION | |
|---|---|---|---|---|---|---|---|---|
| Trial_ID | First_author | Publication_year | Journal | Country | Multicenter | Population_type | Steroid_strategy | Comparator |
| Short study identifier | First author | Year of publication | Journal name | Country, region | yes;no; not reported | pediatric | HC;HC+FC;dexamethasone;methylprednisolone;HAT;mixed | placebo;standard care;other |
| Branco-2014 | Branco | 2014 | Pediatric Cri | Brazil | no | pediatric | HC | placebo |
| deGraaf-2014 | de Graaf | 2014 | Pediatric Cri | United Kingd | yes | pediatric | HC | standard ca |
| El-Nawawy-2017 | El-Nawawy | 2017 | The Pediatric | Egypt | no | pediatric | HC | standard ca |
| Mansour-2012 | Mansour | 2012 | Archives of D | Egypt | no | pediatric | dexamethas | placebo |
| Menon-2017 | Menon | 2017 | Pediatric Cri | Canada | yes | pediatric | HC | placebo |
| Min-1975 | Min | 1975 | The Southea | Myanmar (Bu | no | pediatric | HC | other |
| Nagy-2013 | Nagy | 2013 | Pediatric Pul | Hungary | no | pediatric | methylpredn | placebo |
| Pongpanich-1973 | Pongpanich | 1973 | Journal of th | Thailand | no | pediatric | HC | other |
| Slusher-1996 | Slusher | 1996 | The Pediatric | Nigeria/Keny | yes | pediatric | dexamethas | placebo |
| Sumarmo-1982 | Sumarmo | 1982 | Pediatrics | Indonesia | no | pediatric | HC | placebo |
| Tagarro-2017 | Tagarro | 2017 | The Journal ( | Spain | yes | pediatric | dexamethas | placebo |
| Tassniyom-1993 | Tassniyom | 1993 | Pediatrics | Thailand | yes | pediatric | methylpredn | placebo |
| Valoor-2009 | Valoor | 2009 | Pediatric Cri | India | no | pediatric | HC | other |

| | ELIGIBILITY CRITERIA – ORIGINAL POPULATION | | | | | | | | | | |
|---|---|---|---|---|---|---|---|---|---|---|---|
| Trial_ID | Population_wording_verbatim | Sepsis_term_used | Sepsis_definition_used | If PS-2024 not explicitly reported, complete columns O-T | Infection_required_for_inclusion | Organ_dysfunction_required_for_inclusion | Vasopressor_required | Lactate_required | Mechanical_ventilation_required | Other_organ_failure_required | Eligibility_population_assessment_PS-2024 |
| Short study identifier | Original wording of inclusion criteria | sepsis;severe sepsis;septic shock;infection+SIRS;ARDS;pneumonia;critical illness;COVID-19; other | Pediatric Sepsis Definition 2024 (PS-2024); pre-PS; non-sepsis infection; non-sepsis disease; unclear | reported; not reported | yes;no;unclear | yes;no;unclear | yes;no | yes;no | yes;no | Free text | compatible;likely compatible;unclear;likely incompatible; incompatible |
| Branco-2014 | Double blinc | septic shock | pre-PS | not reported | unclear | unclear | no | no | no | children with | unclear |
| deGraaf-2014 | Children with | septic shock | pre-PS | not reported | unclear | unclear | no | no | no | children with | unclear |
| El-Nawawy-2017 | This study w | septic shock | pre-PS | not reported | yes | yes | no | no | no | Sepsis defini | likely compatible |
| Mansour-2012 | Thirty patien | septic shock | pre-PS | not reported | unclear | unclear | no | no | no | patients with | unclear |
| Menon-2017 | Children age | septic shock | pre-PS | not reported | unclear | yes | yes | no | no | not reported | unclear |
| Min-1975 | Out of 400 ca | other | non-sepsis i | not reported | yes | unclear | no | no | no | dengue shoc | unclear |
| Nagy-2013 | These patien | pneumonia | non-sepsis i | not reported | yes | no | no | no | no | SIRS criteria ( | likely incompatible |
| Pongpanich-1973 | Only serolog | other | non-sepsis i | not reported | yes | unclear | no | no | no | serologically | unclear |
| Slusher-1996 | children with | sepsis; septi | pre-PS | not reported | yes | yes | no | no | no | hypoxemia; c | likely compatible |
| Sumarmo-1982 | All patients s | other | non-sepsis i | not reported | yes | unclear | no | no | no | dengue shoc | unclear |
| Tagarro-2017 | Eligible parti | pneumonia | non-sepsis i | not reported | yes | no | no | no | no | not reported | likely incompatible |
| Tassniyom-1993 | To be eligible | other | non-sepsis i | not reported | yes | unclear | no | no | no | dengue haen | unclear |
| Valoor-2009 | Patients 2 m | septic shock | pre-PS | not reported | yes | yes | yes | no | no | Hypotension | likely compatible |

| | BASELINE – INFECTION | | | | | BASELINE – SEVERITY SCORES | | | | | | | |
|---|---|---|---|---|---|---|---|---|---|---|---|---|---|
| Trial_ID | Infection_described_at_baseline | Infection_confirmation | Infection_source | Infection_type | Positive_microbiology_reported | Phoenix_Sepsis_Score_reported | Phoenix_Sepsis_Score (PSS) | SOFA_reported | SOFA_mean_median | PRISM | PELOD | PIM | If PSS not reported, please fill in the following columns (AI-BB) |
| Short study identifier | yes;no; not reported | suspected;confirmed;mixed;not reported | pulmonary; abdominal; urinary;bloodstream;mixed;not reported | bacterial;viral;fungal;parasitic;mixed;not reported | yes;no; yes (but not for all patients); not reported | yes;no | Numeric value | yes;no | Numeric value | Numeric value | Numeric value | Numeric value | reported; not reported |
| Branco-2014 | no | not reported | not reported | not reported | not reported | no | NR | no | NR | NR | NR | NR | not reported |
| deGraaf-2014 | no | not reported | not reported | not reported | not reported | no | NR | no | NR | NR | NR | 6,3 | not reported |
| El-Nawawy-2017 | not reported | not reported | not reported | not reported | not reported | no | NR | no | NR | NR | 38 ± 11.3 / 31 | 87.0 ± 13.2 / ' | not reported |
| Mansour-2012 | no | not reported | not reported | not reported | not reported | no | NR | no | NR | NR | NR | NR | not reported |
| Menon-2017 | not reported | not reported | not reported | not reported | not reported | no | NR | no | NR | 7 (5, 11) / 6 ( | 6 (3, 9) / 6 (4, | NR | not reported |
| Min-1975 | yes | confirmed | not reported | viral | yes | no | NR | no | NR | NR | NR | NR | not reported |
| Nagy-2013 | yes | mixed | pulmonary | bacterial | yes (but not f | no | NR | no | NR | NR | NR | NR | not reported |
| Pongpanich-1973 | yes | confirmed | not reported | viral | yes | no | NR | no | NR | NR | NR | NR | not reported |
| Slusher-1996 | yes | confirmed | mixed | bacterial | yes (but not f | no | NR | no | NR | NR | NR | NR | not reported |
| Sumarmo-1982 | yes | confirmed | not reported | viral | yes | no | NR | no | NR | NR | NR | NR | not reported |
| Tagarro-2017 | yes | mixed | pulmonary | mixed | yes (but not f | no | NR | no | NR | NR | NR | NR | not reported |
| Tassniyom-1993 | yes | confirmed | not reported | viral | yes | no | NR | no | NR | NR | NR | NR | not reported |
| Valoor-2009 | yes | mixed | mixed | mixed | yes (but not f | no | NR | yes | 7 (3–16) / 8 ( | 10 (3–38) / 13 | NR | NR | not reported |

| | BASELINE – RESPIRATORY PSS DOMAIN | | | | BASELINE – CARDIOVASCULAR PSS DOMAIN | | | | | | BASELINE – COAGULATION PSS DOMAIN | | | | BASELINE – NEUROLOGIC PSS DOMAIN | | PSS RECONSTRUCTION | | | SEPTIC SHOCK RECONSTRUCTION | OVERALL RECLASSIFICATION |
|---|---|---|---|---|---|---|---|---|---|---|---|---|---|---|---|---|---|---|---|---|---|
| Trial_ID | PaO2_FiO2 | SpO2_FiO2 | Respiratory_support | Mechanical_ventilation_baseline | MAP | Hypotension_reported | Vasopressor_use_baseline | Vasopressor_type | Vasopressor_dose_reported | Lactate_baseline | Platelets | INR | D_dimer | Fibrinogen | GCS | Fixed_pupils_bilaterally | PSS_reconstructed_possible | Reconstructed_PSS_minimum | PSS_>=2_compatible | Septic_shock_PSS_compatible | Final_population_classification (whole population) |
| Short study identifier | Numeric value | Numeric value | none;oxygen;HFNC;NIV;IMV;mixed; not reported | yes;no; yes (but not for all patients); not reported | Numeric value | yes;no; age-adjusted | yes;no; yes (but not all patients); not reported | norepinephrine;dopamine;other; not reported | yes;no | Numeric value | Numeric value | Numeric value | Numeric value | Numeric value | Numeric value | yes;no; not reported | yes; partial; no | Numeric value (if reconstruction possible) | yes;likely;unclear;no | yes;partial; unclear;no | PS-2024 compatible;likely compatible;unclear;likely incompatible; incompatible |
| Branco-2014 | NR | NR | not reported | not reported | NR | no | not reported | not reported | no | NR | NR | NR | NR | NR | NR | not reported | no | | unclear | unclear | unclear |
| deGraaf-2014 | NR | NR | not reported | not reported | NR | no | not reported | not reported | no | NR | NR | NR | NR | NR | NR | not reported | no | | unclear | unclear | unclear |
| El-Nawawy-2017 | NR | NR | not reported | not reported | NR | age-adjustec | not reported | not reported | no | NR | 310 ±241.8 / : | 1.9 ±0.9 / 1.6 | NR | NR | NR | not reported | partial | 1 | unclear | unclear | unclear |
| Mansour-2012 | NR | NR | not reported | not reported | NR | no | not reported | not reported | no | NR | NR | NR | NR | NR | NR | not reported | no | | unclear | unclear | unclear |
| Menon-2017 | NR | NR | mixed | yes (but not f | NR | no | yes | not reported | yes | 2.5 (1.2, 3.6) | NR | NR | NR | NR | NR | not reported | partial | 1 | unclear | unclear | unclear |
| Min-1975 | NR | NR | not reported | not reported | NR | yes | no | not reported | no | NR | NR | NR | NR | NR | NR | not reported | no | | unclear | unclear | unclear |
| Nagy-2013 | NR | NR | oxygen | no | NR | no | no | not reported | no | NR | NR | NR | NR | NR | NR | not reported | no | | no | no | likely incompatible |
| Pongpanich-1973 | NR | NR | not reported | not reported | NR | yes | no | not reported | no | NR | NR | NR | NR | NR | NR | not reported | no | | unclear | unclear | unclear |
| Slusher-1996 | NR | NR | not reported | no | NR | yes | no | not reported | no | NR | NR | NR | NR | NR | NR | not reported | no | | unclear | unclear | unclear |
| Sumarmo-1982 | NR | NR | not reported | not reported | NR | yes | no | not reported | no | NR | NR | NR | NR | NR | NR | not reported | no | | unclear | unclear | unclear |
| Tagarro-2017 | NR | NR | oxygen | no | NR | no | no | not reported | no | NR | NR | NR | NR | NR | NR | not reported | no | | no | no | likely incompatible |
| Tassniyom-1993 | NR | NR | not reported | not reported | NR | yes | no | not reported | no | NR | NR | NR | NR | NR | NR | not reported | no | | unclear | unclear | unclear |
| Valoor-2009 | NR | NR | not reported | no | NR | no | yes | norepinephr | no | NR | NR | NR | NR | NR | NR | not reported | partial | 1 | unclear | unclear | likely compatible |

**Supplementary Table S15: Overview of all RCTs classified as likely incompatible or incompatible, including the number and identity of systematic reviews in which each RCT was included**

| RCT ID | classification_status | n_SRs_including_RCT | SRs_including_RCT | clusters |
|---|---|---|---|---|
| Meduri 2007 | likely incompatible | 8 | Annane 2015; Annane 2019; Annane 2025; Fang 2019; Liang 2021; Lin 2019; Pitre 2024; Rochwerg 2018 | 3 |
| Meijvis 2011 | likely incompatible | 8 | Annane 2015; Annane 2019; Annane 2025; Fang 2019; Liang 2021; Lin 2019; Pitre 2024; Rochwerg 2018 | 3 |
| Rezk 2013 | likely incompatible | 8 | Annane 2015; Annane 2019; Annane 2025; Fang 2019; Liang 2021; Lin 2019; Pitre 2024; Rochwerg 2018 | 3 |
| Snijders 2010 | likely incompatible | 8 | Annane 2015; Annane 2019; Annane 2025; Fang 2019; Liang 2021; Lin 2019; Pitre 2024; Rochwerg 2018 | 3 |
| Jamshidi 2021 | likely incompatible | 5 | Annane 2025; Deng 2024; Kato 2023; Lu 2023; Saghafi 2024 | 3; 4; 6 |
| Blum 2015 | likely incompatible | 3 | Annane 2019; Annane 2025; Liang 2021 | 3 |
| Nafae 2013 | likely incompatible | 3 | Annane 2019; Annane 2025; Liang 2021 | 3 |
| Nagy 2013 | likely incompatible | 3 | Annane 2019; Annane 2025; Deng 2025 | 3; 7 |
| McHardy 1972 | likely incompatible | 2 | Annane 2019; Annane 2025 | 3 |
| Tagaro 2017 | likely incompatible | 2 | Annane 2019; Annane 2025 | 3 |
| Corral Gudino 2021 | likely incompatible | 1 | Annane 2025 | 3 |
| Ghanei 2021 | likely incompatible | 1 | Annane 2025 | 3 |
| Horby 2021 | likely incompatible | 1 | Annane 2025 | 3 |
| Villar 2020 | likely incompatible | 1 | Annane 2025 | 3 |
| Wittermans 2020 | likely incompatible | 1 | Annane 2025 | 3 |
| CSG 1963 | incompatible | 6 | Annane 2015; Annane 2019; Annane 2025; Pitre 2024; Rochwerg 2018; Rygard 2018 | 3; 13 |
| Klastersky 1971 | incompatible | 2 | Fang 2019; Liang 2021 | 3 |
| Edalatifard 2020 | incompatible | 1 | Liang 2021 | 3 |

#### Supplementary Table S16: Systematic reviews including at least one RCT classified as likely incompatible or incompatible (n = 14)

| SR | cluster_id | n_RCT_in_SR | n_likely_incompatible | n_incompatible | n_likely_incompatible_or_incompatible | n_no_fulltext | prop_likely_incompatible_or_incompatible |
|---|---|---|---|---|---|---|---|
| Annane 2025 | 3 | 87 | 15 | 1 | 16 | 9 | 0,18390805 |
| Annane 2019 | 3 | 61 | 9 | 1 | 10 | 9 | 0,16393443 |
| Liang 2021 | 3 | 50 | 6 | 2 | 8 | 3 | 0,16 |
| Pitre 2024 | 3 | 45 | 4 | 1 | 5 | 3 | 0,11111111 |
| Rochwerg 2018 | 3 | 42 | 4 | 1 | 5 | 3 | 0,11904762 |
| Fang 2019 | 3 | 37 | 4 | 1 | 5 | 4 | 0,13513514 |
| Annane 2015 | 3 | 33 | 4 | 1 | 5 | 3 | 0,15151515 |
| Lin 2019 | 3 | 30 | 4 | 0 | 4 | 1 | 0,13333333 |
| Rygard 2018 | 13 | 23 | 0 | 1 | 1 | 4 | 0,04347826 |
| Deng 2024 | 6 | 15 | 1 | 0 | 1 | 1 | 0,06666667 |
| Deng 2025 | 7 | 15 | 1 | 0 | 1 | 5 | 0,06666667 |
| Saghafi 2024 | 4 | 11 | 1 | 0 | 1 | 0 | 0,09090909 |
| Kato 2023 | 4 | 9 | 1 | 0 | 1 | 0 | 0,11111111 |
| Lu 2023 | 4 | 8 | 1 | 0 | 1 | 0 | 0,125 |

**Supplementary Table S17: Alignment between PICO profiles and study pool composition - PICO profiling of SRs and clustering according to study pools (n = 42)**

**PICO domains with categories:**

**Population – Age (PA1-4):** Adults (PA1); Children (PA2); Mixed-age populations (PA3); Age not defined (PA4).

**Population – Target (PD1-4):** Sepsis irrespective of shock status (PD1); Sepsis without shock (PD2); Septic shock only (PD3); Other (PD4)

**Intervention – Corticosteroid Type (IT1-5):** Any/multiple corticosteroids without restriction (IT1); Hydrocortisone (IT2); Dexamethasone (IT3); Methylprednisolone (IT4); Other corticosteroids (IT5).

**Intervention – Dose (ID0-2):** Dose not defined (ID0); Low-dose regimen (ID1); High-dose regimen (ID2).

**Intervention – Timing and Schedule (ITM0-3):** Timing or schedule not defined (ITM0); Standard administration schedule (ITM1); Specific timing focus (ITM2); Specific schedule focus (ITM3).

**Intervention – Combination Strategy (IC0-4):** Monotherapy or combination not defined (IC0); Monotherapy (IC1); Hydrocortisone plus fludrocortisone (HC+FC) (IC2); hydrocortisone, ascorbic acid, and thiamine (HAT) (IC3); Other predefined combinations (IC4).

**Comparator (C1-3):** Standard care, placebo, or no corticosteroid (C1); Active corticosteroid comparator (C2); Other, unclear, or not defined comparator (C3).

**Cluster analysis: 17 cluster**

| **SR ID** | **PICO profiles** | | | | | | | **Cluster analysis** | | |
|---|---|---|---|---|---|---|---|---|---|---|
| | I_combination_strategy (IC0-IC4) | I_type (IT1-5) | P_target (PD1-PD4) | P_age (PA1-PA4) | I_timing (ITM0-3) | I_dose (ID0-2) | Comparator (C1-3) | **Cluster rank** | **Cluster number** | **Order dendrogram** |
| Annane 2025 | Mono+HAT | Any/Multiple | Sepsis±Shock | AnyAge(Mixed) | TS_Standard+ScheduleFocus | Low+High | C1+C2 | 1 | **3** | 25 |

| | | | | | | | | | | |
|---|---|---|---|---|---|---|---|---|---|---|
| Pitre 2024 | Mono+HC+FC+HAT | Any/Multiple | Sepsis±Shock | AnyAge(Mixed) | TS_ND | DoseND | C1 | 1 | **3** | 19 |
| Fang 2019 | Mono/ComboND | Any/Multiple | Sepsis±Shock | Adults | TS_ND | DoseND | C1 | 1 | **3** | 21 |
| Liang 2021 | Mono/ComboND | Any/Multiple | Sepsis±Shock | Adults | TS_ND | DoseND | C1 | 1 | **3** | 23 |
| Lin 2019 | Mono/ComboND | Any/Multiple | Sepsis±Shock | Adults | TS_ND | DoseND | C1 | 1 | **3** | 22 |
| Annane 2015 | Mono/ComboND | Any/Multiple | Sepsis±Shock | AnyAge(Mixed) | TS_Standard | Low+High | C1 | 1 | **3** | 18 |
| Annane 2019 | Mono/ComboND | Any/Multiple | Sepsis±Shock | AnyAge(Mixed) | TS_Standard+ScheduleFocus | Low+High | C1+C2 | 1 | **3** | 24 |
| Rochwerg 2018 | Mono/ComboND | Any/Multiple | Sepsis±Shock | AnyAge(Mixed) | TS_ND | DoseND | C1 | 1 | **3** | 20 |
| Assouline 2021 | HAT | HC | Sepsis±Shock | Adults | TS_ND | DoseND | C1+C2 | 2 | **4** | 12 |
| Lu 2023 | HAT | HC | Sepsis±Shock | Adults | TS_Standard | DoseND | C1 | 2 | **4** | 7 |
| Na 2021 | HAT | HC | Sepsis±Shock | Adults | TS_ND | DoseND | C1+C2 | 2 | **4** | 13 |
| Saghafi 2024 | HAT | HC | Sepsis±Shock | Adults | TS_ND | DoseND | C3 | 2 | **4** | 11 |
| Wu 2021 | HAT | HC | Sepsis±Shock | Adults | TS_ND | DoseND | C1+C2 | 2 | **4** | 8 |
| Yao 2021 | HAT | HC | Sepsis±Shock | AgeND | TS_ND | DoseND | C1 | 2 | **4** | 10 |
| Somagutta 2021 | HAT | HC | ShockOnly | Adults | TS_ND | DoseND | C1 | 2 | **4** | 9 |
| Kato 2023 | Mono+HAT | HC | Sepsis±Shock | AgeND | TS_ND | DoseND | C3 | 2 | **4** | 6 |
| Zhou 2018 | Mono+HC+FC+HAT+OtherCombo | Any/Multiple | ShockOnly | Adults | TS_Standard | Low+High | C1 | 3 | **11** | 30 |
| Ni 2019 | Mono/ComboND | Any/Multiple | Sepsis±Shock | Adults | TS_ND | DoseND | C1 | 3 | **11** | 33 |
| Lian 2019 | Mono/ComboND | Any/Multiple | ShockOnly | Adults | TS_ND | DoseND | C1 | 3 | **11** | 31 |
| Wen 2019 | Mono/ComboND | Any/Multiple | ShockOnly | Adults | TS_ND | DoseND | C3 | 3 | **11** | 32 |

| | | | | | | | | | | |
|---|---|---|---|---|---|---|---|---|---|---|
| Lyu 2020 | Mono/ComboND | HC | ShockOnly | Adults | TS_ND | Low | C1 | 3 | **11** | 34 |
| Wu 2020 | Mono/ComboND | HC | ShockOnly | AgeND | TS_ND | Low | C1 | 3 | **11** | 35 |
| Barakakis 2019 | HC+FC | Any/Multiple | ShockOnly | Adults | TS_ND | Low | C1 | 4 | **5** | 36 |
| Yamamoto 2020 | HC+FC | HC | ShockOnly | Adults | TS_ND | DoseND | C1+C2 | 4 | **5** | 38 |
| Wu 2024 | HC+FC | HC | ShockOnly | AgeND | TS_ND | DoseND | C1+C2 | 4 | **5** | 37 |
| Siddiqi 2019 | Mono+HC+FC | HC | ShockOnly | Adults | TS_ND | DoseND | C1 | 5 | **1** | 42 |
| Aletreby 2019 | Mono+HC+FC+OtherCombo | HC | Sepsis±Shock | Adults | TS_Standard | Low+High | C1 | 5 | **1** | 41 |
| Deng 2025 | Mono/ComboND | Any/Multiple | Sepsis±Shock+Other | Children | TS_ND | DoseND | C1 | 6 | **7** | 15 |
| Yang 2020 | Mono/ComboND | Any/Multiple | ShockOnly | Children | TS_ND | DoseND | C3 | 6 | **7** | 16 |
| Zayed 2022 | HAT | HC | Sepsis±Shock | Adults | TS_ND | DoseND | C3 | 7 | **9** | 4 |
| Lee 2022 | HAT | HC | Sepsis±Shock | AgeND | TS_ND | DoseND | C1 | 7 | **9** | 3 |
| Lu 2021 | Mono/ComboND | Any/Multiple | ShockOnly | Adults | TS_Standard | Low+High | C1 | 8 | **12** | 26 |
| Xu 2018 | Mono/ComboND | Any/Multiple | ShockOnly | Adults | TS_Standard | Low | C3 | 8 | **12** | 27 |
| Allen 2019 | Mono/ComboND | Any/Multiple | Sepsis±Shock | Adults | TS_ND | DoseND | C3 | 9 | **2** | 39 |
| Deng 2024 | HAT | HC | Sepsis±Shock | Adults | TS_ND | DoseND | C1+C2 | 10 | **6** | 5 |
| Fratoni 2020 | Mono/ComboND | Dex | Sepsis±Shock | Adults | TS_Standard | Low+High | C1 | 11 | **8** | 17 |
| Li 2025 | Mono/ComboND | HC | ShockOnly | Adults | ScheduleFocus | DoseND | C2 | 12 | **10** | 14 |
| Rygard 2018 | Mono/ComboND | Any/Multiple | ShockOnly | Adults | TS_ND | Low+High | C1 | 13 | **13** | 29 |
| Son 2021 | Mono/ComboND | Any/Multiple | Sepsis±Shock | Adults | TS_ND | DoseND | C1 | 14 | **14** | 28 |
| Song B 2024 | Mono/ComboND | HC | ShockOnly | Adults | TimingFocus | Low | C2 | 15 | **15** | 1 |

| | | | | | | | | | | |
|---|---|---|---|---|---|---|---|---|---|---|
| Song Y 2024 | Mono/ComboND | Any/Multiple | Sepsis±Shock | AgeND | TS_ND | DoseND | C1 | 16 | **16** | 40 |
| Sudaryadi 2023 | HAT | HC | ShockOnly | Adults | TS_ND | DoseND | C2 | 17 | **17** | 2 |

**Supplementary Table S18: Alignment between PICO profiles and study pool composition - PICO profiling of SRs and clustering according to study pools (n = 42)**

| SR ID | Year | Time category | N RCT meta | Effect measure | Point estimate | CI | P value | model | GRADE | Effect category | Intervention combination strategy | Population target | Cluster no. | N RCT SR |
|---|---|---|---|---|---|---|---|---|---|---|---|---|---|---|
| Assouline 2021 | 2021 | 28-d | 8 | RR | 1,02 | 0.86 to 1.20 | 0,85 | Fixed | NR | no evidence of effect | HAT | Sepsis±Shock | 4 | 8 |
| Deng 2024 | 2024 | NR | 12 | RR | 0,81 | 0.63 to 1.06 | NR | Random | Low | no evidence of effect | HAT | Sepsis±Shock | 6 | 15 |
| Lu 2023 | 2023 | 28-d | 8 | RR | 0,96 | 0.83 to 1.11 | 0,6 | Fixed | NR | no evidence of effect | HAT | Sepsis±Shock | 4 | 8 |
| Na 2021 | 2021 | in-hospital | 5 | RR | 1,07 | 0.85 to 1.34 | 0,58 | Random | NR | no evidence of effect | HAT | Sepsis±Shock | 4 | 10 |
| Saghafi 2024 | 2024 | 28-d | 6 | OR | 1,013 | 0.819 to 1.267 | 0,914 | Random | Moderate | no evidence of effect | HAT | Sepsis±Shock | 4 | 11 |
| Wu 2021 | 2021 | 28-d | 4 | RR | 1,04 | 0.81 to 1.33 | 0,76 | Random | NR | no evidence of effect | HAT | Sepsis±Shock | 4 | 6 |
| Zayed 2022 | 2022 | 28-d-mixed | 6 | RR | 1,05 | 0.85 to 1.30 | 0,64 | Random | NR | no evidence of effect | HAT | Sepsis±Shock | 9 | 7 |
| Lee 2022 | 2022 | NA | 0 | NA | NA | NA | NA | NA | NA | no meta-analysis | HAT | Sepsis±Shock | 9 | 6 |
| Yao 2021 | 2021 | 28-d-mixed | 8 | RR | 1,02 | 0.87 to 1.20 | 0,81 | Random | High | no evidence of effect | HAT | Sepsis±Shock | 4 | 8 |

| | | | | | | | | | | | | | | |
|---|---|---|---|---|---|---|---|---|---|---|---|---|---|---|
| Somagutta 2021 | 2021 | in-hospital | 8 | RR | 0,99 | 0.83 to 1.18 | 0,92 | Random | NR | no evidence of effect | HAT | ShockOnly | 4 | 8 |
| Sudaryadi 2023 | 2023 | 28-d | 2 | OR | 0,95 | 0.56 to 1.58 | 0,83 | Fixed | NR | no evidence of effect | HAT | ShockOnly | 17 | 3 |
| Barakakis 2019 | 2019 | 28-d | 2 | OR | 0,79 | 0.64 to 0.97 | NR | Random | NR | benefit | HC+ FC | ShockOnly | 5 | 2 |
| Yamamoto 2020 | 2020 | 28-d | 2 | RR | 0,88 | 0.78 to 0.99 | 0,03 | Random | High | benefit | HC+ FC | ShockOnly | 5 | 3 |
| Wu 2024 | 2024 | in-hospital | 3 | OR | 0,8 | 0.67 to 0.95 | NR | Random | NR | benefit | HC+ FC | ShockOnly | 5 | 3 |
| Annane 2025 | 2025 | 28-d-mixed | 72 | RR | 0,89 | 0.84 to 0.95 | 0,00 07 | Random | Mode rate | benefit | Mono +HAT | Sepsis±Shock | 3 | 87 |
| Kato 2023 | 2023 | 28-d | 3 | OR | 0,88 | 0.56 to 1.39 | 0,59 | Fixed | NR | no evidence of effect | Mono +HAT | Sepsis±Shock | 4 | 9 |
| Siddiqi 2019 | 2019 | 28-d | 5 | RR | 0,92 | 0.85 to 0.99 | 0,04 | Random | NR | benefit | Mono +HC +FC | ShockOnly | 1 | 5 |
| Pitre 2024 | 2024 | 28-d-mixed | 39 | RR | 0,93 | 0.88 to 0.99 | 0,01 | Random | Mode rate | benefit | Mono +HC +FC+ HAT | Sepsis±Shock | 3 | 45 |
| Zhou 2018 | 2018 | 28-d-mixed | 17 | RR | 0,92 | 0.84 to 1.02 | 0,10 4 | Random | NR | no evidence of effect | Mono +HC +FC+ HAT+ Other Com bo | ShockOnly | 11 | 17 |
| Aletreby 2019 | 2019 | 28-d | 9 | RR | 0,93 | 0.86 to 1.01 | 0,70 2 | Random | Very low | no evidence of effect | Mono +HC +FC+ Other Com bo | Sepsis±Shock | 1 | 10 |

| | | | | | | | | | | | | | | |
|---|---|---|---|---|---|---|---|---|---|---|---|---|---|---|
| Allen 2019 | 2019 | NA | 0 | NA | NA | NA | NA | NA | NA | no meta-analysis | Mono/ComboND | Sepsis±Shock | 2 | 7 |
| Fang 2019 | 2019 | 28-d-mixed | 34 | RR | 0,9 | 0.82 to 0.98 | 0,02 | Random | Moderate | benefit | Mono/ComboND | Sepsis±Shock | 3 | 37 |
| Liang 2021 | 2021 | 28-d | 40 | RR | 0,94 | 0.87 to 1.02 | 0,12 | Random | Moderate | no evidence of effect | Mono/ComboND | Sepsis±Shock | 3 | 50 |
| Lin 2019 | 2019 | 28-d | 21 | RR | 0,9 | 0.84 to 0.97 | NR | Fixed | High | benefit | Mono/ComboND | Sepsis±Shock | 3 | 30 |
| Ni 2019 | 2019 | 28-d | 18 | RR | 0,91 | 0.85 to 0.98 | 0,01 | Fixed | NR | benefit | Mono/ComboND | Sepsis±Shock | 11 | 19 |
| Son 2021 | 2021 | 28-d-mixed | 17 | RR | 0,93 | 0.88 to 0.99 | 0,02 | Random | NR | benefit | Mono/ComboND | Sepsis±Shock | 14 | 22 |
| Song Y 2024 | 2024 | 28-d | 7 | OR | 0,87 | 0.78 to 0.98 | 0,03 | Fixed | NR | benefit | Mono/ComboND | Sepsis±Shock | 16 | 9 |
| Annane 2015 | 2015 | 28-d-mixed | 27 | RR | 0,87 | 0.76 to 1.00 | 0,052 | Random | Low | no evidence of effect | Mono/ComboND | Sepsis±Shock | 3 | 33 |
| Annane 2019 | 2019 | 28-d-mixed | 50 | RR | 0,91 | 0.84 to 0.99 | 0,04 | Random | Moderate | benefit | Mono/ComboND | Sepsis±Shock | 3 | 61 |

| | | | | | | | | | | | | | | |
|---|---|---|---|---|---|---|---|---|---|---|---|---|---|---|
| Rochwerg 2018 | 2018 | 28-d | 36 | RR | 0,93 | 0.84 to 1.03 | 0,15 | Random | Low | no evidence of effect | Mono /Com boN D | Sepsis±Shock | 3 | 42 |
| Deng 2025 | 2025 | 28-d-mixed | 13 | RR | 0,65 | 0.50 to 0.85 | 0,56 | Random | Low | benefit | Mono /Com boN D | Sepsis±Shock + NonSepsis (children) | 7 | 15 |
| Lian 2019 | 2019 | 28-d | 21 | RR | 0,93 | 0.88 to 0.99 | 0,02 | Fixed | Mode rate | benefit | Mono /Com boN D | ShockOnly | 11 | 21 |
| Lu 2021 | 2021 | 28-d-mixed | 9 | RR | 0,95 | 0.85 to 1.06 | 0,37 | Random | Mode rate | no evidence of effect | Mono /Com boN D | ShockOnly | 12 | 9 |
| Rygard 2018 | 2018 | 90-d | 22 | RR | 0,96 | 0.91 to 1.02 | 0,21 | Fixed | High | no evidence of effect | Mono /Com boN D | ShockOnly | 13 | 23 |
| Wen 2019 | 2019 | 28-d | 12 | RR | 0,94 | 0.84 to 1.05 | 0,28 5 | Random | NR | no evidence of effect | Mono /Com boN D | ShockOnly | 11 | 18 |
| Xu 2018 | 2018 | 28-d | 9 | RR | 0,96 | 0.85 to 1.09 | 0,53 | Fixed | NR | no evidence of effect | Mono /Com boN D | ShockOnly | 12 | 9 |
| Yang 2020 | 2020 | 28-d-mixed | 12 | OR | 0,67 | 0.46 to 0.98 | 0,41 | Fixed | Low | benefit | Mono /Com boN D | ShockOnly (children) | 7 | 10 |
| Fratoni 2020 | 2020 | NA | 0 | NA | NA | NA | NA | NA | NA | no meta-analysis | Mono /Com boN D | Sepsis±Shock | 8 | 5 |

| | | | | | | | | | | | | | | |
|---|---|---|---|---|---|---|---|---|---|---|---|---|---|---|
| Li 2025 | 2025 | 28-d-mixed | 3 | OR | 0,75 | 0.42 to 1.37 | 0,35 | Fixed | NR | no evidence of effect | Mono/CombоND | ShockOnly | 10 | 3 |
| Lyu 2020 | 2020 | 28-d | 11 | OR | 0,9 | 0.81 to 1.00 | 0,05 | Fixed | NR | no evidence of effect | Mono/ComboND | ShockOnly | 11 | 13 |
| Song B 2024 | 2024 | ICU | 0 | NA | NA | NA | NA | NA | NA | no meta-analysis | Mono/ComboND | ShockOnly | 15 | 0 |
| Wu 2020 | 2020 | 28-d | 10 | RR | 0,96 | 0.88 to 1.05 | 0,37 | Fixed | NR | no evidence of effect | Mono/ComboND | ShockOnly | 11 | 12 |

**Supplementary Table S19: SRxRCT mortality matrix showing randomized controlled trials (n=121) included, meta-analyzed, and not included across systematic reviews (n = 42)**

0: RCT not included in SR

1: RCT included in SR and contributed to meta-analysis of short-term mortality

2: RCT included in SR, but not meta-analyzed

| rct_id | Aletreby 2019 | Allen 2019 | Annane 2015 | Annane 2019 | Annane 2025 | Assouline 2021 | Barakakis 2019 | Deng 2024 | Deng 2025 | Fang 2019 | Fratoni 2020 | Kato 2023 | Lee 2022 | Li 2025 | Lian 2019 | Liang 2021 | Lin 2019 | Lu 2021 | Lu 2023 | Lyu 2020 | Na 2021 | Ni 2019 | Pitre 2024 | Rochwerg 2018 | Rygard 2018 | Saghafi 2024 | Siddiqi 2019 | Somagutta 2021 | Son 2021 | Song B 2024 | Song Y 2024 | Sudaryadi 2023 | Wen 2019 | Wu 2020 | Wu 2021 | Wu 2024 | Xu 2018 | Yamamoto 2020 | Yang 2020 | Yao 2021 | Zayed 2022 | Zhou 2018 |
|---|---|---|---|---|---|---|---|---|---|---|---|---|---|---|---|---|---|---|---|---|---|---|---|---|---|---|---|---|---|---|---|---|---|---|---|---|---|---|---|---|---|---|
| Aboab 2008 | 0 | 0 | 0 | 1 | 1 | 0 | 0 | 0 | 0 | 1 | 0 | 0 | 0 | 0 | 0 | 1 | 0 | 0 | 0 | 0 | 0 | 0 | 0 | 0 | 1 | 0 | 0 | 0 | 0 | 0 | 0 | 0 | 0 | 0 | 0 | 0 | 0 | 0 | 0 | 0 | 0 | 1 |
| Agarwal 2022 | 0 | 0 | 0 | 0 | 1 | 0 | 0 | 0 | 0 | 0 | 0 | 0 | 0 | 0 | 0 | 0 | 0 | 0 | 0 | 0 | 0 | 0 | 1 | 0 | 0 | 0 | 0 | 0 | 0 | 0 | 0 | 0 | 0 | 0 | 0 | 0 | 0 | 0 | 0 | 0 | 0 | 0 |
| Angus 2020 | 0 | 0 | 0 | 0 | 1 | 0 | 0 | 0 | 0 | 0 | 0 | 0 | 0 | 0 | 0 | 2 | 0 | 0 | 0 | 0 | 0 | 0 | 0 | 0 | 0 | 0 | 0 | 0 | 0 | 0 | 0 | 0 | 0 | 0 | 0 | 0 | 0 | 0 | 0 | 0 | 0 | 0 |
| Annane 2002 | 1 | 0 | 1 | 1 | 1 | 0 | 1 | 0 | 0 | 1 | 0 | 0 | 0 | 0 | 1 | 1 | 1 | 1 | 0 | 1 | 0 | 1 | 1 | 1 | 1 | 0 | 1 | 0 | 1 | 0 | 1 | 0 | 1 | 0 | 0 | 1 | 1 | 1 | 0 | 0 | 0 | 1 |
| Annane 2010 | 0 | 0 | 2 | 2 | 2 | 0 | 0 | 0 | 0 | 0 | 0 | 0 | 0 | 0 | 0 | 2 | 0 | 0 | 0 | 0 | 0 | 0 | 0 | 0 | 0 | 0 | 0 | 0 | 0 | 0 | 0 | 0 | 0 | 0 | 0 | 1 | 0 | 2 | 0 | 0 | 0 | 0 |
| Annane 2018 | 1 | 2 | 0 | 1 | 1 | 0 | 1 | 0 | 0 | 1 | 0 | 0 | 0 | 0 | 1 | 1 | 1 | 0 | 0 | 1 | 0 | 1 | 1 | 1 | 1 | 0 | 1 | 0 | 1 | 0 | 1 | 0 | 1 | 0 | 0 | 1 | 0 | 1 | 0 | 0 | 0 | 1 |
| Arabi 2010 | 0 | 0 | 1 | 1 | 1 | 0 | 0 | 0 | 0 | 1 | 0 | 0 | 0 | 0 | 1 | 1 | 1 | 0 | 0 | 1 | 0 | 1 | 1 | 1 | 1 | 0 | 0 | 0 | 0 | 0 | 1 | 0 | 1 | 1 | 0 | 0 | 1 | 0 | 0 | 0 | 0 | 1 |
| Balakrishnan 2018 | 0 | 0 | 0 | 0 | 0 | 0 | 0 | 0 | 0 | 0 | 0 | 0 | 2 | 0 | 0 | 0 | 0 | 0 | 0 | 0 | 2 | 0 | 0 | 0 | 0 | 2 | 0 | 0 | 0 | 0 | 0 | 0 | 0 | 0 | 0 | 0 | 0 | 0 | 0 | 0 | 2 | 0 |
| Birudaraju 2022 | 0 | 0 | 0 | 0 | 1 | 0 | 0 | 0 | 0 | 0 | 0 | 0 | 0 | 0 | 0 | 0 | 0 | 0 | 0 | 0 | 0 | 0 | 1 | 0 | 0 | 0 | 0 | 0 | 0 | 0 | 0 | 0 | 0 | 0 | 0 | 0 | 0 | 0 | 0 | 0 | 0 | 0 |
| Blum 2015 | 0 | 0 | 0 | 1 | 1 | 0 | 0 | 0 | 0 | 0 | 0 | 0 | 0 | 0 | 0 | 1 | 0 | 0 | 0 | 0 | 0 | 0 | 0 | 0 | 0 | 0 | 0 | 0 | 0 | 0 | 0 | 0 | 0 | 0 | 0 | 0 | 0 | 0 | 0 | 0 | 0 | 0 |
| Bollaert 1998 | 0 | 0 | 1 | 1 | 1 | 0 | 0 | 0 | 0 | 1 | 0 | 0 | 0 | 0 | 1 | 1 | 1 | 0 | 0 | 1 | 0 | 1 | 1 | 1 | 1 | 0 | 0 | 0 | 1 | 0 | 0 | 0 | 1 | 1 | 0 | 0 | 1 | 0 | 0 | 0 | 0 | 1 |
| Bone 1987 | 0 | 0 | 1 | 1 | 1 | 0 | 0 | 0 | 0 | 2 | 0 | 0 | 0 | 0 | 1 | 1 | 2 | 0 | 0 | 0 | 0 | 0 | 1 | 1 | 0 | 0 | 0 | 0 | 1 | 0 | 0 | 0 | 2 | 0 | 0 | 0 | 0 | 0 | 0 | 0 | 0 | 0 |
| Branco 2014 | 0 | 0 | 0 | 0 | 0 | 0 | 0 | 0 | 0 | 0 | 0 | 0 | 0 | 0 | 0 | 0 | 0 | 0 | 0 | 0 | 0 | 0 | 2 | 2 | 0 | 0 | 0 | 0 | 0 | 0 | 0 | 0 | 0 | 0 | 0 | 0 | 0 | 0 | 0 | 0 | 0 | 0 |
| Briegel 1999 | 0 | 0 | 1 | 1 | 1 | 0 | 0 | 0 | 0 | 1 | 0 | 0 | 0 | 0 | 1 | 1 | 1 | 1 | 0 | 1 | 0 | 1 | 1 | 1 | 1 | 0 | 0 | 0 | 1 | 0 | 0 | 0 | 2 | 2 | 0 | 0 | 1 | 0 | 0 | 0 | 0 | 1 |
| Briegel 2001 | 0 | 0 | 0 | 0 | 0 | 0 | 0 | 0 | 0 | 0 | 0 | 0 | 0 | 0 | 0 | 0 | 0 | 0 | 0 | 0 | 0 | 0 | 0 | 0 | 0 | 0 | 0 | 0 | 1 | 0 | 0 | 0 | 0 | 0 | 0 | 0 | 0 | 0 | 0 | 0 | 0 | 0 |
| Chang 2020 | 0 | 0 | 0 | 0 | 1 | 1 | 0 | 1 | 0 | 0 | 0 | 1 | 0 | 0 | 0 | 0 | 0 | 0 | 1 | 0 | 2 | 0 | 0 | 0 | 0 | 1 | 0 | 1 | 0 | 0 | 0 | 0 | 0 | 0 | 1 | 0 | 0 | 0 | 0 | 1 | 1 | 0 |
| Chawla 1999 | 0 | 0 | 1 | 1 | 1 | 0 | 0 | 0 | 0 | 1 | 0 | 0 | 0 | 0 | 1 | 1 | 1 | 0 | 0 | 0 | 0 | 0 | 1 | 1 | 1 | 0 | 0 | 0 | 0 | 0 | 0 | 0 | 2 | 0 | 0 | 0 | 0 | 0 | 0 | 0 | 0 | 0 |
| Cicarelli 2006 | 0 | 0 | 0 | 0 | 0 | 0 | 0 | 0 | 0 | 0 | 2 | 0 | 0 | 0 | 0 | 1 | 0 | 0 | 0 | 0 | 0 | 0 | 0 | 0 | 0 | 0 | 0 | 0 | 0 | 0 | 0 | 0 | 0 | 0 | 0 | 0 | 0 | 0 | 0 | 0 | 0 | 0 |
| Cicarelli 2007 | 0 | 0 | 1 | 1 | 1 | 0 | 0 | 0 | 0 | 1 | 2 | 0 | 0 | 0 | 1 | 0 | 1 | 1 | 0 | 0 | 0 | 1 | 1 | 1 | 1 | 0 | 0 | 0 | 1 | 0 | 0 | 0 | 1 | 0 | 0 | 0 | 1 | 0 | 0 | 0 | 0 | 1 |
| Confalonieri 2005 | 0 | 0 | 1 | 1 | 1 | 0 | 0 | 0 | 0 | 1 | 0 | 0 | 0 | 0 | 0 | 1 | 1 | 0 | 0 | 0 | 0 | 1 | 1 | 1 | 0 | 0 | 0 | 0 | 2 | 0 | 0 | 0 | 0 | 0 | 0 | 0 | 0 | 0 | 0 | 0 | 0 | 0 |
| Corral Gudino 2021 | 0 | 0 | 0 | 0 | 1 | 0 | 0 | 0 | 0 | 0 | 0 | 0 | 0 | 0 | 0 | 0 | 0 | 0 | 0 | 0 | 0 | 0 | 0 | 0 | 0 | 0 | 0 | 0 | 0 | 0 | 0 | 0 | 0 | 0 | 0 | 0 | 0 | 0 | 0 | 0 | 0 | 0 |
| CSG 1963 | 0 | 0 | 2 | 2 | 2 | 0 | 0 | 0 | 0 | 0 | 0 | 0 | 0 | 0 | 0 | 0 | 0 | 0 | 0 | 0 | 0 | 0 | 2 | 2 | 1 | 0 | 0 | 0 | 0 | 0 | 0 | 0 | 0 | 0 | 0 | 0 | 0 | 0 | 0 | 0 | 0 | 0 |
| de Graaf 2014 | 0 | 0 | 0 | 0 | 0 | 0 | 0 | 0 | 1 | 0 | 0 | 0 | 0 | 0 | 0 | 0 | 0 | 0 | 0 | 0 | 0 | 0 | 2 | 2 | 0 | 0 | 0 | 0 | 0 | 0 | 0 | 0 | 0 | 0 | 0 | 0 | 0 | 0 | 1 | 0 | 0 | 0 |
| Deng 2011 | 0 | 0 | 0 | 0 | 0 | 0 | 0 | 0 | 0 | 0 | 0 | 0 | 0 | 0 | 0 | 0 | 0 | 0 | 0 | 0 | 0 | 0 | 0 | 0 | 2 | 0 | 0 | 0 | 0 | 0 | 0 | 0 | 0 | 0 | 0 | 0 | 0 | 0 | 0 | 0 | 0 | 0 |
| Dequin 2020 | 0 | 0 | 0 | 0 | 1 | 0 | 0 | 0 | 0 | 0 | 0 | 0 | 0 | 0 | 0 | 2 | 0 | 0 | 0 | 0 | 0 | 0 | 0 | 0 | 0 | 0 | 0 | 0 | 0 | 0 | 0 | 0 | 0 | 0 | 0 | 0 | 0 | 0 | 0 | 0 | 0 | 0 |
| Dequin 2023 | 0 | 0 | 0 | 0 | 1 | 0 | 0 | 0 | 0 | 0 | 0 | 0 | 0 | 0 | 0 | 0 | 0 | 0 | 0 | 0 | 0 | 0 | 0 | 0 | 0 | 0 | 0 | 0 | 0 | 0 | 0 | 0 | 0 | 0 | 0 | 0 | 0 | 0 | 0 | 0 | 0 | 0 |
| Doluee 2018 | 0 | 0 | 0 | 1 | 1 | 0 | 0 | 0 | 0 | 0 | 0 | 0 | 0 | 0 | 1 | 1 | 0 | 1 | 0 | 0 | 0 | 0 | 1 | 0 | 0 | 0 | 0 | 0 | 0 | 0 | 0 | 0 | 0 | 0 | 0 | 0 | 0 | 0 | 0 | 0 | 0 | 0 |
| Edalatifard 2020 | 0 | 0 | 0 | 0 | 0 | 0 | 0 | 0 | 0 | 0 | 0 | 0 | 0 | 0 | 0 | 2 | 0 | 0 | 0 | 0 | 0 | 0 | 0 | 0 | 0 | 0 | 0 | 0 | 0 | 0 | 0 | 0 | 0 | 0 | 0 | 0 | 0 | 0 | 0 | 0 | 0 | 0 |
| El Ghamrawy 2006 | 0 | 0 | 0 | 1 | 1 | 0 | 0 | 0 | 0 | 0 | 0 | 0 | 0 | 0 | 0 | 0 | 0 | 0 | 0 | 0 | 0 | 0 | 0 | 0 | 0 | 0 | 0 | 0 | 0 | 0 | 0 | 0 | 0 | 0 | 0 | 0 | 0 | 0 | 0 | 0 | 0 | 0 |
| El Nawawy 2017 | 0 | 0 | 0 | 2 | 2 | 0 | 0 | 0 | 0 | 0 | 0 | 0 | 0 | 0 | 0 | 0 | 0 | 0 | 0 | 0 | 0 | 0 | 1 | 1 | 0 | 0 | 0 | 0 | 0 | 0 | 0 | 0 | 0 | 0 | 0 | 0 | 0 | 0 | 1 | 0 | 0 | 0 |
| Feng 2023 | 0 | 0 | 0 | 0 | 0 | 0 | 0 | 1 | 0 | 0 | 0 | 0 | 0 | 0 | 0 | 0 | 0 | 0 | 0 | 0 | 0 | 0 | 0 | 0 | 0 | 0 | 0 | 0 | 0 | 0 | 0 | 0 | 0 | 0 | 0 | 0 | 0 | 0 | 0 | 0 | 0 | 0 |
| Fernandez Serrano 2 | 0 | 0 | 0 | 1 | 1 | 0 | 0 | 0 | 0 | 0 | 0 | 0 | 0 | 0 | 0 | 2 | 0 | 0 | 0 | 0 | 0 | 0 | 0 | 0 | 0 | 0 | 0 | 0 | 0 | 0 | 0 | 0 | 0 | 0 | 0 | 0 | 0 | 0 | 0 | 0 | 0 | 0 |
| Fuji 2020 | 0 | 0 | 0 | 0 | 0 | 1 | 0 | 2 | 0 | 0 | 0 | 1 | 2 | 0 | 0 | 0 | 0 | 0 | 0 | 0 | 1 | 0 | 0 | 0 | 0 | 1 | 0 | 1 | 0 | 0 | 0 | 1 | 0 | 0 | 1 | 0 | 0 | 0 | 0 | 1 | 1 | 0 |
| Ghanei 2021 | 0 | 0 | 0 | 0 | 1 | 0 | 0 | 0 | 0 | 0 | 0 | 0 | 0 | 0 | 0 | 0 | 0 | 0 | 0 | 0 | 0 | 0 | 0 | 0 | 0 | 0 | 0 | 0 | 0 | 0 | 0 | 0 | 0 | 0 | 0 | 0 | 0 | 0 | 0 | 0 | 0 | 0 |
| Gordon 2014 | 1 | 0 | 1 | 1 | 1 | 0 | 0 | 0 | 0 | 1 | 0 | 0 | 0 | 0 | 1 | 1 | 0 | 0 | 0 | 1 | 0 | 1 | 1 | 1 | 1 | 0 | 0 | 0 | 0 | 0 | 1 | 0 | 1 | 1 | 0 | 0 | 1 | 0 | 0 | 0 | 0 | 1 |
| Gordon 2016 | 1 | 2 | 0 | 1 | 1 | 0 | 0 | 0 | 0 | 1 | 0 | 0 | 0 | 0 | 1 | 1 | 0 | 0 | 0 | 1 | 0 | 0 | 1 | 1 | 1 | 0 | 0 | 0 | 0 | 0 | 0 | 0 | 0 | 1 | 0 | 0 | 0 | 0 | 0 | 0 | 0 | 1 |
| Horby 2021 | 0 | 0 | 0 | 0 | 1 | 0 | 0 | 0 | 0 | 0 | 0 | 0 | 0 | 0 | 0 | 0 | 0 | 0 | 0 | 0 | 0 | 0 | 0 | 0 | 0 | 0 | 0 | 0 | 0 | 0 | 0 | 0 | 0 | 0 | 0 | 0 | 0 | 0 | 0 | 0 | 0 | 0 |
| Hu 2009 | 0 | 0 | 1 | 1 | 1 | 0 | 0 | 0 | 0 | 1 | 0 | 0 | 0 | 0 | 1 | 1 | 0 | 0 | 0 | 2 | 0 | 2 | 1 | 1 | 1 | 0 | 0 | 0 | 0 | 0 | 0 | 0 | 2 | 2 | 0 | 0 | 0 | 0 | 0 | 0 | 0 | 1 |
| Huang 2014 | 0 | 0 | 0 | 1 | 1 | 0 | 0 | 0 | 0 | 1 | 0 | 0 | 0 | 0 | 0 | 1 | 0 | 0 | 0 | 0 | 0 | 0 | 0 | 0 | 0 | 0 | 0 | 0 | 0 | 0 | 0 | 0 | 0 | 0 | 0 | 0 | 0 | 0 | 0 | 0 | 0 | 0 |
| Huh 2007 | 0 | 0 | 2 | 2 | 2 | 0 | 0 | 0 | 0 | 0 | 0 | 0 | 0 | 0 | 0 | 0 | 0 | 0 | 0 | 0 | 0 | 0 | 0 | 0 | 0 | 0 | 0 | 0 | 0 | 0 | 0 | 0 | 0 | 0 | 0 | 0 | 0 | 0 | 0 | 0 | 0 | 0 |
| Hussein 2021 | 0 | 0 | 0 | 0 | 0 | 0 | 0 | 2 | 0 | 0 | 0 | 1 | 0 | 0 | 0 | 0 | 0 | 0 | 0 | 0 | 2 | 0 | 0 | 0 | 0 | 0 | 0 | 0 | 0 | 0 | 0 | 1 | 0 | 0 | 0 | 0 | 0 | 0 | 0 | 0 | 0 | 0 |
| Hwang 2020 | 0 | 0 | 0 | 0 | 0 | 0 | 0 | 0 | 0 | 0 | 0 | 0 | 2 | 0 | 0 | 0 | 0 | 0 | 0 | 0 | 0 | 0 | 0 | 0 | 0 | 0 | 0 | 0 | 0 | 0 | 0 | 0 | 0 | 0 | 0 | 0 | 0 | 0 | 0 | 1 | 1 | 0 |
| Hyvernat 2016 | 0 | 2 | 0 | 2 | 2 | 0 | 0 | 0 | 0 | 0 | 0 | 0 | 0 | 0 | 0 | 2 | 0 | 0 | 0 | 0 | 0 | 0 | 0 | 0 | 0 | 0 | 0 | 0 | 0 | 0 | 0 | 0 | 0 | 0 | 0 | 0 | 0 | 0 | 0 | 0 | 0 | 0 |
| Iglesias 2020 | 0 | 0 | 0 | 0 | 1 | 1 | 0 | 1 | 0 | 0 | 0 | 2 | 2 | 0 | 0 | 0 | 0 | 0 | 1 | 0 | 1 | 0 | 0 | 0 | 0 | 2 | 0 | 1 | 0 | 0 | 0 | 0 | 0 | 0 | 2 | 0 | 0 | 0 | 0 | 1 | 1 | 0 |
| Jamshidi 2021 | 0 | 0 | 0 | 0 | 2 | 0 | 0 | 1 | 0 | 0 | 0 | 2 | 0 | 0 | 0 | 0 | 0 | 0 | 1 | 0 | 0 | 0 | 0 | 0 | 0 | 2 | 0 | 0 | 0 | 0 | 0 | 0 | 0 | 0 | 0 | 0 | 0 | 0 | 0 | 0 | 0 | 0 |
| Jiang 2019 | 0 | 0 | 0 | 0 | 2 | 0 | 0 | 0 | 0 | 0 | 0 | 0 | 0 | 0 | 0 | 0 | 0 | 0 | 0 | 0 | 0 | 0 | 0 | 0 | 0 | 0 | 0 | 0 | 0 | 0 | 0 | 0 | 0 | 0 | 0 | 0 | 0 | 0 | 0 | 0 | 0 | 0 |
| Karimpour 2019 | 0 | 0 | 0 | 0 | 0 | 0 | 0 | 0 | 0 | 0 | 0 | 0 | 0 | 0 | 0 | 0 | 0 | 0 | 0 | 0 | 0 | 0 | 0 | 0 | 0 | 0 | 0 | 1 | 0 | 0 | 0 | 0 | 0 | 0 | 0 | 0 | 0 | 0 | 0 | 0 | 0 | 0 |
| Kaufmann 2008 | 0 | 0 | 0 | 0 | 0 | 0 | 0 | 0 | 0 | 0 | 0 | 0 | 0 | 0 | 0 | 0 | 0 | 0 | 0 | 0 | 0 | 0 | 0 | 0 | 0 | 0 | 0 | 0 | 2 | 0 | 0 | 0 | 0 | 0 | 0 | 0 | 0 | 0 | 0 | 0 | 0 | 0 |
| Keh 2003 | 0 | 0 | 2 | 2 | 2 | 0 | 0 | 0 | 0 | 2 | 0 | 0 | 0 | 0 | 0 | 1 | 0 | 0 | 0 | 0 | 0 | 0 | 0 | 0 | 0 | 0 | 0 | 0 | 0 | 0 | 0 | 0 | 0 | 0 | 0 | 0 | 0 | 0 | 0 | 0 | 0 | 0 |
| Keh 2016 | 1 | 2 | 0 | 1 | 1 | 0 | 0 | 0 | 0 | 1 | 0 | 0 | 0 | 0 | 0 | 1 | 1 | 0 | 0 | 0 | 0 | 1 | 1 | 1 | 0 | 0 | 0 | 0 | 1 | 0 | 0 | 0 | 1 | 0 | 0 | 0 | 0 | 0 | 0 | 0 | 0 | 0 |
| Klastersky 1971 | 0 | 0 | 0 | 0 | 0 | 0 | 0 | 0 | 0 | 1 | 0 | 0 | 0 | 0 | 0 | 1 | 0 | 0 | 0 | 0 | 0 | 0 | 0 | 0 | 0 | 0 | 0 | 0 | 0 | 0 | 0 | 0 | 0 | 0 | 0 | 0 | 0 | 0 | 0 | 0 | 0 | 0 |
| Kurungundla 2008 | 0 | 0 | 0 | 1 | 1 | 0 | 0 | 0 | 0 | 0 | 0 | 0 | 0 | 0 | 0 | 0 | 0 | 0 | 0 | 0 | 0 | 0 | 0 | 0 | 1 | 0 | 0 | 0 | 0 | 0 | 0 | 0 | 0 | 0 | 0 | 0 | 0 | 0 | 0 | 0 | 0 | 0 |
| Labib 2022 | 0 | 0 | 0 | 0 | 2 | 0 | 0 | 0 | 0 | 0 | 0 | 0 | 0 | 0 | 0 | 0 | 0 | 0 | 0 | 0 | 0 | 0 | 0 | 0 | 0 | 0 | 0 | 0 | 0 | 0 | 0 | 0 | 0 | 0 | 0 | 0 | 0 | 0 | 0 | 0 | 0 | 0 |
| Li 2016 | 0 | 0 | 0 | 1 | 1 | 0 | 0 | 0 | 0 | 0 | 0 | 0 | 0 | 0 | 0 | 0 | 0 | 0 | 0 | 0 | 0 | 0 | 0 | 0 | 0 | 0 | 0 | 0 | 0 | 0 | 0 | 0 | 0 | 0 | 0 | 0 | 0 | 0 | 0 | 0 | 0 | 0 |
| Liu 2012 | 0 | 0 | 1 | 1 | 1 | 0 | 0 | 0 | 0 | 1 | 0 | 0 | 0 | 0 | 0 | 0 | 0 | 0 | 0 | 0 | 0 | 0 | 1 | 1 | 0 | 0 | 0 | 0 | 0 | 0 | 0 | 0 | 0 | 0 | 0 | 0 | 0 | 0 | 0 | 0 | 0 | 0 |
| Loisa 2007 | 0 | 0 | 0 | 2 | 2 | 0 | 0 | 0 | 0 | 0 | 0 | 0 | 0 | 1 | 0 | 2 | 0 | 0 | 0 | 0 | 0 | 0 | 0 | 0 | 0 | 0 | 0 | 0 | 0 | 0 | 0 | 0 | 0 | 0 | 0 | 0 | 0 | 0 | 0 | 0 | 0 | 0 |
| Lucas 1984 | 0 | 0 | 0 | 0 | 0 | 0 | 0 | 0 | 0 | 0 | 2 | 0 | 0 | 0 | 0 | 0 | 0 | 0 | 0 | 0 | 0 | 0 | 0 | 0 | 0 | 0 | 0 | 0 | 0 | 0 | 0 | 0 | 0 | 0 | 0 | 0 | 0 | 0 | 0 | 0 | 0 | 0 |
| Luce 1988 | 0 | 0 | 1 | 1 | 1 | 0 | 0 | 0 | 0 | 1 | 0 | 0 | 0 | 0 | 1 | 1 | 2 | 1 | 0 | 0 | 0 | 0 | 1 | 1 | 0 | 0 | 0 | 0 | 2 | 0 | 0 | 0 | 2 | 0 | 0 | 0 | 0 | 0 | 0 | 0 | 0 | 1 |
| Lv 2017 | 1 | 2 | 0 | 1 | 1 | 0 | 0 | 0 | 0 | 1 | 0 | 0 | 0 | 0 | 1 | 1 | 1 | 1 | 0 | 1 | 0 | 1 | 1 | 1 | 1 | 0 | 1 | 0 | 1 | 0 | 1 | 0 | 1 | 1 | 0 | 0 | 0 | 0 | 0 | 0 | 0 | 1 |
| Lyu 2022 | 0 | 0 | 0 | 0 | 1 | 0 | 0 | 1 | 0 | 0 | 0 | 0 | 0 | 0 | 0 | 0 | 0 | 0 | 1 | 0 | 0 | 0 | 0 | 0 | 0 | 1 | 0 | 0 | 0 | 0 | 0 | 0 | 0 | 0 | 0 | 0 | 0 | 0 | 0 | 0 | 0 | 0 |
| Mansour 2012 | 0 | 0 | 0 | 0 | 0 | 0 | 0 | 0 | 1 | 0 | 0 | 0 | 0 | 0 | 0 | 0 | 0 | 0 | 0 | 0 | 0 | 0 | 0 | 0 | 0 | 0 | 0 | 0 | 0 | 0 | 0 | 0 | 0 | 0 | 0 | 0 | 0 | 0 | 1 | 0 | 0 | 0 |
| McHardy 1972 | 0 | 0 | 0 | 1 | 1 | 0 | 0 | 0 | 0 | 0 | 0 | 0 | 0 | 0 | 0 | 0 | 0 | 0 | 0 | 0 | 0 | 0 | 0 | 0 | 0 | 0 | 0 | 0 | 0 | 0 | 0 | 0 | 0 | 0 | 0 | 0 | 0 | 0 | 0 | 0 | 0 | 0 |
| Meduri 2007 | 0 | 0 | 1 | 1 | 1 | 0 | 0 | 0 | 0 | 1 | 0 | 0 | 0 | 0 | 0 | 1 | 1 | 0 | 0 | 0 | 0 | 0 | 1 | 1 | 0 | 0 | 0 | 0 | 0 | 0 | 0 | 0 | 0 | 0 | 0 | 0 | 0 | 0 | 0 | 0 | 0 | 0 |
| Meduri 2009 | 0 | 0 | 0 | 1 | 1 | 0 | 0 | 0 | 0 | 0 | 0 | 0 | 0 | 0 | 0 | 1 | 0 | 0 | 0 | 0 | 0 | 1 | 1 | 1 | 1 | 0 | 0 | 0 | 0 | 0 | 0 | 0 | 0 | 0 | 0 | 0 | 0 | 0 | 0 | 0 | 0 | 0 |
| Meduri 2022 | 0 | 0 | 0 | 0 | 1 | 0 | 0 | 0 | 0 | 0 | 0 | 0 | 0 | 0 | 0 | 0 | 0 | 0 | 0 | 0 | 0 | 0 | 0 | 0 | 0 | 0 | 0 | 0 | 0 | 0 | 0 | 0 | 0 | 0 | 0 | 0 | 0 | 0 | 0 | 0 | 0 | 0 |
| Meijvis 2011 | 0 | 0 | 1 | 1 | 1 | 0 | 0 | 0 | 0 | 1 | 0 | 0 | 0 | 0 | 0 | 1 | 1 | 0 | 0 | 0 | 0 | 0 | 1 | 1 | 0 | 0 | 0 | 0 | 0 | 0 | 0 | 0 | 0 | 0 | 0 | 0 | 0 | 0 | 0 | 0 | 0 | 0 |
| Menon 2017 | 0 | 0 | 0 | 1 | 1 | 0 | 0 | 0 | 1 | 0 | 0 | 0 | 0 | 0 | 0 | 0 | 0 | 0 | 0 | 0 | 0 | 0 | 1 | 1 | 0 | 0 | 0 | 0 | 0 | 0 | 2 | 0 | 0 | 0 | 0 | 0 | 0 | 0 | 1 | 0 | 0 | 0 |
| Min 1975 | 0 | 0 | 0 | 0 | 0 | 0 | 0 | 0 | 1 | 0 | 0 | 0 | 0 | 0 | 0 | 0 | 0 | 0 | 0 | 0 | 0 | 0 | 0 | 0 | 0 | 0 | 0 | 0 | 0 | 0 | 0 | 0 | 0 | 0 | 0 | 0 | 0 | 0 | 1 | 0 | 0 | 0 |
| Mirea 2014 | 0 | 0 | 0 | 1 | 1 | 0 | 0 | 0 | 0 | 0 | 0 | 0 | 0 | 0 | 0 | 2 | 2 | 0 | 0 | 0 | 0 | 0 | 2 | 2 | 1 | 0 | 0 | 0 | 0 | 0 | 0 | 0 | 0 | 0 | 0 | 0 | 0 | 0 | 0 | 0 | 0 | 0 |
| Mohamed 2020 | 0 | 0 | 0 | 0 | 1 | 1 | 0 | 1 | 0 | 0 | 0 | 2 | 0 | 0 | 0 | 0 | 0 | 0 | 1 | 0 | 1 | 0 | 0 | 0 | 0 | 2 | 0 | 1 | 0 | 0 | 0 | 0 | 0 | 0 | 2 | 0 | 0 | 0 | 0 | 1 | 0 | 0 |
| Mohamed 2023 | 0 | 0 | 0 | 0 | 1 | 0 | 0 | 1 | 0 | 0 | 0 | 0 | 0 | 0 | 0 | 0 | 0 | 0 | 0 | 0 | 0 | 0 | 0 | 0 | 0 | 0 | 0 | 0 | 0 | 0 | 0 | 0 | 0 | 0 | 0 | 0 | 0 | 0 | 0 | 0 | 0 | 0 |
| Moreno 2011 | 0 | 0 | 0 | 0 | 0 | 0 | 0 | 0 | 0 | 0 | 0 | 0 | 0 | 0 | 0 | 0 | 0 | 0 | 0 | 0 | 0 | 0 | 0 | 0 | 0 | 0 | 0 | 0 | 2 | 0 | 0 | 0 | 0 | 0 | 0 | 0 | 0 | 0 | 0 | 0 | 0 | 0 |
| Moskowitz 2020 | 0 | 0 | 0 | 0 | 1 | 1 | 0 | 1 | 0 | 0 | 0 | 2 | 2 | 0 | 0 | 0 | 0 | 0 | 1 | 0 | 1 | 0 | 0 | 0 | 0 | 1 | 0 | 1 | 0 | 0 | 0 | 0 | 0 | 0 | 1 | 0 | 0 | 0 | 0 | 1 | 1 | 0 |
| Mussack 2005 | 0 | 0 | 0 | 0 | 0 | 0 | 0 | 0 | 0 | 0 | 0 | 0 | 0 | 0 | 1 | 0 | 0 | 0 | 0 | 0 | 0 | 0 | 0 | 0 | 0 | 0 | 0 | 0 | 0 | 0 | 0 | 0 | 0 | 1 | 0 | 0 | 0 | 0 | 0 | 0 | 0 | 0 |
| Nafae 2013 | 0 | 0 | 0 | 1 | 1 | 0 | 0 | 0 | 0 | 0 | 0 | 0 | 0 | 0 | 0 | 2 | 0 | 0 | 0 | 0 | 0 | 0 | 0 | 0 | 0 | 0 | 0 | 0 | 0 | 0 | 0 | 0 | 0 | 0 | 0 | 0 | 0 | 0 | 0 | 0 | 0 | 0 |
| Nagy 2013 | 0 | 0 | 0 | 1 | 1 | 0 | 0 | 0 | 1 | 0 | 0 | 0 | 0 | 0 | 0 | 0 | 0 | 0 | 0 | 0 | 0 | 0 | 0 | 0 | 0 | 0 | 0 | 0 | 0 | 0 | 0 | 0 | 0 | 0 | 0 | 0 | 0 | 0 | 0 | 0 | 0 | 0 |
| Ngaosuwan 2018 | 0 | 0 | 0 | 2 | 2 | 0 | 0 | 0 | 0 | 0 | 0 | 0 | 0 | 0 | 0 | 0 | 0 | 0 | 0 | 0 | 0 | 0 | 0 | 0 | 0 | 0 | 0 | 0 | 0 | 0 | 0 | 0 | 0 | 0 | 0 | 0 | 0 | 0 | 0 | 0 | 0 | 0 |
| Oppert 2005 | 1 | 0 | 1 | 1 | 1 | 0 | 0 | 0 | 0 | 1 | 0 | 0 | 0 | 0 | 1 | 1 | 1 | 1 | 0 | 2 | 0 | 1 | 1 | 1 | 1 | 0 | 0 | 0 | 1 | 0 | 0 | 0 | 0 | 1 | 0 | 0 | 1 | 0 | 0 | 0 | 0 | 1 |
| Pierre de Oliveira 2 | 0 | 0 | 0 | 0 | 1 | 0 | 0 | 0 | 0 | 0 | 0 | 0 | 0 | 0 | 0 | 0 | 0 | 0 | 0 | 0 | 0 | 0 | 0 | 0 | 0 | 0 | 0 | 0 | 0 | 0 | 0 | 0 | 0 | 0 | 0 | 0 | 0 | 0 | 0 | 0 | 0 | 0 |
| Pongpanich 1973 | 0 | 0 | 0 | 0 | 0 | 0 | 0 | 0 | 1 | 0 | 0 | 0 | 0 | 0 | 0 | 0 | 0 | 0 | 0 | 0 | 0 | 0 | 0 | 0 | 0 | 0 | 0 | 0 | 0 | 0 | 0 | 0 | 0 | 0 | 0 | 0 | 0 | 0 | 1 | 0 | 0 | 0 |
| Povoa 2015 | 0 | 0 | 0 | 0 | 0 | 0 | 0 | 0 | 0 | 0 | 0 | 0 | 0 | 0 | 1 | 0 | 0 | 0 | 0 | 0 | 0 | 0 | 0 | 0 | 0 | 0 | 0 | 0 | 0 | 0 | 0 | 0 | 1 | 0 | 0 | 0 | 0 | 0 | 0 | 0 | 0 | 0 |
| Raghu 2021 | 0 | 0 | 0 | 0 | 0 | 0 | 0 | 1 | 0 | 0 | 0 | 0 | 0 | 0 | 0 | 0 | 0 | 0 | 0 | 0 | 0 | 0 | 0 | 0 | 0 | 0 | 0 | 0 | 0 | 0 | 0 | 0 | 0 | 0 | 0 | 0 | 0 | 0 | 0 | 0 | 0 | 0 |
| Ram 2022 | 0 | 0 | 0 | 0 | 2 | 0 | 0 | 0 | 0 | 0 | 0 | 0 | 0 | 1 | 0 | 0 | 0 | 0 | 0 | 0 | 0 | 0 | 0 | 0 | 0 | 0 | 0 | 0 | 0 | 0 | 0 | 0 | 0 | 0 | 0 | 0 | 0 | 0 | 0 | 0 | 0 | 0 |
| Reddy 2020 | 0 | 0 | 0 | 0 | 0 | 1 | 0 | 0 | 0 | 0 | 0 | 0 | 0 | 0 | 0 | 0 | 0 | 0 | 0 | 0 | 2 | 0 | 0 | 0 | 0 | 2 | 0 | 0 | 0 | 0 | 0 | 2 | 0 | 0 | 0 | 0 | 0 | 0 | 0 | 0 | 0 | 0 |
| Rezk 2013 | 0 | 0 | 1 | 1 | 1 | 0 | 0 | 0 | 0 | 1 | 0 | 0 | 0 | 0 | 0 | 1 | 2 | 0 | 0 | 0 | 0 | 0 | 1 | 1 | 0 | 0 | 0 | 0 | 0 | 0 | 0 | 0 | 0 | 0 | 0 | 0 | 0 | 0 | 0 | 0 | 0 | 0 |
| Rinaldi 2006 | 2 | 0 | 1 | 1 | 1 | 0 | 0 | 0 | 0 | 1 | 0 | 0 | 0 | 0 | 0 | 1 | 1 | 0 | 0 | 0 | 0 | 1 | 1 | 1 | 0 | 0 | 0 | 0 | 0 | 0 | 0 | 0 | 0 | 0 | 0 | 0 | 0 | 0 | 0 | 0 | 0 | 0 |
| Sabry 2011 | 0 | 0 | 1 | 1 | 1 | 0 | 0 | 0 | 0 | 1 | 0 | 0 | 0 | 0 | 0 | 1 | 2 | 0 | 0 | 0 | 0 | 0 | 1 | 1 | 0 | 0 | 0 | 0 | 0 | 0 | 0 | 0 | 0 | 0 | 0 | 0 | 0 | 0 | 0 | 0 | 0 | 0 |
| Sai 2021 | 0 | 0 | 0 | 0 | 1 | 0 | 0 | 0 | 0 | 0 | 0 | 0 | 0 | 0 | 0 | 0 | 0 | 0 | 0 | 0 | 0 | 0 | 0 | 0 | 0 | 0 | 0 | 0 | 0 | 0 | 0 | 0 | 0 | 0 | 0 | 0 | 0 | 0 | 0 | 0 | 0 | 0 |
| Sang 2020 | 0 | 0 | 0 | 0 | 0 | 0 | 0 | 1 | 0 | 0 | 0 | 0 | 0 | 0 | 0 | 0 | 0 | 0 | 0 | 0 | 0 | 0 | 0 | 0 | 0 | 0 | 0 | 0 | 0 | 0 | 0 | 0 | 0 | 0 | 0 | 0 | 0 | 0 | 0 | 0 | 0 | 0 |
| Schelling 2001 | 0 | 0 | 0 | 0 | 0 | 0 | 0 | 0 | 0 | 0 | 0 | 0 | 0 | 0 | 0 | 0 | 0 | 0 | 0 | 0 | 0 | 0 | 0 | 0 | 0 | 0 | 0 | 0 | 2 | 0 | 0 | 0 | 0 | 0 | 0 | 0 | 0 | 0 | 0 | 0 | 0 | 0 |
| Schumer 1976 | 0 | 0 | 1 | 1 | 1 | 0 | 0 | 0 | 0 | 1 | 2 | 0 | 0 | 0 | 0 | 1 | 2 | 0 | 0 | 0 | 0 | 0 | 1 | 1 | 0 | 0 | 0 | 0 | 1 | 0 | 0 | 0 | 1 | 0 | 0 | 0 | 0 | 0 | 0 | 0 | 0 | 1 |
| Sevransky 2021 | 0 | 0 | 0 | 0 | 1 | 1 | 0 | 1 | 0 | 0 | 0 | 2 | 2 | 0 | 0 | 0 | 0 | 0 | 1 | 0 | 2 | 0 | 0 | 0 | 0 | 1 | 0 | 1 | 0 | 0 | 0 | 0 | 0 | 0 | 0 | 0 | 0 | 0 | 0 | 1 | 0 | 0 |
| Shi 2019 | 0 | 0 | 0 | 0 | 0 | 0 | 0 | 0 | 1 | 0 | 0 | 0 | 0 | 0 | 0 | 0 | 0 | 0 | 0 | 0 | 0 | 0 | 0 | 0 | 0 | 0 | 0 | 0 | 0 | 0 | 0 | 0 | 0 | 0 | 0 | 0 | 0 | 0 | 0 | 0 | 0 | 0 |
| Slusher 1996 | 0 | 0 | 2 | 1 | 1 | 0 | 0 | 0 | 1 | 0 | 0 | 0 | 0 | 0 | 0 | 0 | 0 | 0 | 0 | 0 | 0 | 0 | 2 | 2 | 0 | 0 | 0 | 0 | 0 | 0 | 0 | 0 | 0 | 0 | 0 | 0 | 0 | 0 | 1 | 0 | 0 | 0 |
| Snijders 2010 | 0 | 0 | 1 | 1 | 1 | 0 | 0 | 0 | 0 | 1 | 0 | 0 | 0 | 0 | 0 | 1 | 1 | 0 | 0 | 0 | 0 | 0 | 1 | 1 | 0 | 0 | 0 | 0 | 0 | 0 | 0 | 0 | 0 | 0 | 0 | 0 | 0 | 0 | 0 | 0 | 0 | 0 |
| Sprung 1984 | 0 | 0 | 1 | 1 | 1 | 0 | 0 | 0 | 0 | 1 | 0 | 0 | 0 | 0 | 1 | 1 | 2 | 0 | 0 | 0 | 0 | 0 | 1 | 1 | 0 | 0 | 0 | 0 | 0 | 0 | 0 | 0 | 2 | 0 | 0 | 0 | 0 | 0 | 0 | 0 | 0 | 1 |
| Sprung 2008 | 1 | 0 | 1 | 1 | 1 | 0 | 0 | 0 | 0 | 1 | 2 | 0 | 0 | 0 | 1 | 1 | 1 | 1 | 0 | 1 | 0 | 1 | 1 | 1 | 1 | 0 | 1 | 0 | 1 | 0 | 0 | 0 | 1 | 1 | 0 | 0 | 1 | 0 | 0 | 0 | 0 | 1 |
| Sui 2013 | 0 | 0 | 0 | 2 | 2 | 0 | 0 | 0 | 0 | 0 | 0 | 0 | 0 | 0 | 0 | 0 | 0 | 0 | 0 | 0 | 0 | 0 | 0 | 0 | 0 | 0 | 0 | 0 | 0 | 0 | 0 | 0 | 0 | 0 | 0 | 0 | 0 | 0 | 0 | 0 | 0 | 0 |
| Sumarmo 1982 | 0 | 0 | 0 | 0 | 0 | 0 | 0 | 0 | 1 | 0 | 0 | 0 | 0 | 0 | 0 | 0 | 0 | 0 | 0 | 0 | 0 | 0 | 0 | 0 | 0 | 0 | 0 | 0 | 0 | 0 | 0 | 0 | 0 | 0 | 0 | 0 | 0 | 0 | 1 | 0 | 0 | 0 |
| Tagaro 2017 | 0 | 0 | 0 | 1 | 1 | 0 | 0 | 0 | 0 | 0 | 0 | 0 | 0 | 0 | 0 | 0 | 0 | 0 | 0 | 0 | 0 | 0 | 0 | 0 | 0 | 0 | 0 | 0 | 0 | 0 | 0 | 0 | 0 | 0 | 0 | 0 | 0 | 0 | 0 | 0 | 0 | 0 |
| Tandan 2005 | 0 | 0 | 1 | 1 | 1 | 0 | 0 | 0 | 0 | 1 | 0 | 0 | 0 | 0 | 0 | 1 | 1 | 0 | 0 | 0 | 0 | 0 | 1 | 1 | 1 | 0 | 0 | 0 | 0 | 0 | 0 | 0 | 0 | 0 | 0 | 0 | 0 | 0 | 0 | 0 | 0 | 0 |
| Tassniyom 1993 | 0 | 0 | 0 | 0 | 0 | 0 | 0 | 0 | 1 | 0 | 0 | 0 | 0 | 0 | 0 | 0 | 0 | 0 | 0 | 0 | 0 | 0 | 0 | 0 | 0 | 0 | 0 | 0 | 0 | 0 | 0 | 0 | 0 | 0 | 0 | 0 | 0 | 0 | 1 | 0 | 0 | 0 |
| Tilouche 2019 | 0 | 0 | 0 | 2 | 2 | 0 | 0 | 0 | 0 | 0 | 0 | 0 | 0 | 1 | 0 | 2 | 0 | 0 | 0 | 0 | 0 | 0 | 0 | 0 | 0 | 0 | 0 | 0 | 0 | 0 | 0 | 0 | 0 | 0 | 0 | 0 | 0 | 0 | 0 | 0 | 0 | 0 |
| Tomazini 2020 | 0 | 0 | 0 | 0 | 1 | 0 | 0 | 0 | 0 | 0 | 0 | 0 | 0 | 0 | 0 | 1 | 0 | 0 | 0 | 0 | 0 | 0 | 0 | 0 | 0 | 0 | 0 | 0 | 0 | 0 | 0 | 0 | 0 | 0 | 0 | 0 | 0 | 0 | 0 | 0 | 0 | 0 |
| Tongyoo 2016 | 0 | 2 | 0 | 1 | 1 | 0 | 0 | 0 | 0 | 1 | 0 | 0 | 0 | 0 | 1 | 1 | 1 | 0 | 0 | 1 | 0 | 1 | 1 | 1 | 1 | 0 | 0 | 0 | 1 | 0 | 0 | 0 | 0 | 1 | 0 | 0 | 0 | 0 | 0 | 0 | 0 | 0 |
| Torres 2015 | 0 | 0 | 1 | 1 | 1 | 0 | 0 | 0 | 0 | 1 | 0 | 0 | 0 | 0 | 0 | 1 | 2 | 0 | 0 | 0 | 0 | 0 | 1 | 1 | 1 | 0 | 0 | 0 | 0 | 0 | 2 | 0 | 0 | 0 | 0 | 0 | 0 | 0 | 0 | 0 | 0 | 0 |
| Valoor 2009 | 0 | 0 | 2 | 1 | 1 | 0 | 0 | 0 | 1 | 0 | 0 | 0 | 0 | 0 | 0 | 0 | 0 | 0 | 0 | 0 | 0 | 0 | 2 | 2 | 0 | 0 | 0 | 0 | 0 | 0 | 0 | 0 | 0 | 0 | 0 | 0 | 0 | 0 | 1 | 0 | 0 | 0 |
| VASSCSG 1987 | 0 | 0 | 1 | 1 | 1 | 0 | 0 | 0 | 0 | 2 | 0 | 0 | 0 | 0 | 0 | 1 | 2 | 0 | 0 | 0 | 0 | 0 | 1 | 1 | 0 | 0 | 0 | 0 | 1 | 0 | 0 | 0 | 0 | 0 | 0 | 0 | 0 | 0 | 0 | 0 | 0 | 0 |
| Venkatesh 2018 | 1 | 2 | 0 | 1 | 1 | 0 | 0 | 0 | 0 | 1 | 0 | 0 | 0 | 0 | 1 | 1 | 1 | 0 | 0 | 1 | 0 | 1 | 1 | 1 | 1 | 0 | 1 | 0 | 1 | 0 | 1 | 0 | 1 | 1 | 0 | 0 | 0 | 0 | 0 | 0 | 0 | 1 |
| Villar 2020 | 0 | 0 | 0 | 0 | 1 | 0 | 0 | 0 | 0 | 0 | 0 | 0 | 0 | 0 | 0 | 0 | 0 | 0 | 0 | 0 | 0 | 0 | 0 | 0 | 0 | 0 | 0 | 0 | 0 | 0 | 0 | 0 | 0 | 0 | 0 | 0 | 0 | 0 | 0 | 0 | 0 | 0 |
| Wan 2014 | 0 | 0 | 0 | 0 | 0 | 0 | 0 | 0 | 0 | 0 | 0 | 0 | 0 | 0 | 0 | 0 | 0 | 1 | 0 | 0 | 0 | 0 | 0 | 0 | 0 | 0 | 0 | 0 | 0 | 0 | 0 | 0 | 0 | 0 | 0 | 0 | 1 | 0 | 0 | 0 | 0 | 0 |
| Wang 2022 | 0 | 0 | 0 | 0 | 0 | 0 | 0 | 0 | 2 | 0 | 0 | 0 | 0 | 0 | 0 | 0 | 0 | 0 | 0 | 0 | 0 | 0 | 0 | 0 | 0 | 0 | 0 | 0 | 0 | 0 | 0 | 0 | 0 | 0 | 0 | 0 | 0 | 0 | 0 | 0 | 0 | 0 |
| Wang 2023 | 0 | 0 | 0 | 0 | 0 | 0 | 0 | 2 | 0 | 0 | 0 | 0 | 0 | 0 | 0 | 0 | 0 | 0 | 0 | 0 | 0 | 0 | 0 | 0 | 0 | 0 | 0 | 0 | 0 | 0 | 0 | 0 | 0 | 0 | 0 | 0 | 0 | 0 | 0 | 0 | 0 | 0 |
| Wani 2020 | 0 | 0 | 0 | 0 | 1 | 1 | 0 | 1 | 0 | 0 | 0 | 2 | 0 | 0 | 0 | 0 | 0 | 0 | 1 | 0 | 1 | 0 | 0 | 0 | 0 | 1 | 0 | 1 | 0 | 0 | 0 | 0 | 0 | 0 | 1 | 0 | 0 | 0 | 0 | 1 | 1 | 0 |
| Wen 2004 | 0 | 0 | 0 | 0 | 0 | 0 | 0 | 0 | 2 | 0 | 0 | 0 | 0 | 0 | 0 | 0 | 0 | 0 | 0 | 0 | 0 | 0 | 0 | 0 | 0 | 0 | 0 | 0 | 0 | 0 | 0 | 0 | 0 | 0 | 0 | 0 | 0 | 0 | 0 | 0 | 0 | 0 |
| Wittermans 2020 | 0 | 0 | 0 | 0 | 1 | 0 | 0 | 0 | 0 | 0 | 0 | 0 | 0 | 0 | 0 | 0 | 0 | 0 | 0 | 0 | 0 | 0 | 0 | 0 | 0 | 0 | 0 | 0 | 0 | 0 | 0 | 0 | 0 | 0 | 0 | 0 | 0 | 0 | 0 | 0 | 0 | 0 |
| Yildiz 2002 | 0 | 0 | 1 | 1 | 1 | 0 | 0 | 0 | 0 | 1 | 0 | 0 | 0 | 0 | 0 | 1 | 1 | 0 | 0 | 0 | 0 | 1 | 1 | 1 | 0 | 0 | 0 | 0 | 1 | 0 | 1 | 0 | 0 | 0 | 0 | 0 | 0 | 0 | 0 | 0 | 0 | 0 |
| Yildiz 2011 | 0 | 0 | 1 | 1 | 1 | 0 | 0 | 0 | 0 | 1 | 0 | 0 | 0 | 0 | 0 | 1 | 1 | 0 | 0 | 0 | 0 | 1 | 1 | 1 | 0 | 0 | 0 | 0 | 1 | 0 | 0 | 0 | 0 | 0 | 0 | 0 | 0 | 0 | 0 | 0 | 0 | 0 |
| Yuan 2010 | 0 | 0 | 0 | 0 | 0 | 0 | 0 | 0 | 2 | 0 | 0 | 0 | 0 | 0 | 0 | 0 | 0 | 0 | 0 | 0 | 0 | 0 | 0 | 0 | 0 | 0 | 0 | 0 | 0 | 0 | 0 | 0 | 0 | 0 | 0 | 0 | 0 | 0 | 0 | 0 | 0 | 0 |
| Zeng 2014 | 0 | 0 | 0 | 0 | 0 | 0 | 0 | 0 | 1 | 0 | 0 | 0 | 0 | 0 | 0 | 0 | 0 | 0 | 0 | 0 | 0 | 0 | 0 | 0 | 0 | 0 | 0 | 0 | 0 | 0 | 0 | 0 | 0 | 0 | 0 | 0 | 0 | 0 | 0 | 0 | 0 | 0 |
| Zhou 2015 | 0 | 0 | 0 | 2 | 2 | 0 | 0 | 0 | 0 | 0 | 0 | 0 | 0 | 0 | 0 | 0 | 0 | 0 | 0 | 0 | 0 | 0 | 0 | 0 | 0 | 0 | 0 | 0 | 0 | 0 | 0 | 0 | 0 | 0 | 0 | 0 | 0 | 0 | 0 | 0 | 0 | 0 |

### Supplementary Table S20: Pairwise comparison of systematic reviews addressing non-specific broad corticosteroid strategies in sepsis including Jaccard similarity and classification of conclusions (n = 22)

#### Comparison of median Jaccard between review pairs with concordant and discordant conclusions:

| Same conclusion | N SR pairs | Median Jaccard (J) | IQR_low | IQR_high |
|---|---|---|---|---|
| no | 120 | 0,333333333 | 0,2 | 0,454545455 |
| yes | 111 | 0,324324324 | 0,217483108 | 0,451027397 |

#### Summary of RCT overlap and contribution patterns across SRs:

| Overlap group (J) | Same conclusion | n | percent |
|---|---|---|---|
| high (>=0.80) | no | 1 | 100 |
| low (0.20-0.49) | no | 71 | 53 |
| low (0.20-0.49) | yes | 63 | 47 |
| moderate (0.50-0.79) | no | 19 | 45,2 |
| moderate (0.50-0.79) | yes | 23 | 54,8 |
| very low (<0.20) | no | 29 | 53,7 |
| very low (<0.20) | yes | 25 | 46,3 |

#### Pairwise comparison of SRs according to concordant and discordant conclusions and Jaccard similarity:

| SR1 | SR2 | Effect1 | Effect2 | Same conclusion | Jaccard (J) | Shared trials | Union trials | Overlap group (J) |
|---|---|---|---|---|---|---|---|---|
| Pitre 2024 | Rochwerg 2018 | benefit | no evidence of effect | no | 0,923076923 | 36 | 39 | high (>=0.80) |
| Fang 2019 | Rochwerg 2018 | benefit | no evidence of effect | no | 0,794871795 | 31 | 39 | moderate (0.50-0.79) |

| | | | | | | | | |
|---|---|---|---|---|---|---|---|---|
| Fang 2019 | Liang 2021 | benefit | no evidence of effect | no | 0,761904762 | 32 | 42 | moderate (0.50-0.79) |
| Annane 2015 | Rochwerg 2018 | no evidence of effect | no evidence of effect | yes | 0,75 | 27 | 36 | moderate (0.50-0.79) |
| Fang 2019 | Pitre 2024 | benefit | benefit | yes | 0,738095238 | 31 | 42 | moderate (0.50-0.79) |
| Liang 2021 | Rochwerg 2018 | no evidence of effect | no evidence of effect | yes | 0,727272727 | 32 | 44 | moderate (0.50-0.79) |
| Liang 2021 | Pitre 2024 | no evidence of effect | benefit | no | 0,717391304 | 33 | 46 | moderate (0.50-0.79) |
| Lin 2019 | Ni 2019 | benefit | benefit | yes | 0,695652174 | 16 | 23 | moderate (0.50-0.79) |
| Annane 2015 | Fang 2019 | no evidence of effect | benefit | no | 0,694444444 | 25 | 36 | moderate (0.50-0.79) |
| Annane 2019 | Annane 2025 | benefit | benefit | yes | 0,694444444 | 50 | 72 | moderate (0.50-0.79) |
| Annane 2015 | Pitre 2024 | no evidence of effect | benefit | no | 0,692307692 | 27 | 39 | moderate (0.50-0.79) |
| Annane 2019 | Rochwerg 2018 | benefit | no evidence of effect | no | 0,68627451 | 35 | 51 | moderate (0.50-0.79) |
| Annane 2019 | Pitre 2024 | benefit | benefit | yes | 0,679245283 | 36 | 53 | moderate (0.50-0.79) |
| Annane 2019 | Liang 2021 | benefit | no evidence of effect | no | 0,666666667 | 36 | 54 | moderate (0.50-0.79) |
| Lian 2019 | Zhou 2018 | benefit | no evidence of effect | no | 0,652173913 | 15 | 23 | moderate (0.50-0.79) |
| Annane 2019 | Fang 2019 | benefit | benefit | yes | 0,647058824 | 33 | 51 | moderate (0.50-0.79) |
| Fang 2019 | Lin 2019 | benefit | benefit | yes | 0,617647059 | 21 | 34 | moderate (0.50-0.79) |
| Lyu 2020 | Wu 2020 | no evidence of effect | no evidence of effect | yes | 0,615384615 | 8 | 13 | moderate (0.50-0.79) |

| | | | | | | | | |
|---|---|---|---|---|---|---|---|---|
| Annane 2015 | Liang 2021 | no evidence of effect | no evidence of effect | yes | 0,595238095 | 25 | 42 | moderate (0.50-0.79) |
| Ni 2019 | Son 2021 | benefit | benefit | yes | 0,590909091 | 13 | 22 | moderate (0.50-0.79) |
| Lin 2019 | Rochwerg 2018 | benefit | no evidence of effect | no | 0,583333333 | 21 | 36 | moderate (0.50-0.79) |
| Rygard 2018 | Zhou 2018 | no evidence of effect | no evidence of effect | yes | 0,56 | 14 | 25 | moderate (0.50-0.79) |
| Aletreby 2019 | Siddiqi 2019 | no evidence of effect | benefit | no | 0,555555556 | 5 | 9 | moderate (0.50-0.79) |
| Lyu 2020 | Zhou 2018 | no evidence of effect | no evidence of effect | yes | 0,555555556 | 10 | 18 | moderate (0.50-0.79) |
| Annane 2015 | Annane 2019 | no evidence of effect | benefit | no | 0,54 | 27 | 50 | moderate (0.50-0.79) |
| Aletreby 2019 | Lyu 2020 | no evidence of effect | no evidence of effect | yes | 0,538461538 | 7 | 13 | moderate (0.50-0.79) |
| Lin 2019 | Pitre 2024 | benefit | benefit | yes | 0,538461538 | 21 | 39 | moderate (0.50-0.79) |
| Lian 2019 | Rygard 2018 | benefit | no evidence of effect | no | 0,535714286 | 15 | 28 | moderate (0.50-0.79) |
| Lyu 2020 | Wen 2019 | no evidence of effect | no evidence of effect | yes | 0,533333333 | 8 | 15 | moderate (0.50-0.79) |
| Lyu 2020 | Ni 2019 | no evidence of effect | benefit | no | 0,526315789 | 10 | 19 | moderate (0.50-0.79) |
| Wen 2019 | Zhou 2018 | no evidence of effect | no evidence of effect | yes | 0,526315789 | 10 | 19 | moderate (0.50-0.79) |
| Lian 2019 | Lyu 2020 | benefit | no evidence of effect | no | 0,523809524 | 11 | 21 | moderate (0.50-0.79) |

| | | | | | | | | |
|---|---|---|---|---|---|---|---|---|
| Annane 2025 | Pitre 2024 | benefit | benefit | yes | 0,520547945 | 38 | 73 | moderate (0.50-0.79) |
| Lin 2019 | Son 2021 | benefit | benefit | yes | 0,52 | 13 | 25 | moderate (0.50-0.79) |
| Aletreby 2019 | Wen 2019 | no evidence of effect | no evidence of effect | yes | 0,5 | 7 | 14 | moderate (0.50-0.79) |
| Annane 2015 | Lin 2019 | no evidence of effect | benefit | no | 0,5 | 16 | 32 | moderate (0.50-0.79) |
| Fang 2019 | Zhou 2018 | benefit | no evidence of effect | no | 0,5 | 17 | 34 | moderate (0.50-0.79) |
| Lu 2021 | Xu 2018 | no evidence of effect | no evidence of effect | yes | 0,5 | 6 | 12 | moderate (0.50-0.79) |
| Lyu 2020 | Rygard 2018 | no evidence of effect | no evidence of effect | yes | 0,5 | 11 | 22 | moderate (0.50-0.79) |
| Lyu 2020 | Song Y 2024 | no evidence of effect | benefit | no | 0,5 | 6 | 12 | moderate (0.50-0.79) |
| Ni 2019 | Rochwerg 2018 | benefit | no evidence of effect | no | 0,5 | 18 | 36 | moderate (0.50-0.79) |
| Ni 2019 | Wen 2019 | benefit | no evidence of effect | no | 0,5 | 10 | 20 | moderate (0.50-0.79) |
| Siddiqi 2019 | Song Y 2024 | benefit | benefit | yes | 0,5 | 4 | 8 | moderate (0.50-0.79) |
| Annane 2025 | Liang 2021 | benefit | no evidence of effect | no | 0,493333333 | 37 | 75 | low (0.20-0.49) |
| Liang 2021 | Lin 2019 | no evidence of effect | benefit | no | 0,487804878 | 20 | 41 | low (0.20-0.49) |
| Fang 2019 | Ni 2019 | benefit | benefit | yes | 0,485714286 | 17 | 35 | low (0.20-0.49) |
| Ni 2019 | Rygard 2018 | benefit | no evidence of effect | no | 0,481481481 | 13 | 27 | low (0.20-0.49) |

| | | | | | | | | |
|---|---|---|---|---|---|---|---|---|
| Annane 2025 | Rochwerg 2018 | benefit | no evidence of effect | no | 0,479452055 | 35 | 73 | low (0.20-0.49) |
| Lian 2019 | Wu 2020 | benefit | no evidence of effect | no | 0,476190476 | 10 | 21 | low (0.20-0.49) |
| Fang 2019 | Rygard 2018 | benefit | no evidence of effect | no | 0,473684211 | 18 | 38 | low (0.20-0.49) |
| Lian 2019 | Pitre 2024 | benefit | benefit | yes | 0,463414634 | 19 | 41 | low (0.20-0.49) |
| Aletreby 2019 | Wu 2020 | no evidence of effect | no evidence of effect | yes | 0,461538462 | 6 | 13 | low (0.20-0.49) |
| Lian 2019 | Rochwerg 2018 | benefit | no evidence of effect | no | 0,461538462 | 18 | 39 | low (0.20-0.49) |
| Ni 2019 | Pitre 2024 | benefit | benefit | yes | 0,461538462 | 18 | 39 | low (0.20-0.49) |
| Song Y 2024 | Wen 2019 | benefit | no evidence of effect | no | 0,461538462 | 6 | 13 | low (0.20-0.49) |
| Ni 2019 | Zhou 2018 | benefit | no evidence of effect | no | 0,458333333 | 11 | 24 | low (0.20-0.49) |
| Aletreby 2019 | Song Y 2024 | no evidence of effect | benefit | no | 0,454545455 | 5 | 11 | low (0.20-0.49) |
| Lyu 2020 | Siddiqi 2019 | no evidence of effect | benefit | no | 0,454545455 | 5 | 11 | low (0.20-0.49) |
| Annane 2025 | Fang 2019 | benefit | benefit | yes | 0,452054795 | 33 | 73 | low (0.20-0.49) |
| Rochwerg 2018 | Rygard 2018 | no evidence of effect | no evidence of effect | yes | 0,45 | 18 | 40 | low (0.20-0.49) |
| Son 2021 | Wen 2019 | benefit | no evidence of effect | no | 0,45 | 9 | 20 | low (0.20-0.49) |
| Fang 2019 | Lian 2019 | benefit | benefit | yes | 0,447368421 | 17 | 38 | low (0.20-0.49) |

| | | | | | | | | |
|---|---|---|---|---|---|---|---|---|
| Aletreby 2019 | Zhou 2018 | no evidence of effect | no evidence of effect | yes | 0,444444444 | 8 | 18 | low (0.20-0.49) |
| Lian 2019 | Ni 2019 | benefit | benefit | yes | 0,444444444 | 12 | 27 | low (0.20-0.49) |
| Xu 2018 | Zhou 2018 | no evidence of effect | no evidence of effect | yes | 0,444444444 | 8 | 18 | low (0.20-0.49) |
| Lian 2019 | Wen 2019 | benefit | no evidence of effect | no | 0,434782609 | 10 | 23 | low (0.20-0.49) |
| Lin 2019 | Rygard 2018 | benefit | no evidence of effect | no | 0,433333333 | 13 | 30 | low (0.20-0.49) |
| Rochwerg 2018 | Son 2021 | no evidence of effect | benefit | no | 0,432432432 | 16 | 37 | low (0.20-0.49) |
| Rochwerg 2018 | Zhou 2018 | no evidence of effect | no evidence of effect | yes | 0,432432432 | 16 | 37 | low (0.20-0.49) |
| Lyu 2020 | Xu 2018 | no evidence of effect | no evidence of effect | yes | 0,428571429 | 6 | 14 | low (0.20-0.49) |
| Aletreby 2019 | Ni 2019 | no evidence of effect | benefit | no | 0,421052632 | 8 | 19 | low (0.20-0.49) |
| Ni 2019 | Xu 2018 | benefit | no evidence of effect | no | 0,421052632 | 8 | 19 | low (0.20-0.49) |
| Wu 2020 | Zhou 2018 | no evidence of effect | no evidence of effect | yes | 0,421052632 | 8 | 19 | low (0.20-0.49) |
| Annane 2019 | Lin 2019 | benefit | benefit | yes | 0,42 | 21 | 50 | low (0.20-0.49) |
| Lian 2019 | Liang 2021 | benefit | no evidence of effect | no | 0,418604651 | 18 | 43 | low (0.20-0.49) |
| Pitre 2024 | Rygard 2018 | benefit | no evidence of effect | no | 0,418604651 | 18 | 43 | low (0.20-0.49) |

| | | | | | | | | |
|---|---|---|---|---|---|---|---|---|
| Siddiqi 2019 | Wen 2019 | benefit | no evidence of effect | no | 0,416666667 | 5 | 12 | low (0.20-0.49) |
| Son 2021 | Zhou 2018 | benefit | no evidence of effect | no | 0,416666667 | 10 | 24 | low (0.20-0.49) |
| Liang 2021 | Ni 2019 | no evidence of effect | benefit | no | 0,414634146 | 17 | 41 | low (0.20-0.49) |
| Annane 2019 | Rygard 2018 | benefit | no evidence of effect | no | 0,411764706 | 21 | 51 | low (0.20-0.49) |
| Liang 2021 | Rygard 2018 | no evidence of effect | no evidence of effect | yes | 0,409090909 | 18 | 44 | low (0.20-0.49) |
| Lian 2019 | Son 2021 | benefit | benefit | yes | 0,407407407 | 11 | 27 | low (0.20-0.49) |
| Lian 2019 | Lin 2019 | benefit | benefit | yes | 0,4 | 12 | 30 | low (0.20-0.49) |
| Lyu 2020 | Son 2021 | no evidence of effect | benefit | no | 0,4 | 8 | 20 | low (0.20-0.49) |
| Ni 2019 | Wu 2020 | benefit | no evidence of effect | no | 0,4 | 8 | 20 | low (0.20-0.49) |
| Pitre 2024 | Son 2021 | benefit | benefit | yes | 0,4 | 16 | 40 | low (0.20-0.49) |
| Pitre 2024 | Zhou 2018 | benefit | no evidence of effect | no | 0,4 | 16 | 40 | low (0.20-0.49) |
| Wen 2019 | Xu 2018 | no evidence of effect | no evidence of effect | yes | 0,4 | 6 | 15 | low (0.20-0.49) |
| Lin 2019 | Lyu 2020 | benefit | no evidence of effect | no | 0,391304348 | 9 | 23 | low (0.20-0.49) |
| Rygard 2018 | Wu 2020 | no evidence of effect | no evidence of effect | yes | 0,391304348 | 9 | 23 | low (0.20-0.49) |

| | | | | | | | | |
|---|---|---|---|---|---|---|---|---|
| Liang 2021 | Zhou 2018 | no evidence of effect | no evidence of effect | yes | 0,390243902 | 16 | 41 | low (0.20-0.49) |
| Ni 2019 | Song Y 2024 | benefit | benefit | yes | 0,388888889 | 7 | 18 | low (0.20-0.49) |
| Fang 2019 | Son 2021 | benefit | benefit | yes | 0,378378378 | 14 | 37 | low (0.20-0.49) |
| Annane 2015 | Annane 2025 | no evidence of effect | benefit | no | 0,375 | 27 | 72 | low (0.20-0.49) |
| Annane 2015 | Zhou 2018 | no evidence of effect | no evidence of effect | yes | 0,375 | 12 | 32 | low (0.20-0.49) |
| Lin 2019 | Wen 2019 | benefit | no evidence of effect | no | 0,375 | 9 | 24 | low (0.20-0.49) |
| Wen 2019 | Wu 2020 | no evidence of effect | no evidence of effect | yes | 0,375 | 6 | 16 | low (0.20-0.49) |
| Annane 2015 | Lian 2019 | no evidence of effect | benefit | no | 0,371428571 | 13 | 35 | low (0.20-0.49) |
| Aletreby 2019 | Son 2021 | no evidence of effect | benefit | no | 0,368421053 | 7 | 19 | low (0.20-0.49) |
| Lu 2021 | Zhou 2018 | no evidence of effect | no evidence of effect | yes | 0,368421053 | 7 | 19 | low (0.20-0.49) |
| Annane 2019 | Lian 2019 | benefit | benefit | yes | 0,365384615 | 19 | 52 | low (0.20-0.49) |
| Aletreby 2019 | Lian 2019 | no evidence of effect | benefit | no | 0,363636364 | 8 | 22 | low (0.20-0.49) |
| Annane 2015 | Ni 2019 | no evidence of effect | benefit | no | 0,363636364 | 12 | 33 | low (0.20-0.49) |
| Lian 2019 | Lu 2021 | benefit | no evidence of effect | no | 0,363636364 | 8 | 22 | low (0.20-0.49) |

| | | | | | | | | |
|---|---|---|---|---|---|---|---|---|
| Lian 2019 | Xu 2018 | benefit | no evidence of effect | no | 0,363636364 | 8 | 22 | low (0.20-0.49) |
| Annane 2019 | Ni 2019 | benefit | benefit | yes | 0,36 | 18 | 50 | low (0.20-0.49) |
| Rygard 2018 | Wen 2019 | no evidence of effect | no evidence of effect | yes | 0,36 | 9 | 25 | low (0.20-0.49) |
| Liang 2021 | Son 2021 | no evidence of effect | benefit | no | 0,357142857 | 15 | 42 | low (0.20-0.49) |
| Lin 2019 | Zhou 2018 | benefit | no evidence of effect | no | 0,357142857 | 10 | 28 | low (0.20-0.49) |
| Wu 2020 | Xu 2018 | no evidence of effect | no evidence of effect | yes | 0,357142857 | 5 | 14 | low (0.20-0.49) |
| Aletreby 2019 | Rygard 2018 | no evidence of effect | no evidence of effect | yes | 0,347826087 | 8 | 23 | low (0.20-0.49) |
| Rygard 2018 | Xu 2018 | no evidence of effect | no evidence of effect | yes | 0,347826087 | 8 | 23 | low (0.20-0.49) |
| Rygard 2018 | Son 2021 | no evidence of effect | benefit | no | 0,344827586 | 10 | 29 | low (0.20-0.49) |
| Annane 2019 | Zhou 2018 | benefit | no evidence of effect | no | 0,34 | 17 | 50 | low (0.20-0.49) |
| Annane 2015 | Son 2021 | no evidence of effect | benefit | no | 0,333333333 | 11 | 33 | low (0.20-0.49) |
| Song Y 2024 | Zhou 2018 | benefit | no evidence of effect | no | 0,333333333 | 6 | 18 | low (0.20-0.49) |
| Annane 2015 | Rygard 2018 | no evidence of effect | no evidence of effect | yes | 0,324324324 | 12 | 37 | low (0.20-0.49) |

| | | | | | | | | |
|---|---|---|---|---|---|---|---|---|
| Fang 2019 | Lyu 2020 | benefit | no evidence of effect | no | 0,323529412 | 11 | 34 | low (0.20-0.49) |
| Fang 2019 | Wen 2019 | benefit | no evidence of effect | no | 0,314285714 | 11 | 35 | low (0.20-0.49) |
| Annane 2019 | Son 2021 | benefit | benefit | yes | 0,31372549 | 16 | 51 | low (0.20-0.49) |
| Song Y 2024 | Wu 2020 | benefit | no evidence of effect | no | 0,307692308 | 4 | 13 | low (0.20-0.49) |
| Lyu 2020 | Rochwerg 2018 | no evidence of effect | no evidence of effect | yes | 0,305555556 | 11 | 36 | low (0.20-0.49) |
| Aletreby 2019 | Lin 2019 | no evidence of effect | benefit | no | 0,304347826 | 7 | 23 | low (0.20-0.49) |
| Lin 2019 | Xu 2018 | benefit | no evidence of effect | no | 0,304347826 | 7 | 23 | low (0.20-0.49) |
| Lu 2021 | Son 2021 | no evidence of effect | benefit | no | 0,3 | 6 | 20 | low (0.20-0.49) |
| Son 2021 | Xu 2018 | benefit | no evidence of effect | no | 0,3 | 6 | 20 | low (0.20-0.49) |
| Rochwerg 2018 | Wen 2019 | no evidence of effect | no evidence of effect | yes | 0,297297297 | 11 | 37 | low (0.20-0.49) |
| Siddiqi 2019 | Son 2021 | benefit | benefit | yes | 0,294117647 | 5 | 17 | low (0.20-0.49) |
| Siddiqi 2019 | Zhou 2018 | benefit | no evidence of effect | no | 0,294117647 | 5 | 17 | low (0.20-0.49) |
| Annane 2025 | Lin 2019 | benefit | benefit | yes | 0,291666667 | 21 | 72 | low (0.20-0.49) |
| Lin 2019 | Wu 2020 | benefit | no evidence of effect | no | 0,291666667 | 7 | 24 | low (0.20-0.49) |

| | | | | | | | | |
|---|---|---|---|---|---|---|---|---|
| Annane 2025 | Rygard 2018 | benefit | no evidence of effect | no | 0,287671233 | 21 | 73 | low (0.20-0.49) |
| Aletreby 2019 | Lu 2021 | no evidence of effect | no evidence of effect | yes | 0,285714286 | 4 | 14 | low (0.20-0.49) |
| Aletreby 2019 | Xu 2018 | no evidence of effect | no evidence of effect | yes | 0,285714286 | 4 | 14 | low (0.20-0.49) |
| Annane 2015 | Xu 2018 | no evidence of effect | no evidence of effect | yes | 0,285714286 | 8 | 28 | low (0.20-0.49) |
| Lu 2021 | Ni 2019 | no evidence of effect | benefit | no | 0,285714286 | 6 | 21 | low (0.20-0.49) |
| Son 2021 | Wu 2020 | benefit | no evidence of effect | no | 0,285714286 | 6 | 21 | low (0.20-0.49) |
| Lyu 2020 | Pitre 2024 | no evidence of effect | benefit | no | 0,282051282 | 11 | 39 | low (0.20-0.49) |
| Ni 2019 | Siddiqi 2019 | benefit | benefit | yes | 0,277777778 | 5 | 18 | low (0.20-0.49) |
| Liang 2021 | Lyu 2020 | no evidence of effect | no evidence of effect | yes | 0,275 | 11 | 40 | low (0.20-0.49) |
| Pitre 2024 | Wen 2019 | benefit | no evidence of effect | no | 0,275 | 11 | 40 | low (0.20-0.49) |
| Lian 2019 | Song Y 2024 | benefit | benefit | yes | 0,272727273 | 6 | 22 | low (0.20-0.49) |
| Lin 2019 | Song Y 2024 | benefit | benefit | yes | 0,272727273 | 6 | 22 | low (0.20-0.49) |
| Lu 2021 | Siddiqi 2019 | no evidence of effect | benefit | no | 0,272727273 | 3 | 11 | low (0.20-0.49) |
| Aletreby 2019 | Fang 2019 | no evidence of effect | benefit | no | 0,264705882 | 9 | 34 | low (0.20-0.49) |
| Son 2021 | Song Y 2024 | benefit | benefit | yes | 0,263157895 | 5 | 19 | low (0.20-0.49) |

| | | | | | | | | |
|---|---|---|---|---|---|---|---|---|
| Rygard 2018 | Song Y 2024 | no evidence of effect | benefit | no | 0,260869565 | 6 | 23 | low (0.20-0.49) |
| Fang 2019 | Wu 2020 | benefit | no evidence of effect | no | 0,257142857 | 9 | 35 | low (0.20-0.49) |
| Annane 2025 | Lian 2019 | benefit | benefit | yes | 0,256756757 | 19 | 74 | low (0.20-0.49) |
| Aletreby 2019 | Rochwerg 2018 | no evidence of effect | no evidence of effect | yes | 0,25 | 9 | 36 | low (0.20-0.49) |
| Annane 2025 | Ni 2019 | benefit | benefit | yes | 0,25 | 18 | 72 | low (0.20-0.49) |
| Lin 2019 | Lu 2021 | benefit | no evidence of effect | no | 0,25 | 6 | 24 | low (0.20-0.49) |
| Lu 2021 | Lyu 2020 | no evidence of effect | no evidence of effect | yes | 0,25 | 4 | 16 | low (0.20-0.49) |
| Siddiqi 2019 | Wu 2020 | benefit | no evidence of effect | no | 0,25 | 3 | 12 | low (0.20-0.49) |
| Rochwerg 2018 | Wu 2020 | no evidence of effect | no evidence of effect | yes | 0,243243243 | 9 | 37 | low (0.20-0.49) |
| Lu 2021 | Rygard 2018 | no evidence of effect | no evidence of effect | yes | 0,24 | 6 | 25 | low (0.20-0.49) |
| Lian 2019 | Siddiqi 2019 | benefit | benefit | yes | 0,238095238 | 5 | 21 | low (0.20-0.49) |
| Liang 2021 | Wen 2019 | no evidence of effect | no evidence of effect | yes | 0,238095238 | 10 | 42 | low (0.20-0.49) |
| Lin 2019 | Siddiqi 2019 | benefit | benefit | yes | 0,238095238 | 5 | 21 | low (0.20-0.49) |
| Annane 2025 | Zhou 2018 | benefit | no evidence of effect | no | 0,236111111 | 17 | 72 | low (0.20-0.49) |
| Lu 2021 | Wen 2019 | no evidence of effect | no evidence of effect | yes | 0,235294118 | 4 | 17 | low (0.20-0.49) |

| | | | | | | | | |
|---|---|---|---|---|---|---|---|---|
| Aletreby 2019 | Pitre 2024 | no evidence of effect | benefit | no | 0,230769231 | 9 | 39 | low (0.20-0.49) |
| Song Y 2024 | Xu 2018 | benefit | no evidence of effect | no | 0,230769231 | 3 | 13 | low (0.20-0.49) |
| Fang 2019 | Xu 2018 | benefit | no evidence of effect | no | 0,228571429 | 8 | 35 | low (0.20-0.49) |
| Rygard 2018 | Siddiqi 2019 | no evidence of effect | benefit | no | 0,227272727 | 5 | 22 | low (0.20-0.49) |
| Aletreby 2019 | Liang 2021 | no evidence of effect | no evidence of effect | yes | 0,225 | 9 | 40 | low (0.20-0.49) |
| Pitre 2024 | Wu 2020 | benefit | no evidence of effect | no | 0,225 | 9 | 40 | low (0.20-0.49) |
| Annane 2019 | Lyu 2020 | benefit | no evidence of effect | no | 0,22 | 11 | 50 | low (0.20-0.49) |
| Liang 2021 | Wu 2020 | no evidence of effect | no evidence of effect | yes | 0,219512195 | 9 | 41 | low (0.20-0.49) |
| Annane 2025 | Son 2021 | benefit | benefit | yes | 0,219178082 | 16 | 73 | low (0.20-0.49) |
| Annane 2015 | Wen 2019 | no evidence of effect | no evidence of effect | yes | 0,21875 | 7 | 32 | low (0.20-0.49) |
| Rochwerg 2018 | Xu 2018 | no evidence of effect | no evidence of effect | yes | 0,216216216 | 8 | 37 | low (0.20-0.49) |
| Annane 2019 | Wen 2019 | benefit | no evidence of effect | no | 0,215686275 | 11 | 51 | low (0.20-0.49) |
| Fang 2019 | Song Y 2024 | benefit | benefit | yes | 0,205882353 | 7 | 34 | low (0.20-0.49) |
| Annane 2015 | Lu 2021 | no evidence of effect | no evidence of effect | yes | 0,2 | 6 | 30 | low (0.20-0.49) |

| | | | | | | | | |
|---|---|---|---|---|---|---|---|---|
| Lu 2021 | Pitre 2024 | no evidence of effect | benefit | no | 0,2 | 8 | 40 | low (0.20-0.49) |
| Pitre 2024 | Xu 2018 | benefit | no evidence of effect | no | 0,2 | 8 | 40 | low (0.20-0.49) |
| Fang 2019 | Lu 2021 | benefit | no evidence of effect | no | 0,194444444 | 7 | 36 | very low (<0.20) |
| Rochwerg 2018 | Song Y 2024 | no evidence of effect | benefit | no | 0,194444444 | 7 | 36 | very low (<0.20) |
| Annane 2015 | Lyu 2020 | no evidence of effect | no evidence of effect | yes | 0,1875 | 6 | 32 | very low (<0.20) |
| Lu 2021 | Wu 2020 | no evidence of effect | no evidence of effect | yes | 0,1875 | 3 | 16 | very low (<0.20) |
| Lu 2021 | Rochwerg 2018 | no evidence of effect | no evidence of effect | yes | 0,184210526 | 7 | 38 | very low (<0.20) |
| Aletreby 2019 | Annane 2019 | no evidence of effect | benefit | no | 0,18 | 9 | 50 | very low (<0.20) |
| Pitre 2024 | Song Y 2024 | benefit | benefit | yes | 0,179487179 | 7 | 39 | very low (<0.20) |
| Annane 2019 | Wu 2020 | benefit | no evidence of effect | no | 0,176470588 | 9 | 51 | very low (<0.20) |
| Liang 2021 | Song Y 2024 | no evidence of effect | benefit | no | 0,175 | 7 | 40 | very low (<0.20) |
| Liang 2021 | Lu 2021 | no evidence of effect | no evidence of effect | yes | 0,166666667 | 7 | 42 | very low (<0.20) |
| Liang 2021 | Xu 2018 | no evidence of effect | no evidence of effect | yes | 0,166666667 | 7 | 42 | very low (<0.20) |

| | | | | | | | | |
|---|---|---|---|---|---|---|---|---|
| Siddiqi 2019 | Xu 2018 | benefit | no evidence of effect | no | 0,166666667 | 2 | 12 | very low (<0.20) |
| Annane 2019 | Lu 2021 | benefit | no evidence of effect | no | 0,156862745 | 8 | 51 | very low (<0.20) |
| Annane 2019 | Xu 2018 | benefit | no evidence of effect | no | 0,156862745 | 8 | 51 | very low (<0.20) |
| Annane 2015 | Wu 2020 | no evidence of effect | no evidence of effect | yes | 0,15625 | 5 | 32 | very low (<0.20) |
| Annane 2025 | Lyu 2020 | benefit | no evidence of effect | no | 0,152777778 | 11 | 72 | very low (<0.20) |
| Annane 2025 | Wen 2019 | benefit | no evidence of effect | no | 0,150684932 | 11 | 73 | very low (<0.20) |
| Fang 2019 | Siddiqi 2019 | benefit | benefit | yes | 0,147058824 | 5 | 34 | very low (<0.20) |
| Lu 2021 | Song Y 2024 | no evidence of effect | benefit | no | 0,142857143 | 2 | 14 | very low (<0.20) |
| Annane 2019 | Song Y 2024 | benefit | benefit | yes | 0,14 | 7 | 50 | very low (<0.20) |
| Rochwerg 2018 | Siddiqi 2019 | no evidence of effect | benefit | no | 0,138888889 | 5 | 36 | very low (<0.20) |
| Annane 2015 | Song Y 2024 | no evidence of effect | benefit | no | 0,133333333 | 4 | 30 | very low (<0.20) |
| Pitre 2024 | Siddiqi 2019 | benefit | benefit | yes | 0,128205128 | 5 | 39 | very low (<0.20) |
| Aletreby 2019 | Annane 2015 | no evidence of effect | no evidence of effect | yes | 0,125 | 4 | 32 | very low (<0.20) |
| Aletreby 2019 | Annane 2025 | no evidence of effect | benefit | no | 0,125 | 9 | 72 | very low (<0.20) |

| | | | | | | | | |
|---|---|---|---|---|---|---|---|---|
| Liang 2021 | Siddiqi 2019 | no evidence of effect | benefit | no | 0,125 | 5 | 40 | very low (<0.20) |
| Annane 2025 | Wu 2020 | benefit | no evidence of effect | no | 0,123287671 | 9 | 73 | very low (<0.20) |
| Annane 2025 | Lu 2021 | benefit | no evidence of effect | no | 0,109589041 | 8 | 73 | very low (<0.20) |
| Annane 2025 | Xu 2018 | benefit | no evidence of effect | no | 0,109589041 | 8 | 73 | very low (<0.20) |
| Annane 2019 | Siddiqi 2019 | benefit | benefit | yes | 0,1 | 5 | 50 | very low (<0.20) |
| Annane 2025 | Song Y 2024 | benefit | benefit | yes | 0,097222222 | 7 | 72 | very low (<0.20) |
| Annane 2025 | Siddiqi 2019 | benefit | benefit | yes | 0,069444444 | 5 | 72 | very low (<0.20) |
| Annane 2015 | Siddiqi 2019 | no evidence of effect | benefit | no | 0,066666667 | 2 | 30 | very low (<0.20) |
| Annane 2025 | Kato 2023 | benefit | no evidence of effect | no | 0,013513514 | 1 | 74 | very low (<0.20) |
| Aletreby 2019 | Kato 2023 | no evidence of effect | no evidence of effect | yes | 0 | 0 | 12 | very low (<0.20) |
| Annane 2015 | Kato 2023 | no evidence of effect | no evidence of effect | yes | 0 | 0 | 30 | very low (<0.20) |
| Annane 2019 | Kato 2023 | benefit | no evidence of effect | no | 0 | 0 | 53 | very low (<0.20) |
| Fang 2019 | Kato 2023 | benefit | no evidence of effect | no | 0 | 0 | 37 | very low (<0.20) |
| Kato 2023 | Lian 2019 | no evidence of effect | benefit | no | 0 | 0 | 24 | very low (<0.20) |

| | | | | | | | | |
|---|---|---|---|---|---|---|---|---|
| Kato 2023 | Liang 2021 | no evidence of effect | no evidence of effect | yes | 0 | 0 | 43 | very low (<0.20) |
| Kato 2023 | Lin 2019 | no evidence of effect | benefit | no | 0 | 0 | 24 | very low (<0.20) |
| Kato 2023 | Lu 2021 | no evidence of effect | no evidence of effect | yes | 0 | 0 | 12 | very low (<0.20) |
| Kato 2023 | Lyu 2020 | no evidence of effect | no evidence of effect | yes | 0 | 0 | 14 | very low (<0.20) |
| Kato 2023 | Ni 2019 | no evidence of effect | benefit | no | 0 | 0 | 21 | very low (<0.20) |
| Kato 2023 | Pitre 2024 | no evidence of effect | benefit | no | 0 | 0 | 42 | very low (<0.20) |
| Kato 2023 | Rochwerg 2018 | no evidence of effect | no evidence of effect | yes | 0 | 0 | 39 | very low (<0.20) |
| Kato 2023 | Rygard 2018 | no evidence of effect | no evidence of effect | yes | 0 | 0 | 25 | very low (<0.20) |
| Kato 2023 | Siddiqi 2019 | no evidence of effect | benefit | no | 0 | 0 | 8 | very low (<0.20) |
| Kato 2023 | Son 2021 | no evidence of effect | benefit | no | 0 | 0 | 20 | very low (<0.20) |
| Kato 2023 | Song Y 2024 | no evidence of effect | benefit | no | 0 | 0 | 10 | very low (<0.20) |
| Kato 2023 | Wen 2019 | no evidence of effect | no evidence of effect | yes | 0 | 0 | 15 | very low (<0.20) |

| | | | | | | | | |
|---|---|---|---|---|---|---|---|---|
| Kato 2023 | Wu 2020 | no evidence of effect | no evidence of effect | yes | 0 | 0 | 13 | very low (<0.20) |
| Kato 2023 | Xu 2018 | no evidence of effect | no evidence of effect | yes | 0 | 0 | 12 | very low (<0.20) |
| Kato 2023 | Zhou 2018 | no evidence of effect | no evidence of effect | yes | 0 | 0 | 20 | very low (<0.20) |

**Supplementary Table S21: Frequency of contribution of randomized controlled trials to meta-analysis across systematic reviews dependent on mortality conclusion addressing non-specific corticosteroid strategies in sepsis (n = 22)**

| RCT ID | N benefit | N no effect | Diff_benefit_minus_noeffect | Total contributions | % benefit | % effect |
|---|---|---|---|---|---|---|
| Yildiz 2002 | 8 | 3 | 5 | 11 | 72,7 | 27,3 |
| Yildiz 2011 | 7 | 3 | 4 | 10 | 70 | 30 |
| Annane 2018 | 10 | 7 | 3 | 17 | 58,8 | 41,2 |
| Gordon 2014 | 7 | 10 | -3 | 17 | 41,2 | 58,8 |
| Tongyoo 2016 | 8 | 5 | 3 | 13 | 61,5 | 38,5 |
| Keh 2016 | 7 | 4 | 3 | 11 | 63,6 | 36,4 |
| Confalonieri 2005 | 6 | 3 | 3 | 9 | 66,7 | 33,3 |
| Rinaldi 2006 | 6 | 3 | 3 | 9 | 66,7 | 33,3 |
| Sprung 2008 | 9 | 11 | -2 | 20 | 45 | 55 |
| Venkatesh 2018 | 10 | 8 | 2 | 18 | 55,6 | 44,4 |
| Gordon 2016 | 5 | 7 | -2 | 12 | 41,7 | 58,3 |
| Chawla 1999 | 6 | 4 | 2 | 10 | 60 | 40 |
| Bone 1987 | 5 | 3 | 2 | 8 | 62,5 | 37,5 |
| Meduri 2007 | 5 | 3 | 2 | 8 | 62,5 | 37,5 |
| Meijvis 2011 | 5 | 3 | 2 | 8 | 62,5 | 37,5 |
| Snijders 2010 | 5 | 3 | 2 | 8 | 62,5 | 37,5 |
| Doluee 2018 | 4 | 2 | 2 | 6 | 66,7 | 33,3 |
| Liu 2012 | 4 | 2 | 2 | 6 | 66,7 | 33,3 |
| Huang 2014 | 3 | 1 | 2 | 4 | 75 | 25 |
| Menon 2017 | 3 | 1 | 2 | 4 | 75 | 25 |
| Agarwal 2022 | 2 | 0 | 2 | 2 | 100 | 0 |
| Birudaraju 2022 | 2 | 0 | 2 | 2 | 100 | 0 |
| El Ghamrawy 2006 | 2 | 0 | 2 | 2 | 100 | 0 |
| Fernandez Serrano 2011 | 2 | 0 | 2 | 2 | 100 | 0 |
| Li 2016 | 2 | 0 | 2 | 2 | 100 | 0 |

| | | | | | | |
|---|---|---|---|---|---|---|
| McHardy 1972 | 2 | 0 | 2 | 2 | 100 | 0 |
| Nafae 2013 | 2 | 0 | 2 | 2 | 100 | 0 |
| Nagy 2013 | 2 | 0 | 2 | 2 | 100 | 0 |
| Slusher 1996 | 2 | 0 | 2 | 2 | 100 | 0 |
| Tagaro 2017 | 2 | 0 | 2 | 2 | 100 | 0 |
| Valoor 2009 | 2 | 0 | 2 | 2 | 100 | 0 |
| Wan 2014 | 0 | 2 | -2 | 2 | 0 | 100 |
| Lv 2017 | 10 | 9 | 1 | 19 | 52,6 | 47,4 |
| Arabi 2010 | 8 | 9 | -1 | 17 | 47,1 | 52,9 |
| Bollaert 1998 | 8 | 9 | -1 | 17 | 47,1 | 52,9 |
| Oppert 2005 | 8 | 9 | -1 | 17 | 47,1 | 52,9 |
| Cicarelli 2007 | 8 | 7 | 1 | 15 | 53,3 | 46,7 |
| Sprung 1984 | 5 | 4 | 1 | 9 | 55,6 | 44,4 |
| Tandan 2005 | 5 | 4 | 1 | 9 | 55,6 | 44,4 |
| Meduri 2009 | 4 | 3 | 1 | 7 | 57,1 | 42,9 |
| Rezk 2013 | 4 | 3 | 1 | 7 | 57,1 | 42,9 |
| Sabry 2011 | 4 | 3 | 1 | 7 | 57,1 | 42,9 |
| VASSCSG 1987 | 4 | 3 | 1 | 7 | 57,1 | 42,9 |
| Blum 2015 | 2 | 1 | 1 | 3 | 66,7 | 33,3 |
| Kurungundla 2008 | 2 | 1 | 1 | 3 | 66,7 | 33,3 |
| Mirea 2014 | 2 | 1 | 1 | 3 | 66,7 | 33,3 |
| Angus 2020 | 1 | 0 | 1 | 1 | 100 | 0 |
| Briegel 2001 | 1 | 0 | 1 | 1 | 100 | 0 |
| CSG 1963 | 0 | 1 | -1 | 1 | 0 | 100 |
| Cicarelli 2006 | 0 | 1 | -1 | 1 | 0 | 100 |
| Corral Gudino 2021 | 1 | 0 | 1 | 1 | 100 | 0 |
| Dequin 2020 | 1 | 0 | 1 | 1 | 100 | 0 |
| Dequin 2023 | 1 | 0 | 1 | 1 | 100 | 0 |
| Fuji 2020 | 0 | 1 | -1 | 1 | 0 | 100 |

| | | | | | | |
|---|---|---|---|---|---|---|
| Ghanei 2021 | 1 | 0 | 1 | 1 | 100 | 0 |
| Horby 2021 | 1 | 0 | 1 | 1 | 100 | 0 |
| Hussein 2021 | 0 | 1 | -1 | 1 | 0 | 100 |
| Iglesias 2020 | 1 | 0 | 1 | 1 | 100 | 0 |
| Keh 2003 | 0 | 1 | -1 | 1 | 0 | 100 |
| Lyu 2022 | 1 | 0 | 1 | 1 | 100 | 0 |
| Meduri 2022 | 1 | 0 | 1 | 1 | 100 | 0 |
| Mohamed 2020 | 1 | 0 | 1 | 1 | 100 | 0 |
| Mohamed 2023 | 1 | 0 | 1 | 1 | 100 | 0 |
| Moskowitz 2020 | 1 | 0 | 1 | 1 | 100 | 0 |
| Pierre de Oliveira 2023 | 1 | 0 | 1 | 1 | 100 | 0 |
| Sai 2021 | 1 | 0 | 1 | 1 | 100 | 0 |
| Sevransky 2021 | 1 | 0 | 1 | 1 | 100 | 0 |
| Villar 2020 | 1 | 0 | 1 | 1 | 100 | 0 |
| Wani 2020 | 1 | 0 | 1 | 1 | 100 | 0 |
| Wittermans 2020 | 1 | 0 | 1 | 1 | 100 | 0 |
| Annane 2002 | 10 | 10 | 0 | 20 | 50 | 50 |
| Briegel 1999 | 8 | 8 | 0 | 16 | 50 | 50 |
| Hu 2009 | 5 | 5 | 0 | 10 | 50 | 50 |
| Luce 1988 | 5 | 5 | 0 | 10 | 50 | 50 |
| Schumer 1976 | 5 | 5 | 0 | 10 | 50 | 50 |
| Torres 2015 | 4 | 4 | 0 | 8 | 50 | 50 |
| Aboab 2008 | 3 | 3 | 0 | 6 | 50 | 50 |
| Chang 2020 | 1 | 1 | 0 | 2 | 50 | 50 |
| El Nawawy 2017 | 1 | 1 | 0 | 2 | 50 | 50 |
| Klastersky 1971 | 1 | 1 | 0 | 2 | 50 | 50 |
| Mussack 2005 | 1 | 1 | 0 | 2 | 50 | 50 |
| Povoa 2015 | 1 | 1 | 0 | 2 | 50 | 50 |
| Tomazini 2020 | 1 | 1 | 0 | 2 | 50 | 50 |

#### Supplementary Table S22: Distribution of systematic reviews by population definition (sepsis ± shock vs septic shock only) and effect direction (n = 22)

| Population target group | Effect category | n | percent |
|---|---|---|---|
| Sepsis±Shock | benefit | 8 | 61,5 |
| Sepsis±Shock | no evidence of effect | 5 | 38,5 |
| ShockOnly | benefit | 2 | 22,2 |
| ShockOnly | no evidence of effect | 7 | 77,8 |

| Comparison | group1 | group2 | Benefit group1 | No_effect group1 | Benefit group2 | No_effect group2 | P value | Odds ratio | conf_low | conf_high |
|---|---|---|---|---|---|---|---|---|---|---|
| Effect direction by population group | Sepsis±Shock | ShockOnly | 8 | 5 | 2 | 7 | 0,099071207 | 5,146228898 | 0,624331001 | 70,89448038 |

**Supplementary Table S23: Proportion of systematic reviews including at least one randomized controlled trial classified as likely incompatible or incompatible according to mortality effect direction (benefit vs no evidence of effect) (n = 22)**

| Effect category | (likely) incompatible included in meta | n | Total effect category | percent |
|---|---|---|---|---|
| benefit | no | 5 | 10 | 50 |
| benefit | yes | 5 | 10 | 50 |
| no evidence of effect | no | 8 | 12 | 66,66667 |
| no evidence of effect | yes | 4 | 12 | 33,33333 |

| Comparison | group1 | group2 | Benefit group1 | No_effect group1 | Benefit group2 | No_effect group2 | P value | Odds ratio | conf_low | conf_high |
|---|---|---|---|---|---|---|---|---|---|---|
| At least one likely incompatible/incompatible RCT in mortality meta-analysis: yes vs no | Sepsis±Shock | ShockOnly | 5 | 8 | 5 | 4 | 0,665634675 | 1,936521333 | 0,263752588 | 15,60367287 |

### Supplementary Table S24: Association between methodological and design characteristics and mortality conclusions in systematic reviews addressing non-specific broad concepts of corticosteroid strategies in sepsis (n = 22)

**Distribution of effect category according to mortality time point as defined by the SR (p = 0,377708978):**

| Time category | benefit | no evidence of effect |
|---|---|---|
| 28-d | 5 | 8 |
| 28-d-mixed | 5 | 3 |
| 90-d | 0 | 1 |

**Distribution of effect category according to meta-analytic model used by the SR (p = 1,0):**

| Model | benefit | no evidence of effect |
|---|---|---|
| Fixed | 4 | 5 |
| Random | 6 | 7 |

**Distribution of effect category according to effect measure used by the SR (p = 1,0):**

| Effect measure | benefit | no evidence of effect |
|---|---|---|
| OR | 1 | 2 |
| RR | 9 | 10 |

**Distribution of effect category according to GRADE certainty of evidence of short-term mortality result by the SR (p = 0,47468):**

| GRADE | benefit | no evidence of effect |
|---|---|---|
| High | 1 | 1 |
| Low | 0 | 2 |
| Moderate | 5 | 2 |
| NR | 4 | 6 |

| | | |
|---|---|---|
| Very low | 0 | 1 |

**Distribution of effect category according to AMSTAR 2 rating of the SR (p = 0,571428571):**

| AMSTAR category | benefit | no evidence of effect |
|---|---|---|
| critically low | 8 | 11 |
| low | 2 | 1 |

**Distribution of effect category according to search date (year) of the SR (p = 0,179879049):**

| Effect category | median_year | IQR_low | IQR_high | mean_year |
|---|---|---|---|---|
| benefit | 2019 | 2018 | 2020 | 2019,556 |
| no evidence of effect | 2018 | 2018 | 2019 | 2018,364 |

**Supplementary Table S25: Sensitivity analysis of the short-term meta-analysis of Annane et al. 2025 Cochrane review according to RCT reclassification (Sepsis-3)**

| Study (RCT) | Publication year | Event treat | N treat | Event control | N control | Subgroup defined by Ananne et al. | Reclassification of RCT according to Sepsis-3 definition |
|---|---|---|---|---|---|---|---|
| Aboab 2008 | 2008 | 3 | 10 | 7 | 13 | Septic shock only | unclear |
| Agarwal 2022 | 2022 | 22 | 61 | 25 | 59 | Septic shock only | Sepsis-3 compatible |
| Angus 2020 | 2020 | 55 | 278 | 27 | 101 | COVID-19 | Sepsis-3 compatible |
| Annane 2002 | 2002 | 82 | 151 | 91 | 149 | Septic shock only | Sepsis-3 compatible |
| Annane 2018 | 2018 | 207 | 614 | 244 | 627 | Septic shock only | Sepsis-3 compatible |
| Arabi 2011 | 2011 | 33 | 39 | 26 | 36 | Septic shock only | unclear |
| Blum 2015 | 2015 | 16 | 402 | 13 | 400 | Sepsis and community-acquired pneumonia | likely incompatible |
| Bollaert 1998 | 1998 | 7 | 22 | 12 | 19 | Septic shock only | Sepsis-3 compatible |
| Bone 1987 | 1987 | 65 | 191 | 48 | 190 | Sepsis | unclear |
| Briegel 1999 | 1999 | 3 | 20 | 4 | 20 | Septic shock only | Sepsis-3 compatible |
| Chang 2020 | 2020 | 11 | 40 | 14 | 40 | Sepsis | Sepsis-3 compatible |
| Chawla 1999 | 1999 | 6 | 23 | 10 | 21 | Septic shock only | unclear |
| Cicarelli 2007 | 2007 | 7 | 14 | 12 | 15 | Septic shock only | unclear |
| Confalonieri 2005 | 2005 | 0 | 23 | 6 | 23 | Sepsis and community-acquired pneumonia | likely compatible |
| Corral-Gudino 2021 | 2021 | 7 | 35 | 5 | 29 | COVID-19 | likely incompatible |
| Dequin 2020 | 2020 | 11 | 76 | 20 | 73 | COVID-19 | Sepsis-3 compatible |
| Dequin 2023 | 2023 | 25 | 400 | 47 | 395 | Sepsis and community-acquired pneumonia | unclear |
| Doluee 2018 | 2018 | 54 | 80 | 58 | 80 | Septic shock only | unclear |
| El Ghamrawy 2006 | 2006 | 3 | 17 | 6 | 17 | Sepsis and community-acquired pneumonia | no fulltext |
| Fernández-Serrano 2011 | 2011 | 1 | 28 | 1 | 28 | Sepsis and community-acquired pneumonia | likely compatible |
| Ghanei 2021 | 2021 | 4 | 116 | 12 | 220 | COVID-19 | likely incompatible |
| Gordon 2014 | 2014 | 7 | 31 | 7 | 30 | Septic shock only | Sepsis-3 compatible |

| | | | | | | | |
|---|---|---|---|---|---|---|---|
| Gordon 2016 | 2016 | 62 | 208 | 57 | 213 | Septic shock only | Sepsis-3 compatible |
| Horby 2021 | 2021 | 482 | 2104 | 1110 | 4321 | COVID-19 | likely incompatible |
| Hu 2009 | 2009 | 4 | 38 | 6 | 39 | Septic shock only | no fulltext |
| Huang 2014 | 2014 | 4 | 20 | 7 | 20 | Sepsis | no fulltext |
| Iglesias 2020 | 2020 | 26 | 68 | 14 | 69 | Sepsis | Sepsis-3 compatible |
| Keh 2016 | 2016 | 15 | 171 | 14 | 170 | Sepsis | Sepsis-3 compatible |
| Kurungundla 2008 | 2008 | 4 | 11 | 6 | 10 | Septic shock only | no fulltext |
| Li 2016 | 2016 | 3 | 29 | 3 | 29 | Sepsis and community-acquired pneumonia | no fulltext |
| Liu 2012 | 2012 | 5 | 12 | 10 | 14 | Sepsis and ARDS | unclear |
| Luce 1988 | 1988 | 22 | 38 | 20 | 37 | Sepsis | likely compatible |
| Lv 2017 | 2017 | 23 | 58 | 19 | 60 | Septic shock only | Sepsis-3 compatible |
| Lyu 2022 | 2022 | 79 | 213 | 77 | 213 | Septic shock only | Sepsis-3 compatible |
| McHardy 1972 | 1972 | 3 | 40 | 9 | 86 | Sepsis and community-acquired pneumonia | likely incompatible |
| Meduri 2007 | 2007 | 10 | 42 | 8 | 19 | Sepsis and ARDS | likely incompatible |
| Meduri 2009 | 2009 | 22 | 48 | 4 | 31 | Sepsis | unclear |
| Meduri 2022 | 2022 | 30 | 297 | 34 | 287 | Sepsis and community-acquired pneumonia | likely compatible |
| Meijvis 2011 | 2011 | 9 | 151 | 11 | 153 | Sepsis and community-acquired pneumonia | likely incompatible |
| Menon 2017 | 2017 | 1 | 23 | 3 | 26 | Septic shock only | unclear |
| Mirea 2014 | 2014 | 44 | 117 | 22 | 54 | Septic shock only | unclear |
| Mohamed 2020 | 2020 | 18 | 45 | 19 | 45 | Septic shock only | Sepsis-3 compatible |
| Mohamed 2023 | 2023 | 13 | 53 | 20 | 53 | Septic shock only | Sepsis-3 compatible |
| Moskowitz 2020 | 2020 | 35 | 101 | 29 | 99 | Septic shock only | Sepsis-3 compatible |
| Nafae 2013 | 2013 | 4 | 60 | 6 | 20 | Sepsis and community-acquired pneumonia | likely incompatible |
| Nagy 2013 | 2013 | 0 | 29 | 0 | 30 | Sepsis and community-acquired pneumonia | likely incompatible |
| Oppert 2005 | 2005 | 9 | 23 | 11 | 25 | Septic shock only | Sepsis-3 compatible |

| | | | | | | | |
|---|---|---|---|---|---|---|---|
| Pierre de Oliveira 2023 | 2023 | 14 | 58 | 16 | 54 | COVID-19 | Sepsis-3 compatible |
| Rezk 2013 | 2013 | 0 | 18 | 3 | 9 | Sepsis and ARDS | likely incompatible |
| Rinaldi 2006 | 2006 | 6 | 26 | 7 | 26 | Sepsis | likely compatible |
| Sabry 2011 | 2011 | 2 | 40 | 6 | 40 | Sepsis and community-acquired pneumonia | likely compatible |
| Sai 2021 | 2021 | 41 | 75 | 45 | 75 | Sepsis | unclear |
| Schumer 1976 | 1976 | 9 | 86 | 33 | 86 | Septic shock only | unclear |
| Sevransky 2021 | 2021 | 56 | 252 | 60 | 249 | Septic shock only | Sepsis-3 compatible |
| Slusher 1996 | 1996 | 6 | 36 | 4 | 36 | Sepsis | unclear |
| Snijders 2010 | 2010 | 6 | 104 | 6 | 109 | Sepsis and community-acquired pneumonia | likely incompatible |
| Sprung 1984 | 1984 | 33 | 43 | 11 | 16 | Septic shock only | likely compatible |
| Sprung 2008 | 2008 | 86 | 251 | 78 | 248 | Septic shock only | Sepsis-3 compatible |
| Tagaro 2017 | 2017 | 0 | 30 | 1 | 30 | Sepsis and community-acquired pneumonia | likely incompatible |
| Tandan 2005 | 2005 | 11 | 14 | 13 | 14 | Septic shock only | no fulltext |
| Tomazini 2020 | 2020 | 85 | 151 | 91 | 148 | COVID-19 | Sepsis-3 compatible |
| Tongyoo 2016 | 2016 | 22 | 98 | 27 | 99 | Sepsis and ARDS | Sepsis-3 compatible |
| Torres 2015 | 2015 | 6 | 59 | 9 | 61 | Sepsis and community-acquired pneumonia | unclear |
| Valoor 2009 | 2009 | 7 | 19 | 6 | 19 | Septic shock only | likely compatible |
| VASSCSG 1987 | 1987 | 23 | 112 | 24 | 111 | Sepsis | unclear |
| Venkatesh 2018 | 2018 | 410 | 1853 | 448 | 1860 | Septic shock only | Sepsis-3 compatible |
| Villar 2020 | 2020 | 14 | 96 | 29 | 89 | Sepsis and ARDS | likely incompatible |
| Wani 2020 | 2020 | 20 | 50 | 21 | 50 | Sepsis | Sepsis-3 compatible |
| Wittermans 2021 | 2021 | 4 | 203 | 7 | 198 | Sepsis and community-acquired pneumonia | likely incompatible |
| Yildiz 2002 | 2002 | 8 | 20 | 12 | 20 | Sepsis | unclear |
| Yildiz 2011 | 2011 | 16 | 27 | 15 | 28 | Sepsis | unclear |

**Supplementary Table S26: Sensitivity analysis of the short-term meta-analysis of Pitre et al. 2024 according to RCT reclassification (Sepsis-3)**

| Study (RCT) | Publication year | Event treat | N treat | Event control | N control | Reclassification of RCT according to Sepsis-3 definition |
|---|---|---|---|---|---|---|
| Schumer 1976 | 1976 | 9 | 86 | 33 | 86 | unclear |
| Sprung 1984 | 1984 | 33 | 43 | 11 | 16 | likely compatible |
| Bone 1987 | 1987 | 65 | 191 | 48 | 190 | unclear |
| VASSCSG 1987 | 1987 | 23 | 112 | 24 | 111 | unclear |
| Luce 1988 | 1988 | 22 | 38 | 20 | 37 | likely compatible |
| Bollaert 1998 | 1998 | 7 | 22 | 12 | 19 | Sepsis-3 compatible |
| Briegel 1999 | 1999 | 3 | 20 | 4 | 20 | Sepsis-3 compatible |
| Chawla 1999 | 1999 | 6 | 23 | 10 | 21 | unclear |
| Annane 2002 | 2002 | 82 | 151 | 91 | 149 | Sepsis-3 compatible |
| Yildiz 2002 | 2002 | 8 | 20 | 12 | 20 | unclear |
| Confalonieri 2005 | 2005 | 0 | 23 | 6 | 23 | likely compatible |
| Oppert 2005 | 2005 | 10 | 23 | 11 | 25 | Sepsis-3 compatible |
| Tandan 2005 | 2005 | 11 | 14 | 13 | 14 | no fulltext |
| Rinaldi 2006 | 2006 | 6 | 26 | 7 | 26 | likely compatible |
| Cicarelli 2007 | 2007 | 7 | 14 | 12 | 15 | unclear |
| Meduri 2007 | 2007 | 10 | 42 | 8 | 19 | likely incompatible |
| Sprung 2008 | 2008 | 86 | 251 | 78 | 248 | Sepsis-3 compatible |
| Hu 2009 | 2009 | 4 | 38 | 6 | 39 | no fulltext |
| Meduri 2009 | 2009 | 22 | 48 | 4 | 31 | unclear |
| Snijders 2010 | 2010 | 6 | 104 | 6 | 109 | likely incompatible |
| Arabi 2011 | 2011 | 33 | 39 | 26 | 36 | unclear |
| Meijvis 2011 | 2011 | 9 | 151 | 11 | 153 | likely incompatible |
| Sabry 2011 | 2011 | 2 | 40 | 6 | 40 | likely compatible |
| Yildiz 2011 | 2011 | 16 | 27 | 15 | 28 | unclear |

| | | | | | | |
|---|---|---|---|---|---|---|
| Liu 2012 | 2012 | 3 | 12 | 6 | 14 | unclear |
| Rezk 2013 | 2013 | 0 | 18 | 3 | 9 | likely incompatible |
| Gordon 2014 | 2014 | 7 | 31 | 7 | 30 | Sepsis-3 compatible |
| Torres 2015 | 2015 | 6 | 59 | 9 | 61 | unclear |
| El-Nawawy 2016 | 2016 | 14 | 32 | 26 | 64 | unclear |
| Gordon 2016 | 2016 | 62 | 201 | 57 | 207 | Sepsis-3 compatible |
| Keh 2016 | 2016 | 15 | 171 | 14 | 170 | Sepsis-3 compatible |
| Tongyoo 2016 | 2016 | 22 | 98 | 27 | 99 | Sepsis-3 compatible |
| Lv 2017 | 2017 | 23 | 58 | 19 | 60 | Sepsis-3 compatible |
| Menon 2017 | 2017 | 1 | 23 | 3 | 26 | unclear |
| Annane 2018 | 2018 | 207 | 614 | 244 | 627 | Sepsis-3 compatible |
| Doluee 2018 | 2018 | 54 | 80 | 58 | 80 | unclear |
| Venkatesh 2018 | 2018 | 410 | 1841 | 448 | 1840 | Sepsis-3 compatible |
| Agarwal 2022 | 2022 | 22 | 61 | 25 | 59 | Sepsis-3 compatible |
| Briudaraju 2022 | 2022 | 9 | 21 | 16 | 22 | unclear |
